# Predictive performance of international COVID-19 mortality forecasting models

**DOI:** 10.1101/2020.07.13.20151233

**Authors:** Joseph Friedman, Patrick Liu, Christopher E. Troeger, Austin Carter, Robert C. Reiner, Ryan M. Barber, James Collins, Stephen S. Lim, David M. Pigott, Theo Vos, Simon I. Hay, Christopher J.L. Murray, Emmanuela Gakidou

**Affiliations:** University of California, Los Angeles; Institute For Health Metrics and Evaluation

## Abstract

Forecasts and alternative scenarios of COVID-19 mortality have been critical inputs into a range of policies and decision-makers need information about predictive performance. We identified n=386 public COVID-19 forecasting models and included n=8 that were global in scope and provided public, date-versioned forecasts. For each, we examined the median absolute percent error (MAPE) compared to subsequently observed mortality trends, stratified by weeks of extrapolation, world region, and month of model estimation. Models were also assessed for ability to predict the timing of peak daily mortality. The MAPE among models released in July rose from 1.8% at one week of extrapolation to 24.6% at twelve weeks. The MAPE at six weeks were the highest in Sub-Saharan Africa (34.8%), and the lowest in high-income countries (6.3%). At the global level, several models had about 10% MAPE at six weeks, showing surprisingly good performance despite the complexities of modelling human behavioural responses and government interventions. The framework and publicly available codebase presented here (https://github.com/pyliu47/covidcompare) can be routinely used to compare predictions and evaluate predictive performance in an ongoing fashion.

## Introduction

Forecasts and alternative scenarios of COVID-19 have been critical inputs into a range of important decisions by healthcare providers, local and national government agencies and international organizations and actors^1–4^. For example, hospitals need to prepare for potential surges in the demand for hospital beds, ICU beds and ventilators^1^. National critical response agencies such as the US Federal Emergency Management Agency have scarce resources including ventilators that can be moved to locations in need with sufficient notice^5,6^. Longer range forecasts are important for decisions such as the potential to open schools, universities and workplaces, and under what circumstances^7^. Much longer-range forecasts—six months to a year—are important for a wide range of policy choices, where efforts to reduce disease transmission must be balanced against economic outcomes such as unemployment and poverty^8^. Furthermore, vaccine and new therapeutic trialists need to select locations that will have sufficient transmission to test new products in the time frame when phase three clinical trials are ready to be launched. Nevertheless, hundreds of forecasting models have been published and/or publicly released, and it is often not immediately clear which models have had the best performance, or are most appropriate for predicting a given aspect of the pandemic.

Existing COVID-19 forecasting models differ substantially in methodology, assumptions, range of predictions, and quantities estimated. Furthermore, mortality forecasts for the same location have often differed substantially, in many cases by more than an order of magnitude, even within a six-week forecasting window. The challenge for decision-makers seeking input from models to guide decisions, which can impact many thousands of lives, is therefore not the availability of forecasts, but guidance on which forecasts are likely to be most accurate. Out-of-sample predictive validation—checking how well past versions of forecasting models predict subsequently observed trends—provides insight into future model performance^9^. Although some comparisons have been conducted for models describing the epidemic in the United States^10–13^, to our knowledge similar analyses have not been undertaken for models covering multiple countries, despite the growing global impact of COVID-19.

This paper introduces a publicly available dataset and evaluation framework (https://github.com/pyliu47/covidcompare) for assessing the predictive validity of COVID-19 mortality forecasts. The framework and associated open-access software can be routinely used to track model performance. This will, overtime, serve as a reference for decision-makers on historical model performance, and provide insight into which models should be considered for critical decisions in the future.

## Results

Eight models which fit all inclusion criteria were evaluated (Table 1). These included those modelled by: DELPHI-MIT (Delphi)^14,15^, Youyang Gu (YYG)^10^, the Los Alamos National Laboratory (LANL)^16^, Imperial College London (Imperial)^17^,the SIKJ-Alpha model from the USC Data Science Lab (SIKJalpha)^18^, and three models produced by the Institute for Health Metrics and Evaluation (IHME)^19^ (see methods section for more details). Results are presented in the main text for two main predictive tasks: 1) predicting the magnitude of mortality, and 2) predicting the timing of peak mortality (see methods). Magnitude results are presented in the main text for models that continued to produce forecasts at the time of publication of this article, while peak timing results are presented for models released early enough to capture the first peak in most locations. Results for all historical models are shown in the appendix. Magnitude of mortality results in the main text are presented according to two main analytical approaches. In the “most current” approach, used to select data shown in Figure 3, the most recent 4-week period allowing for the calculation of errors is selected for each extrapolation length. In the “month stratified” approach, used to select data for Figures 4 and 5, models from July were used to calculate errors at each length of extrapolation, with all months shown in the appendix. In each case errors were assessed from one to twelve weeks of forecasting (see methods and Figure 2 for more details).

**Table 1.**
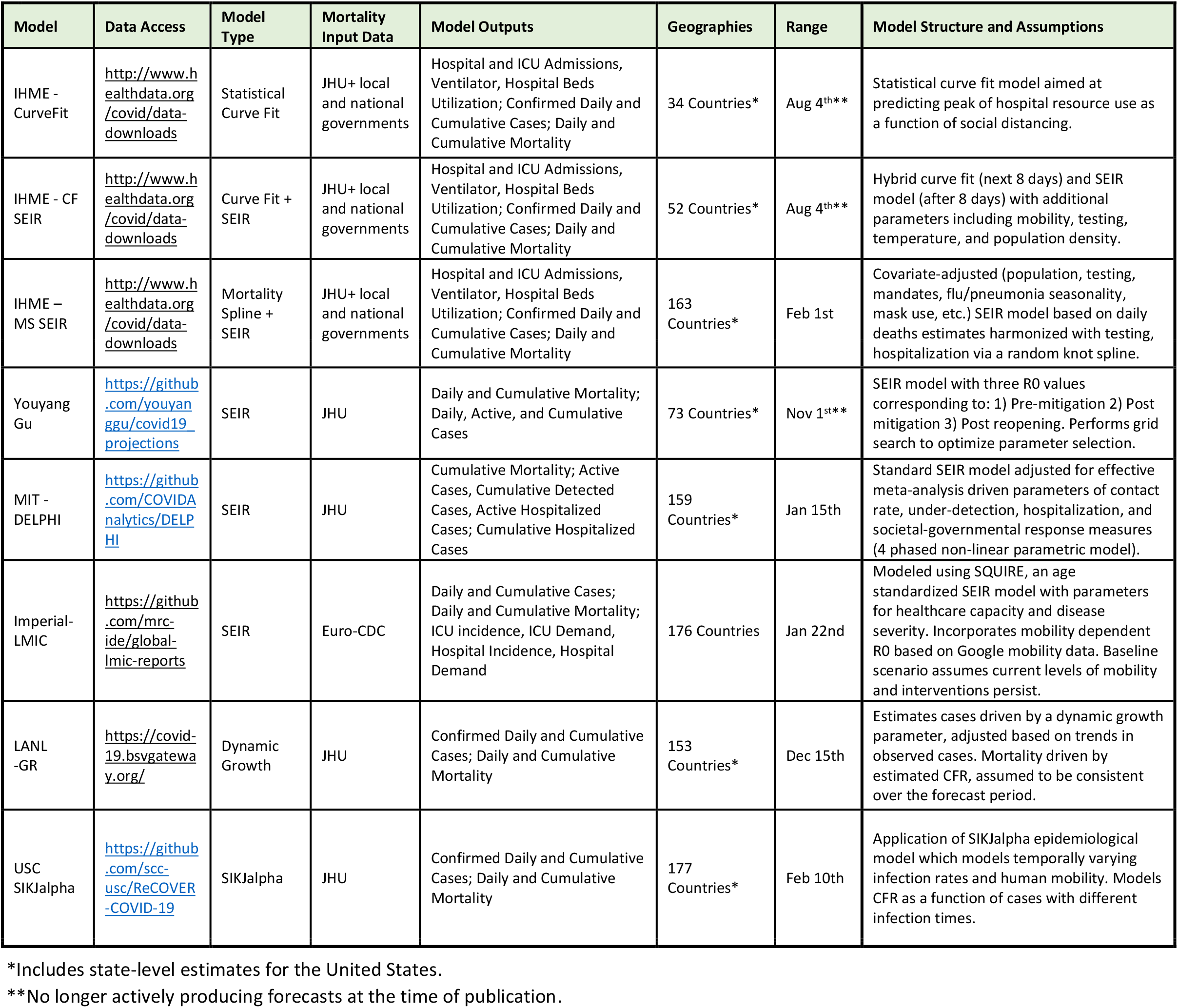
Models Included in the Study. All eight models included in the study are shown. The full list of models assessed for inclusion is shown in the supplemental review file. Range indicates the last date upon which forecasts are available in the most current version of each model.

The evaluation framework developed here for assessing how well models predicted the total number of cumulative deaths is shown in Figure 1 for an example country—the United States—and similar figures for all locations included in the study can be found in the appendix. Figure 1, and similar figures in the appendix, also highlight the direction of error for each model in each location. When looking across iterations of forecasts, a wide range of variation can be observed for nearly all of the models. Nevertheless, in many locations, models largely reached consensus regarding trajectories in the summer of 2020. Models diverged again when predicting trajectories for Fall 2020 and Winter 2021, as some models predicted upticks related to seasonality, while others projected continued slow declines in mortality.

**Figure 1.**
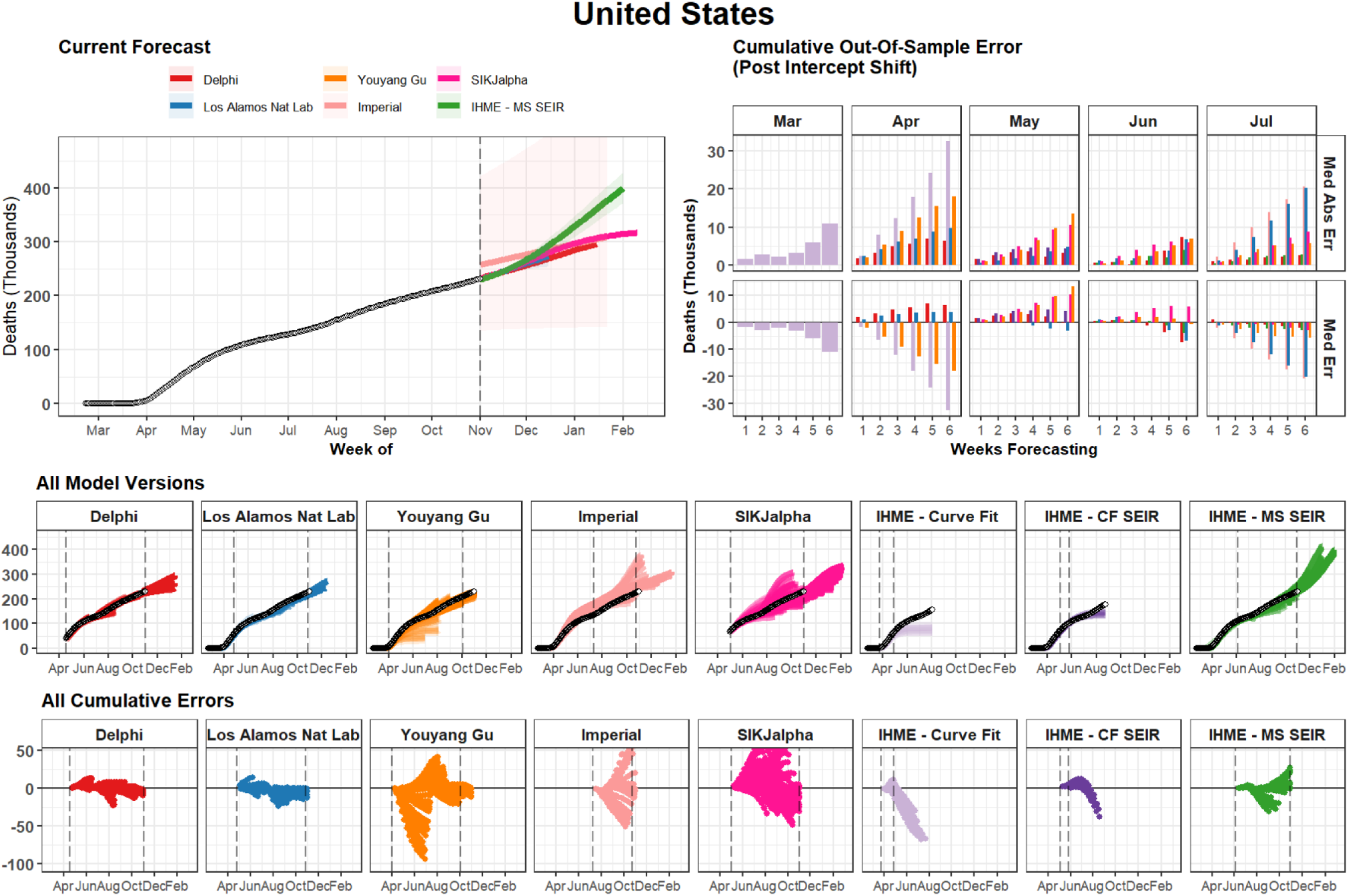
Cumulative Mortality Forecasts and Prediction Errors by Model – Example for United States. The most recent version of each model is shown on the top left. The middle row shows all iterations of each model as separate lines, with the intensity of color indicating model date (darker models are more recent). The vertical dashed lines indicate the first and last model release date for each model. The bottom row shows all errors calculated at weekly intervals. The top right panel summarizes all observed errors, using median error and median absolute error, by weeks of forecasting, and month of model estimation. Errors incorporate an intercept shift to account for differences in each model’s input data. This figure represents an example for the United States of country-specific plots made for all locations examined in this study. Graphs for all geographies can be found in the supplement. Note that while certain model uses different input data source than the other modelling groups causing apparently discordant past trends in the top left panel. We plot raw estimates on the top left panel, however we implement an intercept shift to account for this issue in the calculation of errors.

Figure 2 highlights the most recent errors for each length of extrapolation. For all models, the most recent 1-week errors, reflecting forecasts created in October, ranged from 1% to 2%. The 12-week median absolute percent errors (MAPE), reflecting models produced in July and August, ranged from 22.4% for the SIK-J Alpha model, to 79.9% for the Imperial model. At the global level pooling across models, the most recent 6-week MAPE value was 7.2%.

**Figure 2.**
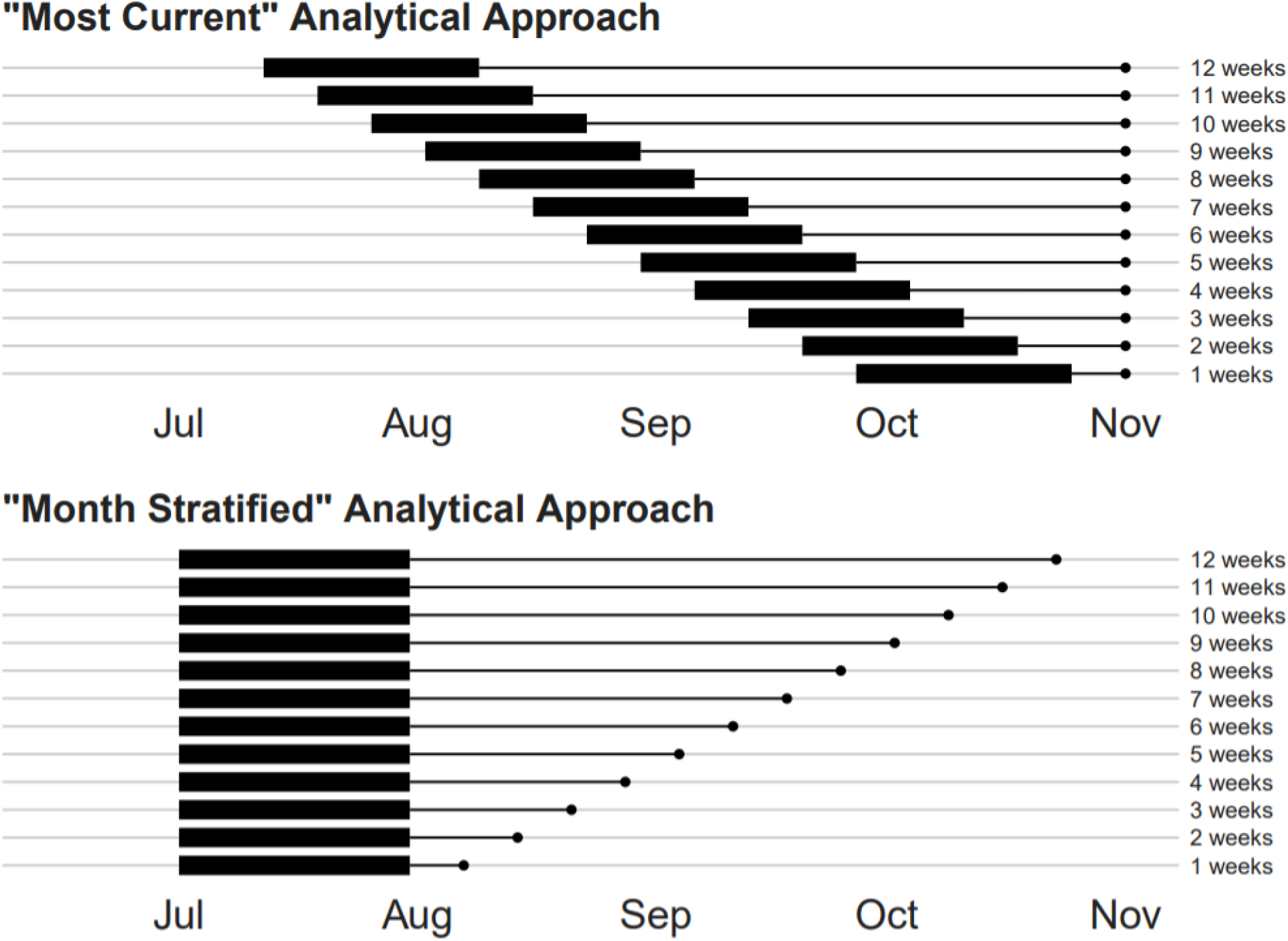
Illustration of Analytical Framework. This figure highlights the analytical framework presented in the main text. Part A highlights the “most current” approach, which is used to select the data shown in Figure 3. Part B highlights the “month stratified” approach used for Figures 4 and 5. The Y axis shows the number of weeks of extrapolation for each scenario, while the x axis shows a range of model date—the date on which a model was released. The thick band in each plot highlights the 4-week window of model dates used for each extrapolation week value. The thin line shows the period for which each set of models is extrapolating before errors are calculated. In the top panel, the most recent four weeks of model dates are used for each extrapolation length. Therefore, for 1-week errors models from October were used, whereas for 12-week errors, models from July and August were used. In the bottom panel, models from July are used in all cases. The analytic strategy highlighted in the top panel provides the most recent evidence possible for each extrapolation length. The strategy in the bottom allows for more reliable assessment of how errors grow with increased extrapolation time.

**Figure 3.**
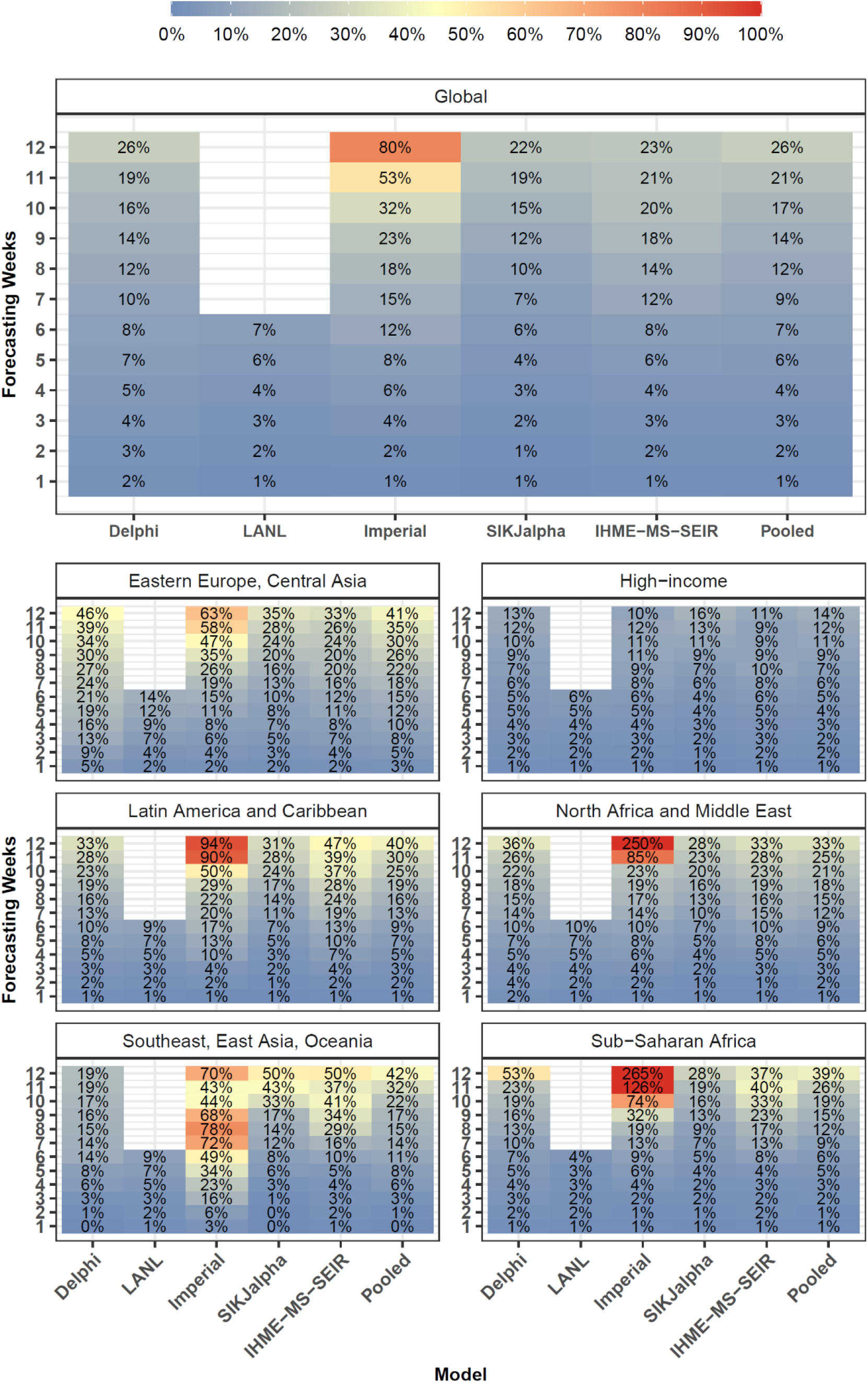
Most Current - Cumulative Mortality Accuracy – Median Absolute Percent Error. Median absolute percent error values, a measure of accuracy, were calculated across all observed errors at weekly intervals, for each model by weeks of forecasting and geographic region. Values that represent fewer than five locations are masked due to small sample size. Models were included in the global average when they included at least five locations in each region. Pooled summary statistics reflect values calculated across all errors from all models, in order to comment on aggregate trends by time or geography. Results are shown here for the most recent four week window allowing for the calculation of errors at each point of extrapolation (see Figure 2 and methods). Results from other months are shown in the supplement.

Systematic assessments of bias for all models produced in July are shown in Figure 4, and Supplemental Figure 2. The Delphi and LANL models from July underestimated mortality, with median percent errors of -5.6% and -8.3% at 6 weeks respectively, while Imperial tended to overestimate (+47.7%), and the remaining models were relatively unbiased.

**Figure 4.**
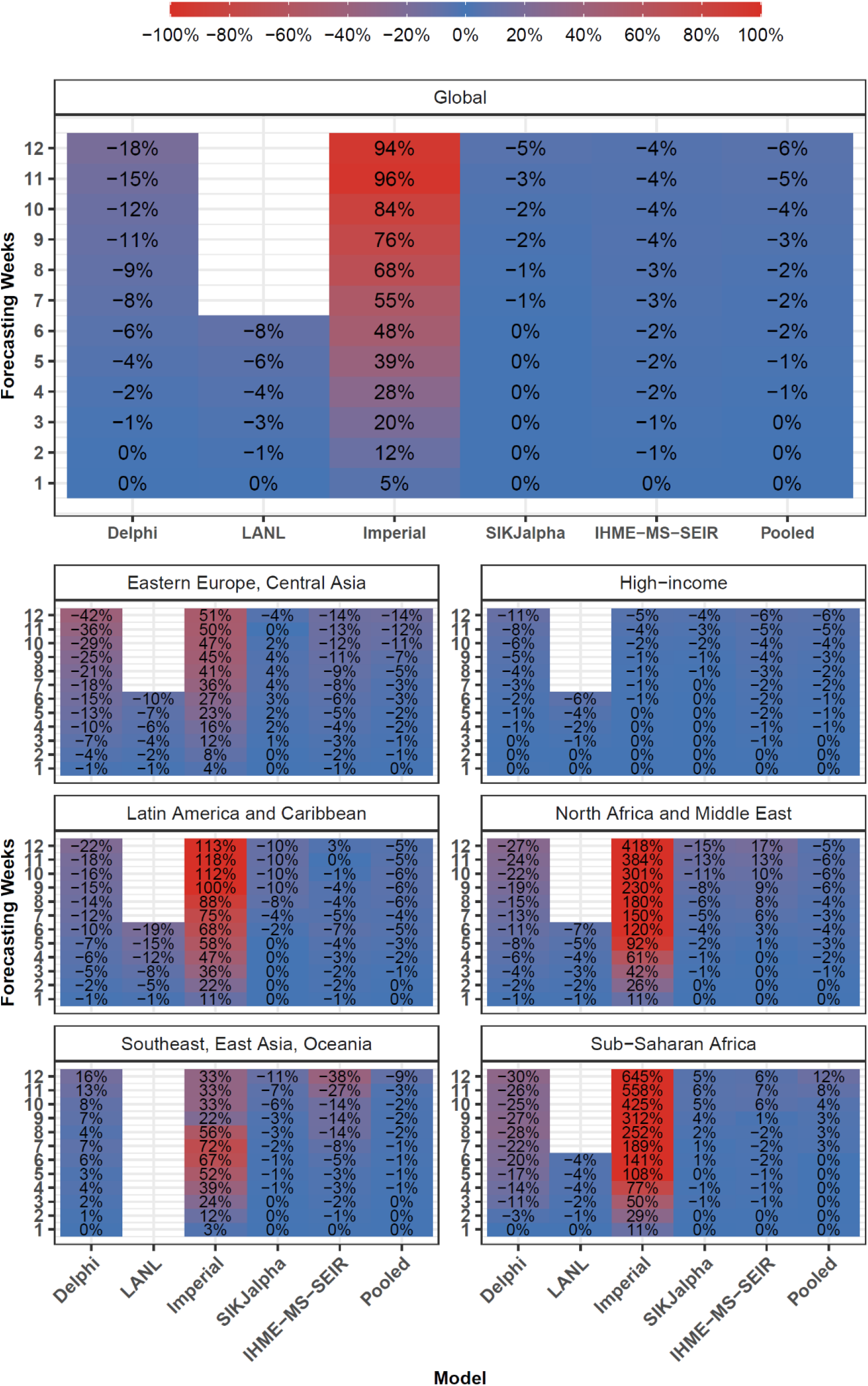
Month Stratified July Models - Cumulative Mortality Bias - Median Percent Error. Median percent error values, a measure of bias, were calculated across all observed errors at weekly intervals, for each model, by weeks of forecasting and geographic region. Values that represent fewer than five locations are masked due to small sample size. Models were included in the global average when they included at least five locations in each region. Pooled summary statistics reflect values calculated across all errors from all models, in order to comment on aggregate trends by time or geography. Results are shown here for models released in July, and results from other months are shown in the appendix.

Overall model performance for models produced in July is shown for cumulative deaths by week in Figure 5. As one might expect, MAPE tends to increase by the number of weeks of extrapolation. Across models released in July the MAPE rose from 1.8% at one week to 24.6% at twelve weeks. Decreases in predictive ability with greater periods of extrapolation were similarly noted for errors in weekly deaths (Supplemental Figure 3). At the global level, MAPE at six weeks was less than 15% for LANL (10.6%), IHME-MS-SEIR (10.6%), SIKJalpha (12.3%) and Delphi (13.6%). The Imperial model had larger errors, about 5-fold higher than other models by six weeks. This appears to be largely driven by the aforementioned tendency to overestimate mortality. At twelve weeks, MAPE values were lowest for the IHME-MS-SEIR (23.7%) model, while the Imperial model had the most elevated MAPE (98.8%). Predictive performance between models was generally similar for median absolute errors (MAEs) (see Supplemental Figure 4). Global MAE values at 12 weeks, among models released in July varied from 204 for the IHME-MS-SEIR model to 1,264 for the Imperial model.

**Figure 5.**
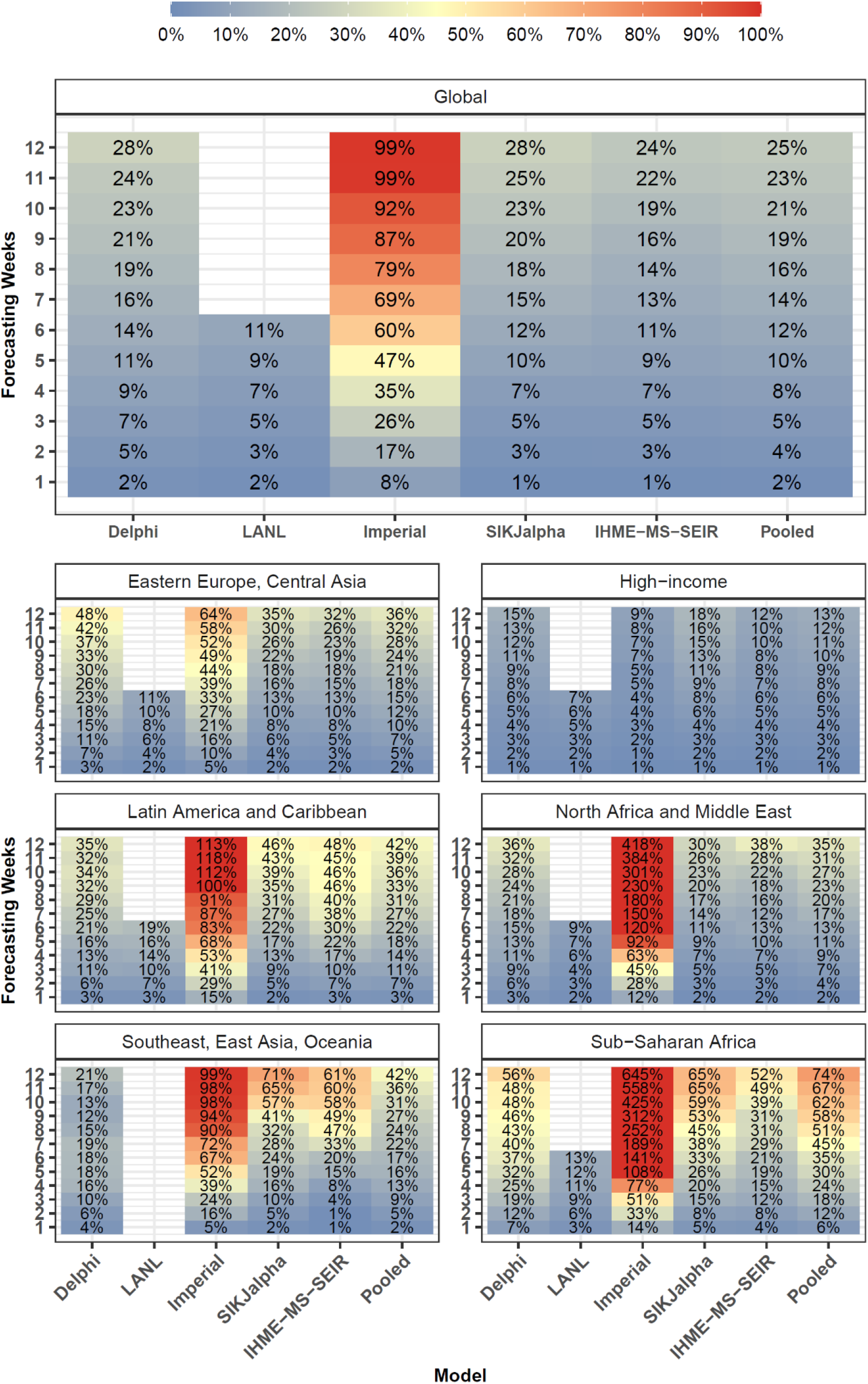
Month Stratified July Models - Cumulative Mortality Accuracy – Median Absolute Percent Error. Median absolute percent error values, a measure of accuracy, were calculated across all observed errors at weekly intervals, for each model by weeks of forecasting and geographic region. Values that represent fewer than five locations are masked due to small sample size. Models were included in the global average when they included at least five locations in each region. Pooled summary statistics reflect values calculated across all errors from all models, in order to comment on aggregate trends by time or geography. Results are shown here for models released in July, and results from other months are shown in the supplement.

Figure 5 also shows that model performance varies substantially by region. The lowest errors across models were observed among high-income countries with a 6-week MAPE values of 6.3%. In contrast, the largest errors were seen in sub-Saharan Africa, with a 6-week MAPE of 34.8%, and Latin America and the Caribbean, with a MAPE of 22.4%. Individual model performance and availability also varied by region.

The evaluation framework for exploring the ability of models to predict the timing of peak mortality accurately—a matter of paramount importance for health service planning—is shown in Figure 6 for an example location, Massachusetts. Similar figures for all locations are shown in the appendix. Median absolute errors (MAE) for peak timing also rose with increased forecasting weeks, from 13 days at one week to 30 days at eight weeks (Figure 7). The MAE at eight weeks ranged from 27 days for the IHME Curve Fit and SIKJ-Alpha models to 54 days for the LANL model, with an overall error across models of 30 days (Figure 7). Models were generally biased towards predicting peak mortality too early (Supplemental Figure 5).

**Figure 6.**
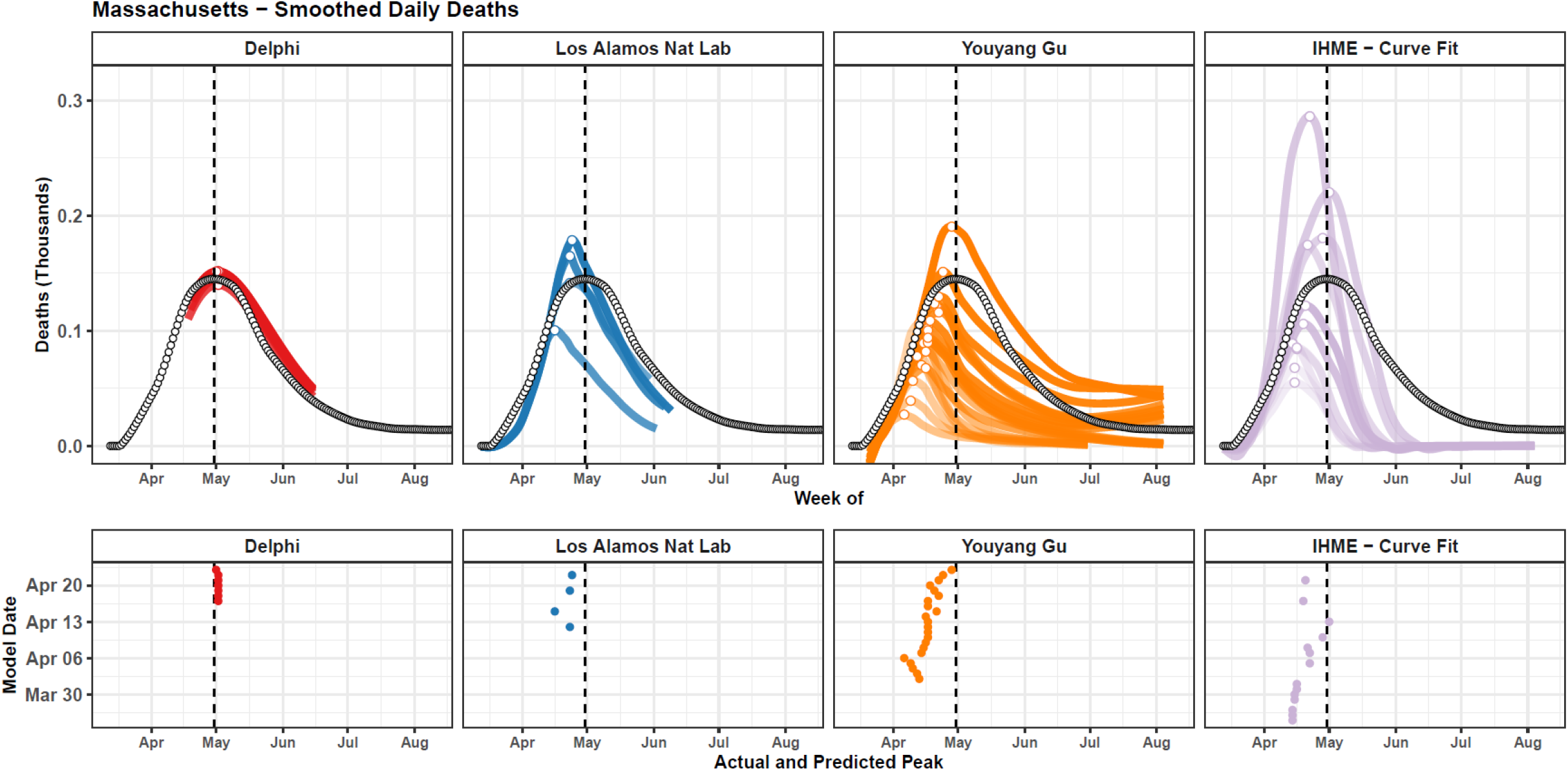
Observed vs Predicted Peak in Daily Deaths– Example for Massachusetts. Observed daily deaths, smoothed using a loess smoother, are shown as black-outlined dots (top). The observed peak in daily deaths is shown with a vertical black line (bottom). Each model version that was released at least one week prior to the observed peak is plotted (top) and its estimated peak is shown with a point (top and bottom). Estimated peaks are shown in the bottom panel with respect to their predicted peak date (x-axis) and model date (y-axis). Values are shown for the Massachusetts, and similar graphs for all other locations are available in the appendix. Massachusetts was chosen as the example location as the United States (used as the example for Figure 1) peaked earlier, only allowing for two models to provide peak timing errors, whereas Massachusetts peaked later, allowing for four models, making for a more illustrative example.

**Figure 7.**
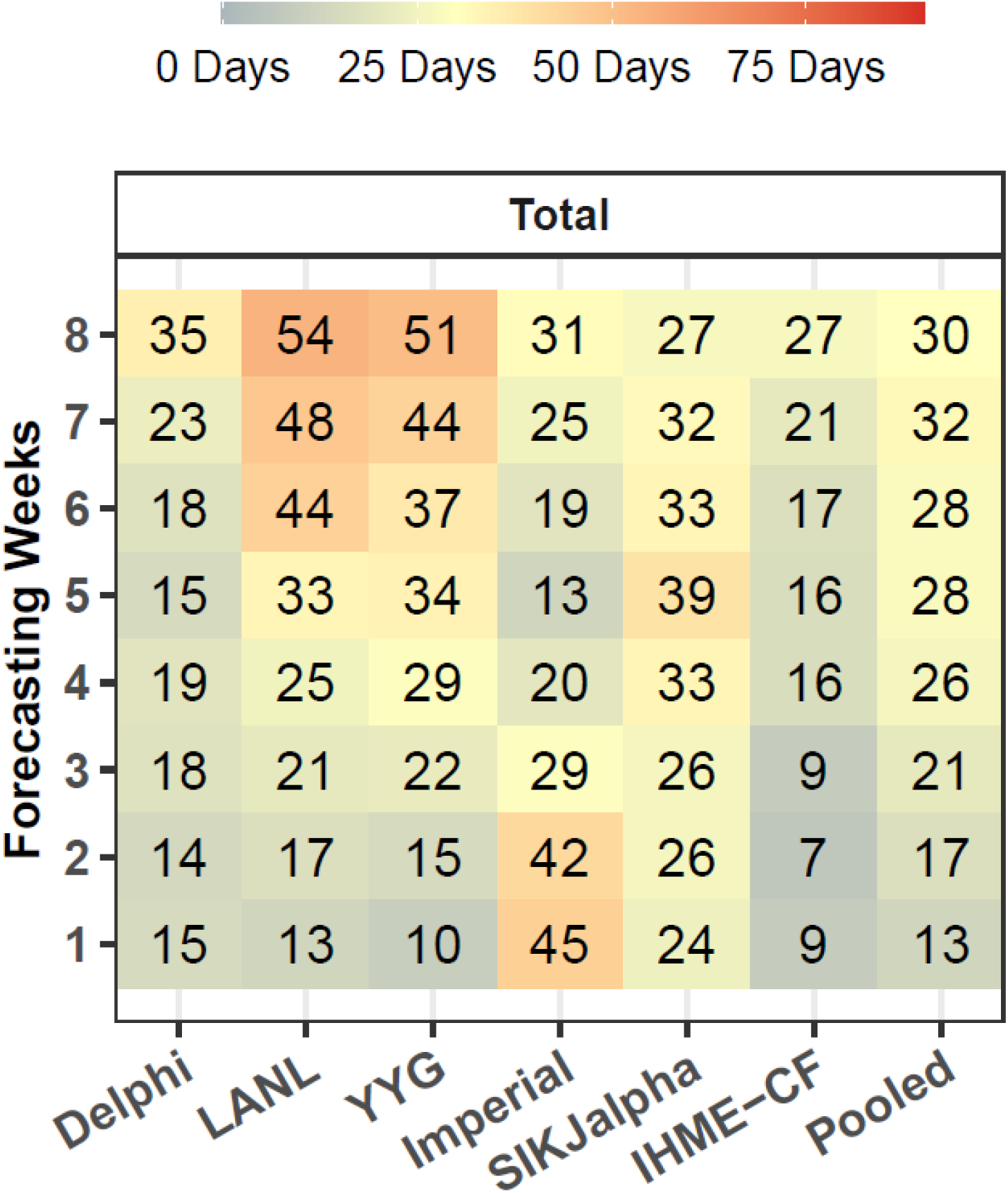
Peak Timing Accuracy – Median Absolute Error in Days. Median absolute error in days is shown by model and number of weeks of forecasting. Models that are not available for at least 40 peak timing predictions are not shown. Errors only reflect models released at least seven days before the observed peak in daily mortality. One week of forecasting refers to errors occurring from seven to 13 days in advance of the observed peak, while two weeks refers to those occurring from 14 to 20 days prior, and so on, up to six weeks, which refers to 42-48 days prior. Errors are pooled across month of estimation, as we found little evidence of change in peak timing performance by month (see appendix).

## Discussion

Eight COVID-19 models were identified that covered more than five countries, were regularly updated, publicly released and provide archived results for past forecasts. Taken together at twelve weeks, the models released in July had a median average percent error of 24.6% percent. Errors tend to increase with longer forecasts, rising from 1.8% at one week to 24.6% at 12 weeks. At twelve weeks of extrapolation, the best predictive performance among models considered at the global level was observed for the IHME-MS-SEIR models, with a MAPE of 23.7%, although the best performing model varied by region. The projections provided by Imperial had considerably higher error (98.8%) and the SIKJalpha and Delphi models had intermediate performance for the same period. In the most current models, the 6-week MAPE across models was 7.2%.

Although models largely converged in their predictions for the summer of 2020 period, forecasts began to diverge again among predictions for Fall 2020 and Winter 2021. These later divergences are likely due to differences in model assumptions related to the effects of seasonality. Although the top performing models are currently performing in a highly comparable fashion, the updated results presented in this framework in an ongoing fashion may highlight major predictive performance differences as the validity of these assumptions are born out in the coming months.

A forecast of the trajectory of the COVID-19 epidemic for a given location depends on three sets of factors: 1) attributes of the virus itself, and characteristics of the location, such as population density and the use of public transport; 2) individual behavioural responses to the pandemic such as avoiding contact with others or wearing a mask; and 3) the actions of governments, such as the imposition of a range of social distancing mandates. Given the complexity of forecasting human and governmental behaviours, especially in the context of a new pandemic, performance of most of the models evaluated here was encouraging. Nevertheless, errors were observed to grow with greater extrapolation time, indicating that governments and planners should recognize the wide uncertainty that comes with longer range forecasts, and strategize accordingly. Hospital administrators may want to hedge on the higher end of the forecast range, while government policymakers may elect to use the mean forecast, depending on their risk tolerance.

We also observed substantial differences in average model predictive performance between regions, which can likely be explained by several factors. Data quality has been shown to vary substantially between countries, and many models were initially calibrated on data from early epidemics in China, Europe, and the United States. Furthermore, different regions are at different stages of their epidemic at any given time. For many of the countries in Sub-Saharan Africa for example, the challenge is predicting if, and when, large outbreaks will occur. It is therefore easier for a model to demonstrate large magnitude errors when it predicts a completely different epidemic trajectory. Contrastingly, in some of the more established epidemics, it is easier to predict the nature of more stabilized, ongoing transmission dynamics.

We also note that the vast majority of COVID-19 forecasting models did not provide sufficient information to be included in this framework, given that publicly available and date-version forecasts were not made available. We would encourage all research groups forecasting COVID-19 mortality to consider providing historical versions of their models in a public platform for all locations, to facilitate ongoing model comparisons. This will improve reproducibility, the speed of development for modelling science, and the ability of policy makers to discriminate between a burgeoning number of models^20^. Many of the models featured in this analysis were generally unbiased, or tended to underestimate future mortality, while other models, such as the Imperial model, as well as many other published models that did not meet our inclusion criteria, tend to substantially overestimate transmission, even within the first four weeks of a forecast. This tendency towards over-estimation among SEIR and other transmission-based models is easy to understand given the potential for the rapid doubling of transmission. Nevertheless, sustained exponential growth in transmission is not often observed, likely due to the behavioural responses of individuals and governments; both react to worsening circumstances in their communities, modifying behaviours and imposing mandates to restrict activities. This endogenous behavioural response is commonly included in economic analyses, however, it has not been routinely featured in transmission dynamics modelling of COVID-19. More explicit modelling of the endogenous response of individuals and governments may improve future model performance for a range of models.

Modelling groups are increasingly providing both reference forecasts, describing likely future trends, and alternative scenarios describing the potential effects of policy choices, such as school openings, timing of mandate re-imposition, or planning for hospital surges. For these scenarios, the error in the reference forecast—which we describe in this manuscript—is actually less important than the error in the effect implied by the difference between the reference forecast and policy scenario. Unfortunately, evaluating the accuracy of these counterfactual scenarios is an extremely difficult task. The validity of such claims depends on the supporting evidence for the assumptions about a policy’s impact on transmission. The best option for decision-makers is likely to examine the impact of these policies as portrayed by a range of modelling groups, especially those that have historically had reasonable predictive performance in their reference forecasts.

Given that a number of very different models demonstrated recent six-week errors for cumulative deaths below 10%, it would likely be worthwhile to construct an ensemble of these models and evaluate the performance the ensemble compared to each component. Although from a logistical standpoint, creating an ensemble of the forecasts would be relatively straightforward, it would be more challenging to integrate such a model pool with scenarios assessing policy options, given that the models have highly different underlying structures. Nevertheless, the inclusion of the models shown here, and future models meeting criteria into an ensemble framework, is an important area for future research.

This analysis of the performance of publicly released COVID-19 forecasting models has limitations. First, we have focused only on forecasts of deaths, as they are available for all models included here. Hospital resource use is also of critical importance, however, and deserves future consideration. Nevertheless, this will be complicated by the heterogeneity in hospital data reporting; many jurisdictions report hospital census counts, others report hospital admissions, and still others do not release hospital data on a regular basis. Without a standardized source for these data, assessment of performance can only be undertaken in an *ad hoc* way. Second, many performance metrics exist which could have been computed for this analysis. We have focused on reporting median absolute percent error, as the metric is frequently used, quite stable, and provides an easily interpreted number that can be communicated to a wide audience. Relative error is an exacting standard, however. For example, a forecast of three deaths in a location that observed only one may represent a 200% error, yet it would be of little policy or planning significance. Conversely, focusing on absolute error would create an assessment dominated by a limited number of locations with large epidemics. Future assessment could consider different metrics that may offer new insights, although the relative rank of performance by model is likely to be similar.

When taking an inclusive approach to including forecasts from various modelling groups, including estimates from a wide range of time periods and geographies, extra care must be taken to ensure comparability between models. We use various techniques to construct fair companions, such as stratifying by region, month of estimation, and weeks of forecasting, and masking summary statistics representing a small number of values. Nevertheless, other researchers may prefer distinct methods of maximizing comparability over a complex and patchy estimate space. Furthermore, the domains assessed here —magnitude of total mortality and peak timing—are not an exhaustive list of all possible dimensions of model performance. By providing an open-access framework to compile forecasts and calculate errors, other researchers can build on the results presented here to provide additional analyses.

COVID-19 mortality forecasts have been used in myriad ways by policymakers as they make difficult decisions about resource management under unprecedented circumstances. Examples include prospectively managing or moving resources between regions such as hospital beds, ICU beds, ventilators, masks and other personal-protective equipment, as well as decisions about social distancing measures, stay-at-home orders, and closing schools, universities and workplaces^1,7^. It is therefore of paramount importance that decision-makers can quickly assess how robust each modelling groups predictions have been historically. Furthermore, we believe a similar approach could be adopted in future pandemics, and for modelling other infectious diseases such as influenza.

Ultimately, policymakers would benefit from considering a multitude of forecasting models as they consider resource planning decisions related to the response to the ongoing COVID-19 pandemic. This study provides a publicly available framework and codebase, which will be updated in an ongoing fashion, to continue to monitor model predictions in a timely manner, and contextualize them with prior predictive performance. It is our hope that this spurs conversation and cooperation amongst researchers, which might lead to more accurate predictions, and ultimately aid in the collective response to COVID-19. As the pandemic continues worldwide and resurges in Europe and North America become more evident, regularly updating models, and continually assessing their predictive validity, will be important in order to provide stakeholders with the best tools for COVID-19 decision-making.

## Methods

### Systematic Review

A total of 386 published and unpublished COVID-19 forecasting models were reviewed (see appendix). Models were excluded from consideration if they did not 1) produce estimates for at least five different countries, 2) did not extrapolate at least four weeks out from the time of estimation, 3) did not estimate mortality, 4) did not provide downloadable, publicly available results, or 5) did not provide date-versioned sets of previously estimated forecasts, which are required to calculate subsequent out-of-sample predictive validity. Eight models which fit all inclusion criteria were evaluated (Table 1). These included those modelled by: DELPHI-MIT (Delphi)^14,15^, Youyang Gu (YYG)^10^, the Los Alamos National Laboratory (LANL)^16^, Imperial College London (Imperial)^17^,the SIKJ-Alpha model from the USC Data Science Lab (SIKJalpha)^18^, and three models produced by the Institute for Health Metrics and Evaluation (IHME)^19^. Beginning March 25^th^, IHME initially produced COVID forecasts using a statistical curve fit model (IHME-CF), which was used through April 29^th^ for publicly released forecasts^1^. On May 4^th^, IHME switched to using a hybrid model, drawing on a statistical curve fit first stage, followed a second-stage epidemiological model with susceptible, exposed, infectious, recovered compartments (SEIR)^21^. This model—referred to herein as the IHME-CF SEIR model—was used through May 26^th^. On May 29^th^, the curve fit stage was replaced by a spline fit to the relationship between log cumulative deaths and log cumulative cases, while the second stage SEIR model remained the same^22^. This model, referred to as the IHME-MS SEIR model, is the basis for recently published work on US State level scenarios of COVID-19 projections in the fall and winter of 2020/2021^23^and was still in use at the time of this publication. The three IHME models rely upon fundamentally different assumptions and core methodologies, and therefore are considered separately. They were also released during different windows of the pandemic, and are therefore compared to models released during similar time periods.

In some cases, numerous scenarios were produced by modelling groups, to describe the potential effects of interventions, or future trajectories under different assumptions. In each case the baseline or status quo scenario was selected to evaluate model performance as that represents the modelers’ best estimate about the most probable course of the pandemic. Table 1 summarizes information about each model assumptions, methodologies, input data, modelled outputs, and forecasting range.

### Model Comparison Framework

In order to conduct a systematic comparison of the out-of-sample predictive validity of international COVID-19 forecasting models, a number of issues must be addressed. Looking across models, a high degree of heterogeneity can be observed in numerous dimensions, including sources of input data, frequency of public releases of model estimates, geographies included in the results, and how far into the future predictions are made available for. Differences in each of these areas must be taken into account, in order to provide a fair and relevant comparison.

#### Input data

A number of sources of input data—describing observed epidemiological trends in COVID-19—exist, and they often do not agree for a given country and time point^24–26^. We chose to use mortality data collected by the Johns Hopkins University Coronavirus Resource Center as the in-sample data against which forecasts were validated at the national level, and data from the New York Times for state-level data for the United States^25,26^. We chose to mainly rely on the Hopkins data as 1) it was the most common input data source used in the different models considered, 2) it covered all countries for which modelling groups produced forecasts, 3) although some quality issues were noted, and managed in our analysis, largely quality was deemed acceptable, and 4) data were made publicly available on a GitHub page and updated daily, which facilitates the maintenance of a timely comparison framework. Locations were excluded from the evaluation (including Ecuador and Peru) where models used alternative data sources, such as excess mortality, in settings with known marked under-registration of COVID-19 deaths and cases^27,28^. We adjusted for differences in model input data using intercept shifts, whereby all models where shifted to perfectly match the in-sample data for the date in which the model was released (see supplemental methods).

#### Frequency of public releases of model estimates

Most forecasting models are updated regularly, but at different intervals, and on different days. Specific days of the week have been associated with a greater number of reported daily deaths. Therefore, previous model comparison efforts in the United States— such as those conducted by the US Centers for Disease Control and Prevention—have required modelers to produce estimates using input data cut-offs from a specific day of the week^29^. For the sake of including all publicly available modelled estimates, we took a more inclusive approach, considering each publicly released iteration of each model. To minimize the effect of day-to-day fluctuations in death reporting, we focus on errors in cumulative and weekly total mortality, which are less sensitive to daily variation.

#### Geographies and time periods included in the results

Each model produces estimates for a different set of national and subnational locations, and extrapolates a variable amount of time from the present. Each model was also first released on a different date, and therefore reflects a different window of the pandemic. Here, we also took an inclusive but stratified approach, and included estimates from all possible locations and time periods. To increase comparability, summary error statistics were stratified by super-region used in the Global Burden of Disease Study^30^, weeks of extrapolation, and month of estimation, and we masked summaries reflecting a small number of locations or time points. Models were included in the global predictive validity results only when they were present for all regions. Estimates were included at the national level for all countries, except the United States, where they were also included at the admin-1 (state) level, as they were available for most models. In order to be considered for inclusion, models were required to forecast at least four weeks into the future.

#### Outcomes

Finally, each model also includes different estimated quantities, including daily and cumulative mortality, number of observed or true underlying cases, and various dimensions of hospital resource utilization. The focus of this analysis is on mortality, as it was the most widely reported outcome, and it also has a high degree of societal, epidemiological and public health importance. We did not focus on forecasts of confirmed cases for several reasons. Certain models we wished to include did not provide an estimate of confirmed cases to subsequently assess predictive performance. Mortality, on the other hand, was available for all models. Furthermore, confirmed cases also depend on testing rates, which vary widely over time and across locations. Modelling confirmed cases, therefore, represents different and perhaps larger challenges. Of course, death numbers also have limitations, but they are generally more reliable than case numbers, at least in the early stages of the pandemic, and in locations with limited capacity to test.

### Comparison of Cumulative Mortality Forecasts

The total magnitude of COVID-19 deaths is a key measure for monitoring the progression of the pandemic. It represents the most commonly produced outcome of COVID-19 forecasting models, and perhaps the most widely debated measure of performance. The main quantity that is considered is errors in total cumulative deaths—as opposed to other metrics such as weekly or daily deaths—as it has been most commonly discussed measure, to-date, in academic and popular press critiques of COVID-19 forecasting models. Nevertheless, alternate measures are presented in the appendix. Errors were assessed for systematic upward or downward bias, and errors for weekly, rather than cumulative deaths, were also assessed. In calculating summary statistics, percent errors were used to control for the large differences in the scale of the epidemic between locations. Medians, rather than means, are calculated due to a small number of large magnitude outliers present in a few time-series. Errors from all models were pooled to calculate overall summary statistics, in order to comment on overarching trends by geography and time.

Results are presented using two analytical strategies in the main text. Both strategies are highlighted in Figure 2. The “most current” approach is used to select the data shown in Figure 3. The “month stratified” approach is used for Figures 3 and 4. In the “most current” approach, the most recent 4 weeks of model dates are used for each extrapolation length. Therefore, for 1-week errors, models from October were used, whereas for 12-week errors, models from July and August were used. This allows for the assessment of the most recent evidence possible for each set of errors displayed. 4-week periods are used to ensure that the results are not unduly biased by featuring only a small number of runs for each model.

In the “month stratified” approach, models from July are used in all cases. This strategy allows for more reliable assessment of certain aspects of predictive validity, as the same models are being compared over time and geographies. For example, the month stratified approach may provide a more comparable assessment of how errors grow with increased length of extrapolation. Models are shown for July in the main text—the most recent month allowing for assessment of errors at twelve weeks of forecasting— and errors stratified for all months are shown in the appendix.

### Comparison of Peak Daily Mortality Forecasts

Each model was also assessed on how well it predicted the timing of peak daily deaths—an additional aspect of COVID-19 epidemiology with acute relevance for resource planning. Peak timing may be better predicted by different models than those best at forecasting the magnitude of mortality, and therefore deserves separate consideration as an outcome of predictive performance. In order to assess peak timing predictive performance, the observed peak of daily deaths in each location was estimated first— a task complicated by the highly volatile nature of reported daily deaths values. Each timeseries of daily deaths was smoothed, and the date of the peak observed in each location, as well as the predicted peak for each iteration of each forecasting model was calculated (see supplemental methods). A LOESS smoother was used, as it was found to be the most robust to daily fluctuations. Results shown here reflect only those locations for which the peak of the epidemic had passed at the time of publication, and for which at least one set of model results was available seven days or more ahead of the peak date. Predictive validity statistics were stratified by the number of weeks in advance of the observed peak that the model was released, as well as the month in which the model was released. Results shown in the main text were pooled across months, as there was little evidence of dramatic differences over time (see appendix). There was insufficient geographic variation to stratify results by regional groupings, although that remains an important topic for further study, which will become feasible as the pandemic peaks in a greater number of countries globally.

### Data and Code Availability

All data and versioned code required to reproduce this analysis its included visualizations are publicly available at (https://github.com/pyliu47/covidcompare).

## Supporting information

Supplemental Figures - Magnitude

Supplemental Figures - Peak Timing

Supplemental Figures - Smoothed Daily Deaths

## Data Availability

All data and code for this analysis are available at: https://github.com/pyliu47/covidcompare

## Acknowledgements

This work was primarily supported by the Bill & Melinda Gates Foundation. J.F. received support from the UCLA Medical Scientist Training program (NIH NIGMS training grant GM008042).

## Author Contributions

JF, PL, TV, SIH, CJLM, and EG conceptualized and designed the study, with substantial input from RR, RB, JC, SL, and DP. JF and PL acquired the data, and JF, PL, CT, and AC wrote the analytical code to conduct the analysis. JF, PL, and CJLM drafted the first draft of the article and all authors meaningfully revised.

SIH, CJLM, and EG supervised the work.

