## Supplement for "Predictive performance of international COVID-19 mortality forecasting models"

#### Supplemental Methods

In order to construct a framework to compare the out-of-sample predictive performance of COVID-19 mortality forecasting models, we conducted a comprehensive review of published and unpublished papers and models related to COVID forecasting. We identified models with date-versioned, publicly available mortality estimates for multiple countries, and create a code base to automatically download and process them into a common framework, as models update daily. We calculated errors for both the magnitude of mortality, and the timing of peak daily mortality. Finally, we calculated summary statistics describing the errors over a number of dimensions, including region, weeks of extrapolation, and month of estimation.

##### Comprehensive Review and Data Compilation

Conducting a comprehensive review of COVID-19 mortality forecasting is complicated by the unpublished nature of many models. In general, the pace of COVID-19 research has been rapid, to provide up-to-date evidence for an evolving situation. We therefore draw on both traditional and non-traditional sources to find models. We used PubMed to identify models published in journal articles, medRxiv to identify models published in preprint articles, and a collection of models curated by the Reich Lab and the US Centers for Disease Control describing forecasts for the United States, many of which also produce estimates internationally[16]. In PubMed and medRxiv we selected any articles with the terms “COVID”, or “Coronavirus” present, as well as one of either term “Project” or “Forecast”. At the time of publication of this article, n=686 articles had been screened. The systematic review framework was created to automatically download new citations for review, and subsequently be updated in a prospective manner, to stay current on future forecasting model efforts.

Each article or model was screened for 5 inclusion criteria:

1) Including a forecasting component (n=383),
2) and making projections for at least five locations (n=59),
3) and making projections for COVID-19 mortality (n=39),
4) and making projections at least four weeks into the future (n=35),
5) and providing publicly available, date-versioned estimates (n=8).

The eight models which fit these criteria are described in the main text (Table 1). All screened articles, including their inclusion and exclusion criteria, are described in the review supplemental file. The code used to compile candidate models and articles, and conduct the systematic review, is presented along with the remainder of the codebase.

For the eight models that were determined to meet all inclusion criteria, a codebase was developed to automatically download each date-versioned set of estimates as they became available. The model date (or estimation date) was assigned as the date on which the estimated became publicly available.

Locations were mapped onto the location hierarchy used by the Global Burden of Disease Study (GBD)[28] that categorizes all countries into regions, and regions into super-regions. When estimates were available at multiple geographic levels, the admin-0 (national) level results were used in all cases, except the United States, where both admin 0 and admin-1 (state) level results were used.

For models that provided only daily deaths, cumulative deaths were calculated as a rolling sum. Similarly, for models that provided only cumulative deaths, daily deaths were calculating by taking the daily first difference of cumulative deaths.

We chose to use mortality data collected by Johns Hopkins University Coronavirus Resource Center as the in-sample data against which forecasts were validated at the national level, and data from the New York Times for state-level data for the United States[20]. Data from both providers were mapped onto the GBD location hierarchy. For all analyses, the most recent set of input data were used, reflecting any potential revisions of historical trends.

#### Mortality Magnitude Predictive Validity

Before out-of-sample errors could be assessed for COVID-19 mortality, differences in input data sources between models were investigated and controlled. Estimates for the same locations, from different models, can differ greatly in magnitude of estimated mortality when they use input data sources that use different methodologies. To create a fair comparison, before errors were calculated, each model-, model-date-, and location-specific timeseries was shifted to match the "true" in-sample data for that model's date of release. This was accomplished by calculating the timeseries-specific difference on the model date, and apply it to the entire timeseries as a fixed intercept shift. Subsequent forecasting errors were calculated using the resulting shifted time-series.

Out-of-sample errors were calculated for each timeseries at weekly intervals, beginning at one week, through six weeks of extrapolation. Summary statistics were first calculated across model-runs for each location, for use in country-specific graphics (Figure 1, for example). Summary statistics included the median absolute error, a measure of accuracy, and median error, a measure of bias. These were calculated separately by model, and by weeks of extrapolation.

Subsequently, errors were summarized between countries. Summary statistics included the median absolute percent error, a measure of accuracy, and median percent error, a measure of bias. Relative error statistics were used for all inter-country comparisons, to account for the substantial differences in magnitude of deaths between locations. Summary statistics were calculated in a stratified manner by regional groupings from the GBD[28], as well as weeks of extrapolation, and month of estimation. Pooled summary statistics were also calculated across models, to provide context about commonalities in trends in predictive performance over time and geographies.

#### Peak Timing

In order to calculate out-of-sample predictive validity statistics on how well each model predicted the timing of peak daily deaths, we smoothed daily death data, which are highly stochastic, applied an algorithm to detect peaks in both observed data and forecasted model estimates, and calculated errors in the difference in number of days between the observed and estimated peaks.

First, observed daily death data were smoothed to provide stable time-series that could be used for local maxima detection. We used various smoothers to accomplish this task, including a LOESS smoother with a span of 0.33, run separately for each location-specific timeseries, a 7-day rolling average, and a 3-day rolling average applied tenfold to the same timeseries. We chose to present results calculated using the LOESS smoother in the main text, as it was found to be the most robust method to daily stochasticity that could introduce false peaks. Although most models produced smooth timeseries of daily deaths, some also demonstrated stochasticity, and so all forecasted daily death timeseries were also smoothed with a LOESS smoother.

Peaks in smoothed, observed daily deaths were calculated according to the following algorithm. A peak was defined as:

1) a local maximum *p* in the timeseries at time *t,*
2) where no other point exists in the next 21 days (*t* through *t*+21*)* that exceeds the *p* by more than 20%,
3) *t* does not fall within the last seven days of the timeseries,
4) where *p* represents at least 5 deaths per day,
5) and if multiple such points *p* exist that meet the above criteria, then the first value will be selected.

Peaks in forecasted trends were also identified with the same algorithm. For a time-series in which no peak was identified using the above algorithm, for a location which did have a peak in observed data, the global maximum value was used. This captured errors among models that failed to ever predict a peak, despite a true peak being observed. Errors for locations with a true peak in observed data, for the model runs in which the model date was at least seven days prior to the true detected peak. Errors were defined as the difference between the date of the true peak and the estimated peak from each forecasting model, in days. Summary statistics included the median absolute error in days, as a measure of accuracy, and the median error in days as a measure of bias. Errors were stratified by model, and weeks of extrapolation, which was defined as:

$$Weeks\ of\ extrapolation\ =\ floor((peak\ date - model\_release\_date)/7)$$

Summary statistics were masked for models that were not released in time to produce peak timing estimates for at least 25 total locations. Due to limited regional coverage it was not possible to stratify results by geography. This will likely become feasible as more locations pass their peak of daily mortality.

### Supplemental Figures

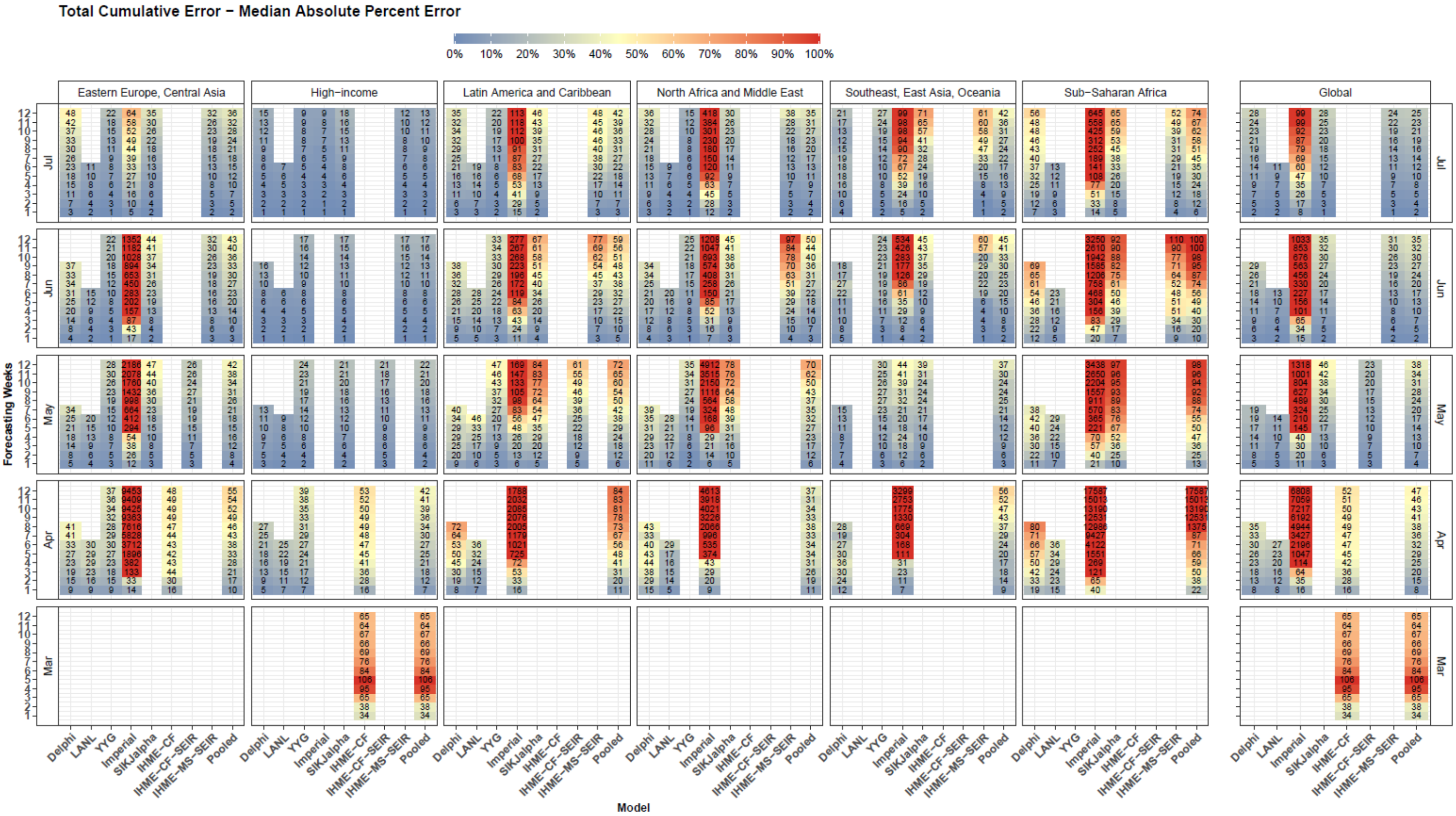


**Supplemental Figure 1. Cumulative Mortality – Median Absolute Percent Error – Month of Estimation**

Median absolute percent error values were calculated across all observed errors at weekly intervals, for each model, by month of estimation, weeks of forecasting, and super regional grouping used in the Global Burden of Disease Study. Values that represent fewer than five locations are masked due to small sample size. Models were included in the global average when they included at least five locations in each region. Pooled summary statistics reflect values calculated across all errors from all models, in order to comment on aggregate trends by time or geography.

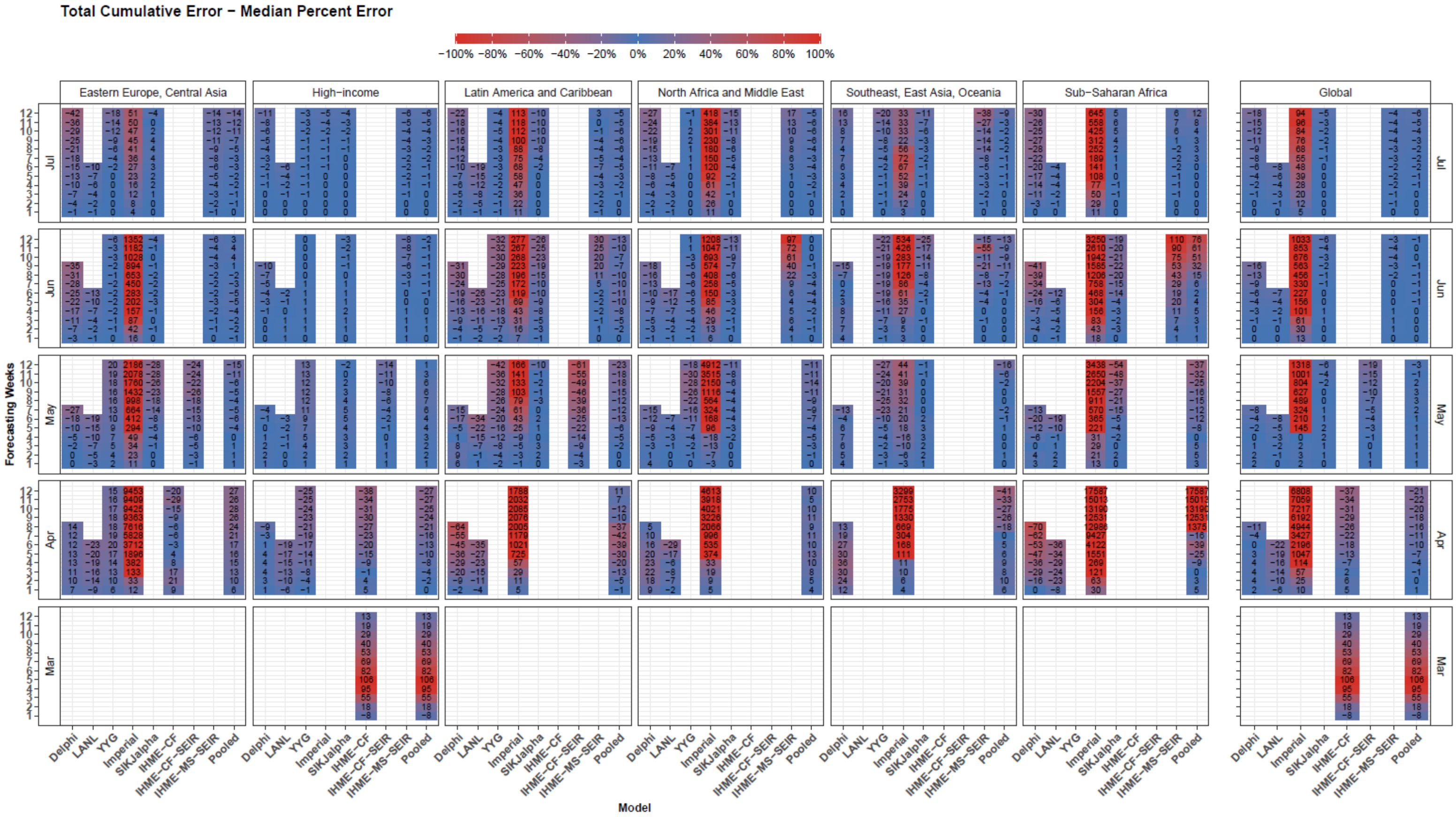


**Supplemental Figure 2. Cumulative Mortality – Median Percent Error – Month of Estimation**

Median percent error values, a measure of bias, were calculated across all observed errors at weekly intervals, for each model, by month of estimation, weeks of forecasting, and super regional grouping used in the Global Burden of Disease Study. Values that represent fewer than five locations are masked due to small sample size. Models were included in the global average when they included at least five locations in each region. Pooled summary statistics reflect values calculated across all errors from all models, in order to comment on aggregate trends by time or geography.

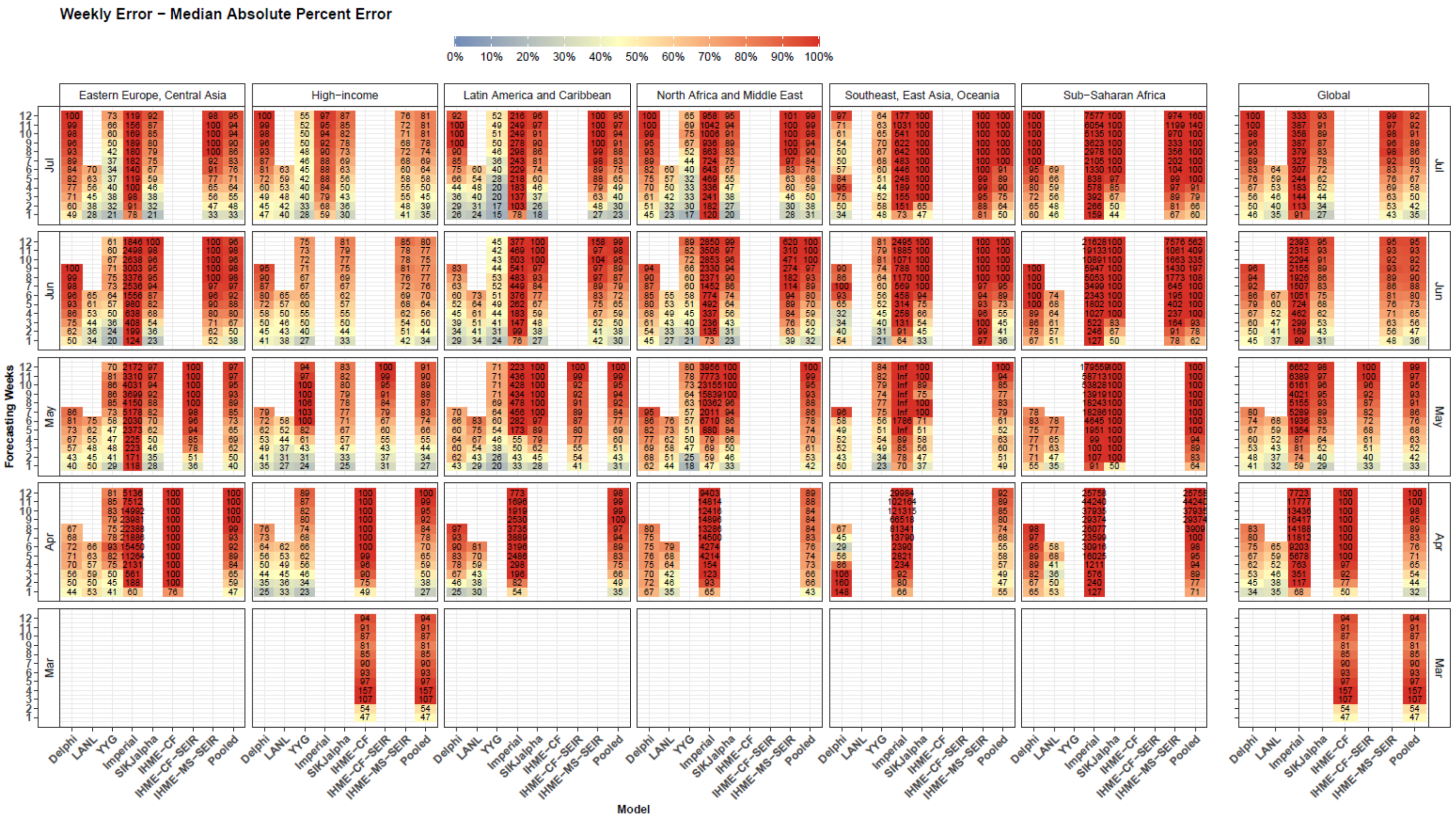


**Supplemental Figure 3. Weekly Mortality – Median Absolute Percent Error – Month of Estimation** Median absolute percent error values were calculated for weekly mortality rates across all observed errors at weekly intervals, for each model, by month of estimation, weeks of forecasting, and super regional grouping used in the Global Burden of Disease Study. Values that represent fewer than 5 locations are masked due to small sample size. Models were included in the global average when they included at least five locations in each region. Pooled summary statistics reflect values calculated across all errors from all models, in order to comment on aggregate trends by time or geography.

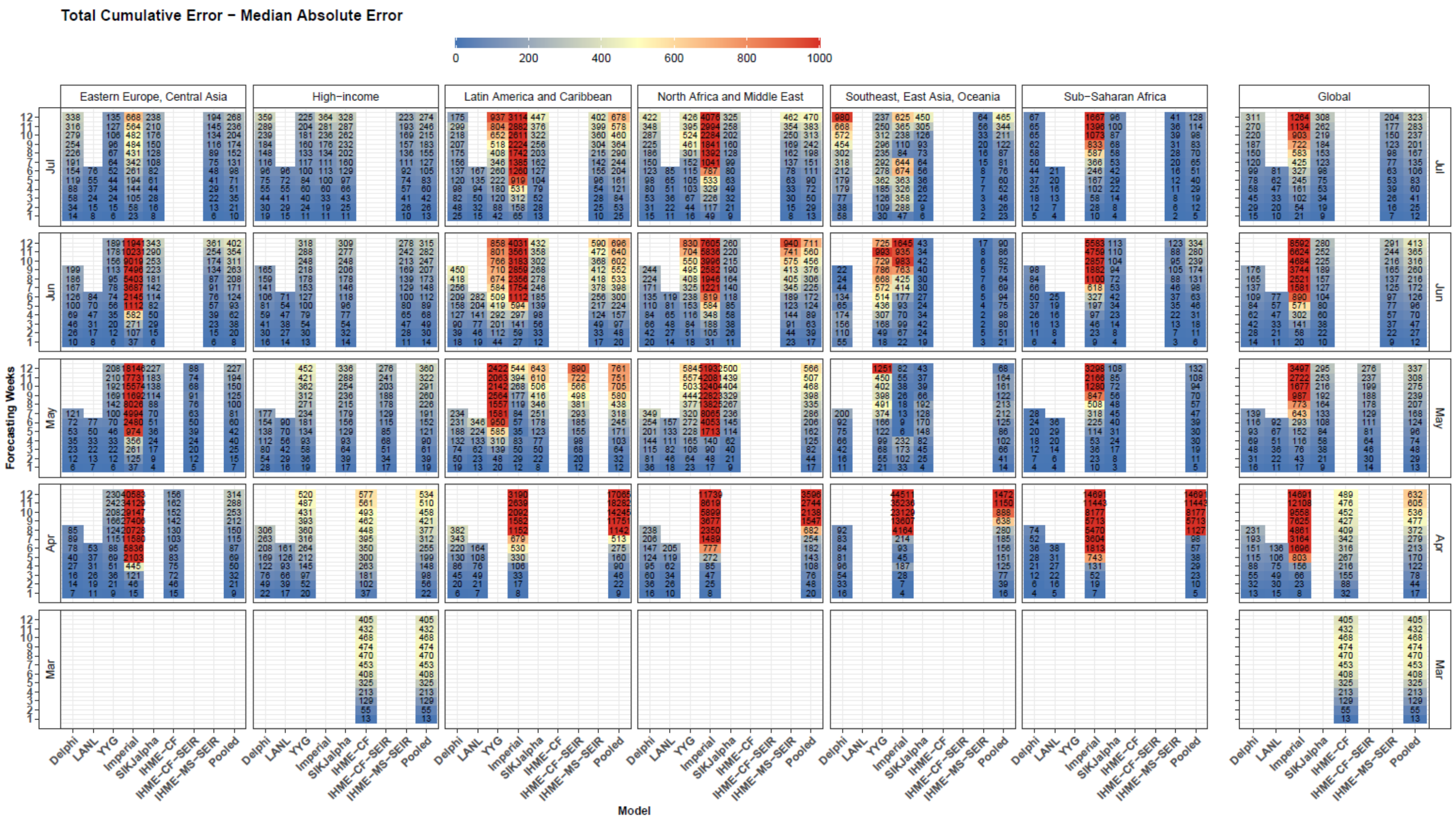


**Supplemental Figure 4. Cumulative Mortality – Median Absolute Error – Month of Estimation** Median absolute error values were calculated for cumulative mortality rates across all observed errors at weekly intervals, for each model, by month of estimation, weeks of forecasting, and super regional grouping used in the Global Burden of Disease Study. Values that represent fewer than 5 locations are masked due to small sample size. Models were included in the global average when they included at least five locations in each region. Pooled summary statistics reflect values calculated across all errors from all models, in order to comment on aggregate trends by time or geography.

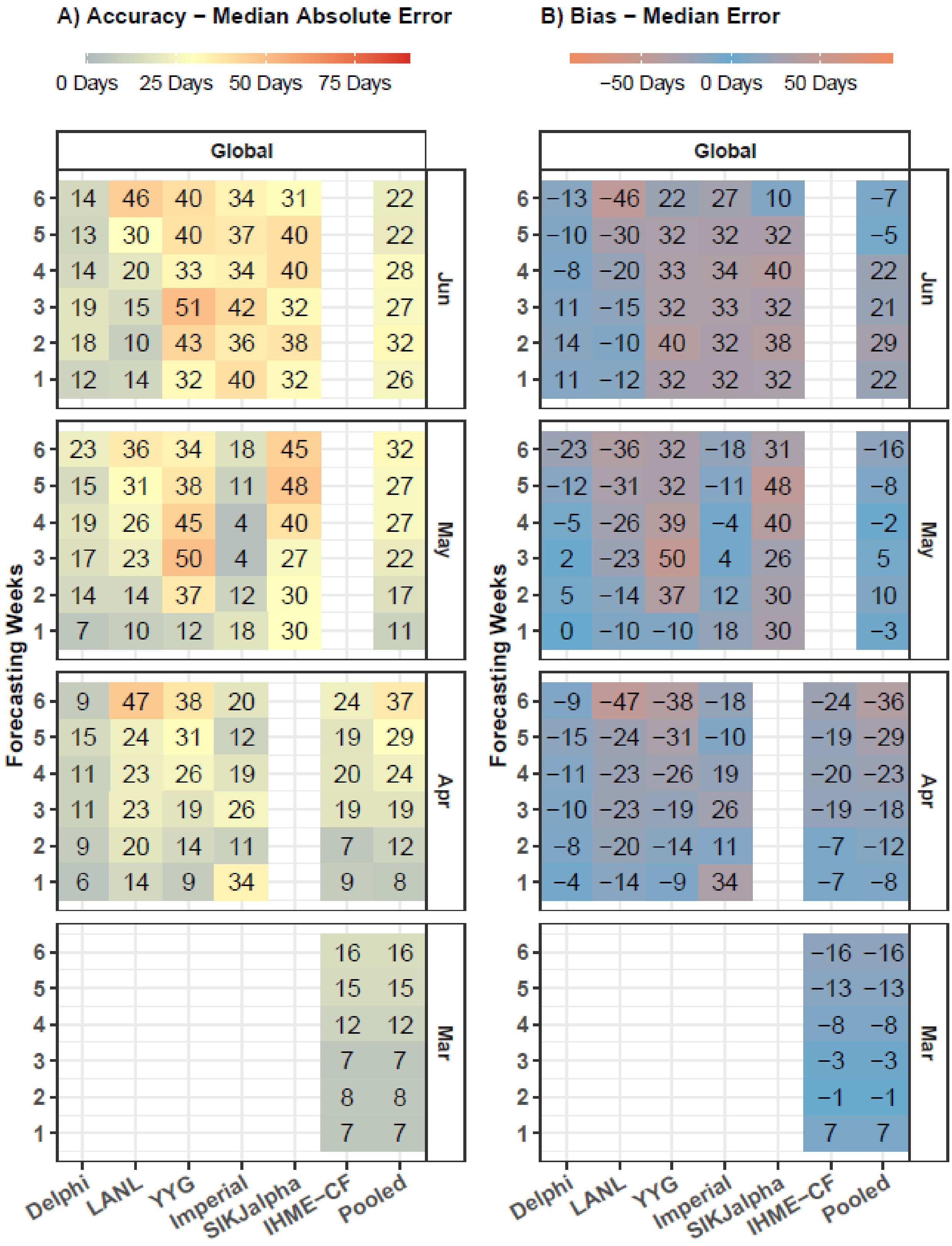


**Supplemental Figure 5. Accuracy and Bias in Peak Timing by Month of Estimation**
Median error in days are shown by model, weeks of forecasting, and estimation month. Models that are not available for at least 40 peak timing predictions are not shown. Errors only reflect models released at least seven days before the observed peak in daily mortality. One week of forecasting refers to errors occurring from seven to 13 days in advance of the observed peak, while two weeks refers to those occurring from 14 to 20 days prior, and so on, up to six weeks, which refers to 42-48 days prior.

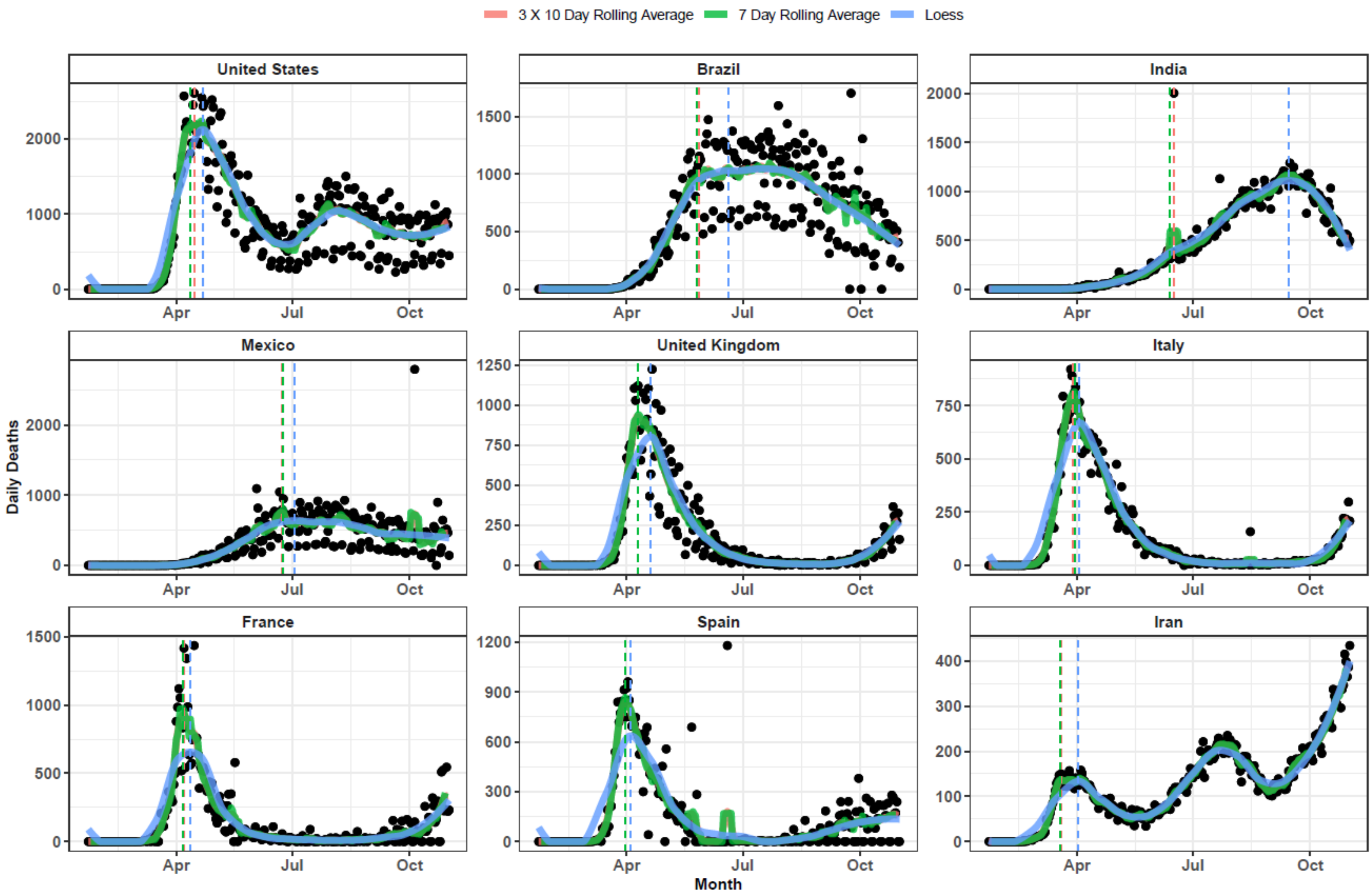


**Supplemental Figure 6. Smoothing Method – Example for nine Countries**

Daily deaths are shown for nine locations, as well as three methods used to smooth them prior to peak date calculation. Calculated peaks from each method are shown with dashed vertical lines. Smoothing method is shown by color.
