## Supplemental Figures - Magnitude for "Predictive performance of international COVID-19 mortality forecasting models"

### United States

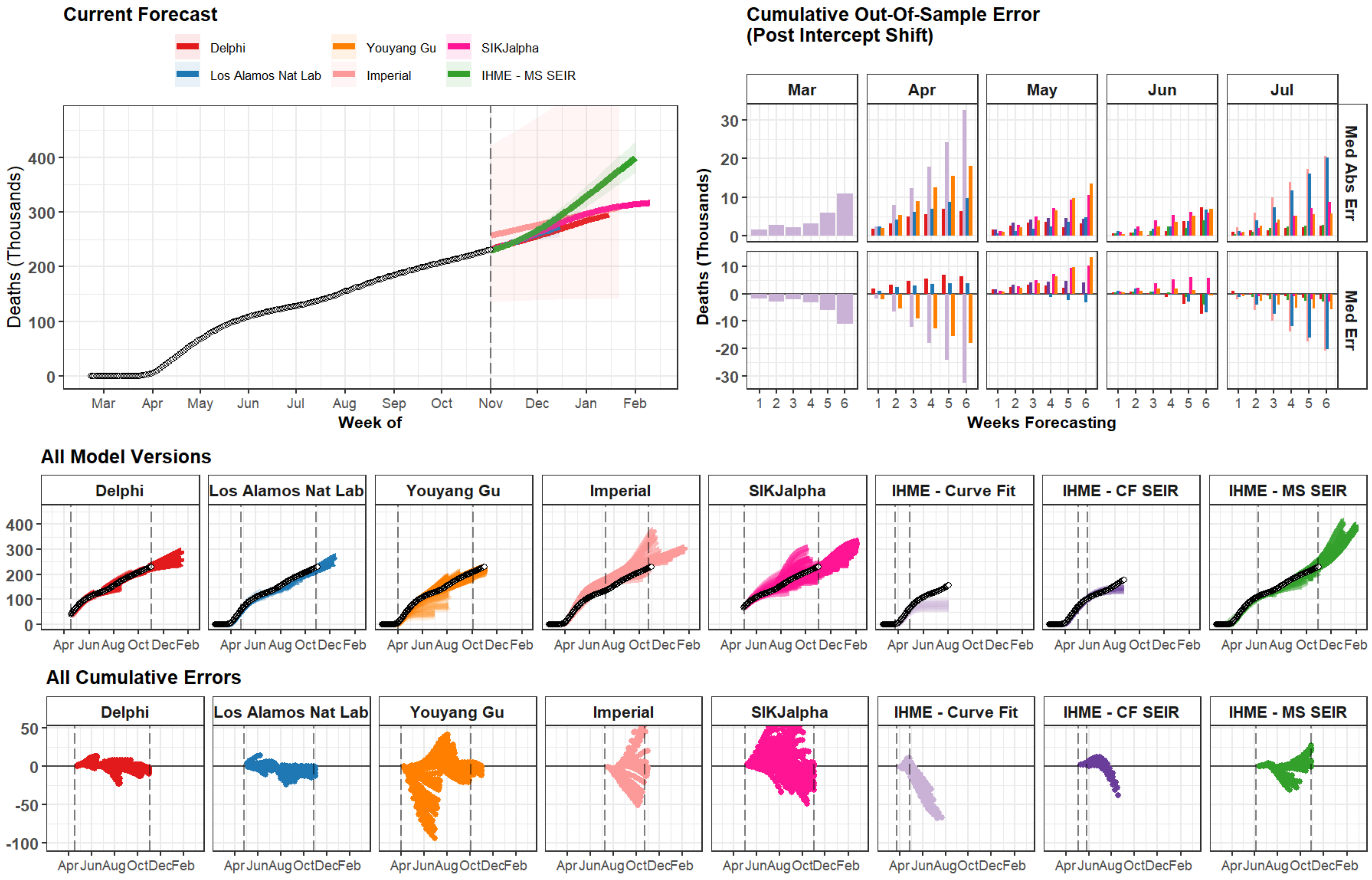

### Brazil

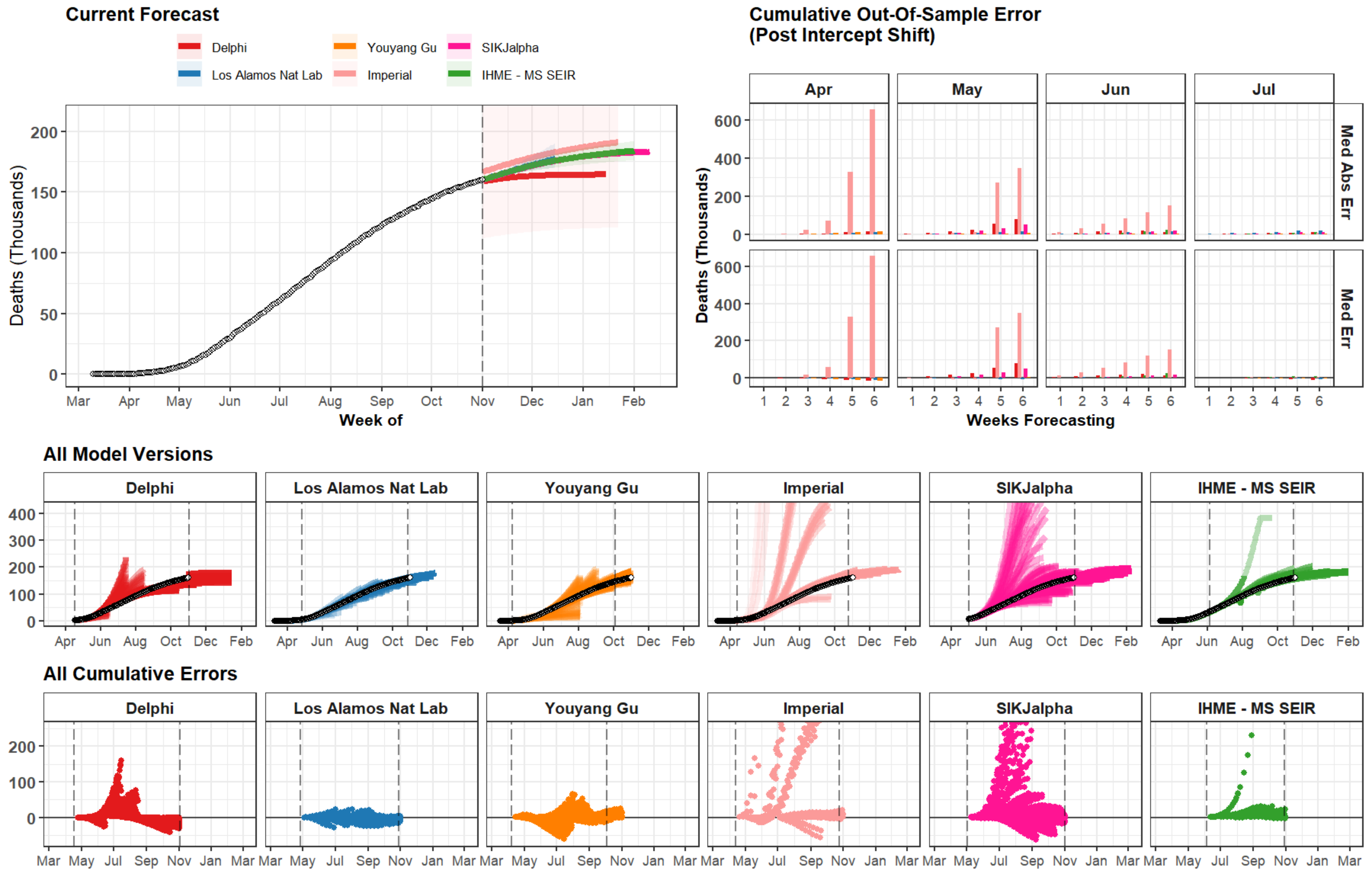
Current Forecast
Delphi
Los Alamos Nat Lab
Youyang Gu
Imperial
SIKJalpha
IHME - MS SEIR
Deaths (Thousands)
Week of
Cumulative Out-Of-Sample Error
(Post Intercept Shift)
Apr
May
Jun
Jul
Med Abs Err
Med Err
Weeks Forecasting
All Model Versions
All Cumulative Errors

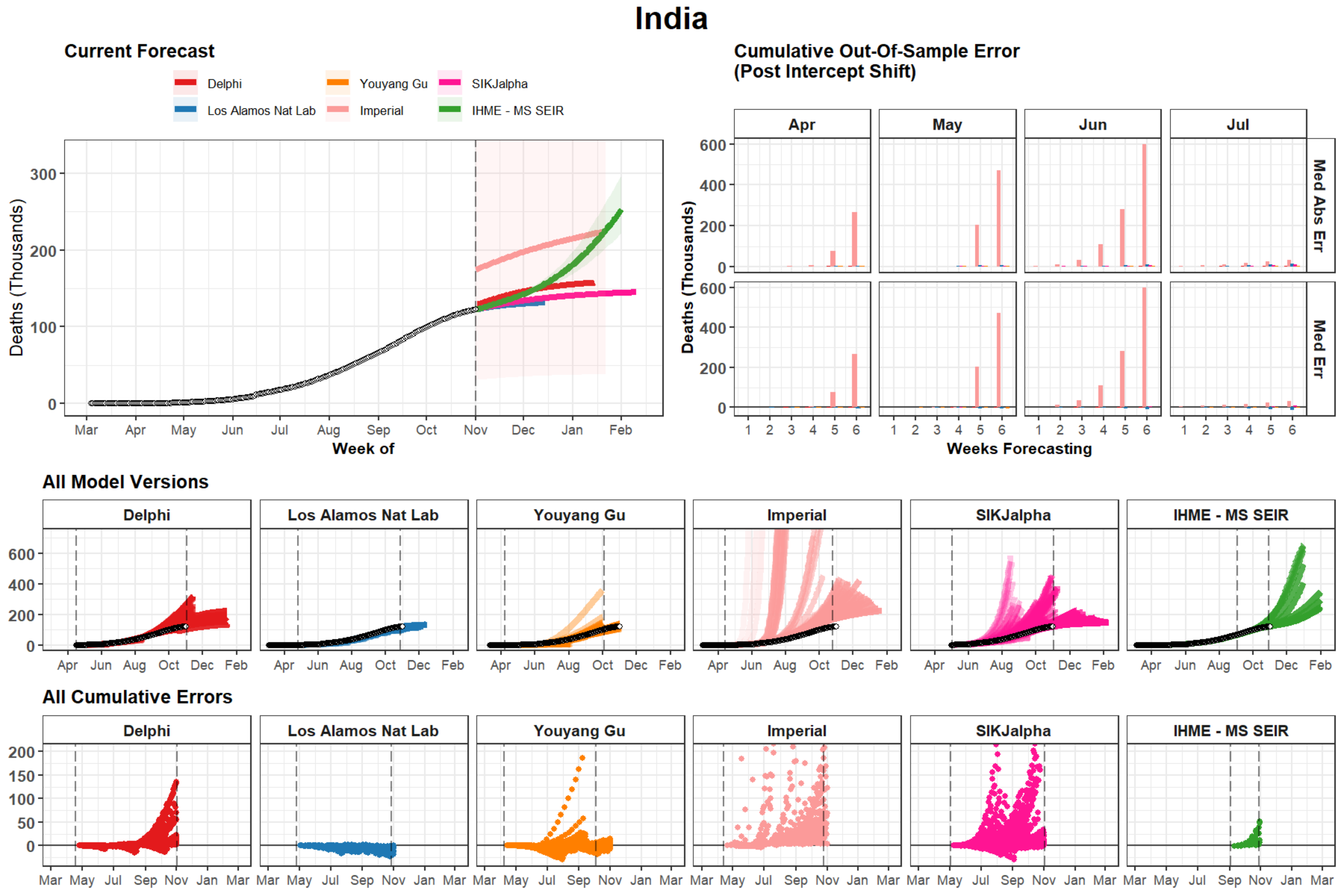

India
Current Forecast
Delphi
Los Alamos Nat Lab
Youyang Gu
Imperial
SIKJalpha
IHME - MS SEIR
Deaths (Thousands)
300
200
100
0
Mar Apr May Jun Jul Aug Sep Oct Nov Dec Jan Feb
Week of
Cumulative Out-Of-Sample Error
(Post Intercept Shift)
Apr May Jun Jul
Med Abs Err
Med Err
600
400
200
0
1 2 3 4 5 6
Weeks Forecasting
All Model Versions
Delphi
Los Alamos Nat Lab
Youyang Gu
Imperial
SIKJalpha
IHME - MS SEIR
600
400
200
0
Apr Jun Aug Oct Dec Feb
All Cumulative Errors
Delphi
Los Alamos Nat Lab
Youyang Gu
Imperial
SIKJalpha
IHME - MS SEIR
200
150
100
50
0
Mar May Jul Sep Nov Jan Mar

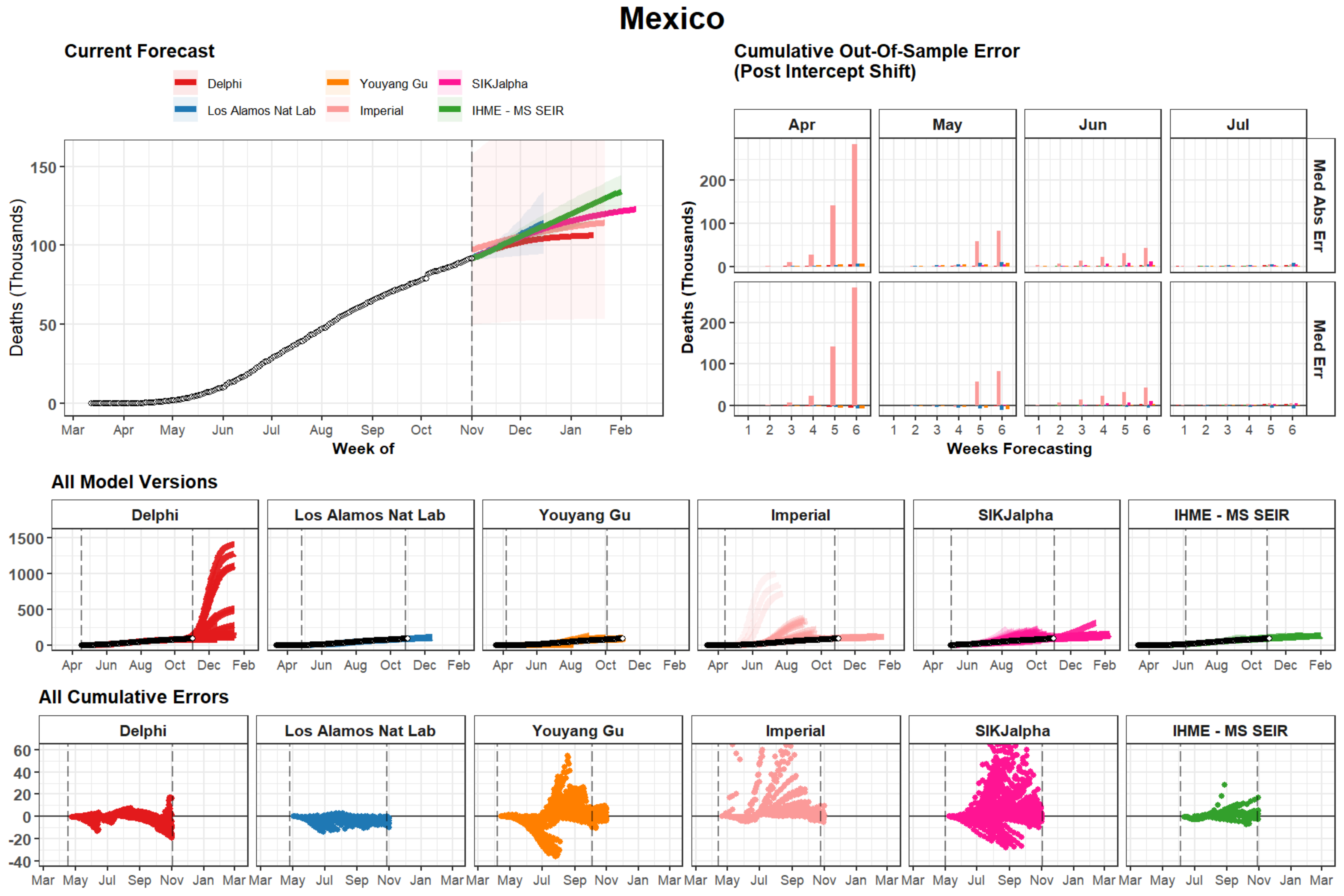
Mexico
Current Forecast
Delphi
Youyang Gu
SIKJalpha
Los Alamos Nat Lab
Imperial
IHME - MS SEIR
Deaths (Thousands)
Week of
Cumulative Out-Of-Sample Error
(Post Intercept Shift)
Apr
May
Jun
Jul
Med Abs Err
Med Err
Weeks Forecasting
All Model Versions
All Cumulative Errors

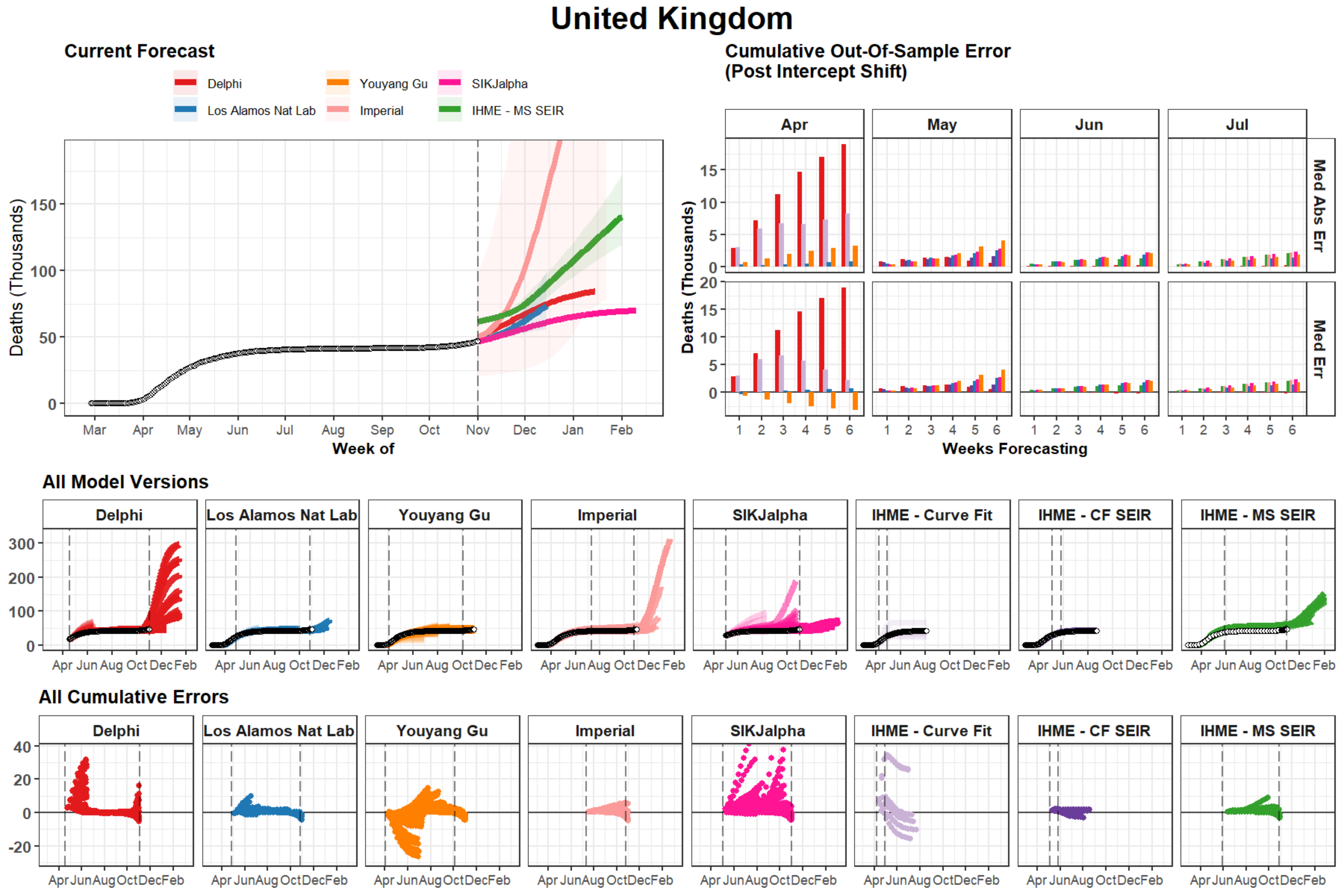
United Kingdom
Current Forecast
Delphi
Los Alamos Nat Lab
Youyang Gu
Imperial
SIKJalpha
IHME - MS SEIR
Deaths (Thousands)
150
100
50
0
Mar
Apr
May
Jun
Jul
Aug
Sep
Oct
Nov
Dec
Jan
Feb
Week of
Cumulative Out-Of-Sample Error
(Post Intercept Shift)
Apr
May
Jun
Jul
Med Abs Err
Med Err
Deaths (Thousands)
15
10
5
0
20
15
10
5
0
1
2
3
4
5
6
Weeks Forecasting
All Model Versions
Delphi
Los Alamos Nat Lab
Youyang Gu
Imperial
SIKJalpha
IHME - Curve Fit
IHME - CF SEIR
IHME - MS SEIR
300
200
100
0
Apr Jun Aug Oct Dec Feb
All Cumulative Errors
Delphi
Los Alamos Nat Lab
Youyang Gu
Imperial
SIKJalpha
IHME - Curve Fit
IHME - CF SEIR
IHME - MS SEIR
40
20
0
-20
Apr Jun Aug Oct Dec Feb

### Italy

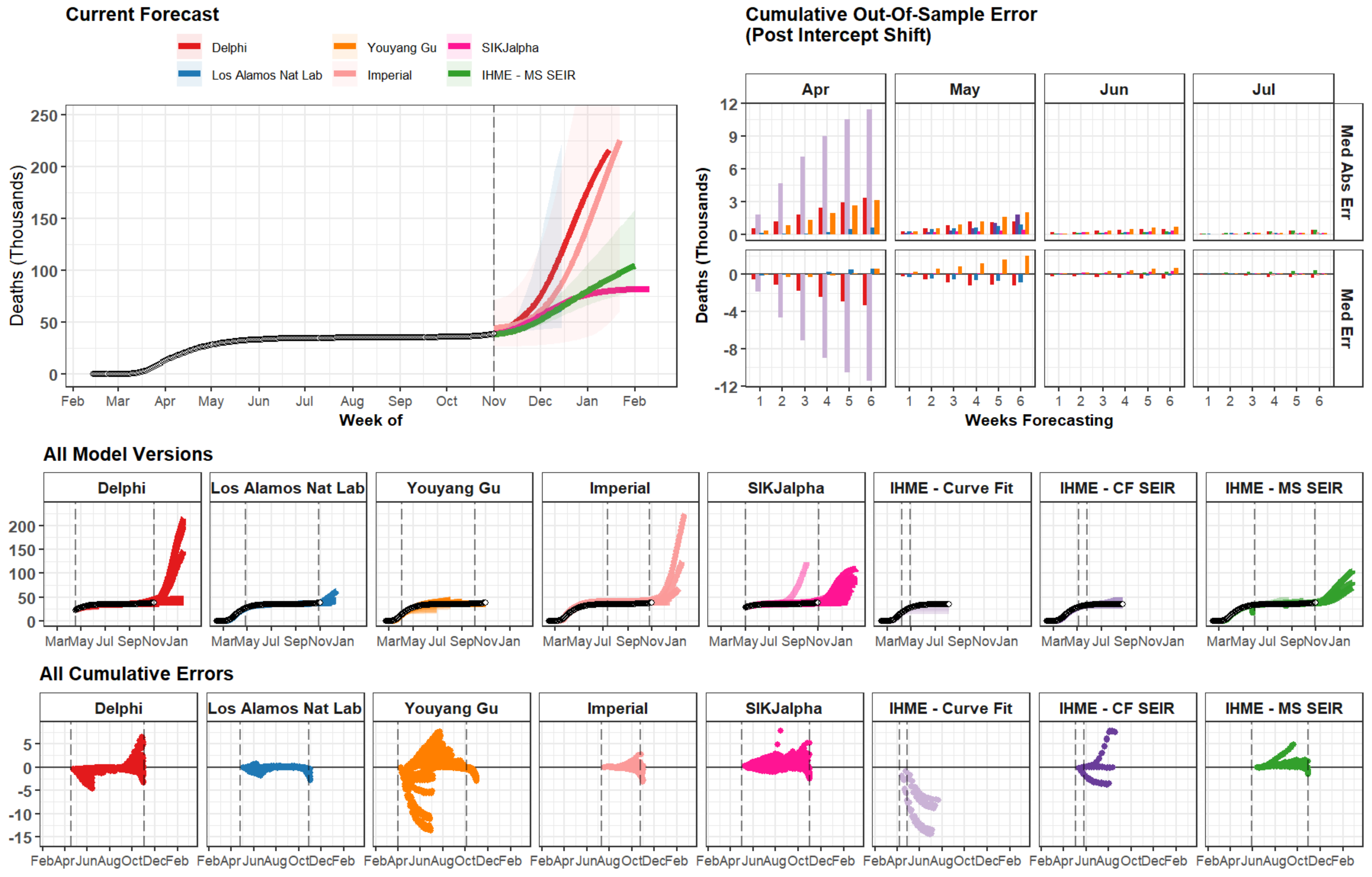
Current Forecast
Delphi
Youyang Gu
SIKJalpha
Los Alamos Nat Lab
Imperial
IHME - MS SEIR
Deaths (Thousands)
Week of
Cumulative Out-Of-Sample Error
(Post Intercept Shift)
Apr
May
Jun
Jul
Med Abs Err
Med Err
Weeks Forecasting
All Model Versions
Delphi
Los Alamos Nat Lab
Youyang Gu
Imperial
SIKJalpha
IHME - Curve Fit
IHME - CF SEIR
IHME - MS SEIR
All Cumulative Errors

### France

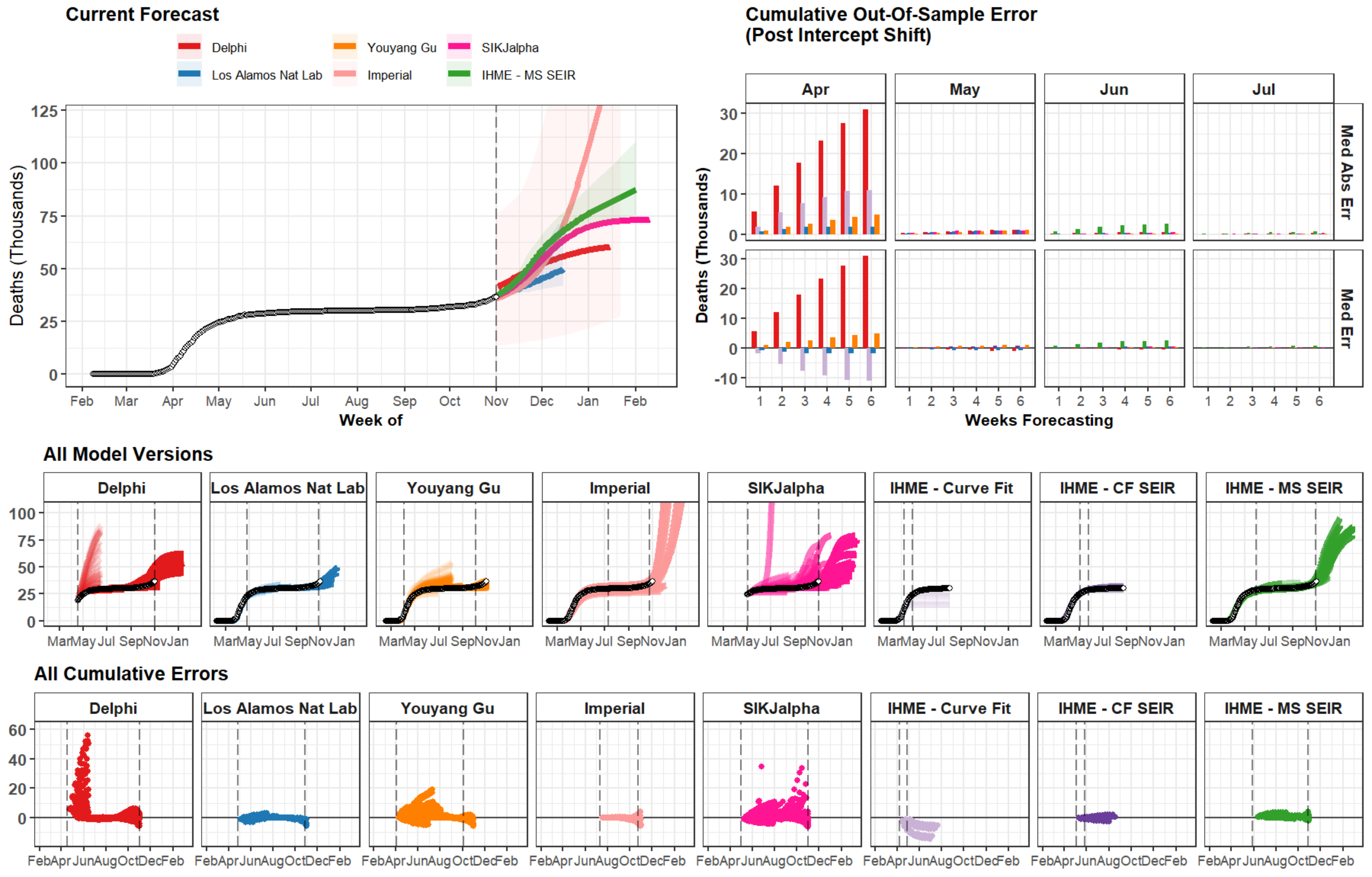

Current Forecast
Delphi
Youyang Gu
SIKJalpha
Los Alamos Nat Lab
Imperial
IHME - MS SEIR
Deaths (Thousands)
Week of
Feb Mar Apr May Jun Jul Aug Sep Oct Nov Dec Jan Feb
Cumulative Out-Of-Sample Error
(Post Intercept Shift)
Apr
May
Jun
Jul
Med Abs Err
Med Err
Weeks Forecasting
All Model Versions
Delphi
Los Alamos Nat Lab
Youyang Gu
Imperial
SIKJalpha
IHME - Curve Fit
IHME - CF SEIR
IHME - MS SEIR
All Cumulative Errors

### Spain

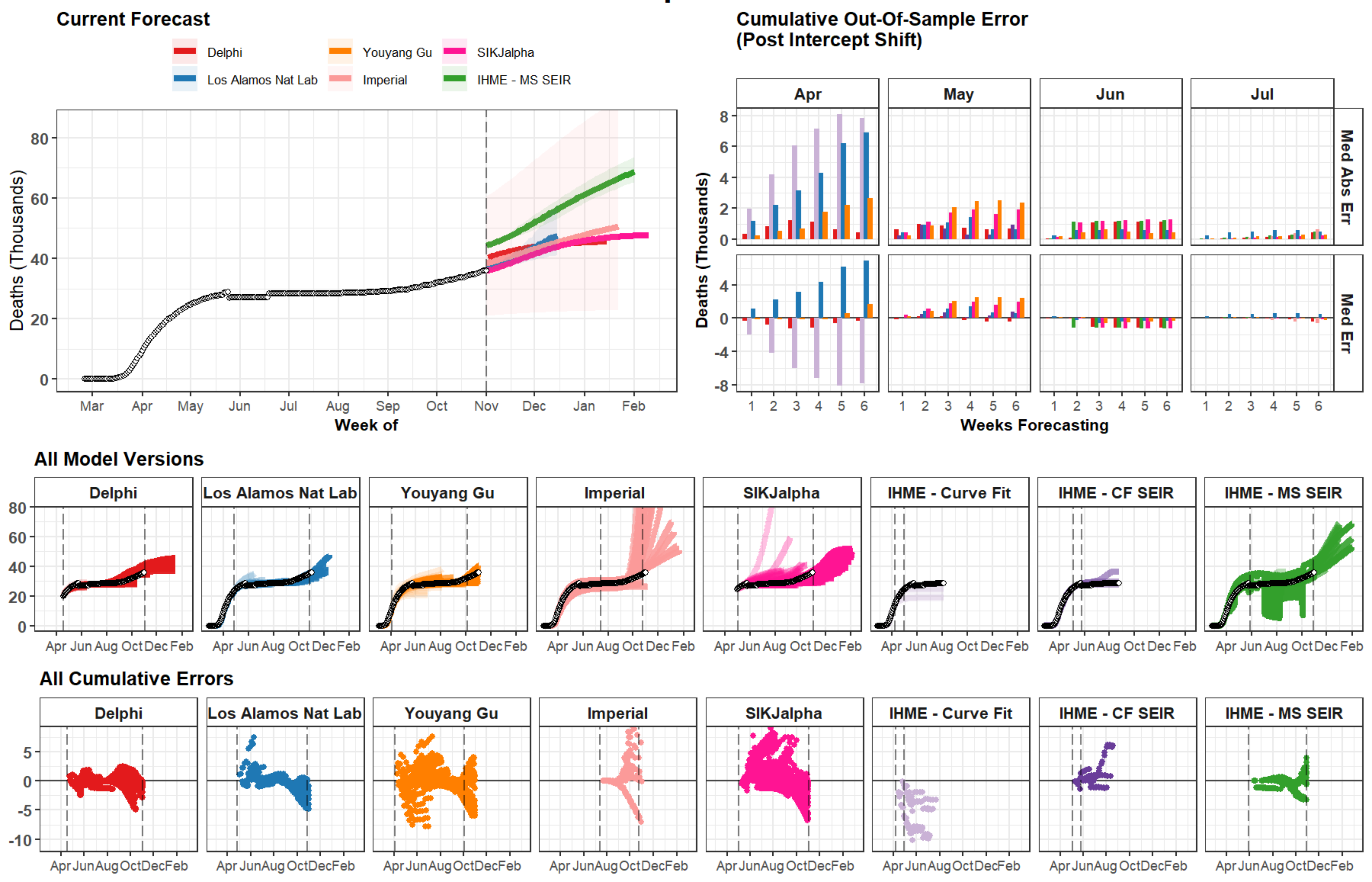
Current Forecast
Delphi
Youyang Gu
SIKJalpha
Los Alamos Nat Lab
Imperial
IHME - MS SEIR
Deaths (Thousands)
Week of
Mar
Apr
May
Jun
Jul
Aug
Sep
Oct
Nov
Dec
Jan
Feb
Cumulative Out-Of-Sample Error
(Post Intercept Shift)
Apr
May
Jun
Jul
Med Abs Err
Med Err
Weeks Forecasting
All Model Versions
Delphi
Los Alamos Nat Lab
Youyang Gu
Imperial
SIKJalpha
IHME - Curve Fit
IHME - CF SEIR
IHME - MS SEIR
All Cumulative Errors

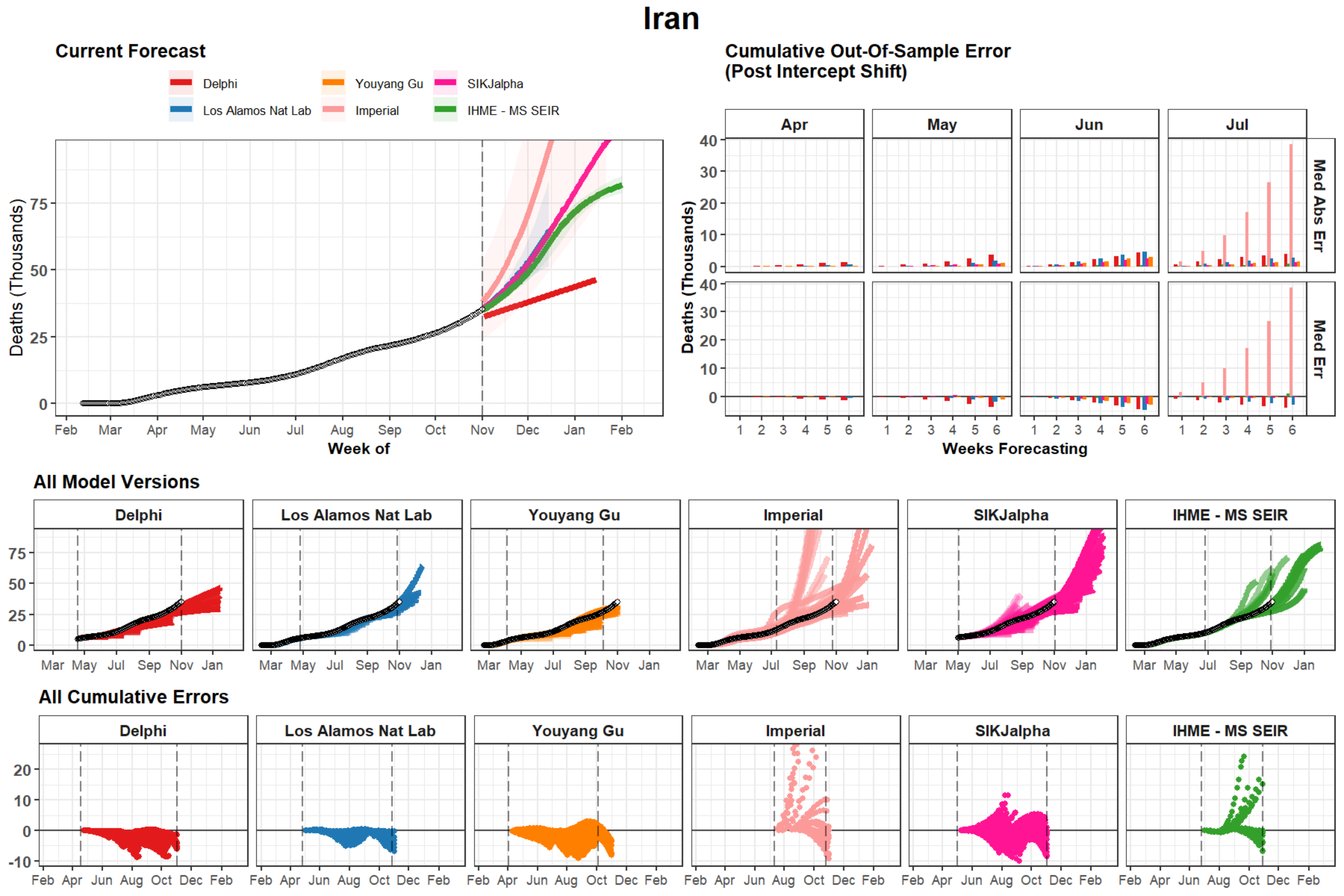

Iran
Current Forecast
Delphi
Los Alamos Nat Lab
Youyang Gu
Imperial
SIKJalpha
IHME - MS SEIR
Deaths (Thousands)
Week of
Feb Mar Apr May Jun Jul Aug Sep Oct Nov Dec Jan Feb
Cumulative Out-Of-Sample Error
(Post Intercept Shift)
Apr May Jun Jul
Med Abs Err
Med Err
Weeks Forecasting
All Model Versions
Delphi Los Alamos Nat Lab Youyang Gu Imperial SIKJalpha IHME - MS SEIR
Mar May Jul Sep Nov Jan
All Cumulative Errors
Feb Apr Jun Aug Oct Dec Feb

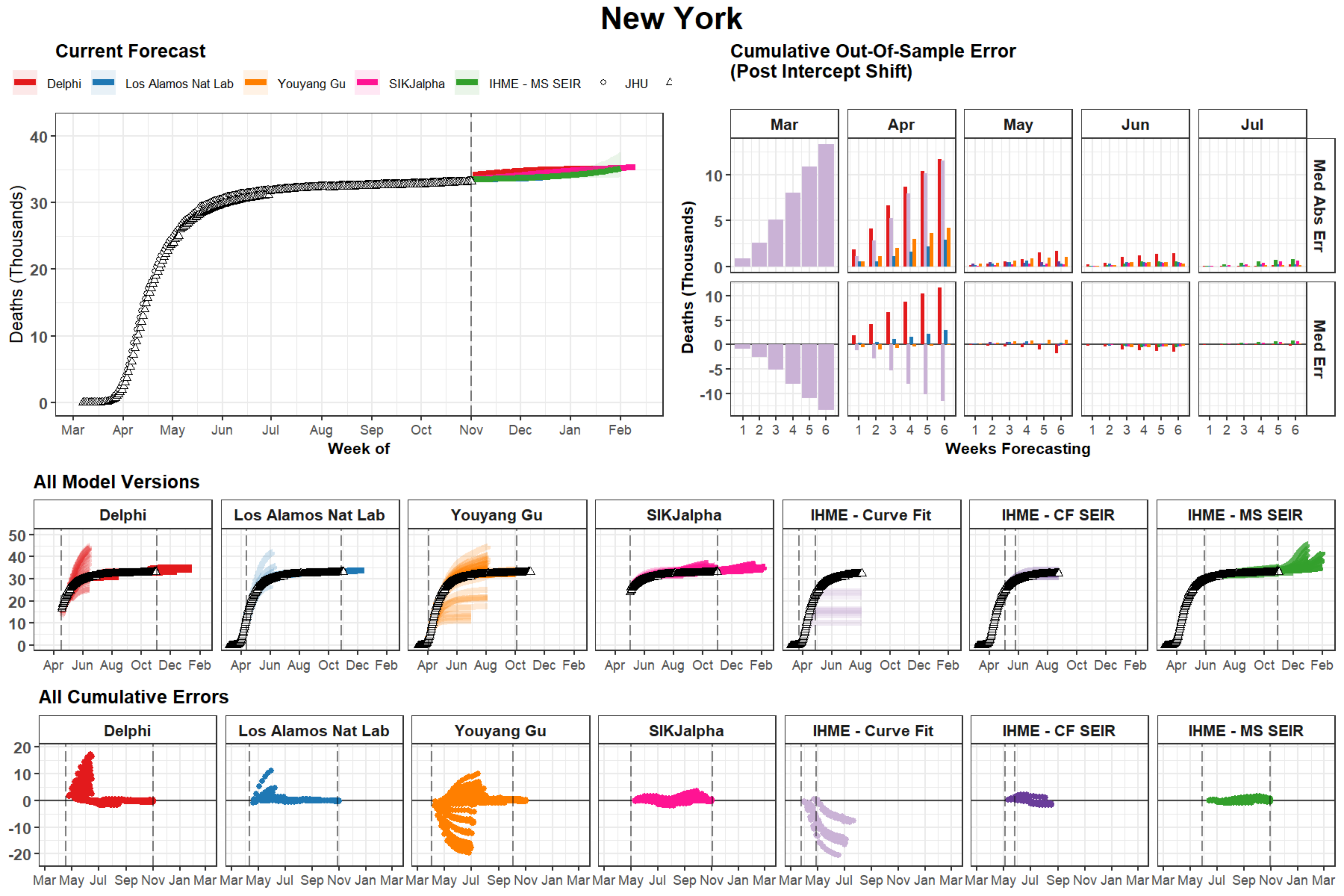
New York
Current Forecast
Delphi
Los Alamos Nat Lab
Youyang Gu
SIKJalpha
IHME - MS SEIR
JHU
Deaths (Thousands)
Week of
Mar
Apr
May
Jun
Jul
Aug
Sep
Oct
Nov
Dec
Jan
Feb
Cumulative Out-Of-Sample Error
(Post Intercept Shift)
Med Abs Err
Med Err
Weeks Forecasting
All Model Versions
IHME - Curve Fit
IHME - CF SEIR
All Cumulative Errors

### Colombia

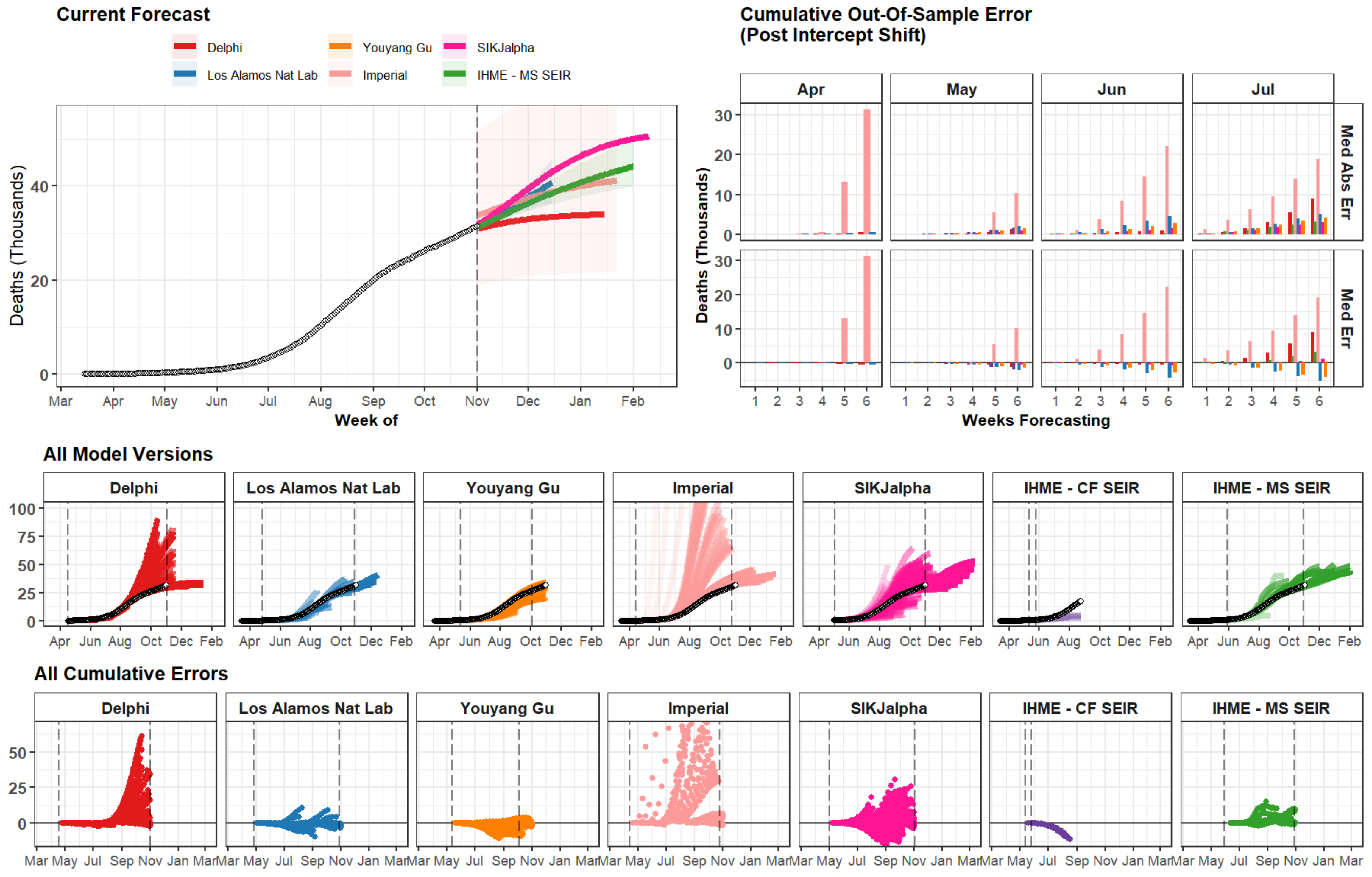

Current Forecast
Delphi
Los Alamos Nat Lab
Youyang Gu
Imperial
SIKJalpha
IHME - MS SEIR
Deaths (Thousands)
Week of
Cumulative Out-Of-Sample Error
(Post Intercept Shift)
Apr
May
Jun
Jul
Med Abs Err
Med Err
Weeks Forecasting
All Model Versions
Delphi
Los Alamos Nat Lab
Youyang Gu
Imperial
SIKJalpha
IHME - CF SEIR
IHME - MS SEIR
All Cumulative Errors

### Argentina

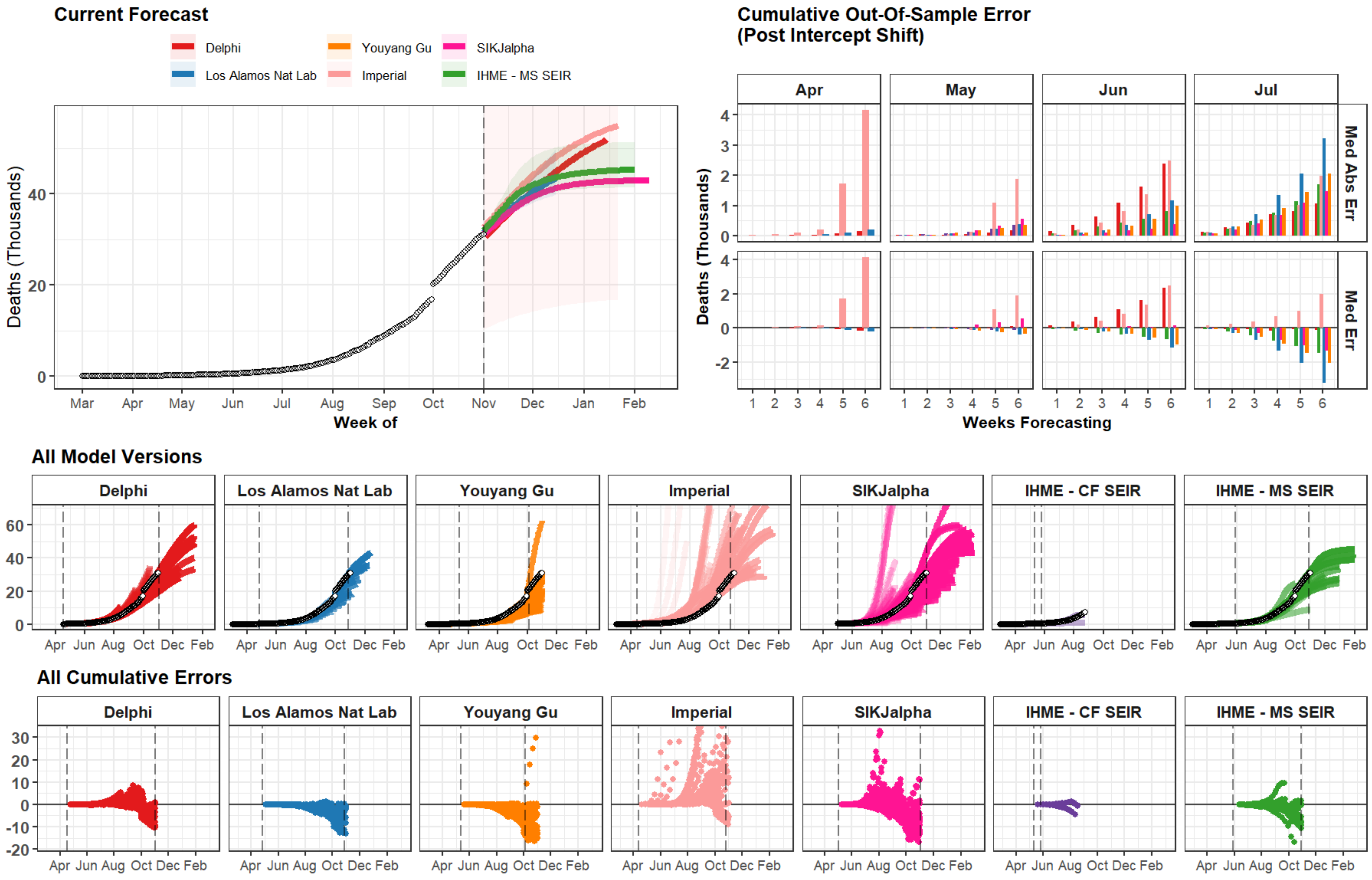

Current Forecast
Delphi
Los Alamos Nat Lab
Youyang Gu
Imperial
SIKJalpha
IHME - MS SEIR
Deaths (Thousands)
Week of
Mar Apr May Jun Jul Aug Sep Oct Nov Dec Jan Feb
Cumulative Out-Of-Sample Error
(Post Intercept Shift)
Apr May Jun Jul
Med Abs Err
Med Err
Weeks Forecasting
All Model Versions
Delphi
Los Alamos Nat Lab
Youyang Gu
Imperial
SIKJalpha
IHME - CF SEIR
IHME - MS SEIR
Apr Jun Aug Oct Dec Feb
All Cumulative Errors

### Russian Federation

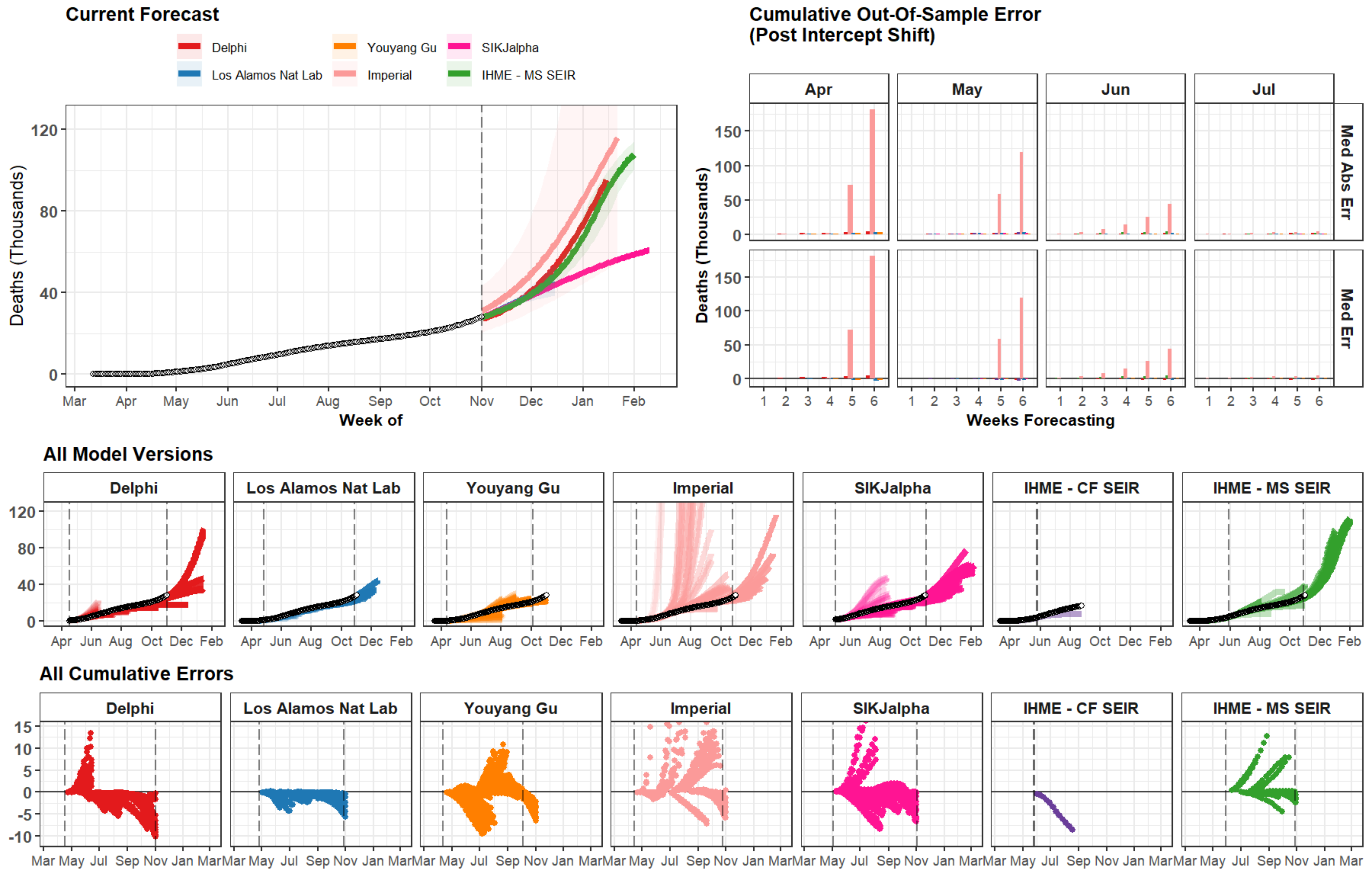

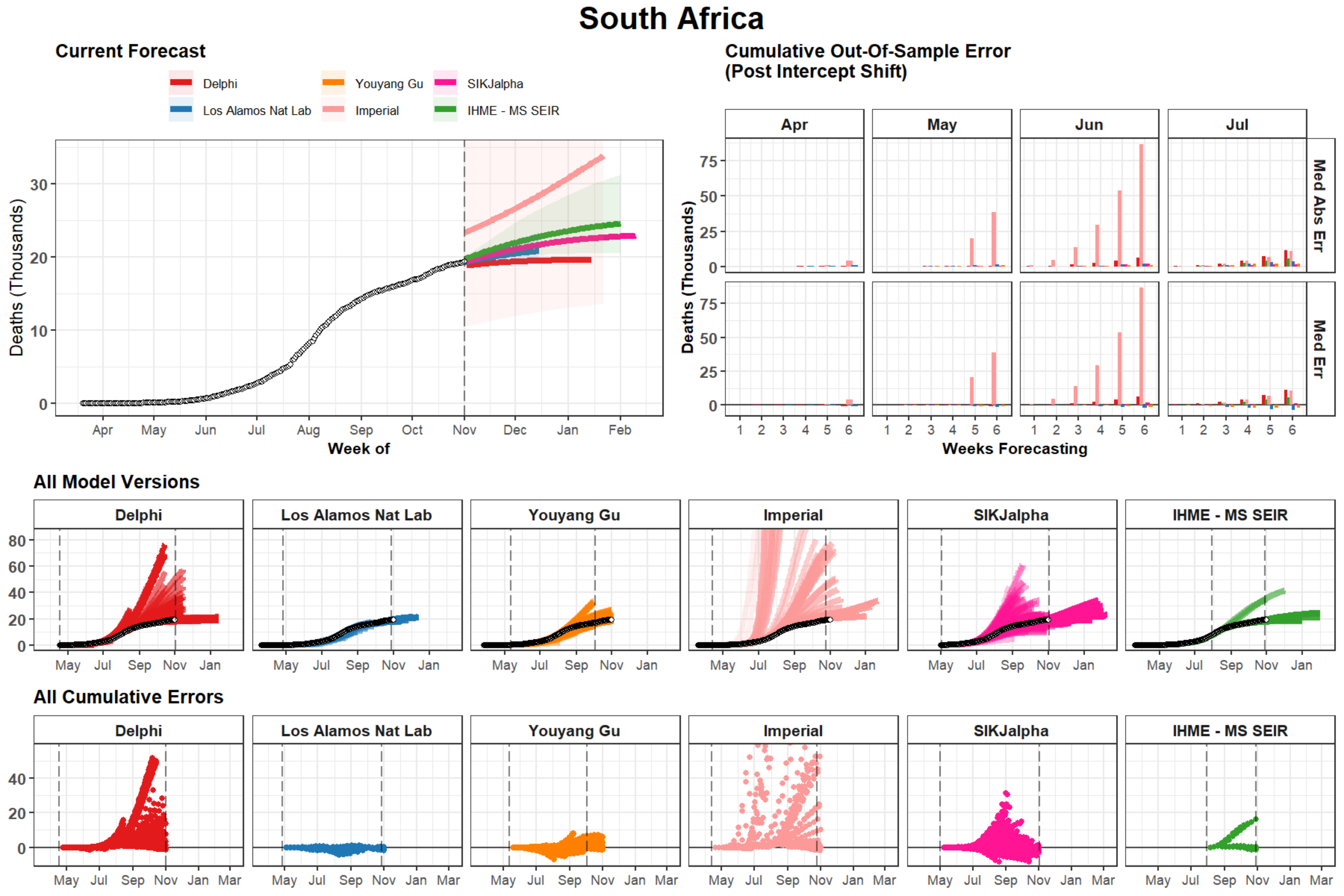

South Africa
Current Forecast
Delphi
Los Alamos Nat Lab
Youyang Gu
Imperial
SIKJalpha
IHME - MS SEIR
Deaths (Thousands)
Week of
Apr
May
Jun
Jul
Aug
Sep
Oct
Nov
Dec
Jan
Feb
Cumulative Out-Of-Sample Error
(Post Intercept Shift)
Med Abs Err
Med Err
Weeks Forecasting
All Model Versions
All Cumulative Errors
Mar

### Texas

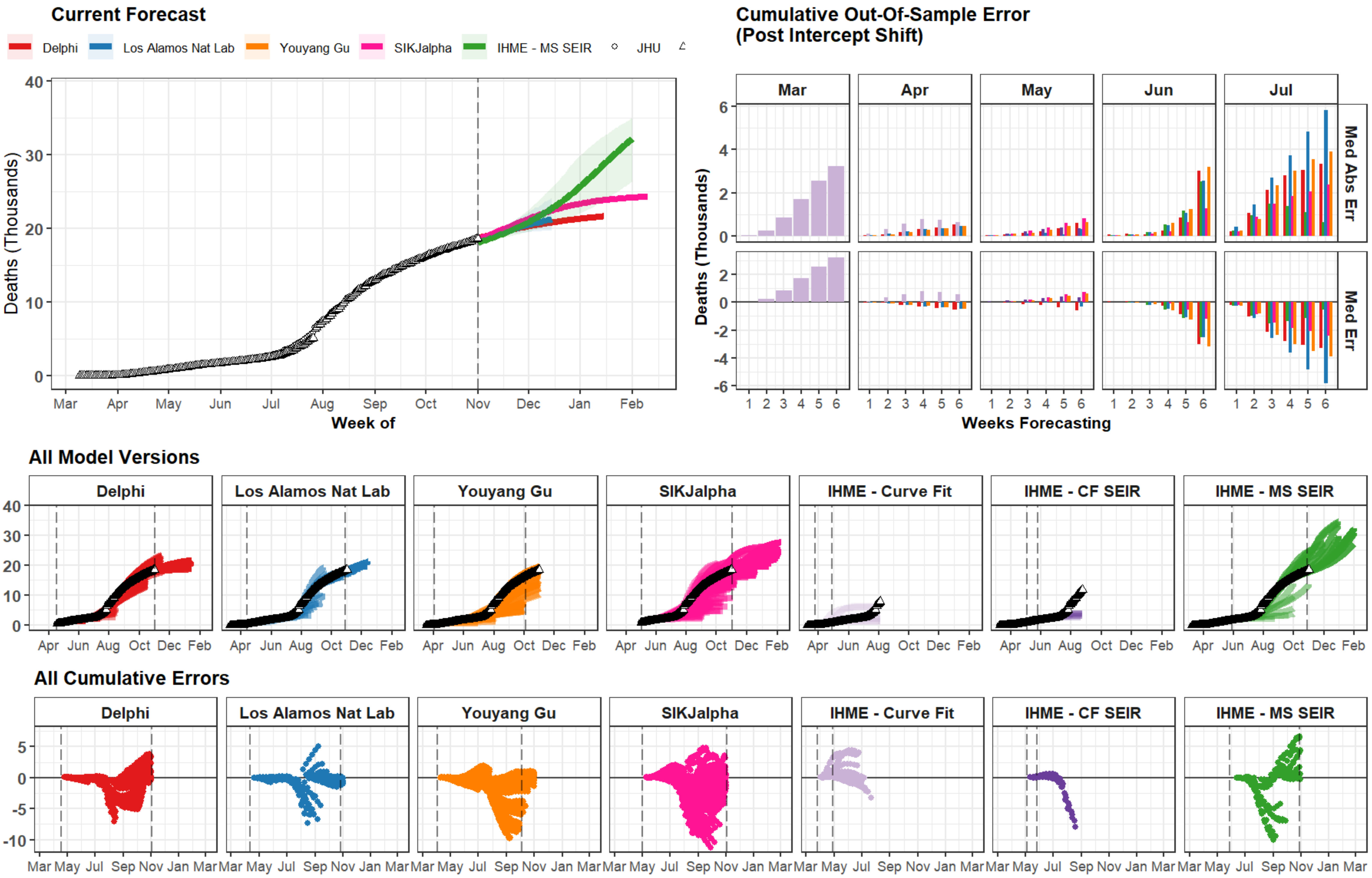
Current Forecast
Delphi
Los Alamos Nat Lab
Youyang Gu
SIKJalpha
IHME - MS SEIR
JHU
Deaths (Thousands)
Week of
Mar
Apr
May
Jun
Jul
Aug
Sep
Oct
Nov
Dec
Jan
Feb
Cumulative Out-Of-Sample Error
(Post Intercept Shift)
Med Abs Err
Med Err
Weeks Forecasting
All Model Versions
IHME - Curve Fit
IHME - CF SEIR
All Cumulative Errors

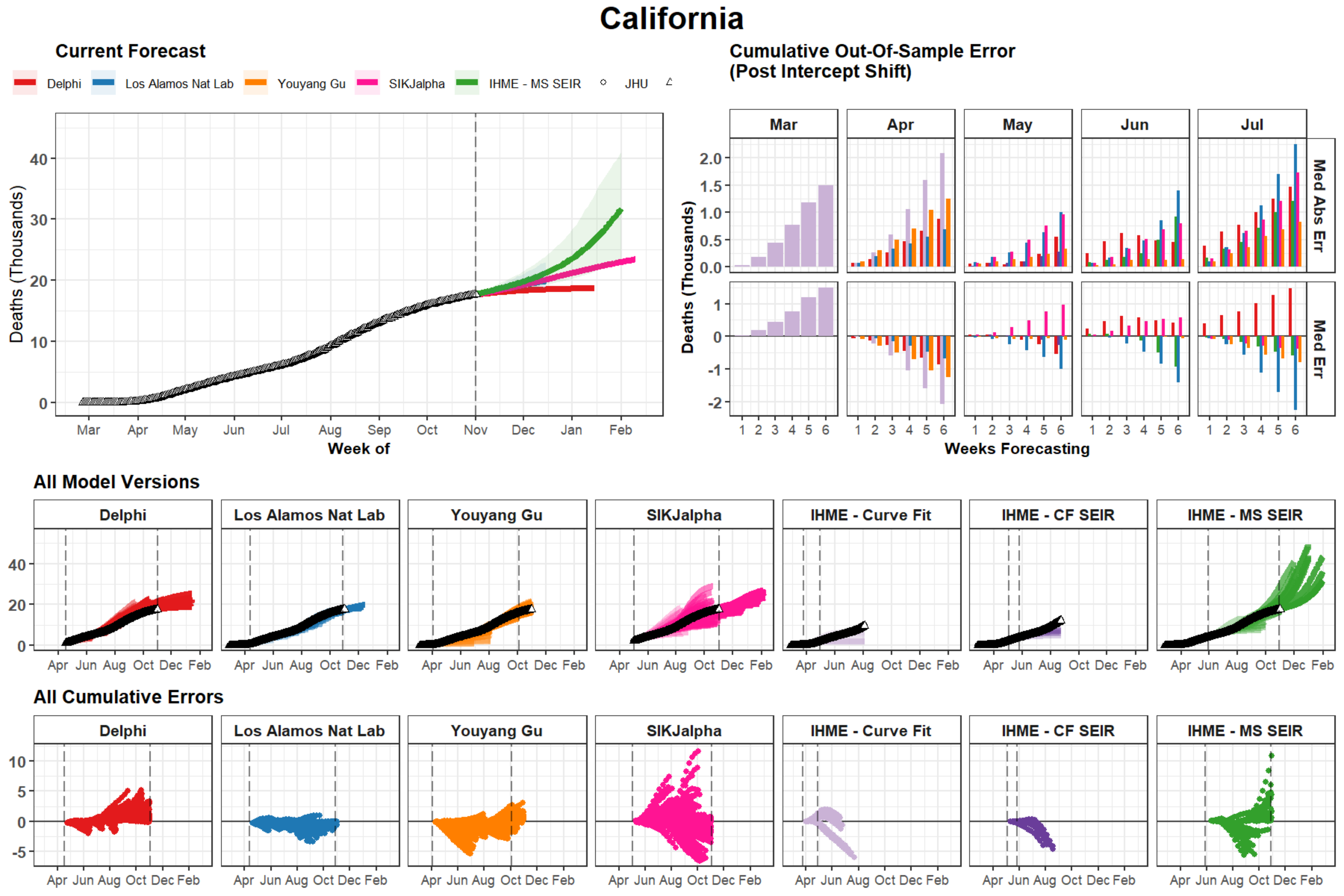
California
Current Forecast
Delphi
Los Alamos Nat Lab
Youyang Gu
SIKJalpha
IHME - MS SEIR
JHU
Deaths (Thousands)
Week of
Mar Apr May Jun Jul Aug Sep Oct Nov Dec Jan Feb
Cumulative Out-Of-Sample Error
(Post Intercept Shift)
Mar Apr May Jun Jul
Med Abs Err
Med Err
Weeks Forecasting
All Model Versions
Delphi
Los Alamos Nat Lab
Youyang Gu
SIKJalpha
IHME - Curve Fit
IHME - CF SEIR
IHME - MS SEIR
Apr Jun Aug Oct Dec Feb
All Cumulative Errors
Delphi
Los Alamos Nat Lab
Youyang Gu
SIKJalpha
IHME - Curve Fit
IHME - CF SEIR
IHME - MS SEIR
Apr Jun Aug Oct Dec Feb

### Florida

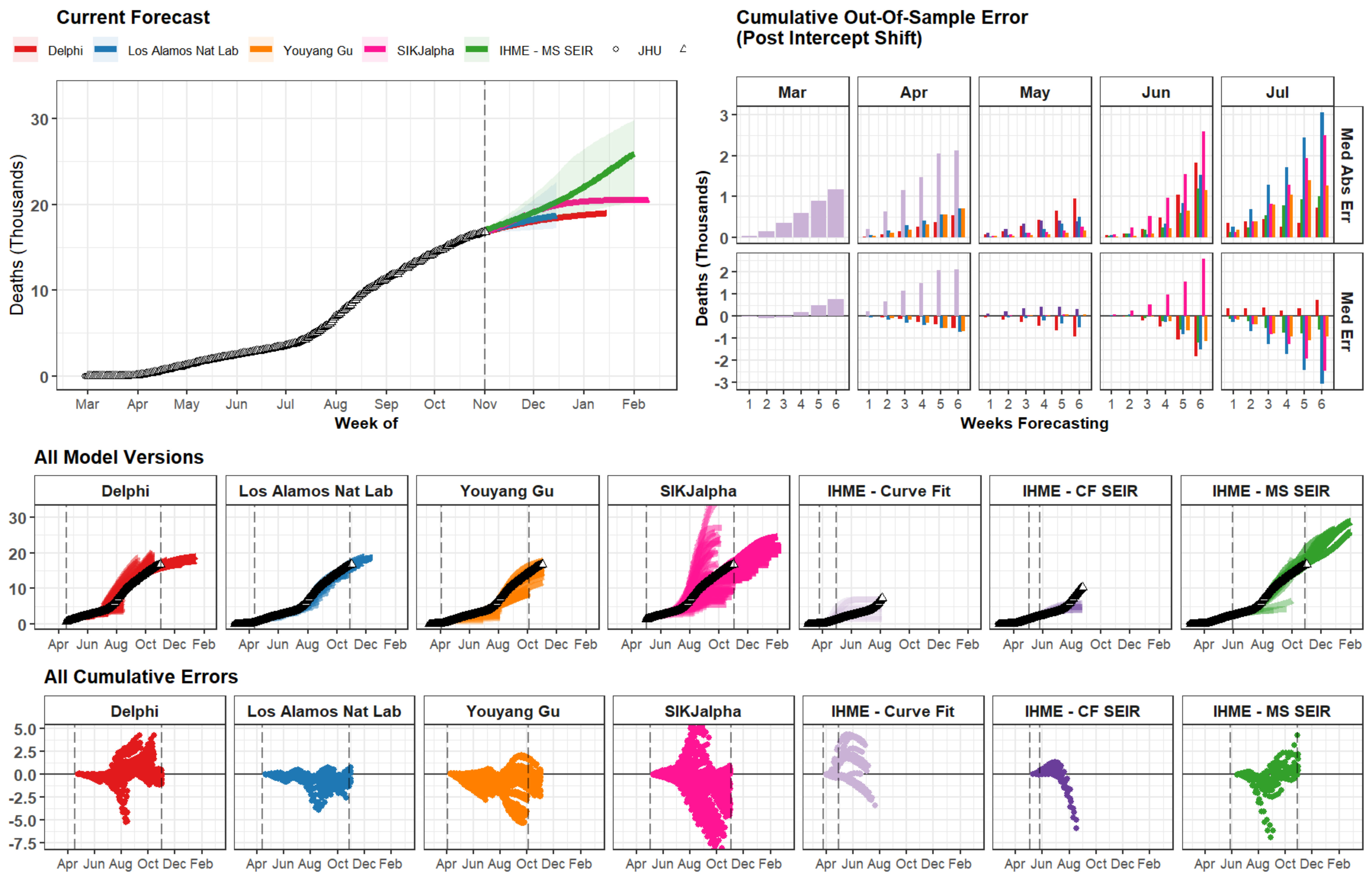

Current Forecast
Delphi
Los Alamos Nat Lab
Youyang Gu
SIKJalpha
IHME - MS SEIR
JHU
Deaths (Thousands)
30
20
10
0
Mar Apr May Jun Jul Aug Sep Oct Nov Dec Jan Feb
Week of
Cumulative Out-Of-Sample Error
(Post Intercept Shift)
Mar
Apr
May
Jun
Jul
Med Abs Err
Med Err
Deaths (Thousands)
3 2 1 0
2 1 0 -1 -2 -3
1 2 3 4 5 6
Weeks Forecasting
All Model Versions
Delphi
Los Alamos Nat Lab
Youyang Gu
SIKJalpha
IHME - Curve Fit
IHME - CF SEIR
IHME - MS SEIR
30 20 10 0
Apr Jun Aug Oct Dec Feb
All Cumulative Errors
Delphi
Los Alamos Nat Lab
Youyang Gu
SIKJalpha
IHME - Curve Fit
IHME - CF SEIR
IHME - MS SEIR
5.0 2.5 0.0 -2.5 -5.0 -7.5
Apr Jun Aug Oct Dec Feb

### New Jersey

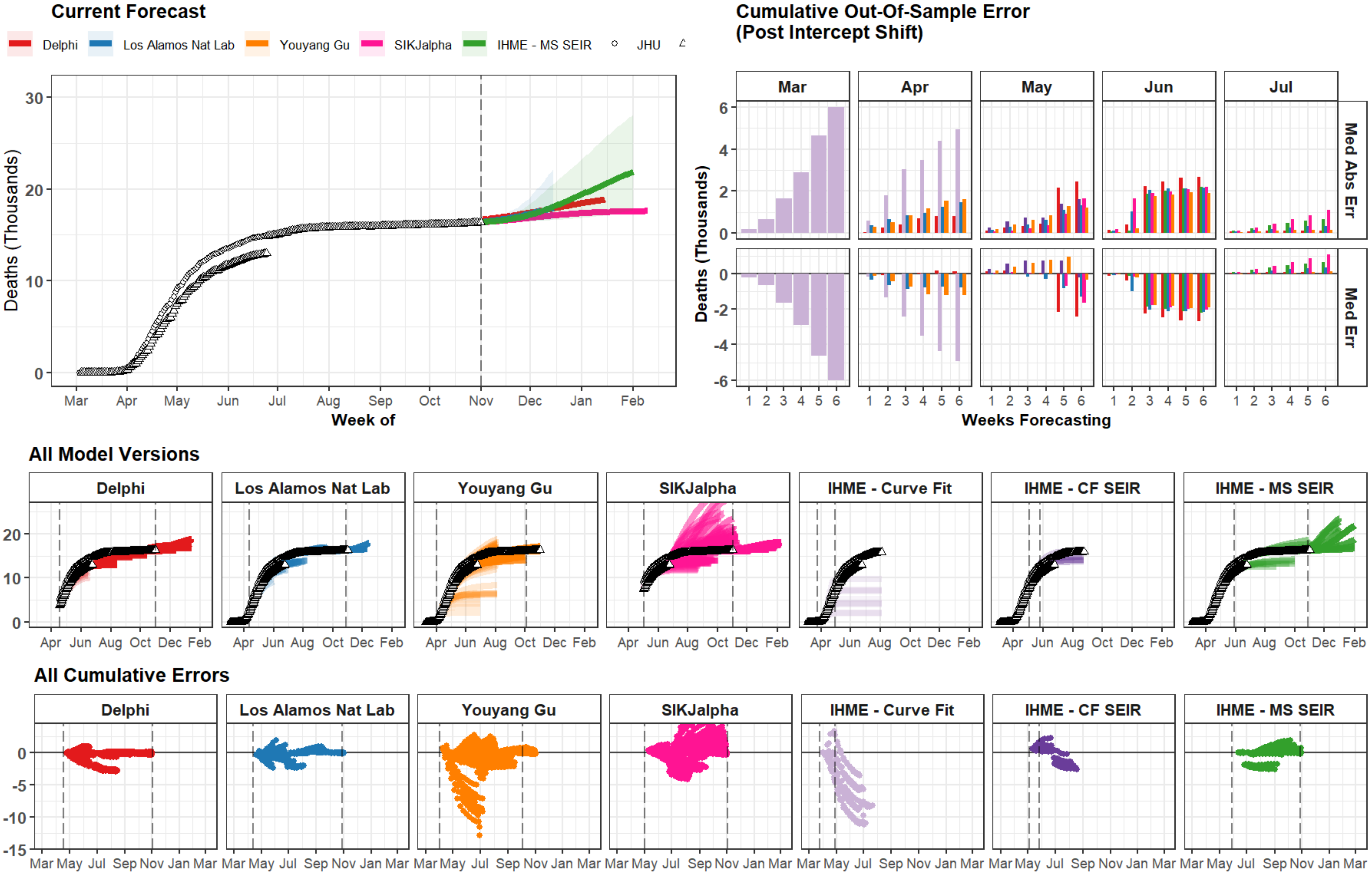
Current Forecast
Delphi
Los Alamos Nat Lab
Youyang Gu
SIKJalpha
IHME - MS SEIR
JHU
Deaths (Thousands)
Week of
Mar
Apr
May
Jun
Jul
Aug
Sep
Oct
Nov
Dec
Jan
Feb
Cumulative Out-Of-Sample Error
(Post Intercept Shift)
Med Abs Err
Med Err
Weeks Forecasting
All Model Versions
IHME - Curve Fit
IHME - CF SEIR
All Cumulative Errors

### Chile

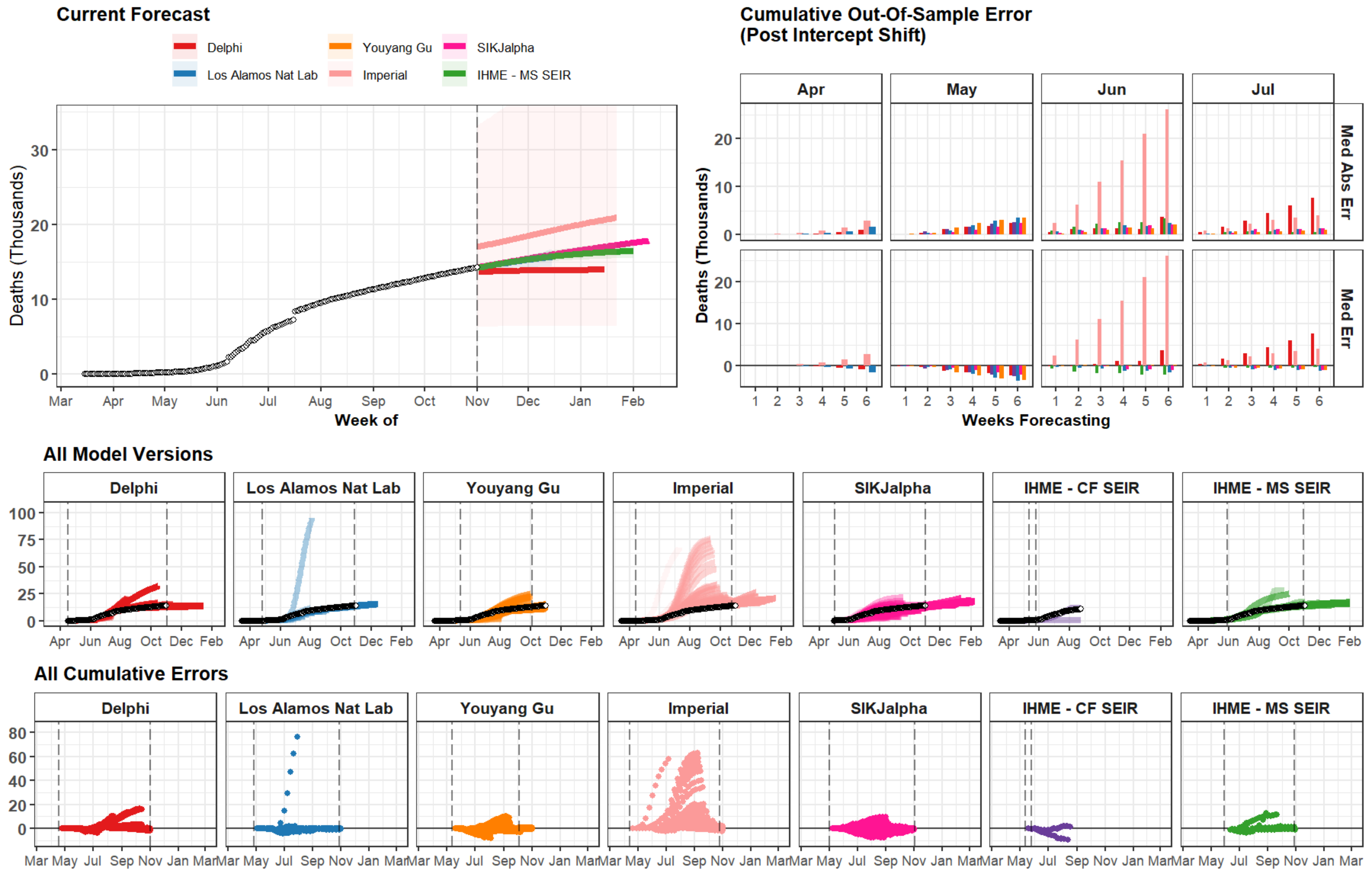

Current Forecast
Delphi
Youyang Gu
SIKJalpha
Los Alamos Nat Lab
Imperial
IHME - MS SEIR
Deaths (Thousands)
Week of
Mar Apr May Jun Jul Aug Sep Oct Nov Dec Jan Feb
Cumulative Out-Of-Sample Error (Post Intercept Shift)
Apr May Jun Jul
Med Abs Err
Med Err
Weeks Forecasting
All Model Versions
Delphi
Los Alamos Nat Lab
Youyang Gu
Imperial
SIKJalpha
IHME - CF SEIR
IHME - MS SEIR
All Cumulative Errors

### Indonesia

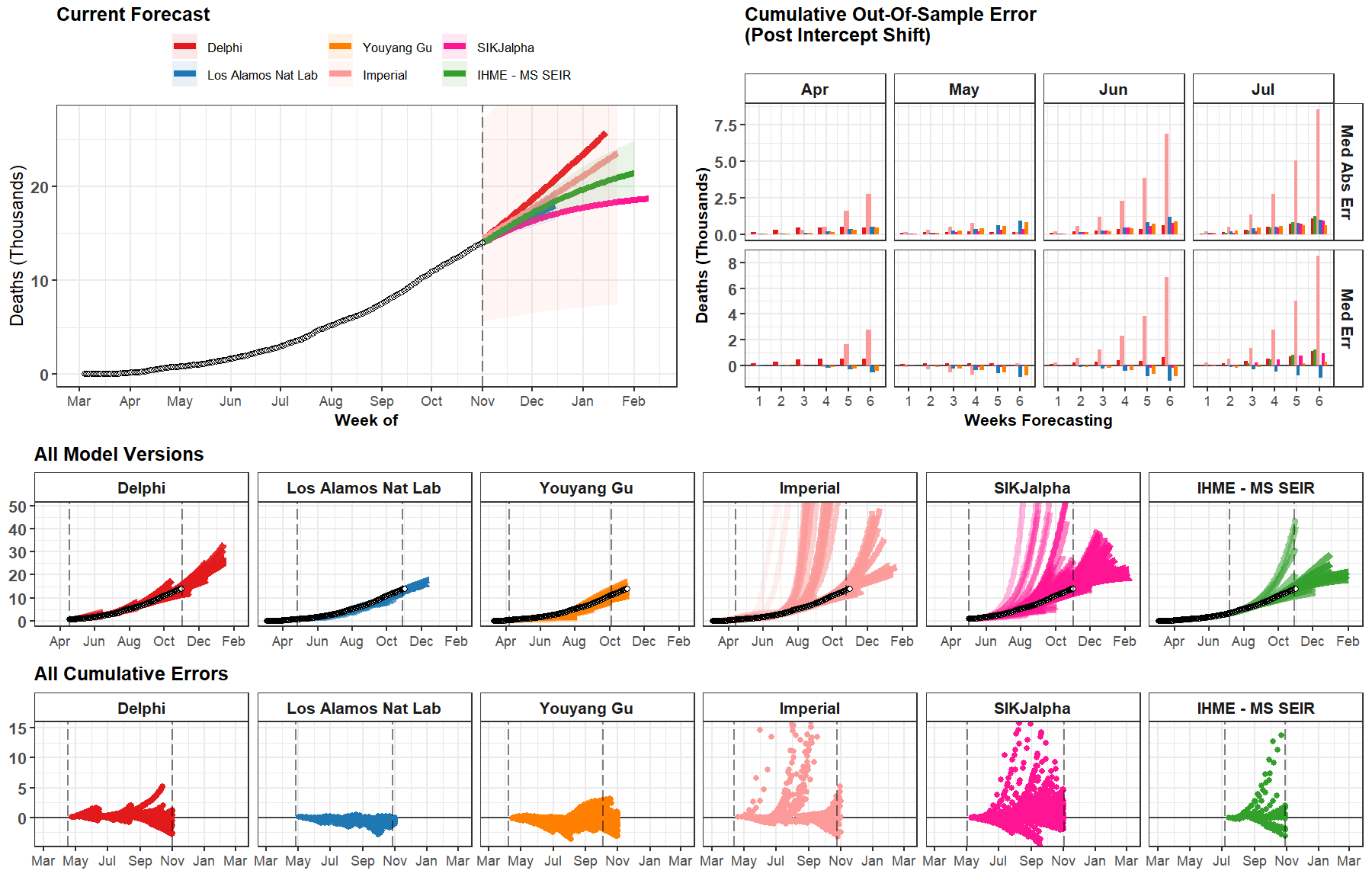

Current Forecast
Delphi
Los Alamos Nat Lab
Youyang Gu
Imperial
SIKJalpha
IHME - MS SEIR
Deaths (Thousands)
0
10
20
Mar
Apr
May
Jun
Jul
Aug
Sep
Oct
Nov
Dec
Jan
Feb
Week of
Cumulative Out-Of-Sample Error
(Post Intercept Shift)
Apr
May
Jun
Jul
Med Abs Err
Med Err
0.0
2.5
5.0
7.5
0
2
4
6
8
1
2
3
4
5
6
Weeks Forecasting
All Model Versions
Delphi
Los Alamos Nat Lab
Youyang Gu
Imperial
SIKJalpha
IHME - MS SEIR
0
10
20
30
40
50
Apr
Jun
Aug
Oct
Dec
Feb
All Cumulative Errors
Delphi
Los Alamos Nat Lab
Youyang Gu
Imperial
SIKJalpha
IHME - MS SEIR
0
5
10
15
Mar
May
Jul
Sep
Nov
Jan
Mar

### Belgium

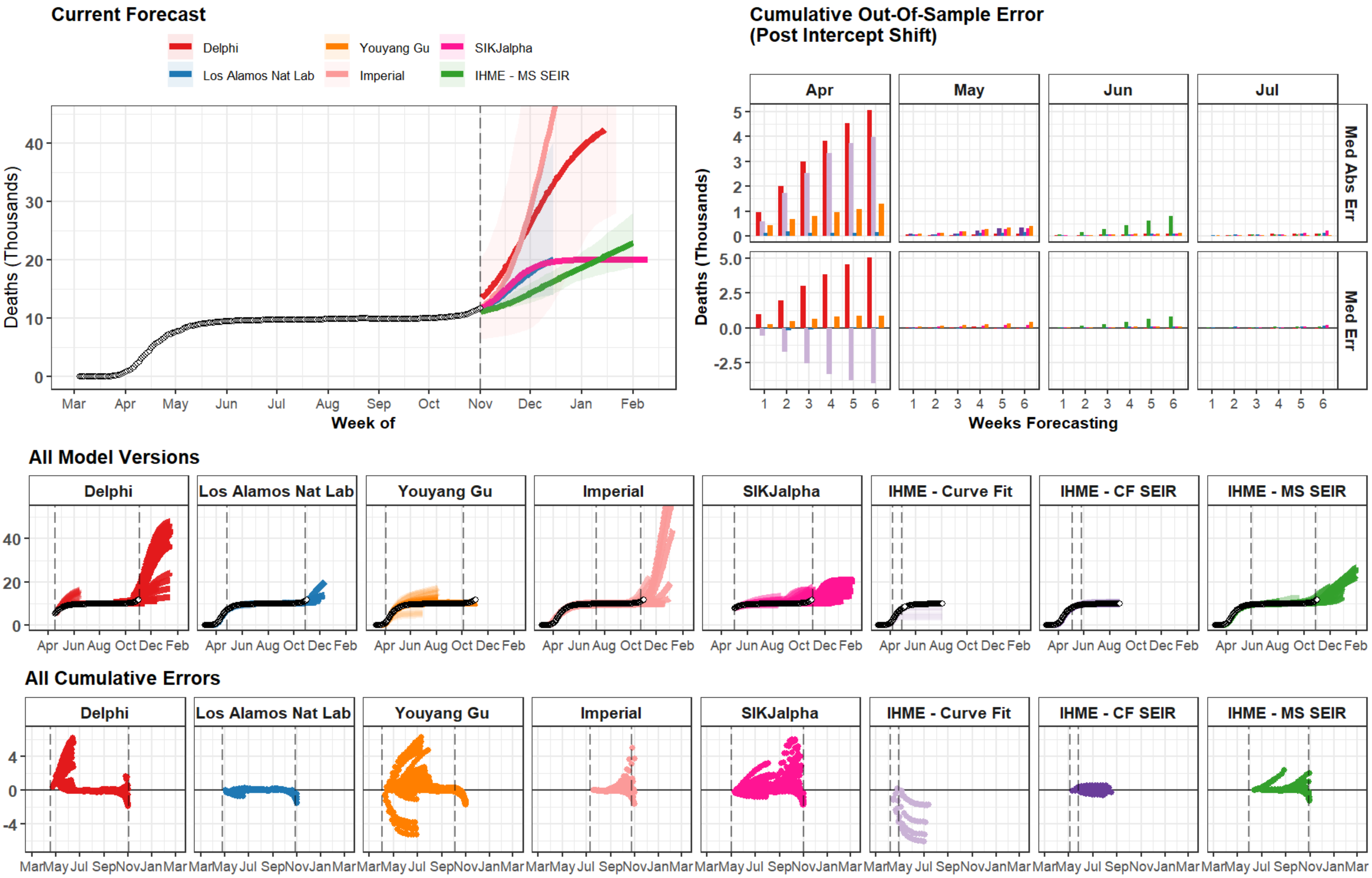
Current Forecast
Delphi
Los Alamos Nat Lab
Youyang Gu
Imperial
SIKJalpha
IHME - MS SEIR
Deaths (Thousands)
Week of
Mar Apr May Jun Jul Aug Sep Oct Nov Dec Jan Feb
Cumulative Out-Of-Sample Error
(Post Intercept Shift)
Apr
May
Jun
Jul
Med Abs Err
Med Err
Weeks Forecasting
All Model Versions
Delphi
Los Alamos Nat Lab
Youyang Gu
Imperial
SIKJalpha
IHME - Curve Fit
IHME - CF SEIR
IHME - MS SEIR
All Cumulative Errors

### Iraq

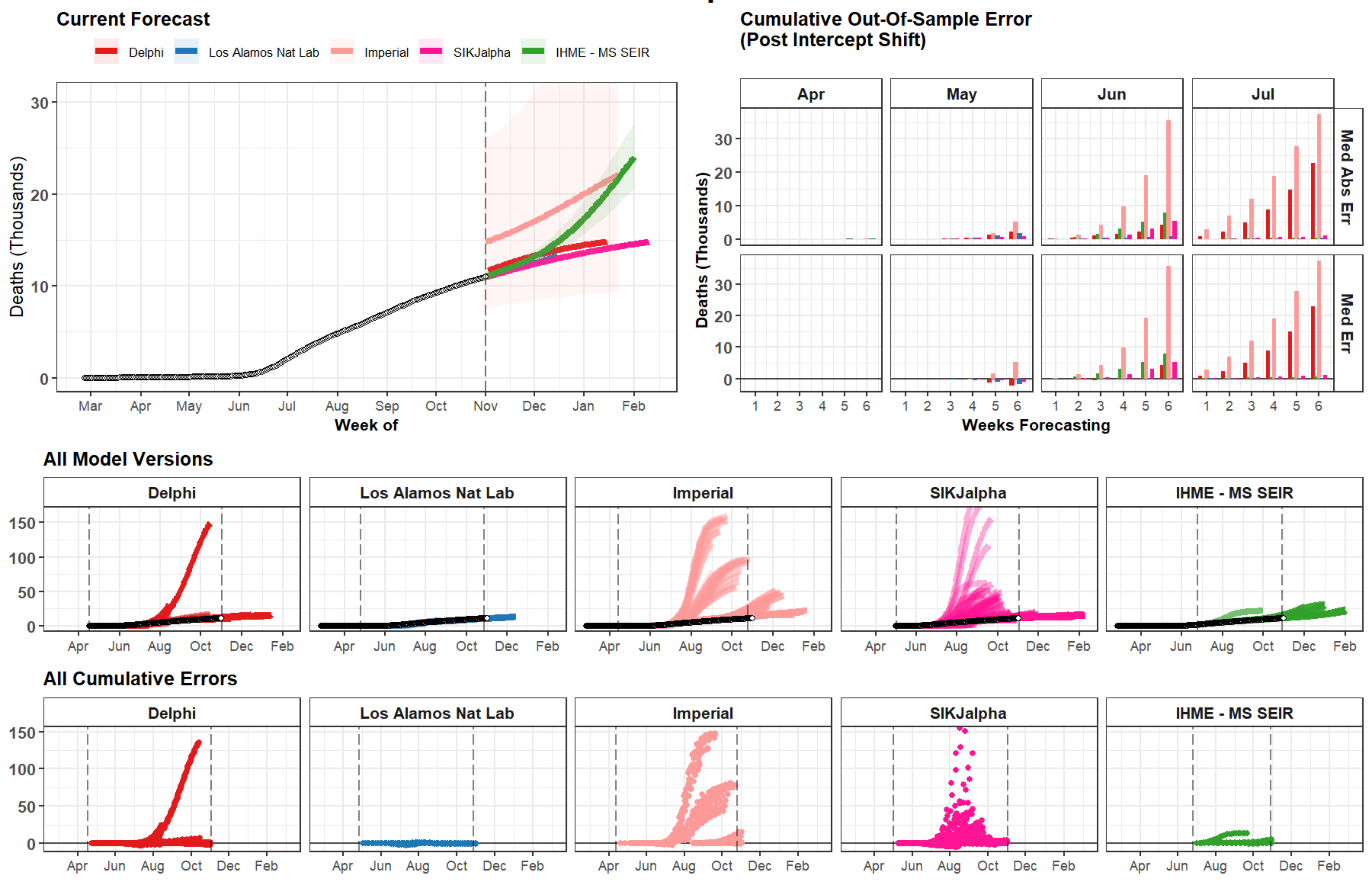
Current Forecast
Delphi
Los Alamos Nat Lab
Imperial
SIKJalpha
IHME - MS SEIR
Deaths (Thousands)
Week of
Mar
Apr
May
Jun
Jul
Aug
Sep
Oct
Nov
Dec
Jan
Feb
Cumulative Out-Of-Sample Error
(Post Intercept Shift)
Apr
May
Jun
Jul
Med Abs Err
Med Err
Weeks Forecasting
All Model Versions
All Cumulative Errors

### Germany

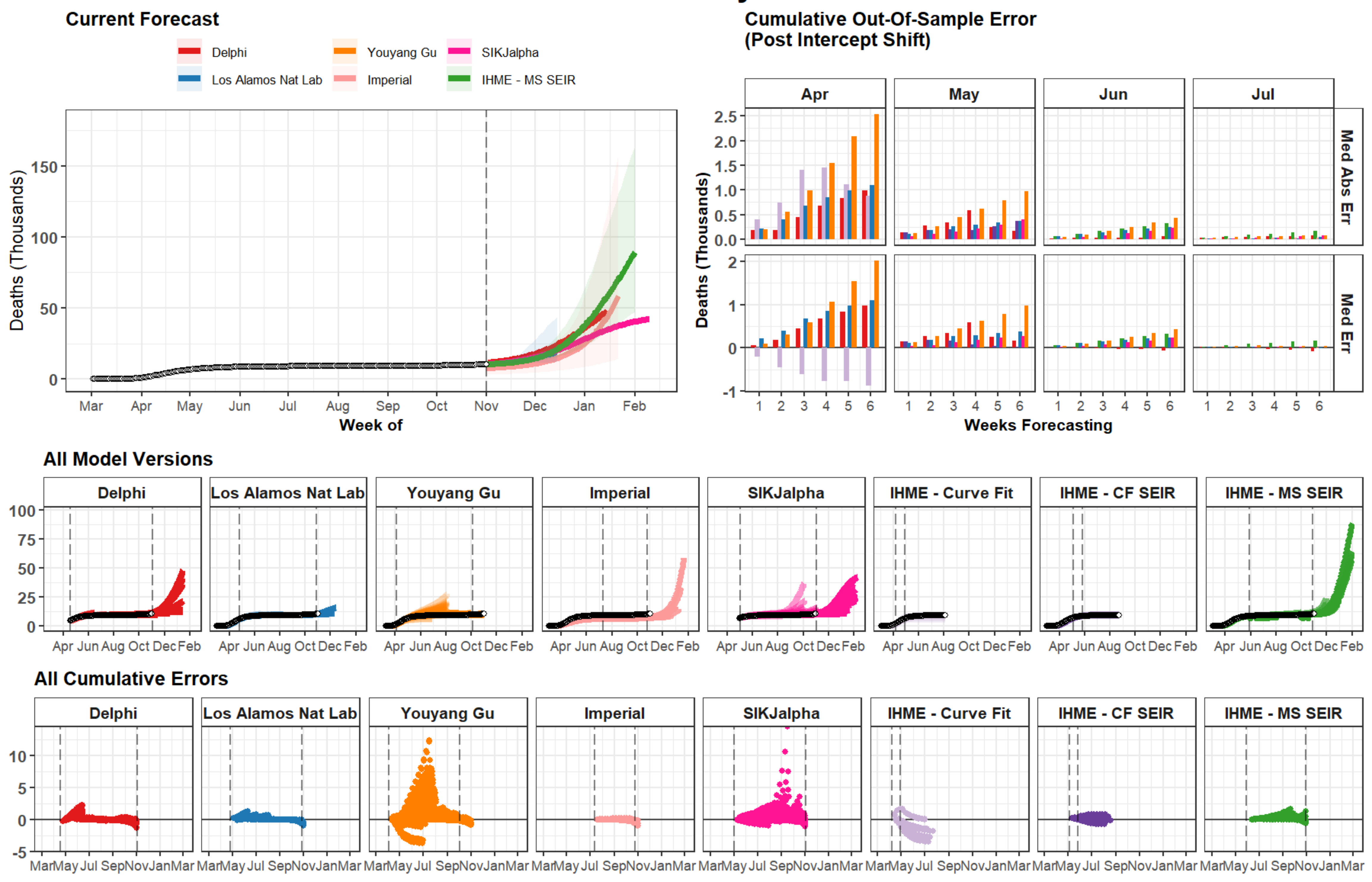
Germany
Current Forecast
Delphi
Youyang Gu
SIKJalpha
Los Alamos Nat Lab
Imperial
IHME - MS SEIR
Deaths (Thousands)
Week of
Mar Apr May Jun Jul Aug Sep Oct Nov Dec Jan Feb
Cumulative Out-Of-Sample Error
(Post Intercept Shift)
Apr May Jun Jul
Med Abs Err
Med Err
Weeks Forecasting
All Model Versions
Delphi
Los Alamos Nat Lab
Youyang Gu
Imperial
SIKJalpha
IHME - Curve Fit
IHME - CF SEIR
IHME - MS SEIR
All Cumulative Errors

### Turkey

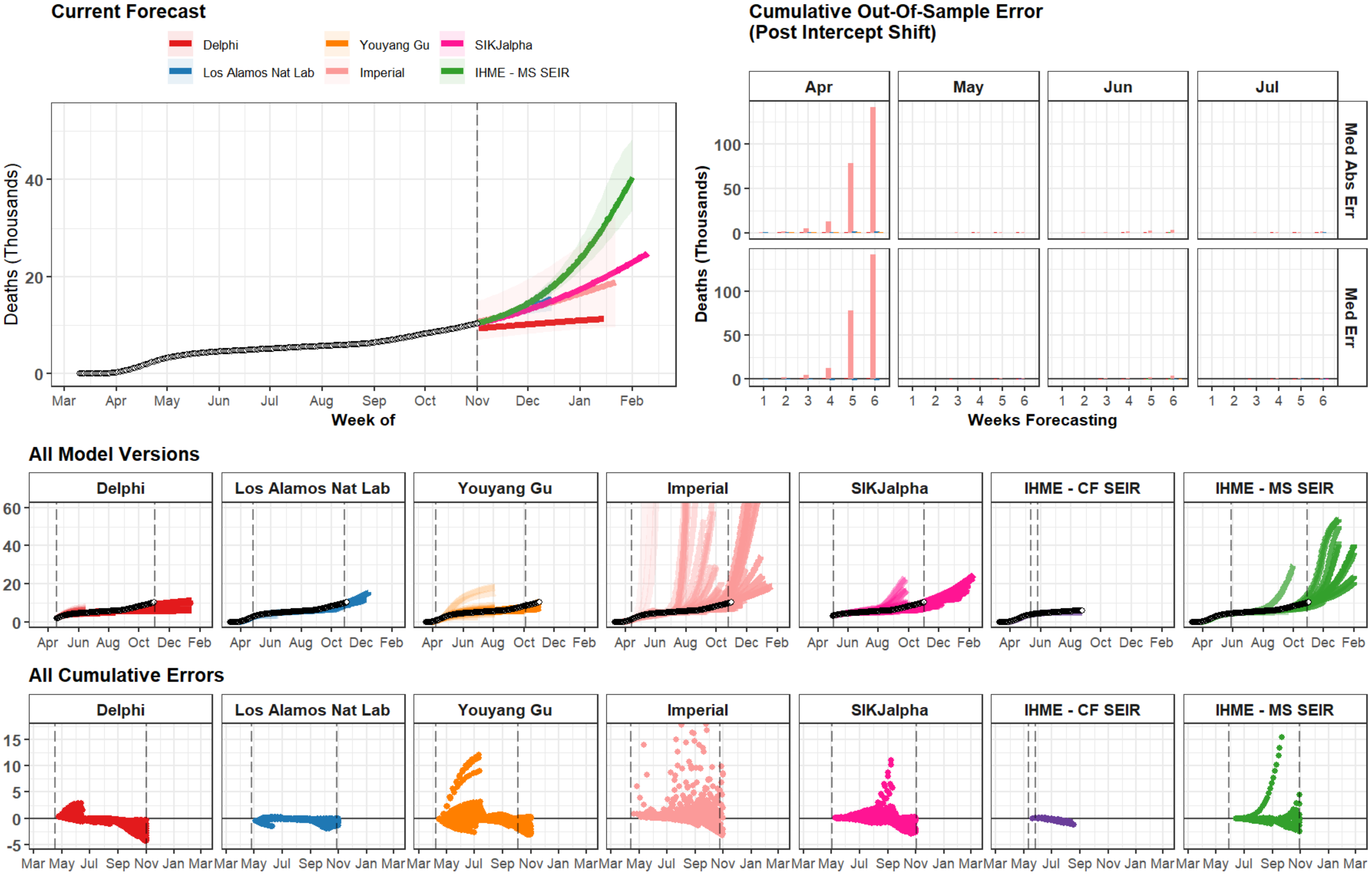
Current Forecast
Delphi
Los Alamos Nat Lab
Youyang Gu
Imperial
SIKJalpha
IHME - MS SEIR
Deaths (Thousands)
Week of
Mar Apr May Jun Jul Aug Sep Oct Nov Dec Jan Feb
Cumulative Out-Of-Sample Error
(Post Intercept Shift)
Apr
May
Jun
Jul
Med Abs Err
Med Err
Weeks Forecasting
All Model Versions
Delphi
Los Alamos Nat Lab
Youyang Gu
Imperial
SIKJalpha
IHME - CF SEIR
IHME - MS SEIR
All Cumulative Errors

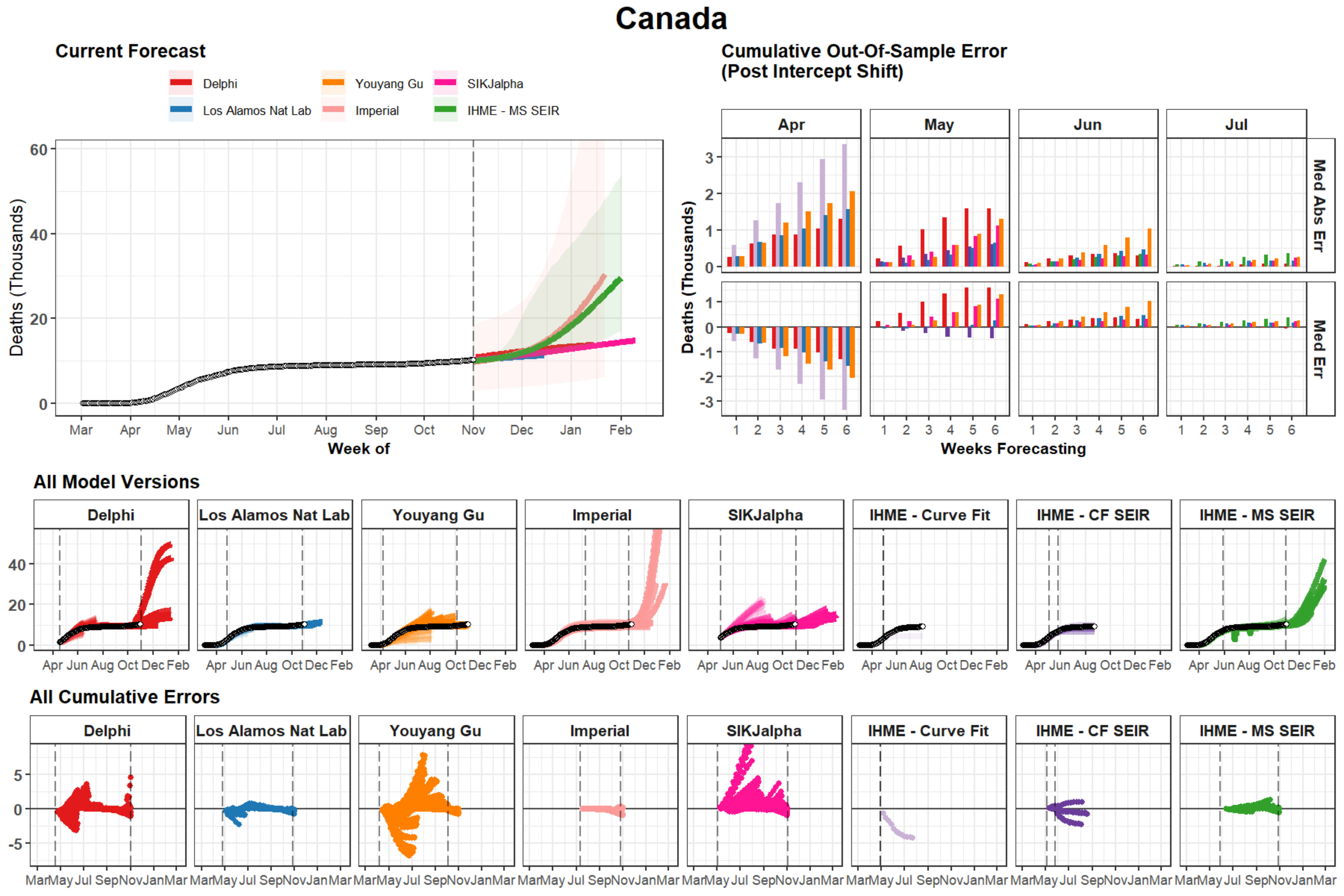
Canada
Current Forecast
Delphi
Youyang Gu
SIKJalpha
Los Alamos Nat Lab
Imperial
IHME - MS SEIR
Deaths (Thousands)
Week of
Mar Apr May Jun Jul Aug Sep Oct Nov Dec Jan Feb
Cumulative Out-Of-Sample Error
(Post Intercept Shift)
Apr May Jun Jul
Med Abs Err
Med Err
Weeks Forecasting
All Model Versions
Delphi
Los Alamos Nat Lab
Youyang Gu
Imperial
SIKJalpha
IHME - Curve Fit
IHME - CF SEIR
IHME - MS SEIR
All Cumulative Errors

### Illinois

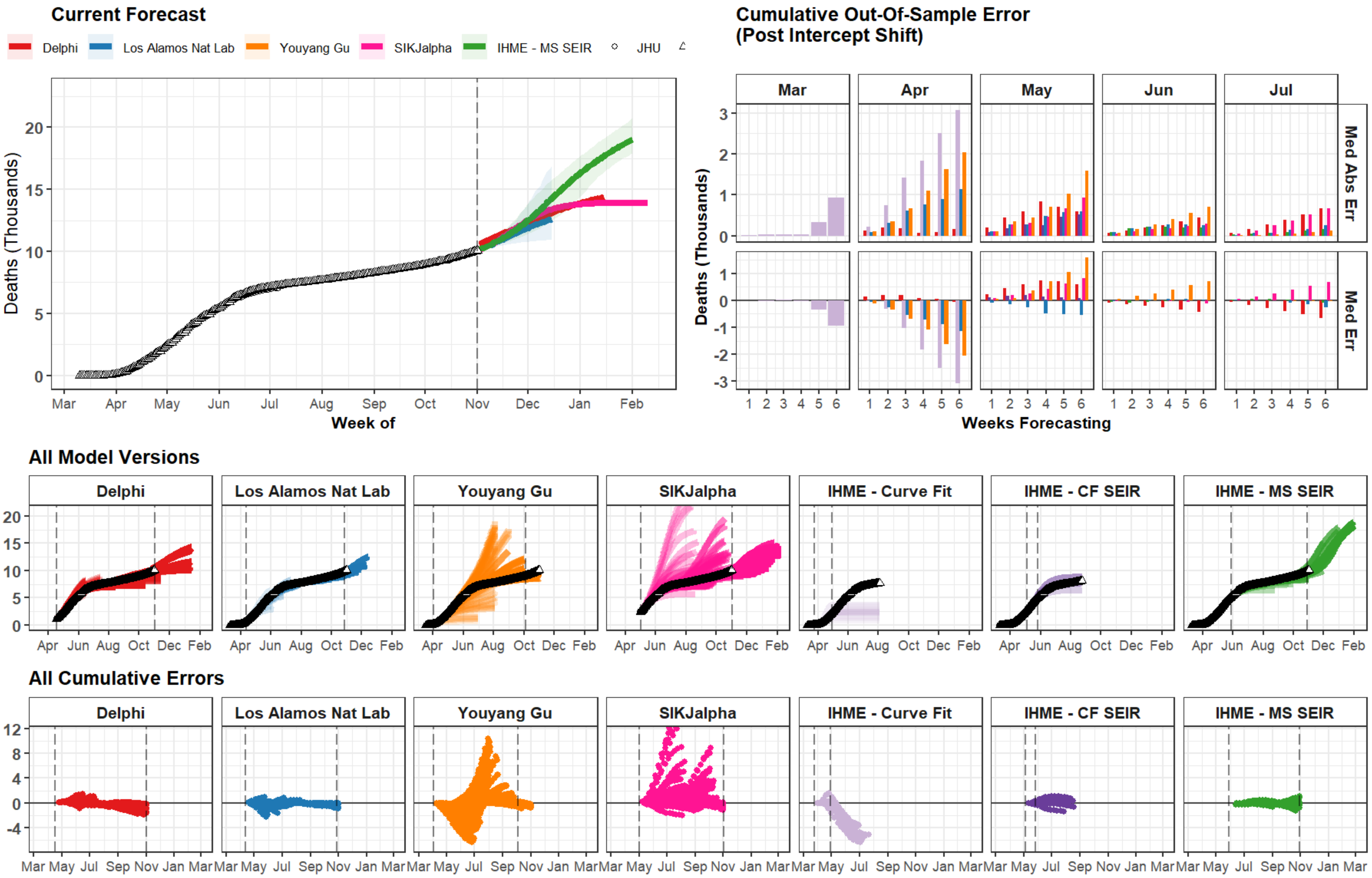
Current Forecast
Delphi
Los Alamos Nat Lab
Youyang Gu
SIKJalpha
IHME - MS SEIR
JHU
Deaths (Thousands)
Week of
Cumulative Out-Of-Sample Error
(Post Intercept Shift)
Mar
Apr
May
Jun
Jul
Med Abs Err
Med Err
Weeks Forecasting
All Model Versions
Delphi
Los Alamos Nat Lab
Youyang Gu
SIKJalpha
IHME - Curve Fit
IHME - CF SEIR
IHME - MS SEIR
All Cumulative Errors

### Massachusetts

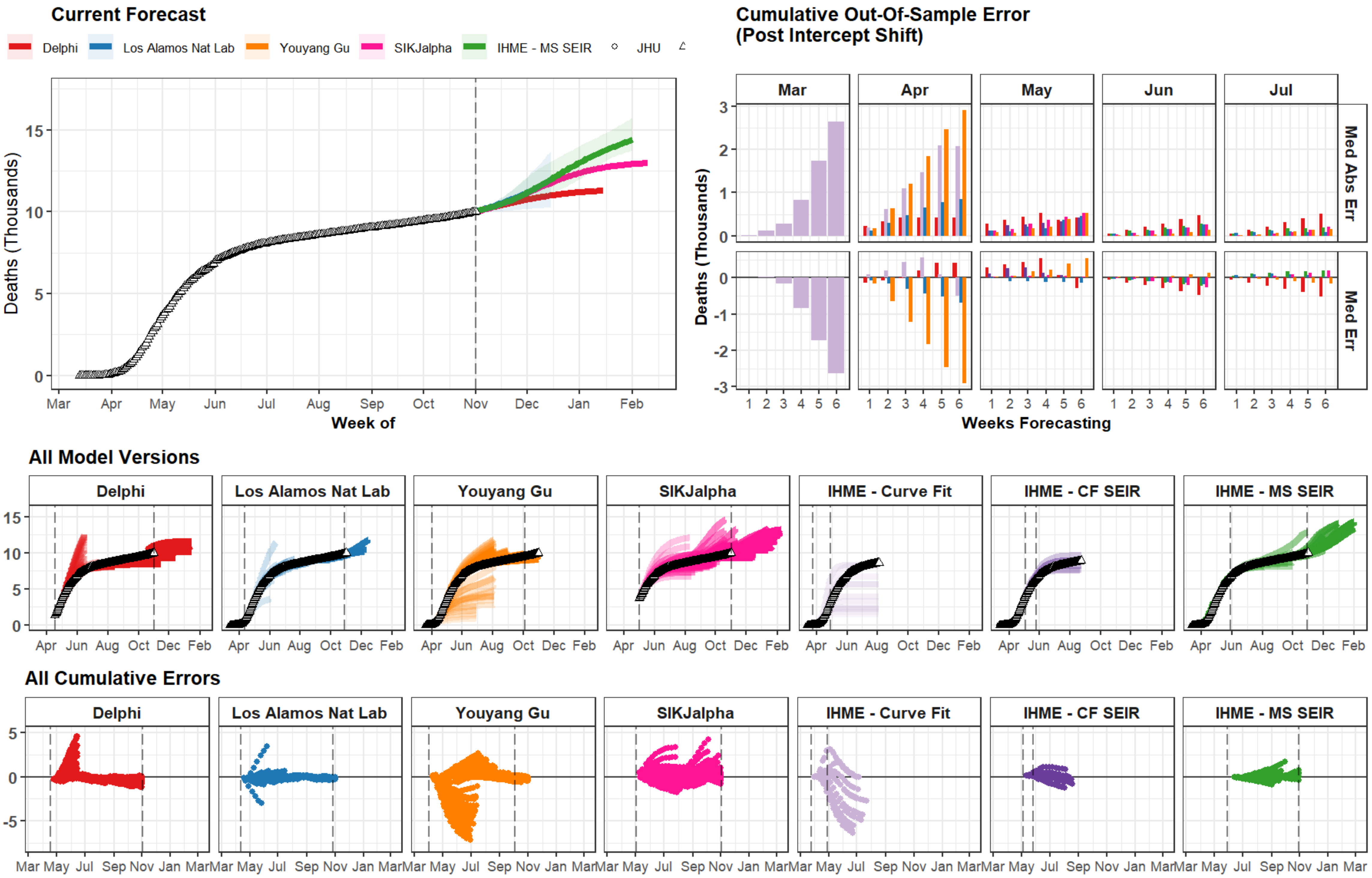

### Pennsylvania

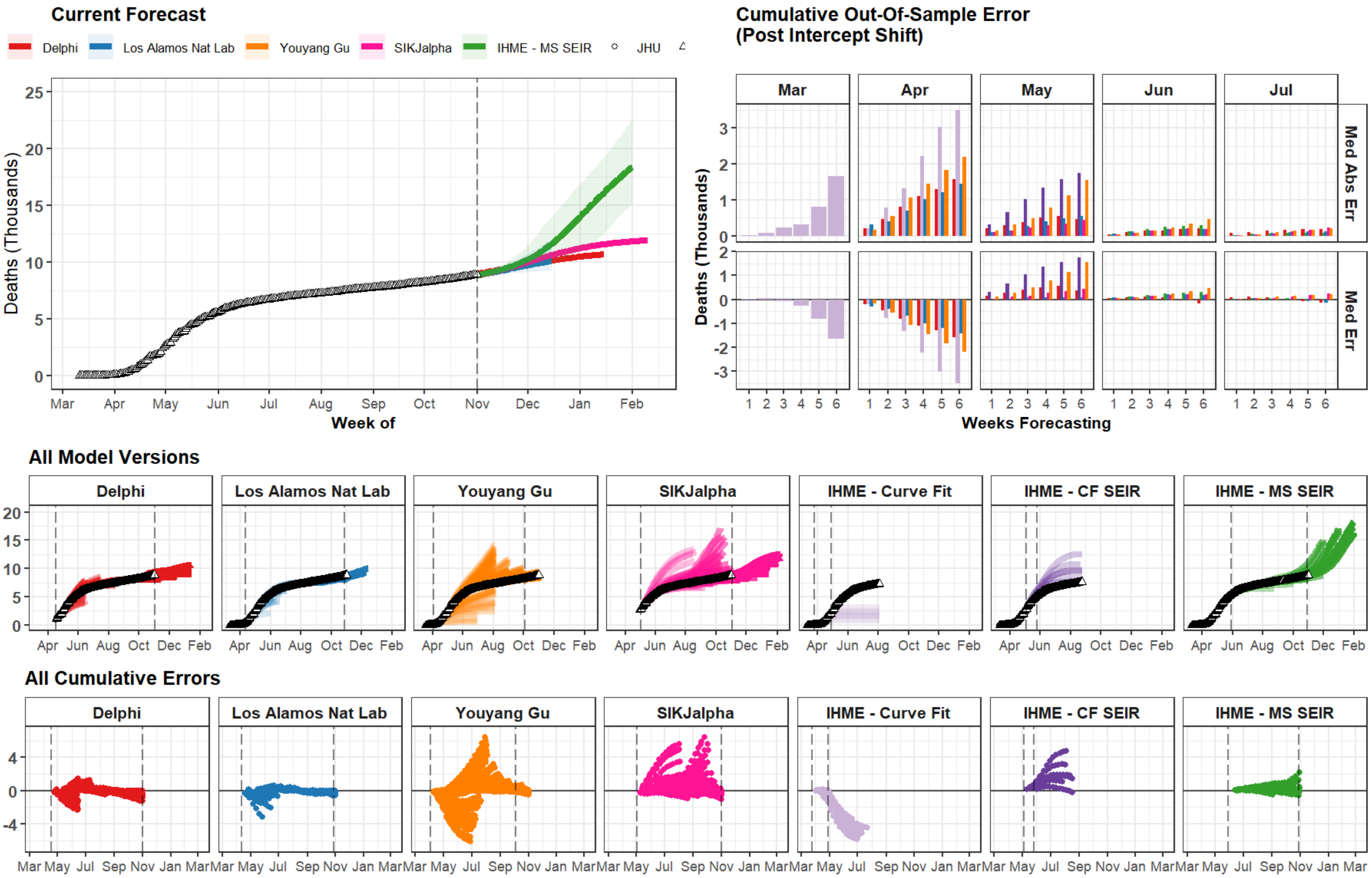

### Bolivia

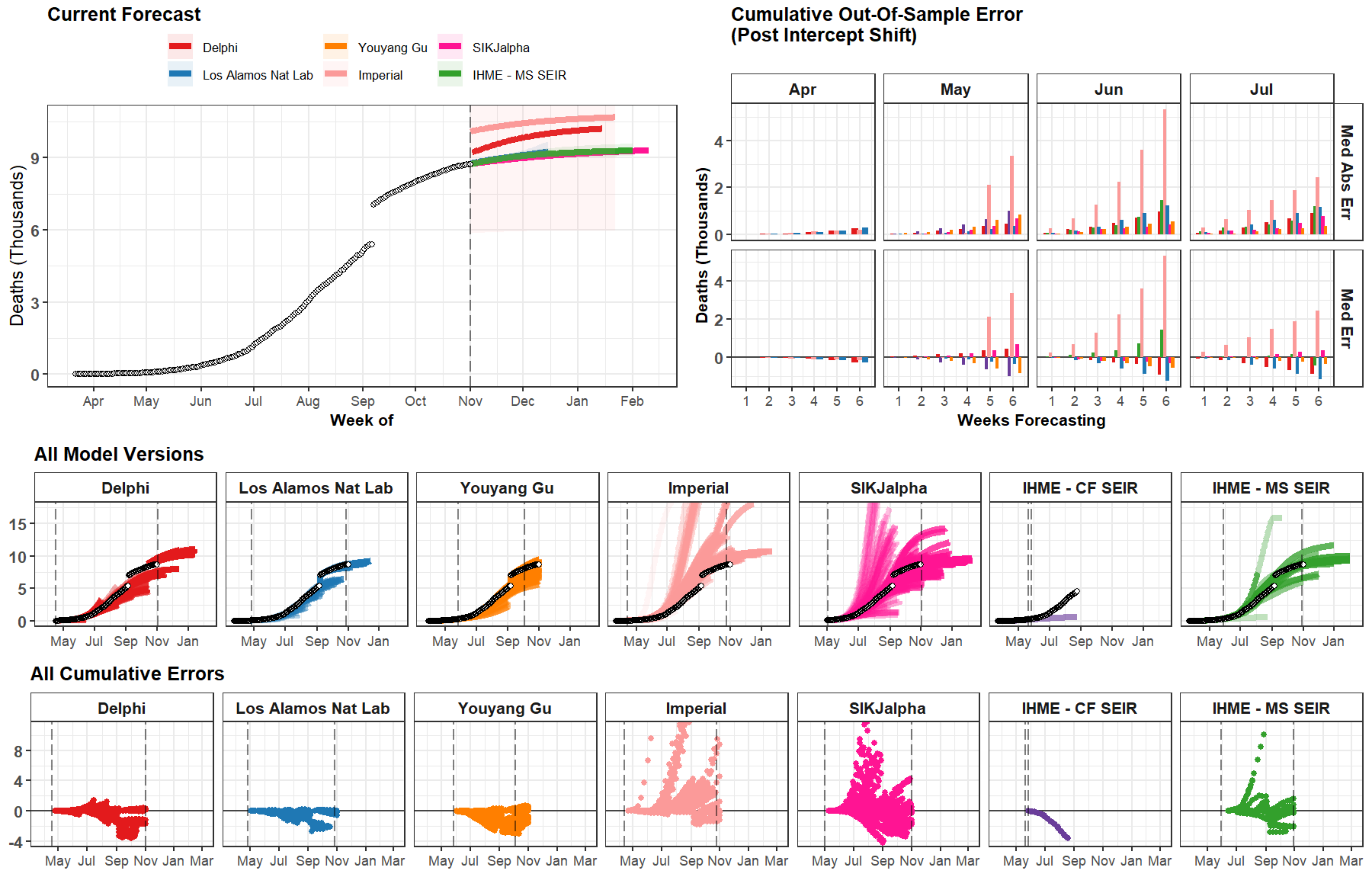
Current Forecast
Delphi
Los Alamos Nat Lab
Youyang Gu
Imperial
SIKJalpha
IHME - MS SEIR
Deaths (Thousands)
Week of
Apr
May
Jun
Jul
Aug
Sep
Oct
Nov
Dec
Jan
Feb
Cumulative Out-Of-Sample Error
(Post Intercept Shift)
Med Abs Err
Med Err
Weeks Forecasting
All Model Versions
IHME - CF SEIR
All Cumulative Errors
Mar

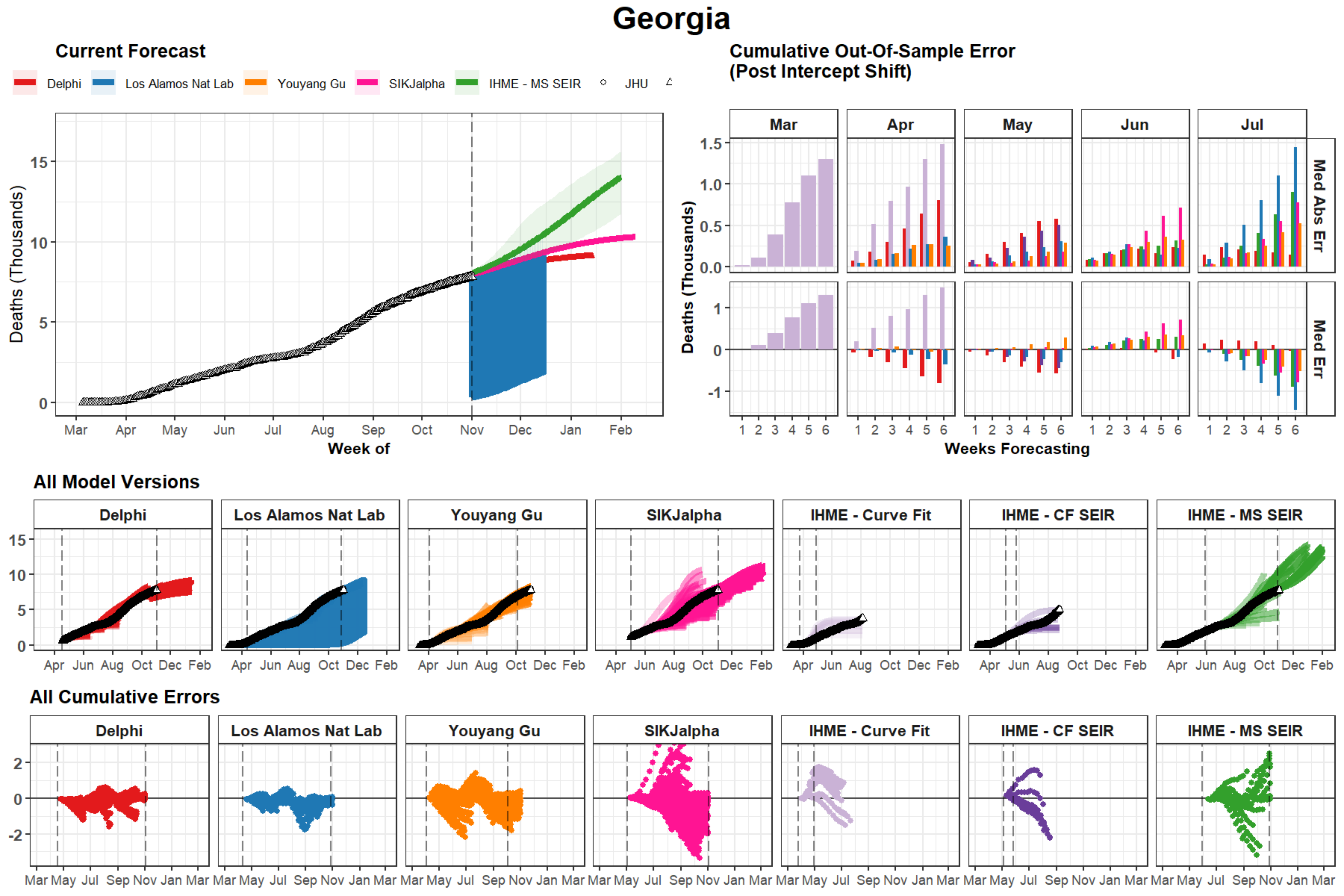
Georgia
Current Forecast
Delphi
Los Alamos Nat Lab
Youyang Gu
SIKJalpha
IHME - MS SEIR
JHU
Deaths (Thousands)
Week of
Mar Apr May Jun Jul Aug Sep Oct Nov Dec Jan Feb
Cumulative Out-Of-Sample Error
(Post Intercept Shift)
Mar Apr May Jun Jul
Med Abs Err
Med Err
Weeks Forecasting
All Model Versions
Delphi
Los Alamos Nat Lab
Youyang Gu
SIKJalpha
IHME - Curve Fit
IHME - CF SEIR
IHME - MS SEIR
Apr Jun Aug Oct Dec Feb
All Cumulative Errors
Mar May Jul Sep Nov Jan Mar

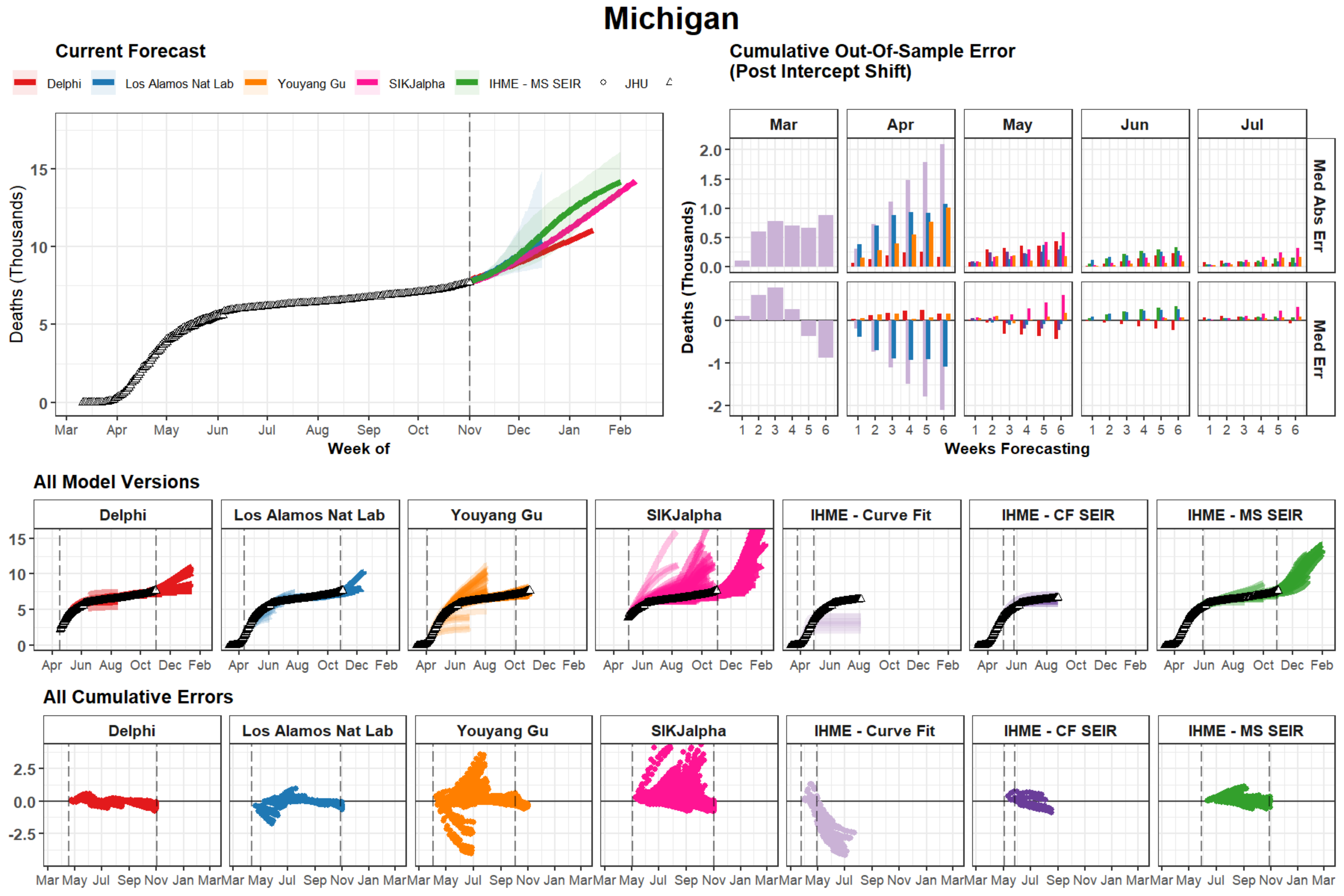

Michigan
Current Forecast
Delphi
Los Alamos Nat Lab
Youyang Gu
SIKJalpha
IHME - MS SEIR
JHU
Deaths (Thousands)
Week of
Mar
Apr
May
Jun
Jul
Aug
Sep
Oct
Nov
Dec
Jan
Feb
Cumulative Out-Of-Sample Error
(Post Intercept Shift)
Med Abs Err
Med Err
Weeks Forecasting
All Model Versions
Delphi
Los Alamos Nat Lab
Youyang Gu
SIKJalpha
IHME - Curve Fit
IHME - CF SEIR
IHME - MS SEIR
All Cumulative Errors

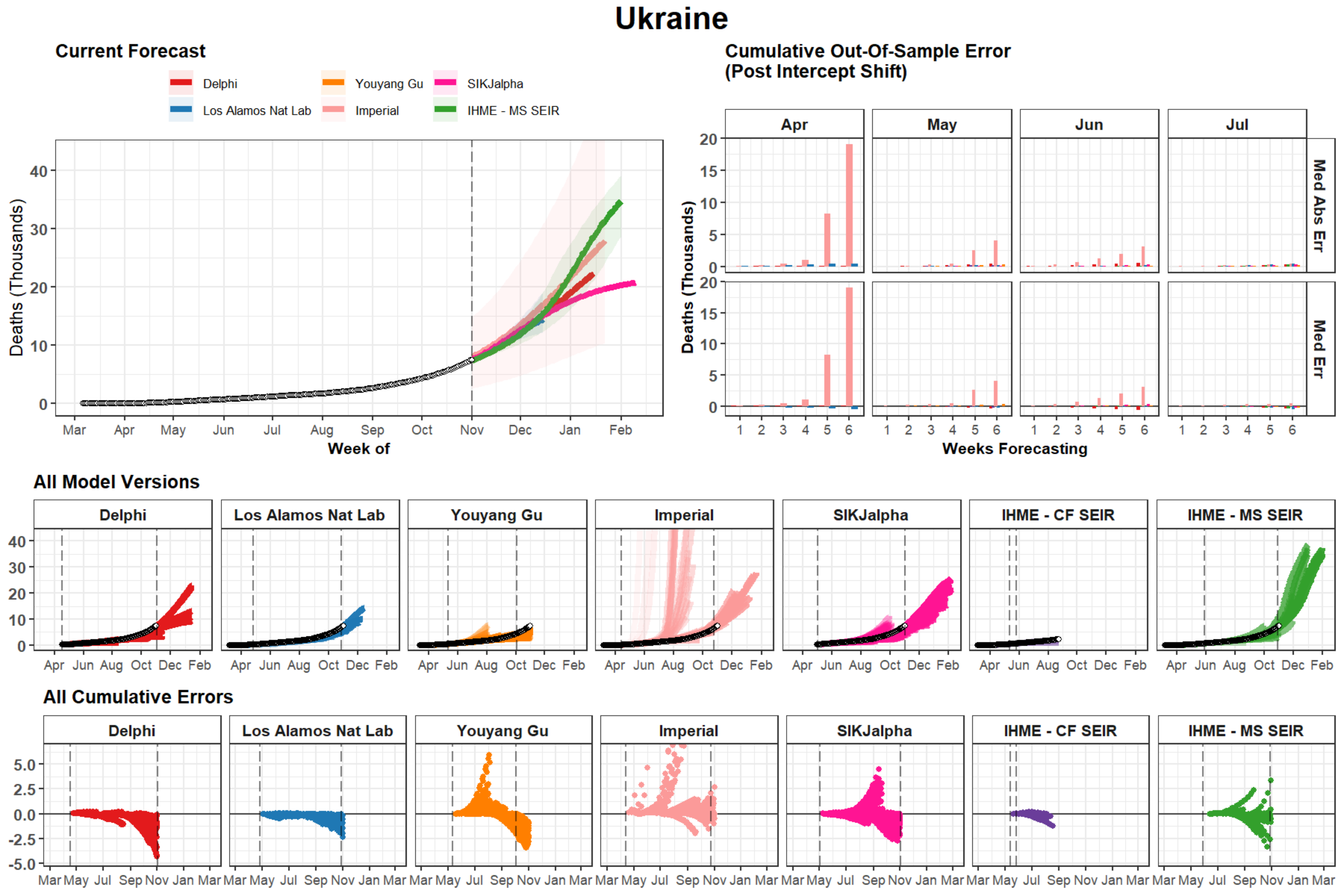

Ukraine
Current Forecast
Delphi
Los Alamos Nat Lab
Youyang Gu
Imperial
SIKJalpha
IHME - MS SEIR
Deaths (Thousands)
Week of
Mar Apr May Jun Jul Aug Sep Oct Nov Dec Jan Feb
Cumulative Out-Of-Sample Error
(Post Intercept Shift)
Apr
May
Jun
Jul
Med Abs Err
Med Err
Weeks Forecasting
All Model Versions
Delphi
Los Alamos Nat Lab
Youyang Gu
Imperial
SIKJalpha
IHME - CF SEIR
IHME - MS SEIR
Apr Jun Aug Oct Dec Feb
All Cumulative Errors
Mar May Jul Sep Nov Jan Mar

### Netherlands

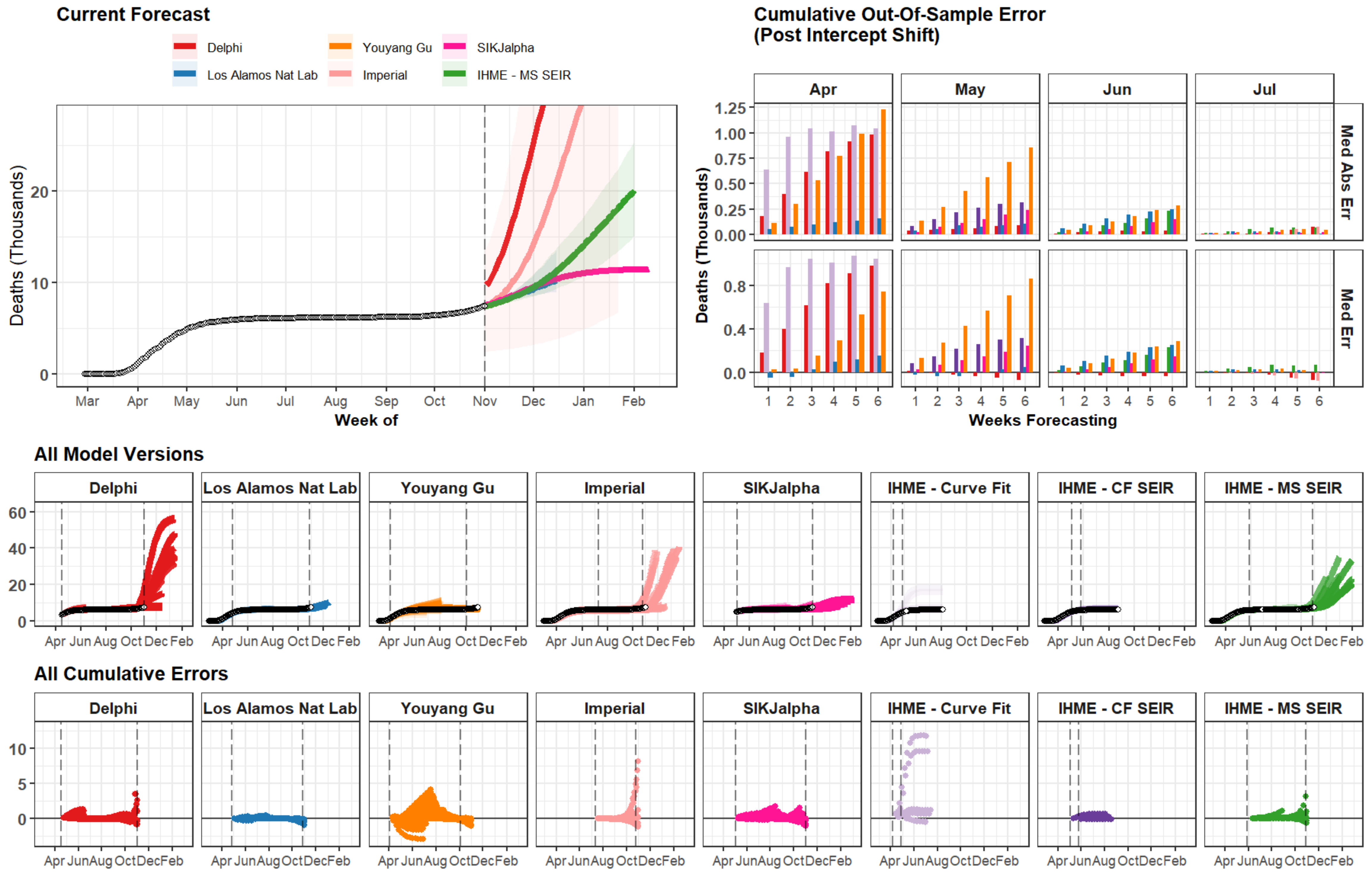

### Philippines

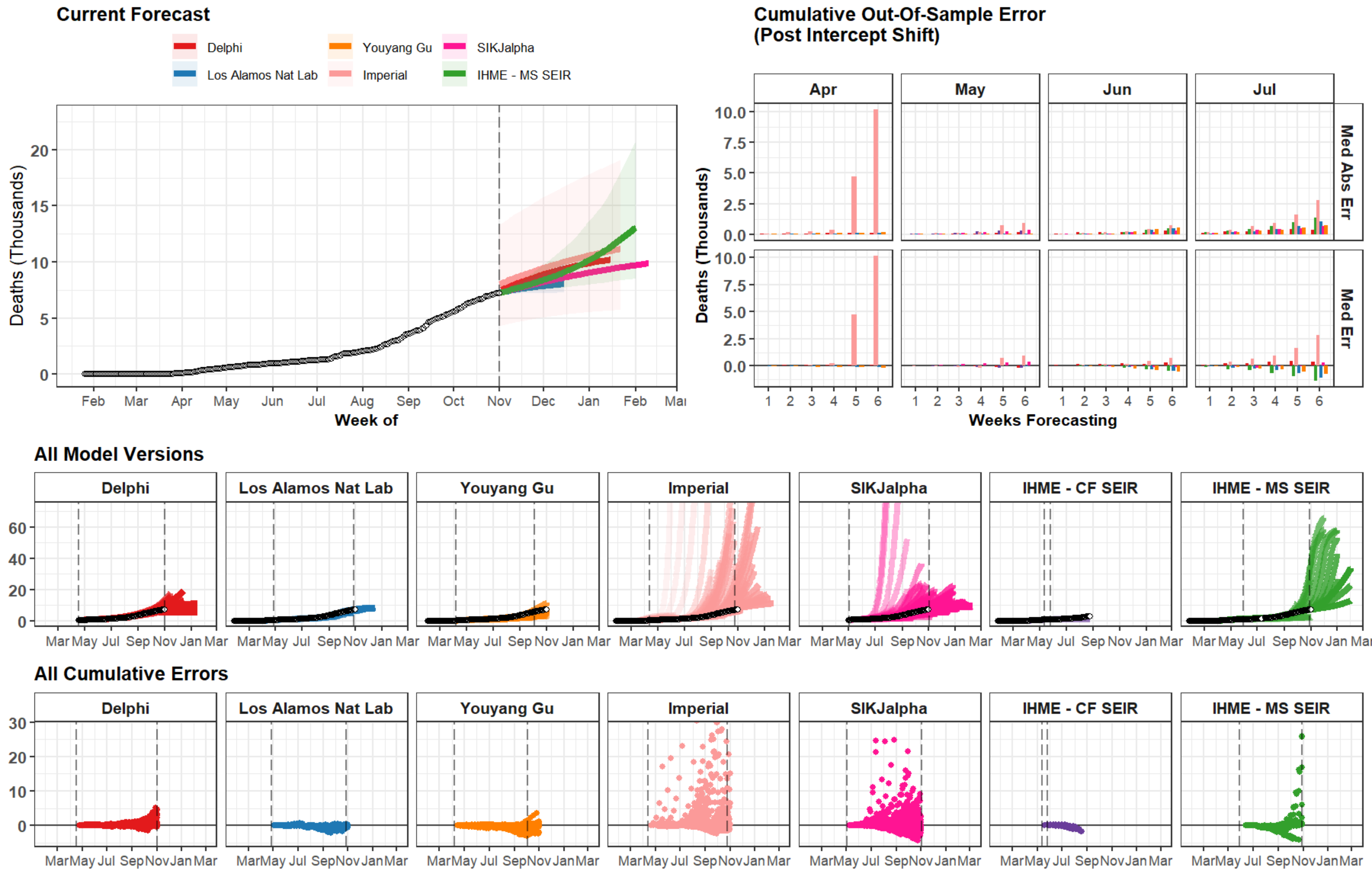

### Romania

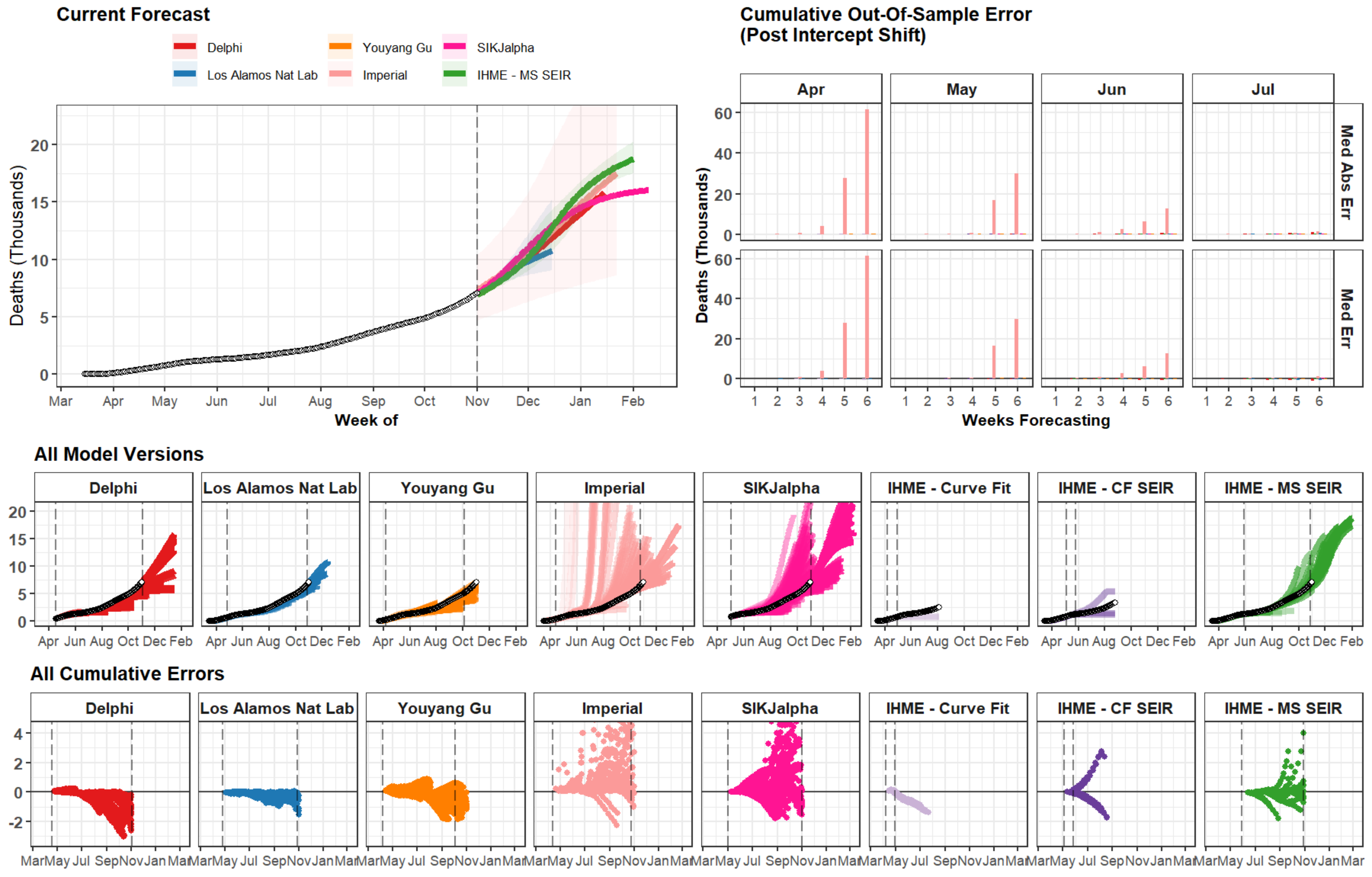

Current Forecast
Delphi
Youyang Gu
SIKJalpha
Los Alamos Nat Lab
Imperial
IHME - MS SEIR
Deaths (Thousands)
Week of
Mar
Apr
May
Jun
Jul
Aug
Sep
Oct
Nov
Dec
Jan
Feb
Cumulative Out-Of-Sample Error
(Post Intercept Shift)
Med Abs Err
Med Err
Weeks Forecasting
All Model Versions
Delphi
Los Alamos Nat Lab
Youyang Gu
Imperial
SIKJalpha
IHME - Curve Fit
IHME - CF SEIR
IHME - MS SEIR
All Cumulative Errors

### Pakistan

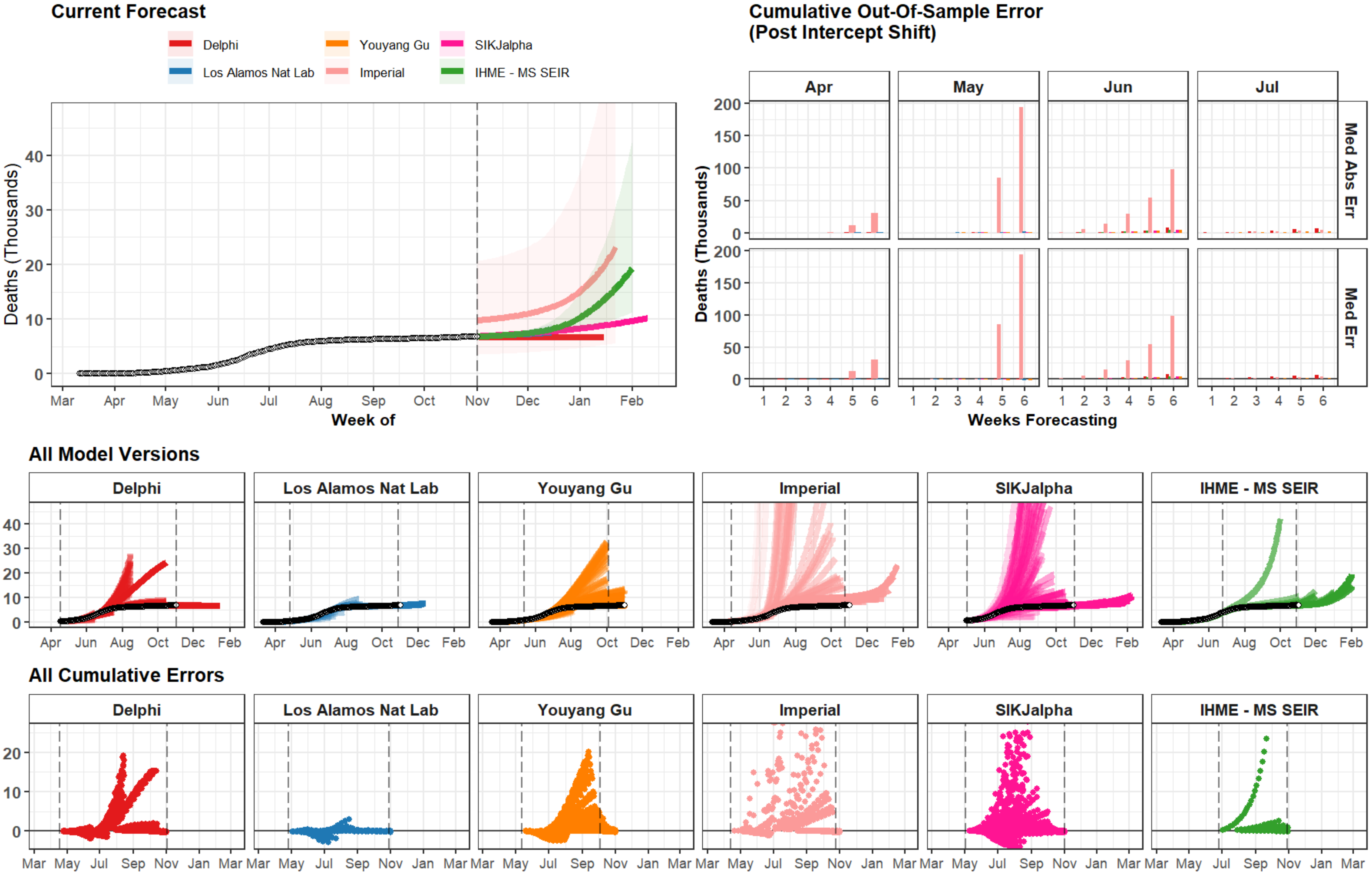

Current Forecast
Delphi
Los Alamos Nat Lab
Youyang Gu
Imperial
SIKJalpha
IHME - MS SEIR
Deaths (Thousands)
Week of
Mar Apr May Jun Jul Aug Sep Oct Nov Dec Jan Feb
Cumulative Out-Of-Sample Error
(Post Intercept Shift)
Apr May Jun Jul
Med Abs Err
Med Err
Weeks Forecasting
All Model Versions
All Cumulative Errors

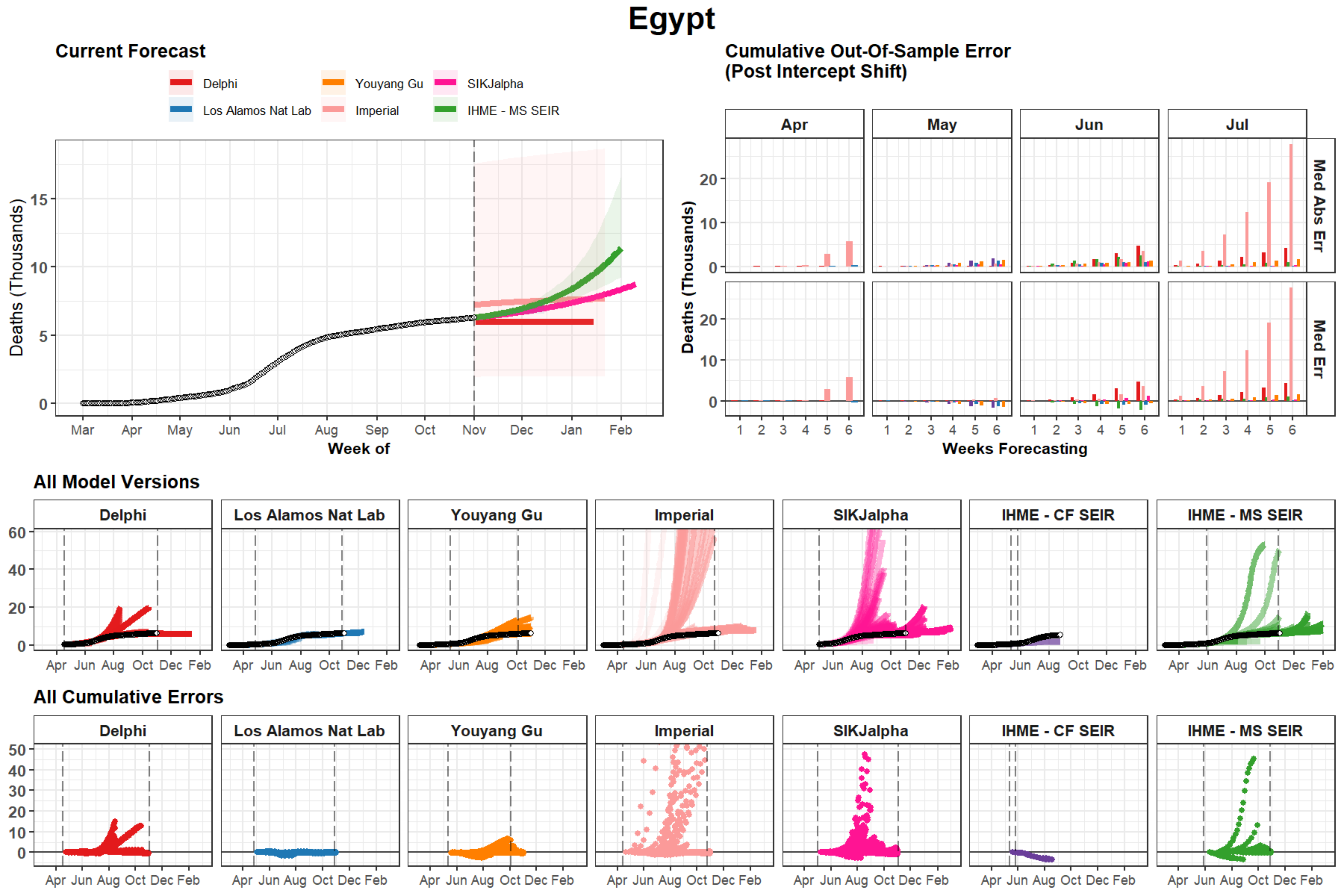

Egypt
Current Forecast
Delphi
Los Alamos Nat Lab
Youyang Gu
Imperial
SIKJalpha
IHME - MS SEIR
Deaths (Thousands)
Week of
Mar Apr May Jun Jul Aug Sep Oct Nov Dec Jan Feb
Cumulative Out-Of-Sample Error
(Post Intercept Shift)
Apr May Jun Jul
Med Abs Err
Med Err
Weeks Forecasting
All Model Versions
Delphi
Los Alamos Nat Lab
Youyang Gu
Imperial
SIKJalpha
IHME - CF SEIR
IHME - MS SEIR
Apr Jun Aug Oct Dec Feb
All Cumulative Errors

### Arizona

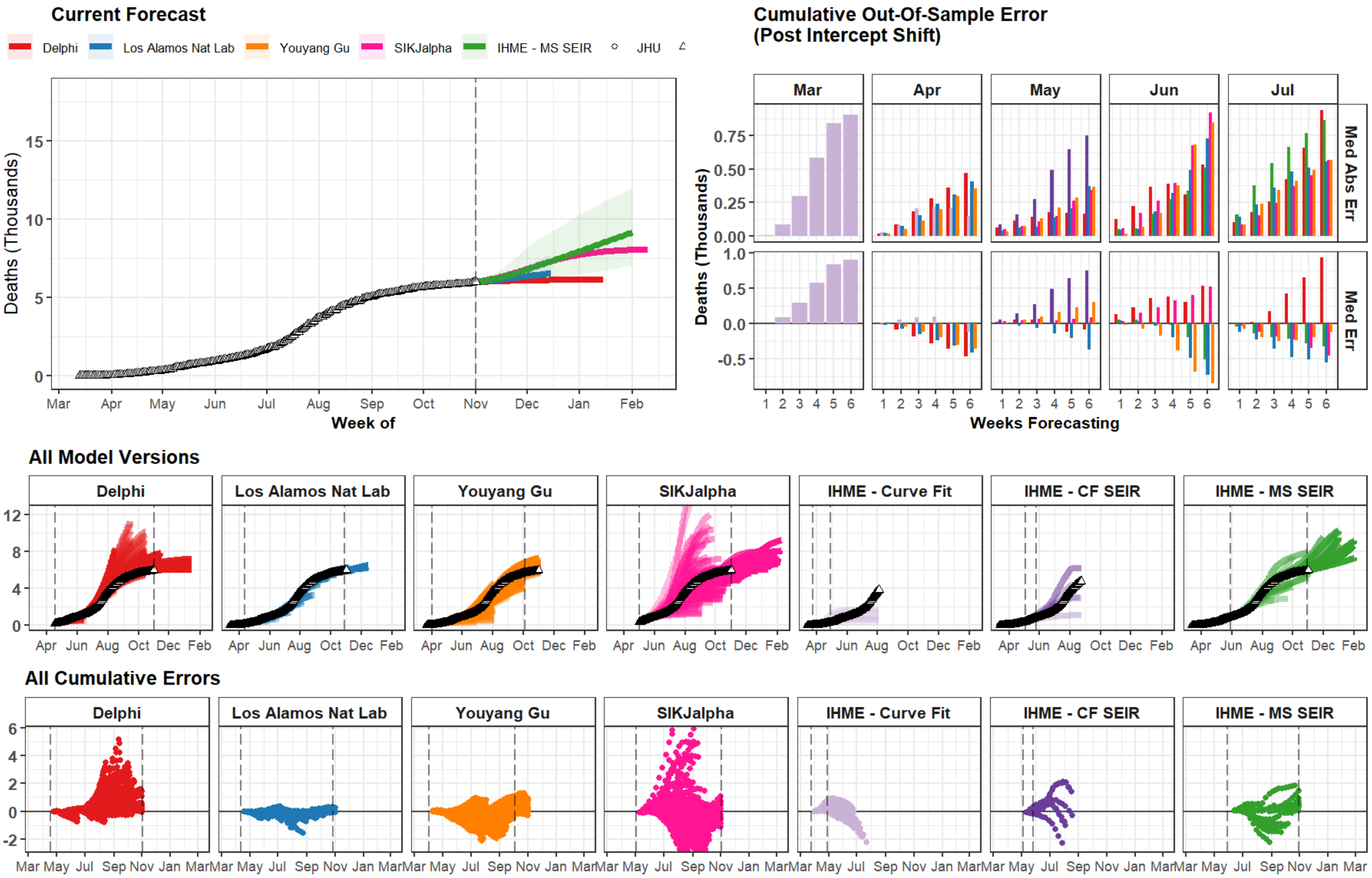
Current Forecast
Delphi
Los Alamos Nat Lab
Youyang Gu
SIKJalpha
IHME - MS SEIR
JHU
Deaths (Thousands)
Week of
Cumulative Out-Of-Sample Error
(Post Intercept Shift)
Mar
Apr
May
Jun
Jul
Med Abs Err
Med Err
Weeks Forecasting
All Model Versions
Delphi
Los Alamos Nat Lab
Youyang Gu
SIKJalpha
IHME - Curve Fit
IHME - CF SEIR
IHME - MS SEIR
All Cumulative Errors

### Bangladesh

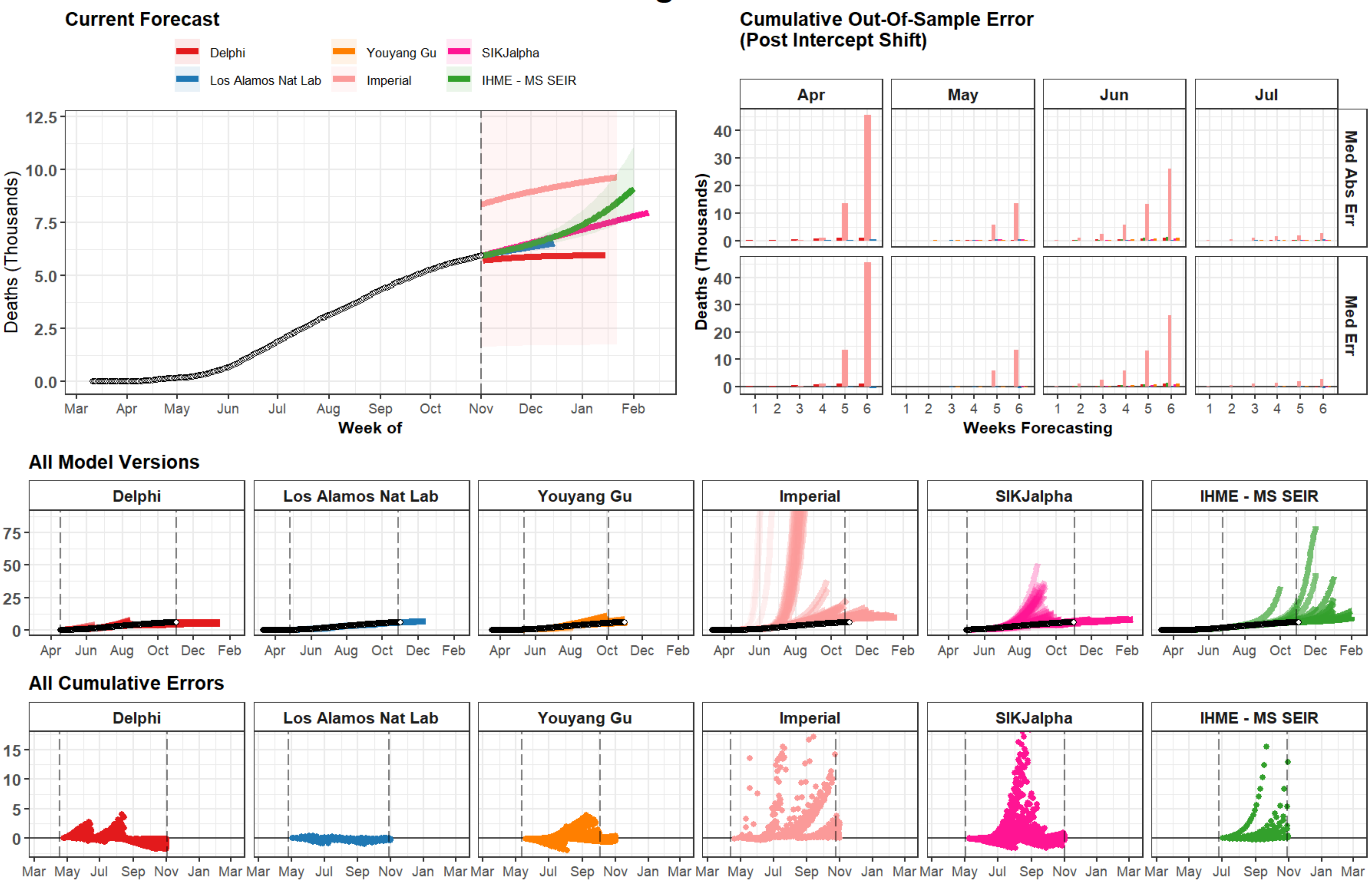

Current Forecast
Delphi
Los Alamos Nat Lab
Youyang Gu
Imperial
SIKJalpha
IHME - MS SEIR
Deaths (Thousands)
Week of
Cumulative Out-Of-Sample Error
(Post Intercept Shift)
Apr
May
Jun
Jul
Med Abs Err
Med Err
Weeks Forecasting
All Model Versions
All Cumulative Errors

### Sweden

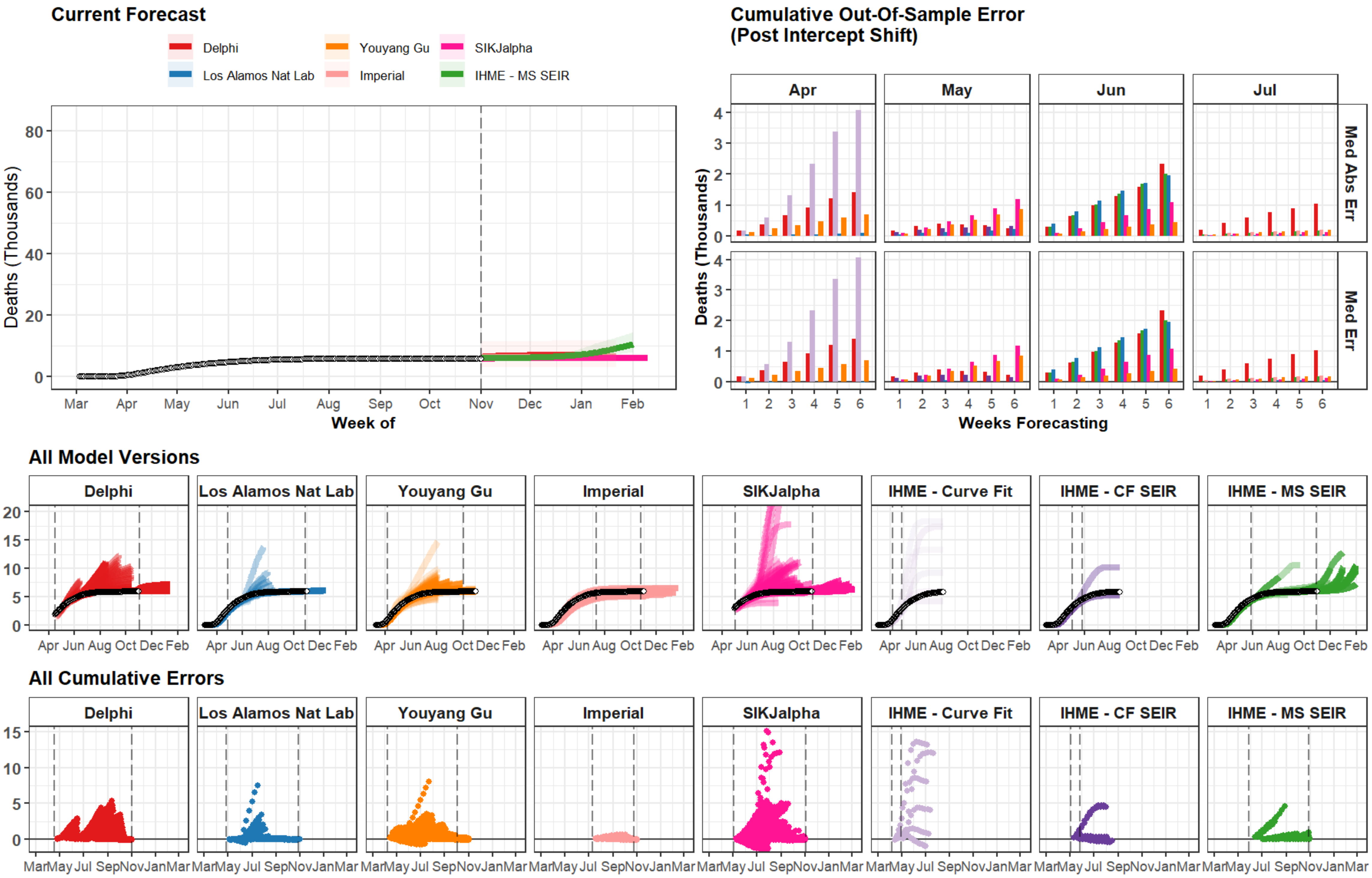
Current Forecast
Delphi
Los Alamos Nat Lab
Youyang Gu
Imperial
SIKJalpha
IHME - MS SEIR
Deaths (Thousands)
0
20
40
60
80
Mar
Apr
May
Jun
Jul
Aug
Sep
Oct
Nov
Dec
Jan
Feb
Week of
Cumulative Out-Of-Sample Error
(Post Intercept Shift)
Apr
May
Jun
Jul
Med Abs Err
Med Err
Deaths (Thousands)
Weeks Forecasting
All Model Versions
Delphi
Los Alamos Nat Lab
Youyang Gu
Imperial
SIKJalpha
IHME - Curve Fit
IHME - CF SEIR
IHME - MS SEIR
Apr Jun Aug Oct Dec Feb
All Cumulative Errors
Mar May Jul Sep Nov Jan Mar

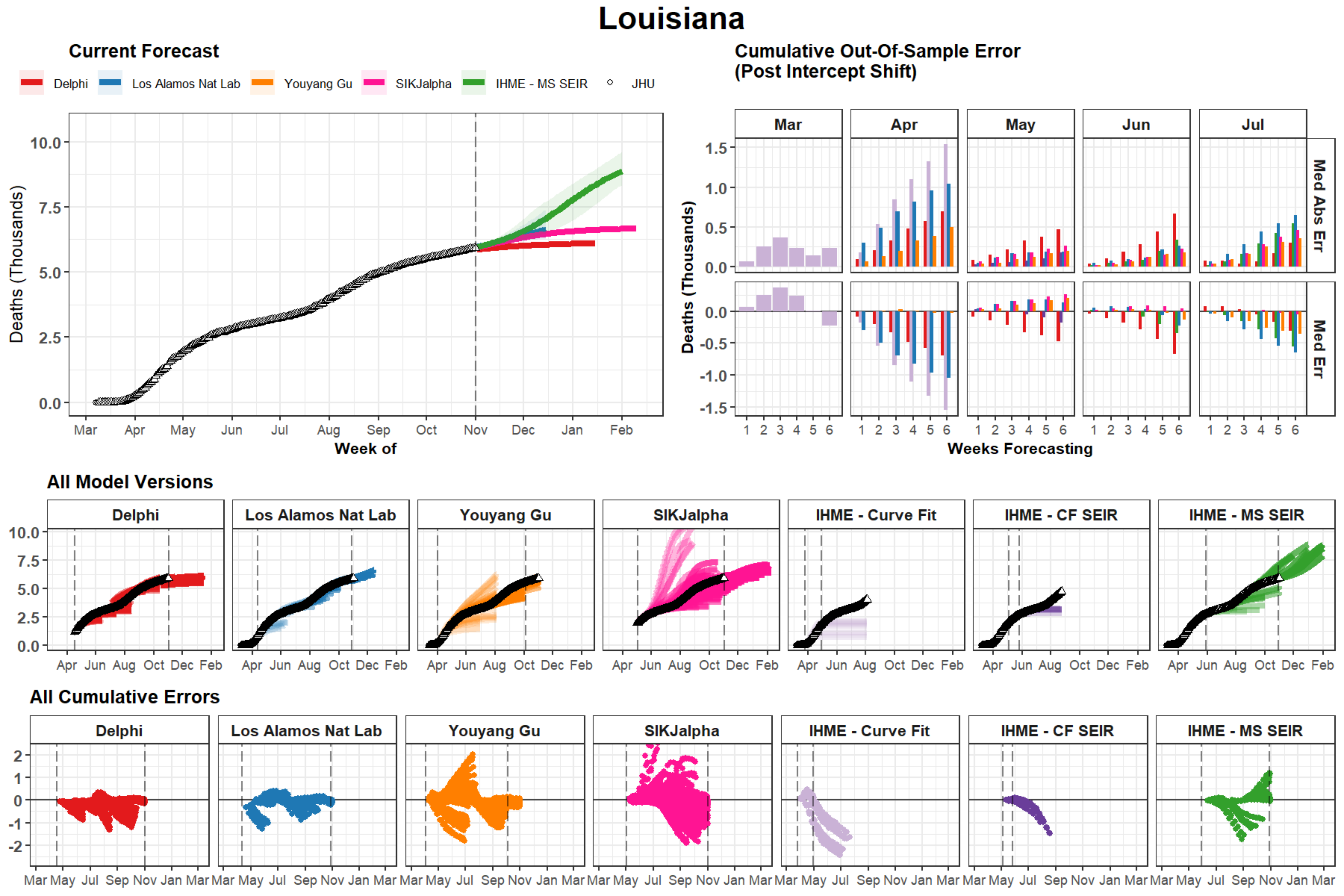

Louisiana
Current Forecast
Delphi
Los Alamos Nat Lab
Youyang Gu
SIKJalpha
IHME - MS SEIR
JHU
Deaths (Thousands)
Week of
Mar Apr May Jun Jul Aug Sep Oct Nov Dec Jan Feb
Cumulative Out-Of-Sample Error
(Post Intercept Shift)
Mar Apr May Jun Jul
Med Abs Err
Med Err
Weeks Forecasting
All Model Versions
Delphi
Los Alamos Nat Lab
Youyang Gu
SIKJalpha
IHME - Curve Fit
IHME - CF SEIR
IHME - MS SEIR
All Cumulative Errors

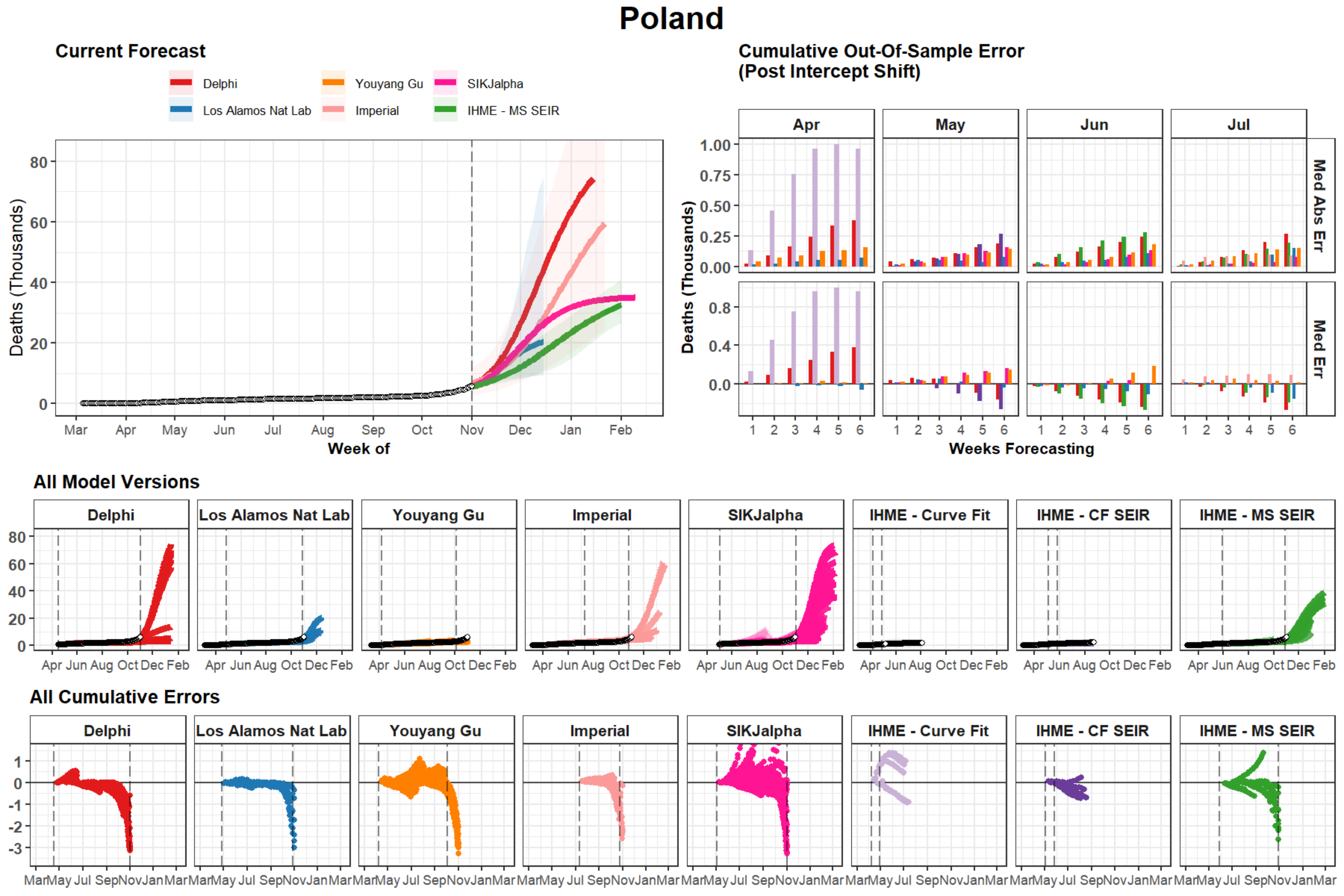
Poland
Current Forecast
Delphi
Los Alamos Nat Lab
Youyang Gu
Imperial
SIKJalpha
IHME - MS SEIR
Deaths (Thousands)
Week of
Mar Apr May Jun Jul Aug Sep Oct Nov Dec Jan Feb
Cumulative Out-Of-Sample Error
(Post Intercept Shift)
Apr
May
Jun
Jul
Med Abs Err
Med Err
Weeks Forecasting
All Model Versions
Delphi
Los Alamos Nat Lab
Youyang Gu
Imperial
SIKJalpha
IHME - Curve Fit
IHME - CF SEIR
IHME - MS SEIR
All Cumulative Errors

### Saudi Arabia

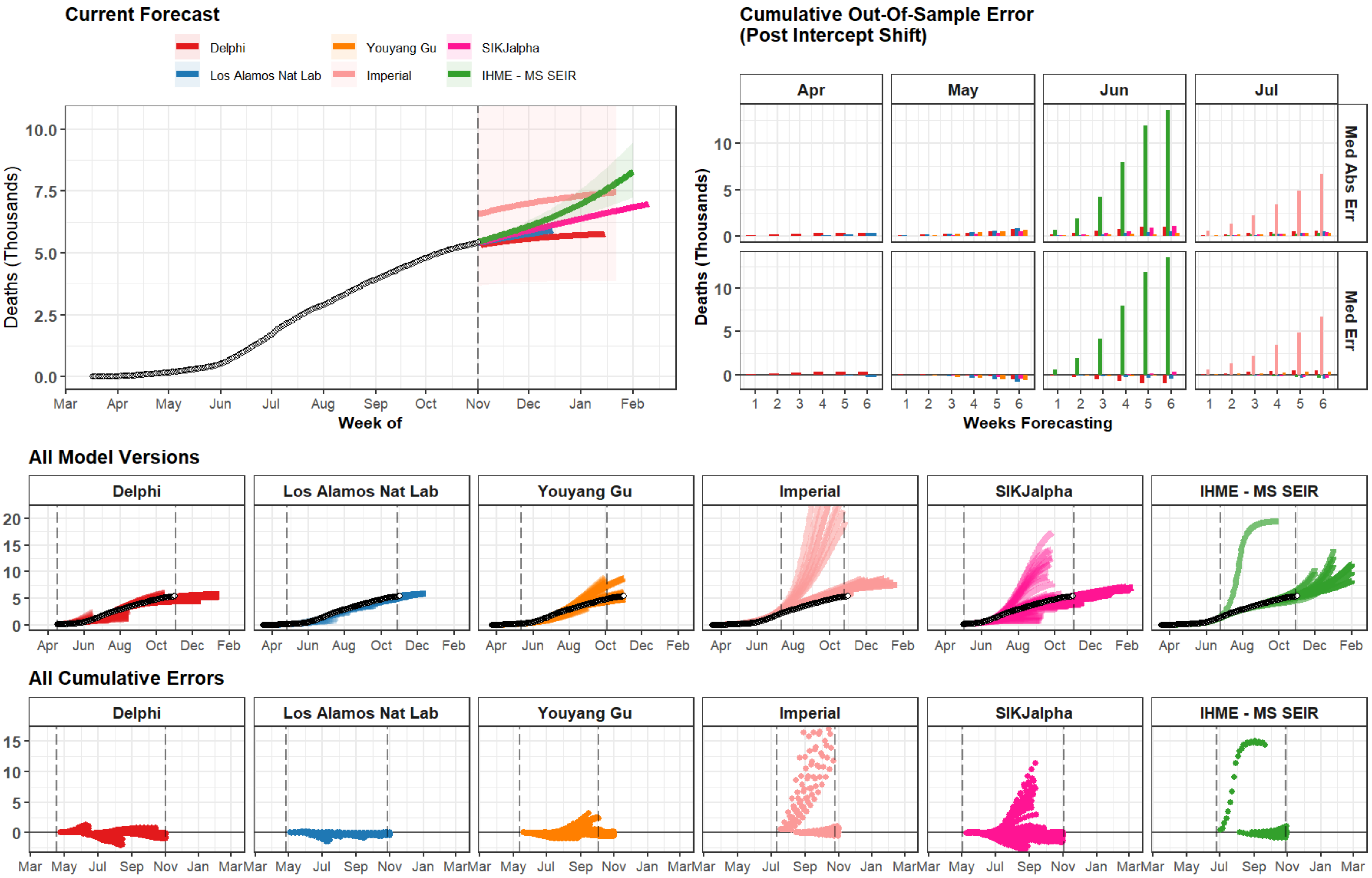

### Ohio

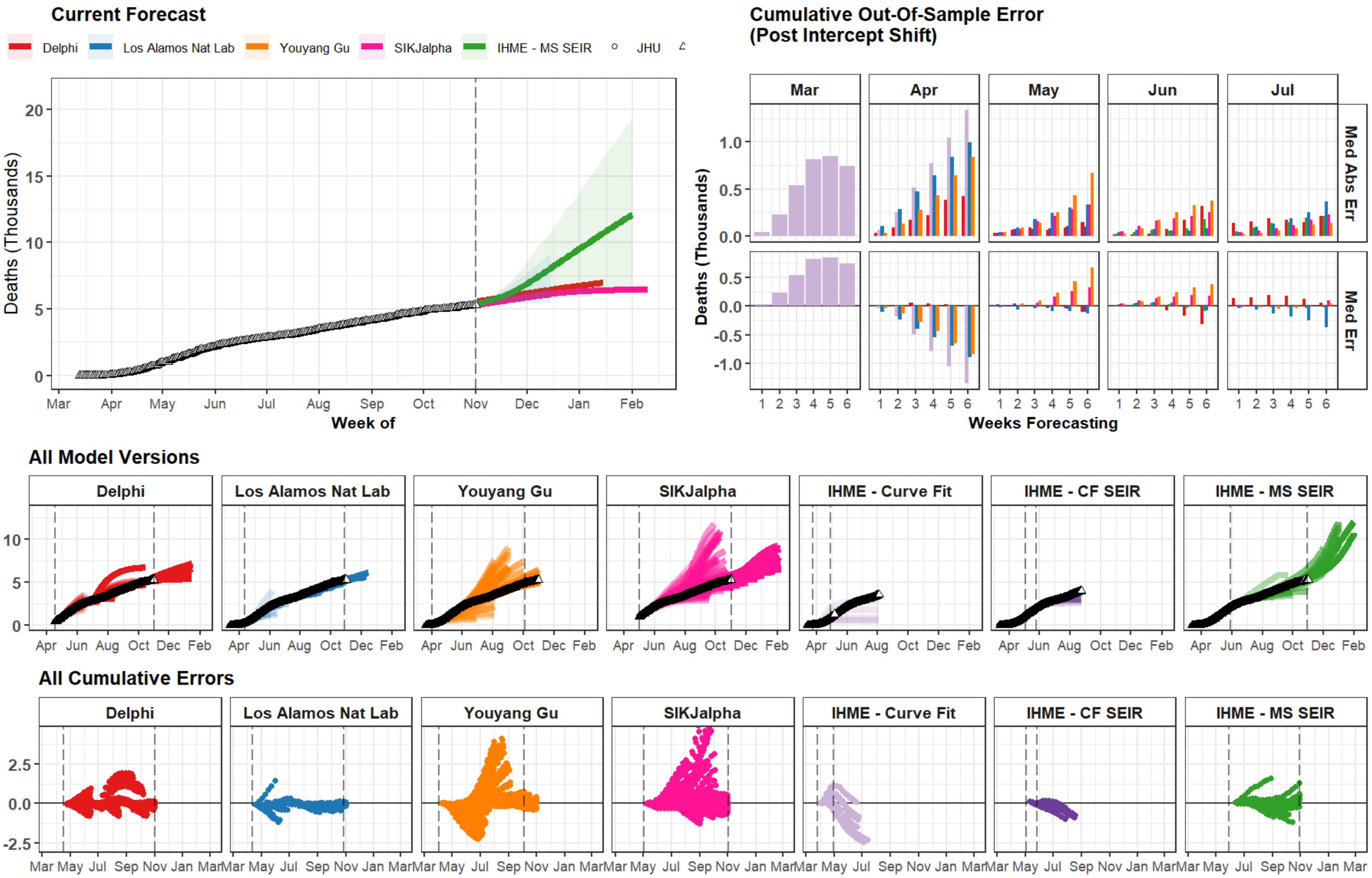

Current Forecast
Delphi
Los Alamos Nat Lab
Youyang Gu
SIKJalpha
IHME - MS SEIR
JHU
Deaths (Thousands)
Week of
Mar Apr May Jun Jul Aug Sep Oct Nov Dec Jan Feb
Cumulative Out-Of-Sample Error
(Post Intercept Shift)
Mar Apr May Jun Jul
Med Abs Err
Med Err
Weeks Forecasting
All Model Versions
Delphi
Los Alamos Nat Lab
Youyang Gu
SIKJalpha
IHME - Curve Fit
IHME - CF SEIR
IHME - MS SEIR
Apr Jun Aug Oct Dec Feb
All Cumulative Errors
Mar May Jul Sep Nov Jan Mar

### China

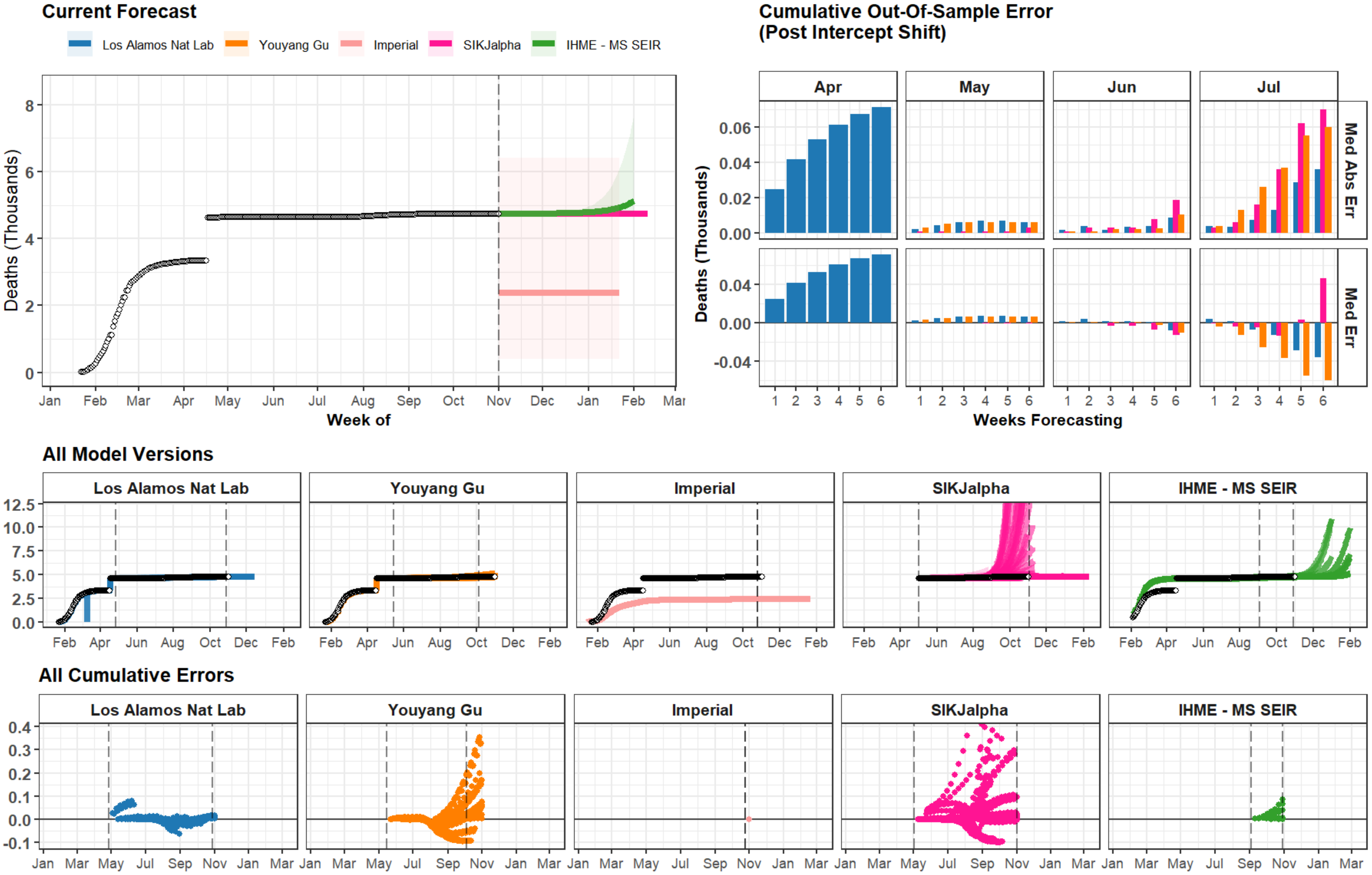
Current Forecast
Los Alamos Nat Lab
Youyang Gu
Imperial
SIKJalpha
IHME - MS SEIR
Deaths (Thousands)
Week of
Cumulative Out-Of-Sample Error
(Post Intercept Shift)
Apr
May
Jun
Jul
Med Abs Err
Med Err
Weeks Forecasting
All Model Versions
All Cumulative Errors

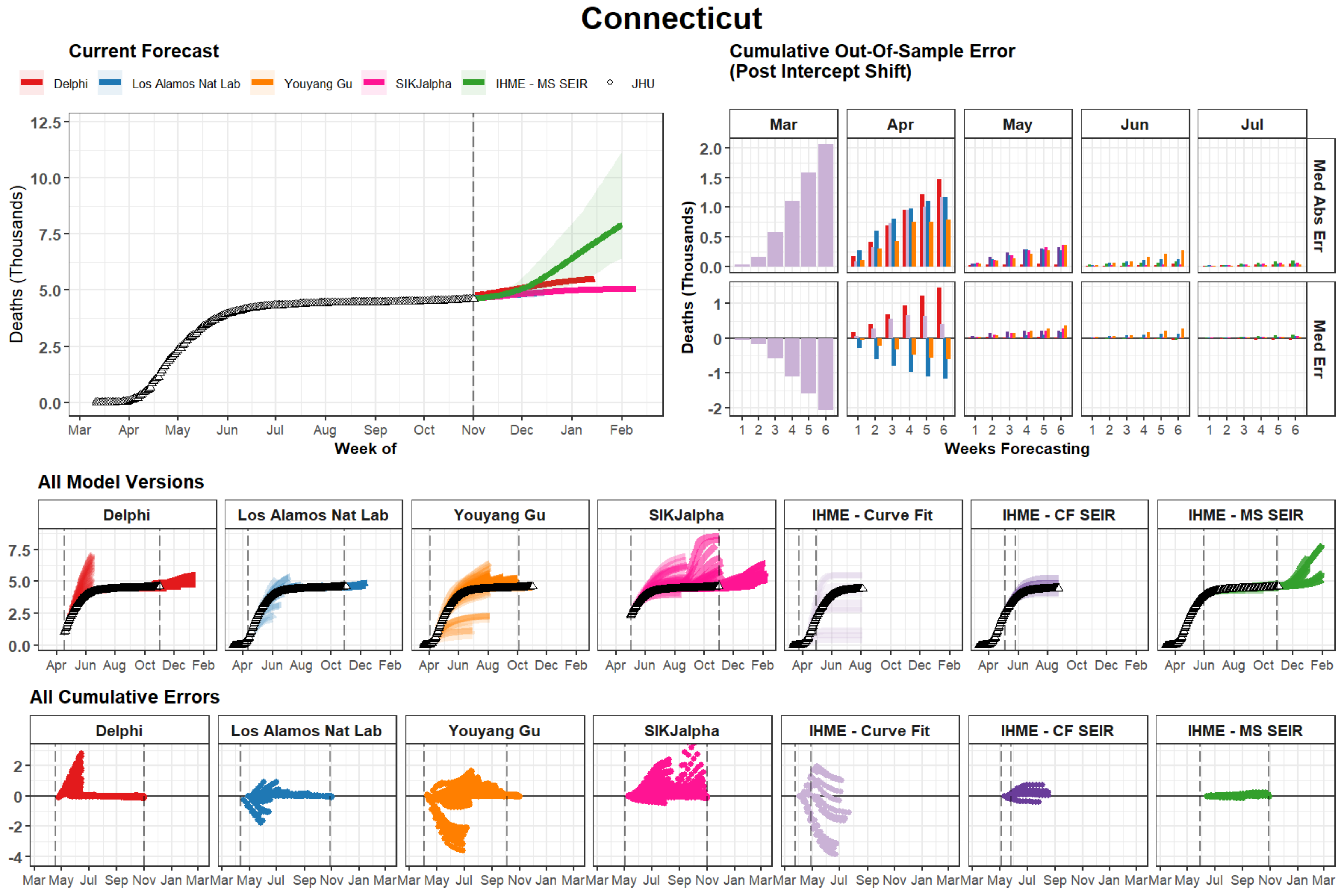
Connecticut
Current Forecast
Delphi
Los Alamos Nat Lab
Youyang Gu
SIKJalpha
IHME - MS SEIR
JHU
Deaths (Thousands)
12.5
10.0
7.5
5.0
2.5
0.0
Mar Apr May Jun Jul Aug Sep Oct Nov Dec Jan Feb
Week of
Cumulative Out-Of-Sample Error
(Post Intercept Shift)
Mar Apr May Jun Jul
Med Abs Err
Med Err
Deaths (Thousands)
Weeks Forecasting
1 2 3 4 5 6
All Model Versions
Delphi
Los Alamos Nat Lab
Youyang Gu
SIKJalpha
IHME - Curve Fit
IHME - CF SEIR
IHME - MS SEIR
Apr Jun Aug Oct Dec Feb
All Cumulative Errors
Delphi
Los Alamos Nat Lab
Youyang Gu
SIKJalpha
IHME - Curve Fit
IHME - CF SEIR
IHME - MS SEIR
Mar May Jul Sep Nov Jan Mar

### North Carolina

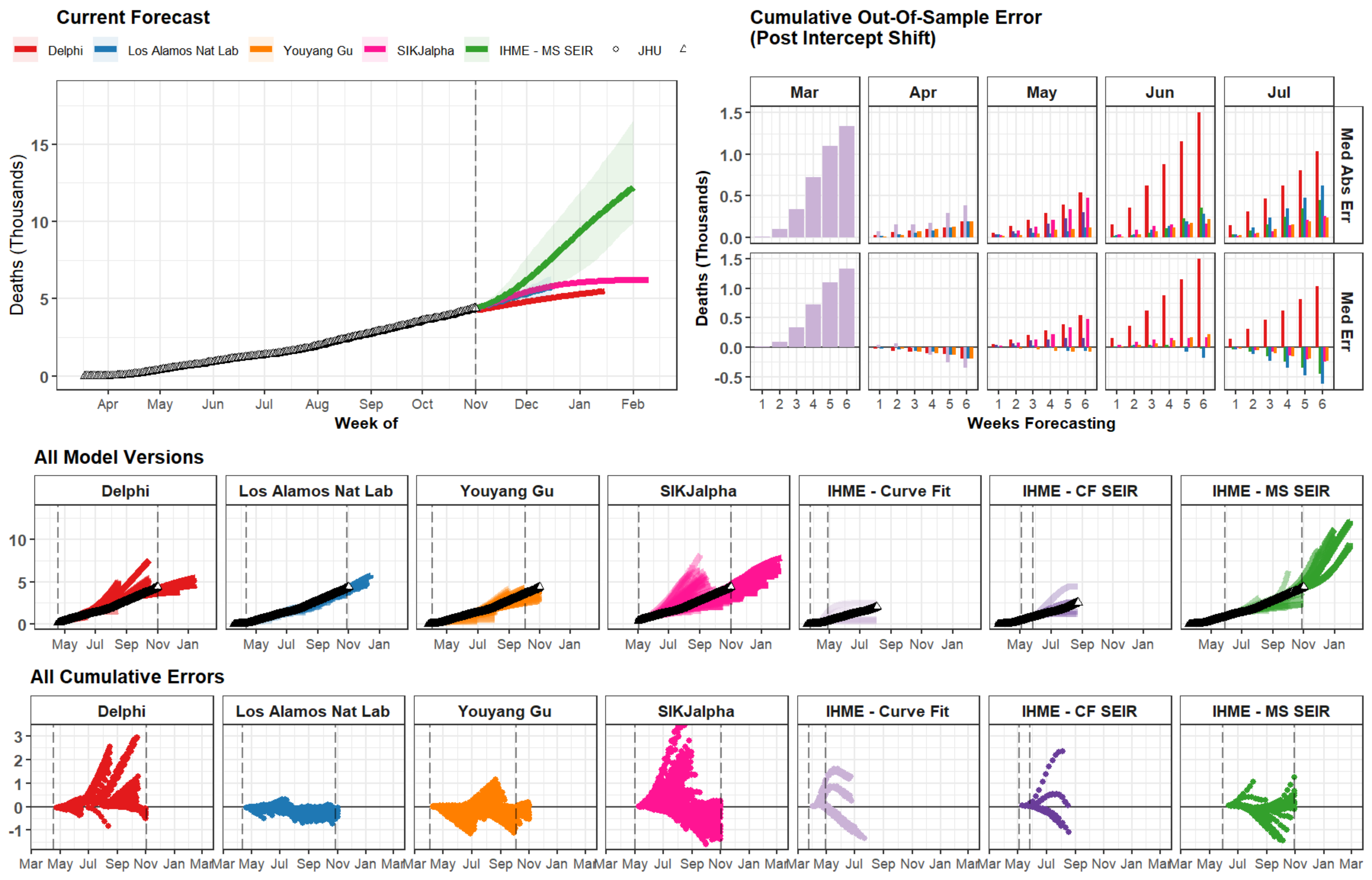

### Indiana

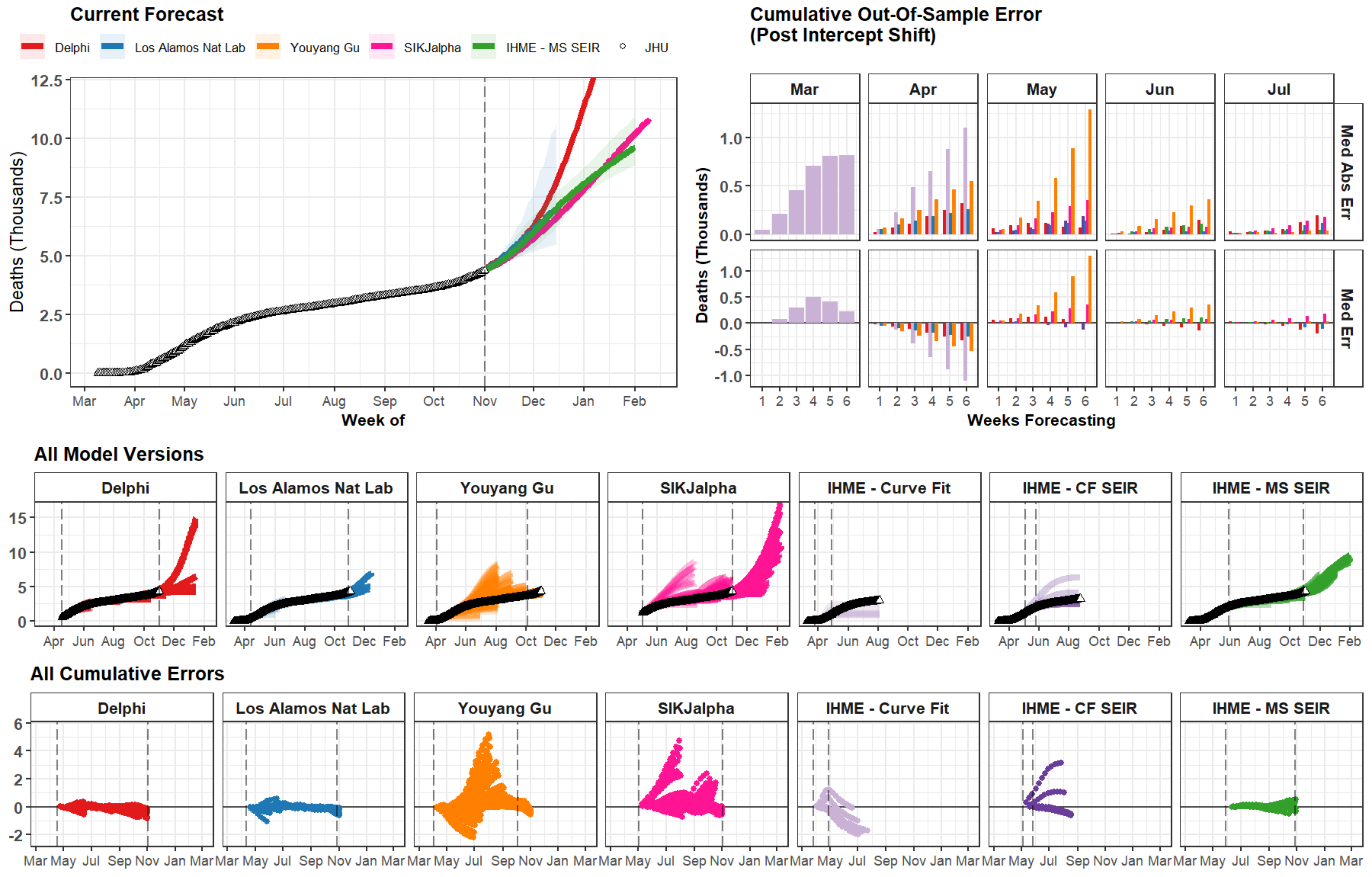

Current Forecast
Delphi
Los Alamos Nat Lab
Youyang Gu
SIKJalpha
IHME - MS SEIR
JHU
Deaths (Thousands)
Week of
Cumulative Out-Of-Sample Error
(Post Intercept Shift)
Mar
Apr
May
Jun
Jul
Med Abs Err
Med Err
Weeks Forecasting
All Model Versions
Delphi
Los Alamos Nat Lab
Youyang Gu
SIKJalpha
IHME - Curve Fit
IHME - CF SEIR
IHME - MS SEIR
All Cumulative Errors

### Maryland

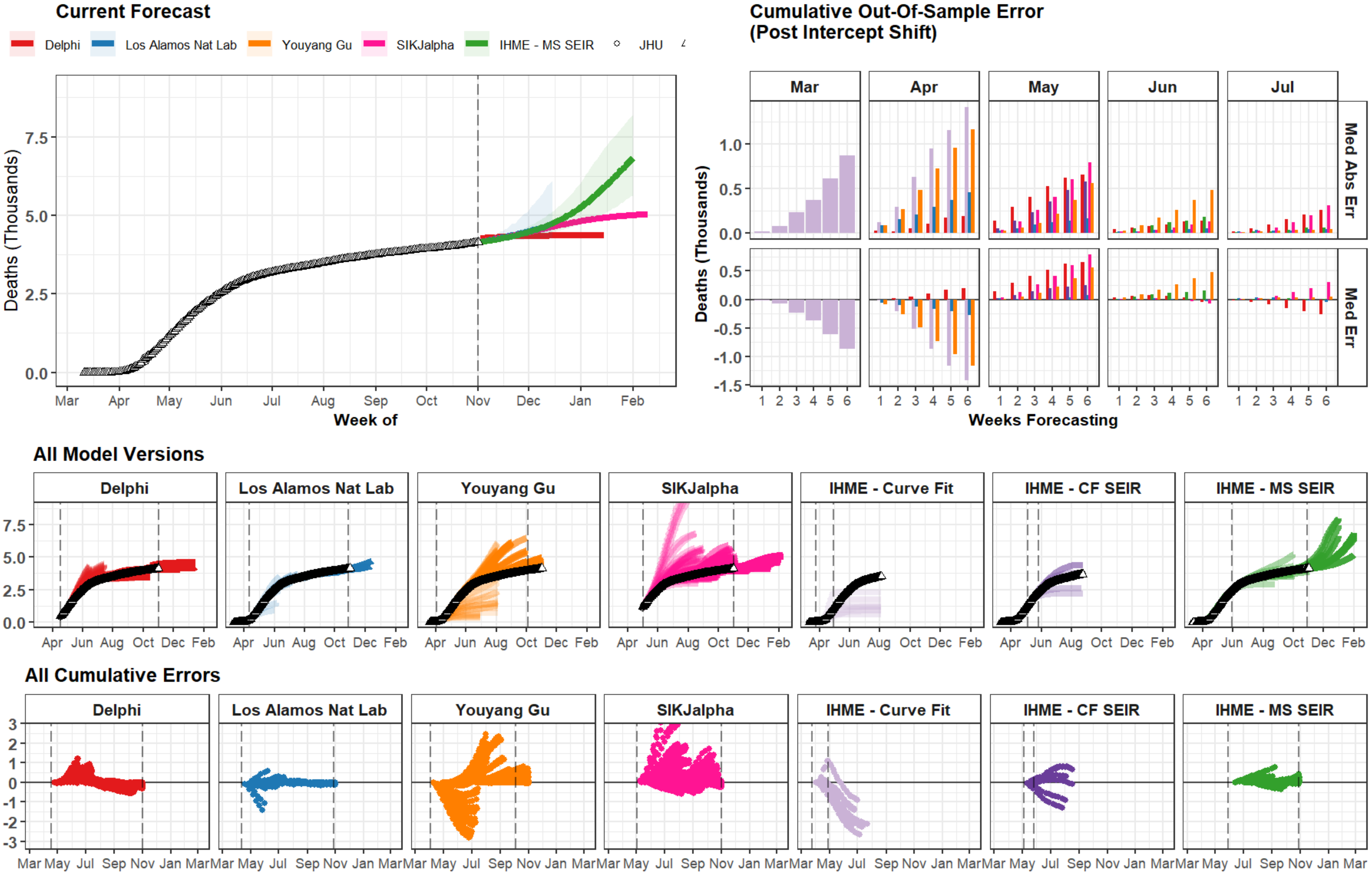

Current Forecast
Delphi
Los Alamos Nat Lab
Youyang Gu
SIKJalpha
IHME - MS SEIR
JHU
Deaths (Thousands)
Week of
Mar Apr May Jun Jul Aug Sep Oct Nov Dec Jan Feb
Cumulative Out-Of-Sample Error
(Post Intercept Shift)
Mar
Apr
May
Jun
Jul
Med Abs Err
Med Err
Weeks Forecasting
All Model Versions
Delphi
Los Alamos Nat Lab
Youyang Gu
SIKJalpha
IHME - Curve Fit
IHME - CF SEIR
IHME - MS SEIR
All Cumulative Errors

### South Carolina

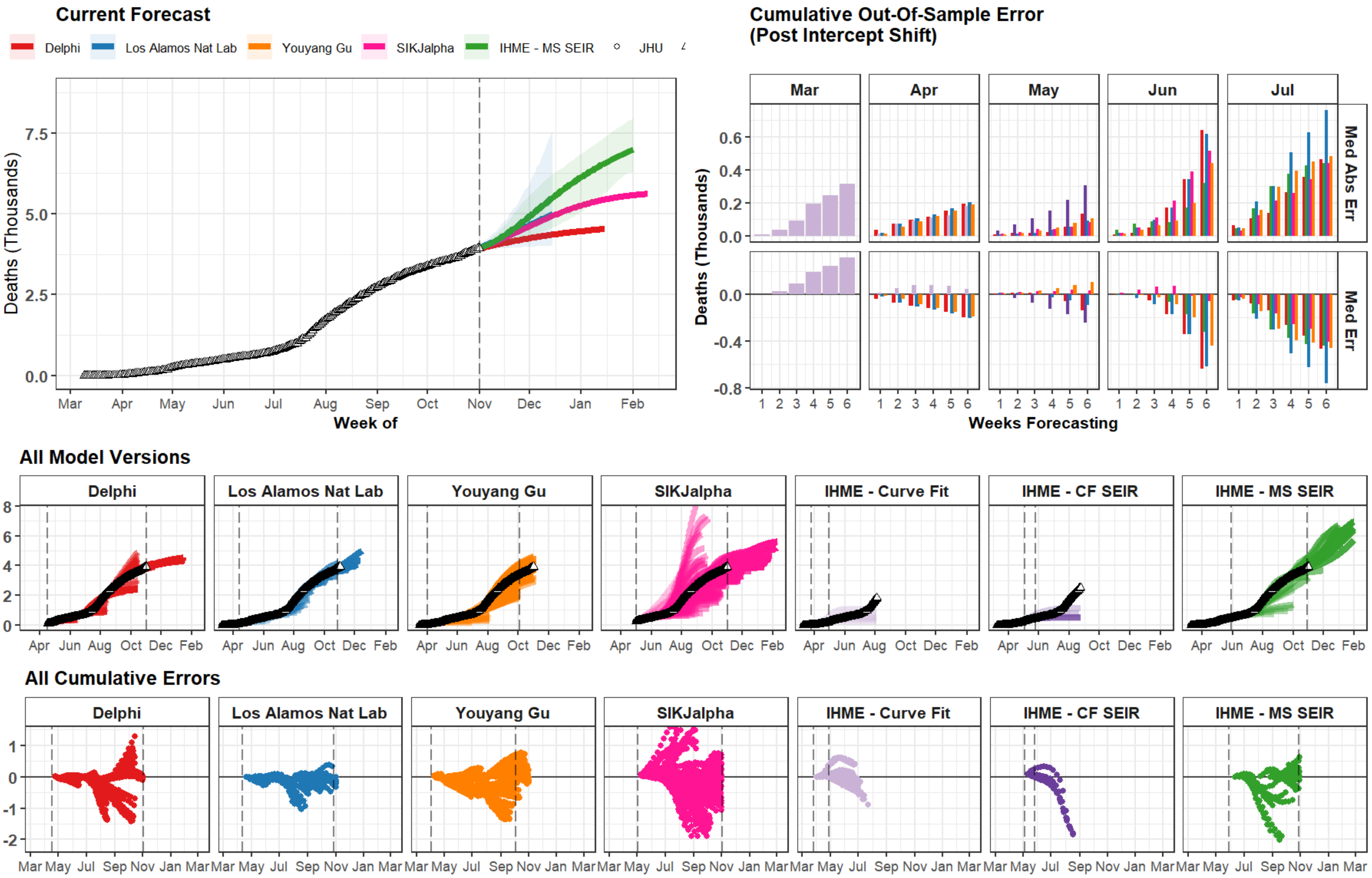

### Morocco

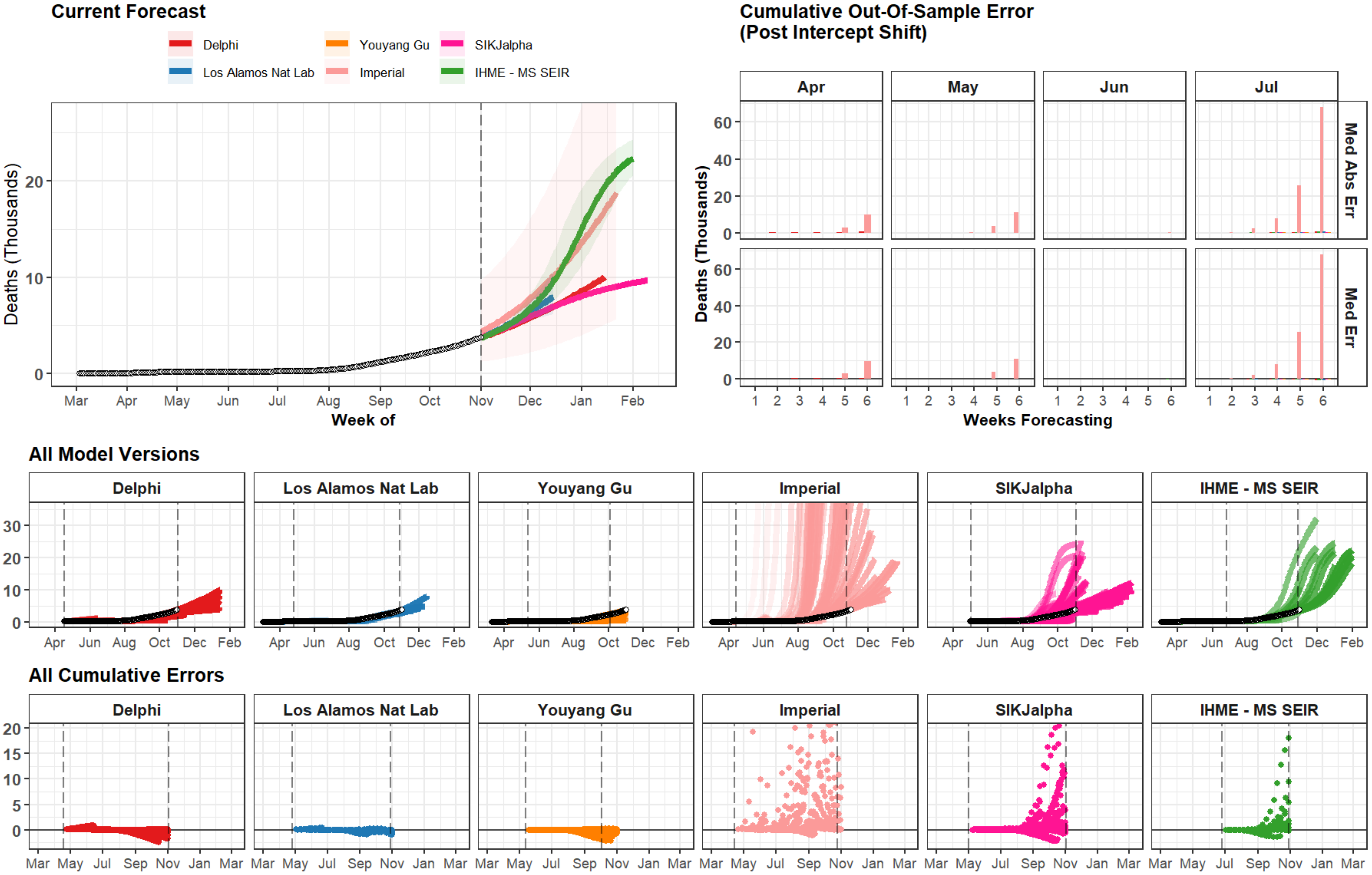

Current Forecast
Delphi
Los Alamos Nat Lab
Youyang Gu
Imperial
SIKJalpha
IHME - MS SEIR
Deaths (Thousands)
Week of
Mar Apr May Jun Jul Aug Sep Oct Nov Dec Jan Feb
Cumulative Out-Of-Sample Error
(Post Intercept Shift)
Apr May Jun Jul
Med Abs Err
Med Err
Weeks Forecasting
All Model Versions
All Cumulative Errors

### Guatemala

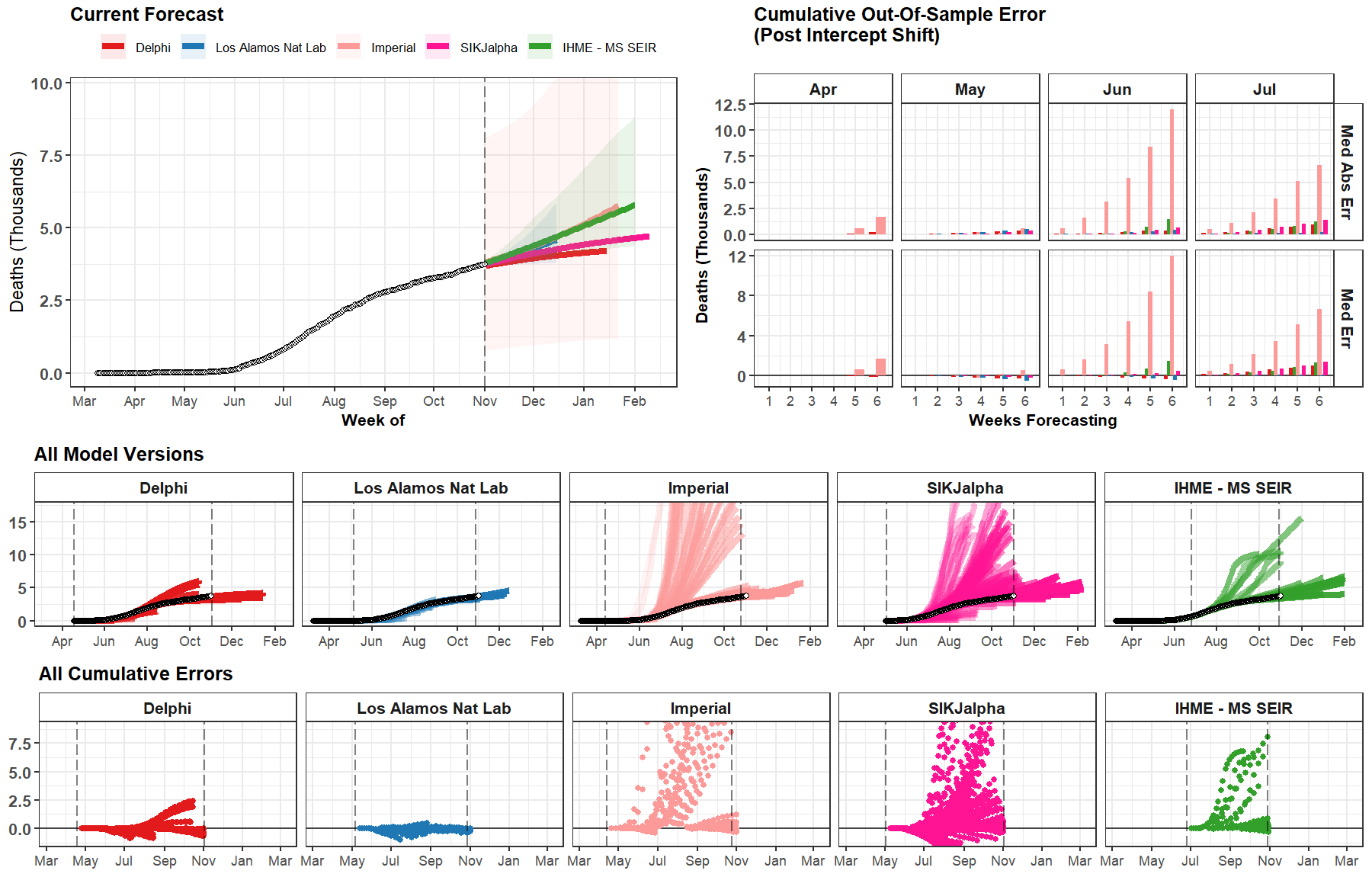
Current Forecast
Delphi
Los Alamos Nat Lab
Imperial
SIKJalpha
IHME - MS SEIR
Deaths (Thousands)
Week of
Cumulative Out-Of-Sample Error
(Post Intercept Shift)
Apr
May
Jun
Jul
Med Abs Err
Med Err
Weeks Forecasting
All Model Versions
All Cumulative Errors

### Virginia

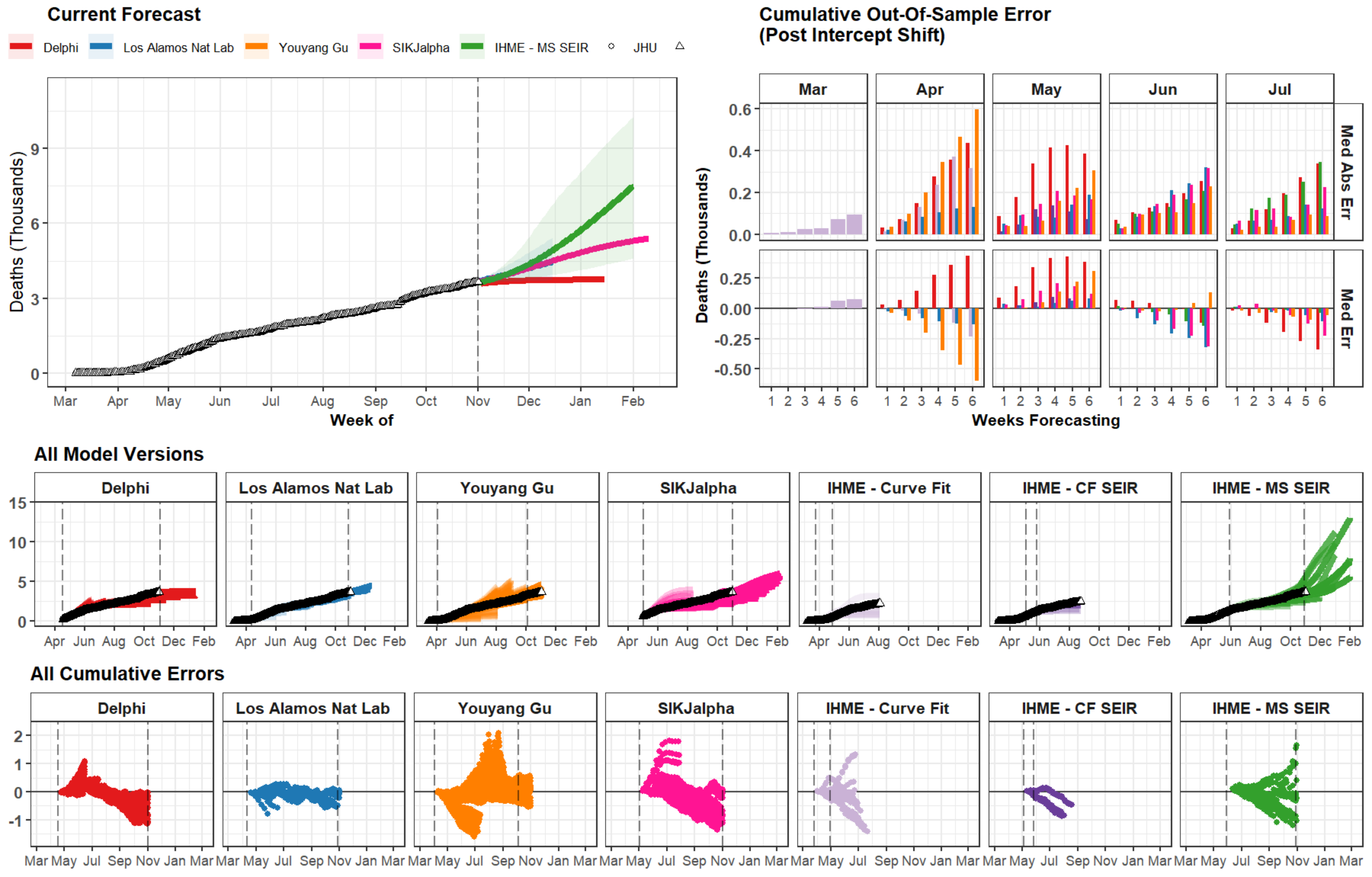

Current Forecast
Delphi
Los Alamos Nat Lab
Youyang Gu
SIKJalpha
IHME - MS SEIR
JHU
Deaths (Thousands)
Week of
Mar Apr May Jun Jul Aug Sep Oct Nov Dec Jan Feb
Cumulative Out-Of-Sample Error
(Post Intercept Shift)
Mar Apr May Jun Jul
Med Abs Err
Med Err
Weeks Forecasting
All Model Versions
Delphi
Los Alamos Nat Lab
Youyang Gu
SIKJalpha
IHME - Curve Fit
IHME - CF SEIR
IHME - MS SEIR
All Cumulative Errors

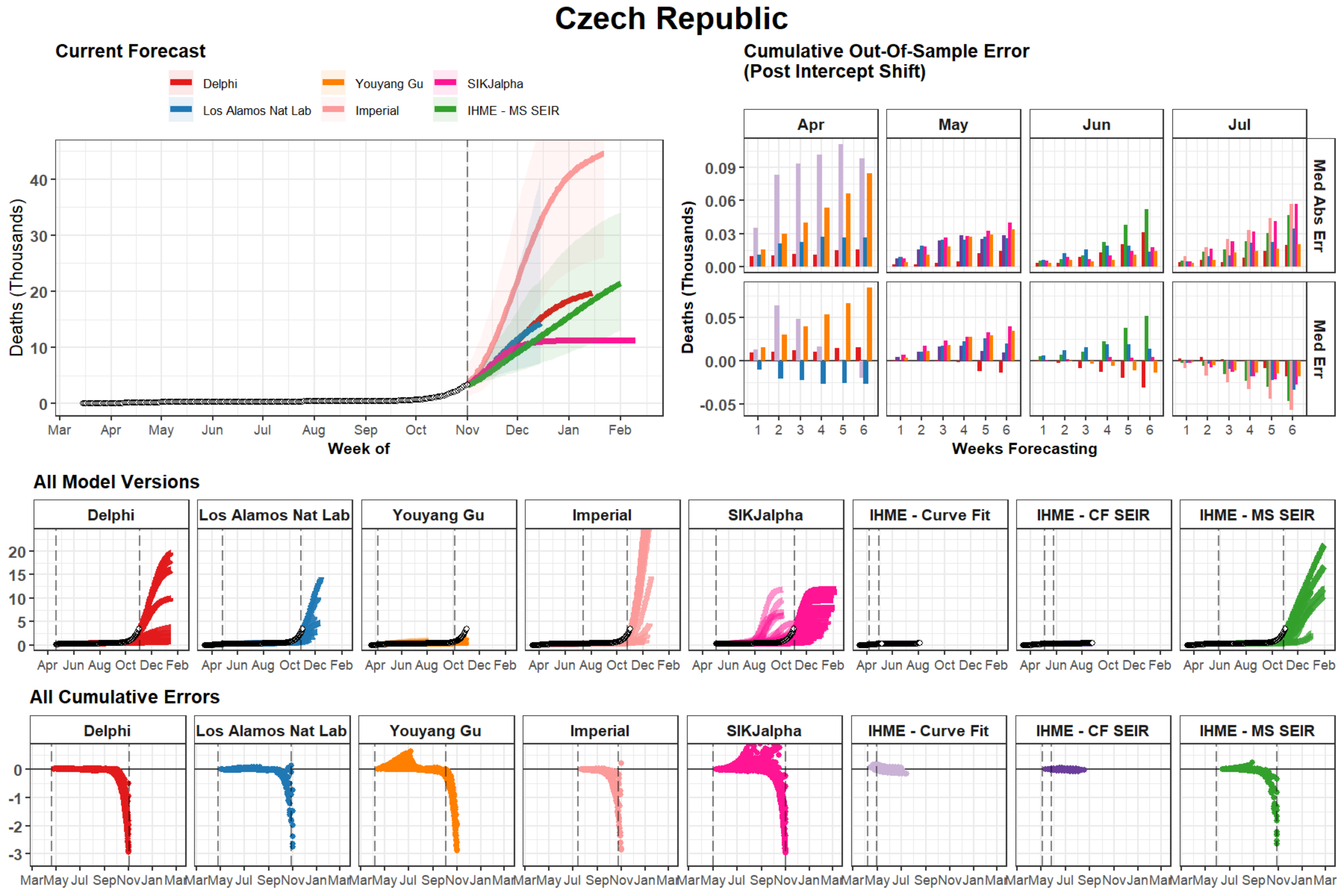
Czech Republic
Current Forecast
Delphi
Los Alamos Nat Lab
Youyang Gu
Imperial
SIKJalpha
IHME - MS SEIR
Deaths (Thousands)
Week of
Mar Apr May Jun Jul Aug Sep Oct Nov Dec Jan Feb
Cumulative Out-Of-Sample Error
(Post Intercept Shift)
Apr May Jun Jul
Med Abs Err
Med Err
Weeks Forecasting
All Model Versions
Delphi
Los Alamos Nat Lab
Youyang Gu
Imperial
SIKJalpha
IHME - Curve Fit
IHME - CF SEIR
IHME - MS SEIR
All Cumulative Errors

### Mississippi

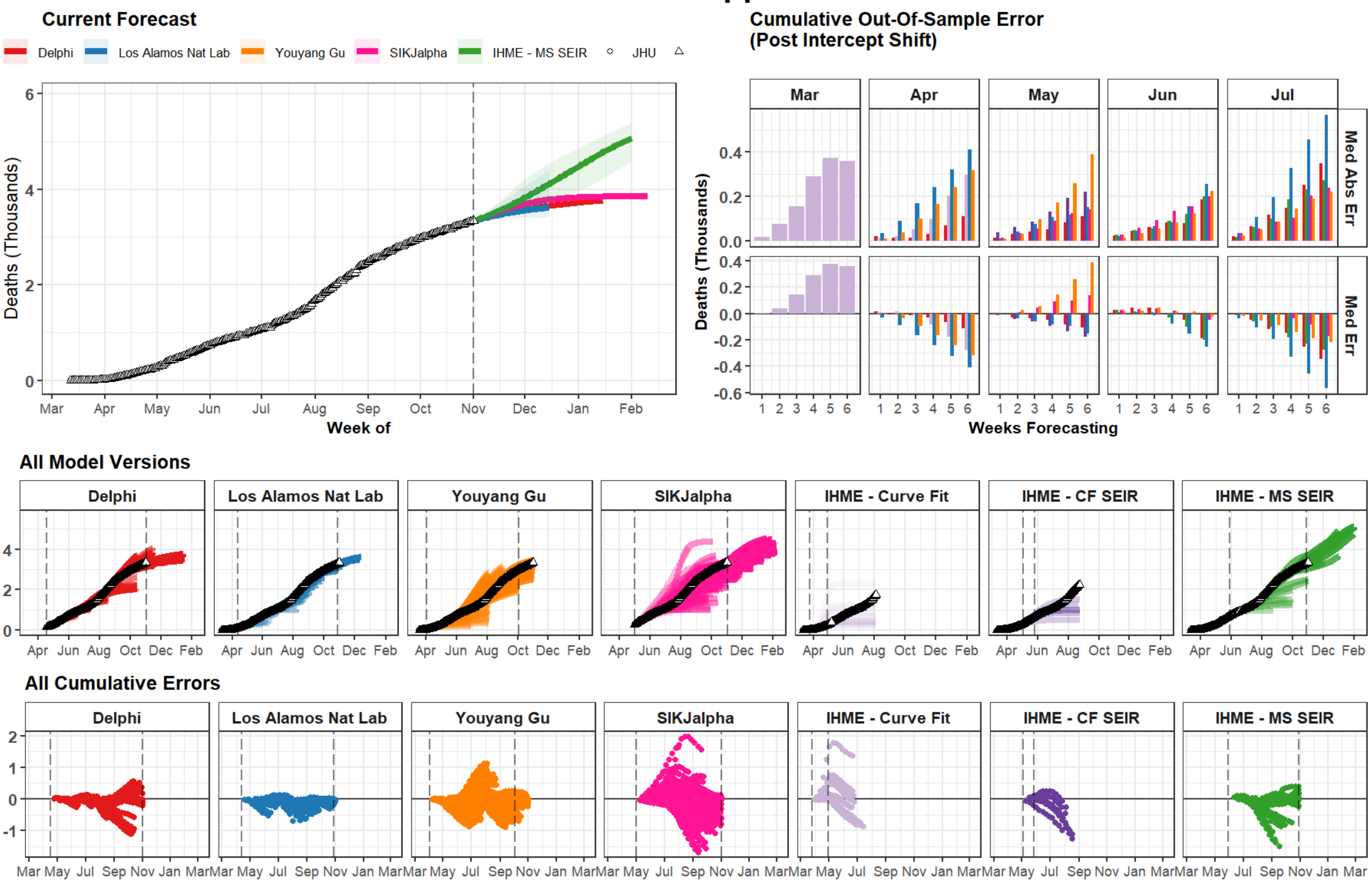

### Tennessee

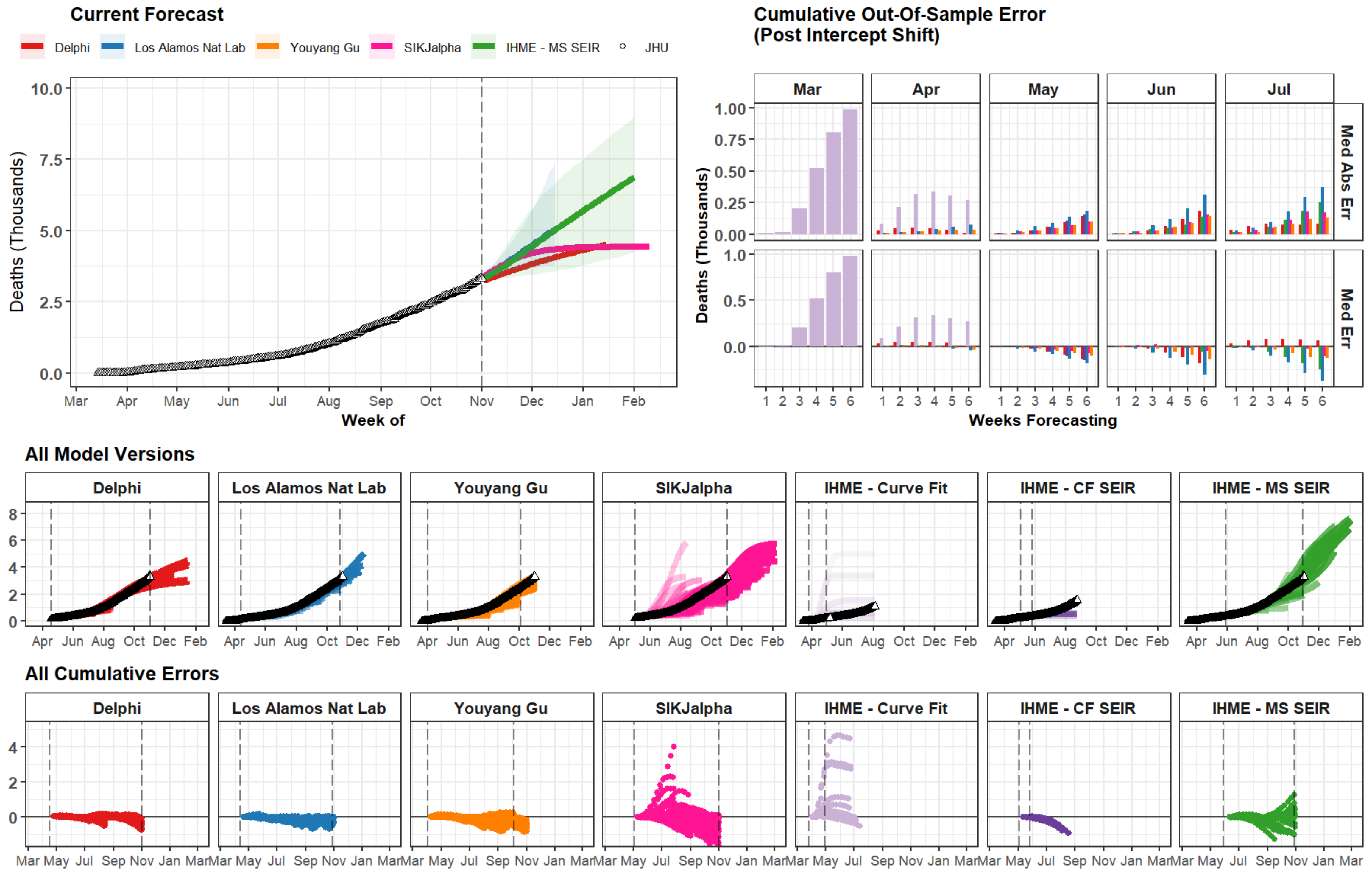

Current Forecast
Delphi
Los Alamos Nat Lab
Youyang Gu
SIKJalpha
IHME - MS SEIR
JHU
Deaths (Thousands)
Week of
Cumulative Out-Of-Sample Error
(Post Intercept Shift)
Mar
Apr
May
Jun
Jul
Med Abs Err
Med Err
Weeks Forecasting
All Model Versions
Delphi
Los Alamos Nat Lab
Youyang Gu
SIKJalpha
IHME - Curve Fit
IHME - CF SEIR
IHME - MS SEIR
All Cumulative Errors

### Missouri

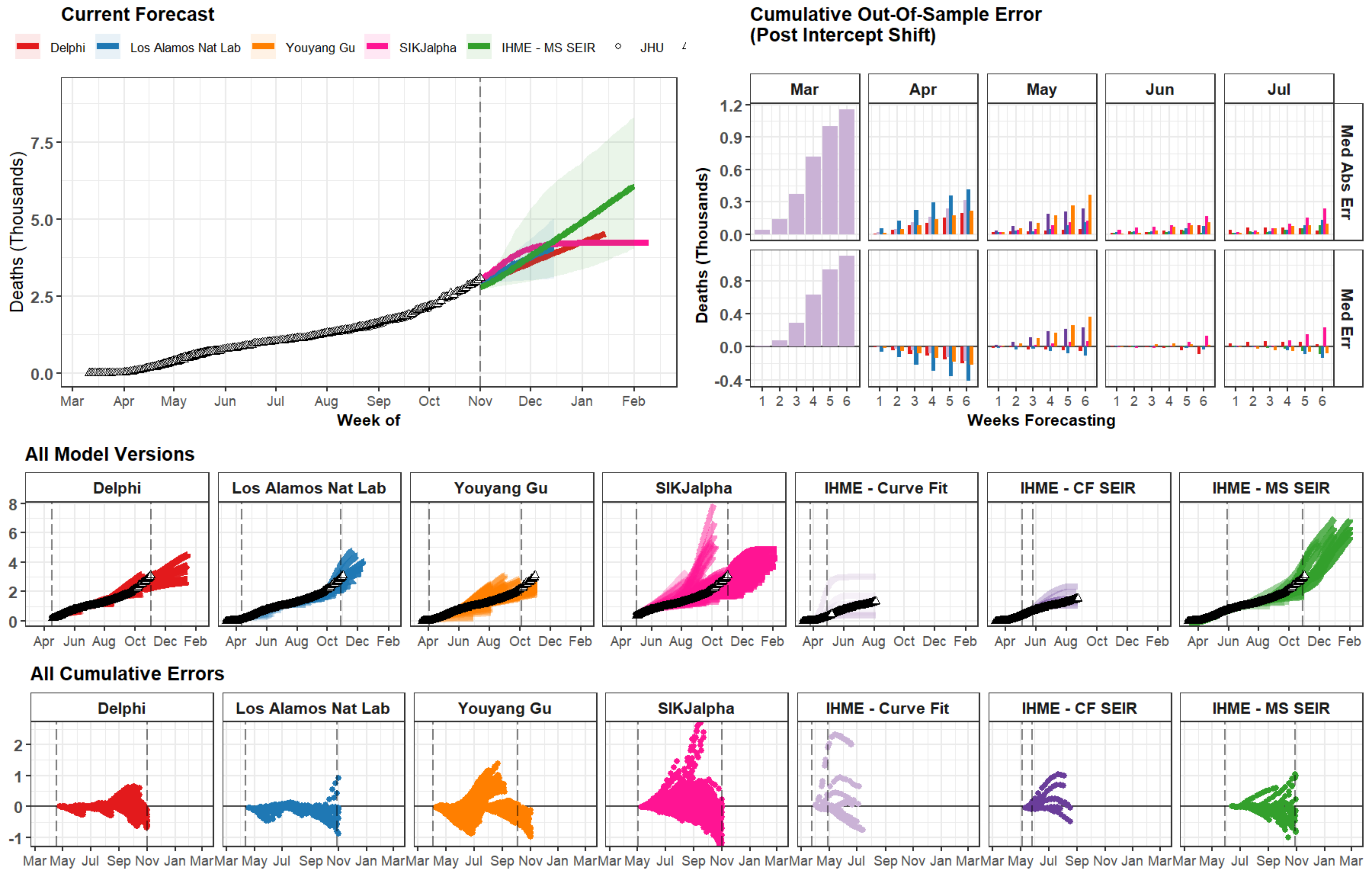
Current Forecast
Delphi
Los Alamos Nat Lab
Youyang Gu
SIKJalpha
IHME - MS SEIR
JHU
Deaths (Thousands)
Week of
Cumulative Out-Of-Sample Error
(Post Intercept Shift)
Mar
Apr
May
Jun
Jul
Med Abs Err
Med Err
Weeks Forecasting
All Model Versions
Delphi
Los Alamos Nat Lab
Youyang Gu
SIKJalpha
IHME - Curve Fit
IHME - CF SEIR
IHME - MS SEIR
All Cumulative Errors

### Alabama

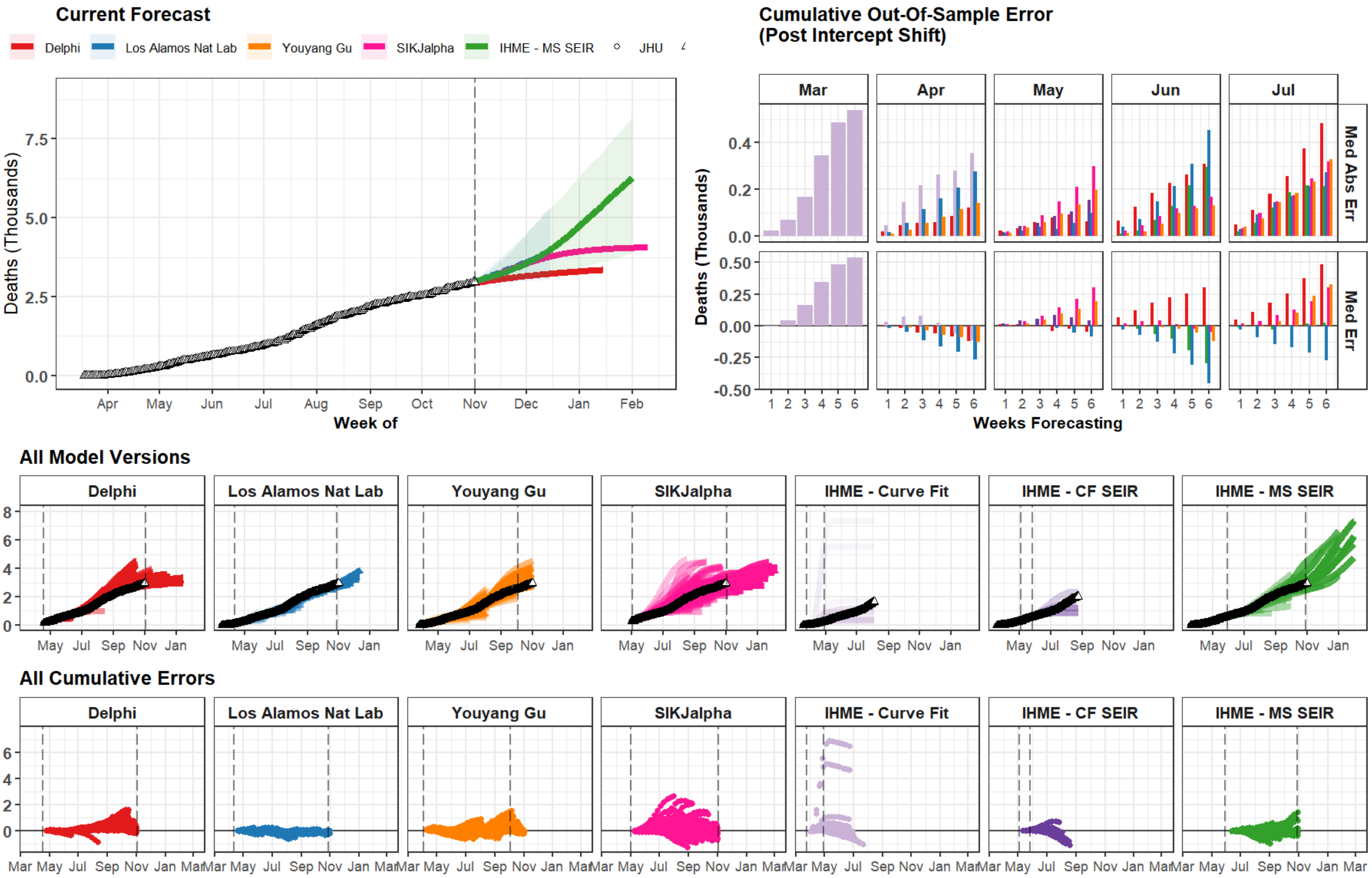
Current Forecast
Delphi
Los Alamos Nat Lab
Youyang Gu
SIKJalpha
IHME - MS SEIR
JHU
Deaths (Thousands)
7.5
5.0
2.5
0.0
Apr May Jun Jul Aug Sep Oct Nov Dec Jan Feb
Week of
Cumulative Out-Of-Sample Error
(Post Intercept Shift)
Mar Apr May Jun Jul
Med Abs Err
Med Err
0.4
0.2
0.0
0.50
0.25
0.00
-0.25
-0.50
1 2 3 4 5 6
Weeks Forecasting
All Model Versions
Delphi
Los Alamos Nat Lab
Youyang Gu
SIKJalpha
IHME - Curve Fit
IHME - CF SEIR
IHME - MS SEIR
8
6
4
2
0
May Jul Sep Nov Jan
All Cumulative Errors
6
4
2
0
Mar May Jul Sep Nov Jan Mar

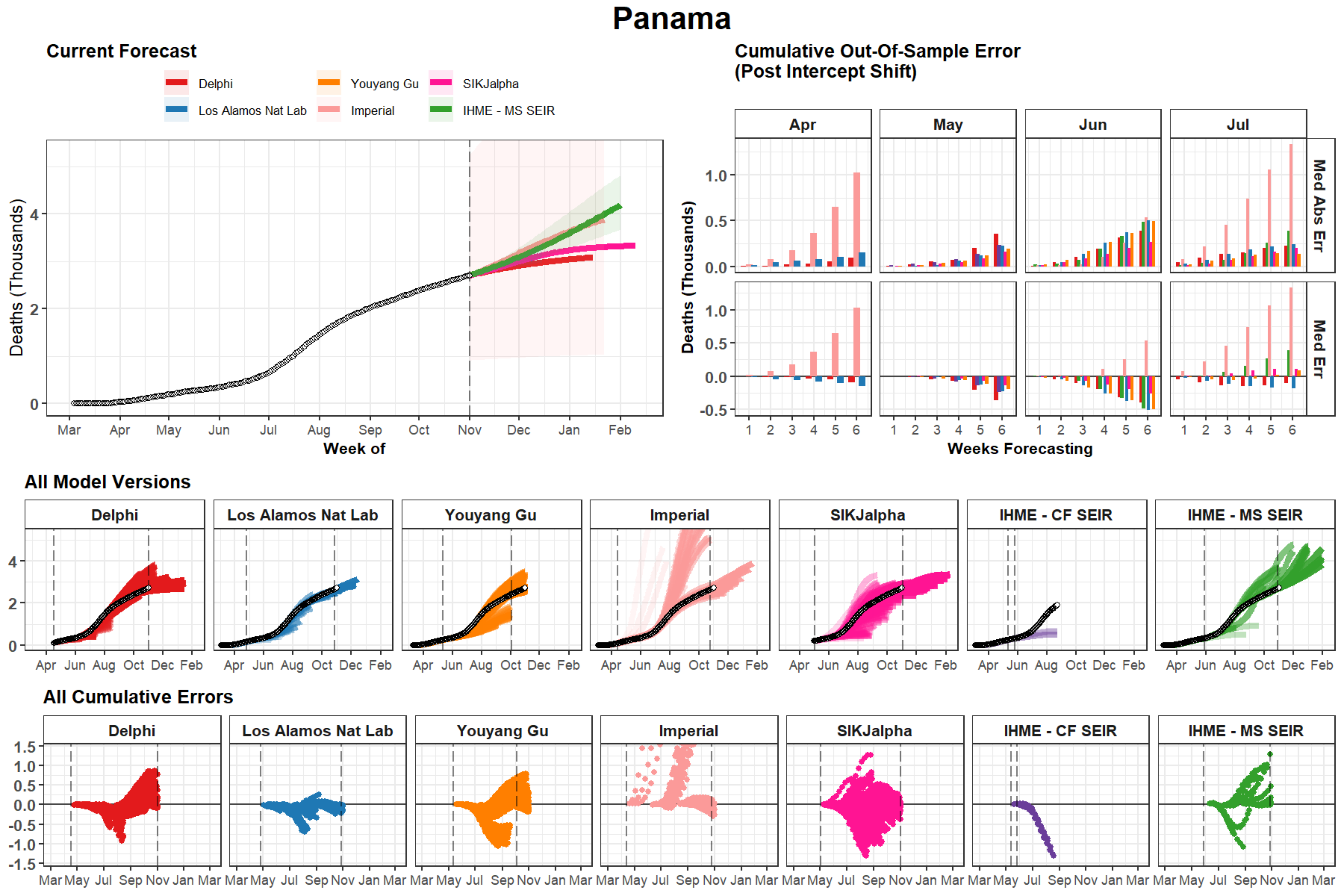

Panama
Current Forecast
Delphi
Youyang Gu
SIKJalpha
Los Alamos Nat Lab
Imperial
IHME - MS SEIR
Deaths (Thousands)
Week of
Mar Apr May Jun Jul Aug Sep Oct Nov Dec Jan Feb
Cumulative Out-Of-Sample Error
(Post Intercept Shift)
Apr
May
Jun
Jul
Med Abs Err
Med Err
Weeks Forecasting
All Model Versions
Delphi
Los Alamos Nat Lab
Youyang Gu
Imperial
SIKJalpha
IHME - CF SEIR
IHME - MS SEIR
All Cumulative Errors

### Honduras

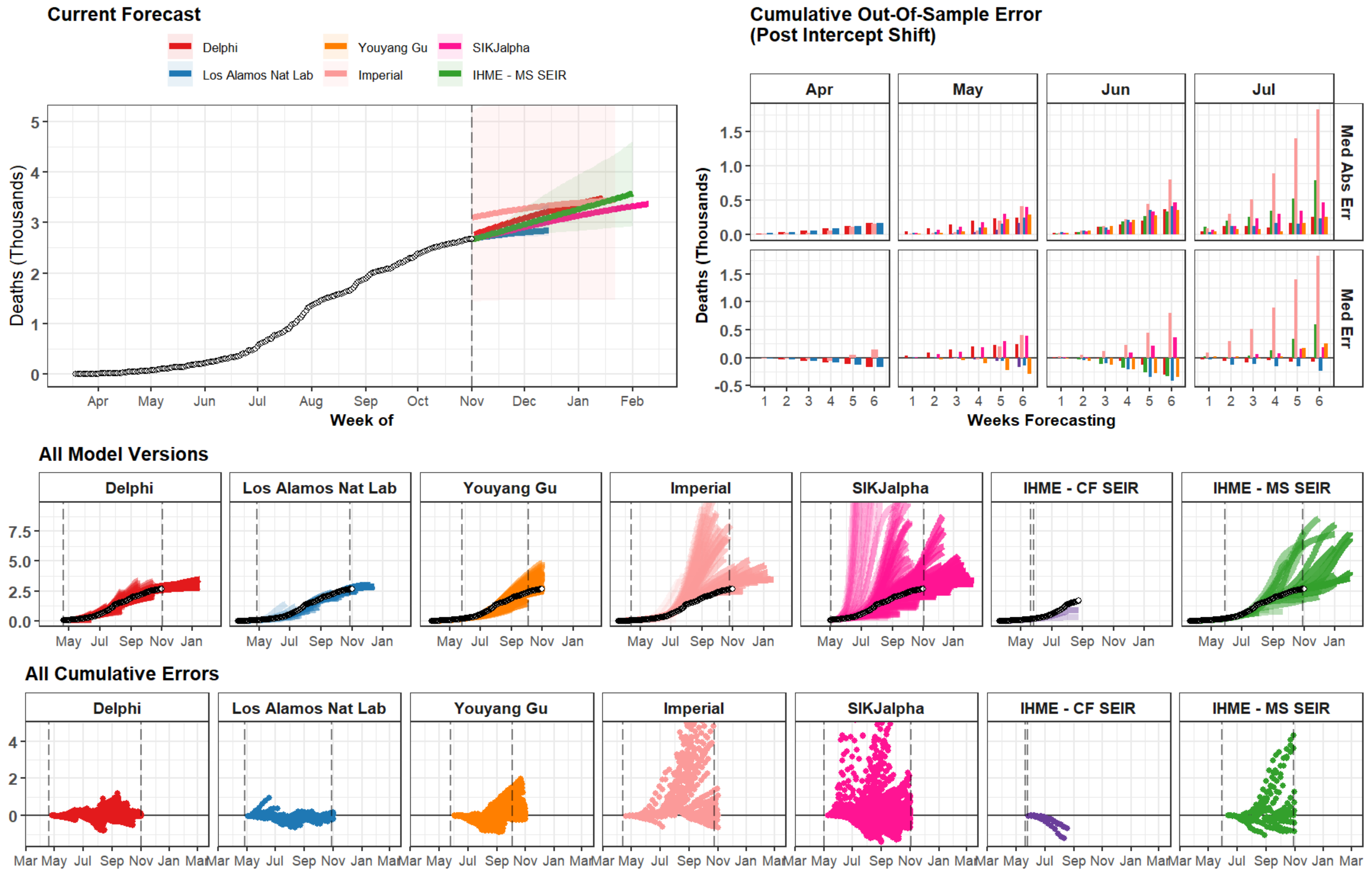
Current Forecast
Delphi
Los Alamos Nat Lab
Youyang Gu
Imperial
SIKJalpha
IHME - MS SEIR
Deaths (Thousands)
Week of
Apr
May
Jun
Jul
Aug
Sep
Oct
Nov
Dec
Jan
Feb
Cumulative Out-Of-Sample Error
(Post Intercept Shift)
Apr
May
Jun
Jul
Med Abs Err
Med Err
Weeks Forecasting
All Model Versions
Delphi
Los Alamos Nat Lab
Youyang Gu
Imperial
SIKJalpha
IHME - CF SEIR
IHME - MS SEIR
All Cumulative Errors

### Israel

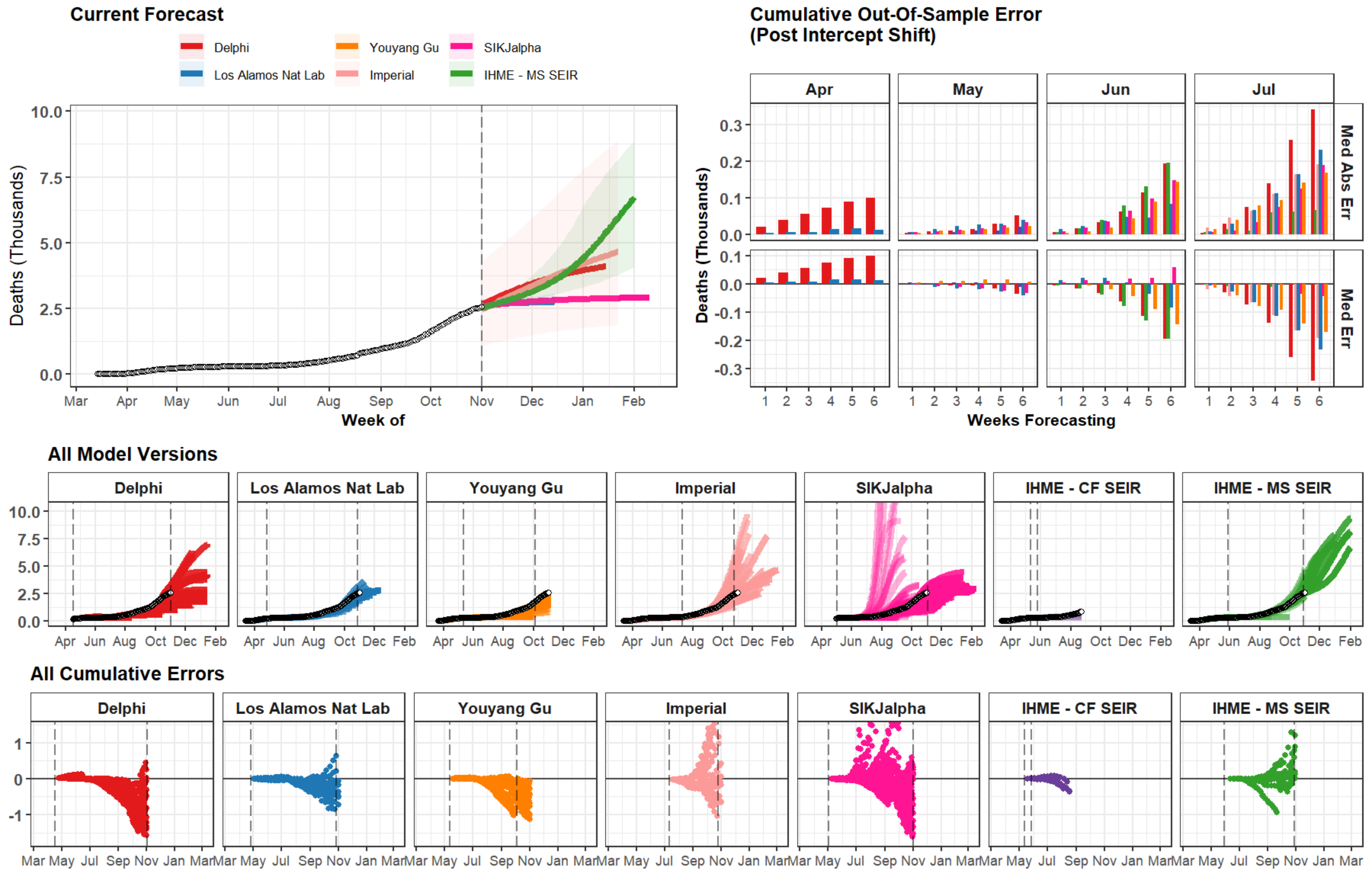

Current Forecast
Delphi
Youyang Gu
SIKJalpha
Los Alamos Nat Lab
Imperial
IHME - MS SEIR
Deaths (Thousands)
Week of
Cumulative Out-Of-Sample Error
(Post Intercept Shift)
Apr
May
Jun
Jul
Med Abs Err
Med Err
Weeks Forecasting
All Model Versions
Delphi
Los Alamos Nat Lab
Youyang Gu
Imperial
SIKJalpha
IHME - CF SEIR
IHME - MS SEIR
All Cumulative Errors

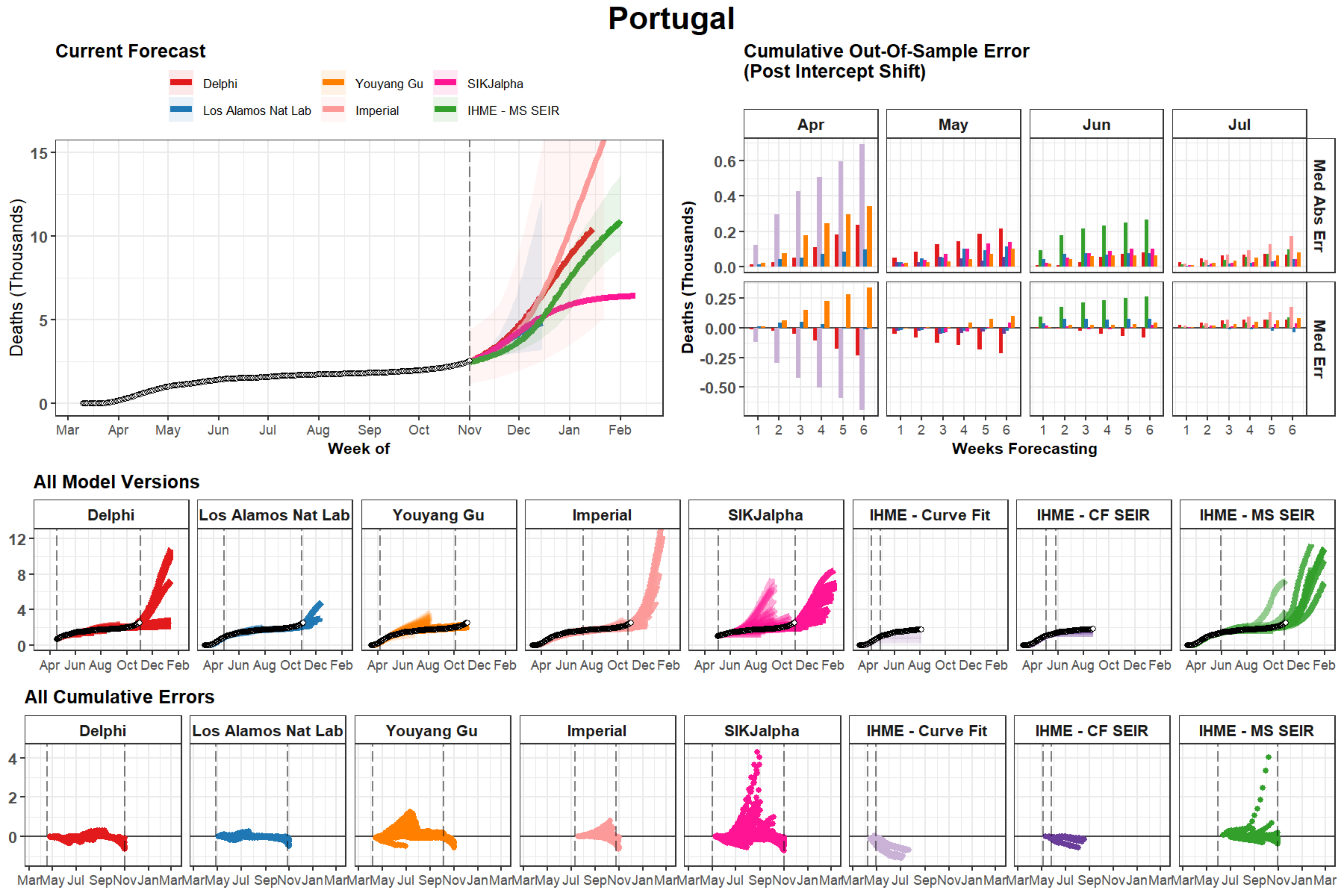

Portugal
Current Forecast
Delphi
Los Alamos Nat Lab
Youyang Gu
Imperial
SIKJalpha
IHME - MS SEIR
Deaths (Thousands)
Week of
Mar Apr May Jun Jul Aug Sep Oct Nov Dec Jan Feb
Cumulative Out-Of-Sample Error
(Post Intercept Shift)
Apr
May
Jun
Jul
Med Abs Err
Med Err
Weeks Forecasting
All Model Versions
Delphi
Los Alamos Nat Lab
Youyang Gu
Imperial
SIKJalpha
IHME - Curve Fit
IHME - CF SEIR
IHME - MS SEIR
Apr Jun Aug Oct Dec Feb
All Cumulative Errors
Mar May Jul Sep Nov Jan Mar

### Minnesota

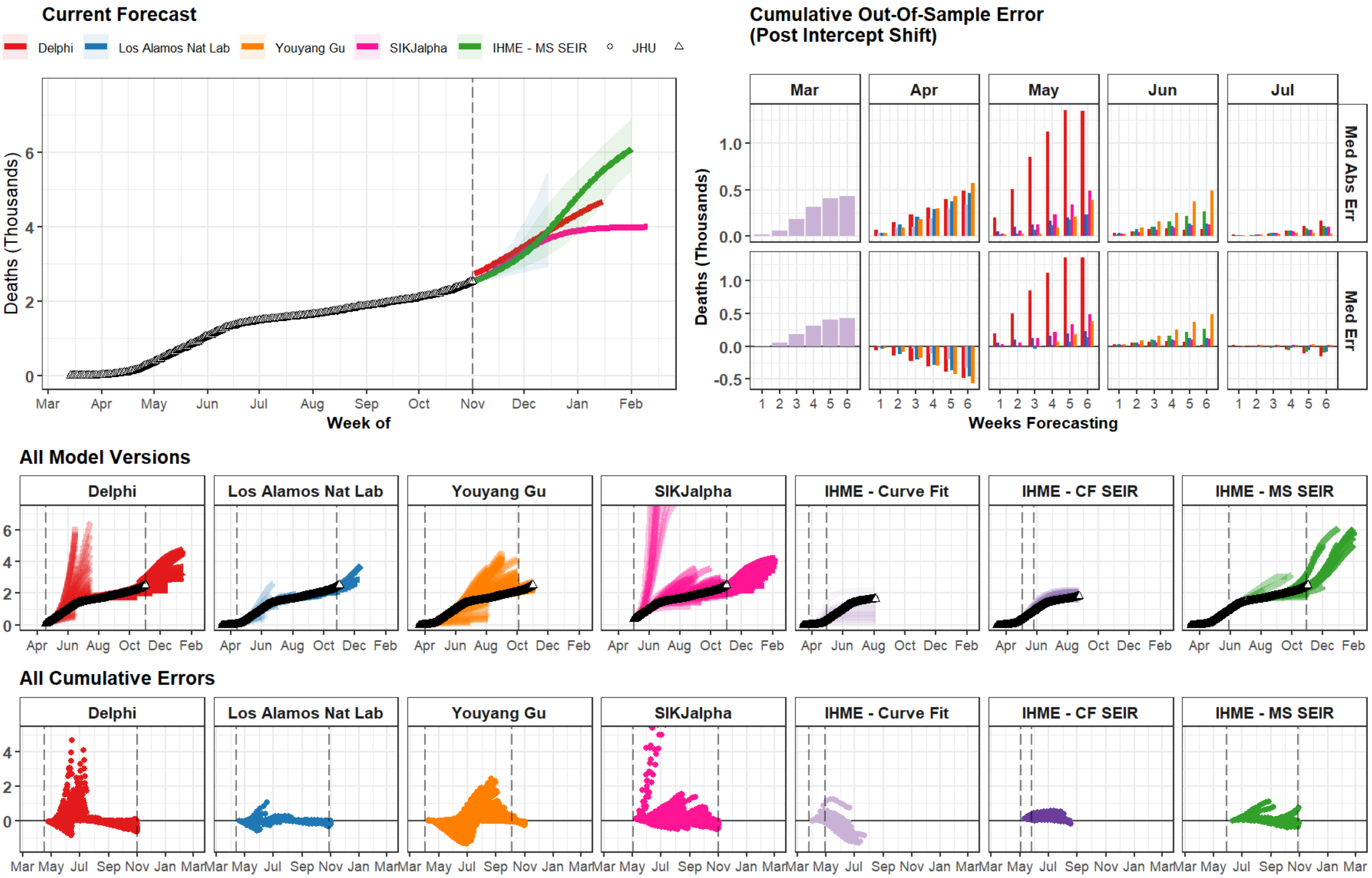

Current Forecast
Delphi
Los Alamos Nat Lab
Youyang Gu
SIKJalpha
IHME - MS SEIR
JHU
Deaths (Thousands)
Week of
Mar Apr May Jun Jul Aug Sep Oct Nov Dec Jan Feb
Cumulative Out-Of-Sample Error
(Post Intercept Shift)
Mar Apr May Jun Jul
Med Abs Err
Med Err
Weeks Forecasting
All Model Versions
Delphi
Los Alamos Nat Lab
Youyang Gu
SIKJalpha
IHME - Curve Fit
IHME - CF SEIR
IHME - MS SEIR
Apr Jun Aug Oct Dec Feb
All Cumulative Errors
Mar May Jul Sep Nov Jan Mar

### Washington

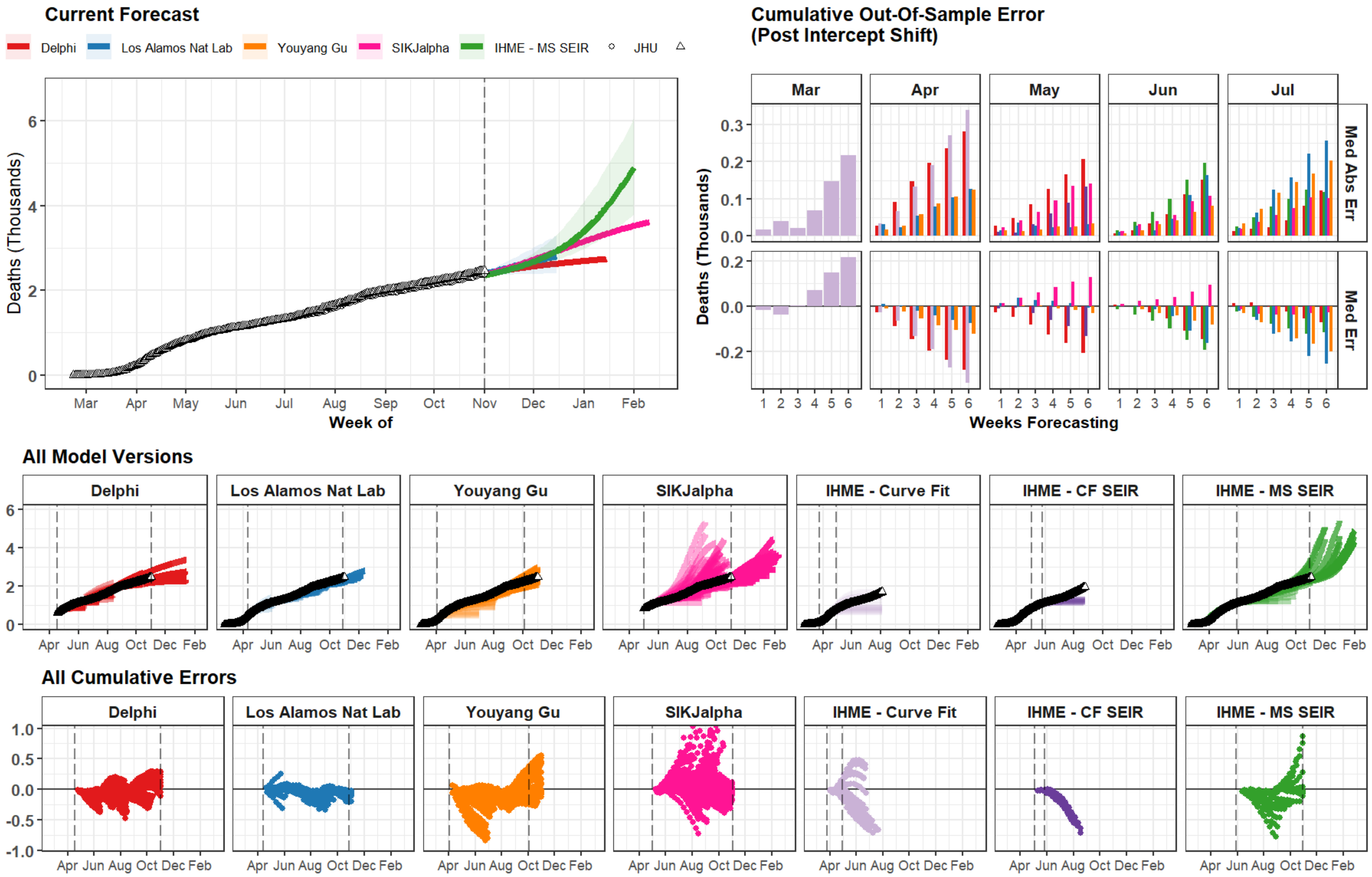

Current Forecast
Delphi
Los Alamos Nat Lab
Youyang Gu
SIKJalpha
IHME - MS SEIR
JHU
Deaths (Thousands)
Week of
Mar
Apr
May
Jun
Jul
Aug
Sep
Oct
Nov
Dec
Jan
Feb
Cumulative Out-Of-Sample Error
(Post Intercept Shift)
Med Abs Err
Med Err
Weeks Forecasting
All Model Versions
IHME - Curve Fit
IHME - CF SEIR
All Cumulative Errors

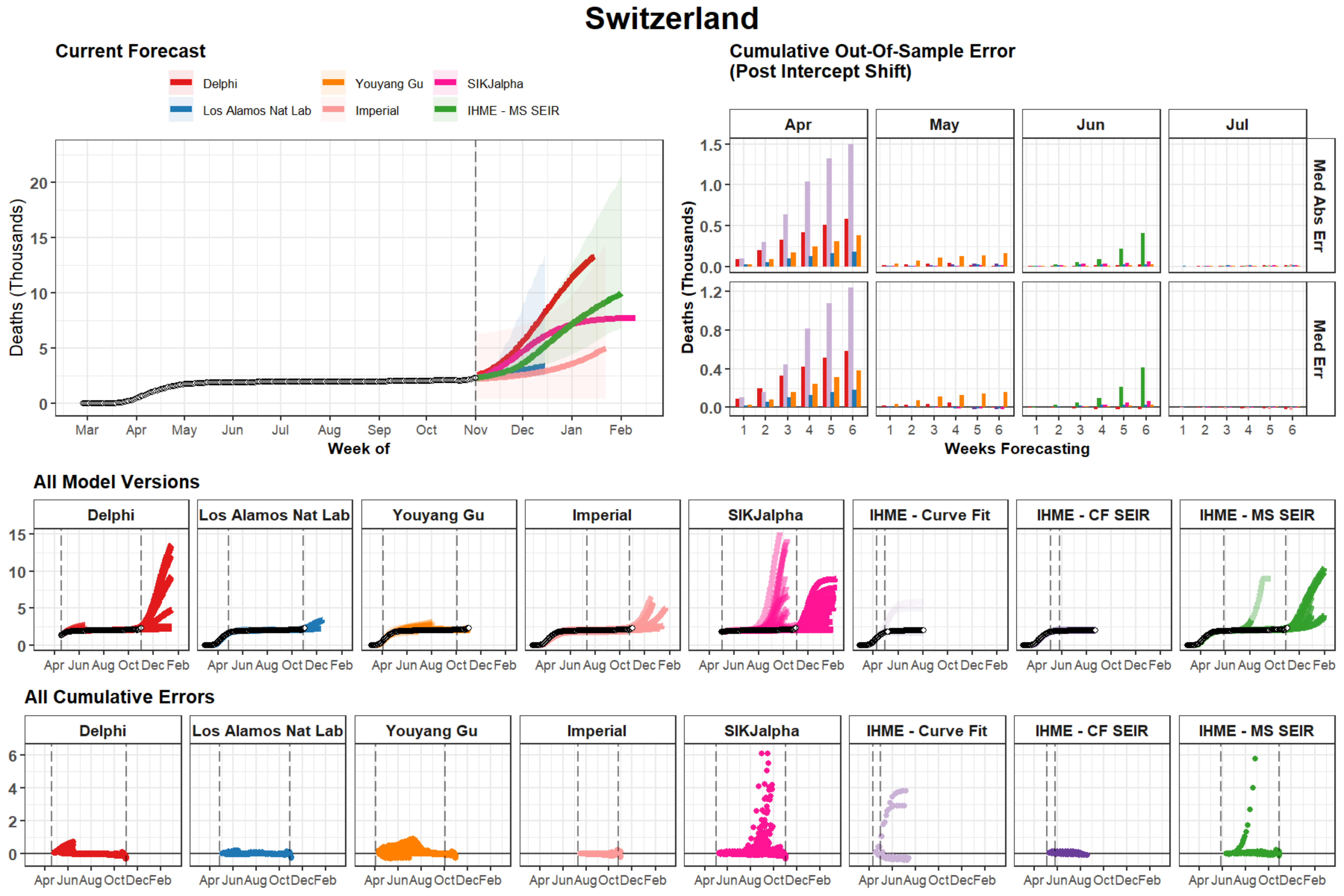
Switzerland
Current Forecast
Delphi
Los Alamos Nat Lab
Youyang Gu
Imperial
SIKJalpha
IHME - MS SEIR
Deaths (Thousands)
Week of
Mar Apr May Jun Jul Aug Sep Oct Nov Dec Jan Feb
Cumulative Out-Of-Sample Error
(Post Intercept Shift)
Apr
May
Jun
Jul
Med Abs Err
Med Err
Weeks Forecasting
All Model Versions
Delphi
Los Alamos Nat Lab
Youyang Gu
Imperial
SIKJalpha
IHME - Curve Fit
IHME - CF SEIR
IHME - MS SEIR
Apr Jun Aug Oct Dec Feb
All Cumulative Errors

### Colorado

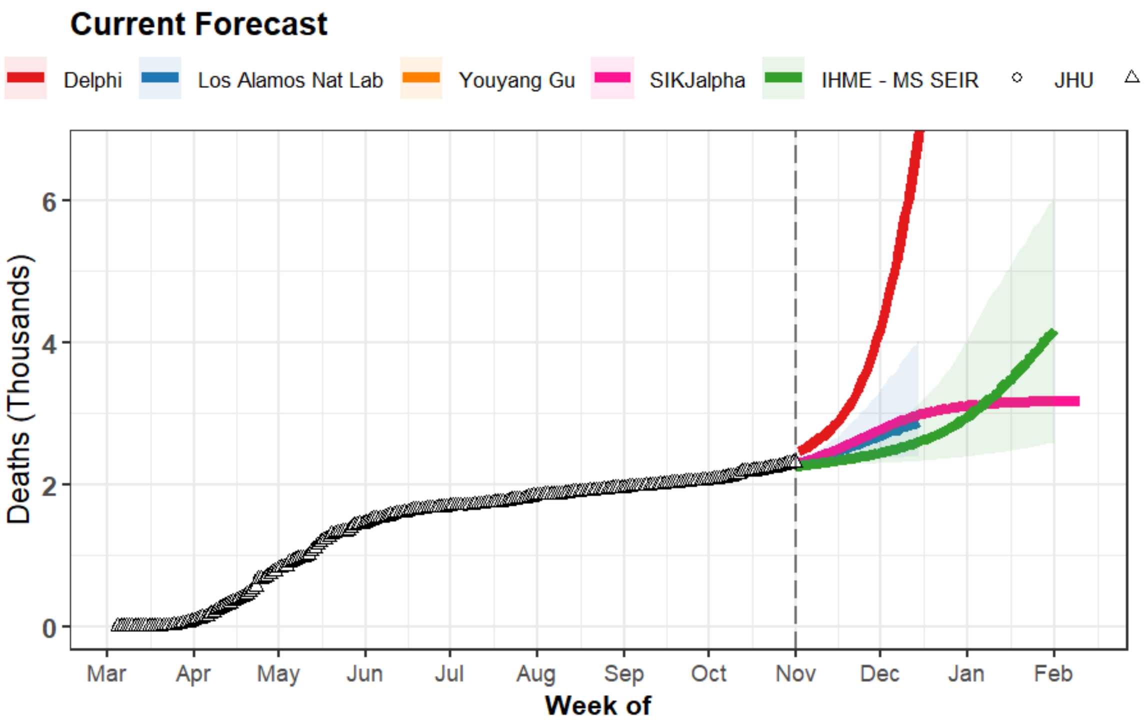

Current Forecast
Delphi
Los Alamos Nat Lab
Youyang Gu
SIKJalpha
IHME - MS SEIR
JHU
Deaths (Thousands)
Week of
Mar Apr May Jun Jul Aug Sep Oct Nov Dec Jan Feb


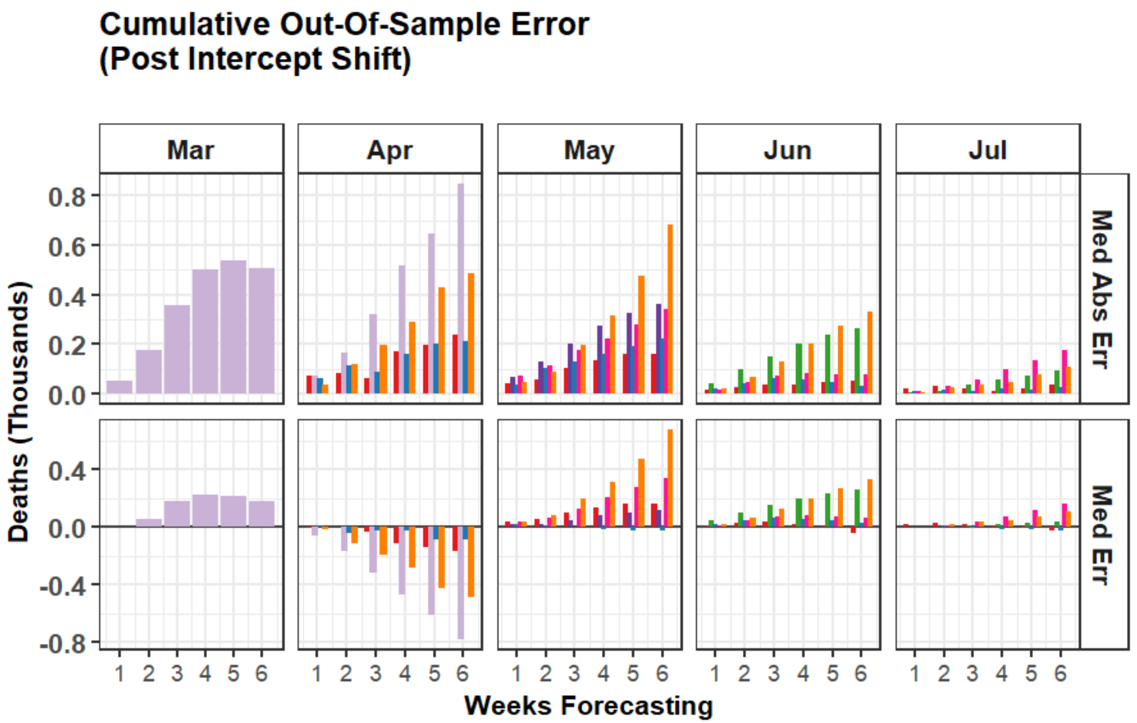

Cumulative Out-Of-Sample Error
(Post Intercept Shift)
Mar
Apr
May
Jun
Jul
Med Abs Err
Med Err
Deaths (Thousands)
Weeks Forecasting


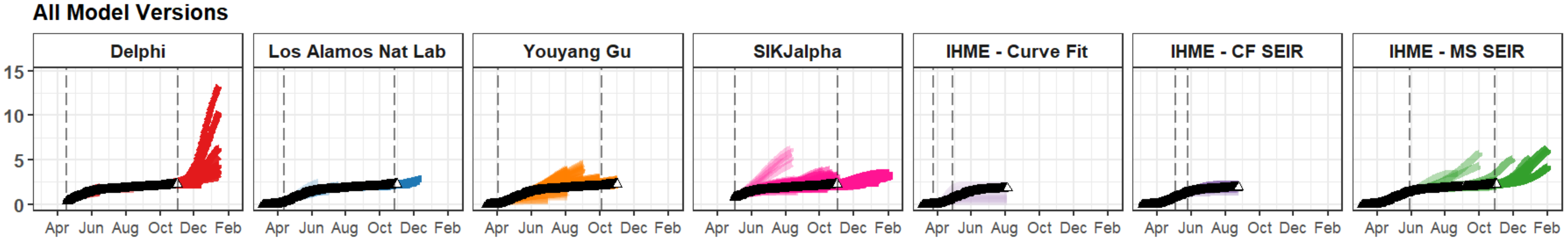

All Model Versions
Delphi
Los Alamos Nat Lab
Youyang Gu
SIKJalpha
IHME - Curve Fit
IHME - CF SEIR
IHME - MS SEIR
Apr Jun Aug Oct Dec Feb


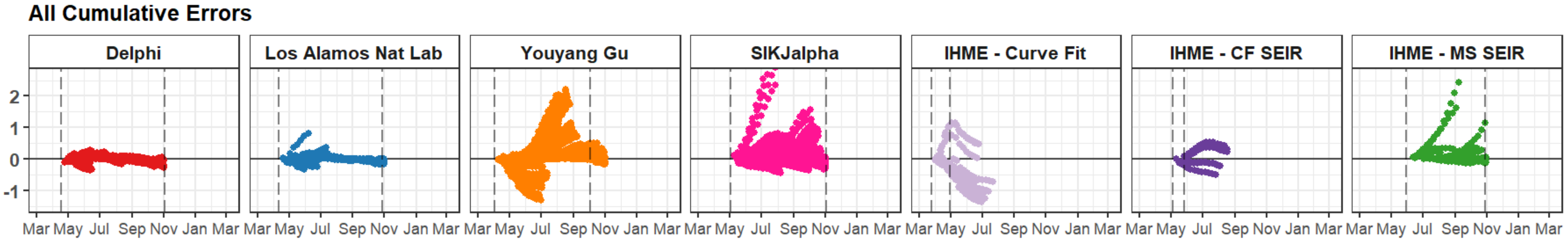

All Cumulative Errors
Delphi
Los Alamos Nat Lab
Youyang Gu
SIKJalpha
IHME - Curve Fit
IHME - CF SEIR
IHME - MS SEIR
Mar May Jul Sep Nov Jan Mar

### Dominican Republic

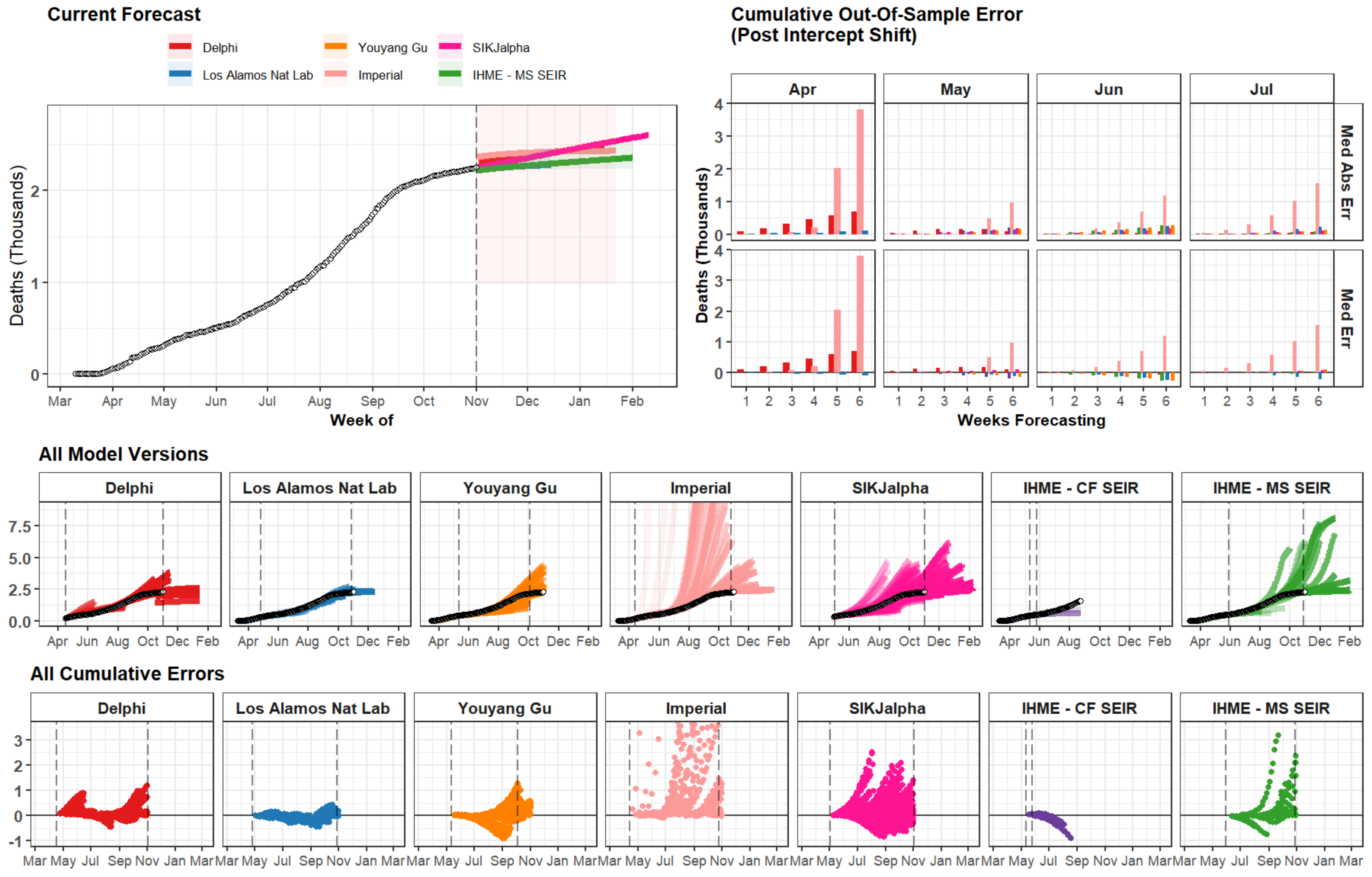

### Wisconsin

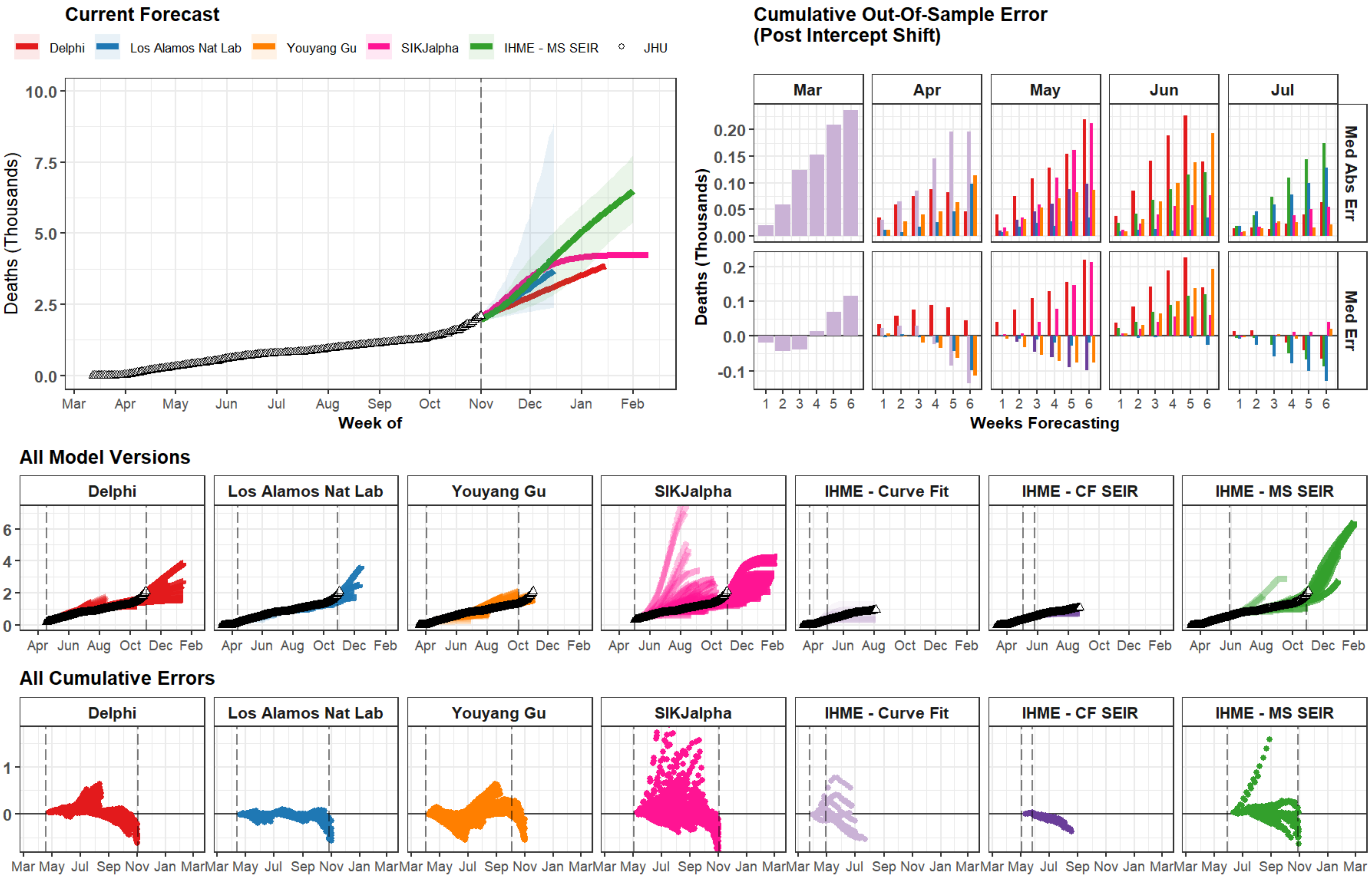

Current Forecast
Delphi
Los Alamos Nat Lab
Youyang Gu
SIKJalpha
IHME - MS SEIR
JHU
Deaths (Thousands)
Week of
Mar Apr May Jun Jul Aug Sep Oct Nov Dec Jan Feb
Cumulative Out-Of-Sample Error
(Post Intercept Shift)
Mar
Apr
May
Jun
Jul
Med Abs Err
Med Err
Deaths (Thousands)
Weeks Forecasting
All Model Versions
Delphi
Los Alamos Nat Lab
Youyang Gu
SIKJalpha
IHME - Curve Fit
IHME - CF SEIR
IHME - MS SEIR
All Cumulative Errors

### Algeria

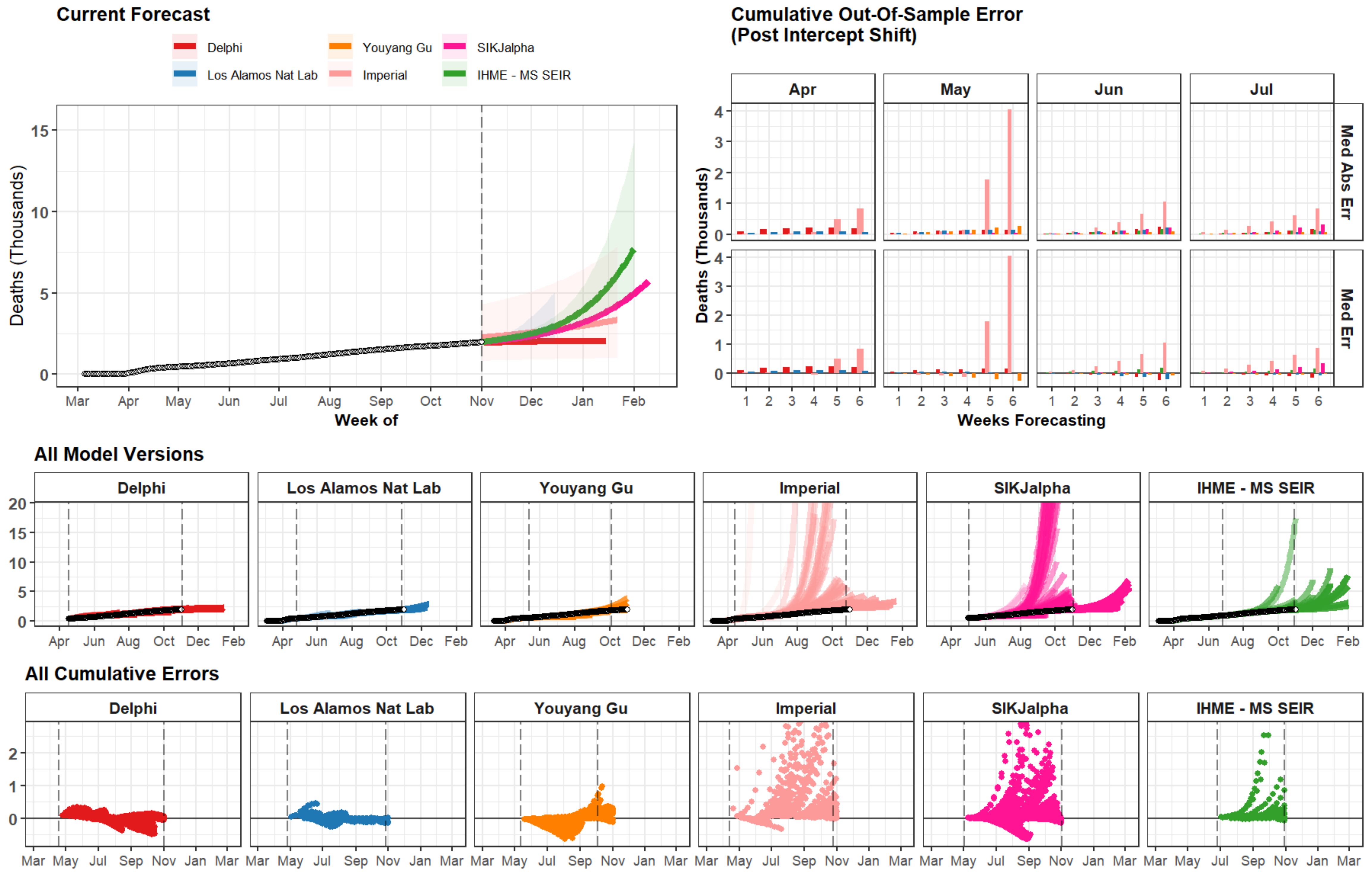
Current Forecast
Delphi
Los Alamos Nat Lab
Youyang Gu
Imperial
SIKJalpha
IHME - MS SEIR
Deaths (Thousands)
Week of
Cumulative Out-Of-Sample Error
(Post Intercept Shift)
Apr
May
Jun
Jul
Med Abs Err
Med Err
Weeks Forecasting
All Model Versions
All Cumulative Errors

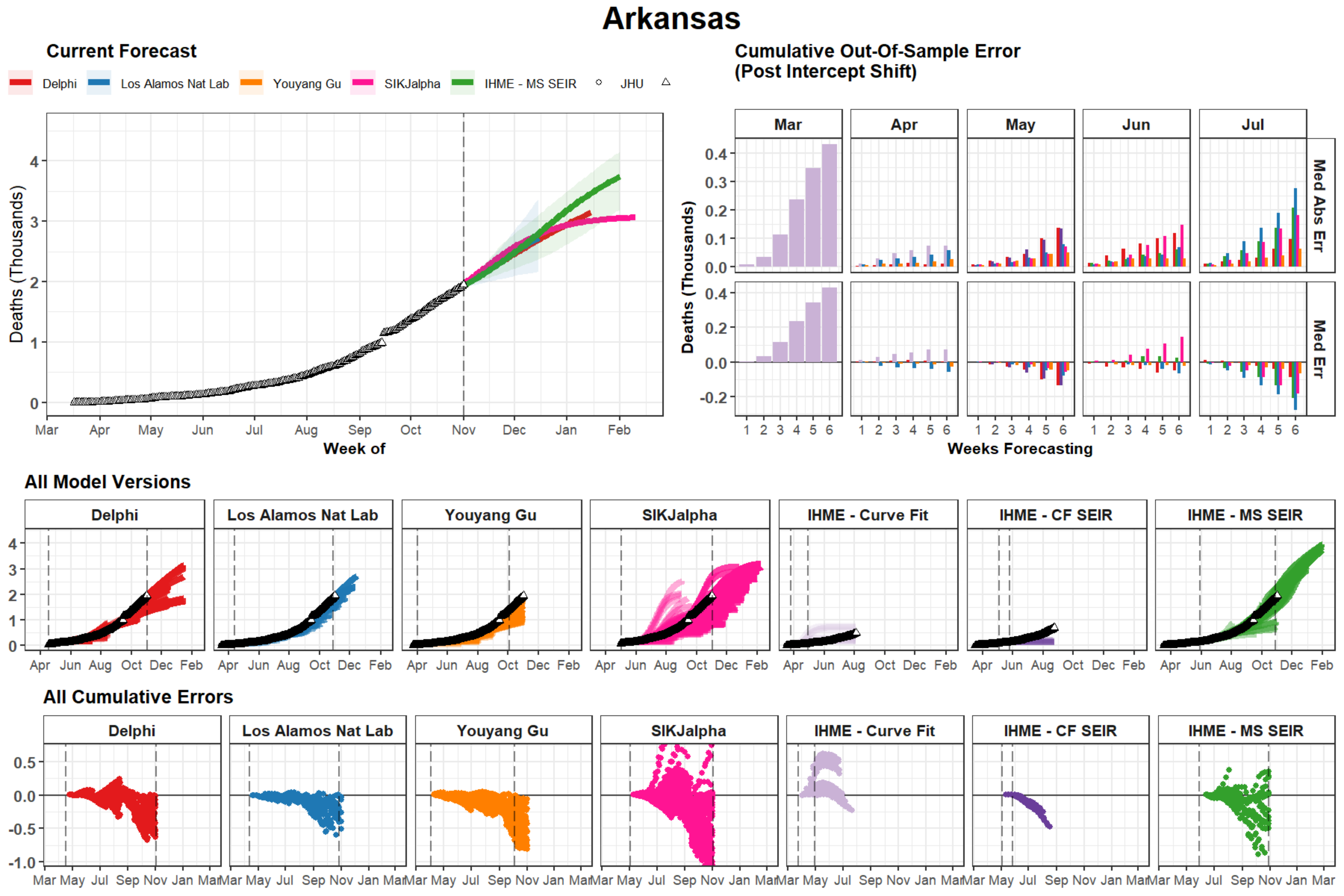
Arkansas
Current Forecast
Delphi
Los Alamos Nat Lab
Youyang Gu
SIKJalpha
IHME - MS SEIR
JHU
Deaths (Thousands)
Week of
Mar Apr May Jun Jul Aug Sep Oct Nov Dec Jan Feb
Cumulative Out-Of-Sample Error
(Post Intercept Shift)
Mar Apr May Jun Jul
Med Abs Err
Med Err
Weeks Forecasting
All Model Versions
Delphi
Los Alamos Nat Lab
Youyang Gu
SIKJalpha
IHME - Curve Fit
IHME - CF SEIR
IHME - MS SEIR
All Cumulative Errors

### Ireland

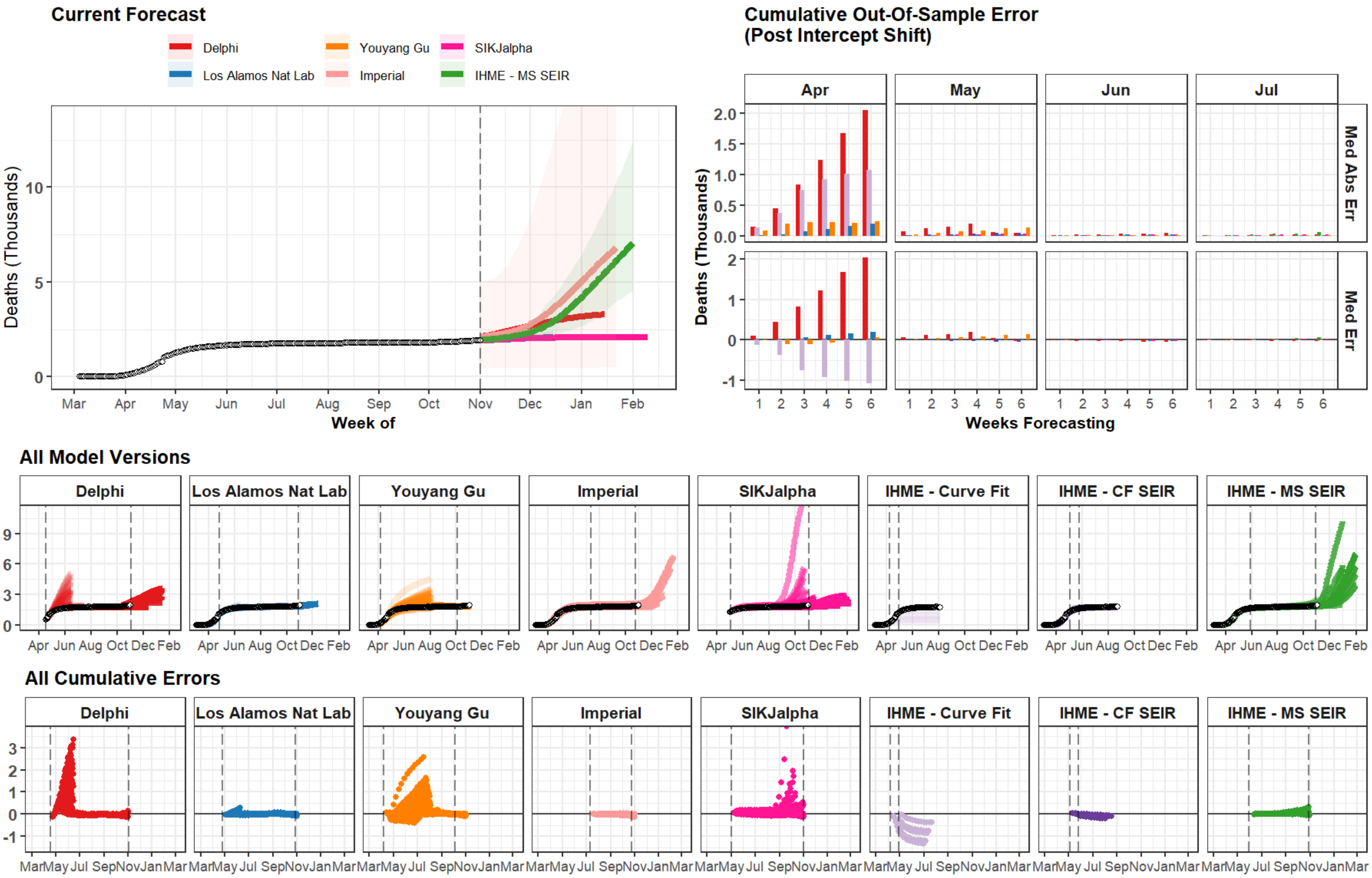

Current Forecast
Delphi
Los Alamos Nat Lab
Youyang Gu
Imperial
SIKJalpha
IHME - MS SEIR
Deaths (Thousands)
Week of
Mar Apr May Jun Jul Aug Sep Oct Nov Dec Jan Feb
Cumulative Out-Of-Sample Error
(Post Intercept Shift)
Apr
May
Jun
Jul
Med Abs Err
Med Err
Weeks Forecasting
All Model Versions
Delphi
Los Alamos Nat Lab
Youyang Gu
Imperial
SIKJalpha
IHME - Curve Fit
IHME - CF SEIR
IHME - MS SEIR
Apr Jun Aug Oct Dec Feb
All Cumulative Errors
Mar May Jul Sep Nov Jan Mar

### Kazakhstan

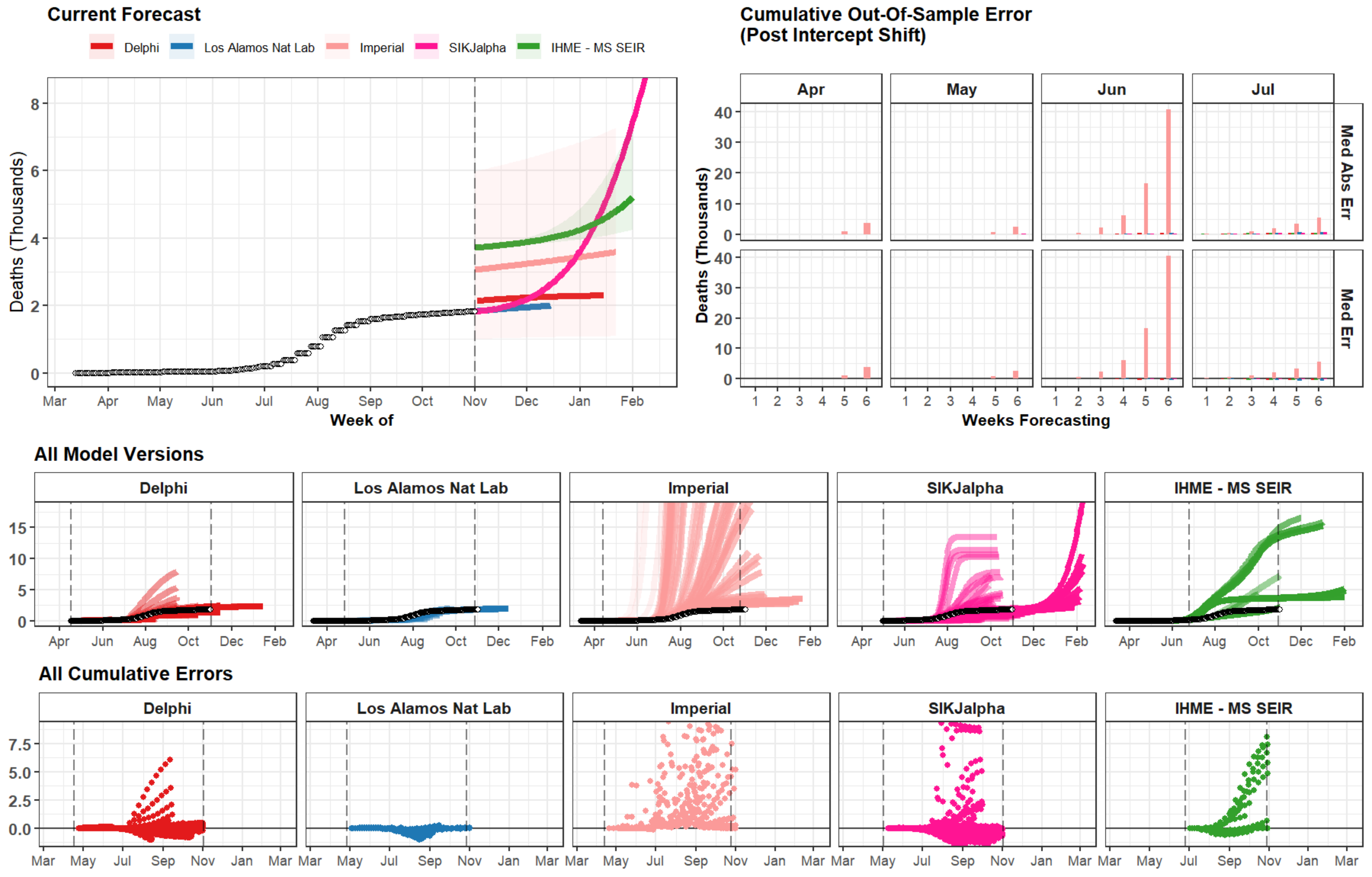

Current Forecast
Delphi
Los Alamos Nat Lab
Imperial
SIKJalpha
IHME - MS SEIR
Deaths (Thousands)
0
2
4
6
8
Mar
Apr
May
Jun
Jul
Aug
Sep
Oct
Nov
Dec
Jan
Feb
Week of
Cumulative Out-Of-Sample Error
(Post Intercept Shift)
Apr
May
Jun
Jul
Med Abs Err
Med Err
0
10
20
30
40
1
2
3
4
5
6
Weeks Forecasting
All Model Versions
Delphi
Los Alamos Nat Lab
Imperial
SIKJalpha
IHME - MS SEIR
0
5
10
15
Apr
Jun
Aug
Oct
Dec
Feb
All Cumulative Errors
Delphi
Los Alamos Nat Lab
Imperial
SIKJalpha
IHME - MS SEIR
0.0
2.5
5.0
7.5
Mar
May
Jul
Sep
Nov
Jan
Mar

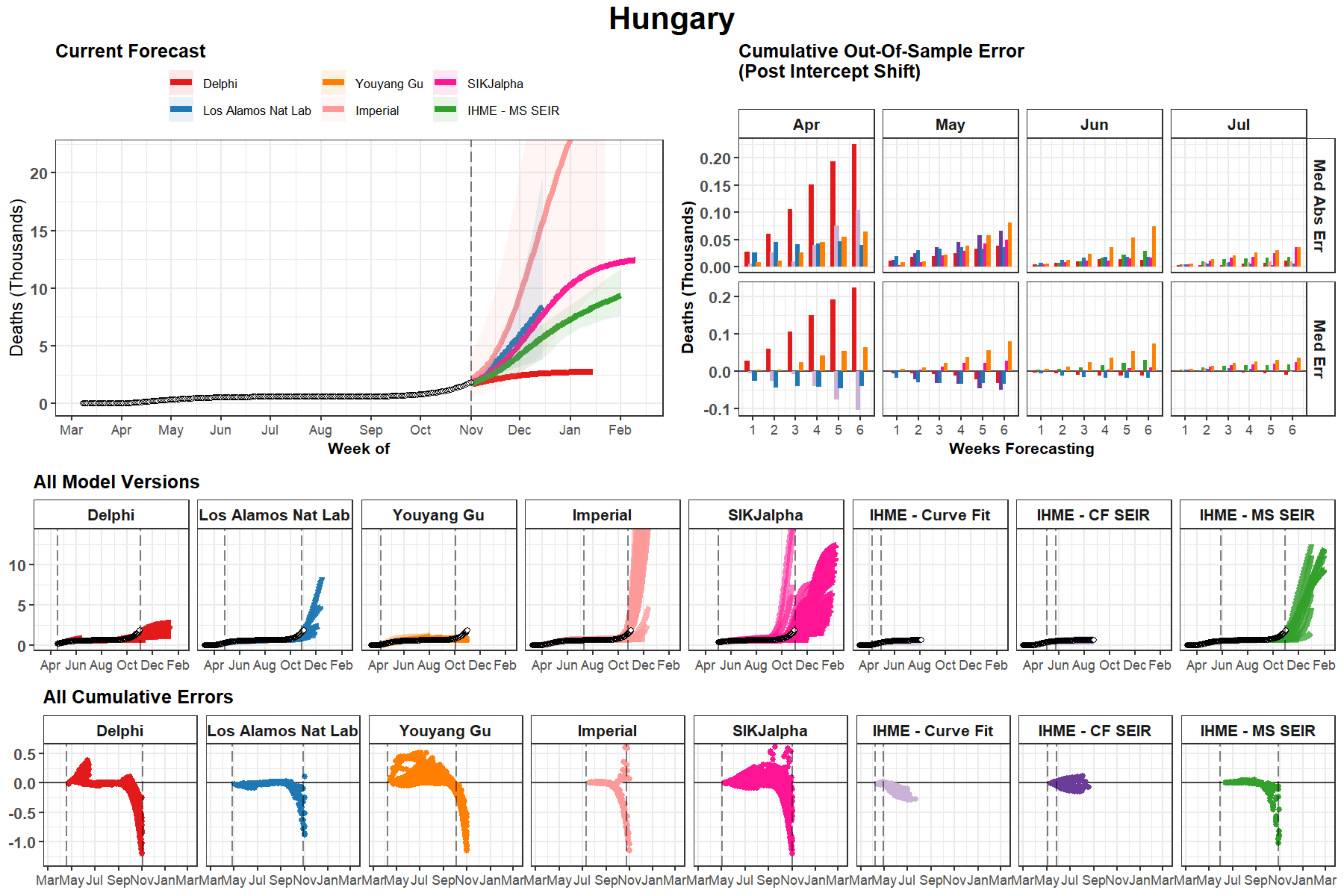
Hungary
Current Forecast
Delphi
Los Alamos Nat Lab
Youyang Gu
Imperial
SIKJalpha
IHME - MS SEIR
Deaths (Thousands)
Week of
Mar Apr May Jun Jul Aug Sep Oct Nov Dec Jan Feb
Cumulative Out-Of-Sample Error
(Post Intercept Shift)
Apr May Jun Jul
Med Abs Err
Med Err
Weeks Forecasting
All Model Versions
Delphi
Los Alamos Nat Lab
Youyang Gu
Imperial
SIKJalpha
IHME - Curve Fit
IHME - CF SEIR
IHME - MS SEIR
All Cumulative Errors

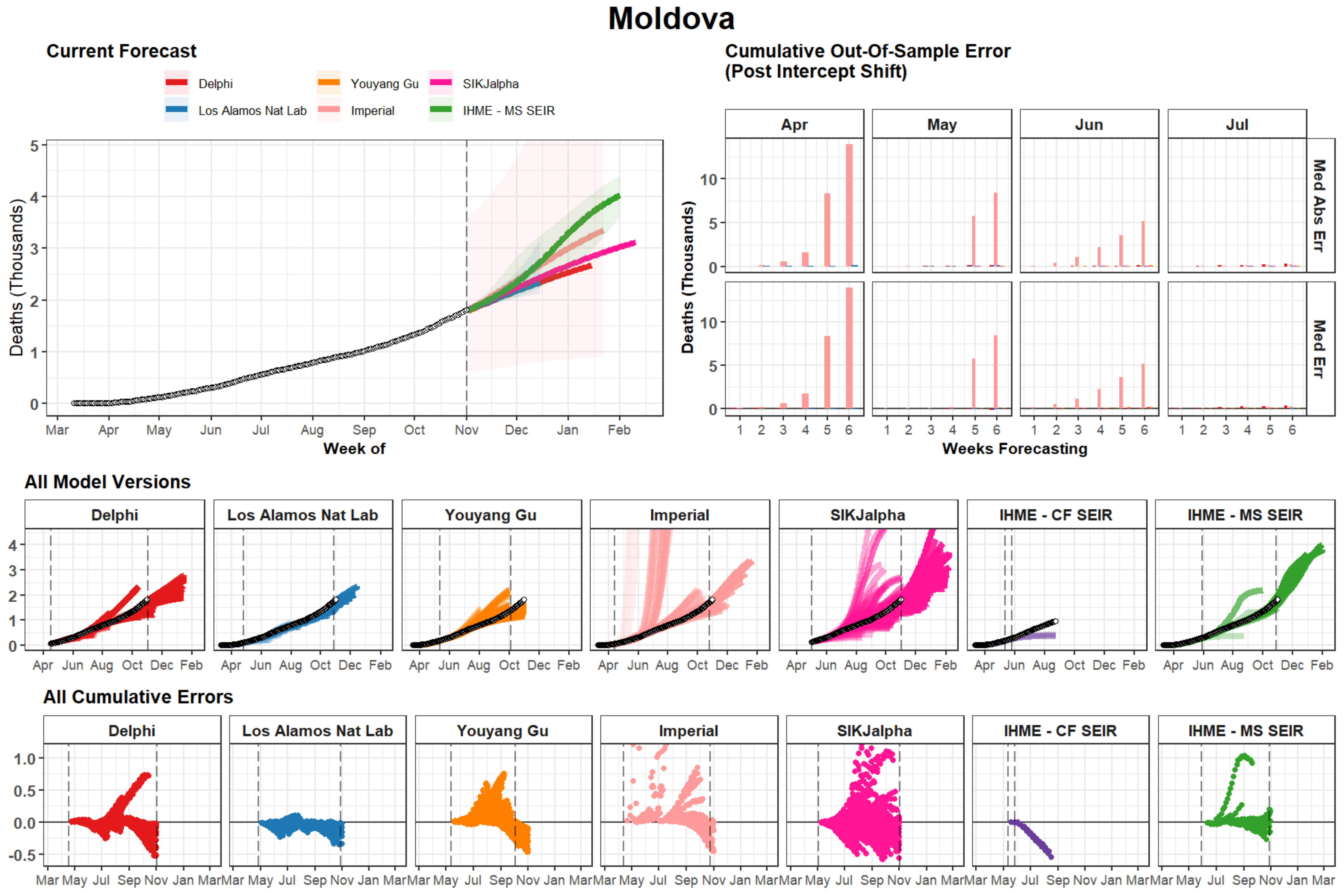
Moldova
Current Forecast
Delphi
Los Alamos Nat Lab
Youyang Gu
Imperial
SIKJalpha
IHME - MS SEIR
Deaths (Thousands)
Week of
Mar
Apr
May
Jun
Jul
Aug
Sep
Oct
Nov
Dec
Jan
Feb
Cumulative Out-Of-Sample Error
(Post Intercept Shift)
Med Abs Err
Med Err
Weeks Forecasting
All Model Versions
IHME - CF SEIR
All Cumulative Errors

### Nevada

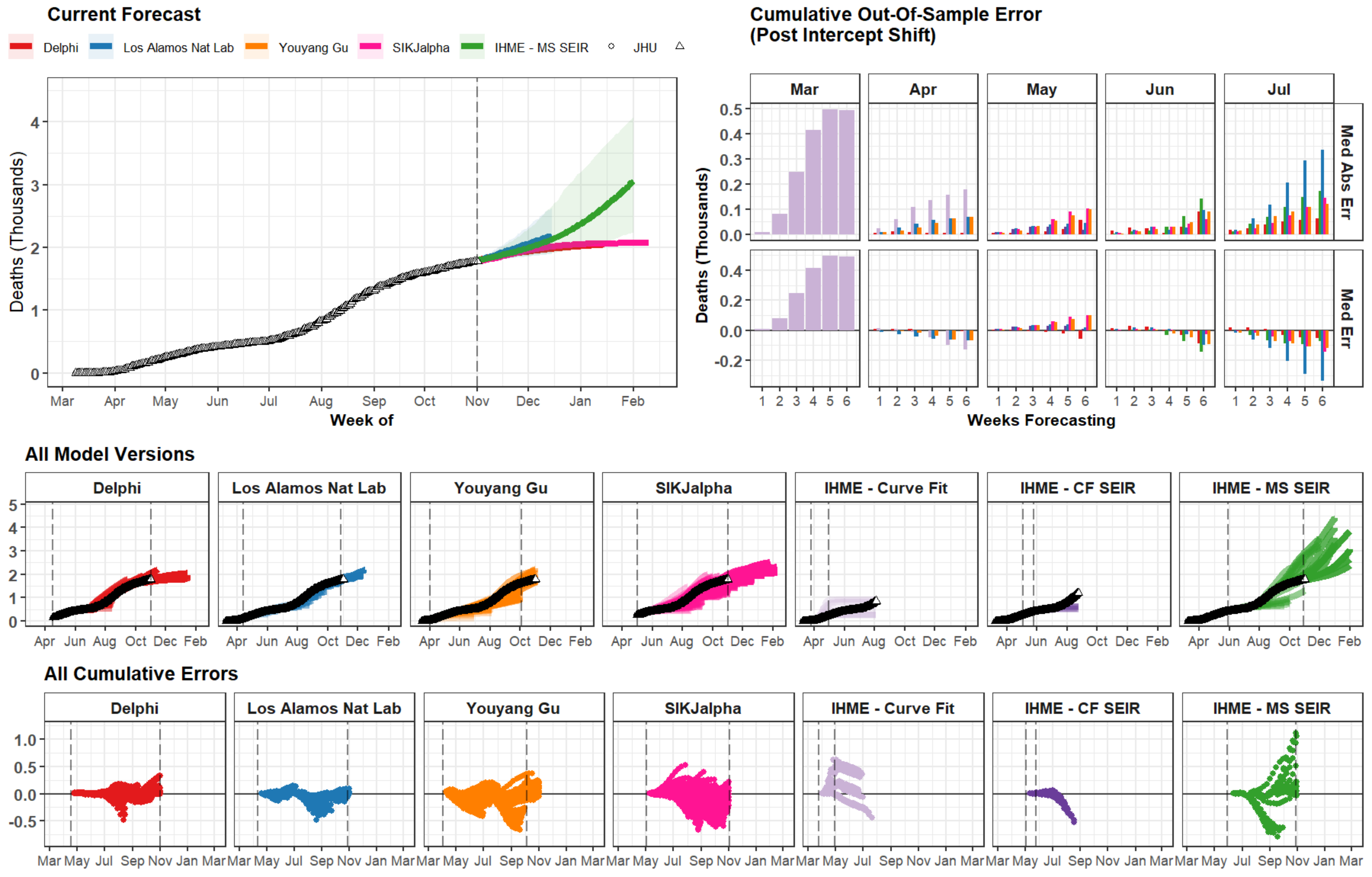

Current Forecast
Delphi
Los Alamos Nat Lab
Youyang Gu
SIKJalpha
IHME - MS SEIR
JHU
Deaths (Thousands)
Week of
Mar
Apr
May
Jun
Jul
Aug
Sep
Oct
Nov
Dec
Jan
Feb
Cumulative Out-Of-Sample Error
(Post Intercept Shift)
Med Abs Err
Med Err
Weeks Forecasting
All Model Versions
Delphi
Los Alamos Nat Lab
Youyang Gu
SIKJalpha
IHME - Curve Fit
IHME - CF SEIR
IHME - MS SEIR
All Cumulative Errors

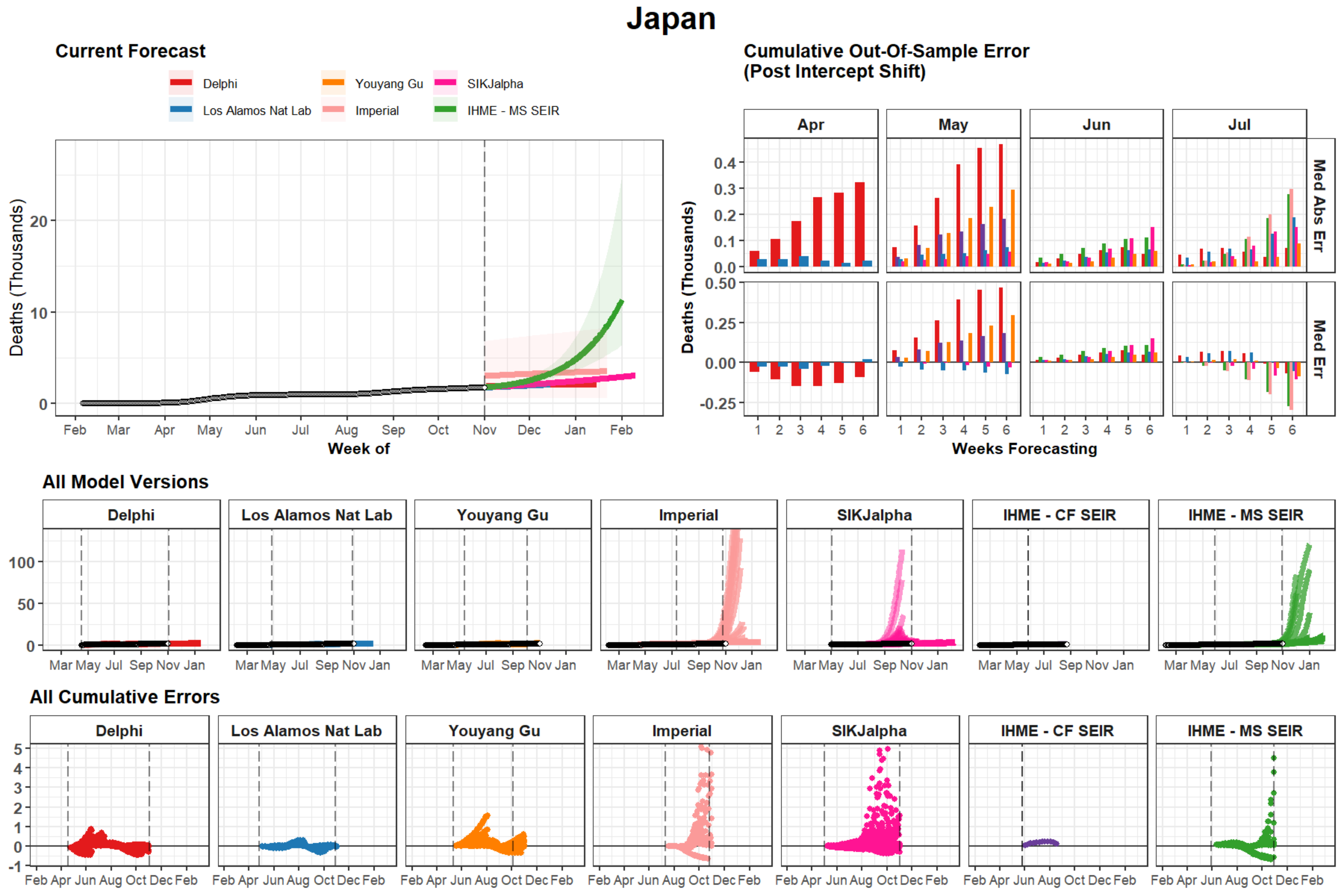

Japan
Current Forecast
Delphi
Los Alamos Nat Lab
Youyang Gu
Imperial
SIKJalpha
IHME - MS SEIR
Deaths (Thousands)
0
10
20
Feb Mar Apr May Jun Jul Aug Sep Oct Nov Dec Jan Feb
Week of
Cumulative Out-Of-Sample Error
(Post Intercept Shift)
Apr
May
Jun
Jul
Med Abs Err
Med Err
0.0
0.1
0.2
0.3
0.4
0.50
0.25
0.00
-0.25
1 2 3 4 5 6
Weeks Forecasting
All Model Versions
Delphi
Los Alamos Nat Lab
Youyang Gu
Imperial
SIKJalpha
IHME - CF SEIR
IHME - MS SEIR
0
50
100
Mar May Jul Sep Nov Jan
All Cumulative Errors
-1
0
1
2
3
4
5
Feb Apr Jun Aug Oct Dec Feb

### Iowa

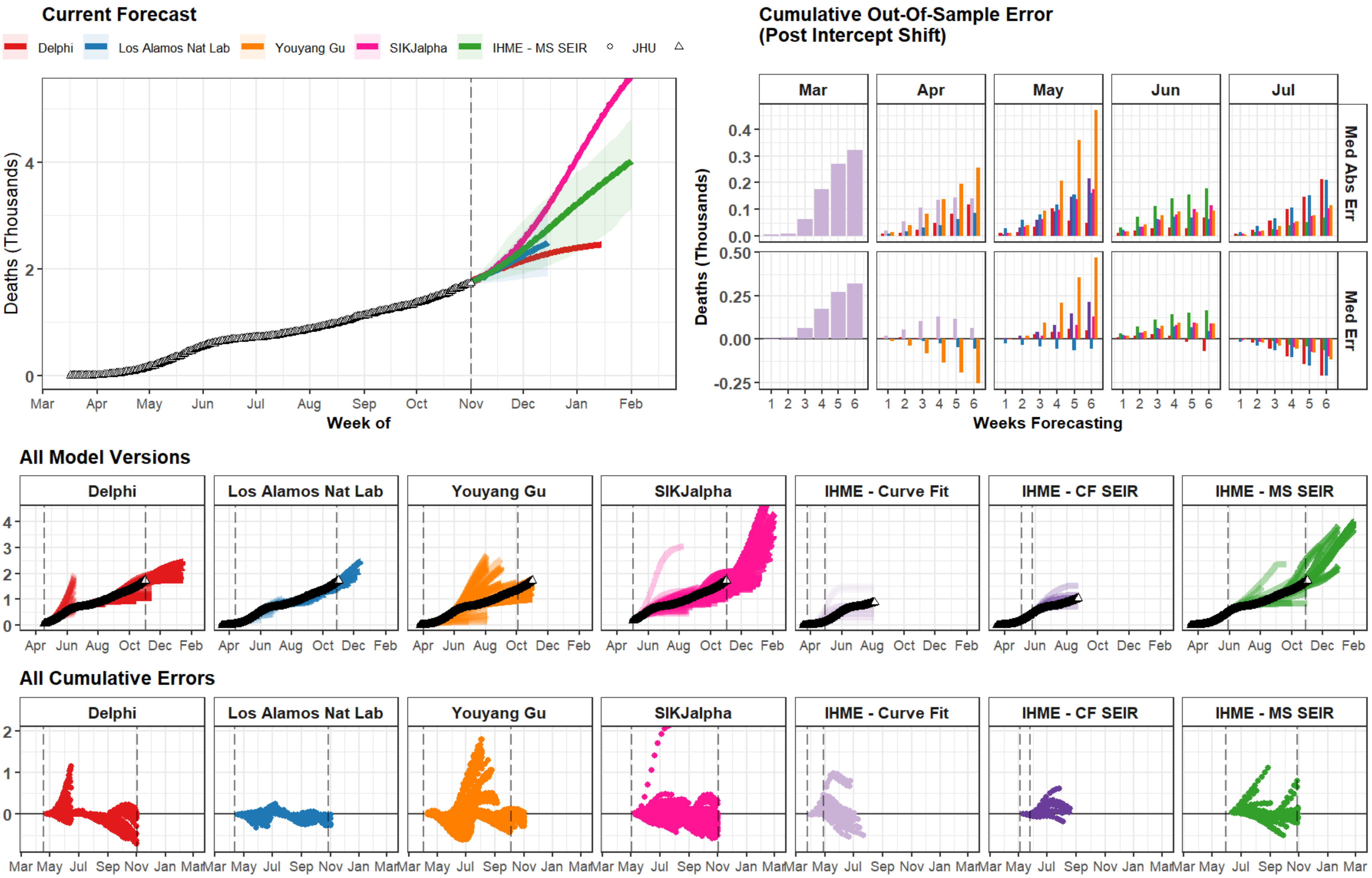
Current Forecast
Delphi
Los Alamos Nat Lab
Youyang Gu
SIKJalpha
IHME - MS SEIR
JHU
Deaths (Thousands)
0
2
4
Mar
Apr
May
Jun
Jul
Aug
Sep
Oct
Nov
Dec
Jan
Feb
Week of
Cumulative Out-Of-Sample Error
(Post Intercept Shift)
Mar
Apr
May
Jun
Jul
Med Abs Err
Med Err
Deaths (Thousands)
0.0
0.1
0.2
0.3
0.4
-0.25
0.00
0.25
0.50
1 2 3 4 5 6
Weeks Forecasting
All Model Versions
Delphi
Los Alamos Nat Lab
Youyang Gu
SIKJalpha
IHME - Curve Fit
IHME - CF SEIR
IHME - MS SEIR
0
1
2
3
4
Apr Jun Aug Oct Dec Feb
All Cumulative Errors
Delphi
Los Alamos Nat Lab
Youyang Gu
SIKJalpha
IHME - Curve Fit
IHME - CF SEIR
IHME - MS SEIR
0
1
2
Mar May Jul Sep Nov Jan Mar

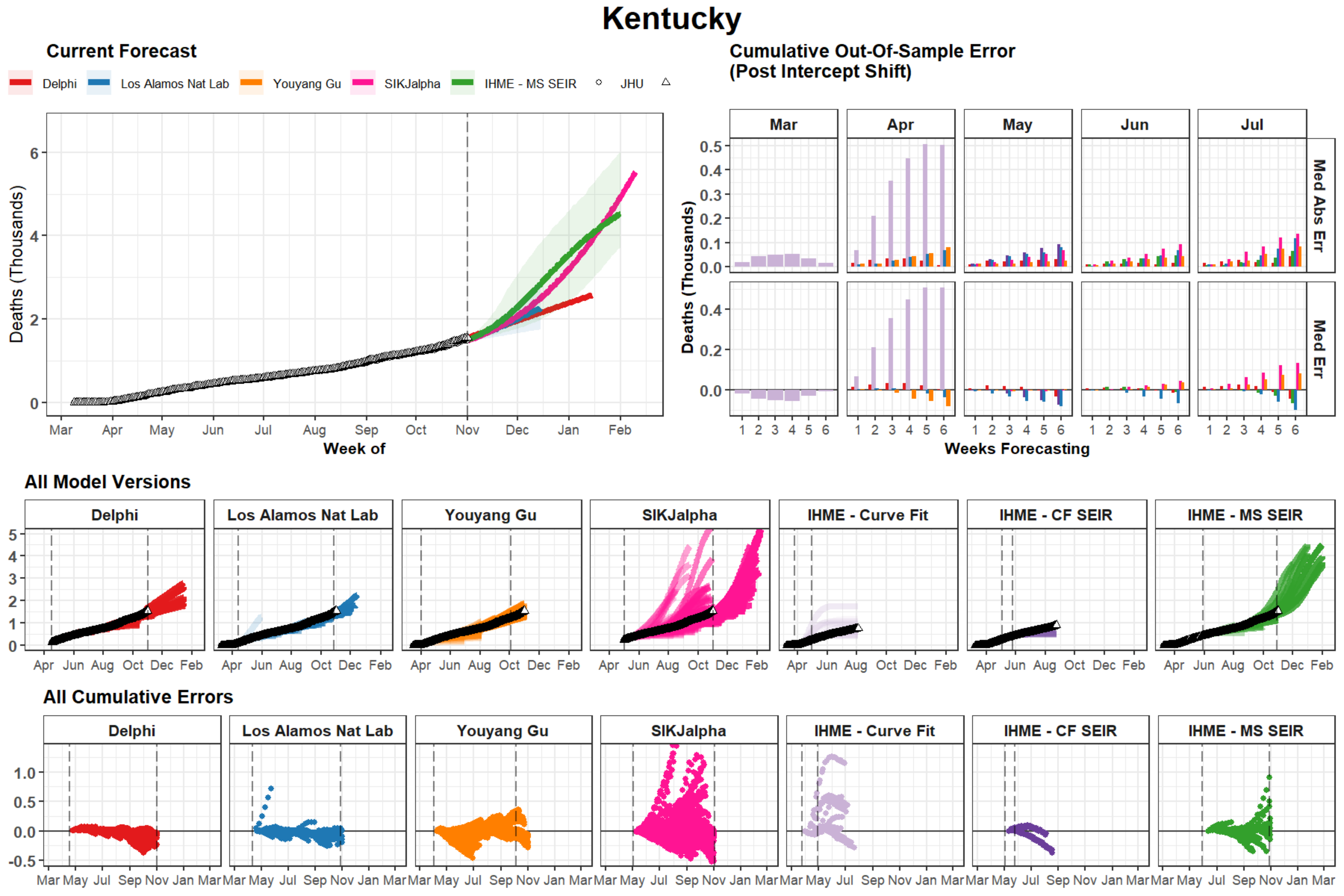
Kentucky
Current Forecast
Delphi
Los Alamos Nat Lab
Youyang Gu
SIKJalpha
IHME - MS SEIR
JHU
Deaths (Thousands)
Week of
Mar Apr May Jun Jul Aug Sep Oct Nov Dec Jan Feb
Cumulative Out-Of-Sample Error
(Post Intercept Shift)
Mar Apr May Jun Jul
Med Abs Err
Med Err
Weeks Forecasting
All Model Versions
Delphi
Los Alamos Nat Lab
Youyang Gu
SIKJalpha
IHME - Curve Fit
IHME - CF SEIR
IHME - MS SEIR
All Cumulative Errors
Delphi
Los Alamos Nat Lab
Youyang Gu
SIKJalpha
IHME - Curve Fit
IHME - CF SEIR
IHME - MS SEIR

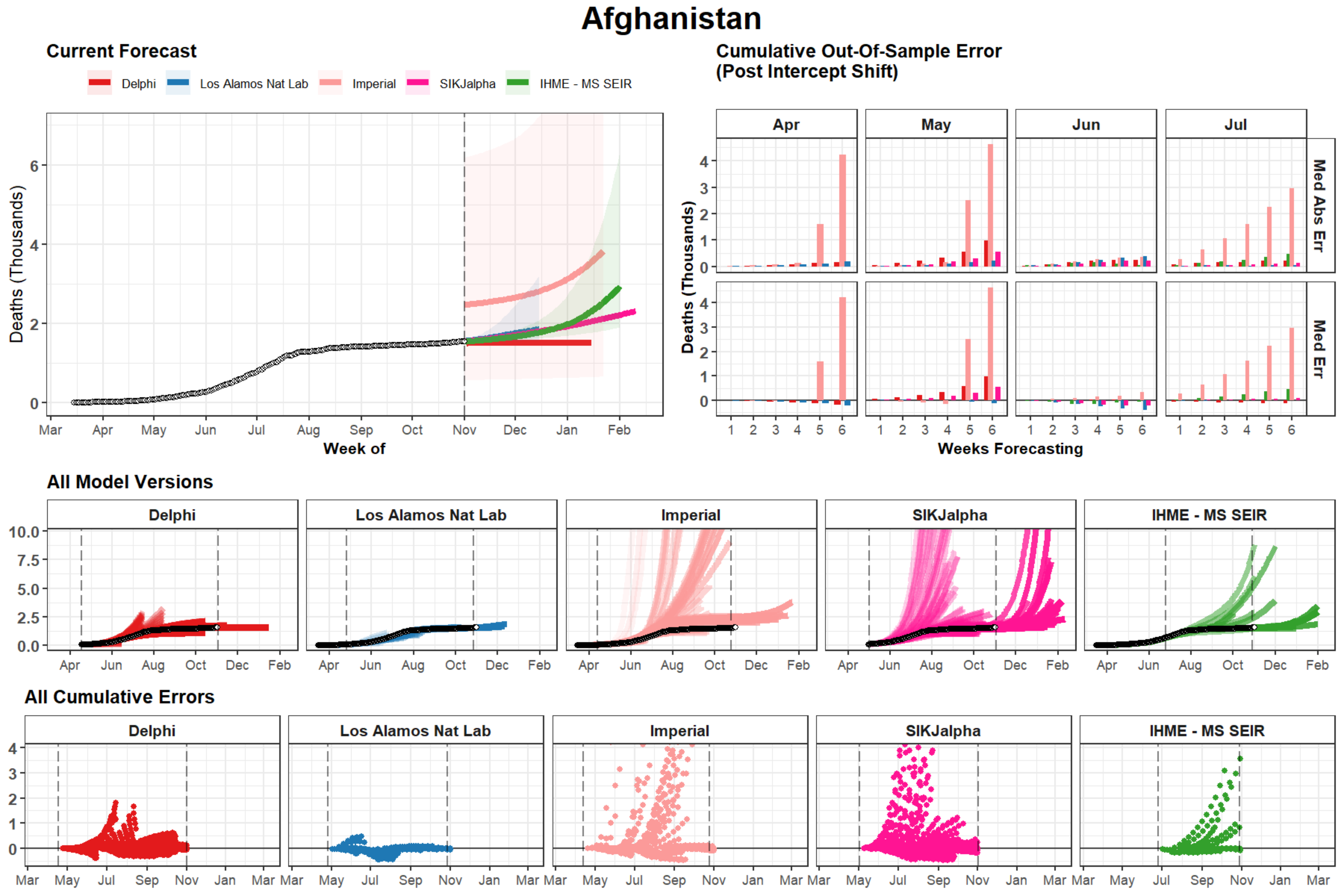

Afghanistan
Current Forecast
Delphi
Los Alamos Nat Lab
Imperial
SIKJalpha
IHME - MS SEIR
Deaths (Thousands)
Week of
Mar
Apr
May
Jun
Jul
Aug
Sep
Oct
Nov
Dec
Jan
Feb
Cumulative Out-Of-Sample Error
(Post Intercept Shift)
Apr
May
Jun
Jul
Med Abs Err
Med Err
Weeks Forecasting
All Model Versions
All Cumulative Errors

### Ethiopia

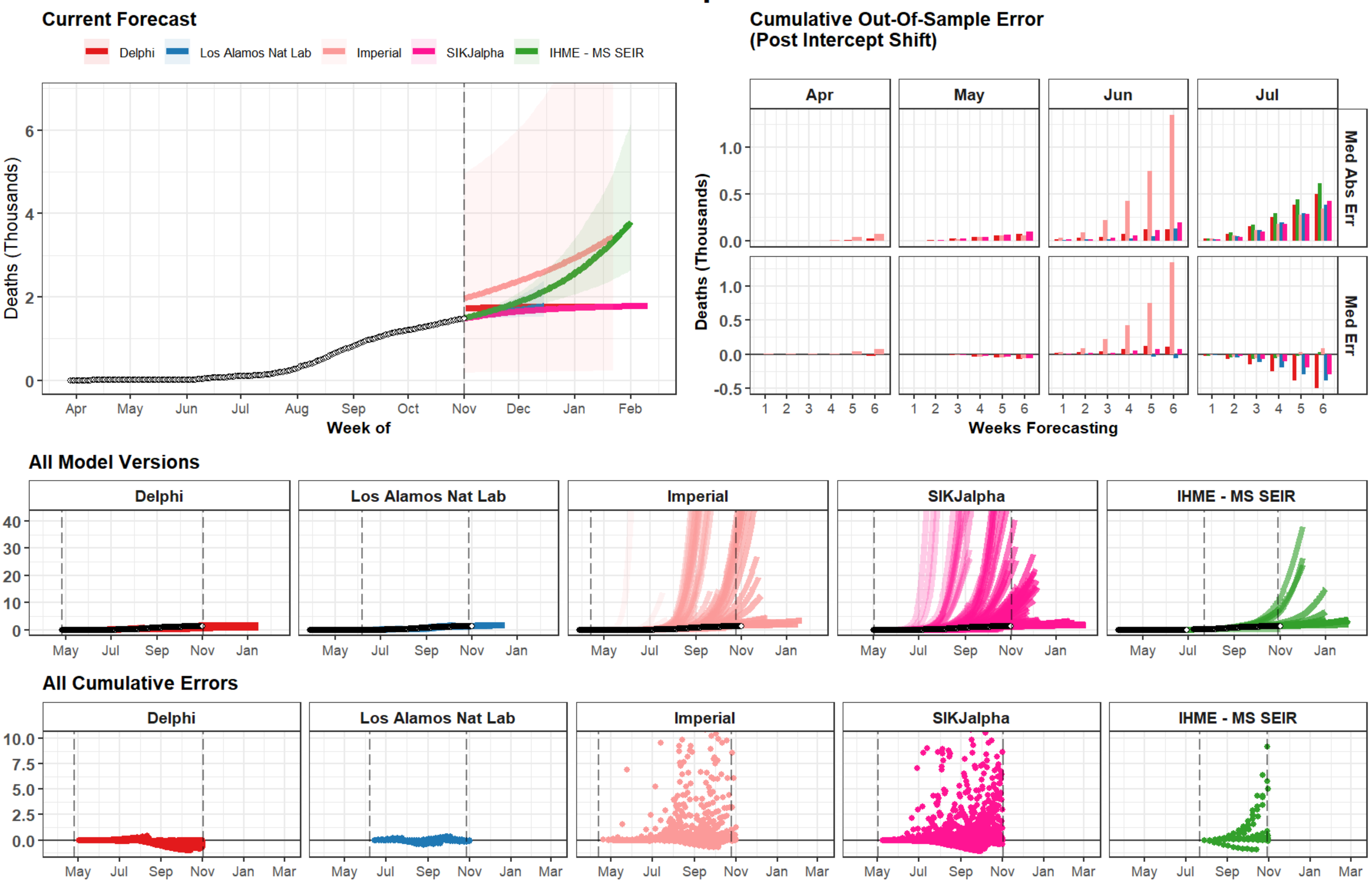

Current Forecast
Delphi
Los Alamos Nat Lab
Imperial
SIKJalpha
IHME - MS SEIR
Deaths (Thousands)
Week of
Apr May Jun Jul Aug Sep Oct Nov Dec Jan Feb
Cumulative Out-Of-Sample Error
(Post Intercept Shift)
Apr
May
Jun
Jul
Med Abs Err
Med Err
Weeks Forecasting
All Model Versions
All Cumulative Errors
May Jul Sep Nov Jan Mar

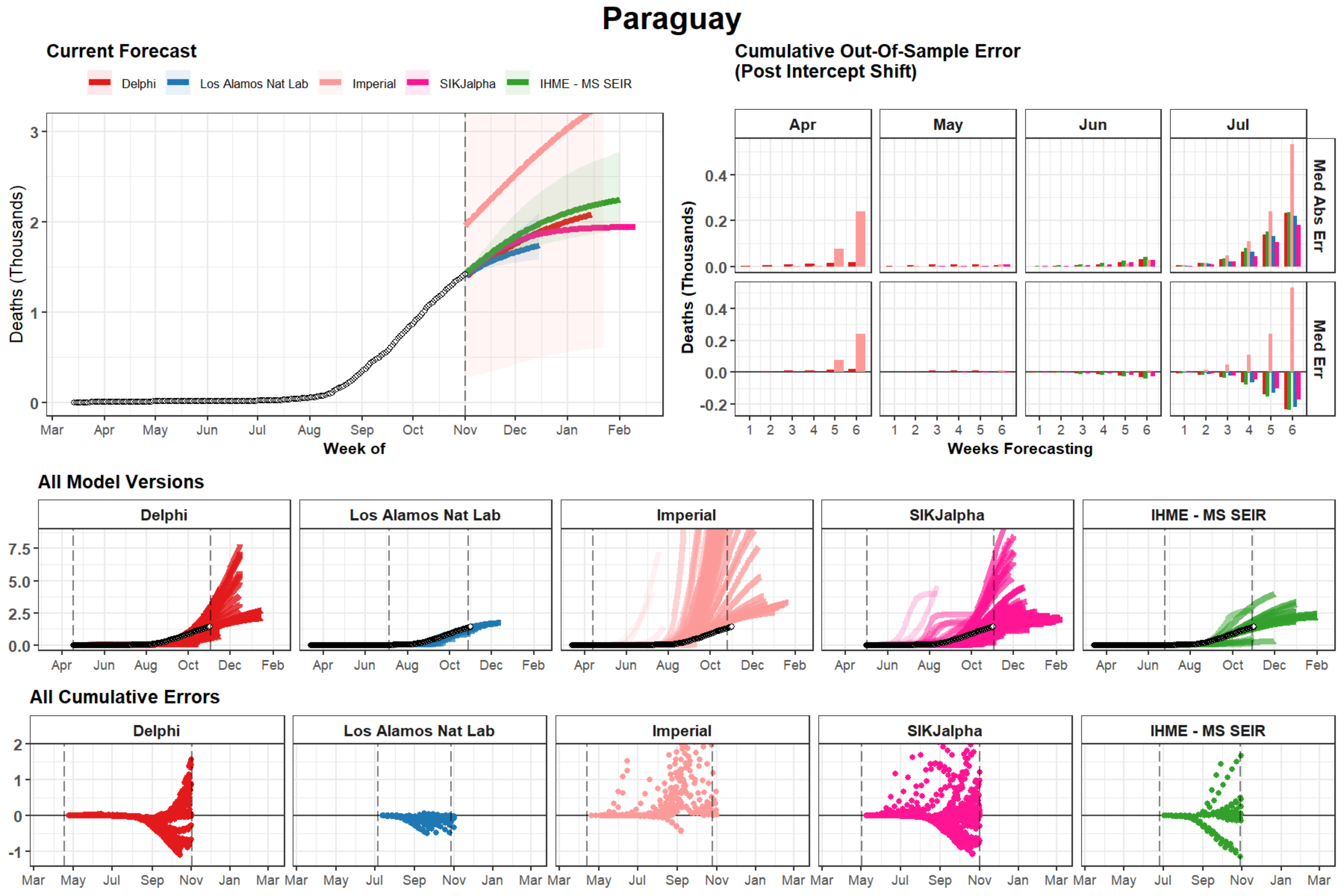

Paraguay
Current Forecast
Delphi
Los Alamos Nat Lab
Imperial
SIKJalpha
IHME - MS SEIR
Deaths (Thousands)
Week of
Mar Apr May Jun Jul Aug Sep Oct Nov Dec Jan Feb
Cumulative Out-Of-Sample Error
(Post Intercept Shift)
Apr May Jun Jul
Med Abs Err
Med Err
Weeks Forecasting
All Model Versions
All Cumulative Errors

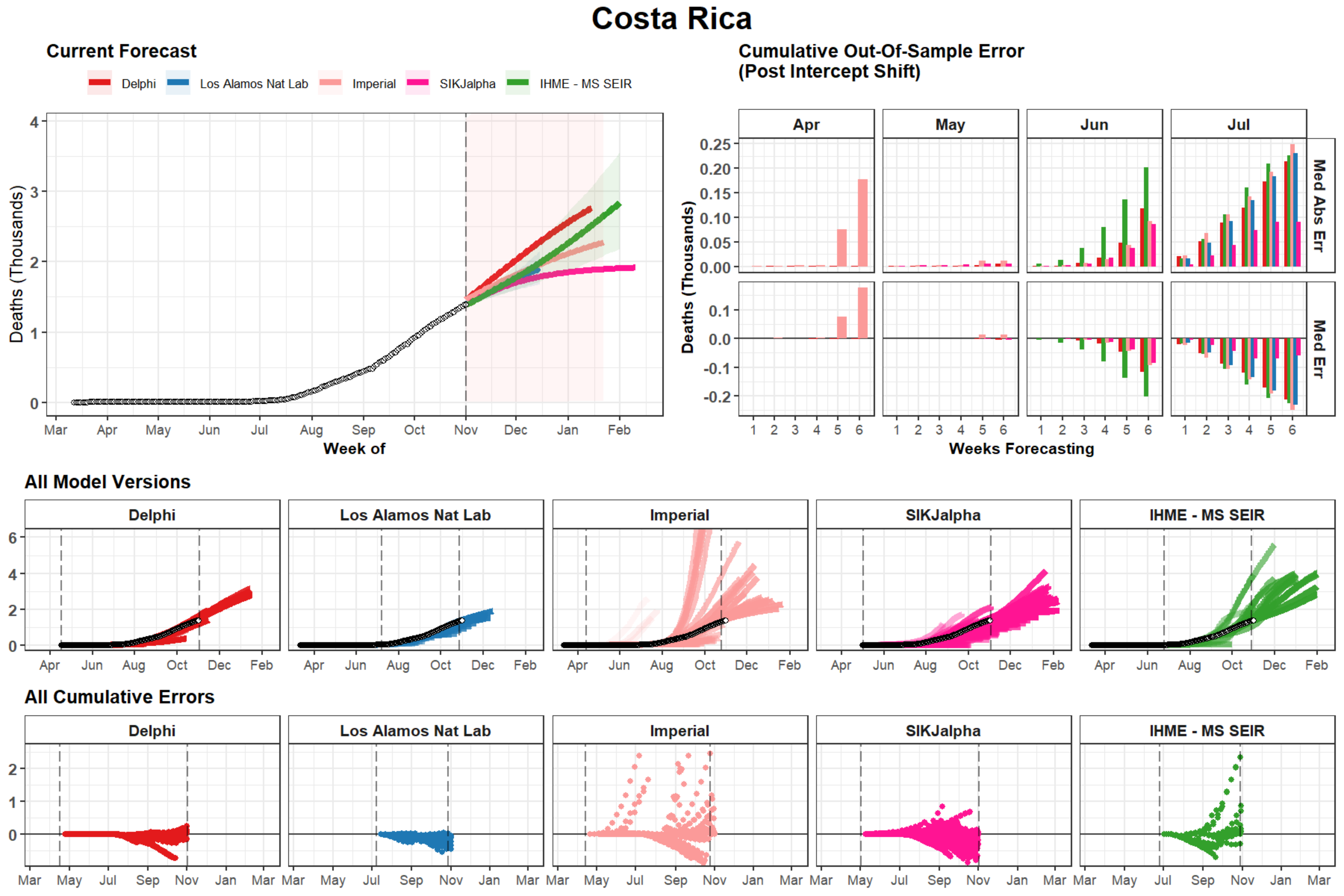

Costa Rica
Current Forecast
Delphi
Los Alamos Nat Lab
Imperial
SIKJalpha
IHME - MS SEIR
Deaths (Thousands)
Week of
Mar Apr May Jun Jul Aug Sep Oct Nov Dec Jan Feb
Cumulative Out-Of-Sample Error
(Post Intercept Shift)
Apr May Jun Jul
Med Abs Err
Med Err
Weeks Forecasting
All Model Versions
All Cumulative Errors

### Armenia

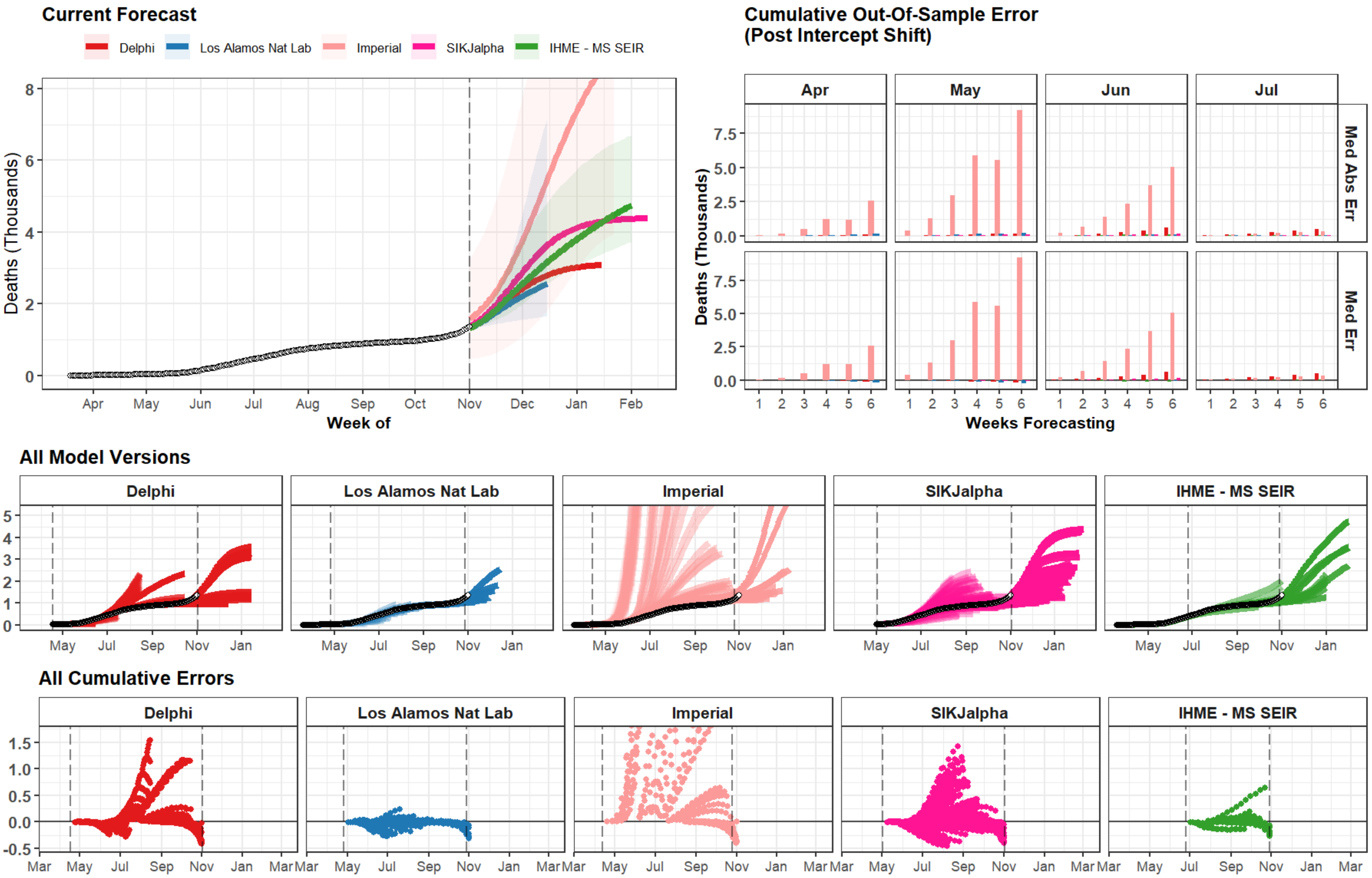

Current Forecast
Delphi
Los Alamos Nat Lab
Imperial
SIKJalpha
IHME - MS SEIR
Deaths (Thousands)
0
2
4
6
8
Apr
May
Jun
Jul
Aug
Sep
Oct
Nov
Dec
Jan
Feb
Week of
Cumulative Out-Of-Sample Error
(Post Intercept Shift)
Apr
May
Jun
Jul
Med Abs Err
Med Err
Deaths (Thousands)
0.0
2.5
5.0
7.5
1
2
3
4
5
6
Weeks Forecasting
All Model Versions
Delphi
Los Alamos Nat Lab
Imperial
SIKJalpha
IHME - MS SEIR
0
1
2
3
4
5
May
Jul
Sep
Nov
Jan
All Cumulative Errors
Delphi
Los Alamos Nat Lab
Imperial
SIKJalpha
IHME - MS SEIR
-0.5
0.0
0.5
1.0
1.5
Mar
May
Jul
Sep
Nov
Jan
Mar

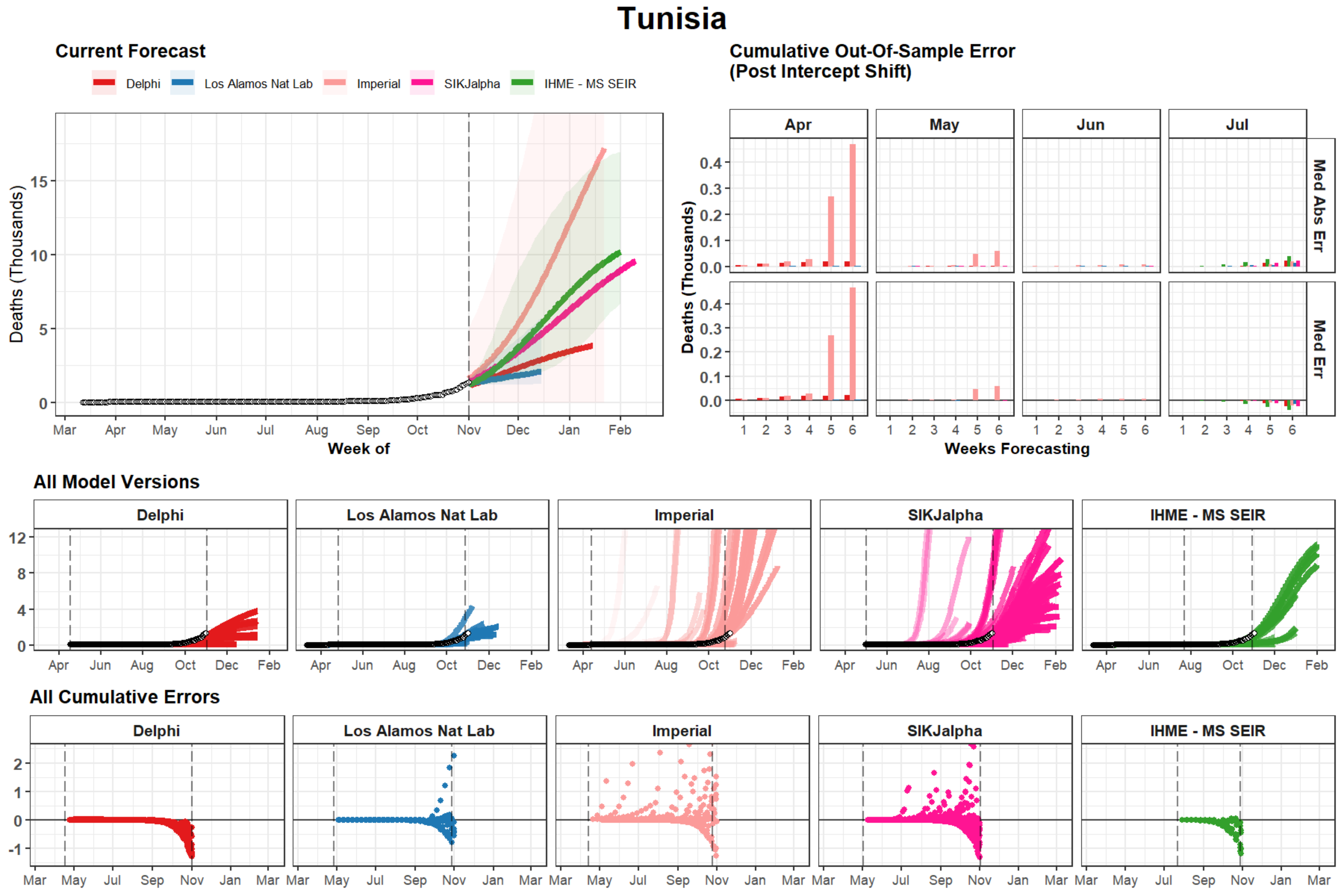
Tunisia
Current Forecast
Delphi
Los Alamos Nat Lab
Imperial
SIKJalpha
IHME - MS SEIR
Deaths (Thousands)
Week of
Cumulative Out-Of-Sample Error
(Post Intercept Shift)
Apr
May
Jun
Jul
Med Abs Err
Med Err
Weeks Forecasting
All Model Versions
All Cumulative Errors

### Oklahoma

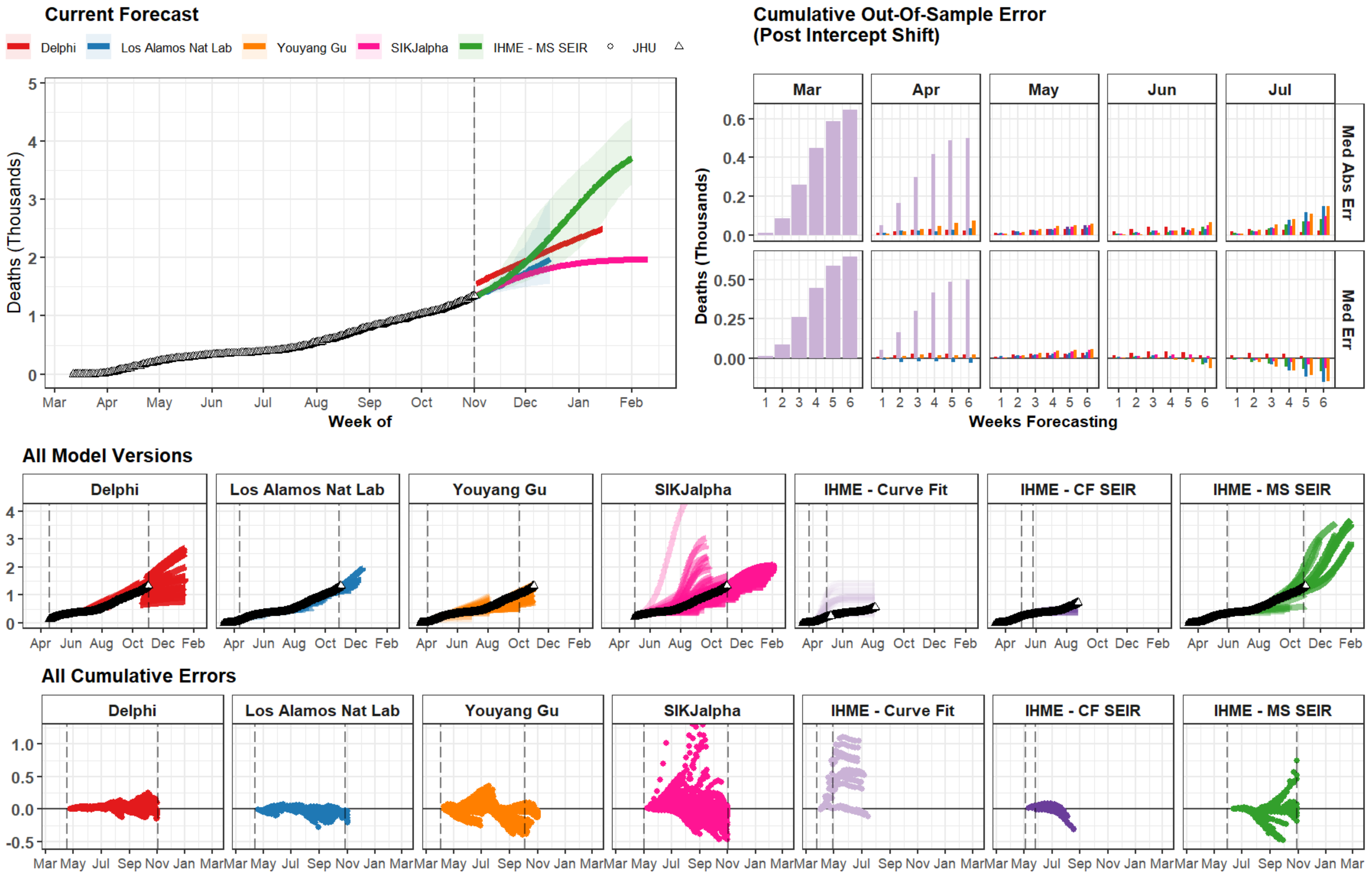

Current Forecast
Delphi
Los Alamos Nat Lab
Youyang Gu
SIKJalpha
IHME - MS SEIR
JHU
Deaths (Thousands)
Week of
Mar Apr May Jun Jul Aug Sep Oct Nov Dec Jan Feb
Cumulative Out-Of-Sample Error (Post Intercept Shift)
Mar Apr May Jun Jul
Med Abs Err
Med Err
Weeks Forecasting
All Model Versions
Delphi
Los Alamos Nat Lab
Youyang Gu
SIKJalpha
IHME - Curve Fit
IHME - CF SEIR
IHME - MS SEIR
All Cumulative Errors

### Bulgaria

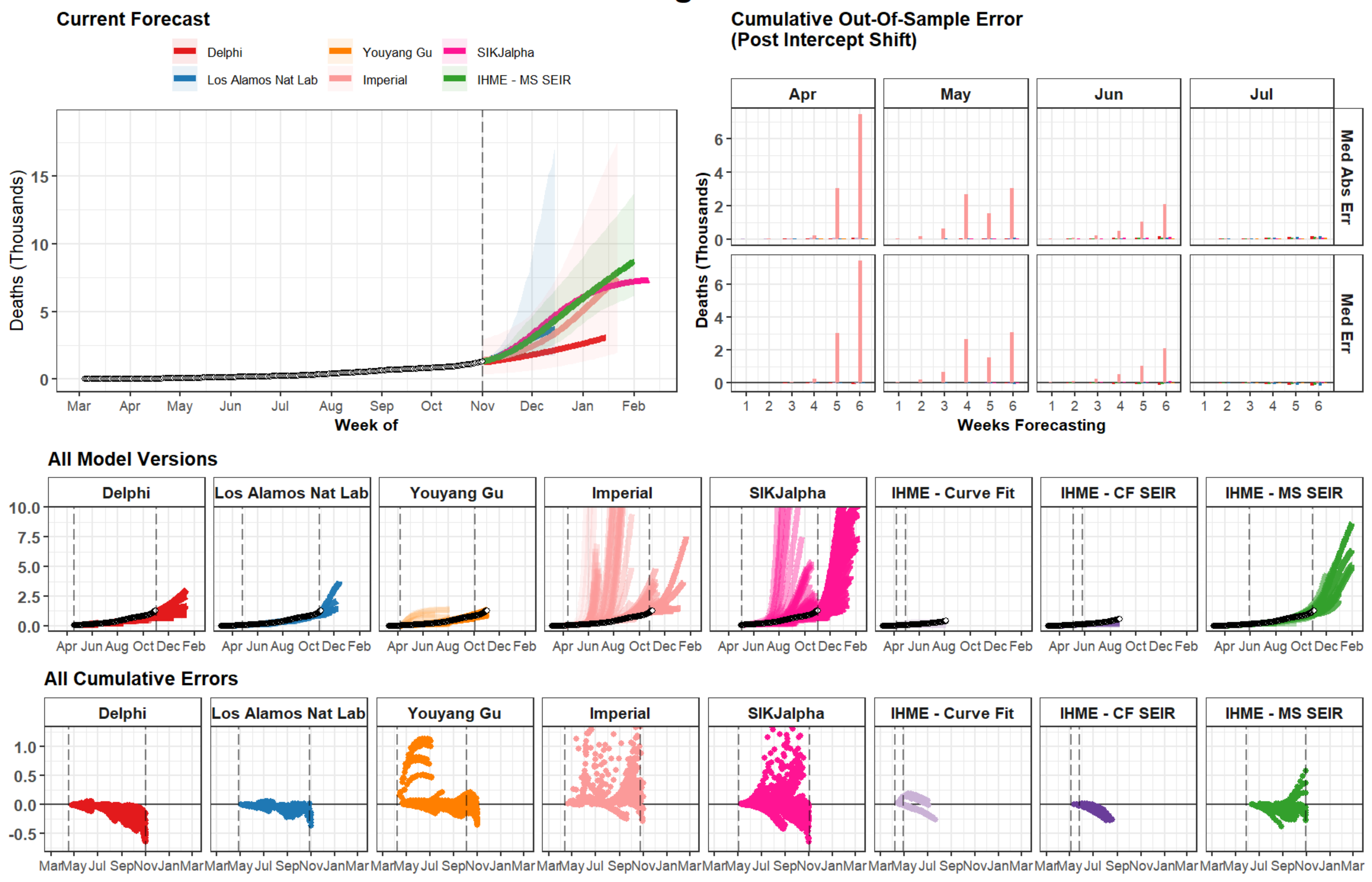

Current Forecast
Delphi
Los Alamos Nat Lab
Youyang Gu
Imperial
SIKJalpha
IHME - MS SEIR
Deaths (Thousands)
Week of
Mar Apr May Jun Jul Aug Sep Oct Nov Dec Jan Feb
Cumulative Out-Of-Sample Error
(Post Intercept Shift)
Apr
May
Jun
Jul
Med Abs Err
Med Err
Weeks Forecasting
All Model Versions
Delphi
Los Alamos Nat Lab
Youyang Gu
Imperial
SIKJalpha
IHME - Curve Fit
IHME - CF SEIR
IHME - MS SEIR
Apr Jun Aug Oct Dec Feb
All Cumulative Errors
Mar May Jul Sep Nov Jan Mar

### Myanmar

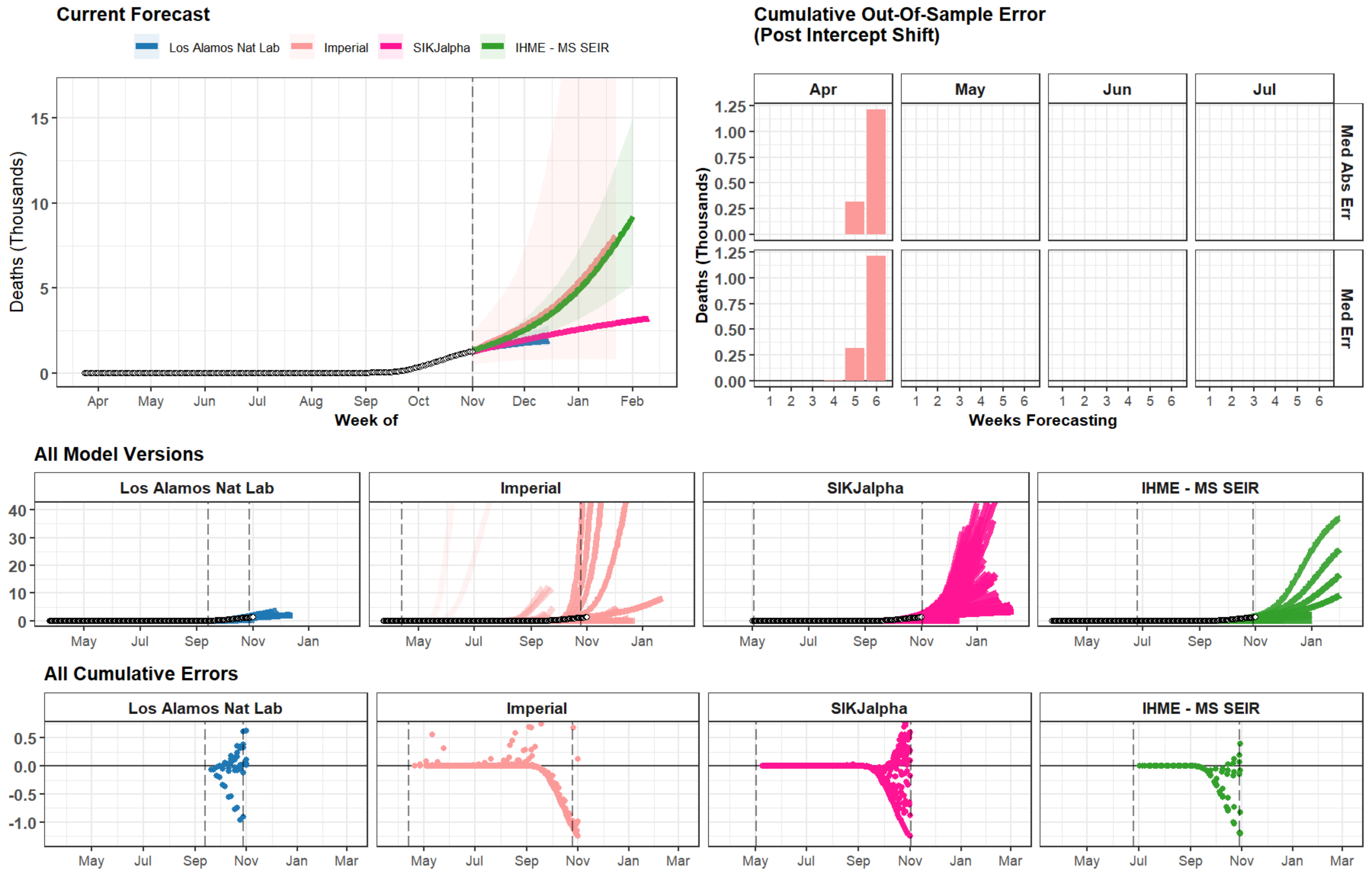
Current Forecast
Los Alamos Nat Lab
Imperial
SIKJalpha
IHME - MS SEIR
Deaths (Thousands)
0
5
10
15
Apr
May
Jun
Jul
Aug
Sep
Oct
Nov
Dec
Jan
Feb
Week of
Cumulative Out-Of-Sample Error
(Post Intercept Shift)
Apr
May
Jun
Jul
Med Abs Err
Med Err
Deaths (Thousands)
0.00
0.25
0.50
0.75
1.00
1.25
1 2 3 4 5 6
Weeks Forecasting
All Model Versions
Los Alamos Nat Lab
Imperial
SIKJalpha
IHME - MS SEIR
0
10
20
30
40
May
Jul
Sep
Nov
Jan
All Cumulative Errors
Los Alamos Nat Lab
Imperial
SIKJalpha
IHME - MS SEIR
0.5
0.0
-0.5
-1.0
May
Jul
Sep
Nov
Jan
Mar

### Bosnia and Herzegovina

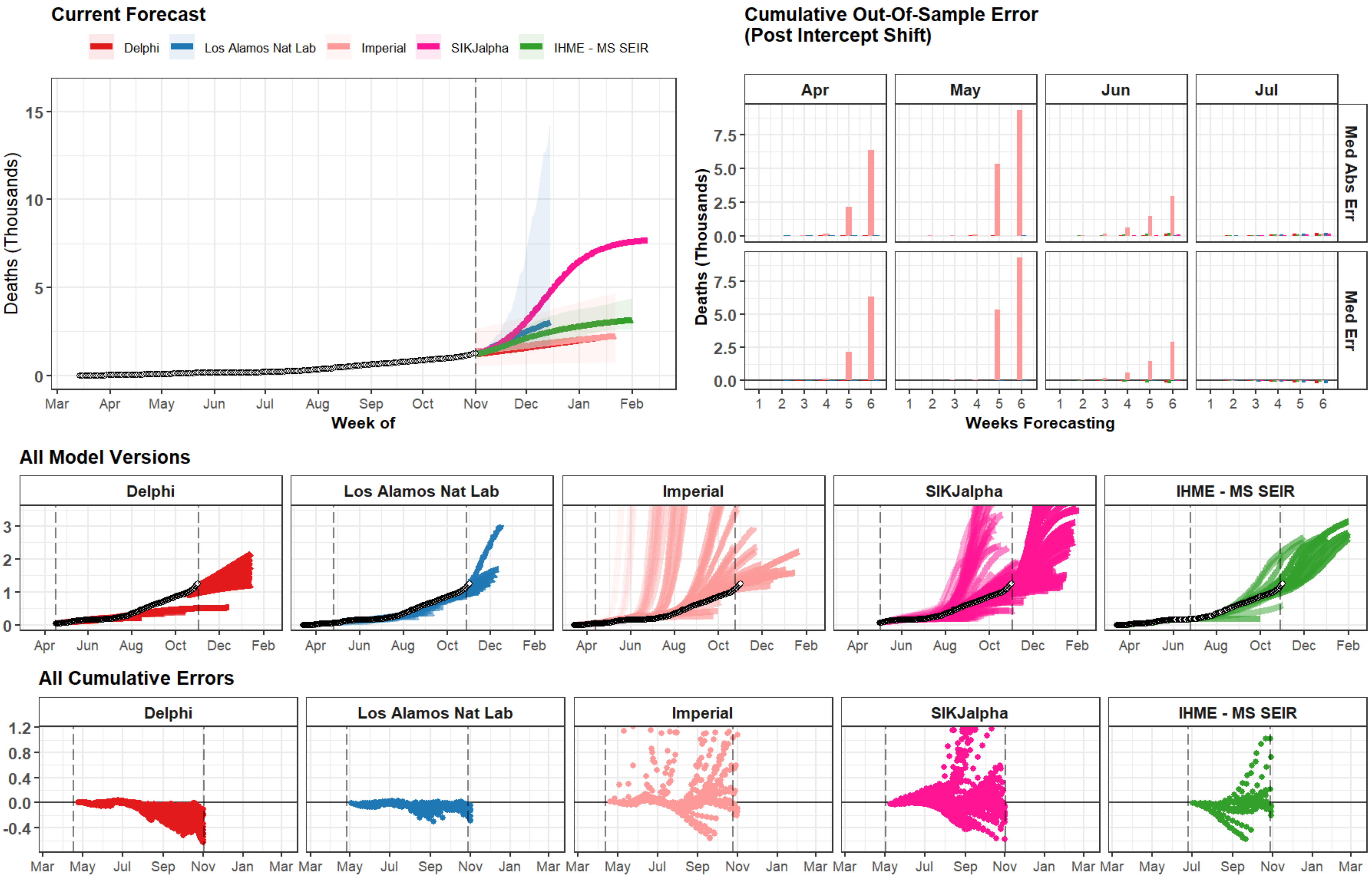

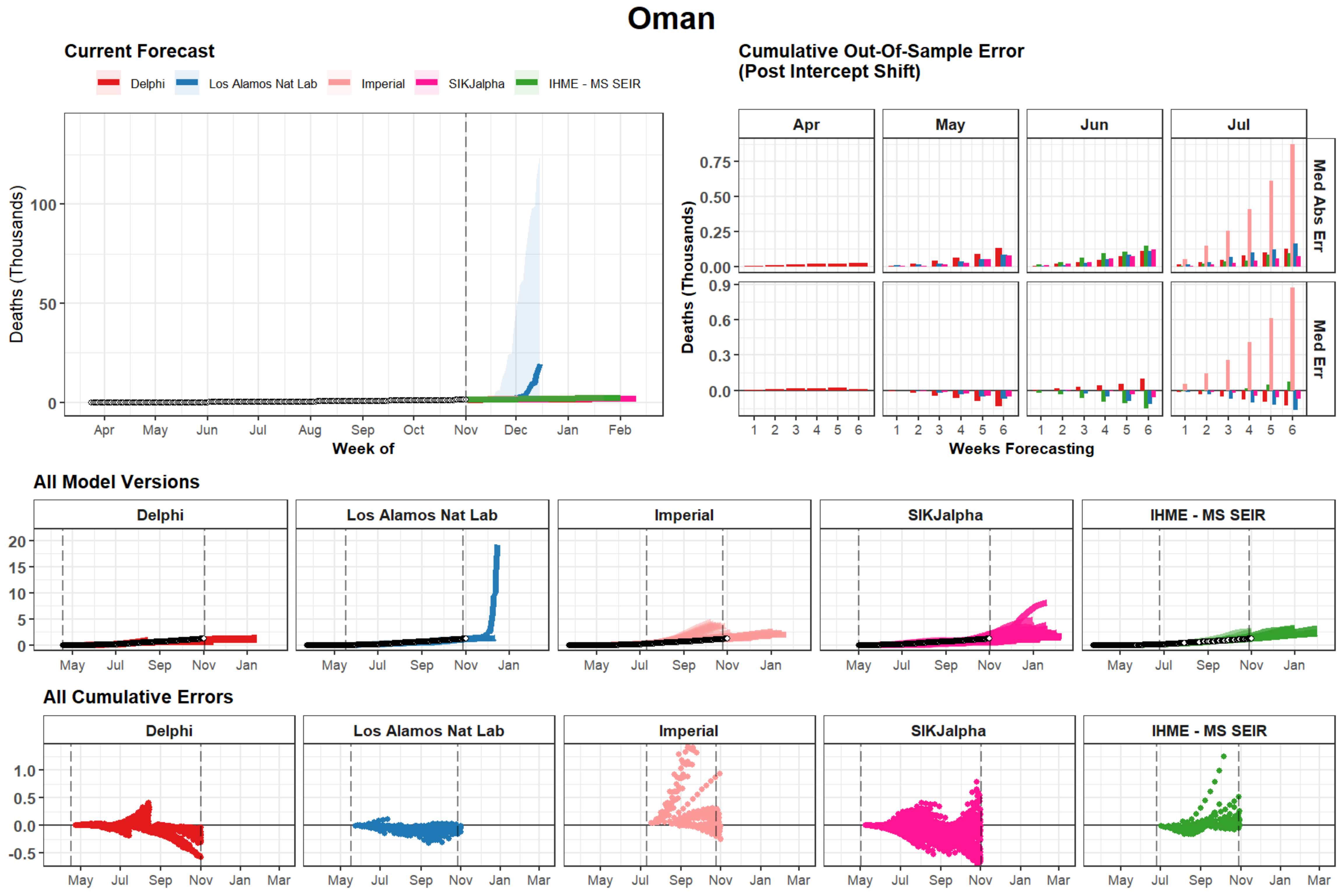

Oman
Current Forecast
Delphi
Los Alamos Nat Lab
Imperial
SIKJalpha
IHME - MS SEIR
Deaths (Thousands)
Week of
Cumulative Out-Of-Sample Error
(Post Intercept Shift)
Apr
May
Jun
Jul
Med Abs Err
Med Err
Weeks Forecasting
All Model Versions
All Cumulative Errors

### Rhode Island

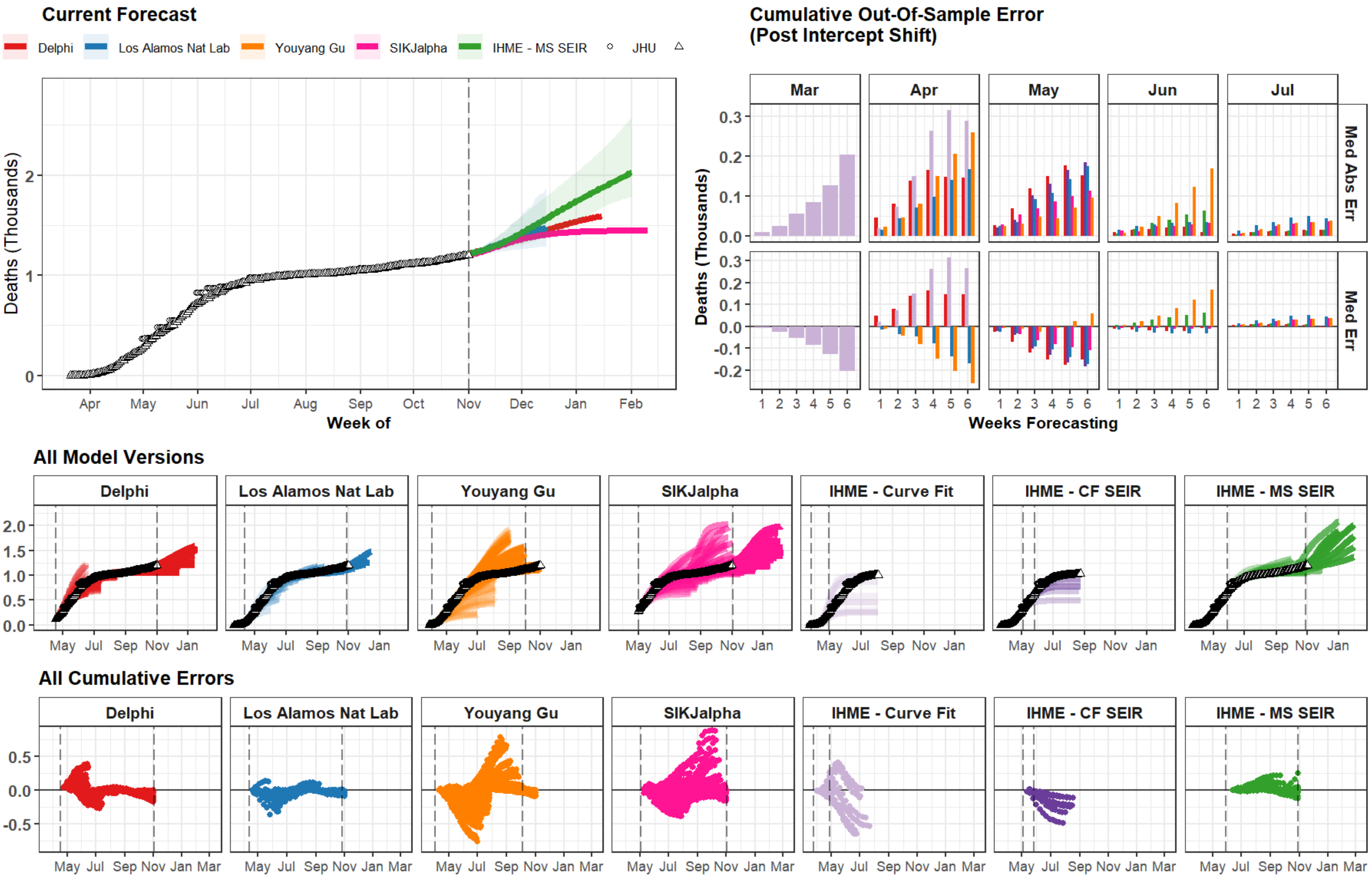

### Nigeria

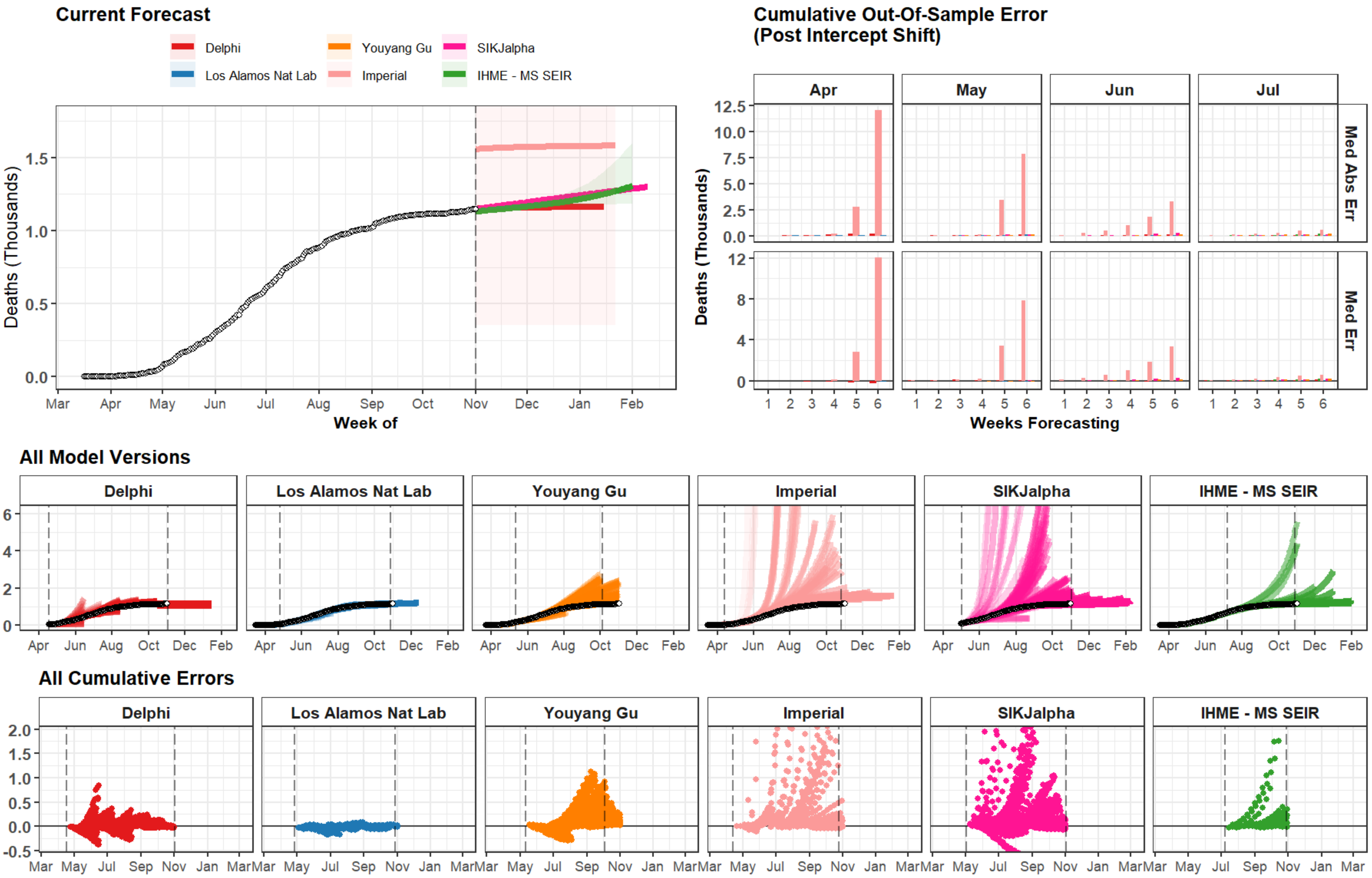

Current Forecast
Delphi
Youyang Gu
SIKJalpha
Los Alamos Nat Lab
Imperial
IHME - MS SEIR
Deaths (Thousands)
Week of
Mar
Apr
May
Jun
Jul
Aug
Sep
Oct
Nov
Dec
Jan
Feb
Cumulative Out-Of-Sample Error
(Post Intercept Shift)
Apr
May
Jun
Jul
Med Abs Err
Med Err
Weeks Forecasting
All Model Versions
All Cumulative Errors

### Kyrgyzstan

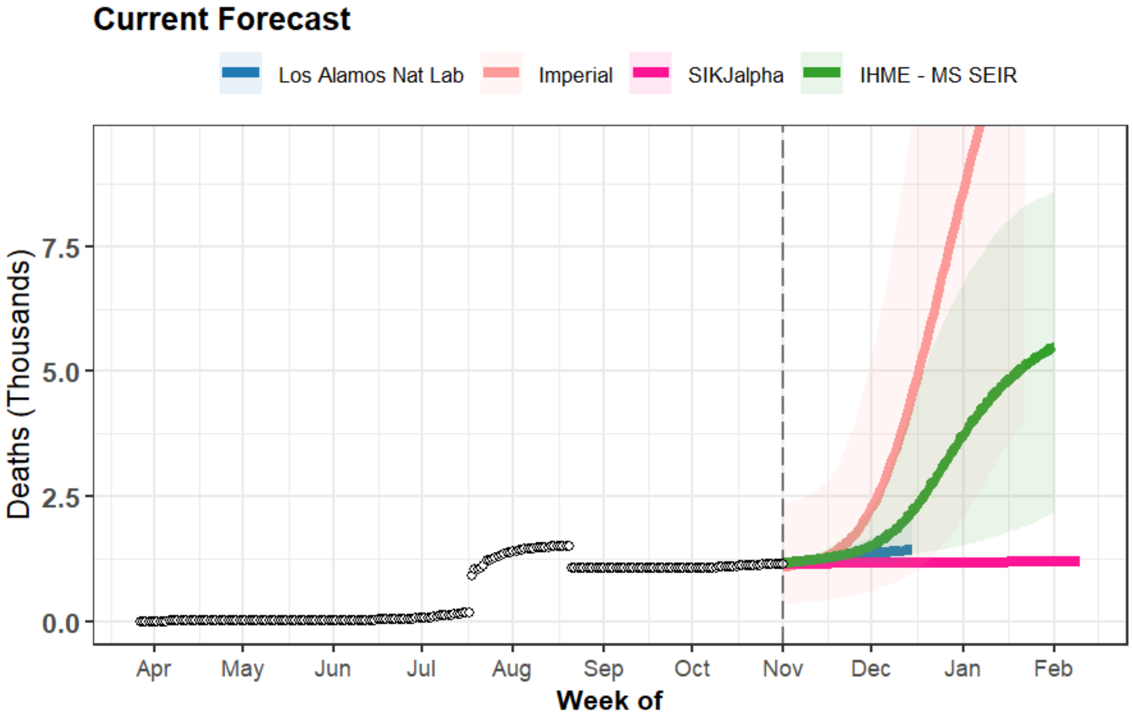
Current Forecast
Los Alamos Nat Lab
Imperial
SIKJalpha
IHME - MS SEIR
Deaths (Thousands)
7.5
5.0
2.5
0.0
Apr
May
Jun
Jul
Aug
Sep
Oct
Nov
Dec
Jan
Feb
Week of

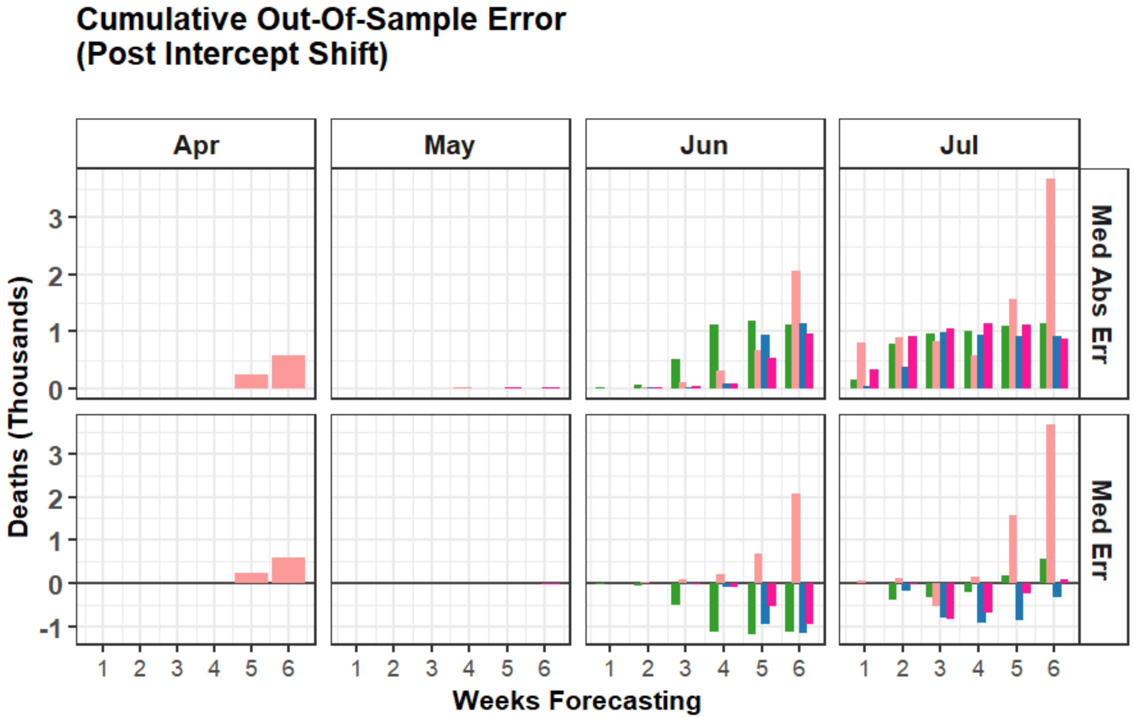
Cumulative Out-Of-Sample Error
(Post Intercept Shift)
Apr
May
Jun
Jul
Med Abs Err
Med Err
Deaths (Thousands)
Weeks Forecasting

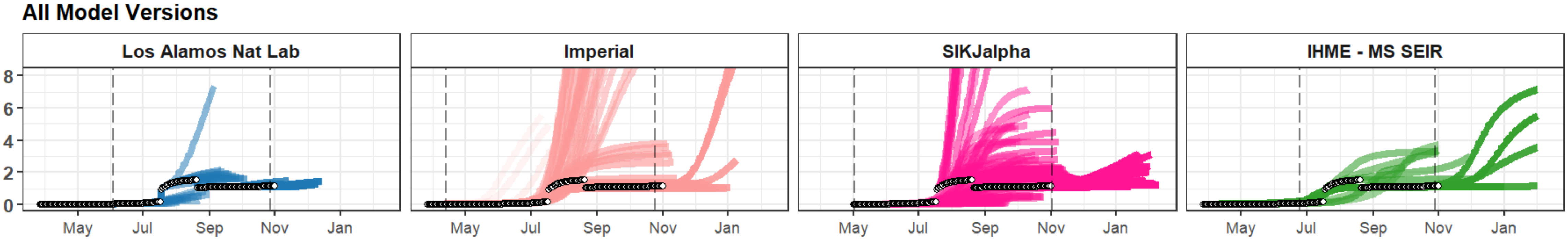
All Model Versions
Los Alamos Nat Lab
Imperial
SIKJalpha
IHME - MS SEIR
May
Jul
Sep
Nov
Jan

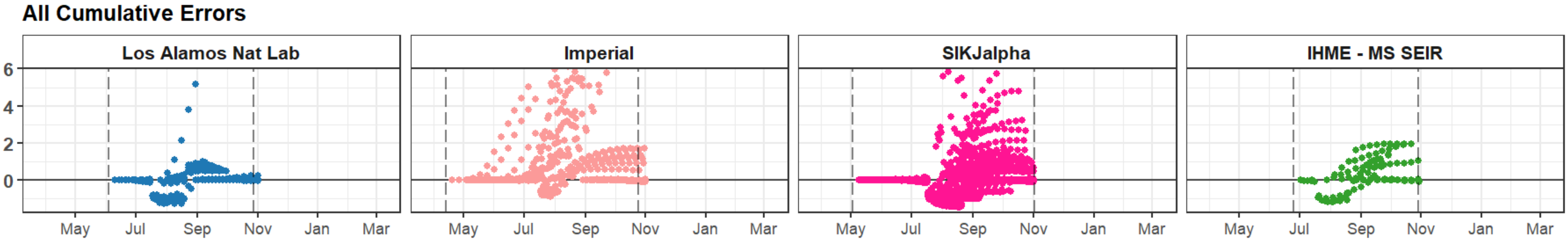
All Cumulative Errors
Los Alamos Nat Lab
Imperial
SIKJalpha
IHME - MS SEIR
May
Jul
Sep
Nov
Jan
Mar

### Austria

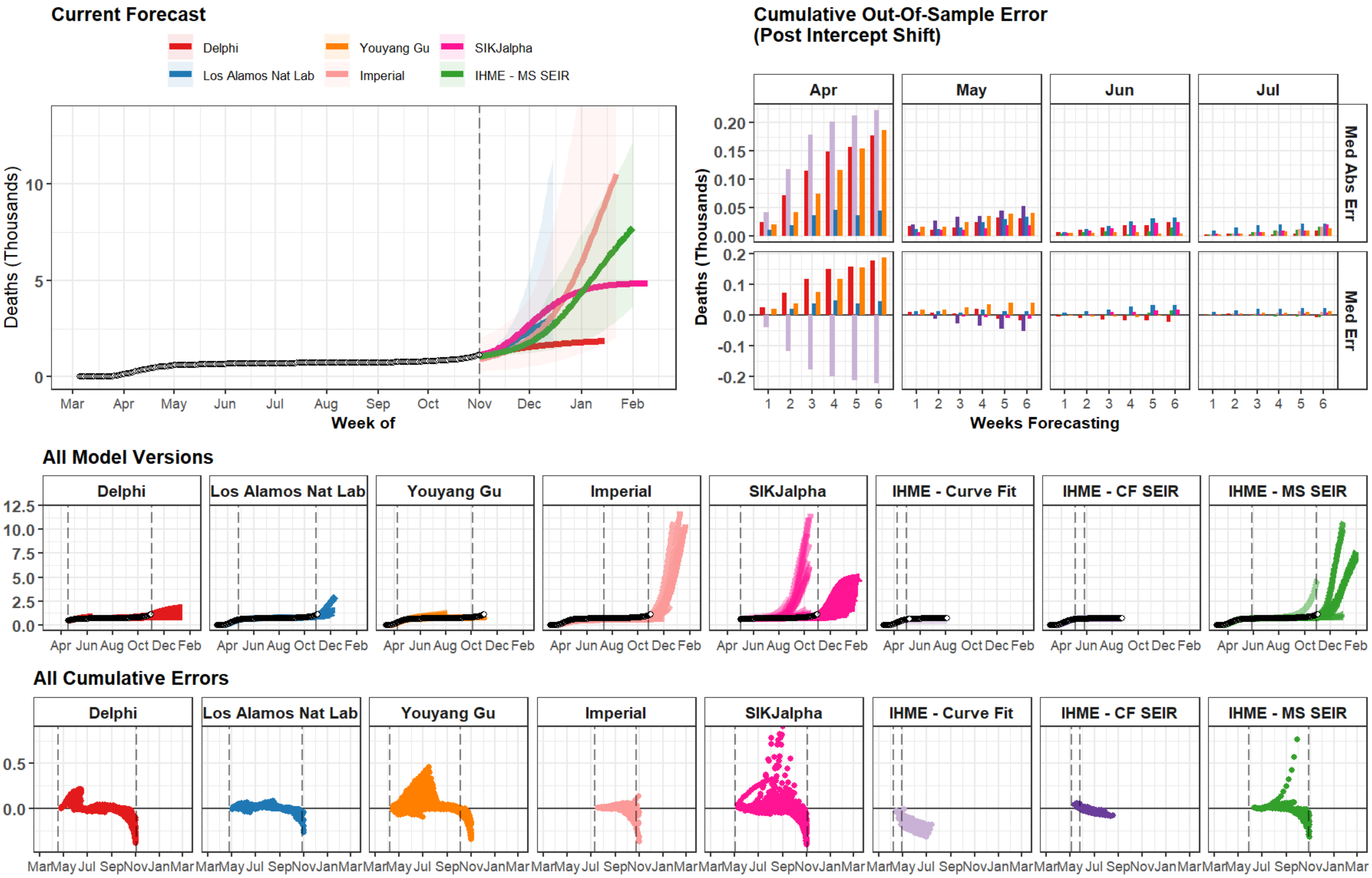
Current Forecast
Delphi
Los Alamos Nat Lab
Youyang Gu
Imperial
SIKJalpha
IHME - MS SEIR
Deaths (Thousands)
Week of
Mar Apr May Jun Jul Aug Sep Oct Nov Dec Jan Feb
Cumulative Out-Of-Sample Error
(Post Intercept Shift)
Apr
May
Jun
Jul
Med Abs Err
Med Err
Deaths (Thousands)
Weeks Forecasting
All Model Versions
Delphi
Los Alamos Nat Lab
Youyang Gu
Imperial
SIKJalpha
IHME - Curve Fit
IHME - CF SEIR
IHME - MS SEIR
Apr Jun Aug Oct Dec Feb
All Cumulative Errors
Delphi
Los Alamos Nat Lab
Youyang Gu
Imperial
SIKJalpha
IHME - Curve Fit
IHME - CF SEIR
IHME - MS SEIR
Mar May Jul Sep Nov Jan Mar

### Kansas

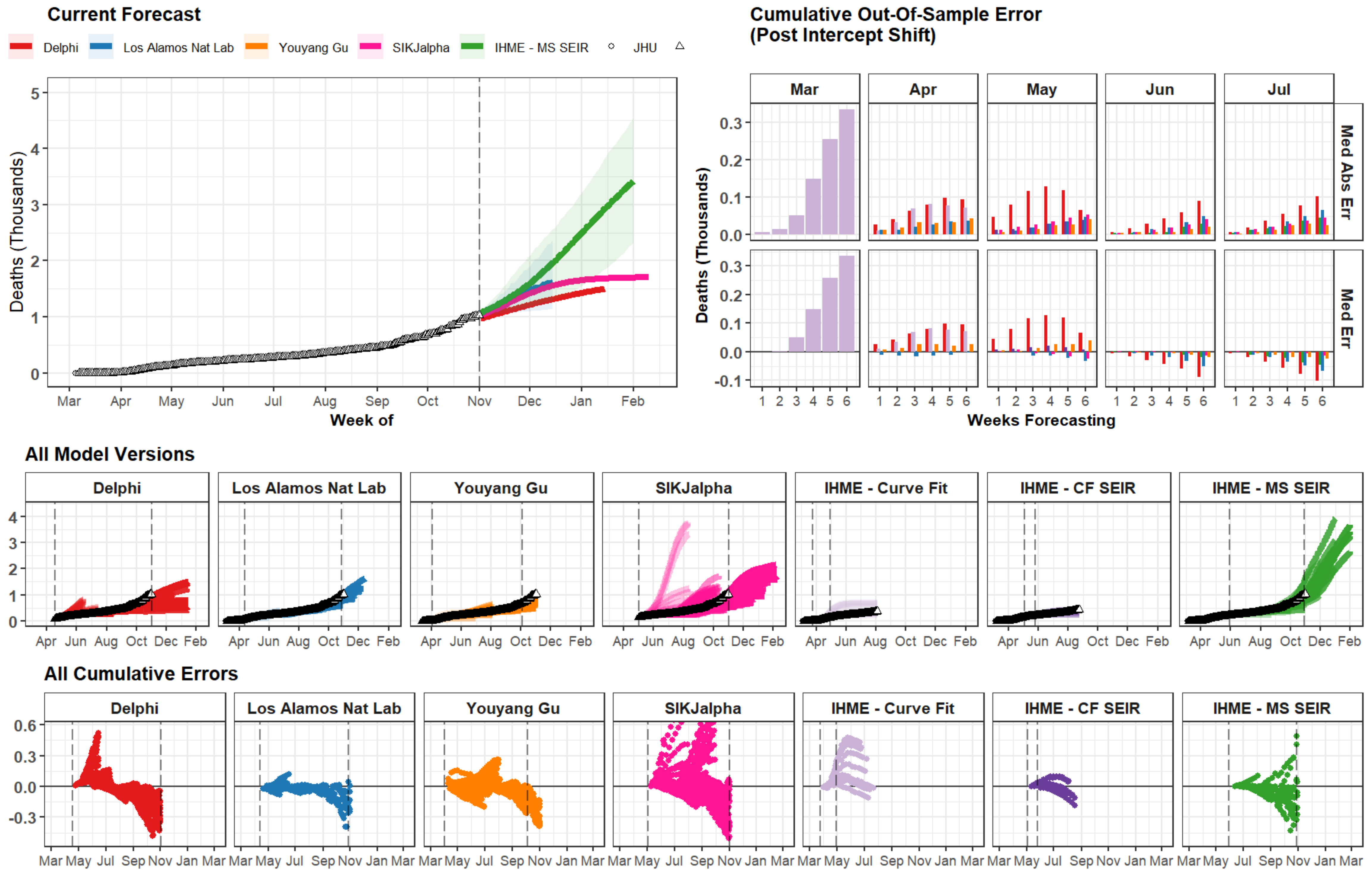

Current Forecast
Delphi
Los Alamos Nat Lab
Youyang Gu
SIKJalpha
IHME - MS SEIR
JHU
Deaths (Thousands)
Week of
Mar Apr May Jun Jul Aug Sep Oct Nov Dec Jan Feb
Cumulative Out-Of-Sample Error
(Post Intercept Shift)
Mar Apr May Jun Jul
Med Abs Err
Med Err
Weeks Forecasting
All Model Versions
Delphi
Los Alamos Nat Lab
Youyang Gu
SIKJalpha
IHME - Curve Fit
IHME - CF SEIR
IHME - MS SEIR
Apr Jun Aug Oct Dec Feb
All Cumulative Errors
Mar May Jul Sep Nov Jan Mar

### New Mexico

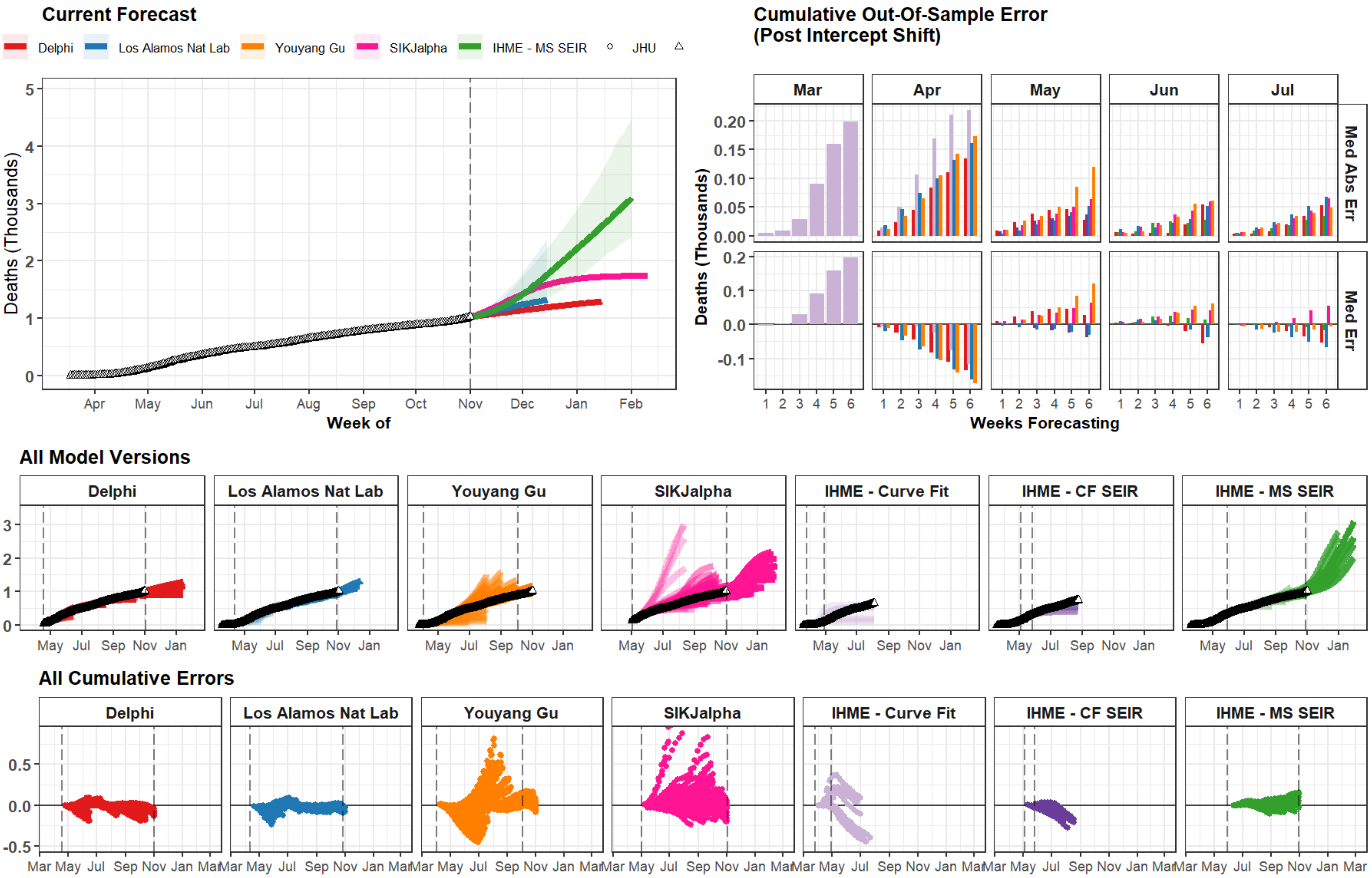
Current Forecast
Delphi
Los Alamos Nat Lab
Youyang Gu
SIKJalpha
IHME - MS SEIR
JHU
Deaths (Thousands)
Week of
Apr May Jun Jul Aug Sep Oct Nov Dec Jan Feb
Cumulative Out-Of-Sample Error
(Post Intercept Shift)
Mar Apr May Jun Jul
Med Abs Err
Med Err
Weeks Forecasting
All Model Versions
Delphi
Los Alamos Nat Lab
Youyang Gu
SIKJalpha
IHME - Curve Fit
IHME - CF SEIR
IHME - MS SEIR
May Jul Sep Nov Jan
All Cumulative Errors
Mar May Jul Sep Nov Jan Mar

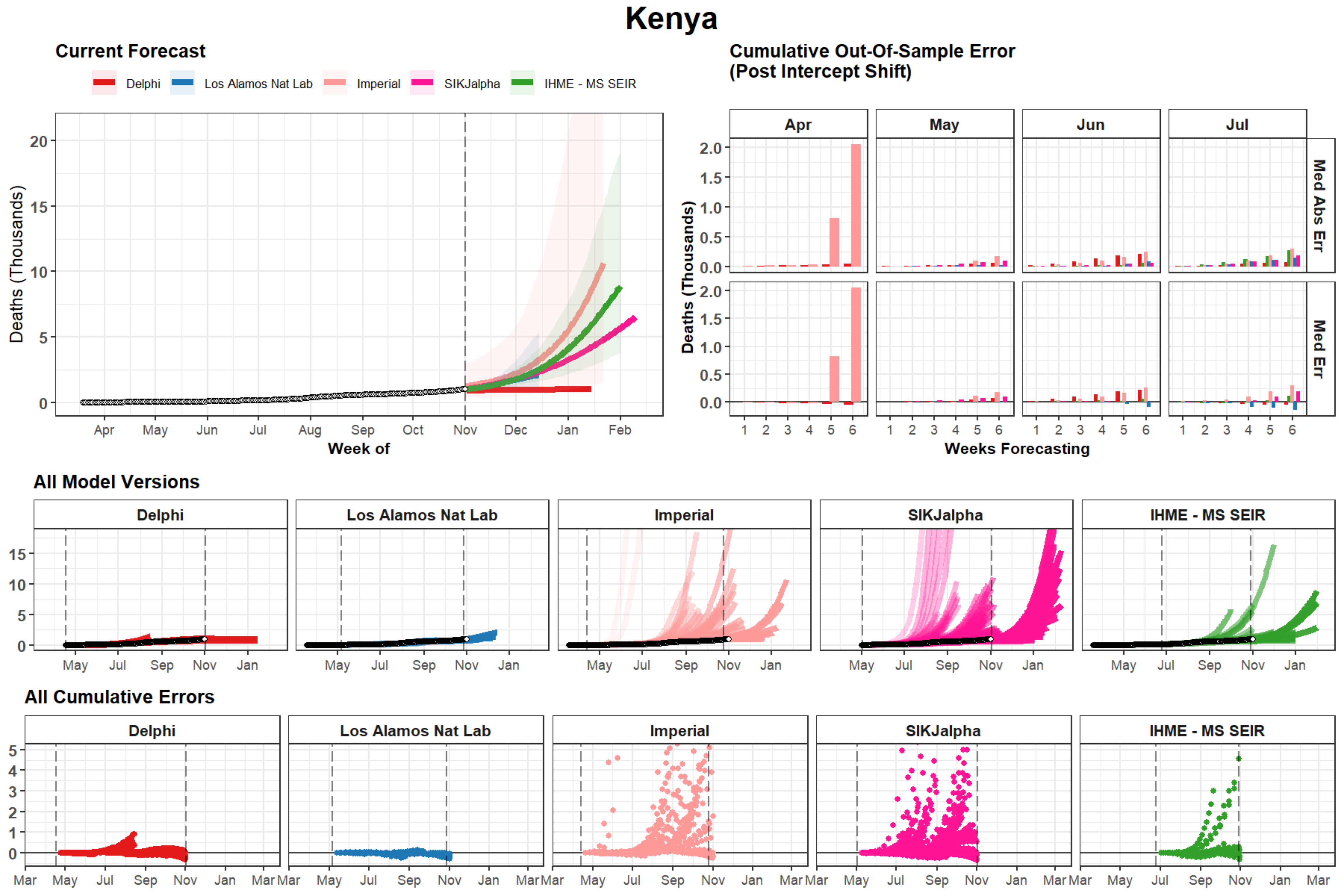

Kenya
Current Forecast
Delphi
Los Alamos Nat Lab
Imperial
SIKJalpha
IHME - MS SEIR
Deaths (Thousands)
Week of
Apr
May
Jun
Jul
Aug
Sep
Oct
Nov
Dec
Jan
Feb
Cumulative Out-Of-Sample Error
(Post Intercept Shift)
Med Abs Err
Med Err
Weeks Forecasting
All Model Versions
All Cumulative Errors
Mar

### Macedonia

#### Current Forecast

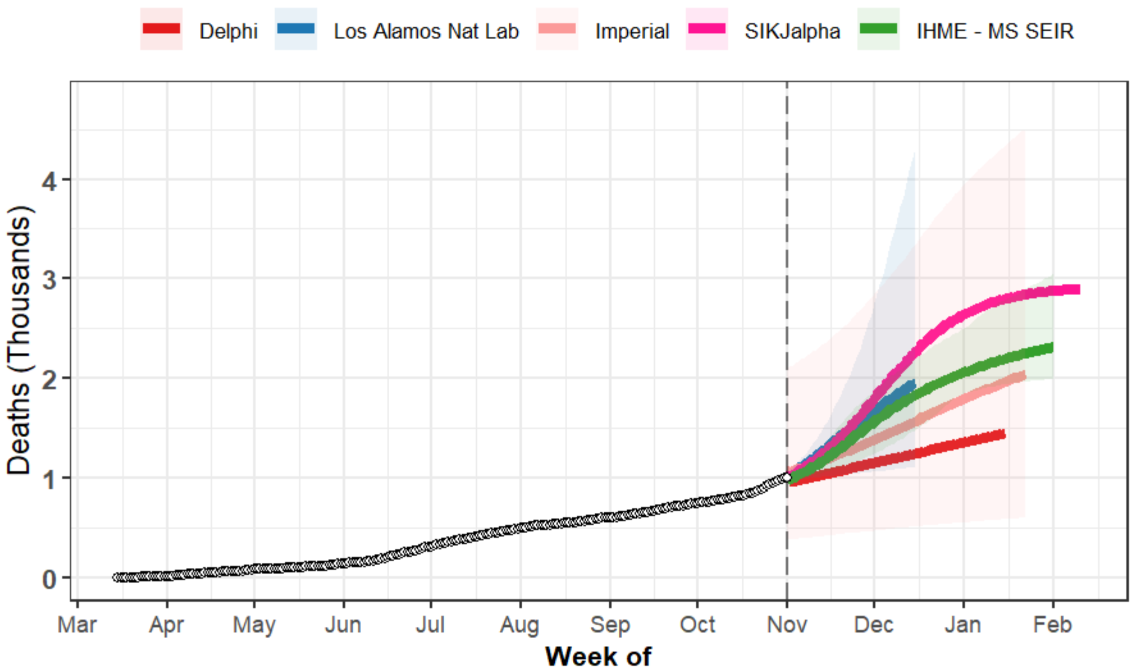


#### Cumulative Out-Of-Sample Error (Post Intercept Shift)

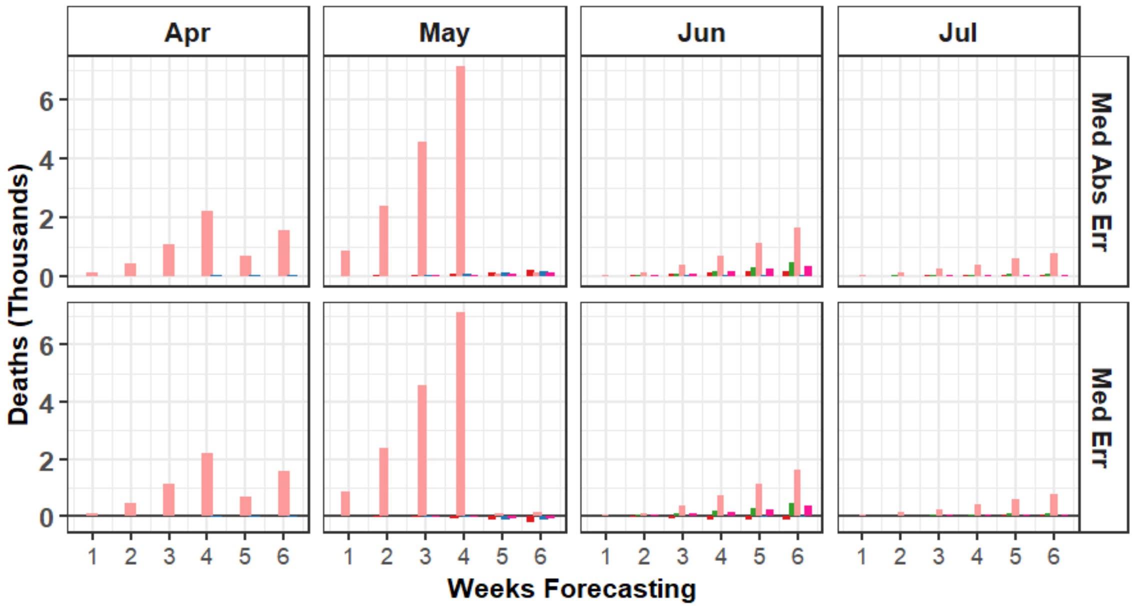


#### All Model Versions

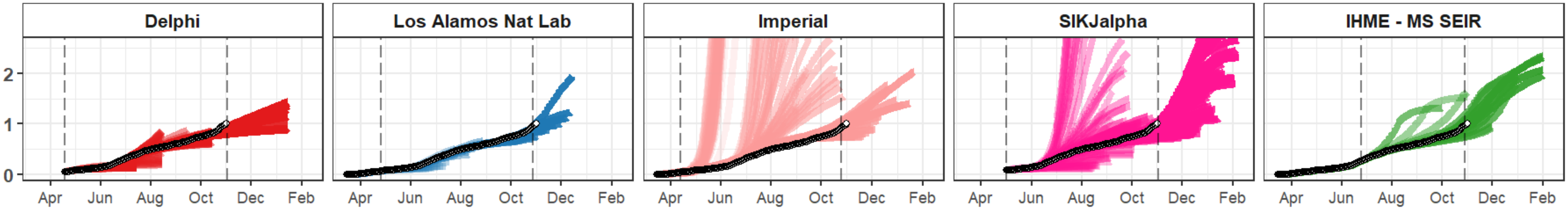


#### All Cumulative Errors

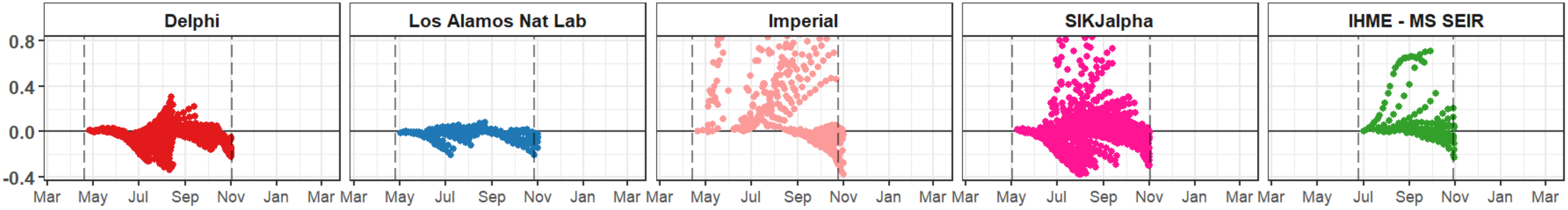

### Belarus

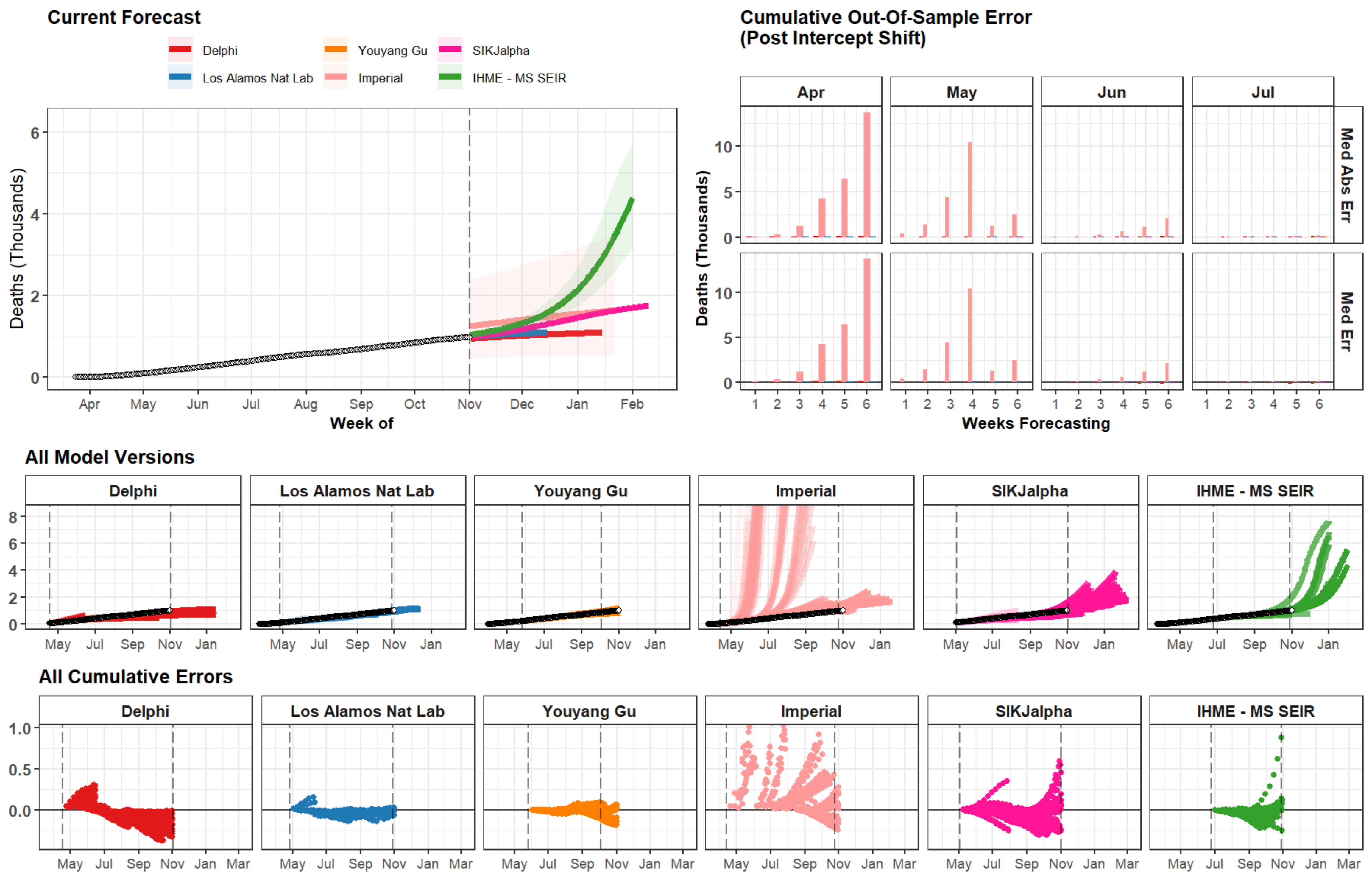

Current Forecast
Delphi
Los Alamos Nat Lab
Youyang Gu
Imperial
SIKJalpha
IHME - MS SEIR
Deaths (Thousands)
Week of
Apr May Jun Jul Aug Sep Oct Nov Dec Jan Feb
Cumulative Out-Of-Sample Error
(Post Intercept Shift)
Apr
May
Jun
Jul
Med Abs Err
Med Err
Weeks Forecasting
All Model Versions
All Cumulative Errors
May Jul Sep Nov Jan Mar

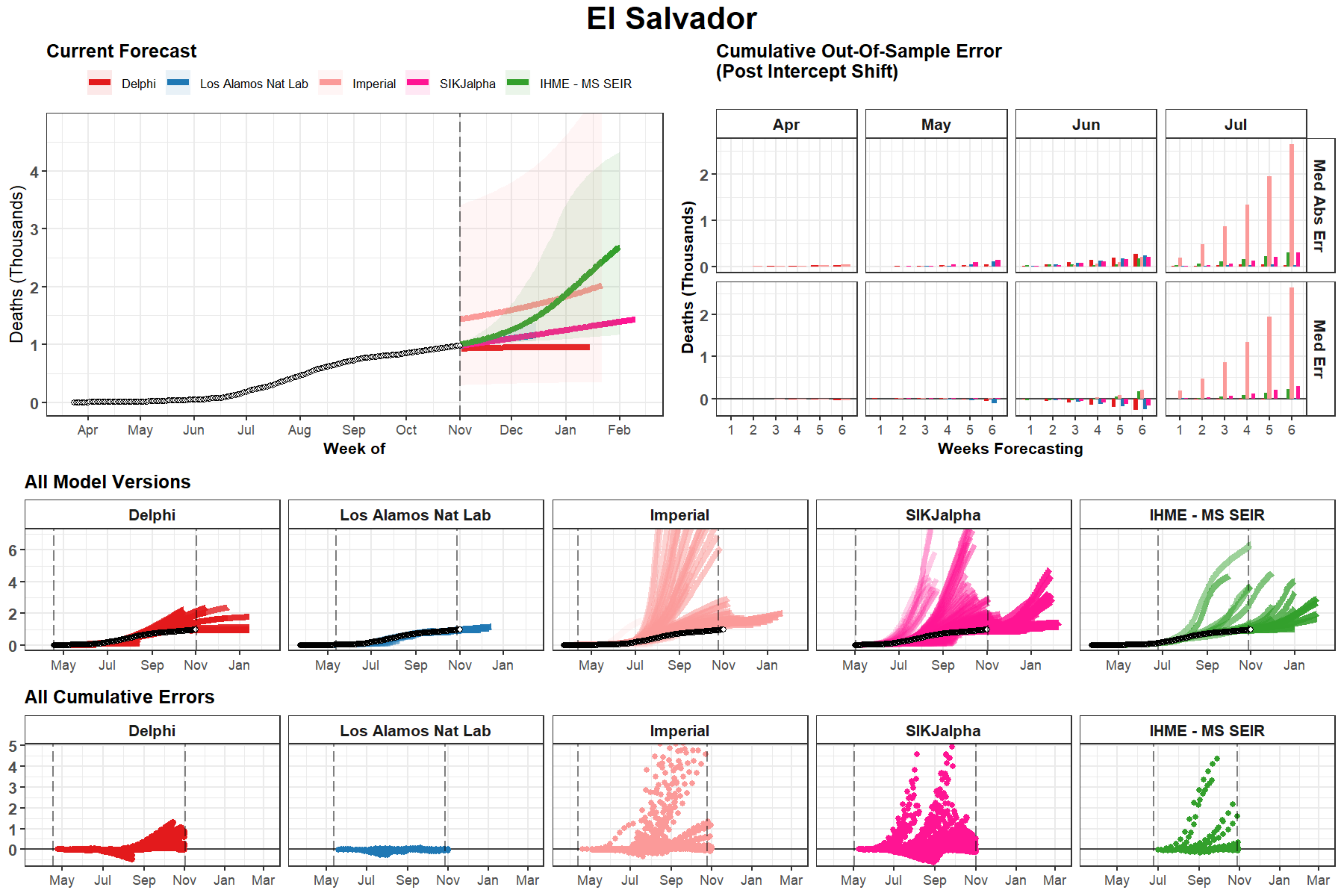
El Salvador
Current Forecast
Delphi
Los Alamos Nat Lab
Imperial
SIKJalpha
IHME - MS SEIR
Deaths (Thousands)
Week of
Cumulative Out-Of-Sample Error
(Post Intercept Shift)
Apr
May
Jun
Jul
Med Abs Err
Med Err
Weeks Forecasting
All Model Versions
All Cumulative Errors

### Nepal

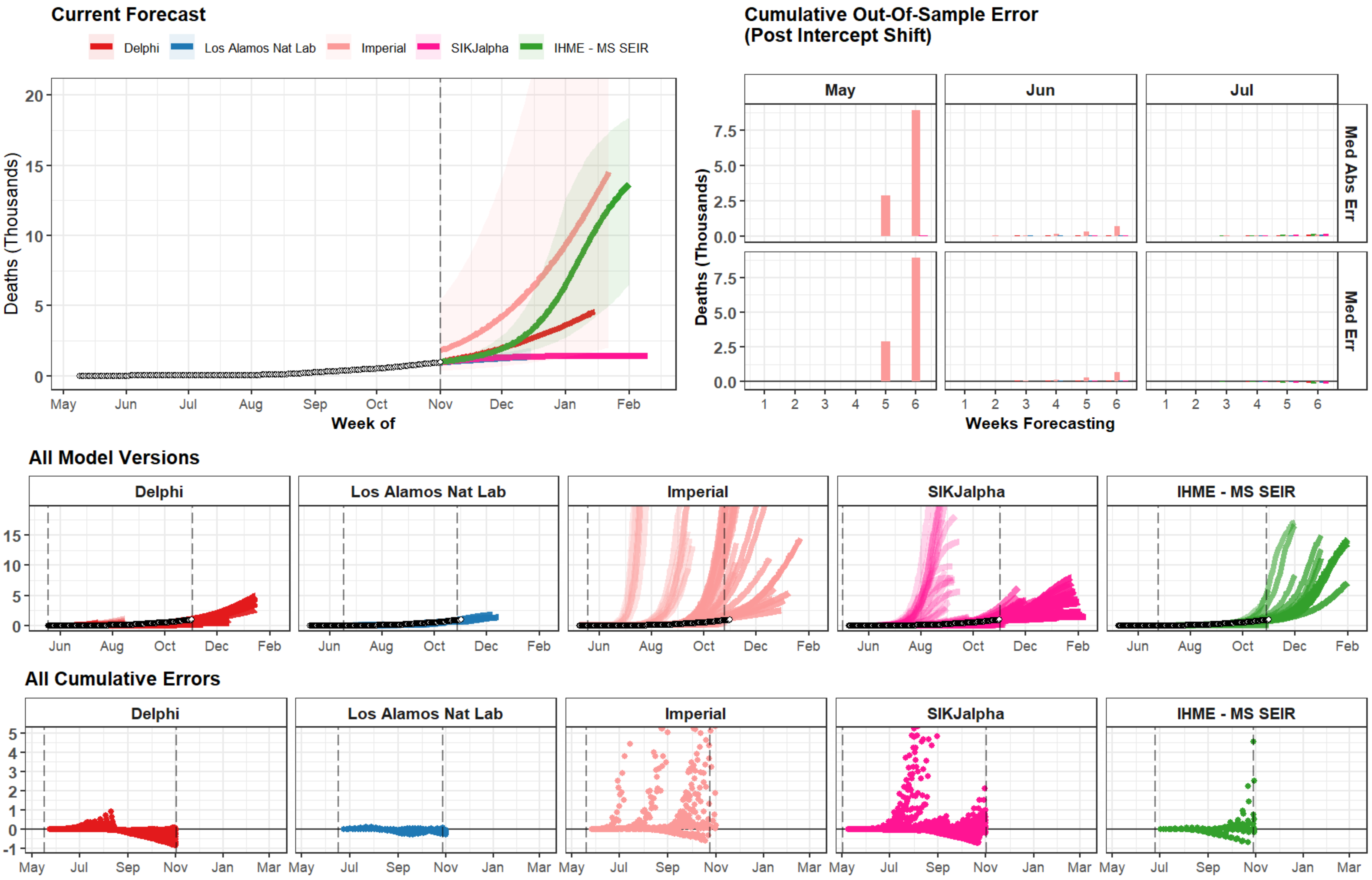
Current Forecast
Delphi
Los Alamos Nat Lab
Imperial
SIKJalpha
IHME - MS SEIR
Deaths (Thousands)
Week of
May
Jun
Jul
Aug
Sep
Oct
Nov
Dec
Jan
Feb
Cumulative Out-Of-Sample Error
(Post Intercept Shift)
May
Jun
Jul
Med Abs Err
Med Err
Deaths (Thousands)
Weeks Forecasting
All Model Versions
Delphi
Los Alamos Nat Lab
Imperial
SIKJalpha
IHME - MS SEIR
All Cumulative Errors
Delphi
Los Alamos Nat Lab
Imperial
SIKJalpha
IHME - MS SEIR

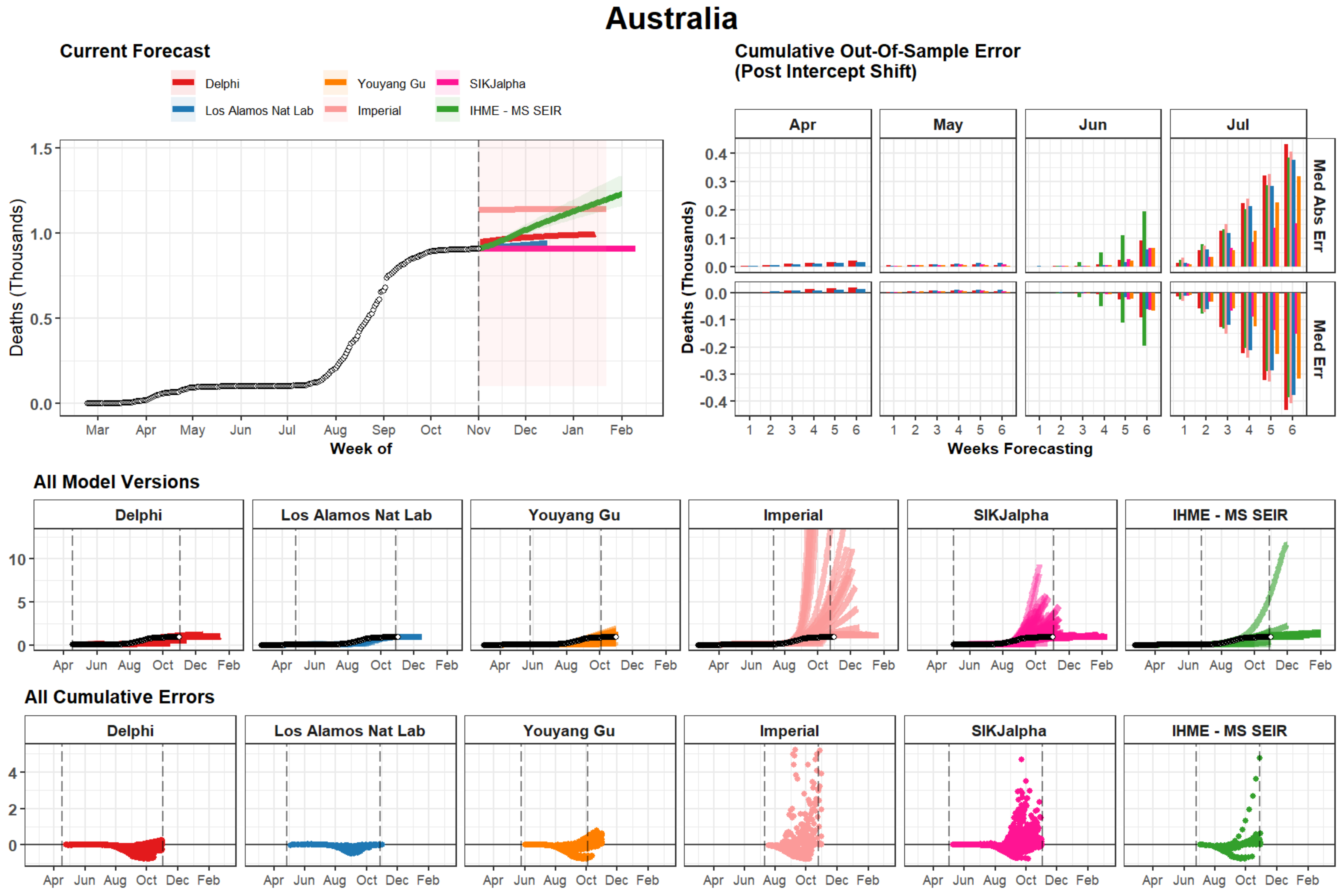
Australia
Current Forecast
Delphi
Youyang Gu
SIKJalpha
Los Alamos Nat Lab
Imperial
IHME - MS SEIR
Deaths (Thousands)
Week of
Mar
Apr
May
Jun
Jul
Aug
Sep
Oct
Nov
Dec
Jan
Feb
Cumulative Out-Of-Sample Error
(Post Intercept Shift)
Apr
May
Jun
Jul
Med Abs Err
Med Err
Weeks Forecasting
All Model Versions
All Cumulative Errors

### Libya

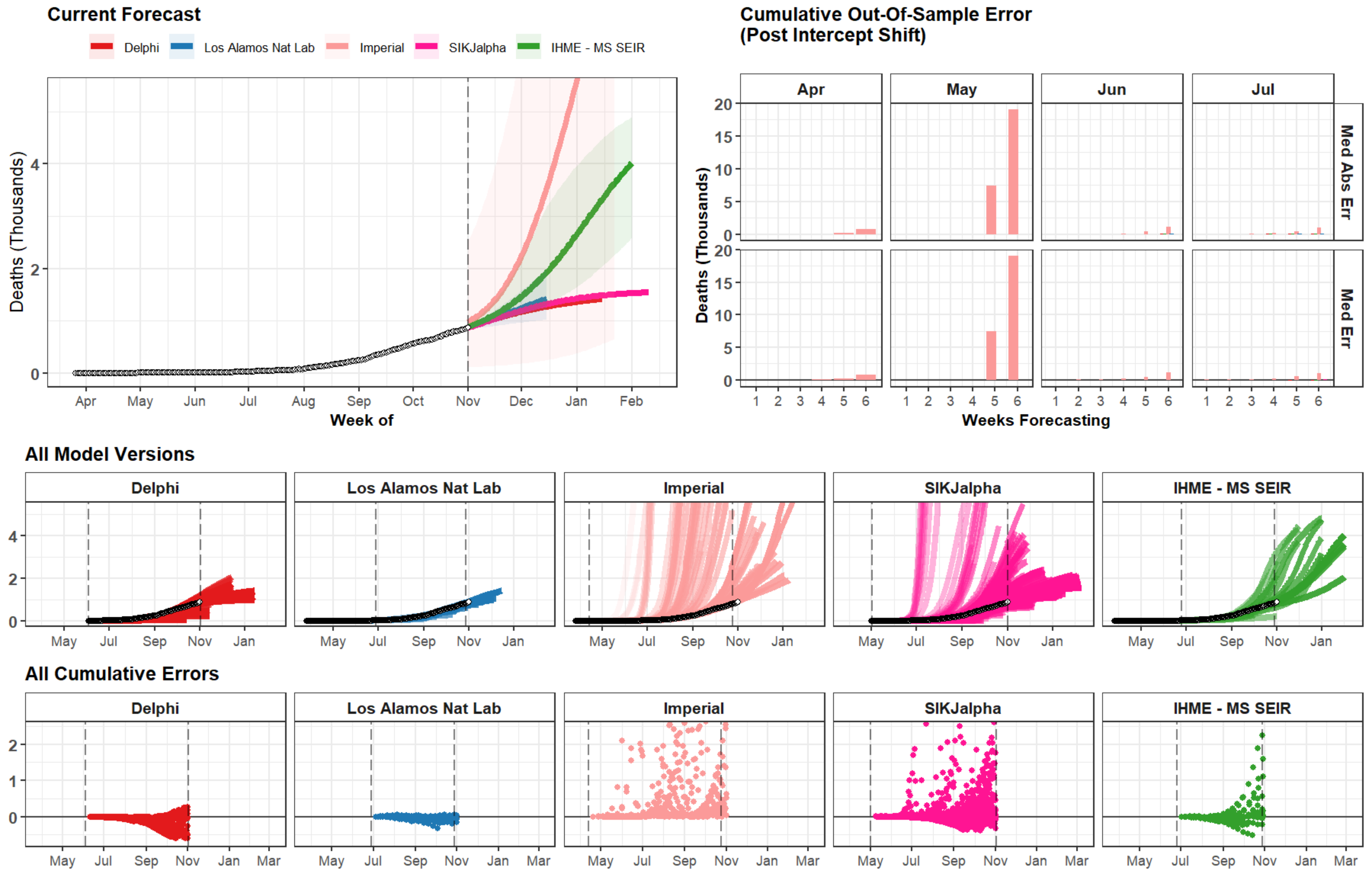
Current Forecast
Delphi
Los Alamos Nat Lab
Imperial
SIKJalpha
IHME - MS SEIR
Deaths (Thousands)
Week of
Cumulative Out-Of-Sample Error
(Post Intercept Shift)
Apr
May
Jun
Jul
Med Abs Err
Med Err
Weeks Forecasting
All Model Versions
All Cumulative Errors

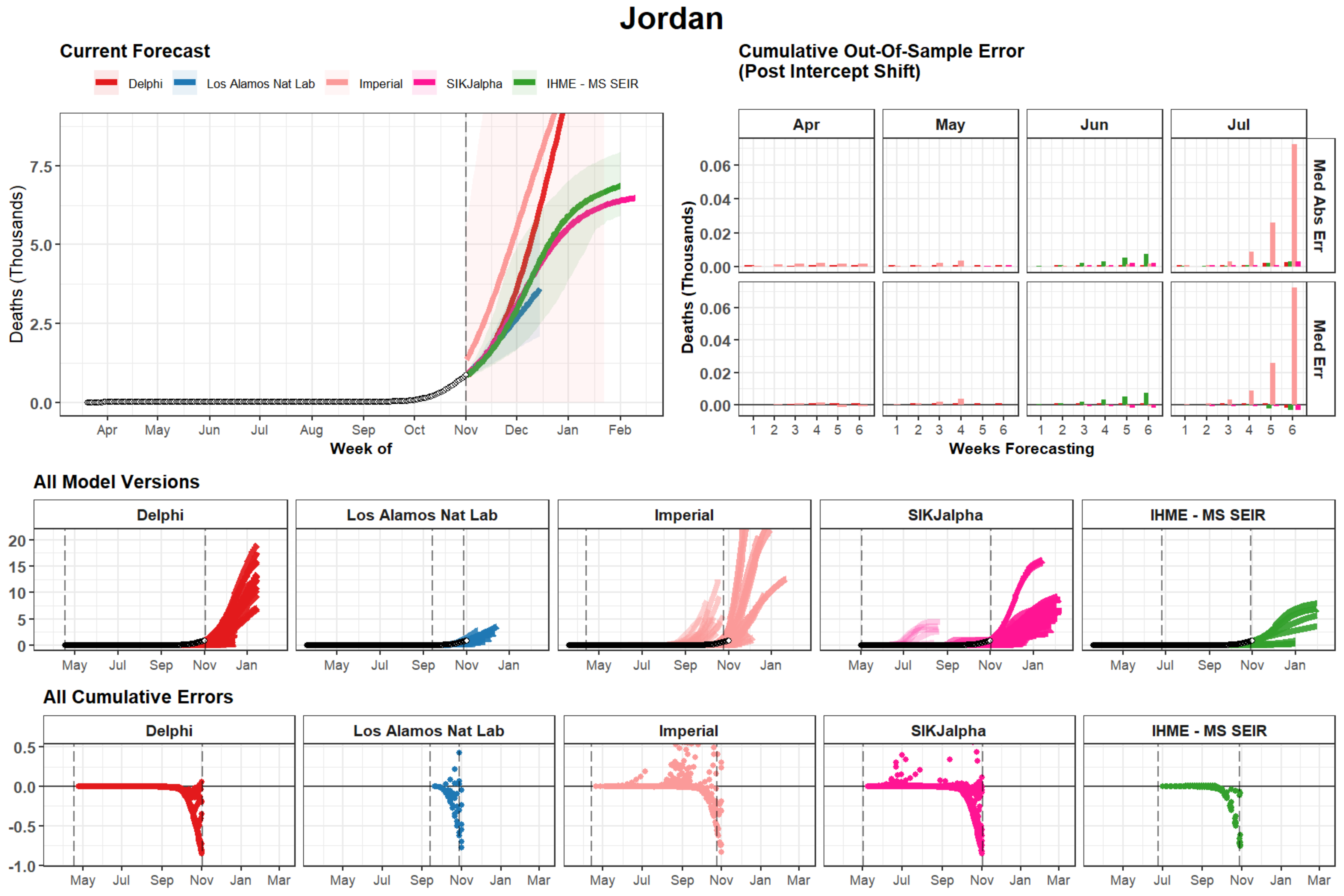
Jordan
Current Forecast
Delphi
Los Alamos Nat Lab
Imperial
SIKJalpha
IHME - MS SEIR
Deaths (Thousands)
Week of
Cumulative Out-Of-Sample Error
(Post Intercept Shift)
Apr
May
Jun
Jul
Med Abs Err
Med Err
Weeks Forecasting
All Model Versions
All Cumulative Errors

### Sudan

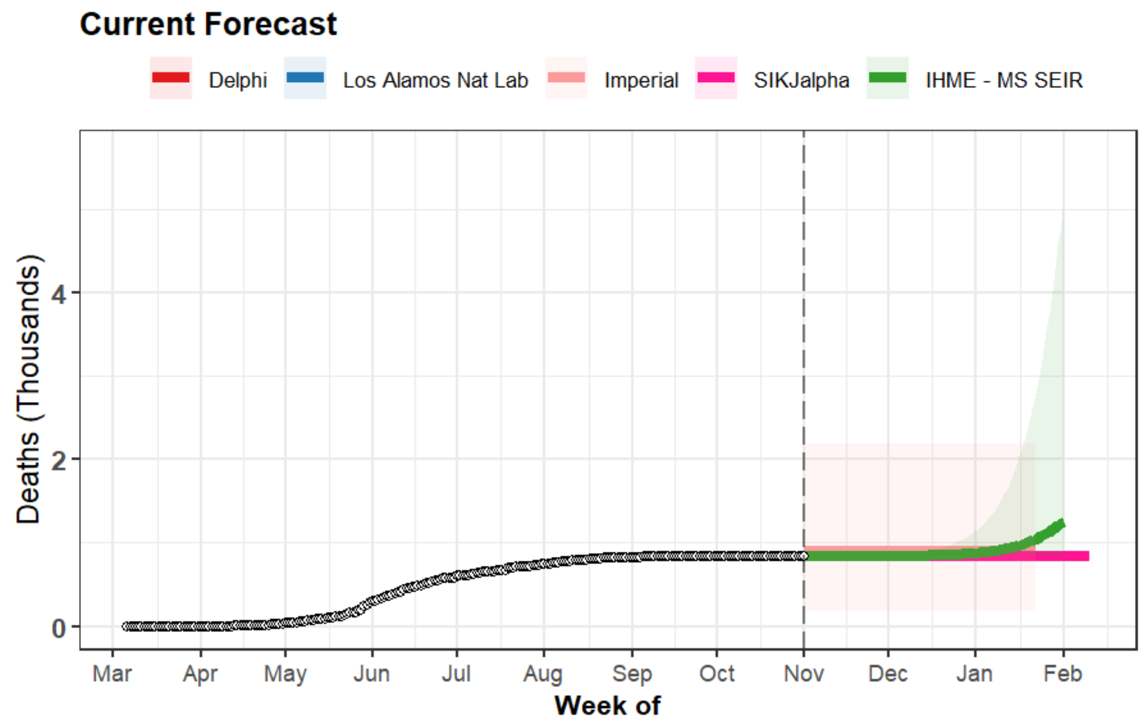
Current Forecast
Delphi
Los Alamos Nat Lab
Imperial
SIKJalpha
IHME - MS SEIR
Deaths (Thousands)
0
2
4
Mar
Apr
May
Jun
Jul
Aug
Sep
Oct
Nov
Dec
Jan
Feb
Week of

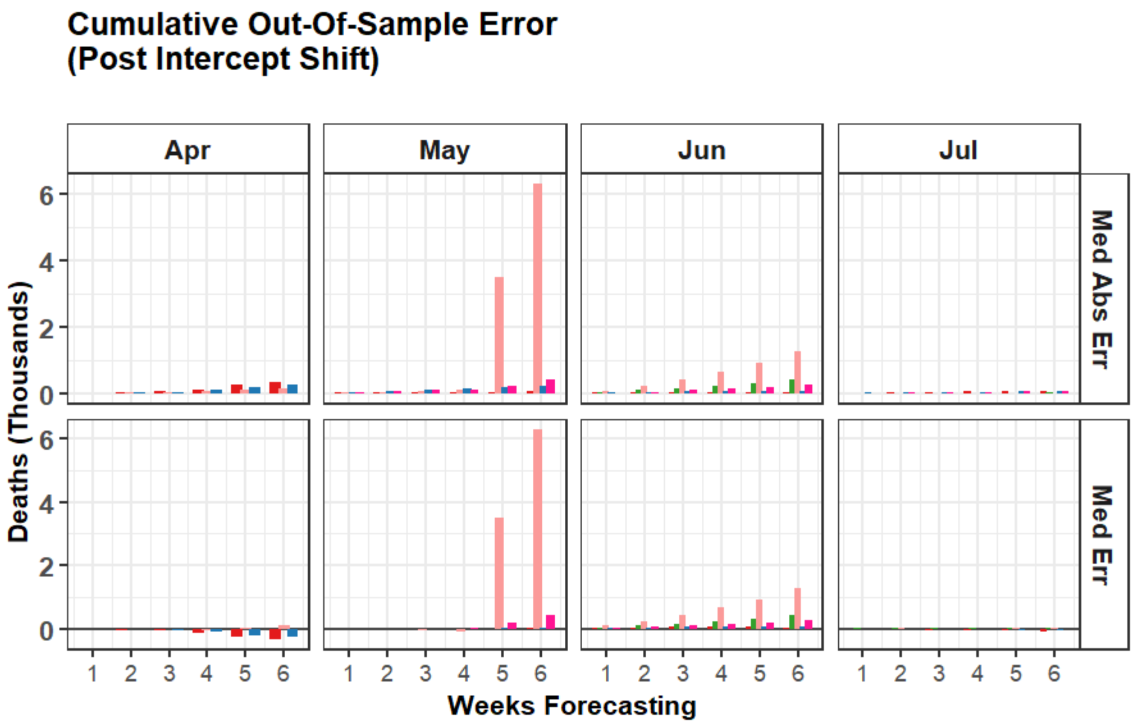
Cumulative Out-Of-Sample Error
(Post Intercept Shift)
Apr
May
Jun
Jul
Med Abs Err
Med Err
Deaths (Thousands)
0
2
4
6
1
2
3
4
5
6
Weeks Forecasting

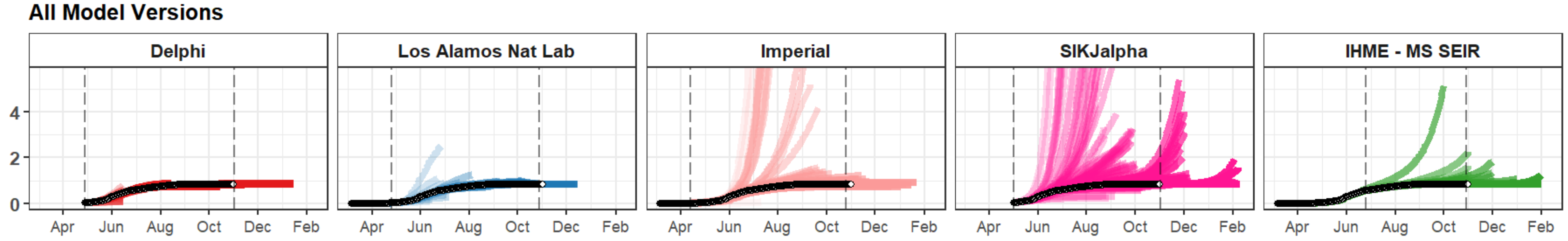
All Model Versions
Delphi
Los Alamos Nat Lab
Imperial
SIKJalpha
IHME - MS SEIR
0
2
4
Apr
Jun
Aug
Oct
Dec
Feb

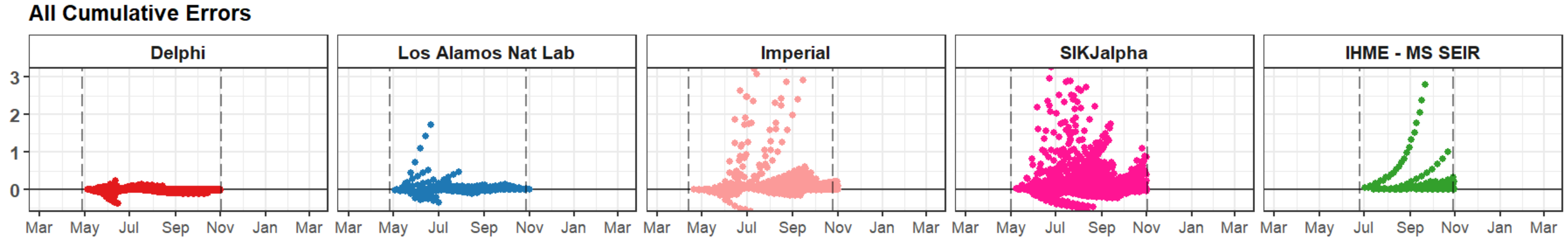
All Cumulative Errors
Delphi
Los Alamos Nat Lab
Imperial
SIKJalpha
IHME - MS SEIR
0
1
2
3
Mar
May
Jul
Sep
Nov
Jan
Mar

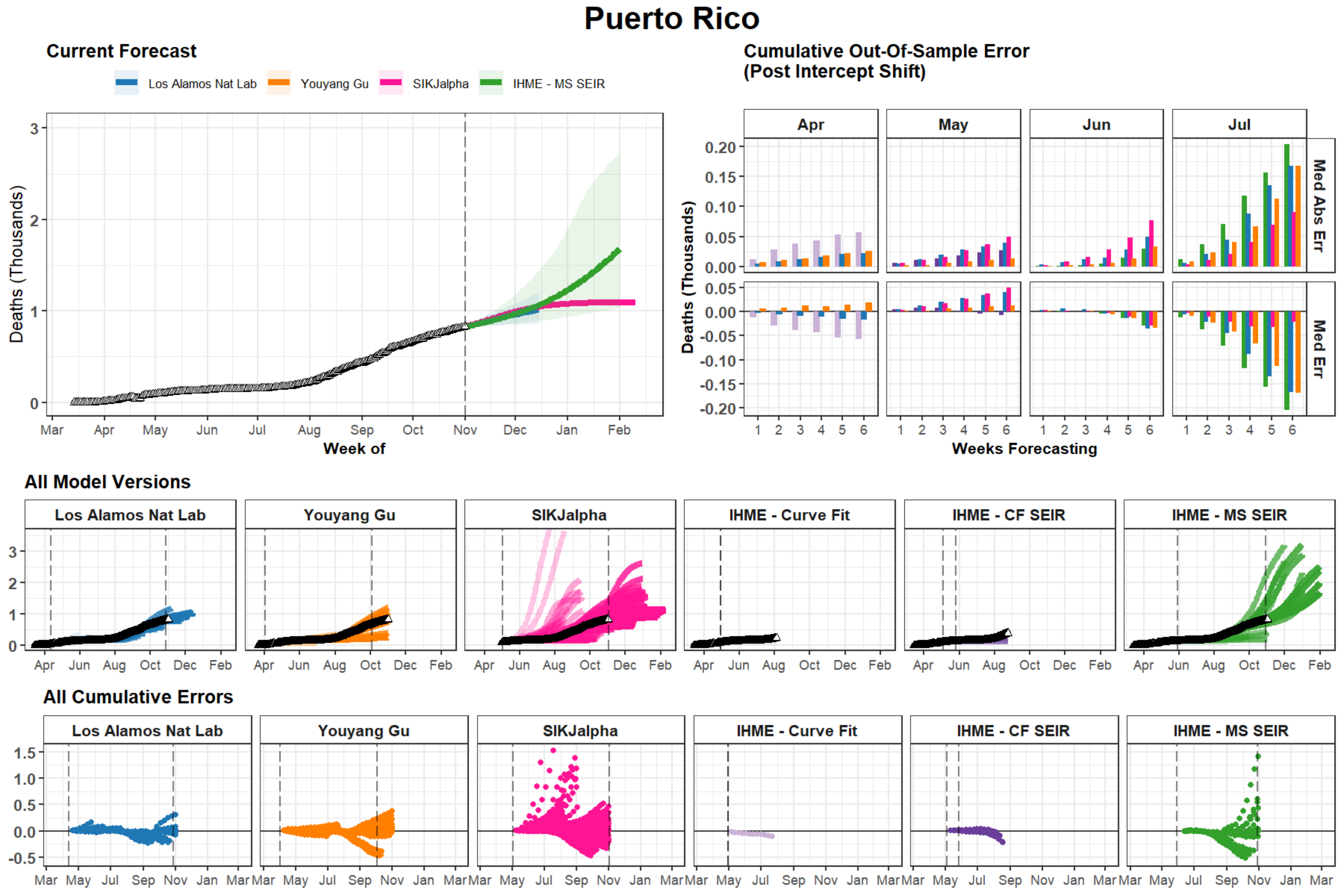
Puerto Rico
Current Forecast
Los Alamos Nat Lab
Youyang Gu
SIKJalpha
IHME - MS SEIR
Deaths (Thousands)
Week of
Mar
Apr
May
Jun
Jul
Aug
Sep
Oct
Nov
Dec
Jan
Feb
Cumulative Out-Of-Sample Error
(Post Intercept Shift)
Med Abs Err
Med Err
Weeks Forecasting
All Model Versions
IHME - Curve Fit
IHME - CF SEIR
All Cumulative Errors

### Serbia

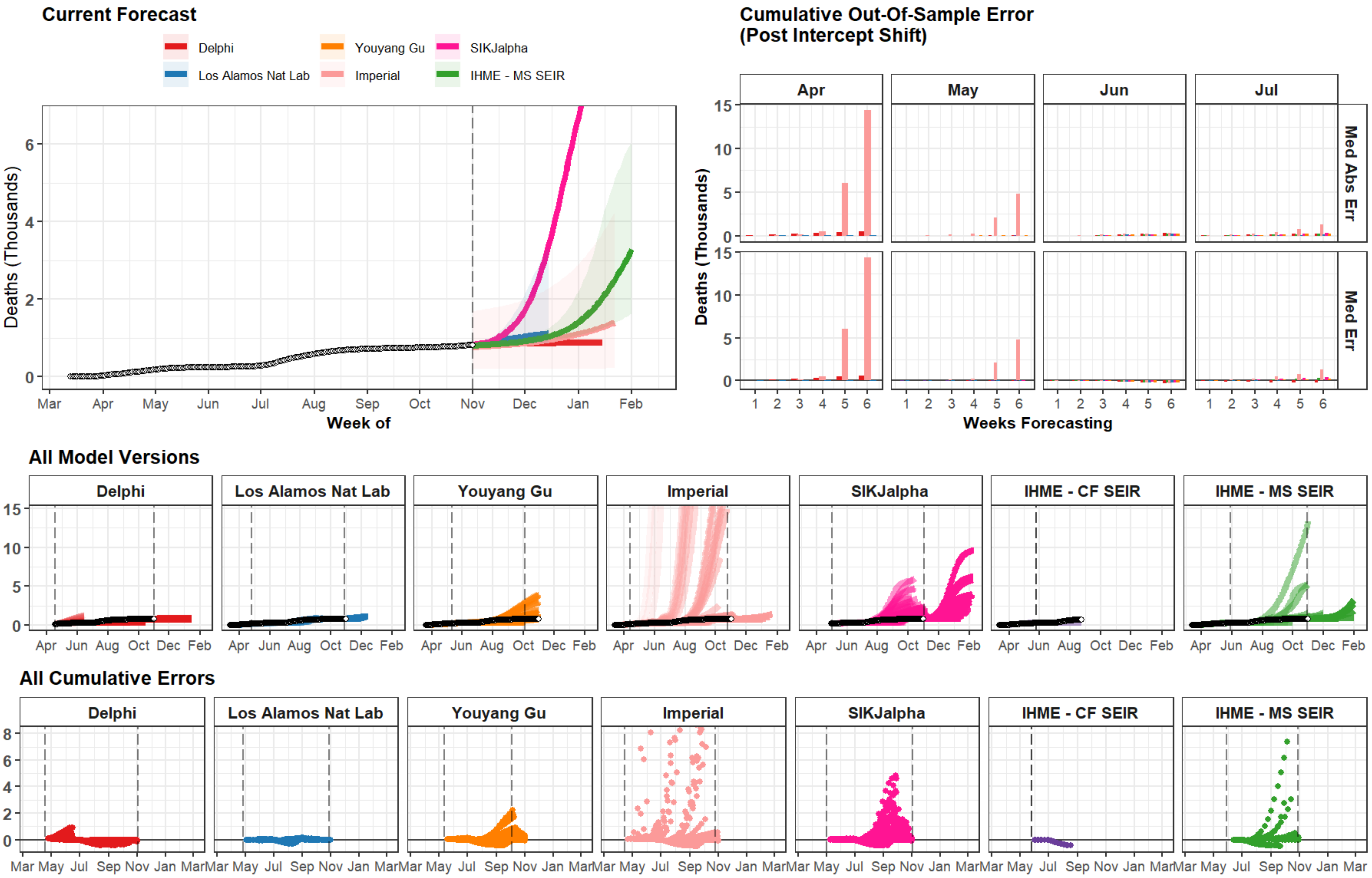
Current Forecast
Delphi
Los Alamos Nat Lab
Youyang Gu
Imperial
SIKJalpha
IHME - MS SEIR
Deaths (Thousands)
Week of
Mar Apr May Jun Jul Aug Sep Oct Nov Dec Jan Feb
Cumulative Out-Of-Sample Error
(Post Intercept Shift)
Apr May Jun Jul
Med Abs Err
Med Err
Weeks Forecasting
All Model Versions
Delphi
Los Alamos Nat Lab
Youyang Gu
Imperial
SIKJalpha
IHME - CF SEIR
IHME - MS SEIR
All Cumulative Errors

### Venezuela

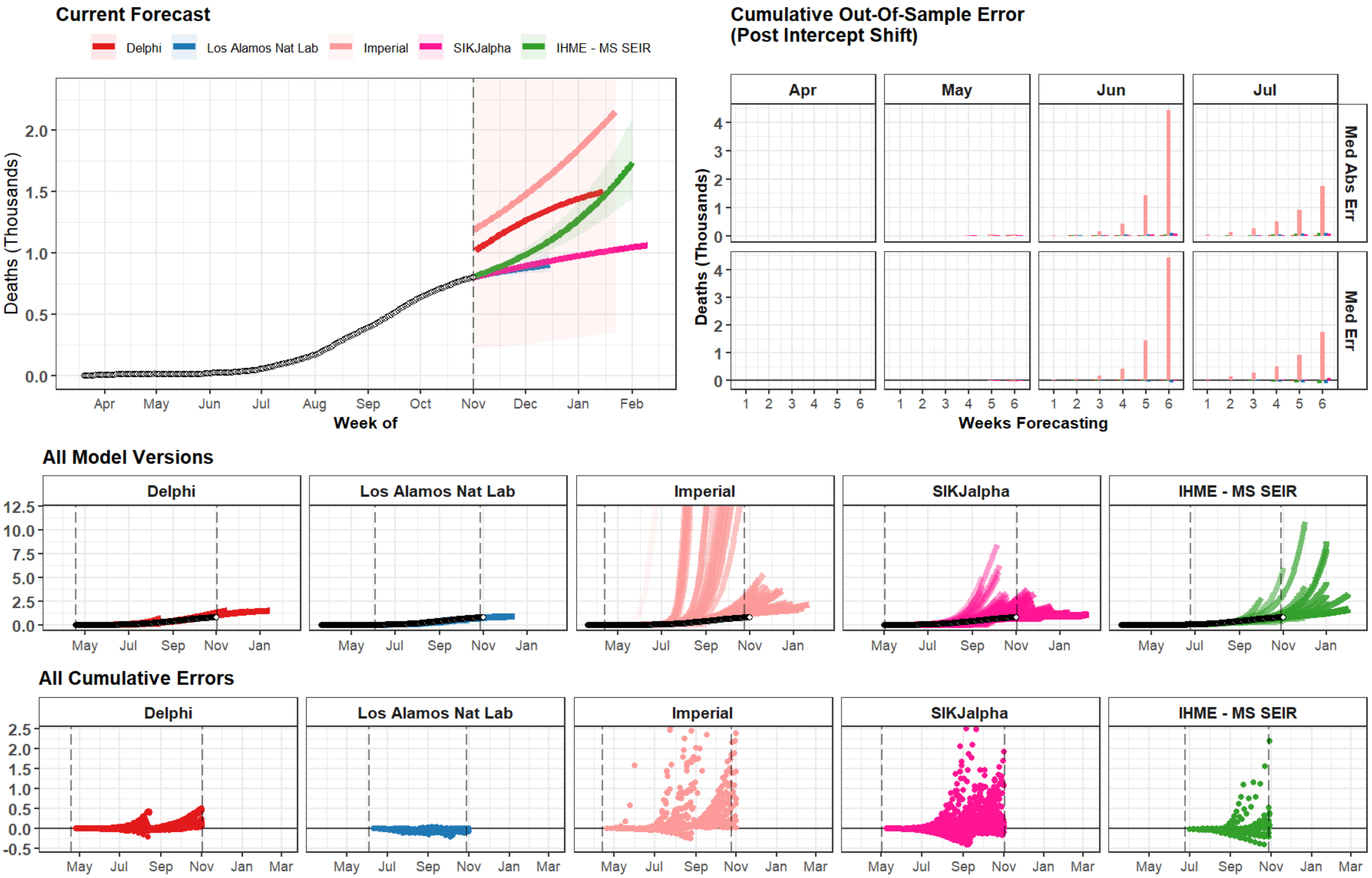
Current Forecast
Delphi
Los Alamos Nat Lab
Imperial
SIKJalpha
IHME - MS SEIR
Deaths (Thousands)
Week of
Apr
May
Jun
Jul
Aug
Sep
Oct
Nov
Dec
Jan
Feb
Cumulative Out-Of-Sample Error
(Post Intercept Shift)
Med Abs Err
Med Err
Weeks Forecasting
All Model Versions
All Cumulative Errors
Mar

### Kuwait

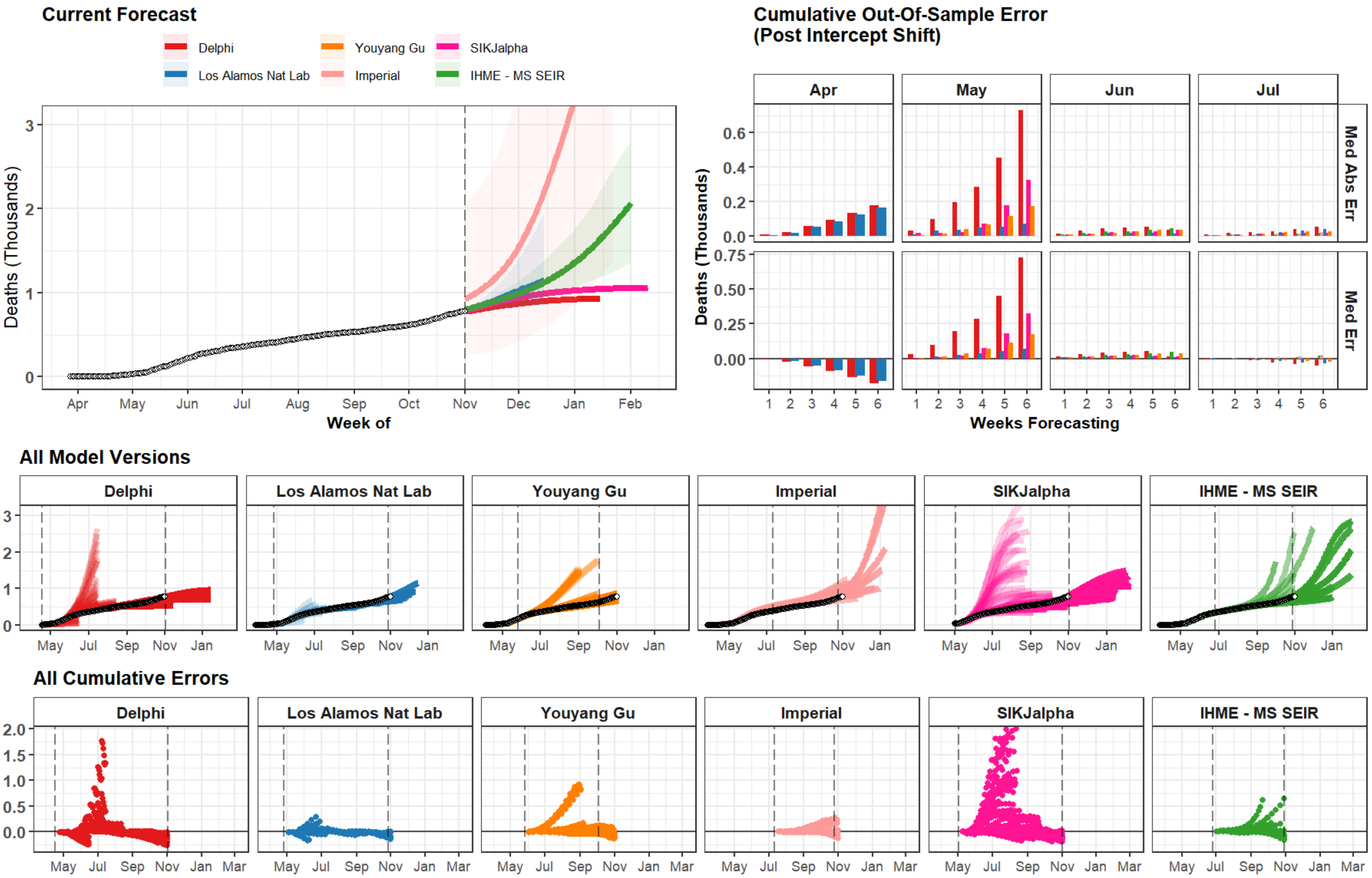
Current Forecast
Delphi
Los Alamos Nat Lab
Youyang Gu
Imperial
SIKJalpha
IHME - MS SEIR
Deaths (Thousands)
Week of
Apr
May
Jun
Jul
Aug
Sep
Oct
Nov
Dec
Jan
Feb
Cumulative Out-Of-Sample Error
(Post Intercept Shift)
Med Abs Err
Med Err
Weeks Forecasting
All Model Versions
All Cumulative Errors
Mar

### Azerbaijan

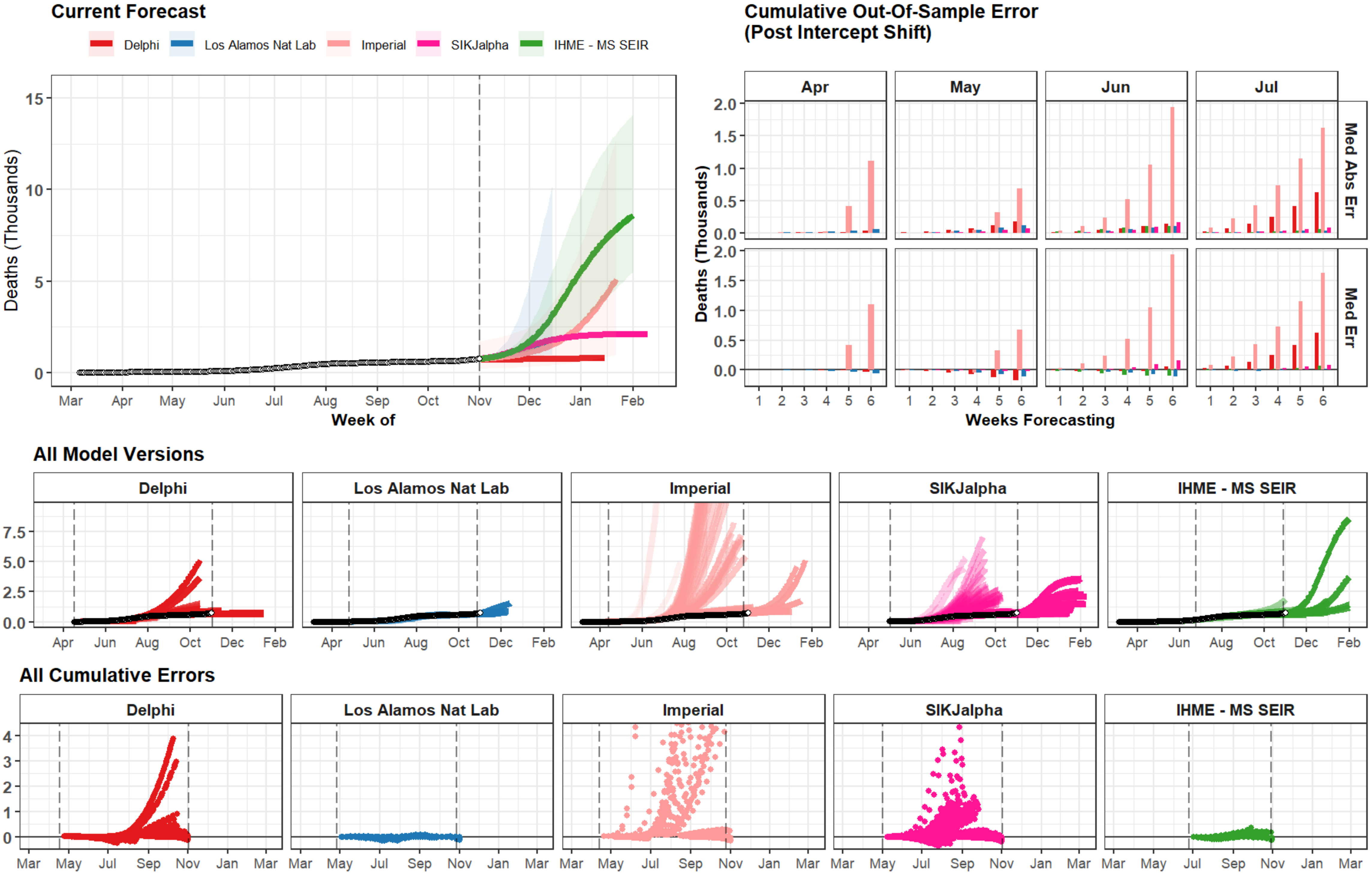

Current Forecast
Delphi
Los Alamos Nat Lab
Imperial
SIKJalpha
IHME - MS SEIR
Deaths (Thousands)
15
10
5
0
Mar
Apr
May
Jun
Jul
Aug
Sep
Oct
Nov
Dec
Jan
Feb
Week of
Cumulative Out-Of-Sample Error
(Post Intercept Shift)
Apr
May
Jun
Jul
Med Abs Err
Med Err
2.0
1.5
1.0
0.5
0.0
1
2
3
4
5
6
Weeks Forecasting
All Model Versions
Delphi
Los Alamos Nat Lab
Imperial
SIKJalpha
IHME - MS SEIR
7.5
5.0
2.5
0.0
Apr
Jun
Aug
Oct
Dec
Feb
All Cumulative Errors
Delphi
Los Alamos Nat Lab
Imperial
SIKJalpha
IHME - MS SEIR
4
3
2
1
0
Mar
May
Jul
Sep
Nov
Jan
Mar

### Denmark

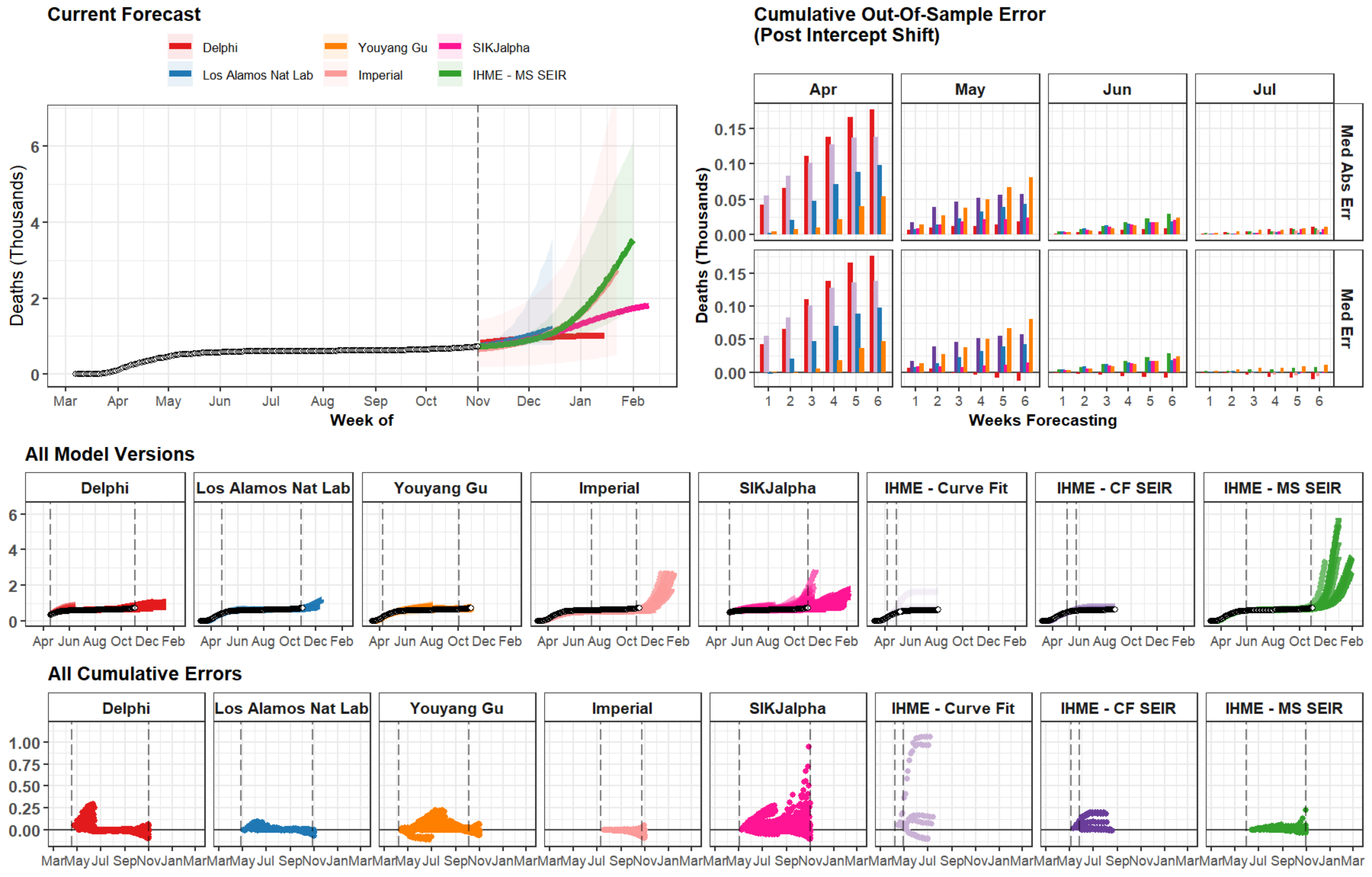
Current Forecast
Delphi
Los Alamos Nat Lab
Youyang Gu
Imperial
SIKJalpha
IHME - MS SEIR
Deaths (Thousands)
Week of
Cumulative Out-Of-Sample Error
(Post Intercept Shift)
Apr
May
Jun
Jul
Med Abs Err
Med Err
Weeks Forecasting
All Model Versions
Delphi
Los Alamos Nat Lab
Youyang Gu
Imperial
SIKJalpha
IHME - Curve Fit
IHME - CF SEIR
IHME - MS SEIR
All Cumulative Errors

### Delaware

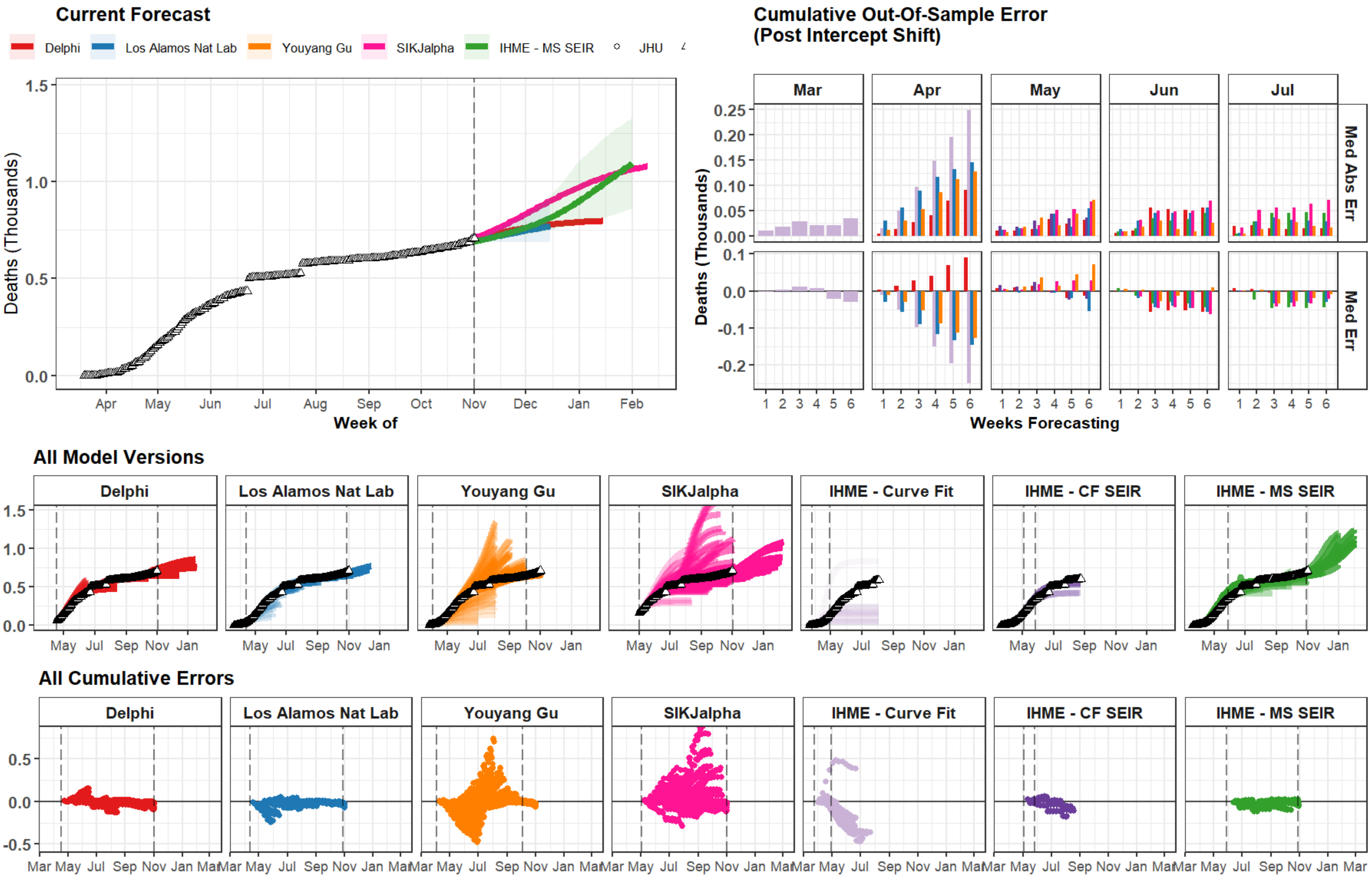
Current Forecast
Delphi
Los Alamos Nat Lab
Youyang Gu
SIKJalpha
IHME - MS SEIR
JHU
Deaths (Thousands)
Week of
Apr May Jun Jul Aug Sep Oct Nov Dec Jan Feb
Cumulative Out-Of-Sample Error
(Post Intercept Shift)
Mar
Apr
May
Jun
Jul
Med Abs Err
Med Err
Weeks Forecasting
All Model Versions
Delphi
Los Alamos Nat Lab
Youyang Gu
SIKJalpha
IHME - Curve Fit
IHME - CF SEIR
IHME - MS SEIR
All Cumulative Errors

### Oregon

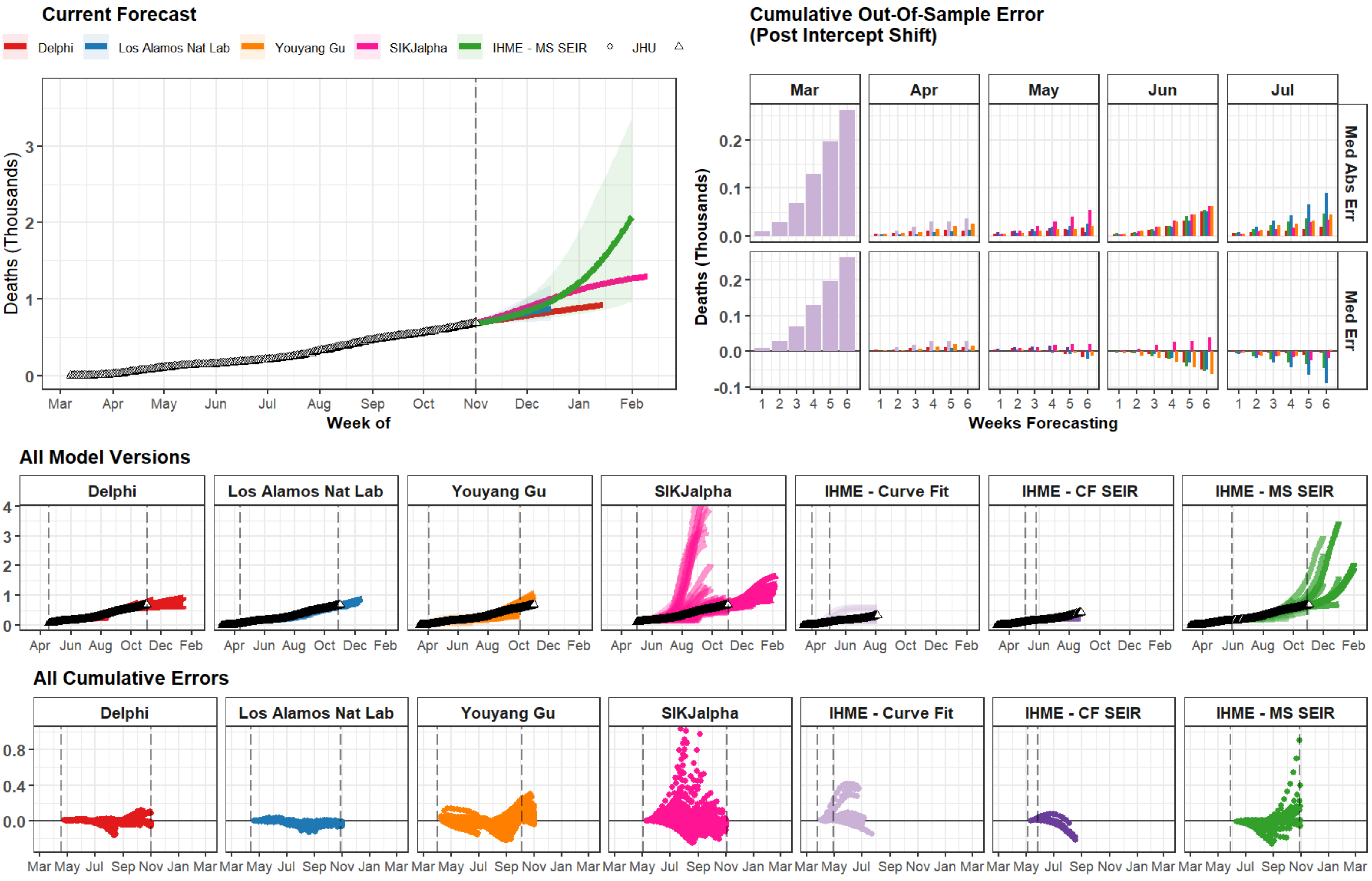

Current Forecast
Delphi
Los Alamos Nat Lab
Youyang Gu
SIKJalpha
IHME - MS SEIR
JHU
Deaths (Thousands)
Week of
Mar Apr May Jun Jul Aug Sep Oct Nov Dec Jan Feb
Cumulative Out-Of-Sample Error (Post Intercept Shift)
Mar Apr May Jun Jul
Med Abs Err
Med Err
Weeks Forecasting
All Model Versions
Delphi
Los Alamos Nat Lab
Youyang Gu
SIKJalpha
IHME - Curve Fit
IHME - CF SEIR
IHME - MS SEIR
All Cumulative Errors

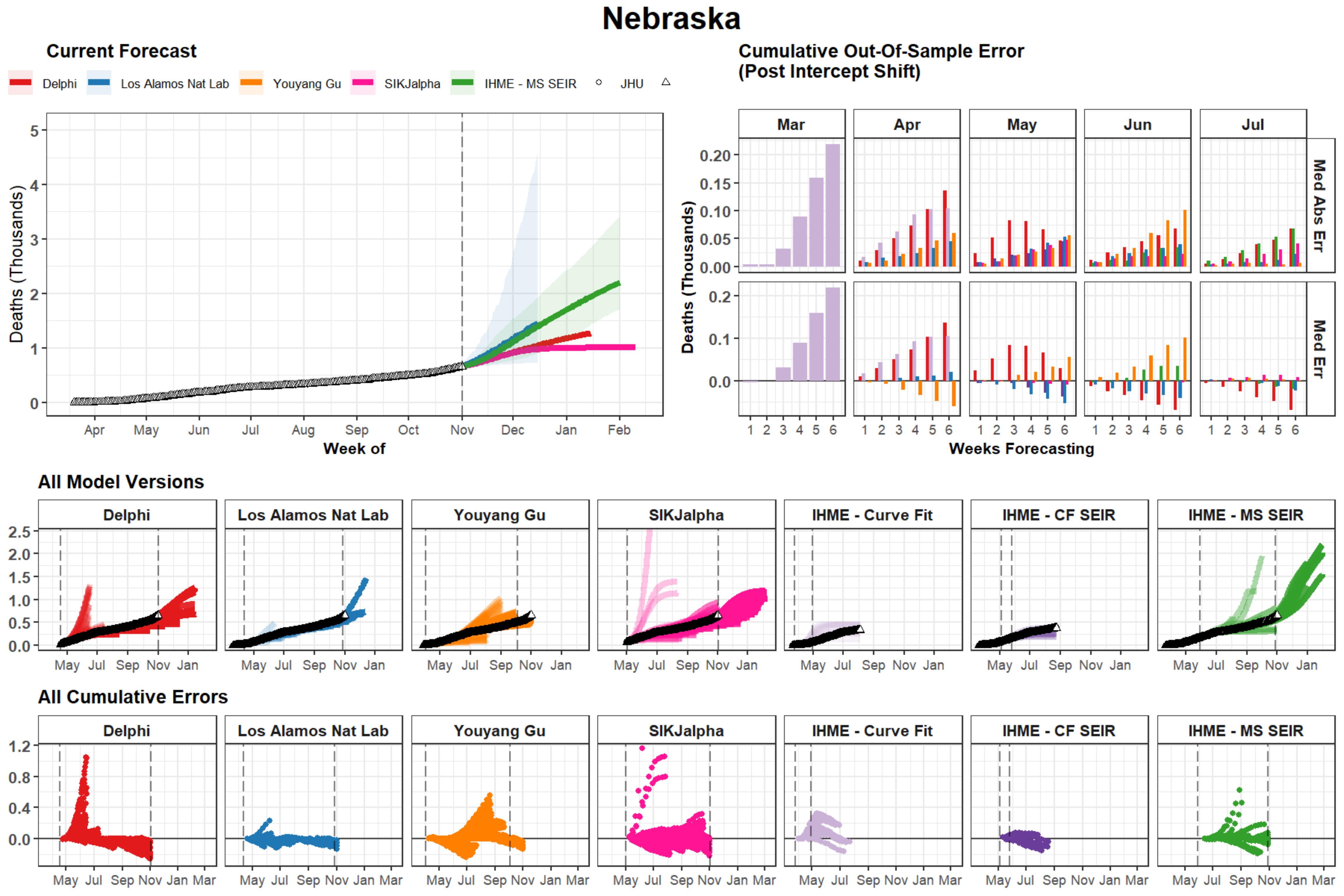
Nebraska
Current Forecast
Delphi
Los Alamos Nat Lab
Youyang Gu
SIKJalpha
IHME - MS SEIR
JHU
Deaths (Thousands)
Week of
Apr May Jun Jul Aug Sep Oct Nov Dec Jan Feb
Cumulative Out-Of-Sample Error
(Post Intercept Shift)
Mar Apr May Jun Jul
Med Abs Err
Med Err
Weeks Forecasting
All Model Versions
Delphi
Los Alamos Nat Lab
Youyang Gu
SIKJalpha
IHME - Curve Fit
IHME - CF SEIR
IHME - MS SEIR
All Cumulative Errors

### District of Columbia

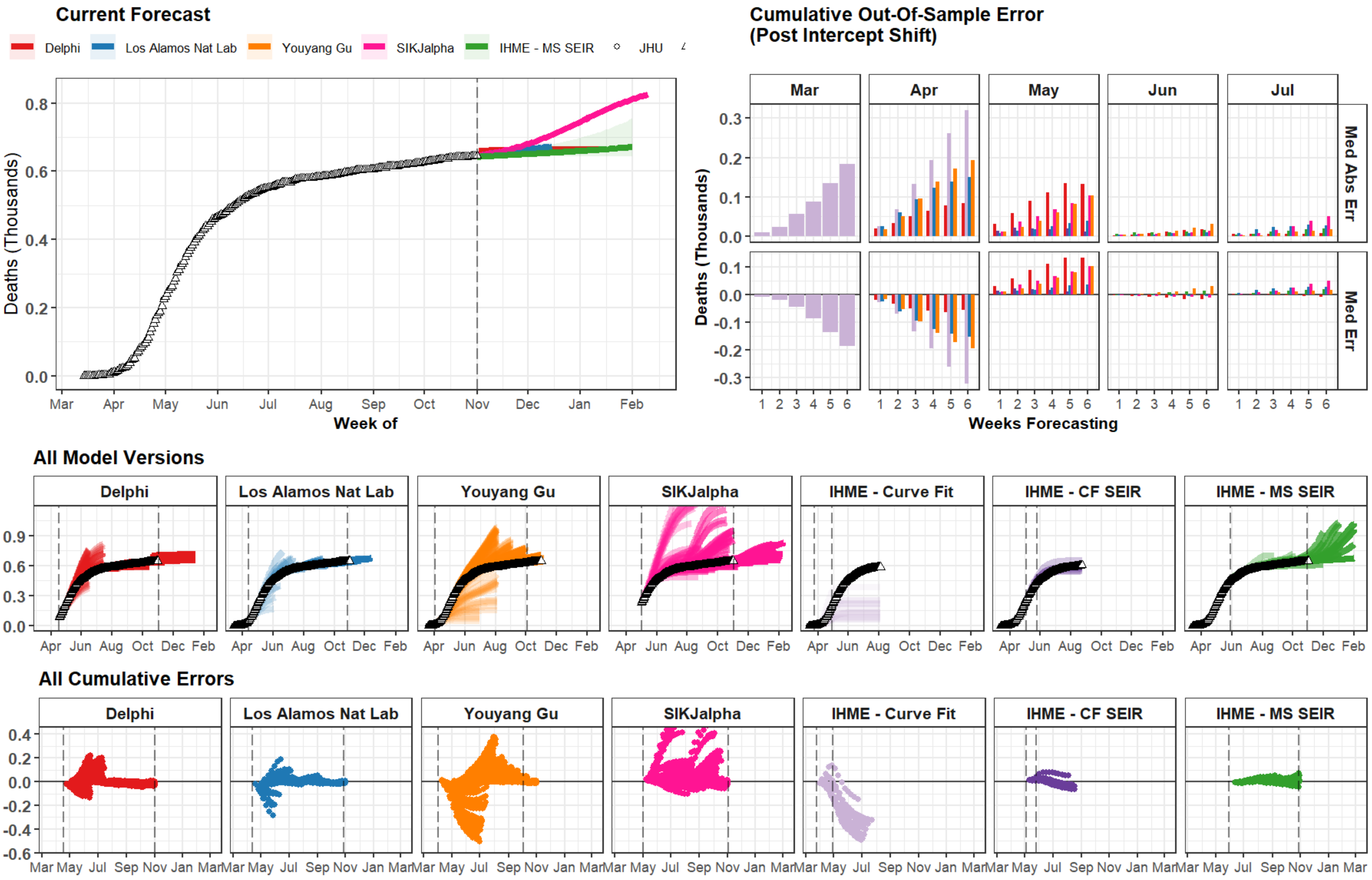

### Lebanon

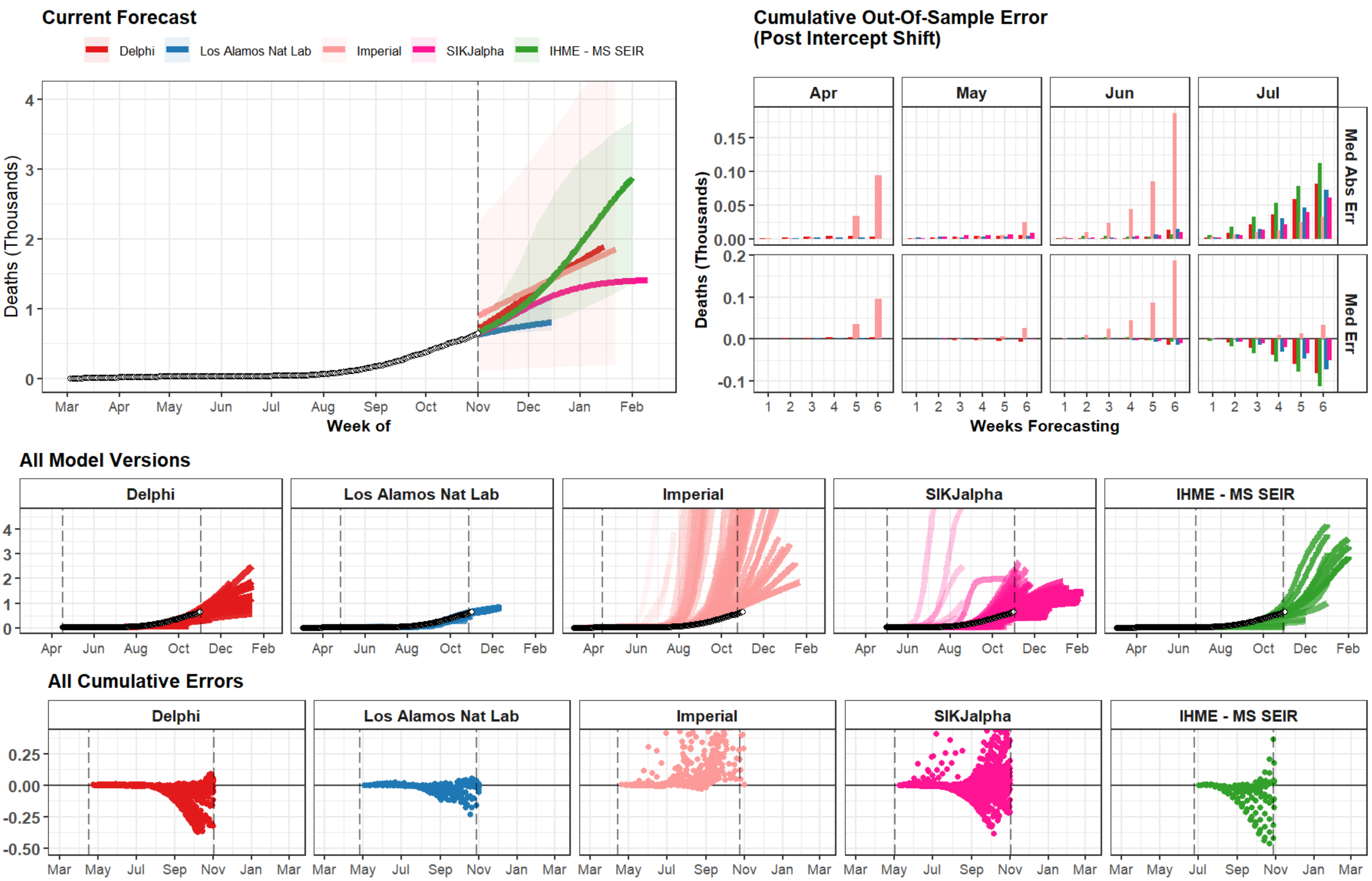

Current Forecast
Delphi
Los Alamos Nat Lab
Imperial
SIKJalpha
IHME - MS SEIR
Deaths (Thousands)
Week of
Mar
Apr
May
Jun
Jul
Aug
Sep
Oct
Nov
Dec
Jan
Feb
Cumulative Out-Of-Sample Error
(Post Intercept Shift)
Apr
May
Jun
Jul
Med Abs Err
Med Err
Weeks Forecasting
All Model Versions
All Cumulative Errors

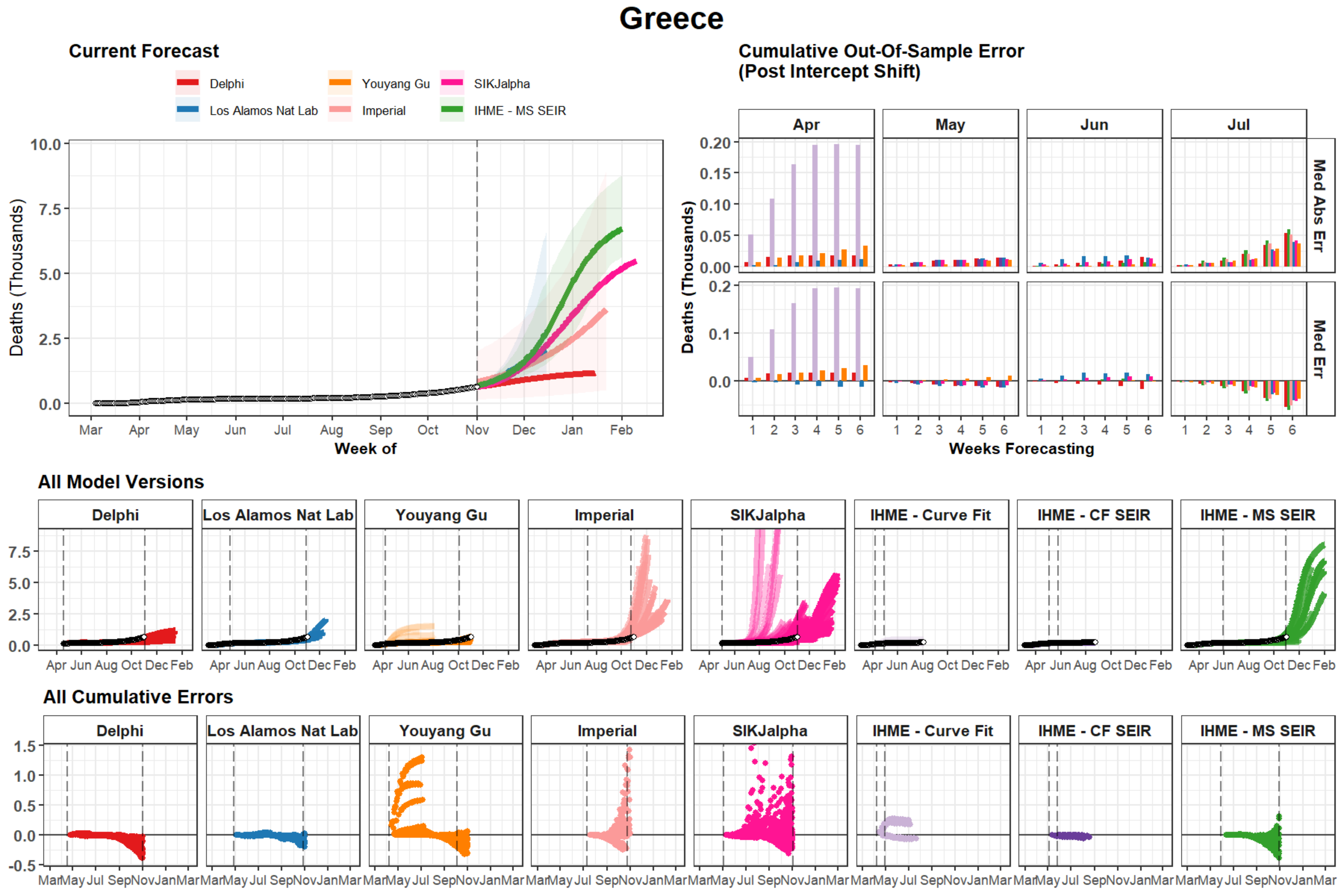

Greece
Current Forecast
Delphi
Los Alamos Nat Lab
Youyang Gu
Imperial
SIKJalpha
IHME - MS SEIR
Deaths (Thousands)
Week of
Cumulative Out-Of-Sample Error
(Post Intercept Shift)
Apr
May
Jun
Jul
Med Abs Err
Med Err
Weeks Forecasting
All Model Versions
Delphi
Los Alamos Nat Lab
Youyang Gu
Imperial
SIKJalpha
IHME - Curve Fit
IHME - CF SEIR
IHME - MS SEIR
All Cumulative Errors

### Idaho

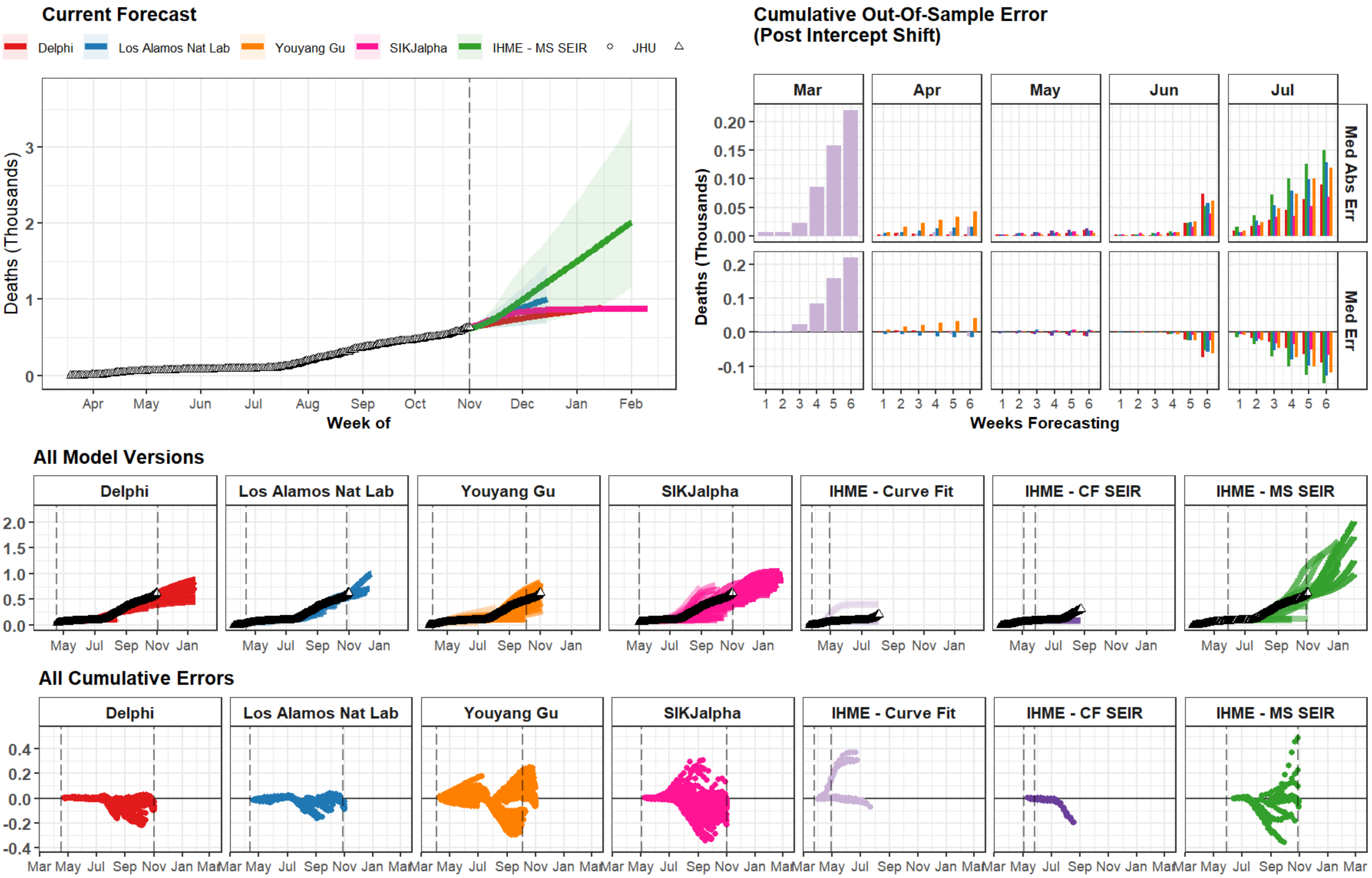
Current Forecast
Delphi
Los Alamos Nat Lab
Youyang Gu
SIKJalpha
IHME - MS SEIR
JHU
Deaths (Thousands)
Week of
Apr May Jun Jul Aug Sep Oct Nov Dec Jan Feb
Cumulative Out-Of-Sample Error
(Post Intercept Shift)
Mar Apr May Jun Jul
Med Abs Err
Med Err
Weeks Forecasting
All Model Versions
Delphi
Los Alamos Nat Lab
Youyang Gu
SIKJalpha
IHME - Curve Fit
IHME - CF SEIR
IHME - MS SEIR
All Cumulative Errors

### Utah

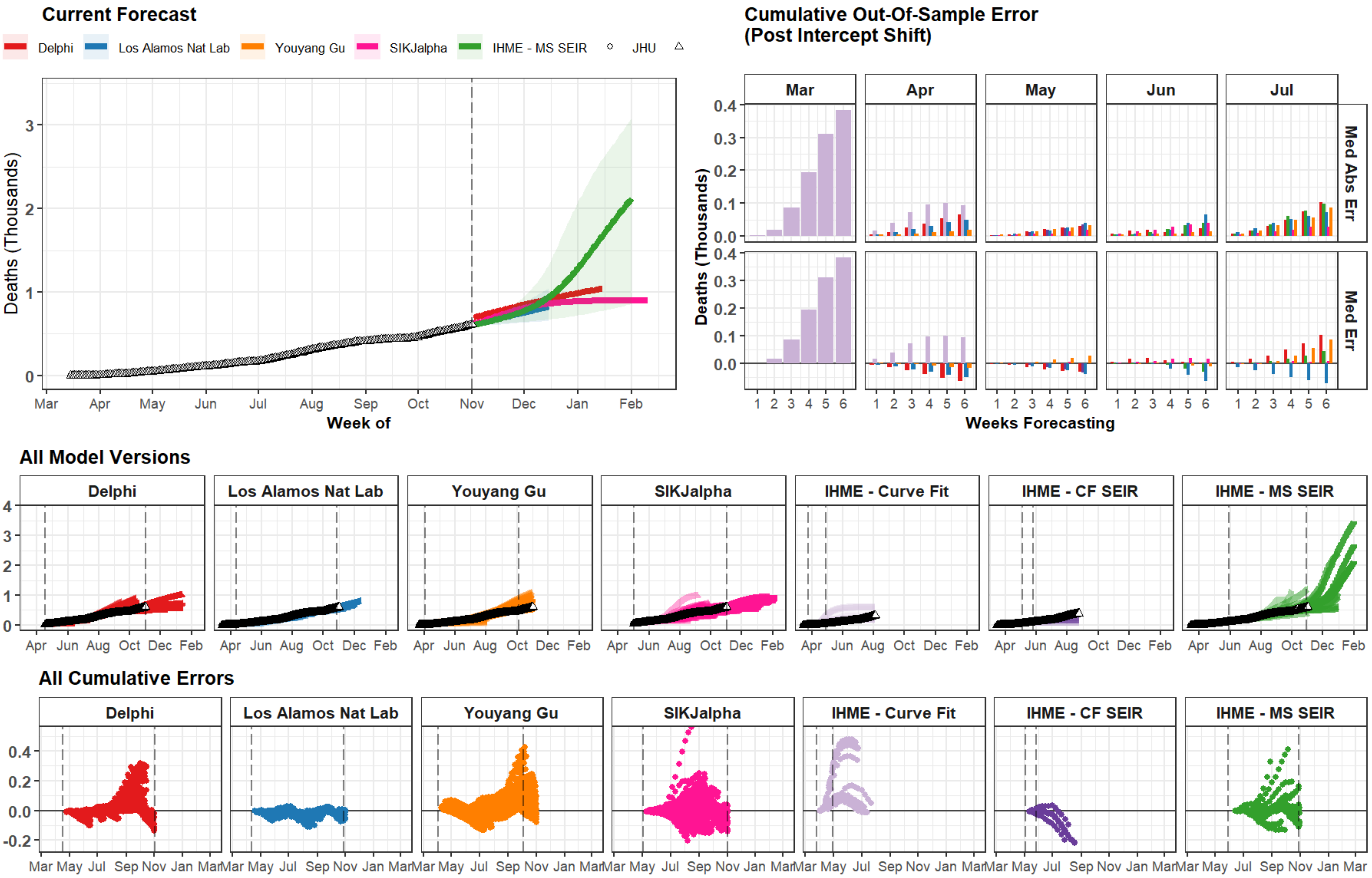

Current Forecast
Delphi
Los Alamos Nat Lab
Youyang Gu
SIKJalpha
IHME - MS SEIR
JHU
Deaths (Thousands)
0
1
2
3
Mar
Apr
May
Jun
Jul
Aug
Sep
Oct
Nov
Dec
Jan
Feb
Week of
Cumulative Out-Of-Sample Error
(Post Intercept Shift)
Mar
Apr
May
Jun
Jul
Med Abs Err
Med Err
0.0
0.1
0.2
0.3
0.4
1 2 3 4 5 6
Weeks Forecasting
All Model Versions
Delphi
Los Alamos Nat Lab
Youyang Gu
SIKJalpha
IHME - Curve Fit
IHME - CF SEIR
IHME - MS SEIR
0
1
2
3
4
Apr Jun Aug Oct Dec Feb
All Cumulative Errors
-0.2
0.0
0.2
0.4
Mar May Jul Sep Nov Jan Mar

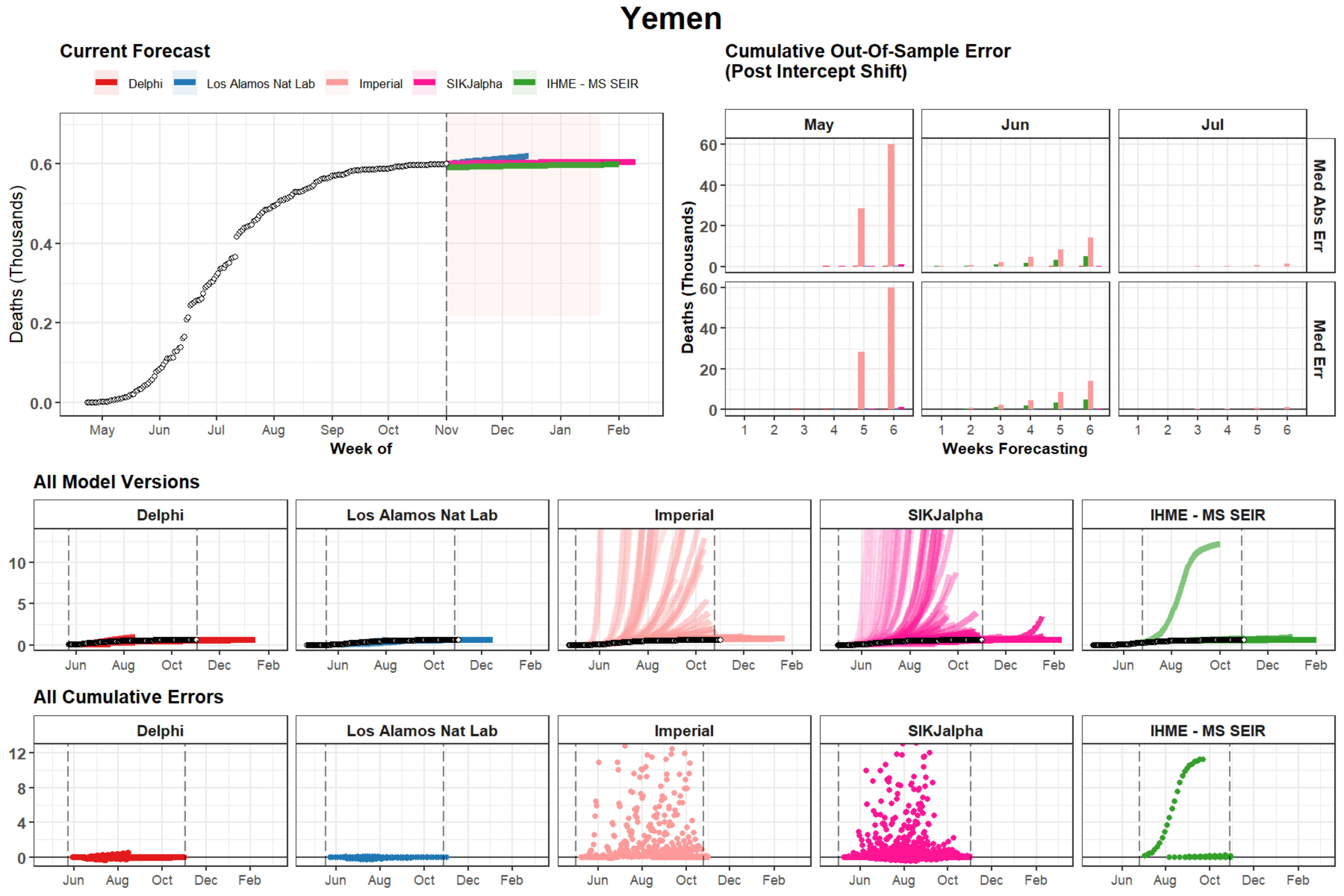
Yemen
Current Forecast
Delphi
Los Alamos Nat Lab
Imperial
SIKJalpha
IHME - MS SEIR
Deaths (Thousands)
Week of
May
Jun
Jul
Aug
Sep
Oct
Nov
Dec
Jan
Feb
Cumulative Out-Of-Sample Error
(Post Intercept Shift)
Med Abs Err
Med Err
Weeks Forecasting
All Model Versions
All Cumulative Errors

### Uzbekistan

#### Current Forecast

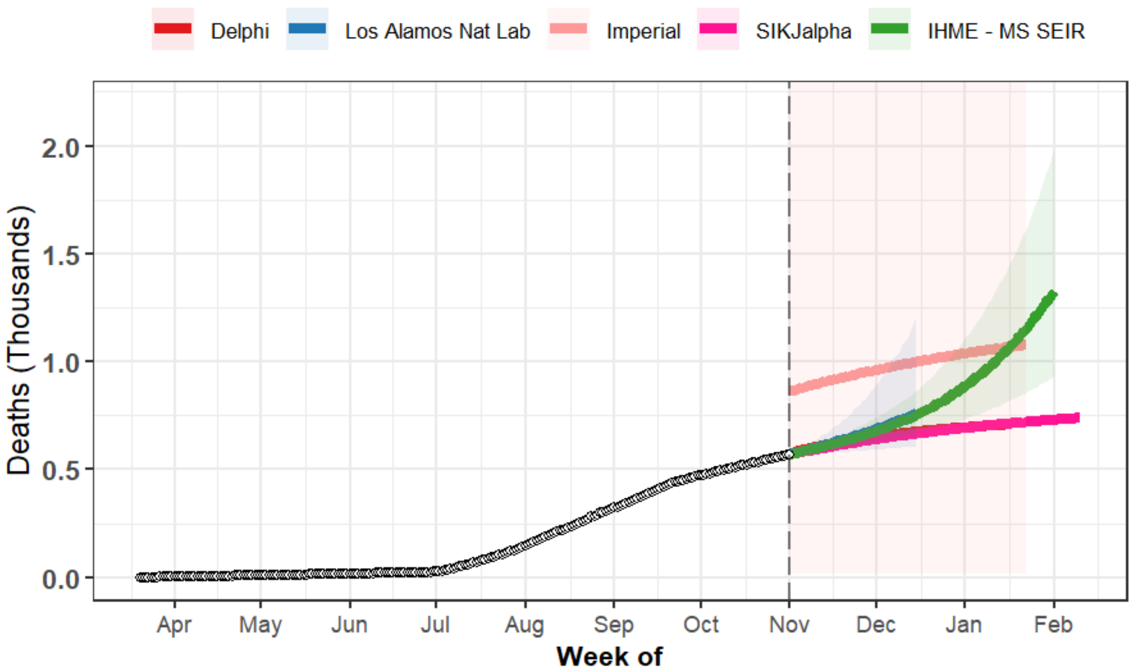


#### Cumulative Out-Of-Sample Error (Post Intercept Shift)

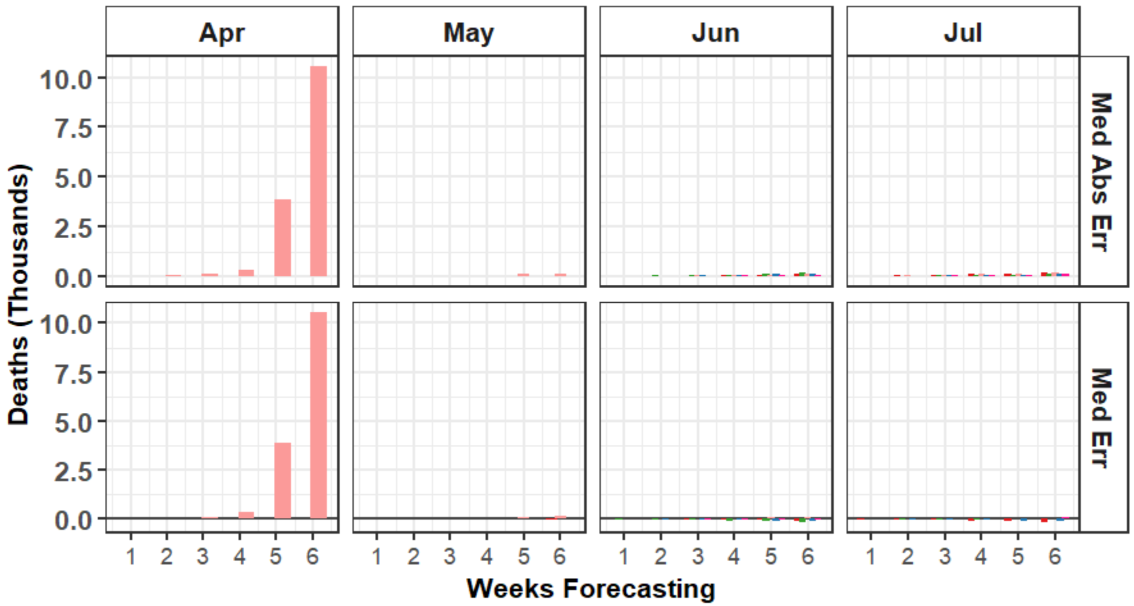


#### All Model Versions

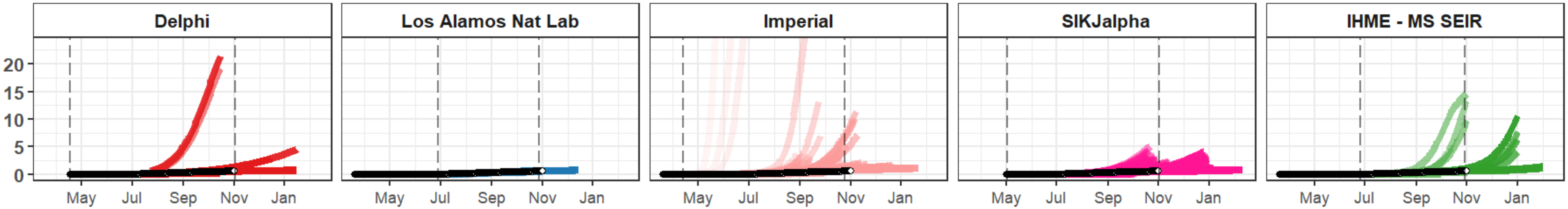


#### All Cumulative Errors

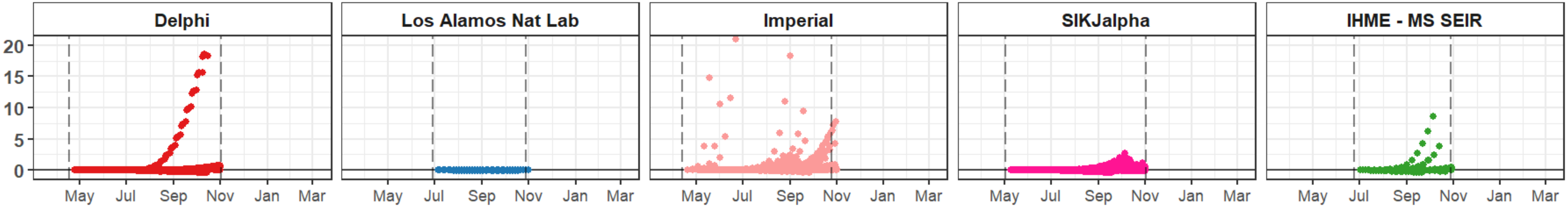

### Croatia

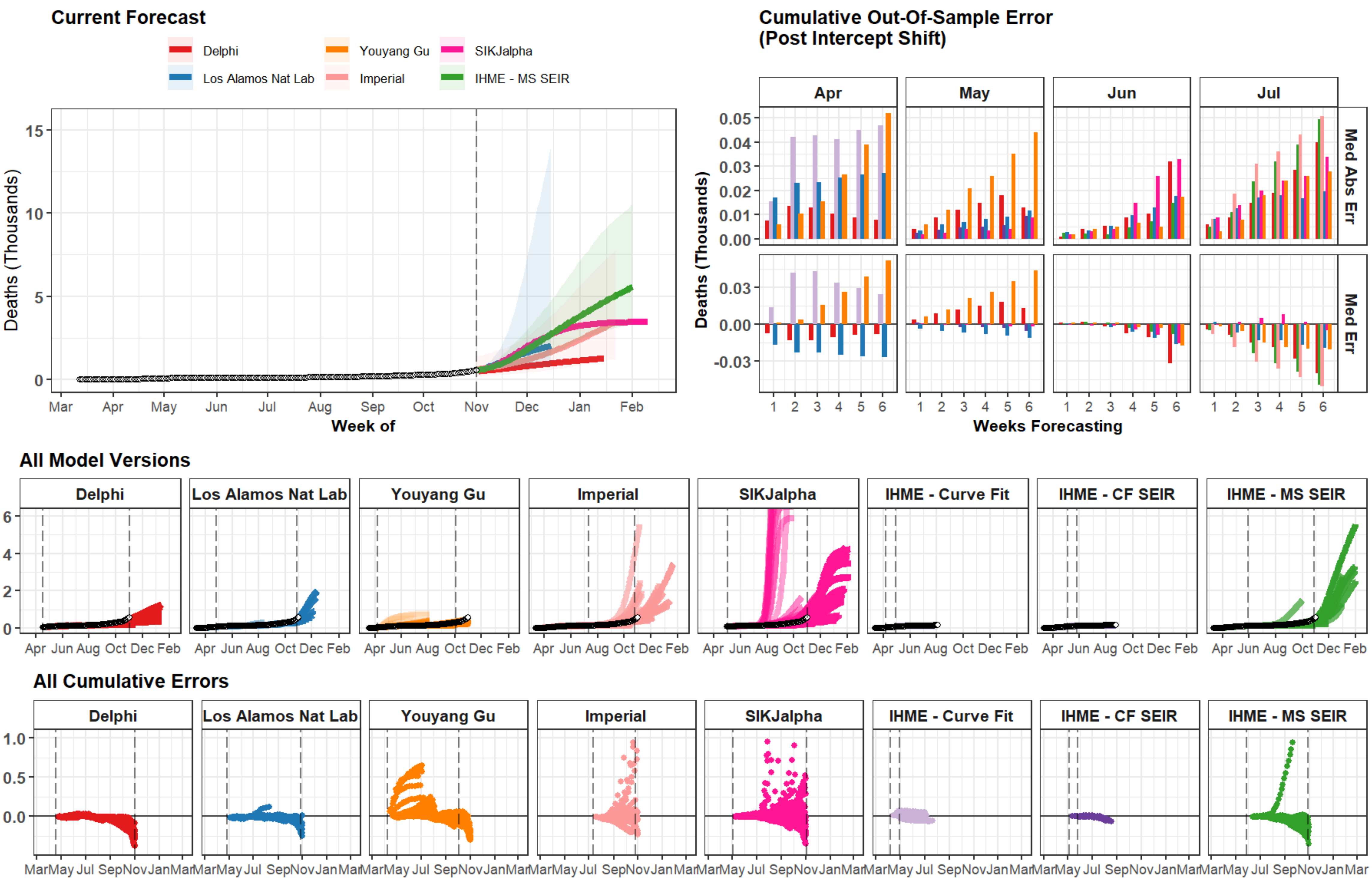
Current Forecast
Delphi
Los Alamos Nat Lab
Youyang Gu
Imperial
SIKJalpha
IHME - MS SEIR
Deaths (Thousands)
Week of
Mar Apr May Jun Jul Aug Sep Oct Nov Dec Jan Feb
Cumulative Out-Of-Sample Error
(Post Intercept Shift)
Apr May Jun Jul
Med Abs Err
Med Err
Weeks Forecasting
All Model Versions
Delphi
Los Alamos Nat Lab
Youyang Gu
Imperial
SIKJalpha
IHME - Curve Fit
IHME - CF SEIR
IHME - MS SEIR
Apr Jun Aug Oct Dec Feb
All Cumulative Errors
MarMay Jul SepNovJanMar

### North Dakota

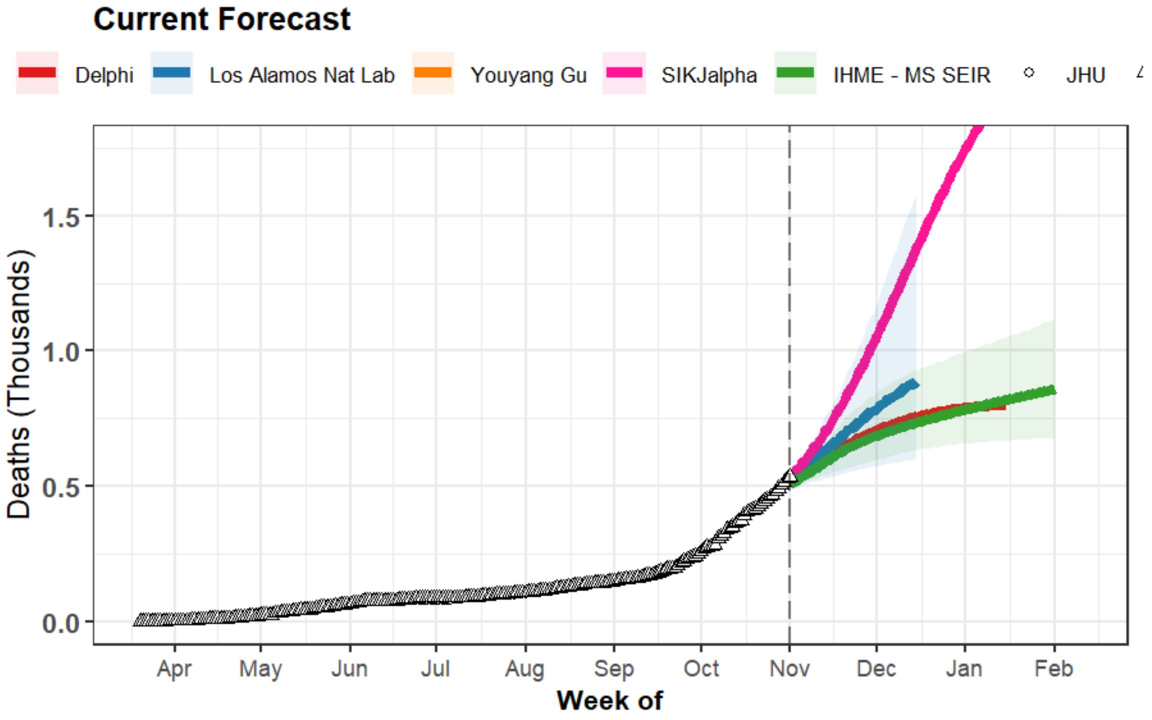


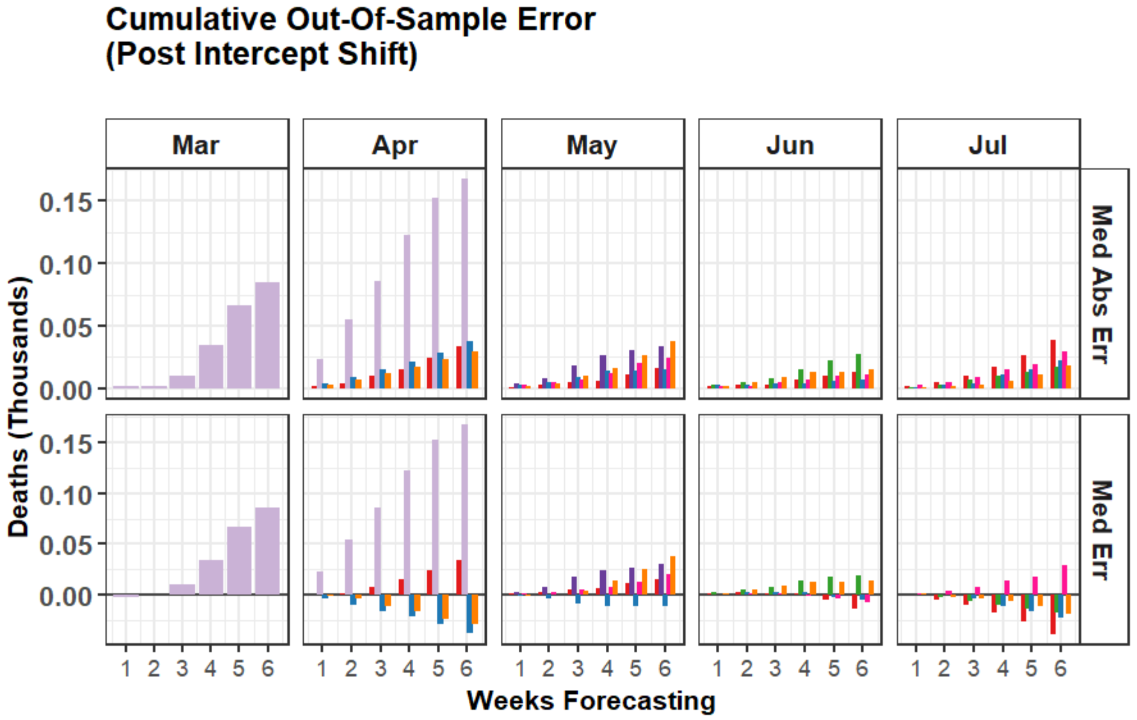


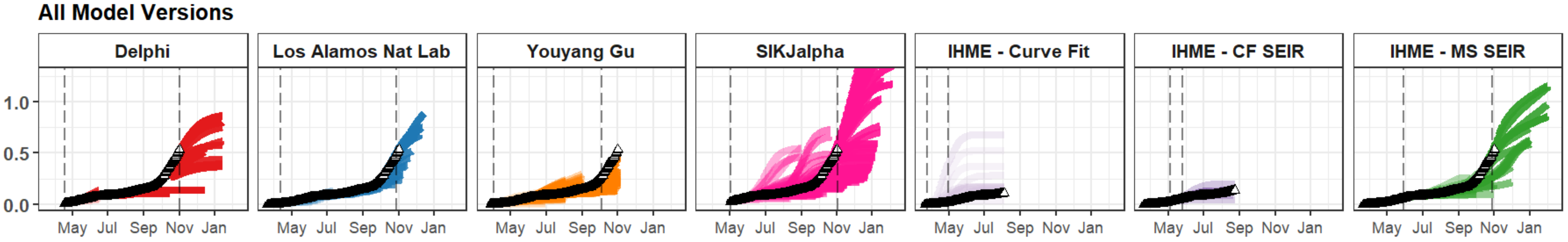


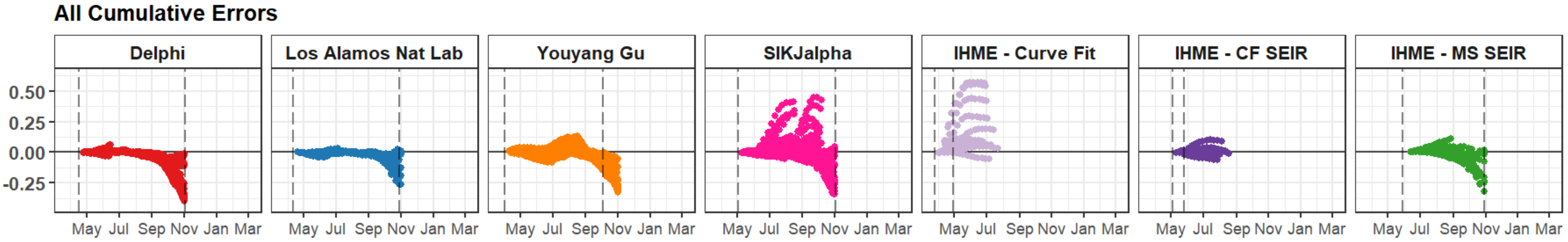

### Albania

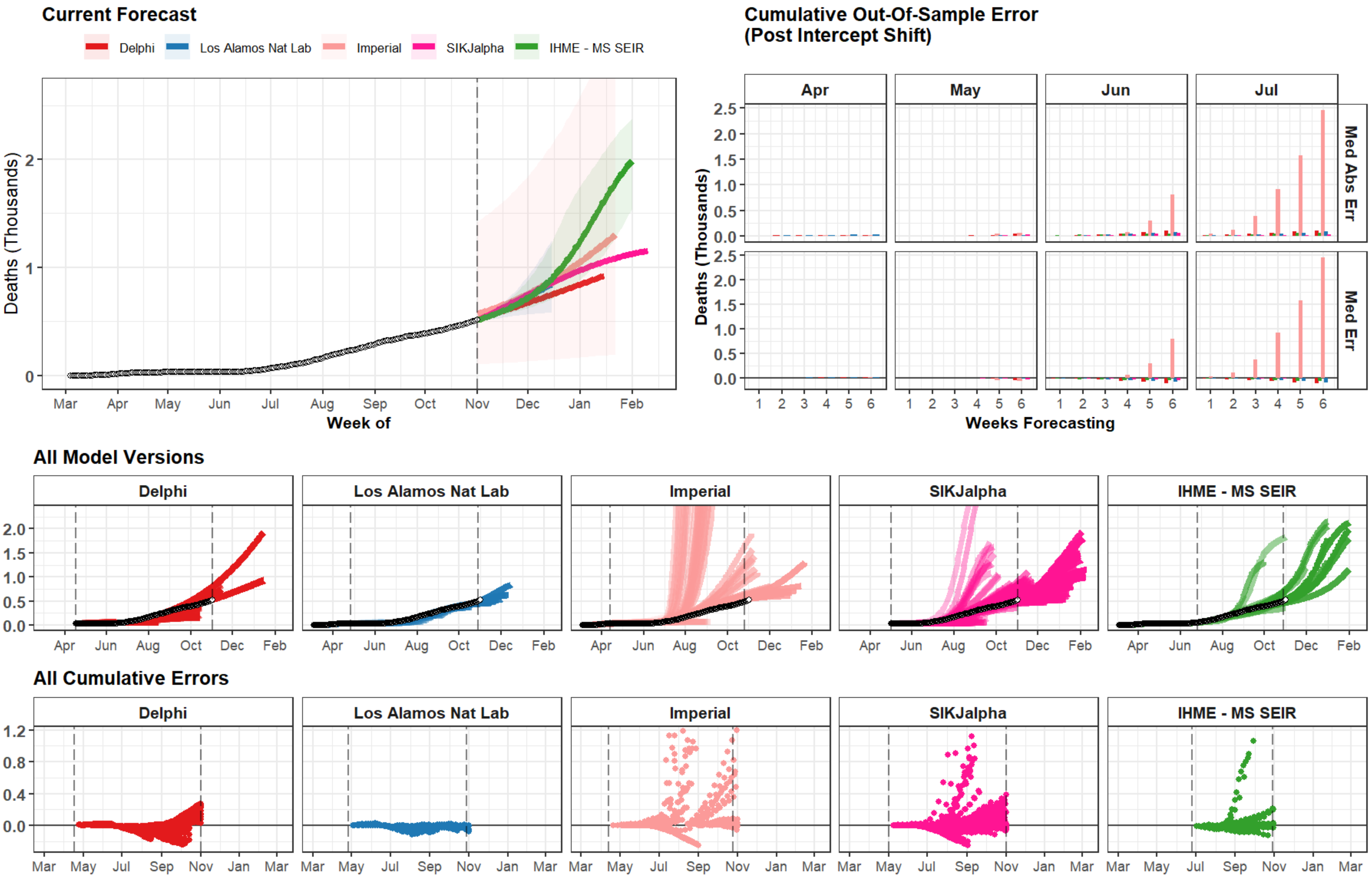

Current Forecast
Delphi
Los Alamos Nat Lab
Imperial
SIKJalpha
IHME - MS SEIR
Deaths (Thousands)
0
1
2
Mar
Apr
May
Jun
Jul
Aug
Sep
Oct
Nov
Dec
Jan
Feb
Week of
Cumulative Out-Of-Sample Error
(Post Intercept Shift)
Apr
May
Jun
Jul
Med Abs Err
Med Err
0.0
0.5
1.0
1.5
2.0
2.5
1
2
3
4
5
6
Weeks Forecasting
All Model Versions
Delphi
Los Alamos Nat Lab
Imperial
SIKJalpha
IHME - MS SEIR
0.0
0.5
1.0
1.5
2.0
Apr
Jun
Aug
Oct
Dec
Feb
All Cumulative Errors
Delphi
Los Alamos Nat Lab
Imperial
SIKJalpha
IHME - MS SEIR
1.2
0.8
0.4
0.0
Mar
May
Jul
Sep
Nov
Jan
Mar

### United Arab Emirates

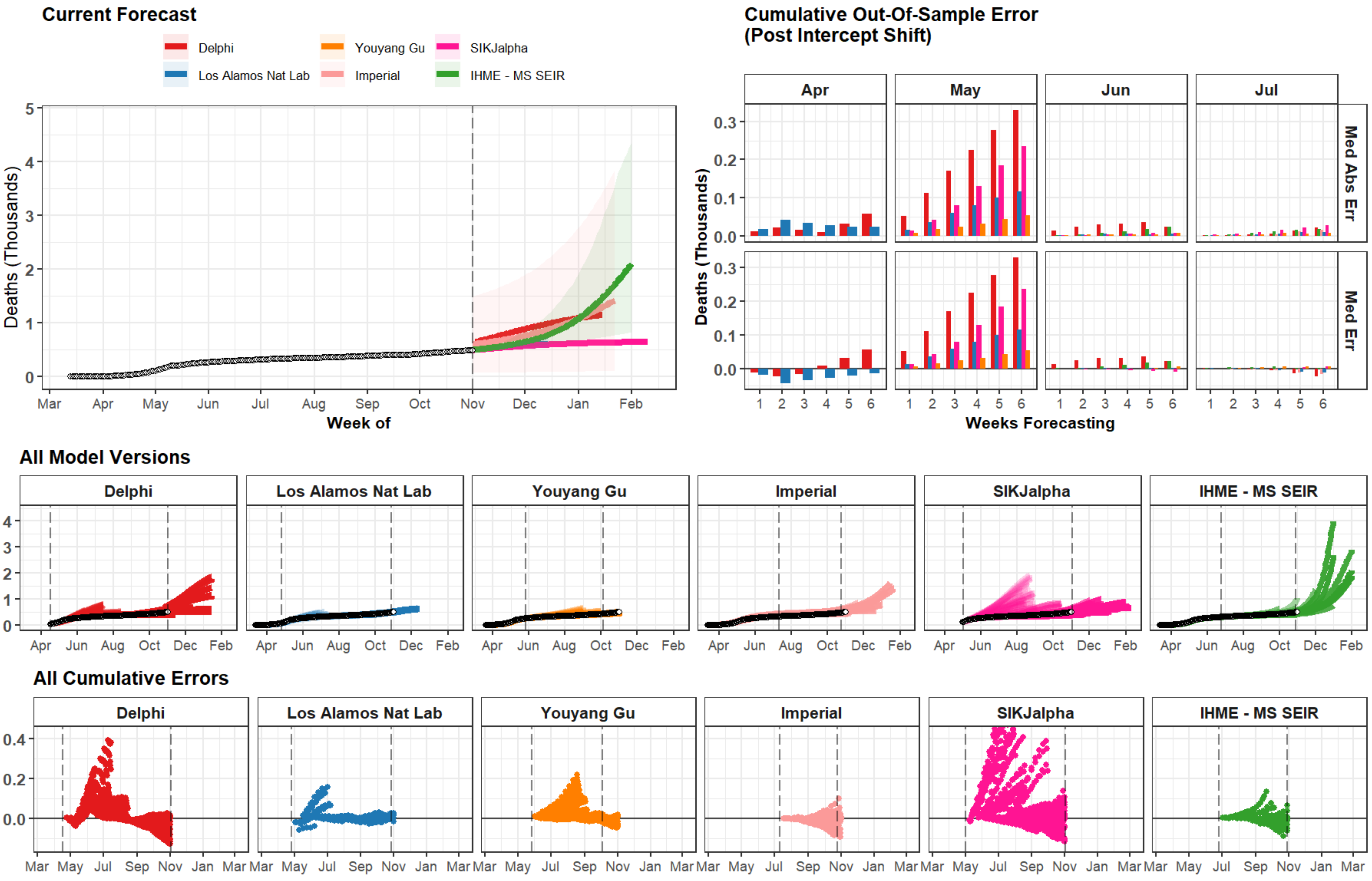

### New Hampshire

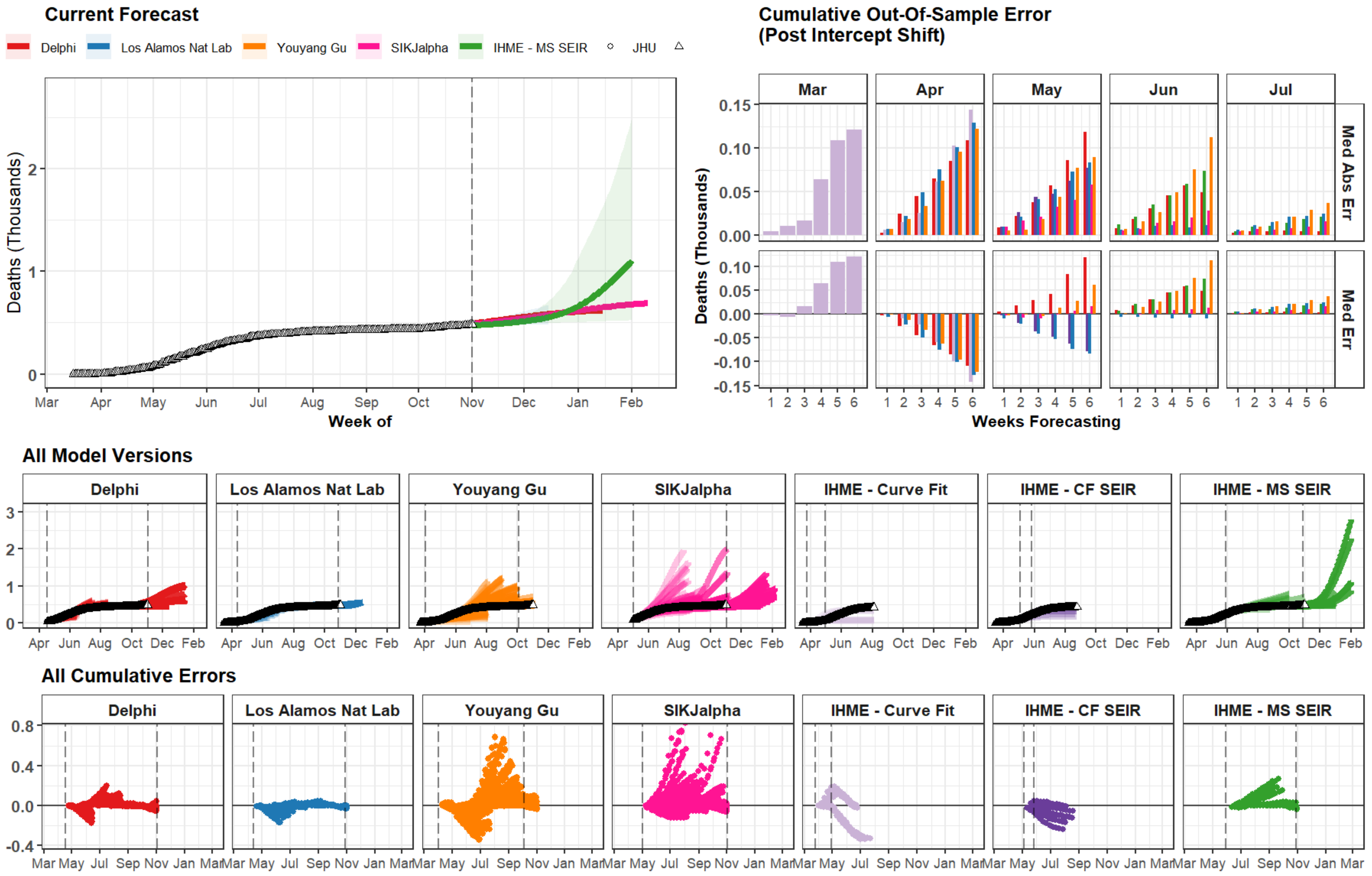

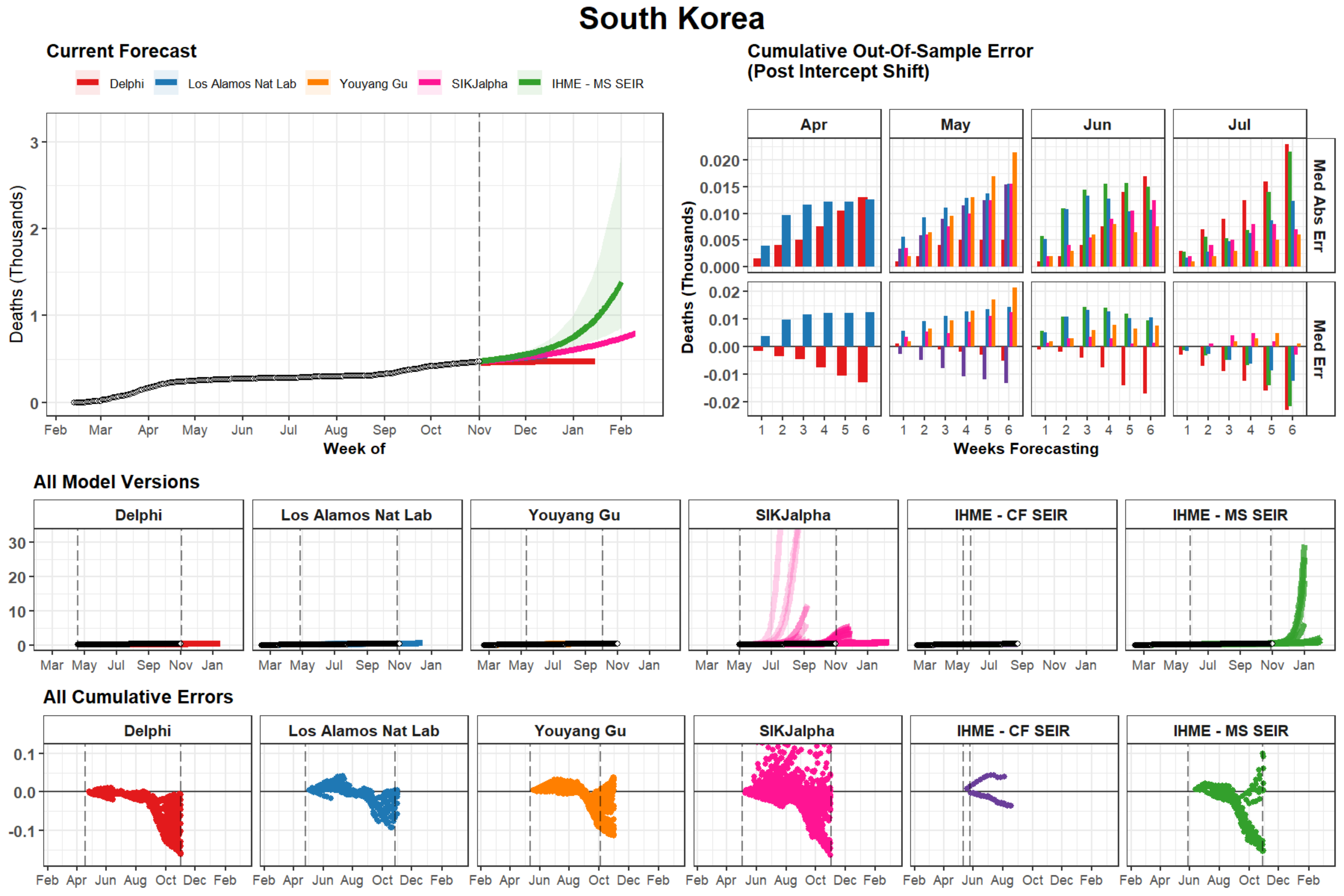
South Korea
Current Forecast
Delphi
Los Alamos Nat Lab
Youyang Gu
SIKJalpha
IHME - MS SEIR
Deaths (Thousands)
Week of
Feb Mar Apr May Jun Jul Aug Sep Oct Nov Dec Jan Feb
Cumulative Out-Of-Sample Error
(Post Intercept Shift)
Apr
May
Jun
Jul
Med Abs Err
Med Err
Weeks Forecasting
All Model Versions
Delphi
Los Alamos Nat Lab
Youyang Gu
SIKJalpha
IHME - CF SEIR
IHME - MS SEIR
Mar May Jul Sep Nov Jan
All Cumulative Errors
Feb Apr Jun Aug Oct Dec Feb

### West Virginia

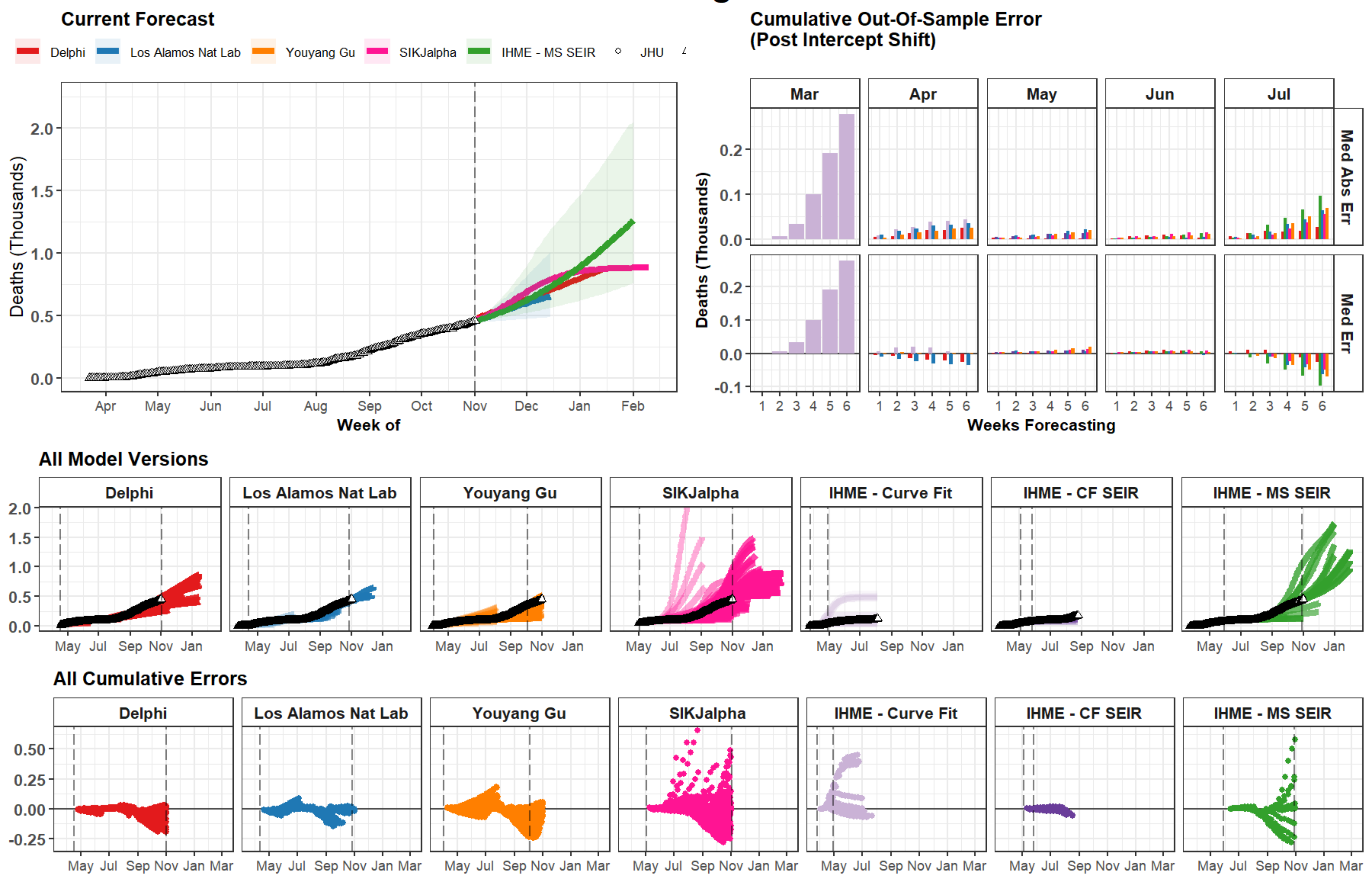

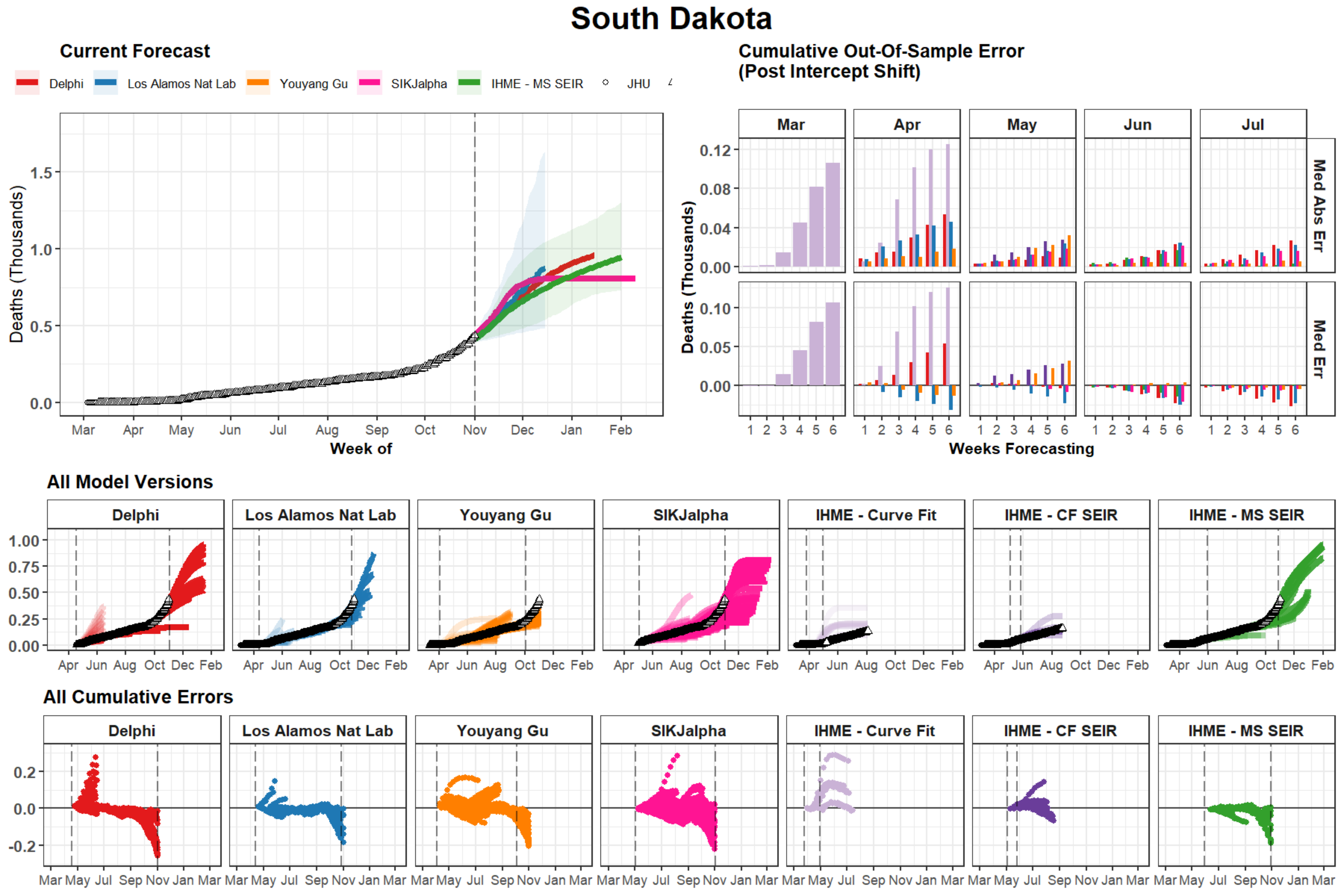

South Dakota
Current Forecast
Delphi
Los Alamos Nat Lab
Youyang Gu
SIKJalpha
IHME - MS SEIR
JHU
Deaths (Thousands)
0.0
0.5
1.0
1.5
Mar
Apr
May
Jun
Jul
Aug
Sep
Oct
Nov
Dec
Jan
Feb
Week of
Cumulative Out-Of-Sample Error
(Post Intercept Shift)
Mar
Apr
May
Jun
Jul
Med Abs Err
Med Err
0.00
0.04
0.08
0.12
0.05
0.10
1 2 3 4 5 6
Weeks Forecasting
All Model Versions
Delphi
Los Alamos Nat Lab
Youyang Gu
SIKJalpha
IHME - Curve Fit
IHME - CF SEIR
IHME - MS SEIR
0.00
0.25
0.50
0.75
1.00
Apr Jun Aug Oct Dec Feb
All Cumulative Errors
-0.2
0.0
0.2
Mar May Jul Sep Nov Jan Mar

### Cameroon

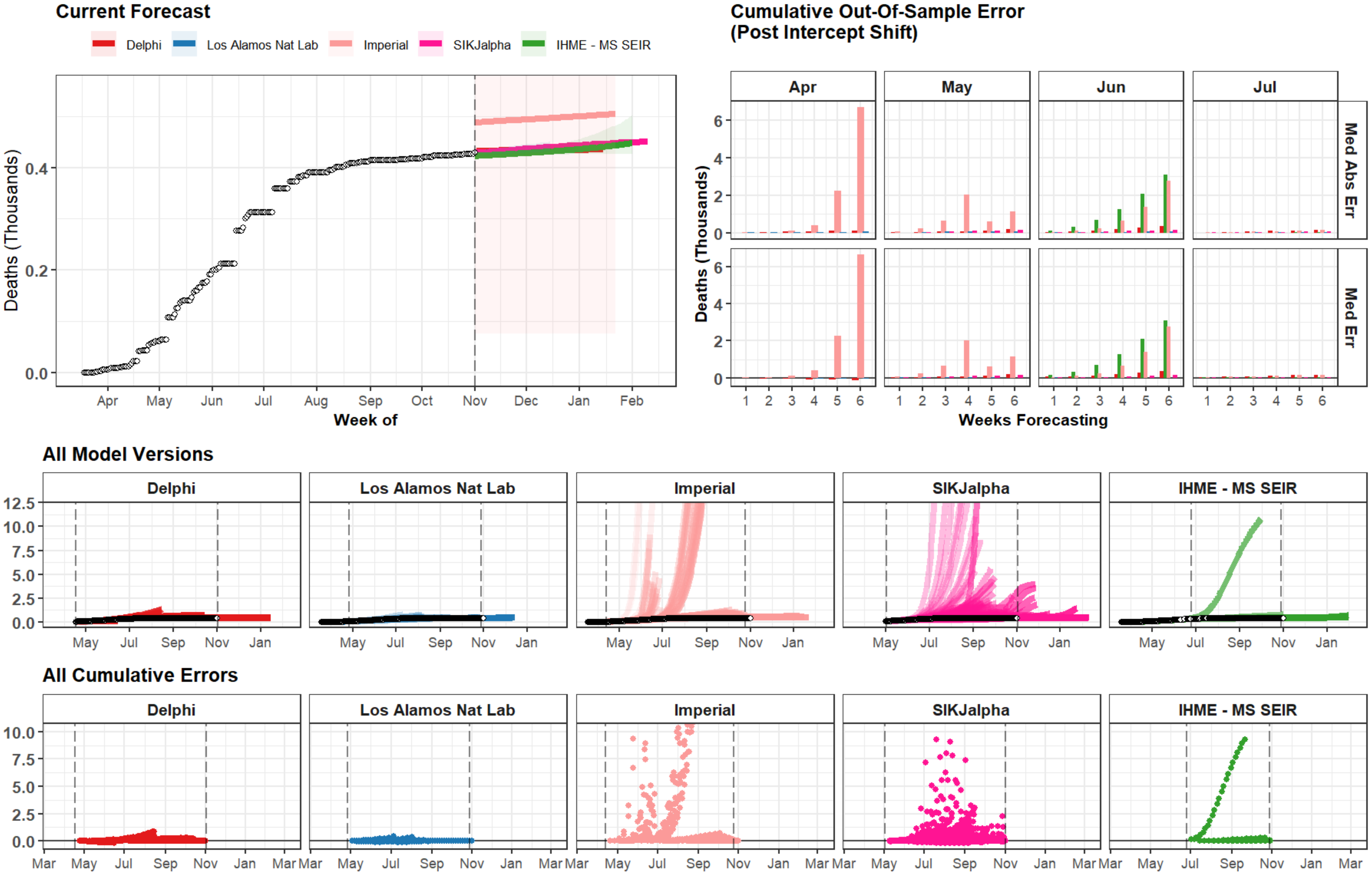
Current Forecast
Delphi
Los Alamos Nat Lab
Imperial
SIKJalpha
IHME - MS SEIR
Deaths (Thousands)
Week of
Apr
May
Jun
Jul
Aug
Sep
Oct
Nov
Dec
Jan
Feb
Cumulative Out-Of-Sample Error
(Post Intercept Shift)
Med Abs Err
Med Err
Weeks Forecasting
All Model Versions
All Cumulative Errors

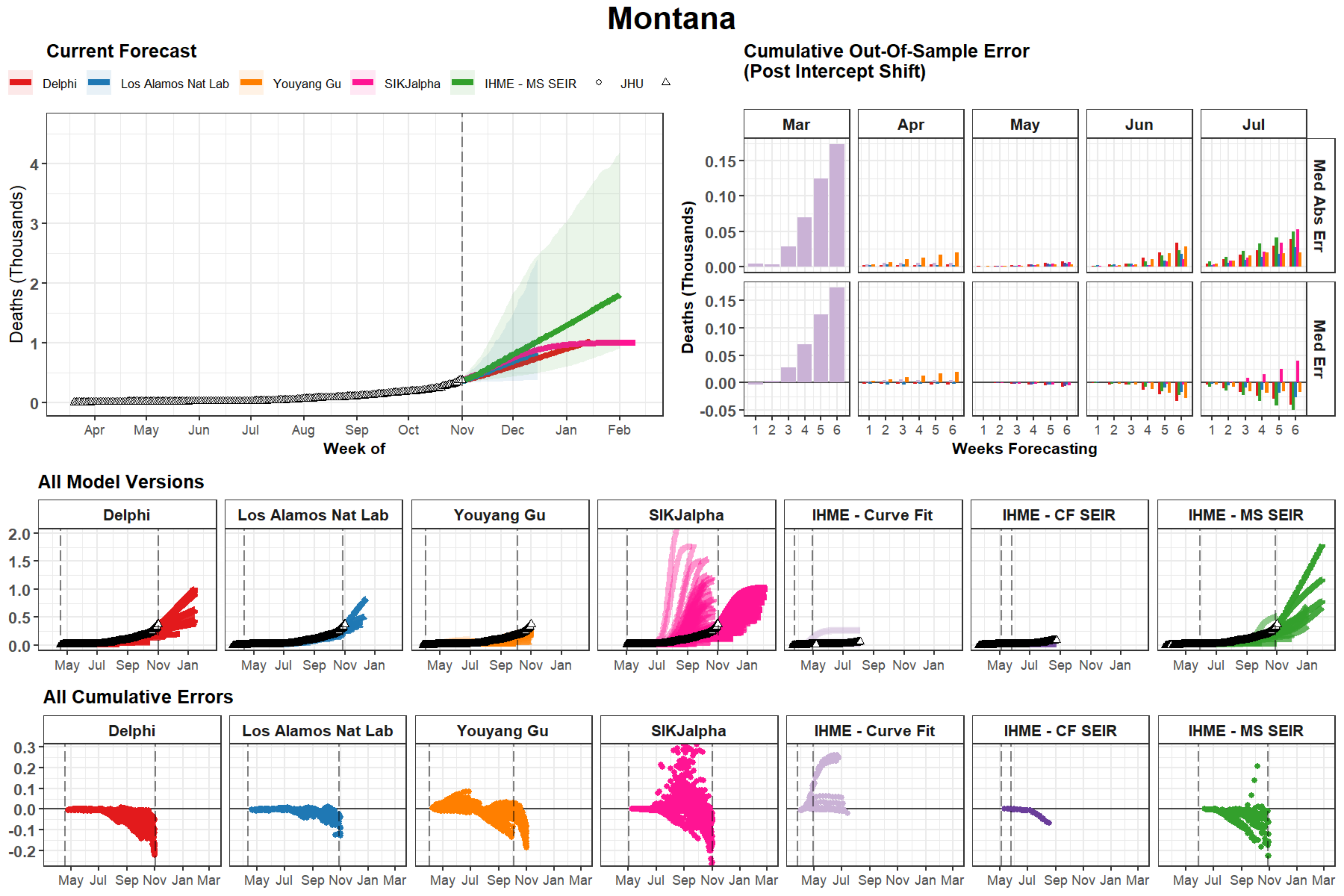
Montana
Current Forecast
Delphi
Los Alamos Nat Lab
Youyang Gu
SIKJalpha
IHME - MS SEIR
JHU
Deaths (Thousands)
Week of
Apr May Jun Jul Aug Sep Oct Nov Dec Jan Feb
Cumulative Out-Of-Sample Error
(Post Intercept Shift)
Mar Apr May Jun Jul
Med Abs Err
Med Err
Weeks Forecasting
All Model Versions
Delphi
Los Alamos Nat Lab
Youyang Gu
SIKJalpha
IHME - Curve Fit
IHME - CF SEIR
IHME - MS SEIR
May Jul Sep Nov Jan
All Cumulative Errors
May Jul Sep Nov Jan Mar

### Slovenia

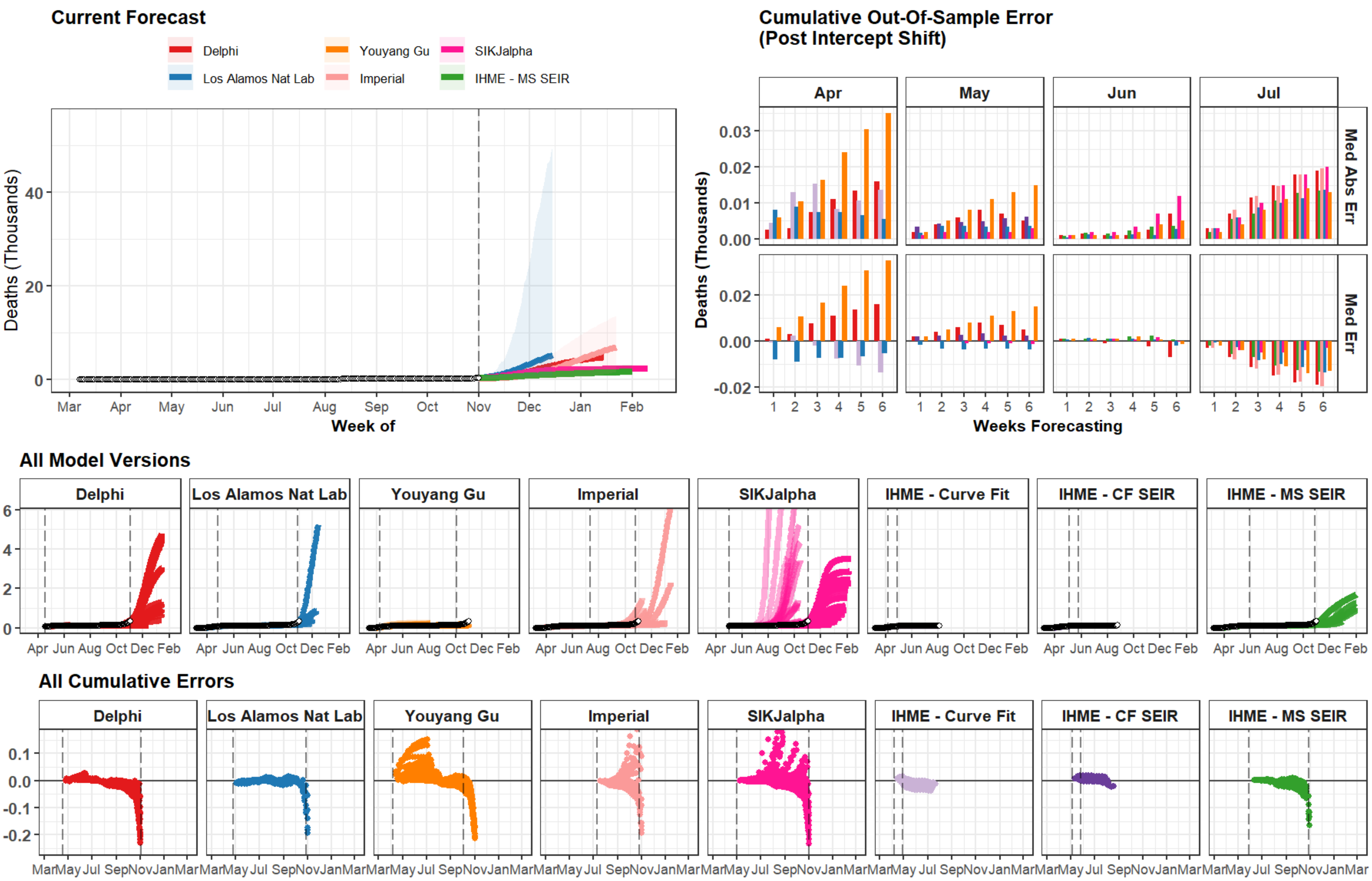
Current Forecast
Delphi
Youyang Gu
SIKJalpha
Los Alamos Nat Lab
Imperial
IHME - MS SEIR
Deaths (Thousands)
Week of
Mar Apr May Jun Jul Aug Sep Oct Nov Dec Jan Feb
Cumulative Out-Of-Sample Error (Post Intercept Shift)
Apr May Jun Jul
Med Abs Err
Med Err
Weeks Forecasting
All Model Versions
Delphi
Los Alamos Nat Lab
Youyang Gu
Imperial
SIKJalpha
IHME - Curve Fit
IHME - CF SEIR
IHME - MS SEIR
All Cumulative Errors

### Finland

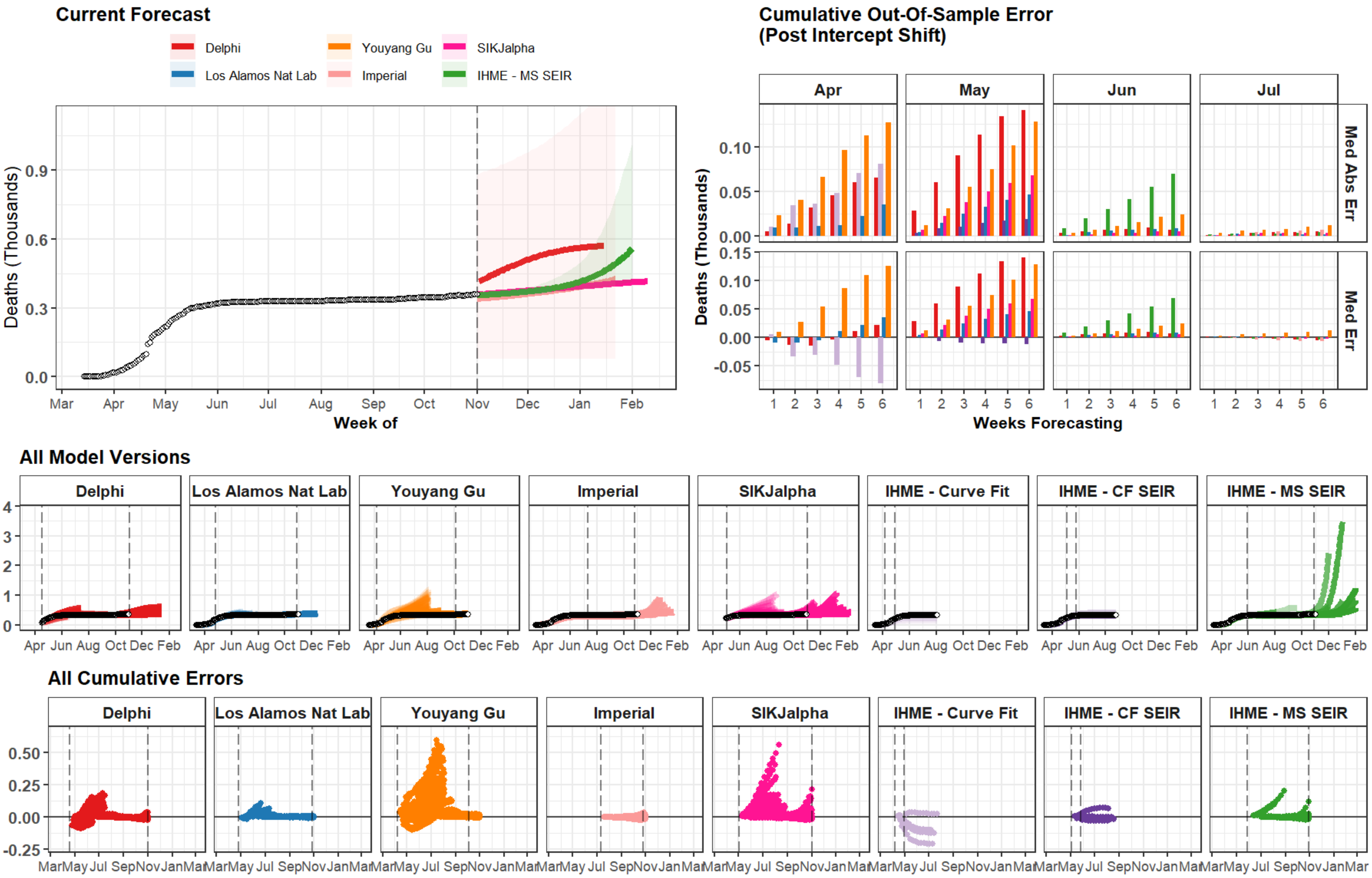

Current Forecast
Delphi
Youyang Gu
SIKJalpha
Los Alamos Nat Lab
Imperial
IHME - MS SEIR
Deaths (Thousands)
0.0
0.3
0.6
0.9
Mar Apr May Jun Jul Aug Sep Oct Nov Dec Jan Feb
Week of
Cumulative Out-Of-Sample Error
(Post Intercept Shift)
Apr
May
Jun
Jul
Med Abs Err
Med Err
0.00
0.05
0.10
0.15
-0.05
1 2 3 4 5 6
Weeks Forecasting
All Model Versions
Delphi
Los Alamos Nat Lab
Youyang Gu
Imperial
SIKJalpha
IHME - Curve Fit
IHME - CF SEIR
IHME - MS SEIR
0 1 2 3 4
Apr Jun Aug Oct Dec Feb
All Cumulative Errors
-0.25 0.00 0.25 0.50
MarMay Jul SepNovJanMar

### Zambia

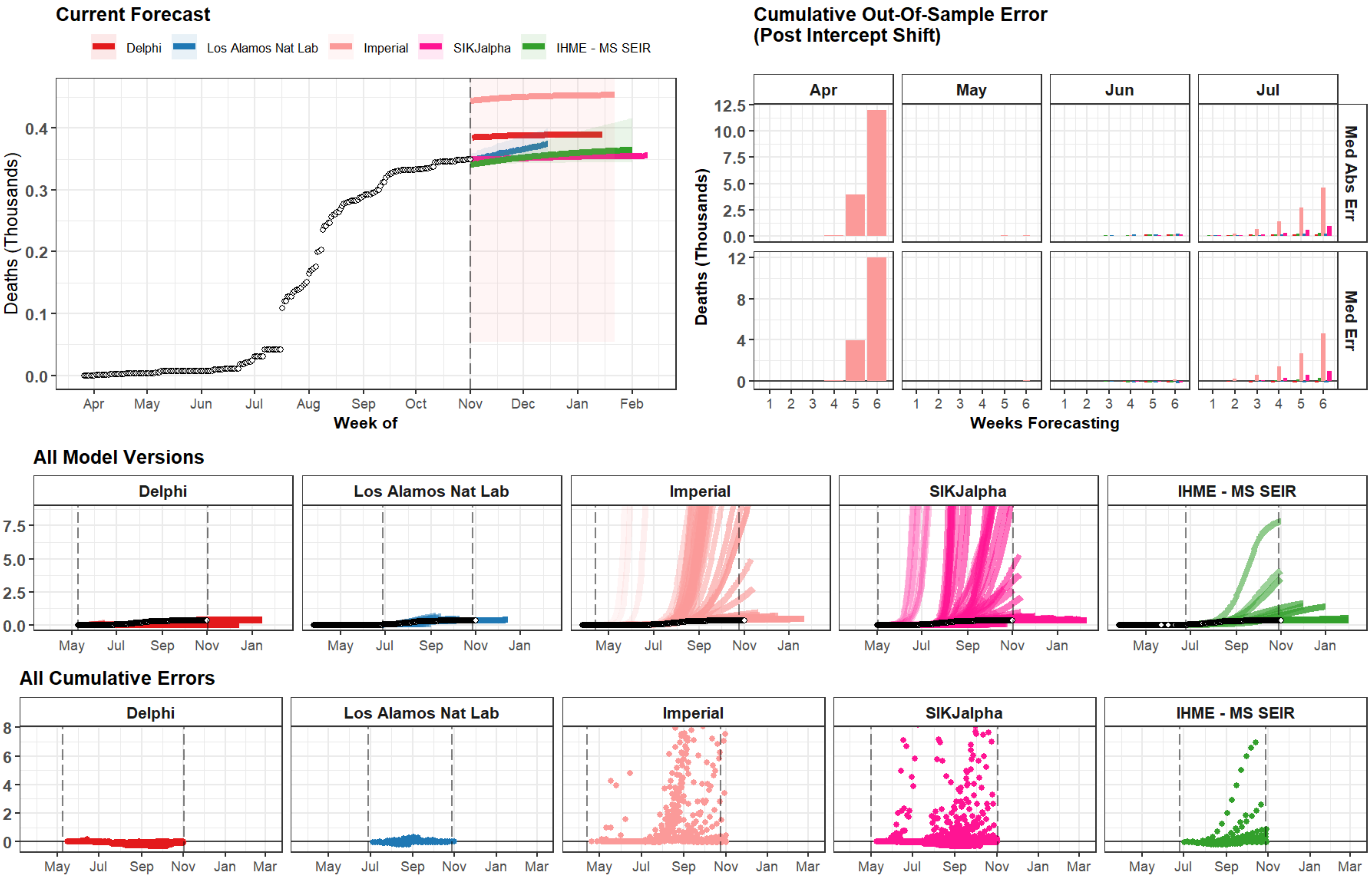
Current Forecast
Delphi
Los Alamos Nat Lab
Imperial
SIKJalpha
IHME - MS SEIR
Deaths (Thousands)
Week of
Apr
May
Jun
Jul
Aug
Sep
Oct
Nov
Dec
Jan
Feb
Cumulative Out-Of-Sample Error
(Post Intercept Shift)
Med Abs Err
Med Err
Weeks Forecasting
All Model Versions
All Cumulative Errors
Mar

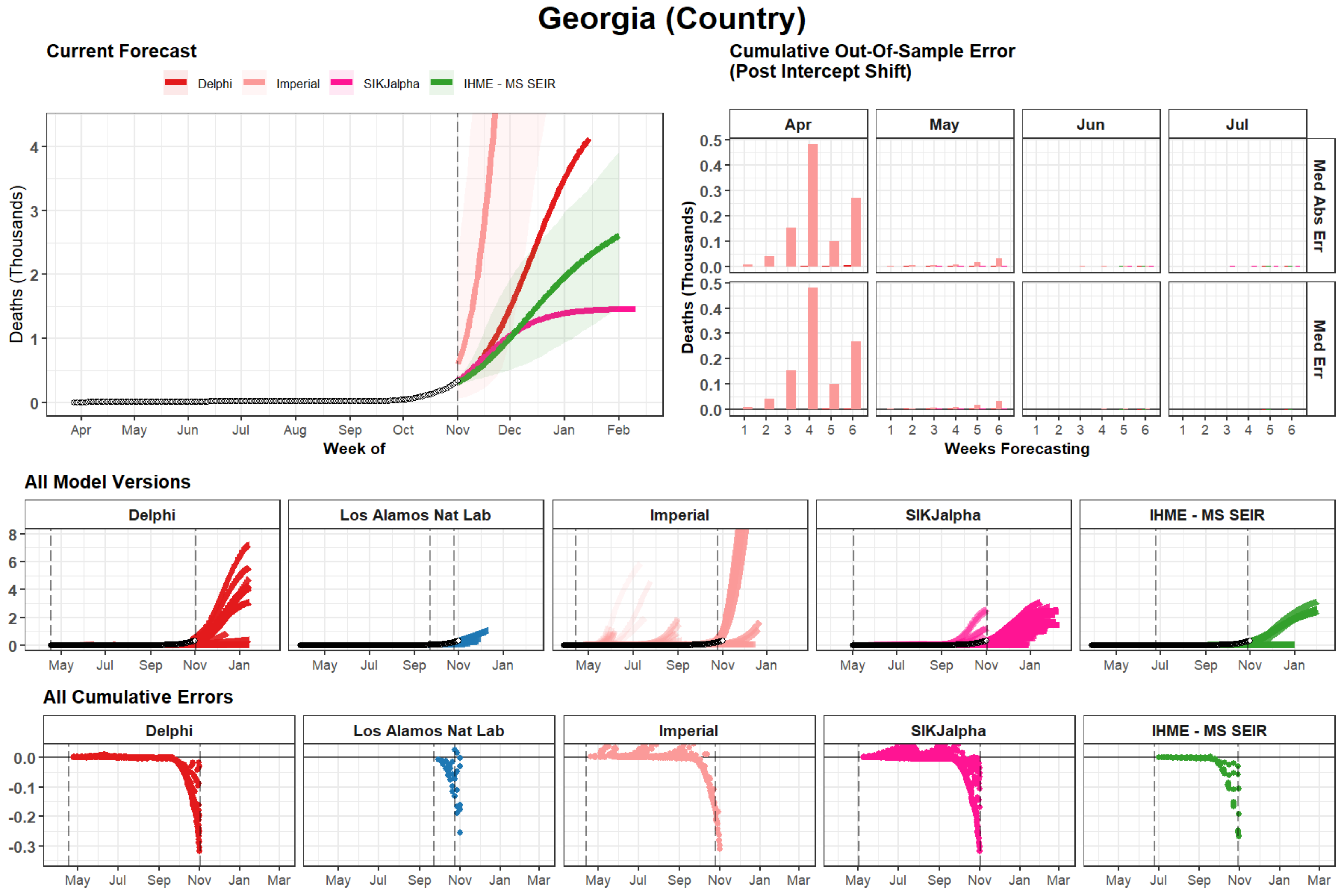

Georgia (Country)
Current Forecast
Delphi
Imperial
SIKJalpha
IHME - MS SEIR
Deaths (Thousands)
Week of
Apr
May
Jun
Jul
Aug
Sep
Oct
Nov
Dec
Jan
Feb
Cumulative Out-Of-Sample Error
(Post Intercept Shift)
Med Abs Err
Med Err
Weeks Forecasting
All Model Versions
Delphi
Los Alamos Nat Lab
Imperial
SIKJalpha
IHME - MS SEIR
All Cumulative Errors

### Senegal

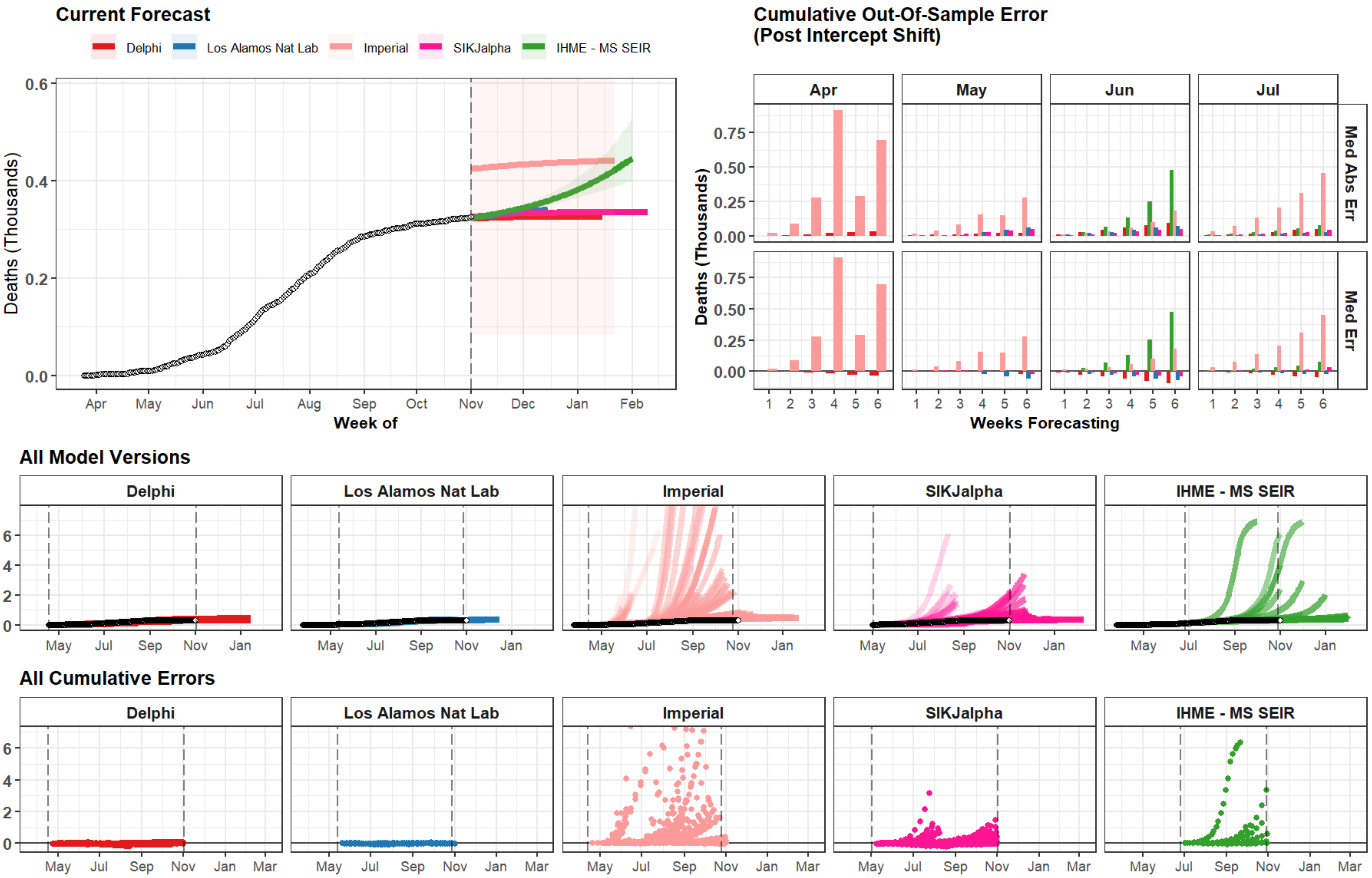
Current Forecast
Delphi
Los Alamos Nat Lab
Imperial
SIKJalpha
IHME - MS SEIR
Deaths (Thousands)
Week of
Cumulative Out-Of-Sample Error
(Post Intercept Shift)
Apr
May
Jun
Jul
Med Abs Err
Med Err
Weeks Forecasting
All Model Versions
All Cumulative Errors

### Bahrain

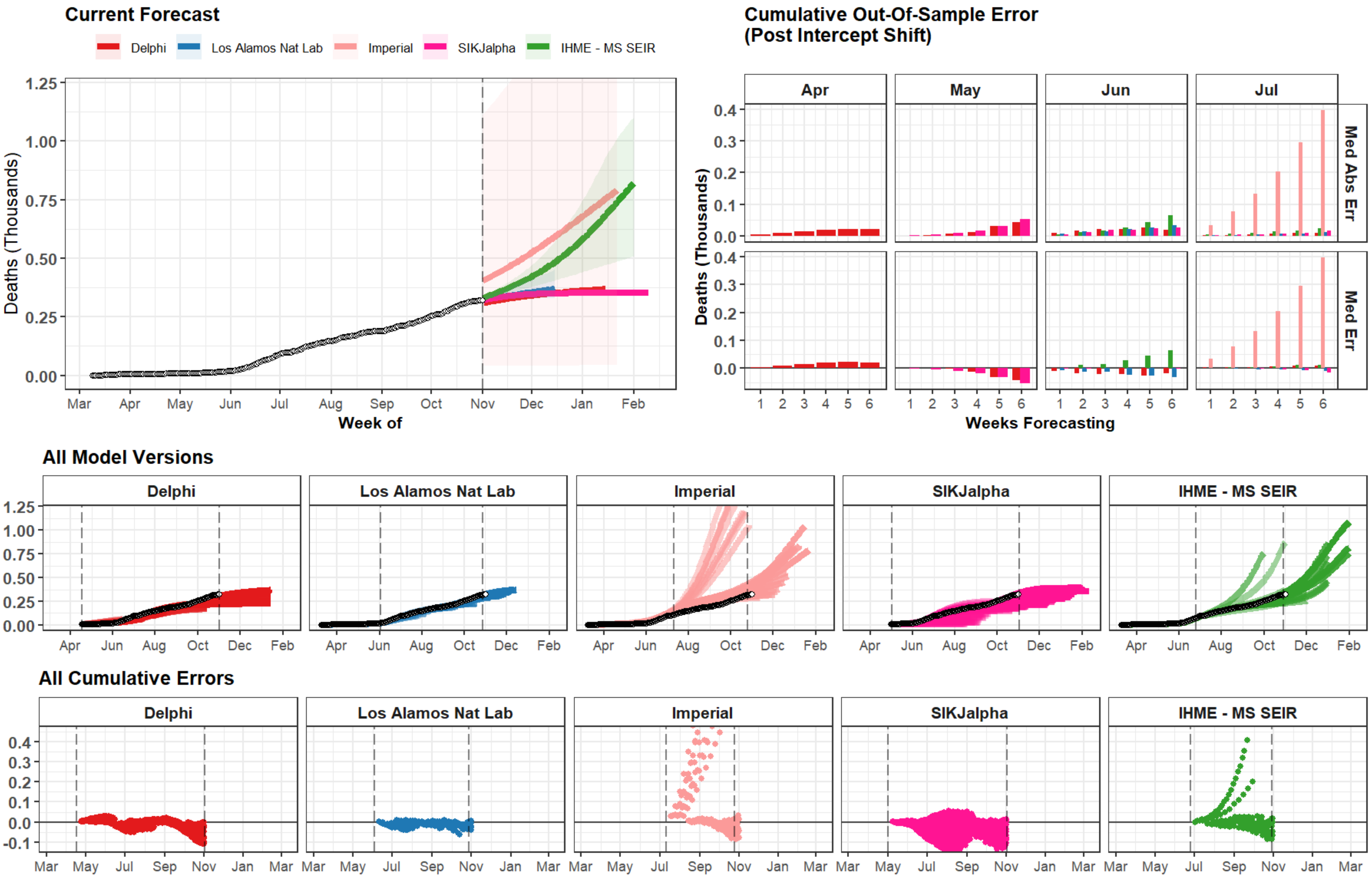

Current Forecast
Delphi
Los Alamos Nat Lab
Imperial
SIKJalpha
IHME - MS SEIR
Deaths (Thousands)
1.25
1.00
0.75
0.50
0.25
0.00
Mar Apr May Jun Jul Aug Sep Oct Nov Dec Jan Feb
Week of
Cumulative Out-Of-Sample Error
(Post Intercept Shift)
Apr
May
Jun
Jul
Med Abs Err
Med Err
Deaths (Thousands)
0.4
0.3
0.2
0.1
0.0
1 2 3 4 5 6
Weeks Forecasting
All Model Versions
Delphi
Los Alamos Nat Lab
Imperial
SIKJalpha
IHME - MS SEIR
1.25
1.00
0.75
0.50
0.25
0.00
Apr Jun Aug Oct Dec Feb
All Cumulative Errors
Delphi
Los Alamos Nat Lab
Imperial
SIKJalpha
IHME - MS SEIR
0.4
0.3
0.2
0.1
0.0
-0.1
Mar May Jul Sep Nov Jan Mar

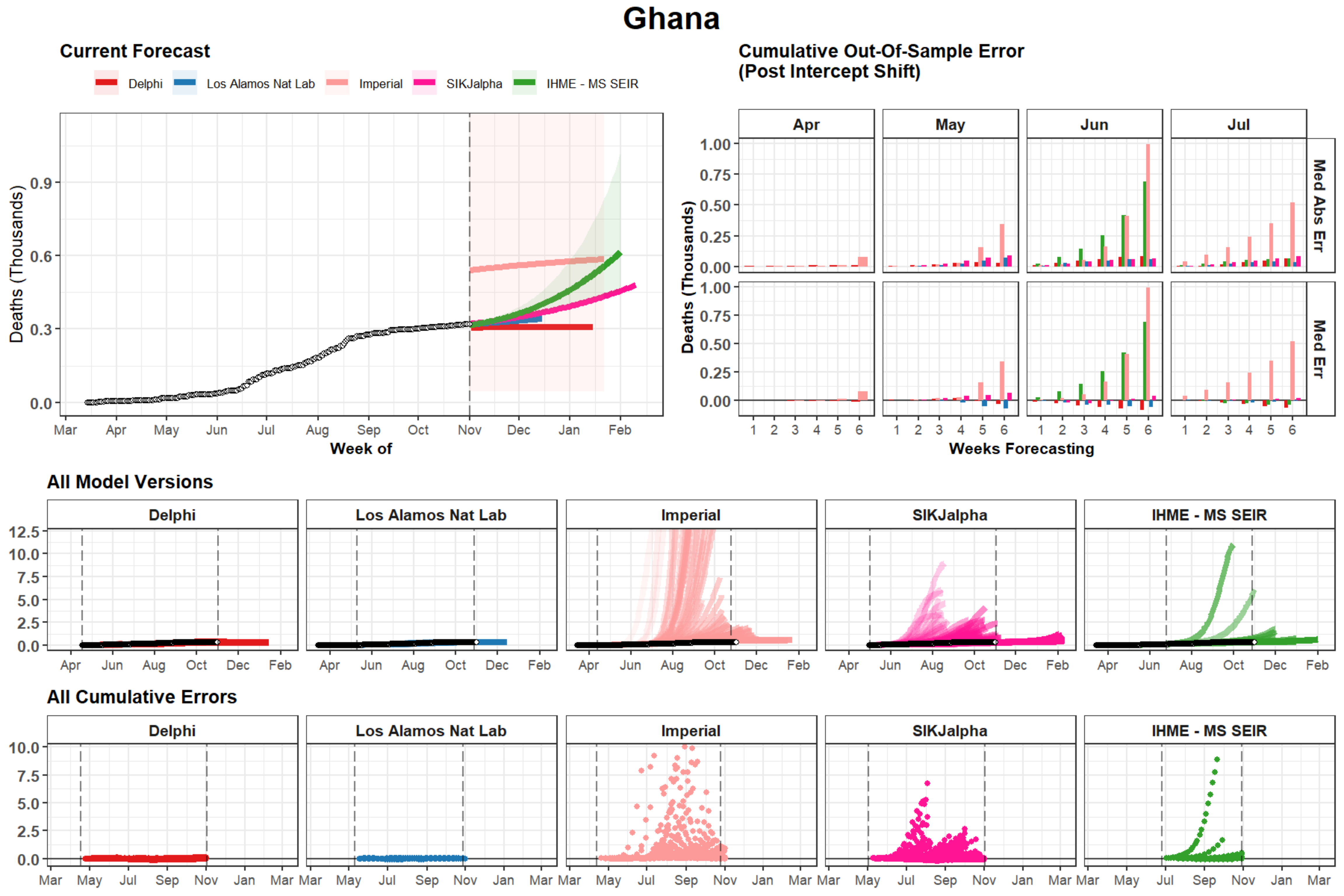

Ghana
Current Forecast
Delphi
Los Alamos Nat Lab
Imperial
SIKJalpha
IHME - MS SEIR
Deaths (Thousands)
Week of
Cumulative Out-Of-Sample Error
(Post Intercept Shift)
Apr
May
Jun
Jul
Med Abs Err
Med Err
Weeks Forecasting
All Model Versions
All Cumulative Errors

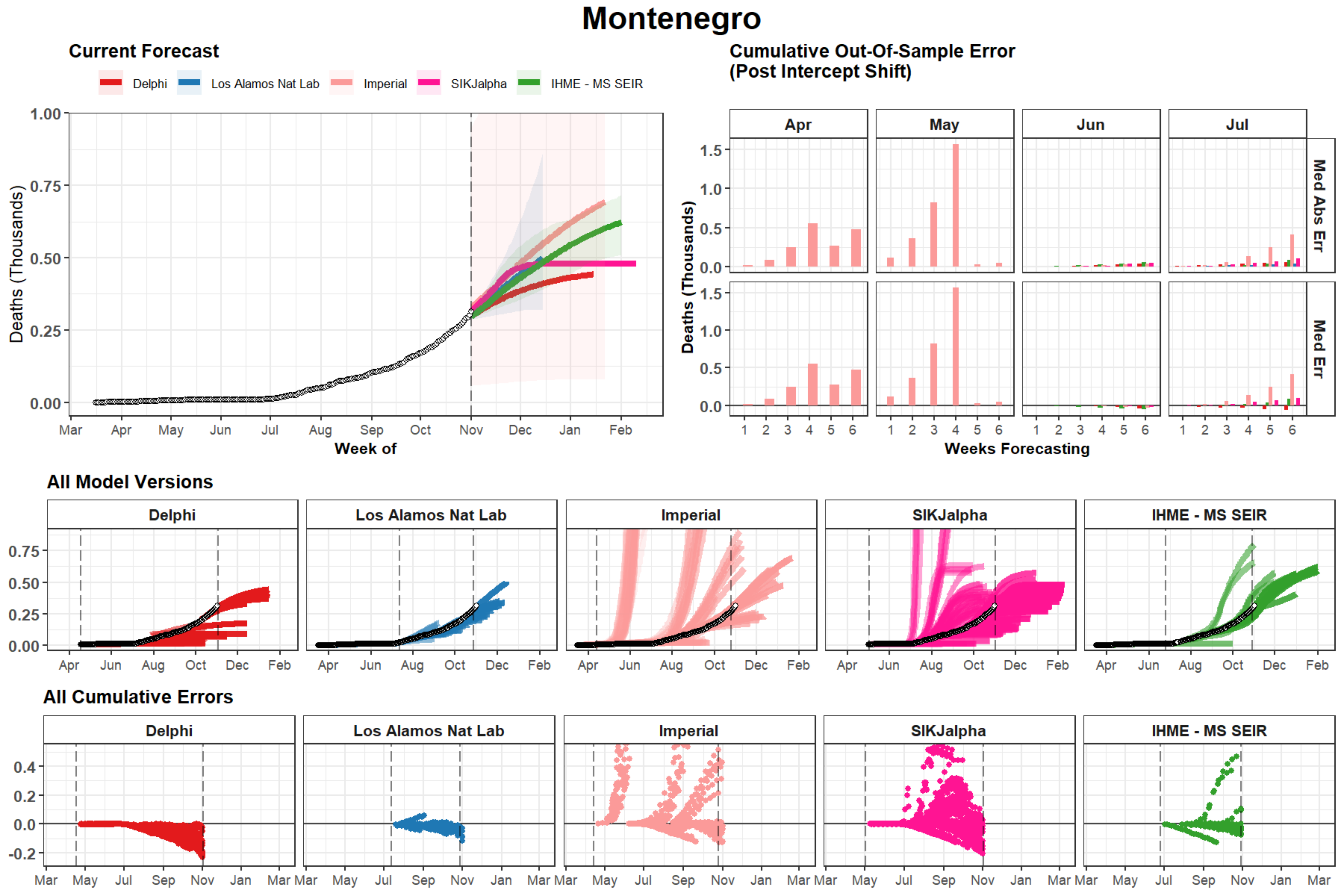
Montenegro
Current Forecast
Delphi
Los Alamos Nat Lab
Imperial
SIKJalpha
IHME - MS SEIR
Deaths (Thousands)
Week of
Cumulative Out-Of-Sample Error
(Post Intercept Shift)
Apr
May
Jun
Jul
Med Abs Err
Med Err
Weeks Forecasting
All Model Versions
All Cumulative Errors

### Democratic Republic of the Congo

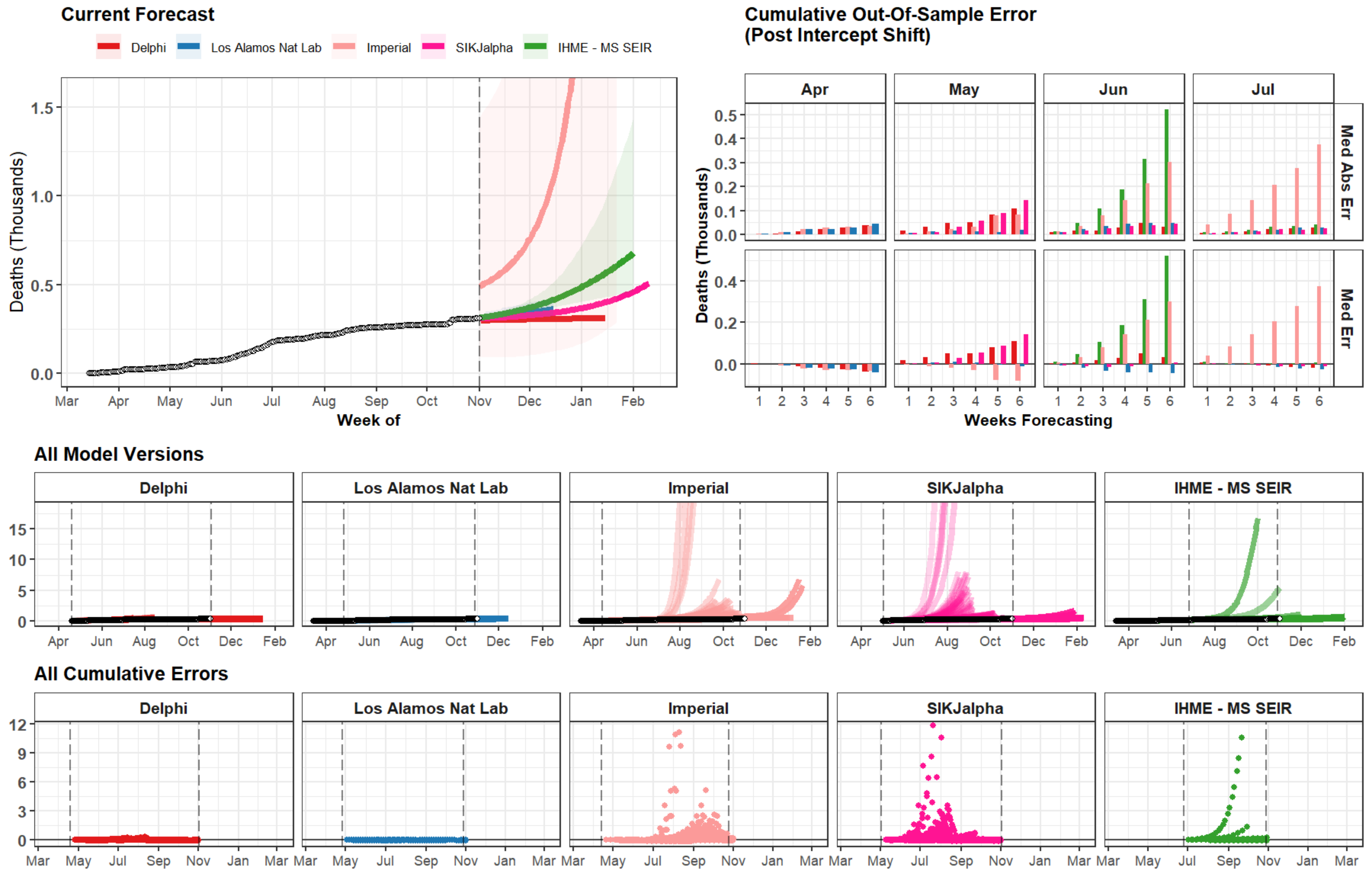

### Syria

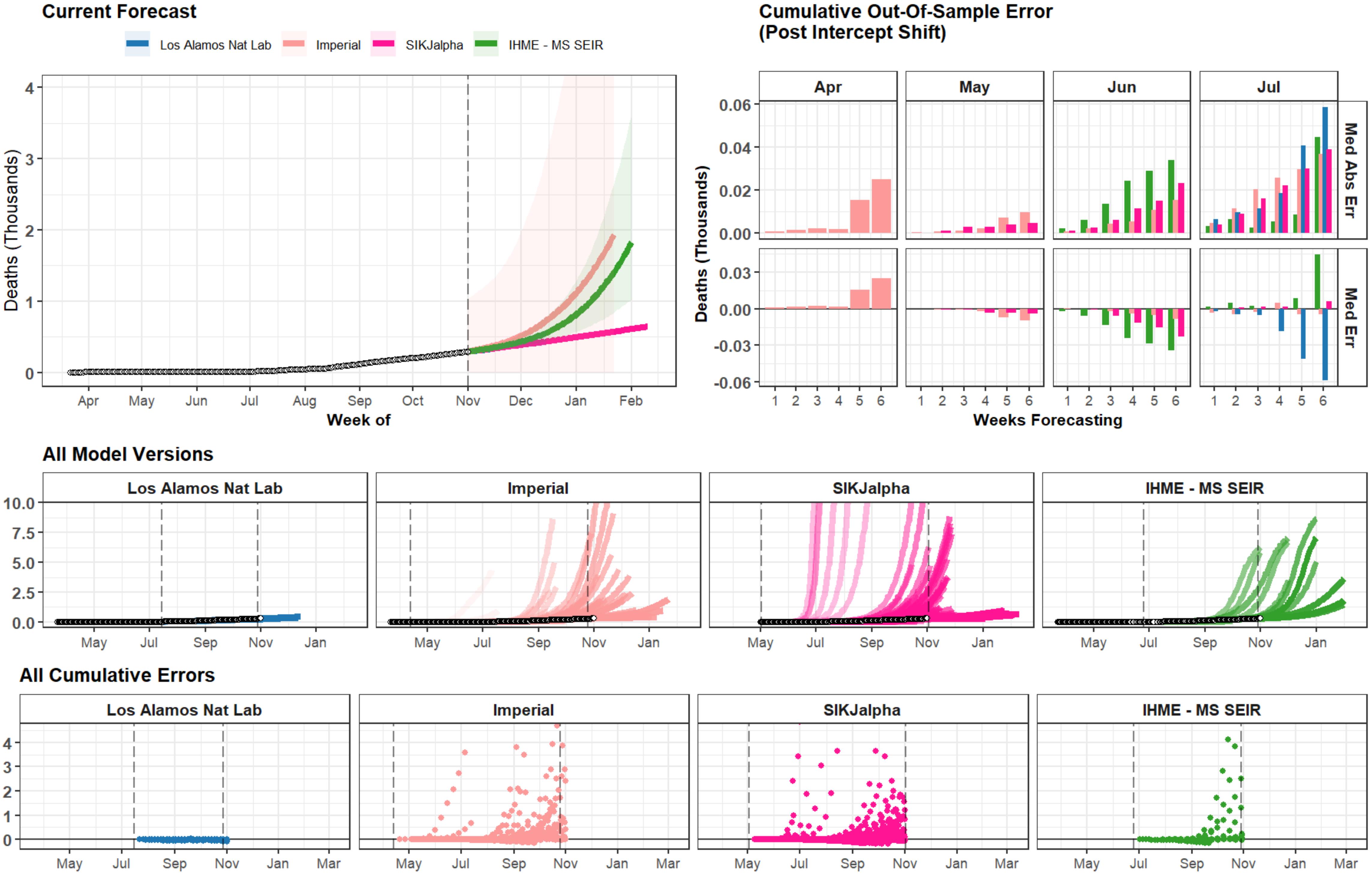
Current Forecast
Los Alamos Nat Lab
Imperial
SIKJalpha
IHME - MS SEIR
Deaths (Thousands)
Week of
Apr May Jun Jul Aug Sep Oct Nov Dec Jan Feb
Cumulative Out-Of-Sample Error
(Post Intercept Shift)
Apr
May
Jun
Jul
Med Abs Err
Med Err
Deaths (Thousands)
Weeks Forecasting
All Model Versions
Los Alamos Nat Lab
Imperial
SIKJalpha
IHME - MS SEIR
May Jul Sep Nov Jan
All Cumulative Errors
Los Alamos Nat Lab
Imperial
SIKJalpha
IHME - MS SEIR
May Jul Sep Nov Jan Mar

### Angola

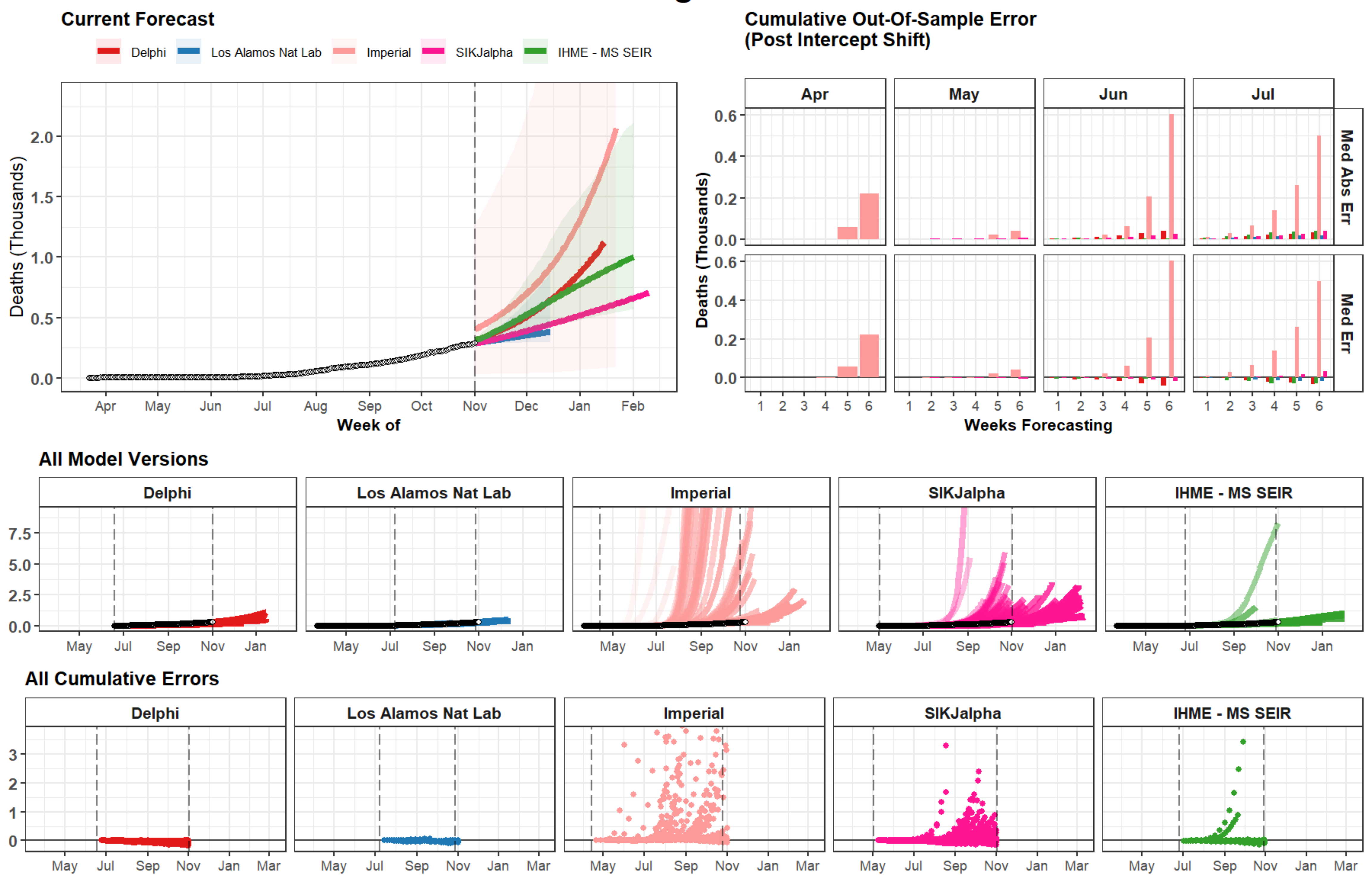

Current Forecast
Delphi
Los Alamos Nat Lab
Imperial
SIKJalpha
IHME - MS SEIR
Deaths (Thousands)
2.0
1.5
1.0
0.5
0.0
Apr
May
Jun
Jul
Aug
Sep
Oct
Nov
Dec
Jan
Feb
Week of
Cumulative Out-Of-Sample Error
(Post Intercept Shift)
Apr
May
Jun
Jul
Med Abs Err
Med Err
0.6
0.4
0.2
0.0
1
2
3
4
5
6
Weeks Forecasting
All Model Versions
Delphi
Los Alamos Nat Lab
Imperial
SIKJalpha
IHME - MS SEIR
7.5
5.0
2.5
0.0
May
Jul
Sep
Nov
Jan
All Cumulative Errors
Delphi
Los Alamos Nat Lab
Imperial
SIKJalpha
IHME - MS SEIR
3
2
1
0
May
Jul
Sep
Nov
Jan
Mar

### Norway

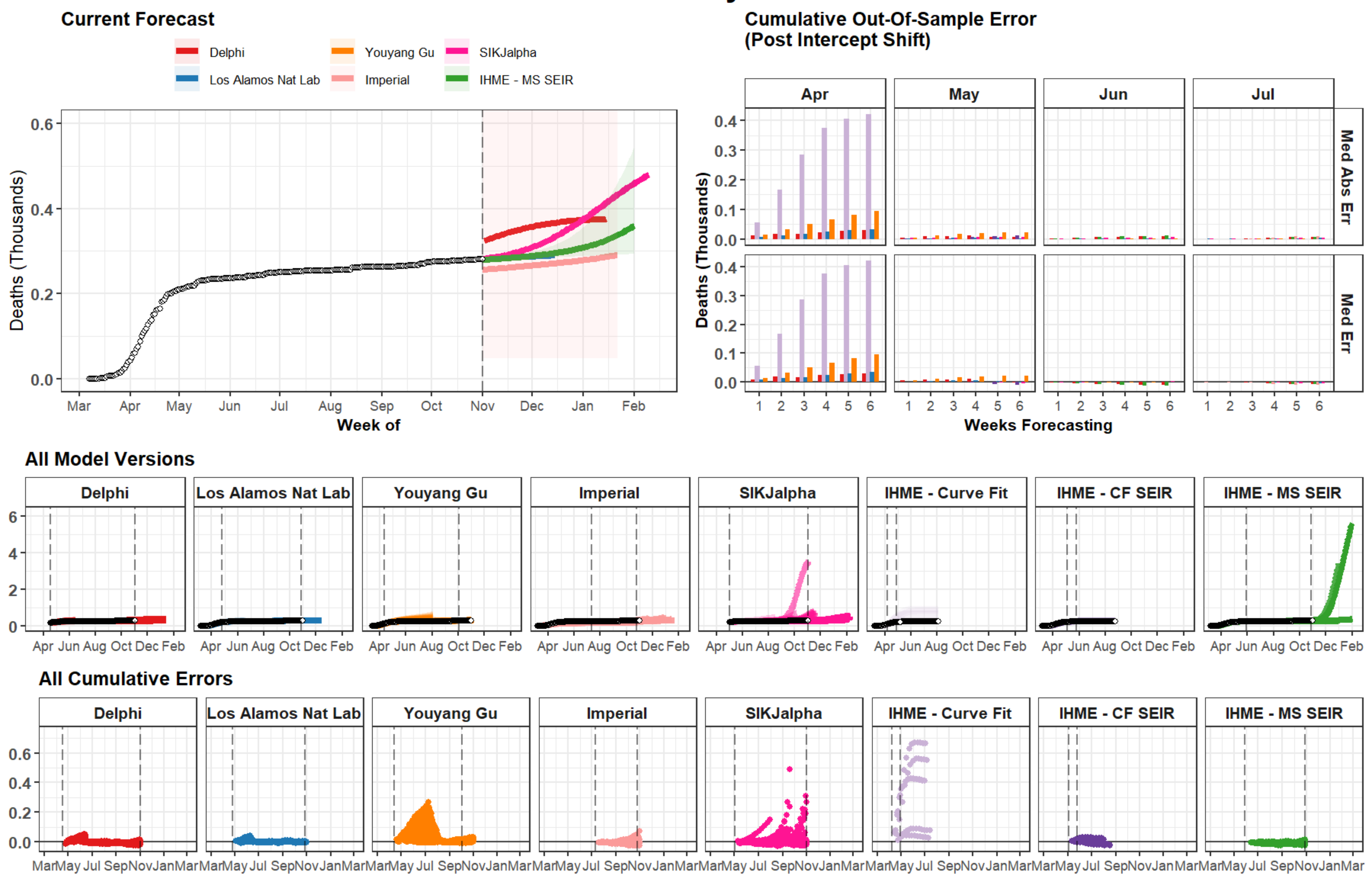

Current Forecast
Delphi
Los Alamos Nat Lab
Youyang Gu
Imperial
SIKJalpha
IHME - MS SEIR
Deaths (Thousands)
Week of
Cumulative Out-Of-Sample Error
(Post Intercept Shift)
Apr
May
Jun
Jul
Med Abs Err
Med Err
Weeks Forecasting
All Model Versions
Delphi
Los Alamos Nat Lab
Youyang Gu
Imperial
SIKJalpha
IHME - Curve Fit
IHME - CF SEIR
IHME - MS SEIR
All Cumulative Errors

### Malaysia

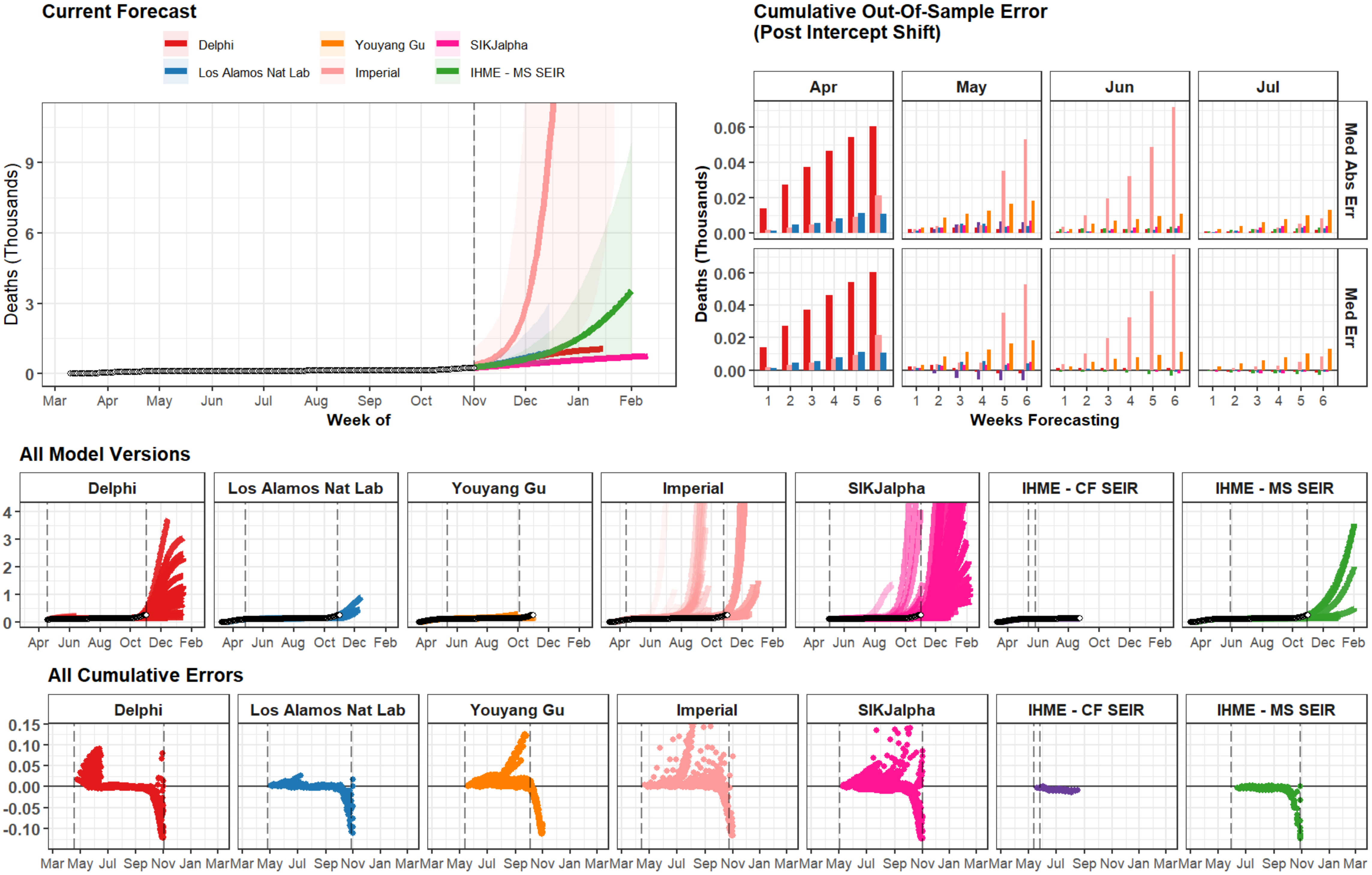
Current Forecast
Delphi
Los Alamos Nat Lab
Youyang Gu
Imperial
SIKJalpha
IHME - MS SEIR
Deaths (Thousands)
Week of
Mar Apr May Jun Jul Aug Sep Oct Nov Dec Jan Feb
Cumulative Out-Of-Sample Error
(Post Intercept Shift)
Apr May Jun Jul
Med Abs Err
Med Err
Weeks Forecasting
All Model Versions
Delphi
Los Alamos Nat Lab
Youyang Gu
Imperial
SIKJalpha
IHME - CF SEIR
IHME - MS SEIR
All Cumulative Errors

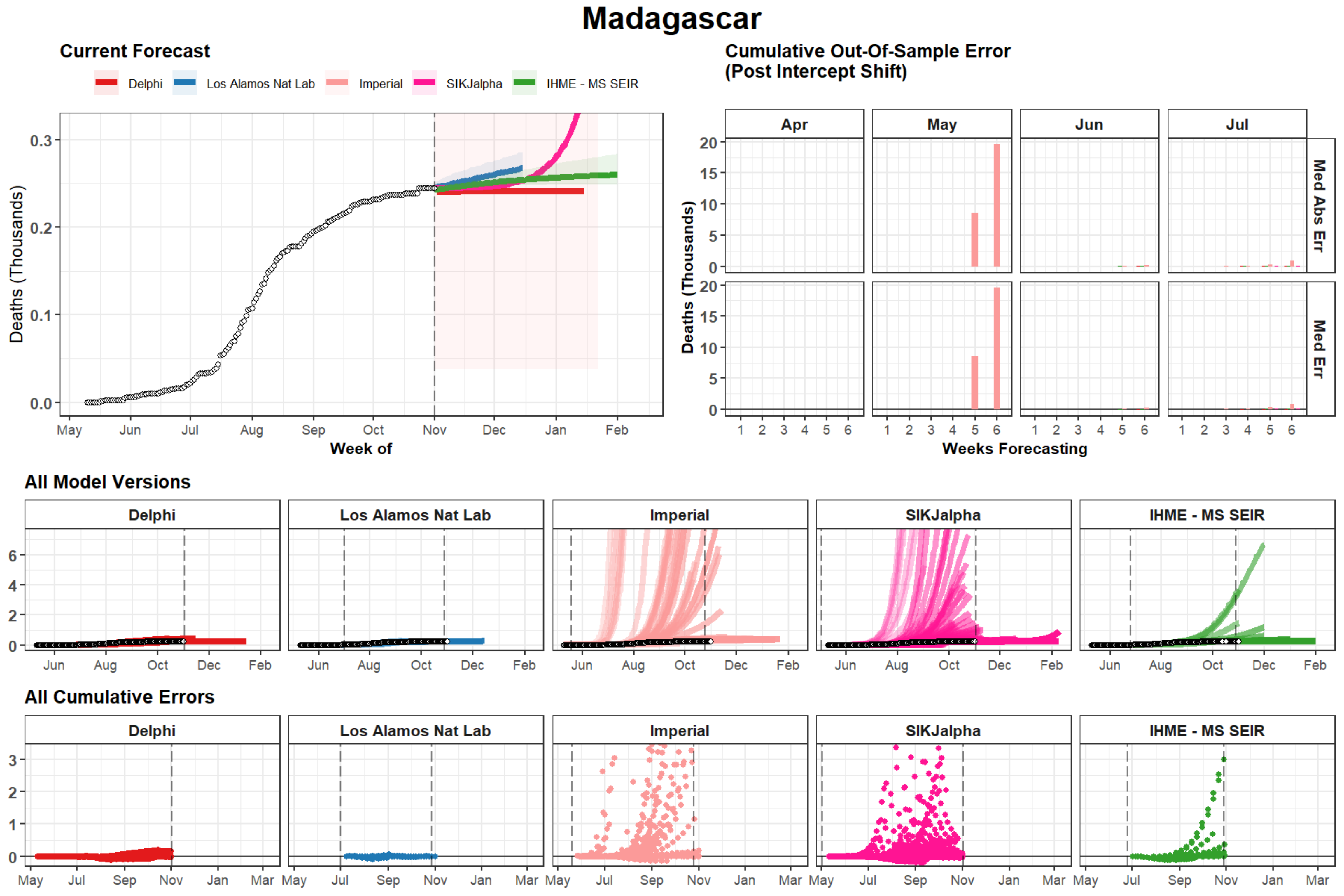
Madagascar
Current Forecast
Delphi
Los Alamos Nat Lab
Imperial
SIKJalpha
IHME - MS SEIR
Deaths (Thousands)
Week of
May
Jun
Jul
Aug
Sep
Oct
Nov
Dec
Jan
Feb
Cumulative Out-Of-Sample Error
(Post Intercept Shift)
Apr
May
Jun
Jul
Med Abs Err
Med Err
Weeks Forecasting
All Model Versions
All Cumulative Errors
Mar

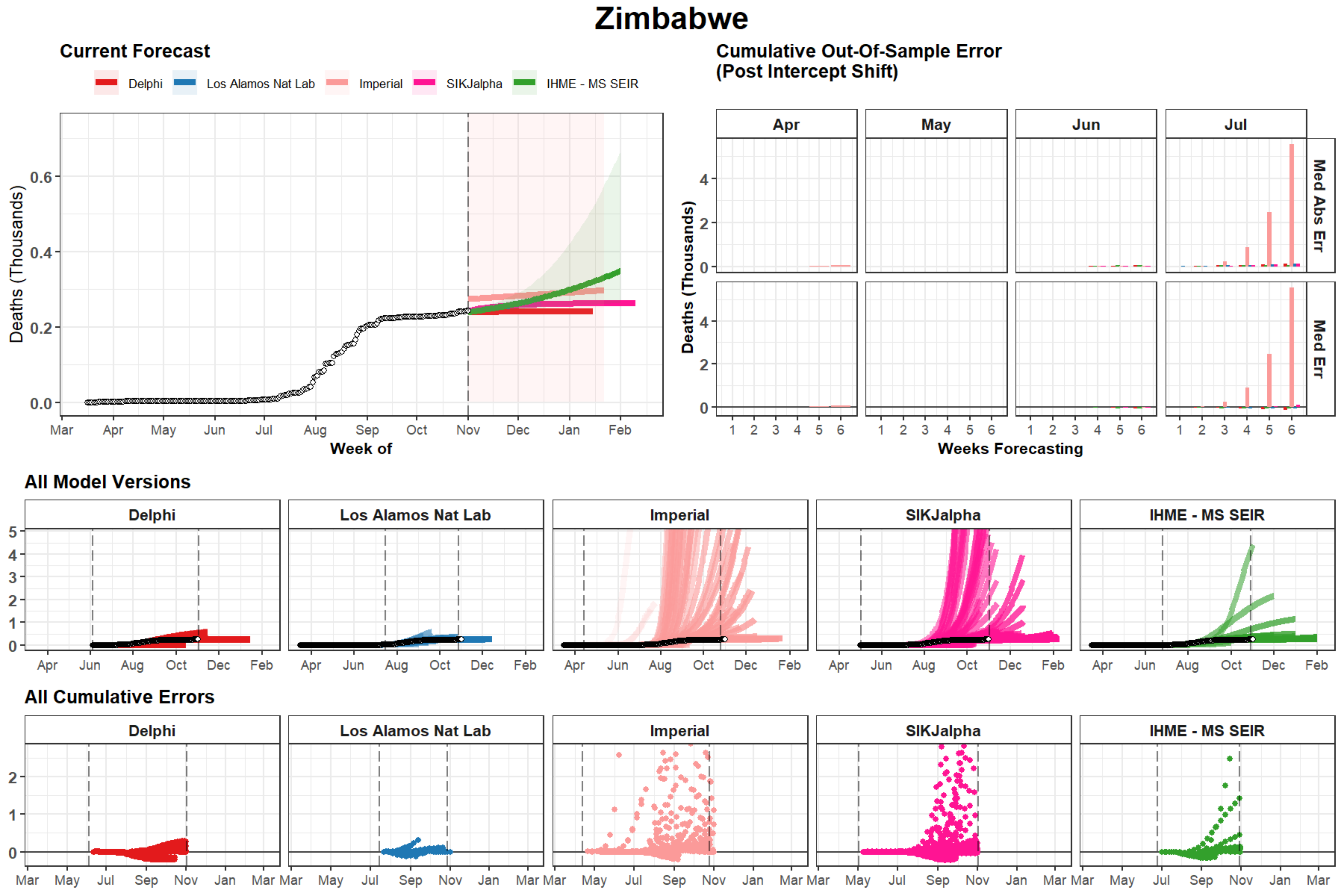

Zimbabwe
Current Forecast
Delphi
Los Alamos Nat Lab
Imperial
SIKJalpha
IHME - MS SEIR
Deaths (Thousands)
Week of
Cumulative Out-Of-Sample Error
(Post Intercept Shift)
Apr
May
Jun
Jul
Med Abs Err
Med Err
Weeks Forecasting
All Model Versions
All Cumulative Errors

### Haiti

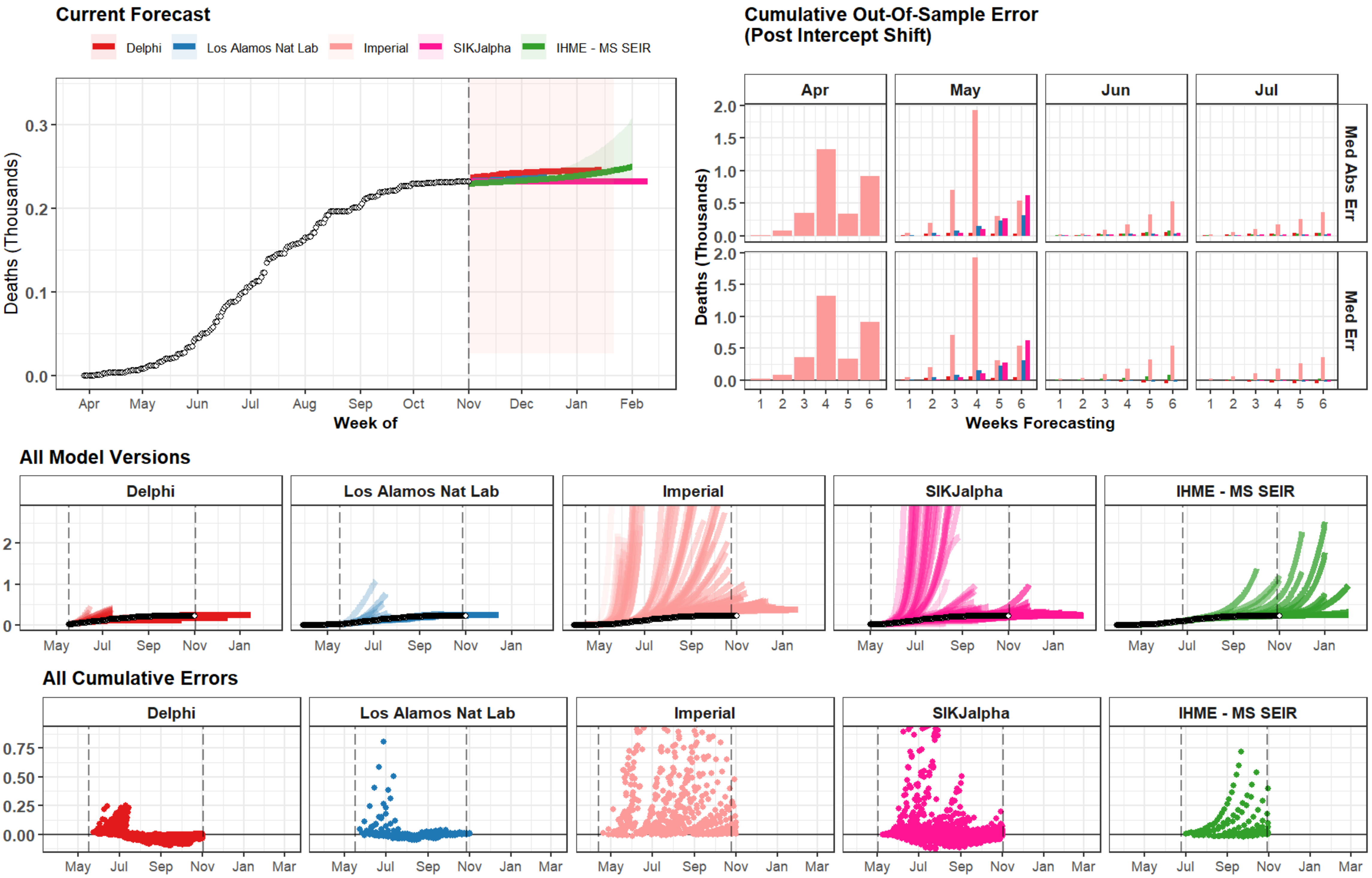

Current Forecast
Delphi
Los Alamos Nat Lab
Imperial
SIKJalpha
IHME - MS SEIR
Deaths (Thousands)
Week of
Cumulative Out-Of-Sample Error
(Post Intercept Shift)
Apr
May
Jun
Jul
Med Abs Err
Med Err
Weeks Forecasting
All Model Versions
All Cumulative Errors

### Qatar

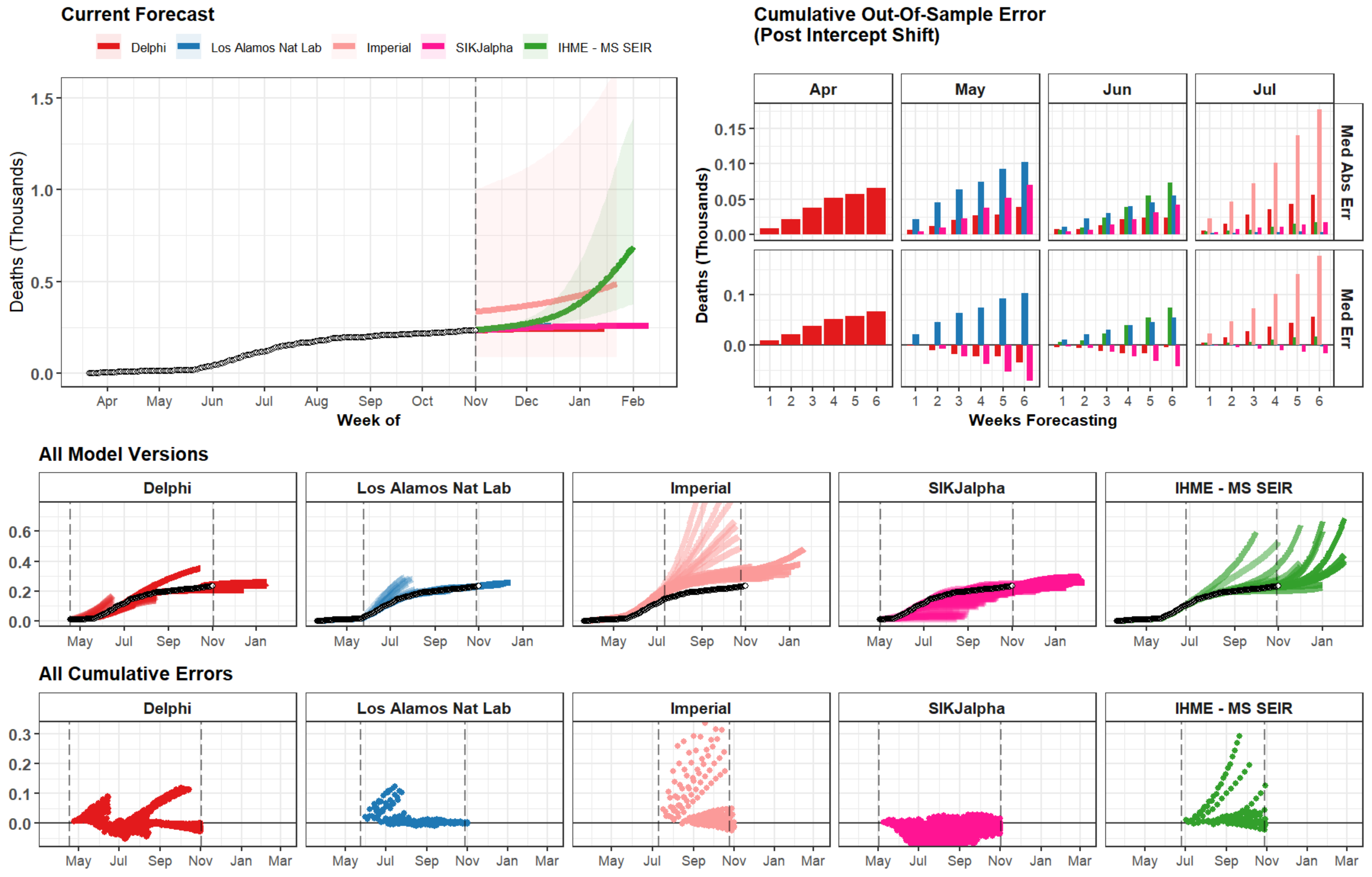
Current Forecast
Delphi
Los Alamos Nat Lab
Imperial
SIKJalpha
IHME - MS SEIR
Deaths (Thousands)
1.5
1.0
0.5
0.0
Apr
May
Jun
Jul
Aug
Sep
Oct
Nov
Dec
Jan
Feb
Week of
Cumulative Out-Of-Sample Error
(Post Intercept Shift)
Apr
May
Jun
Jul
Med Abs Err
Med Err
0.15
0.10
0.05
0.00
0.1
0.0
1
2
3
4
5
6
Weeks Forecasting
All Model Versions
Delphi
Los Alamos Nat Lab
Imperial
SIKJalpha
IHME - MS SEIR
0.6
0.4
0.2
0.0
May
Jul
Sep
Nov
Jan
All Cumulative Errors
Delphi
Los Alamos Nat Lab
Imperial
SIKJalpha
IHME - MS SEIR
0.3
0.2
0.1
0.0
May
Jul
Sep
Nov
Jan
Mar

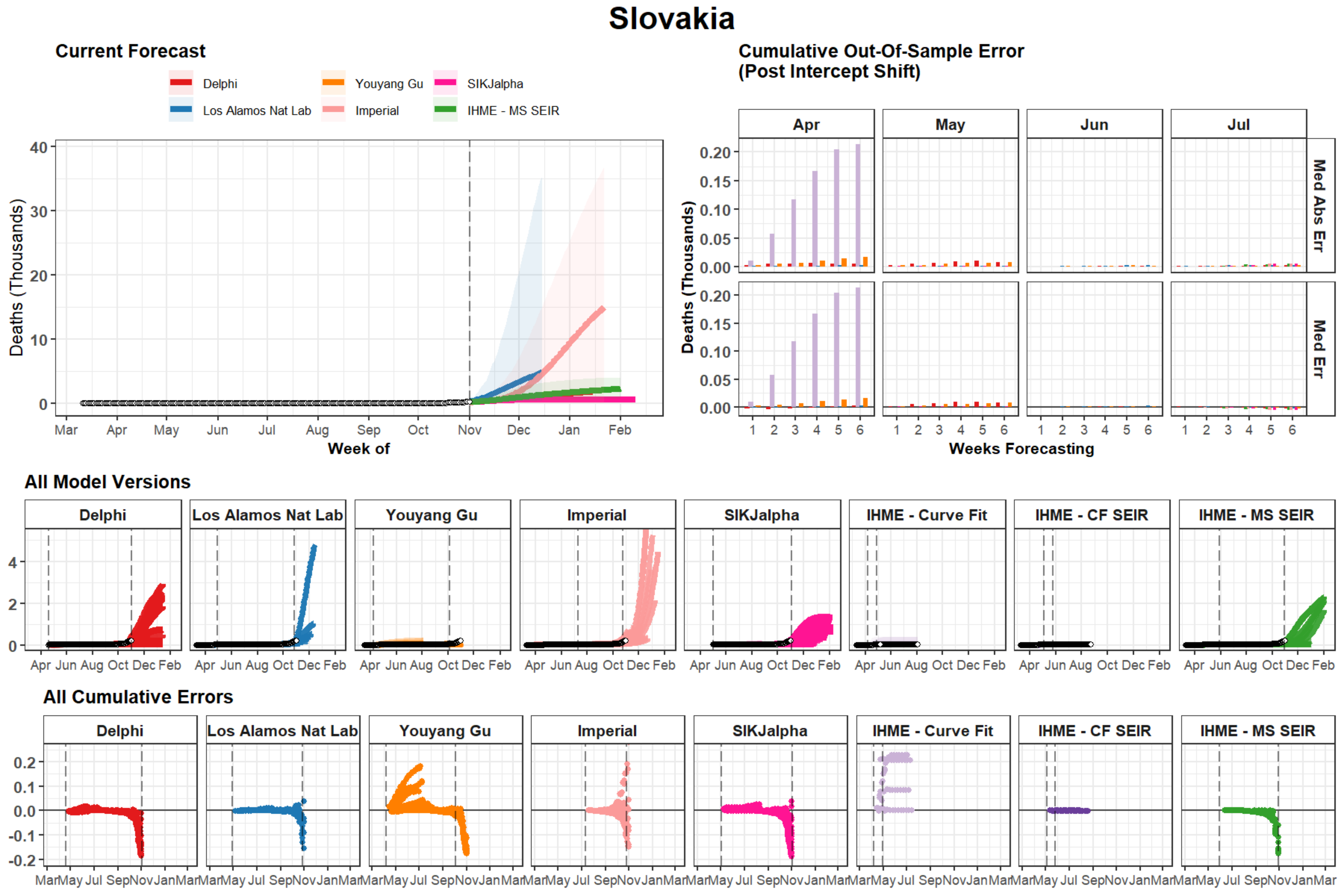
Slovakia
Current Forecast
Delphi
Los Alamos Nat Lab
Youyang Gu
Imperial
SIKJalpha
IHME - MS SEIR
Deaths (Thousands)
Week of
Mar Apr May Jun Jul Aug Sep Oct Nov Dec Jan Feb
Cumulative Out-Of-Sample Error
(Post Intercept Shift)
Apr
May
Jun
Jul
Med Abs Err
Med Err
Deaths (Thousands)
Weeks Forecasting
All Model Versions
Delphi
Los Alamos Nat Lab
Youyang Gu
Imperial
SIKJalpha
IHME - Curve Fit
IHME - CF SEIR
IHME - MS SEIR
Apr Jun Aug Oct Dec Feb
All Cumulative Errors
Mar May Jul Sep Nov Jan Mar

### Hawaii

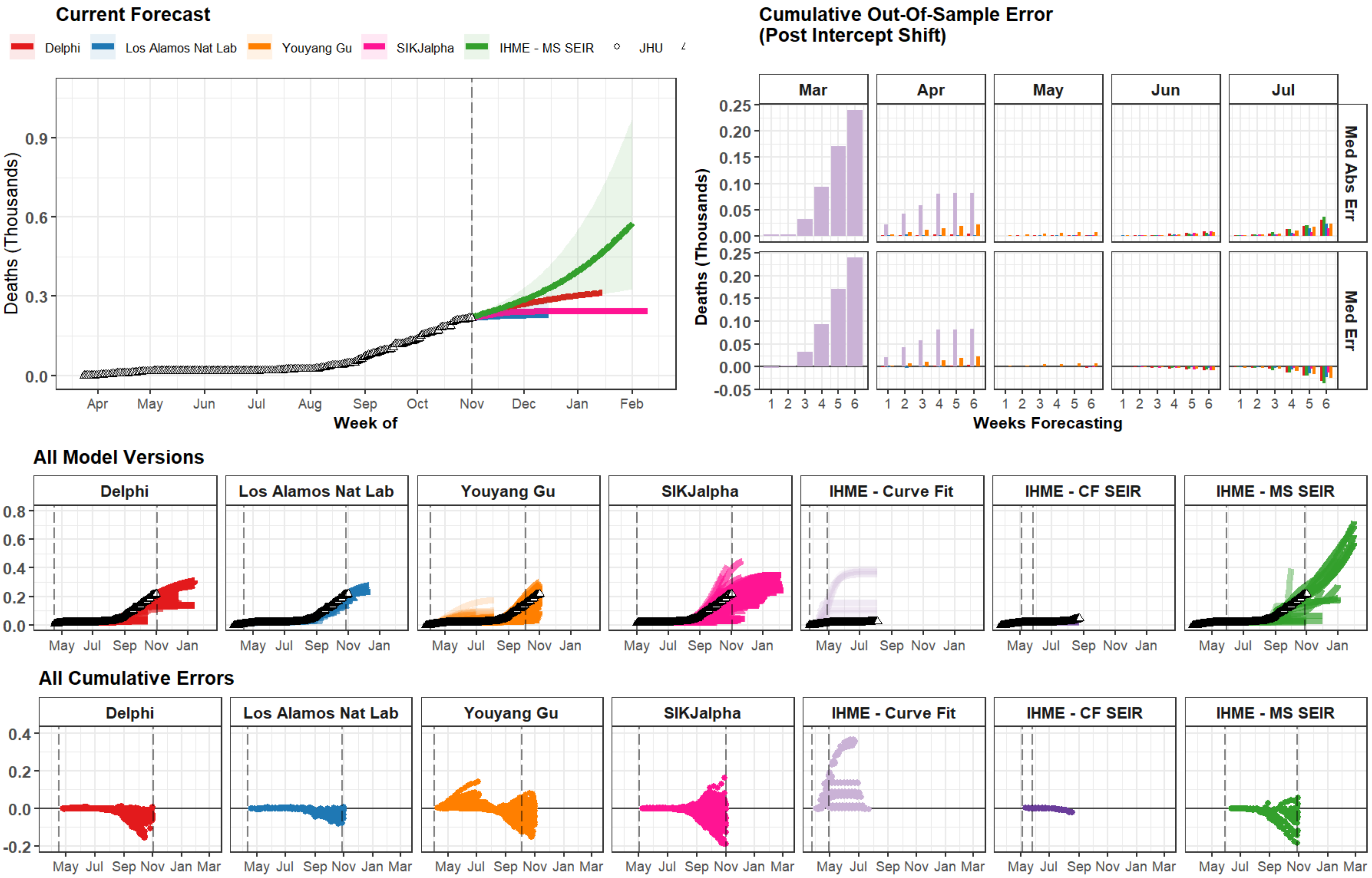

Current Forecast
Delphi
Los Alamos Nat Lab
Youyang Gu
SIKJalpha
IHME - MS SEIR
JHU
Deaths (Thousands)
Week of
Apr May Jun Jul Aug Sep Oct Nov Dec Jan Feb
Cumulative Out-Of-Sample Error
(Post Intercept Shift)
Mar Apr May Jun Jul
Med Abs Err
Med Err
Weeks Forecasting
All Model Versions
Delphi
Los Alamos Nat Lab
Youyang Gu
SIKJalpha
IHME - Curve Fit
IHME - CF SEIR
IHME - MS SEIR
All Cumulative Errors

### Jamaica

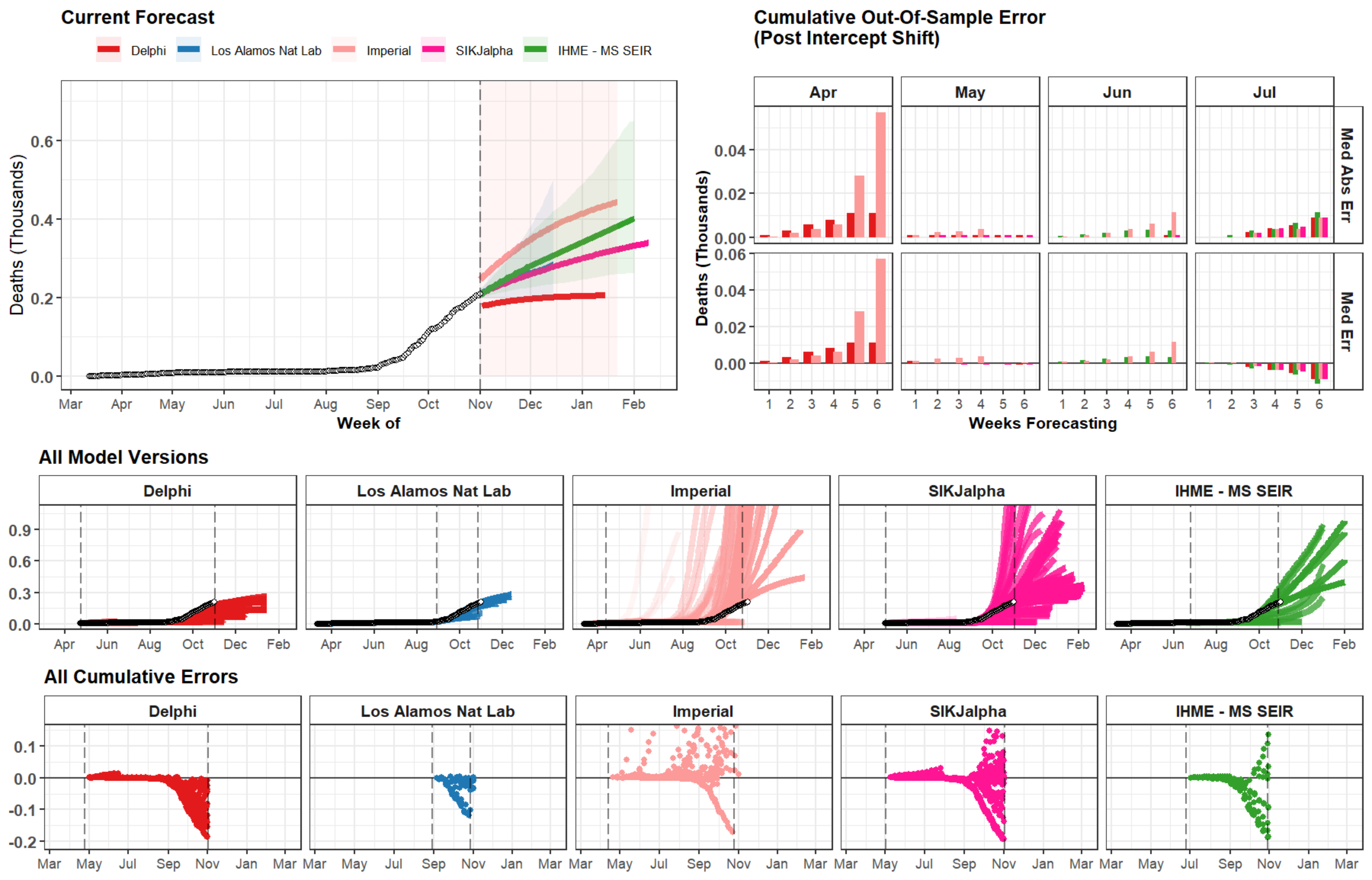

Current Forecast
Delphi
Los Alamos Nat Lab
Imperial
SIKJalpha
IHME - MS SEIR
Deaths (Thousands)
0.6
0.4
0.2
0.0
Mar
Apr
May
Jun
Jul
Aug
Sep
Oct
Nov
Dec
Jan
Feb
Week of
Cumulative Out-Of-Sample Error
(Post Intercept Shift)
Apr
May
Jun
Jul
Med Abs Err
Med Err
0.04
0.02
0.00
0.06
Weeks Forecasting
1
2
3
4
5
6
All Model Versions
0.9
0.3
All Cumulative Errors
0.1
-0.1
-0.2

### Malawi

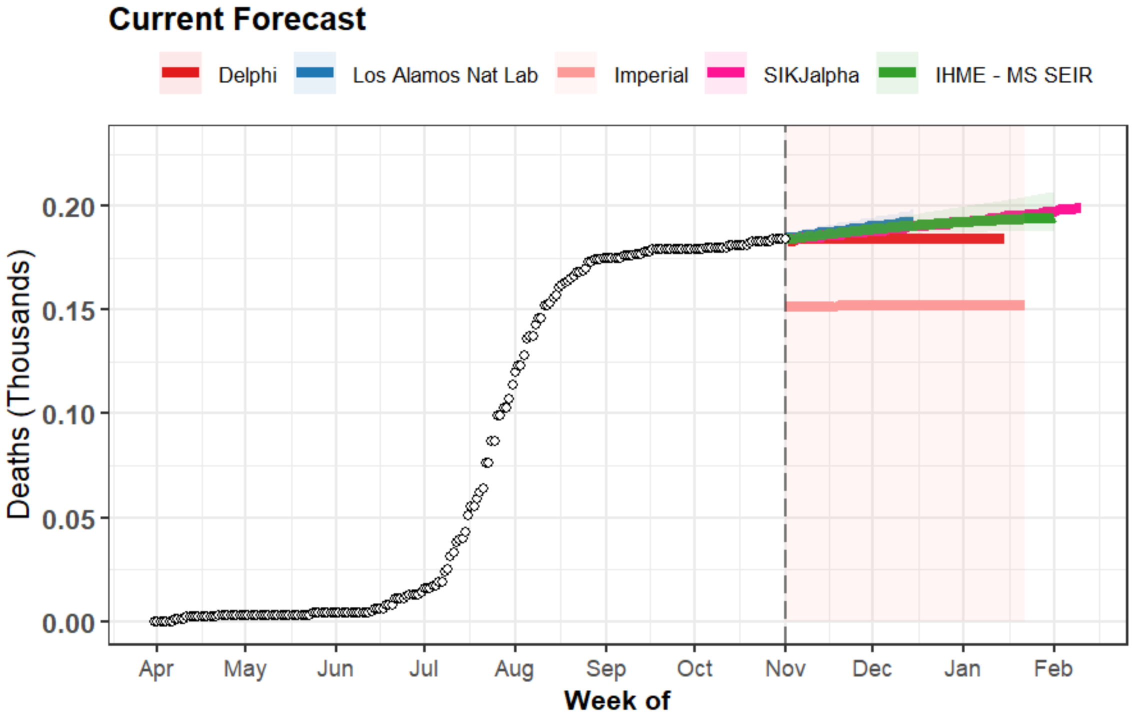

Current Forecast
Delphi
Los Alamos Nat Lab
Imperial
SIKJalpha
IHME - MS SEIR
Deaths (Thousands)
0.20
0.15
0.10
0.05
0.00
Apr
May
Jun
Jul
Aug
Sep
Oct
Nov
Dec
Jan
Feb
Week of


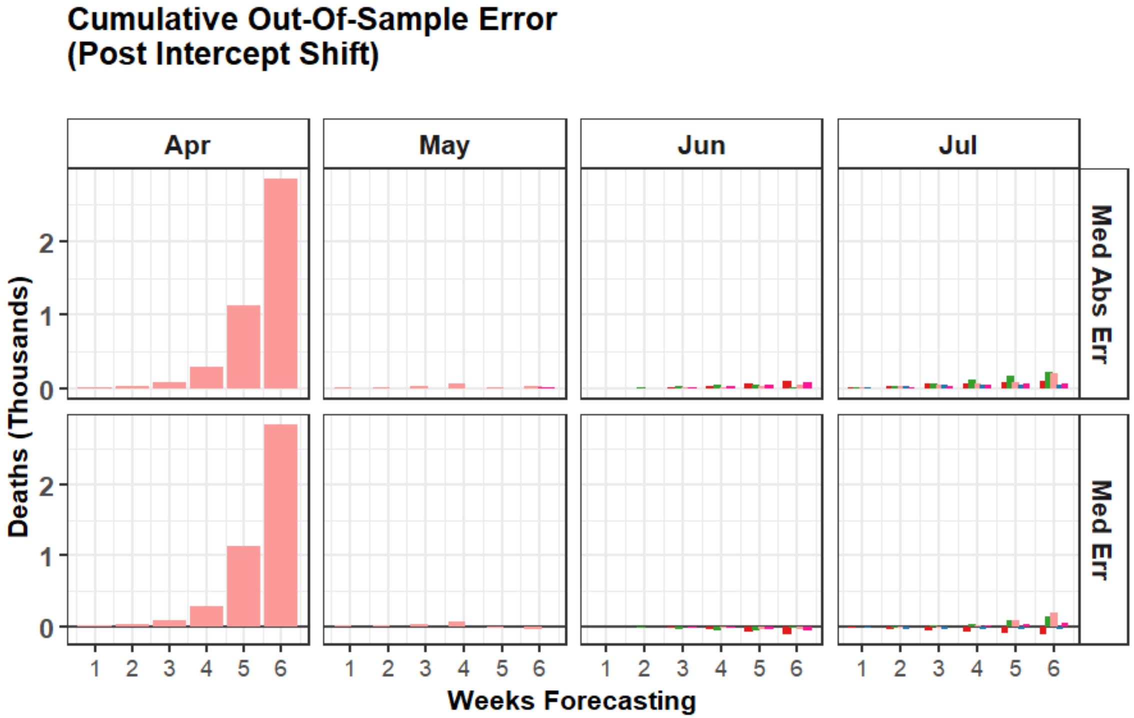

Cumulative Out-Of-Sample Error
(Post Intercept Shift)
Apr
May
Jun
Jul
Med Abs Err
Med Err
Deaths (Thousands)
2
1
0
1
2
3
4
5
6
Weeks Forecasting


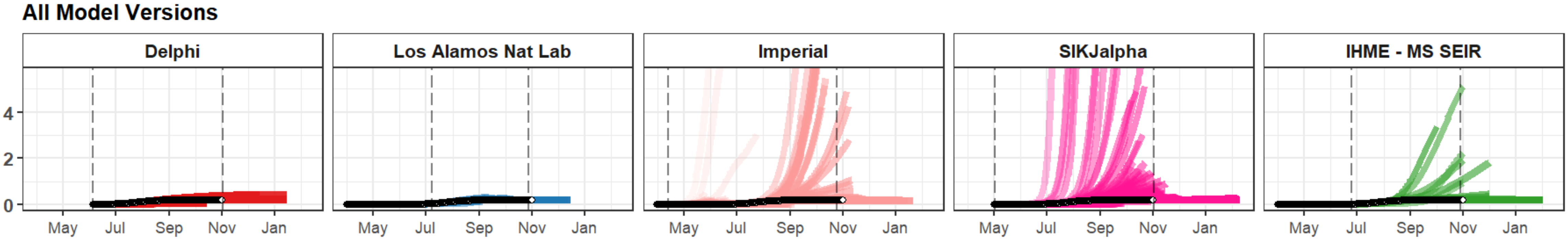

All Model Versions
Delphi
Los Alamos Nat Lab
Imperial
SIKJalpha
IHME - MS SEIR
4
2
0
May
Jul
Sep
Nov
Jan


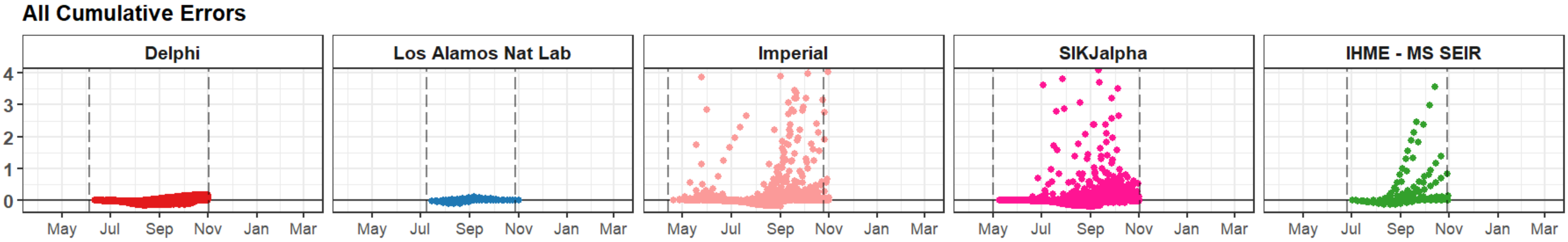

All Cumulative Errors
Delphi
Los Alamos Nat Lab
Imperial
SIKJalpha
IHME - MS SEIR
4
3
2
1
0
May
Jul
Sep
Nov
Jan
Mar

### Lithuania

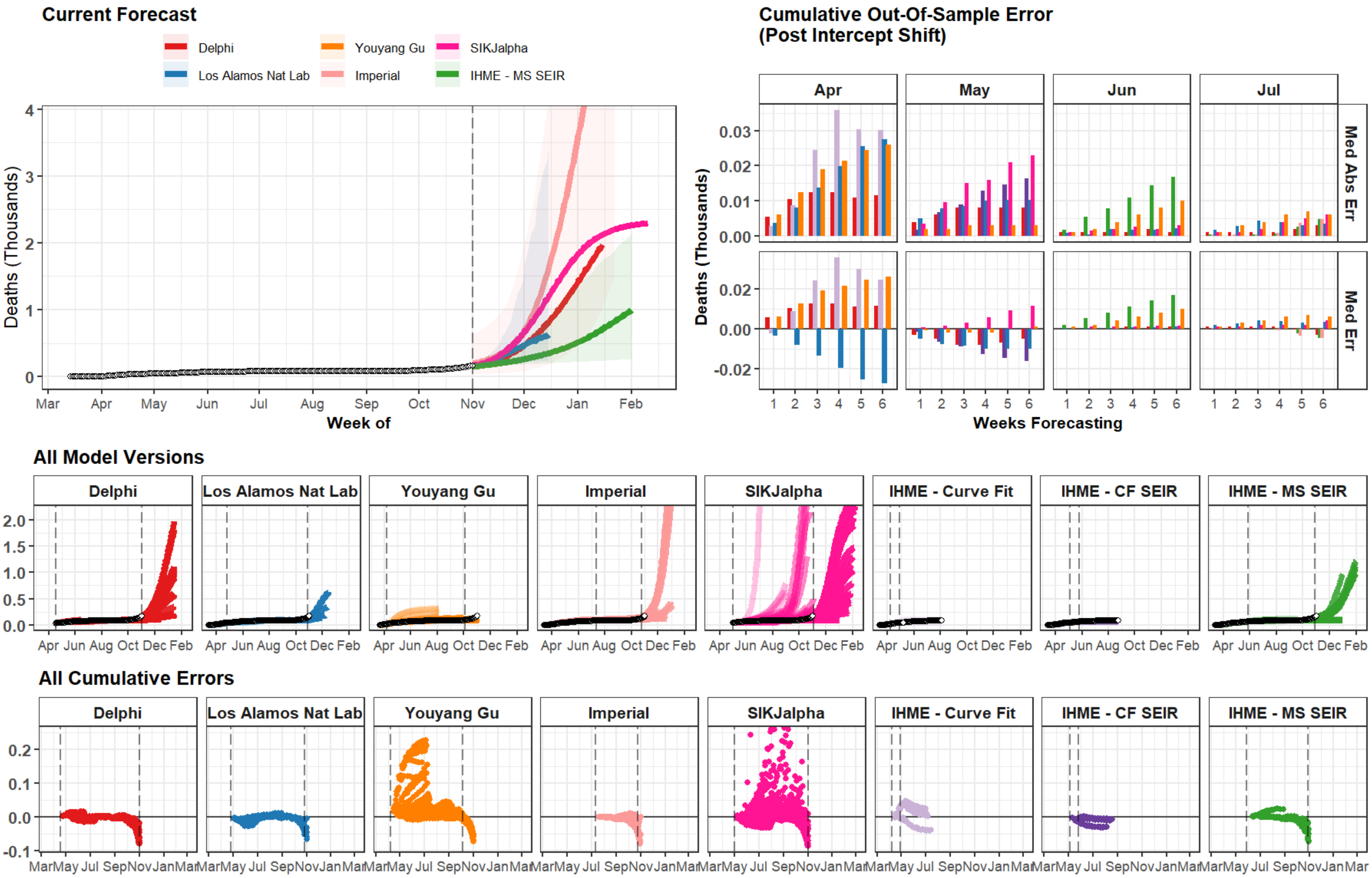

Current Forecast
Delphi
Youyang Gu
SIKJalpha
Los Alamos Nat Lab
Imperial
IHME - MS SEIR
Deaths (Thousands)
Week of
Mar Apr May Jun Jul Aug Sep Oct Nov Dec Jan Feb
Cumulative Out-Of-Sample Error
(Post Intercept Shift)
Apr
May
Jun
Jul
Med Abs Err
Med Err
Weeks Forecasting
All Model Versions
Delphi
Los Alamos Nat Lab
Youyang Gu
Imperial
SIKJalpha
IHME - Curve Fit
IHME - CF SEIR
IHME - MS SEIR
All Cumulative Errors

### Mauritania

#### Current Forecast

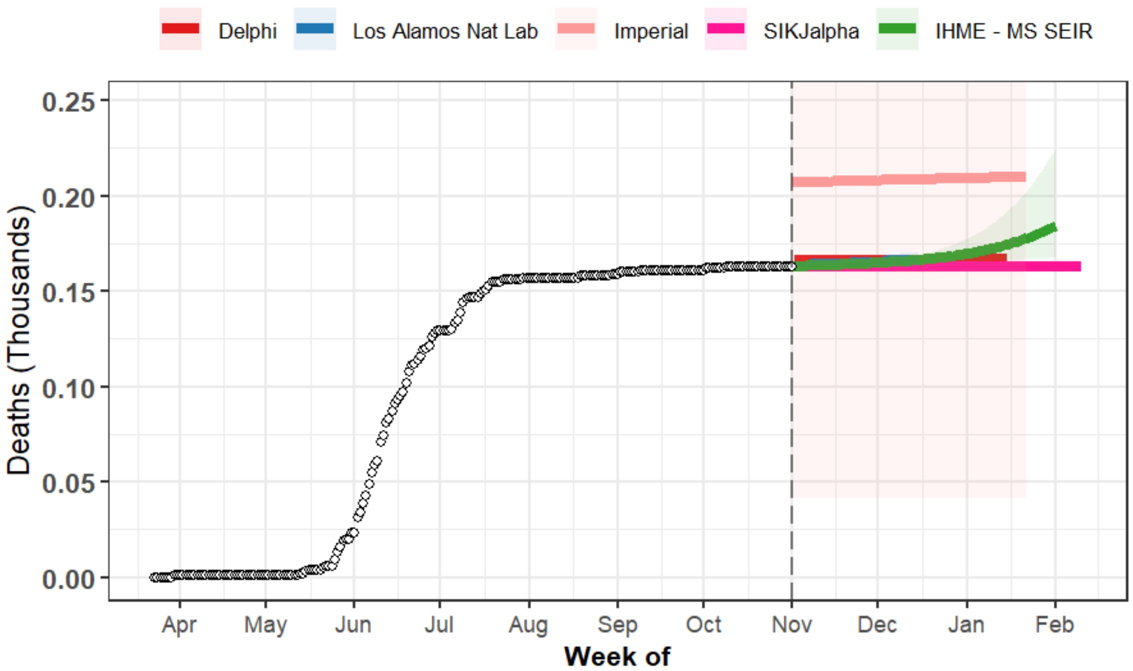


#### Cumulative Out-Of-Sample Error (Post Intercept Shift)

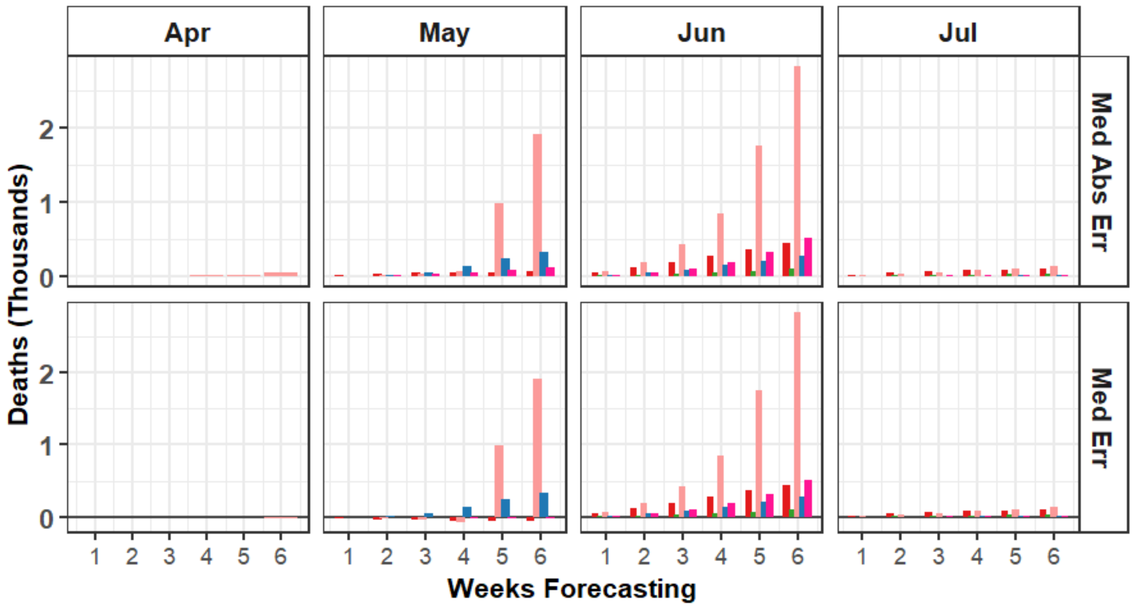


#### All Model Versions

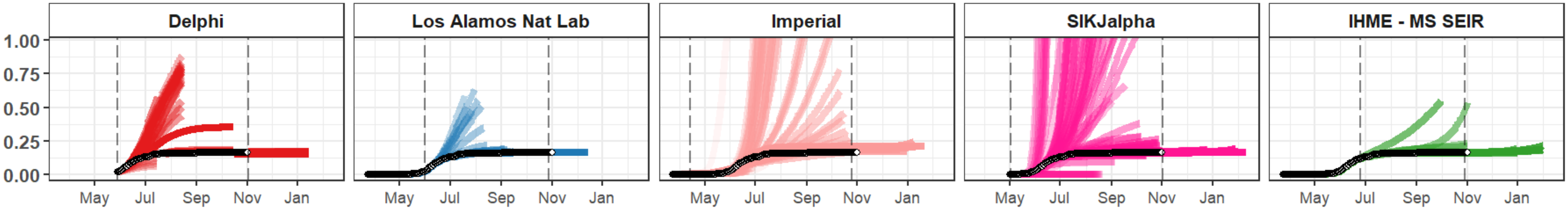


#### All Cumulative Errors

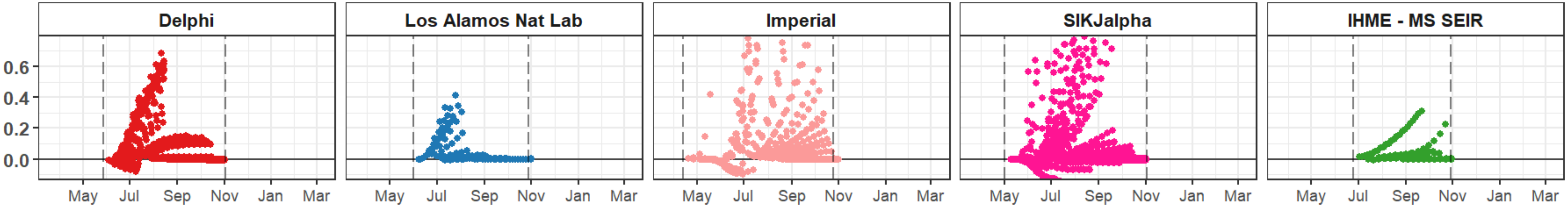

### Nicaragua

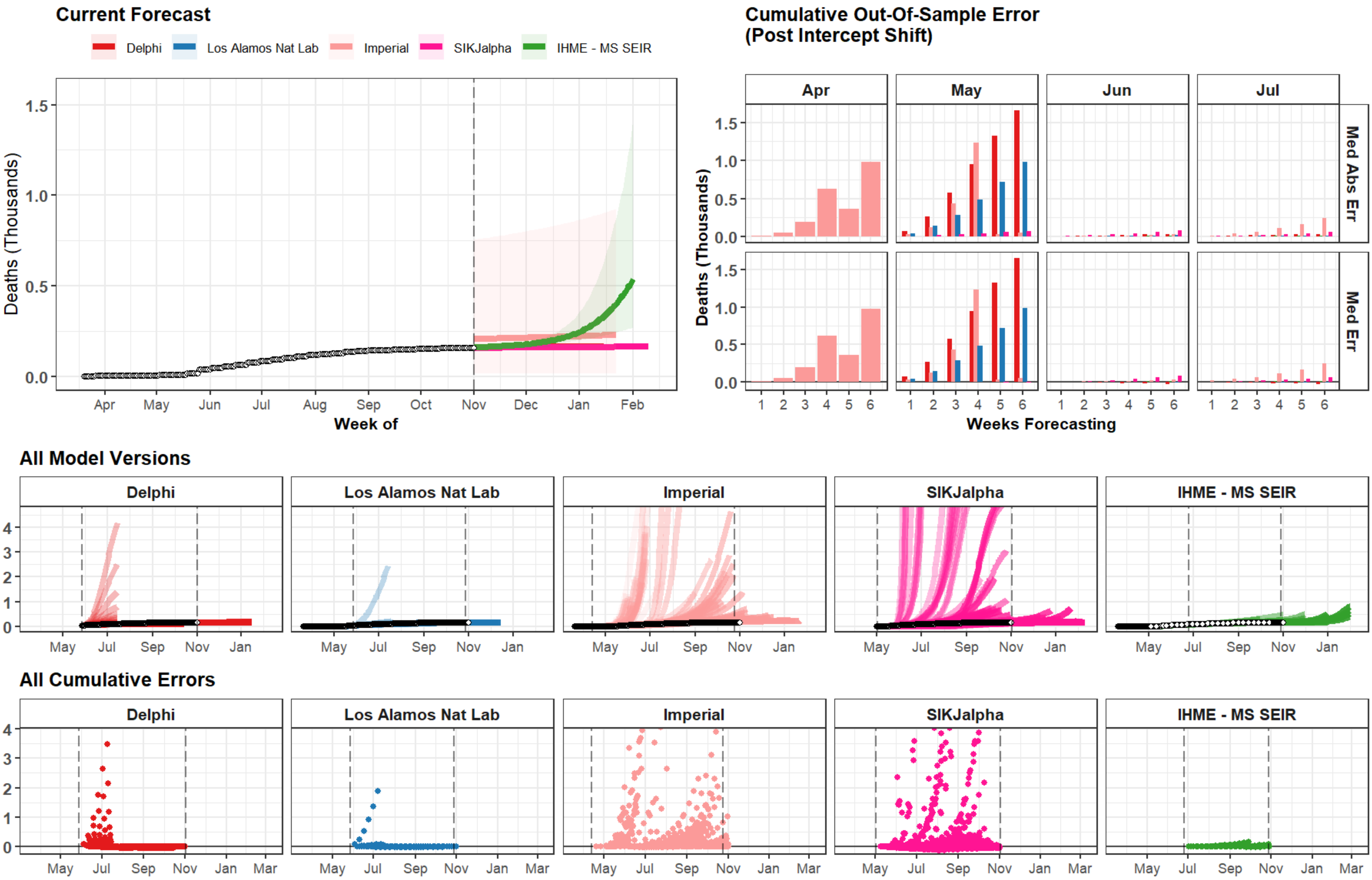
Current Forecast
Delphi
Los Alamos Nat Lab
Imperial
SIKJalpha
IHME - MS SEIR
Deaths (Thousands)
Week of
Cumulative Out-Of-Sample Error
(Post Intercept Shift)
Apr
May
Jun
Jul
Med Abs Err
Med Err
Weeks Forecasting
All Model Versions
All Cumulative Errors

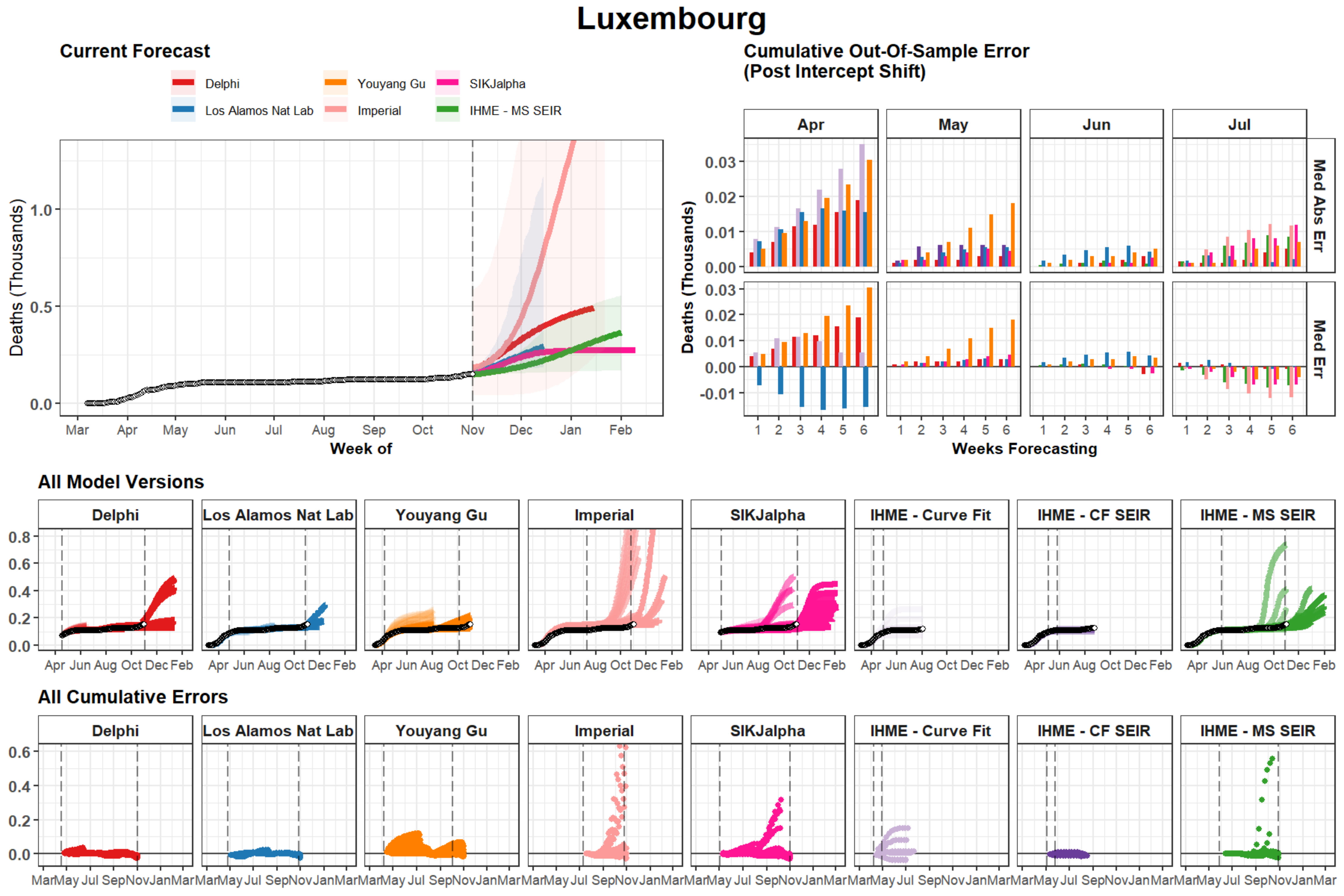

Luxembourg
Current Forecast
Delphi
Los Alamos Nat Lab
Youyang Gu
Imperial
SIKJalpha
IHME - MS SEIR
Deaths (Thousands)
Week of
Cumulative Out-Of-Sample Error (Post Intercept Shift)
Apr
May
Jun
Jul
Med Abs Err
Med Err
Weeks Forecasting
All Model Versions
Delphi
Los Alamos Nat Lab
Youyang Gu
Imperial
SIKJalpha
IHME - Curve Fit
IHME - CF SEIR
IHME - MS SEIR
All Cumulative Errors

### Maine

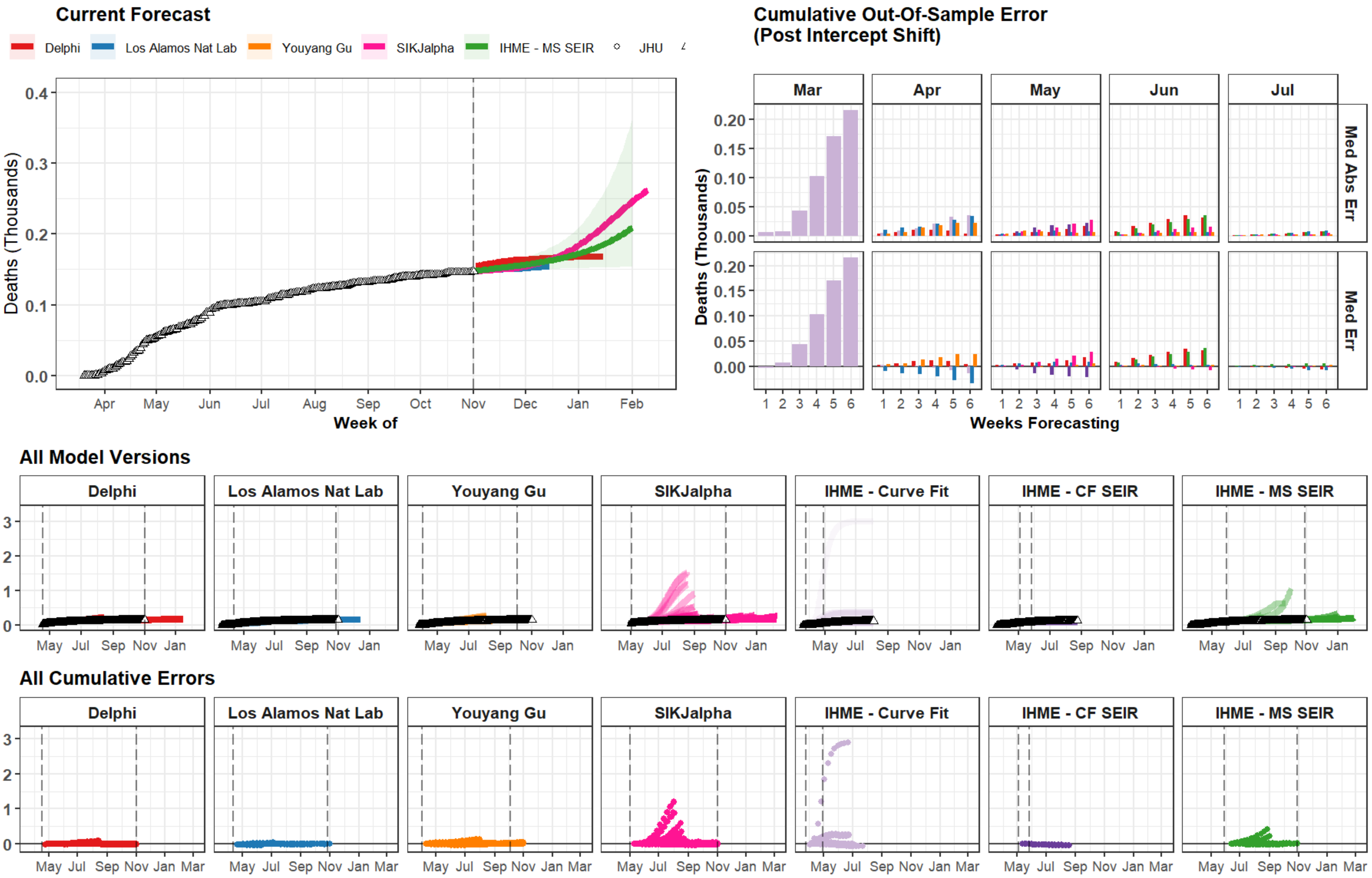
Current Forecast
Delphi
Los Alamos Nat Lab
Youyang Gu
SIKJalpha
IHME - MS SEIR
JHU
Deaths (Thousands)
Week of
Apr May Jun Jul Aug Sep Oct Nov Dec Jan Feb
Cumulative Out-Of-Sample Error
(Post Intercept Shift)
Mar Apr May Jun Jul
Med Abs Err
Med Err
Weeks Forecasting
All Model Versions
Delphi
Los Alamos Nat Lab
Youyang Gu
SIKJalpha
IHME - Curve Fit
IHME - CF SEIR
IHME - MS SEIR
May Jul Sep Nov Jan
All Cumulative Errors
May Jul Sep Nov Jan Mar

### Mali

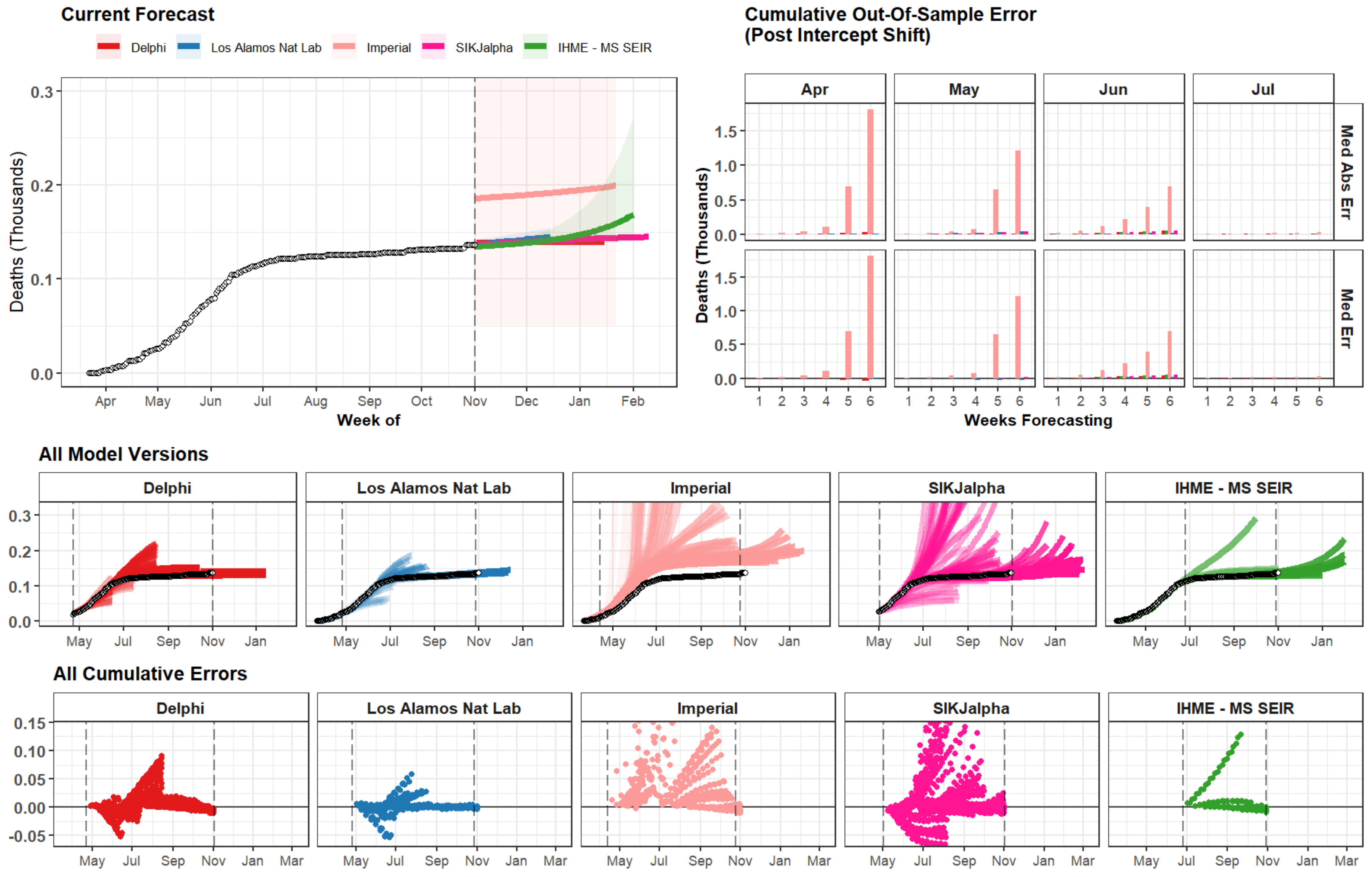

Current Forecast
Delphi
Los Alamos Nat Lab
Imperial
SIKJalpha
IHME - MS SEIR
Deaths (Thousands)
0.3
0.2
0.1
0.0
Apr
May
Jun
Jul
Aug
Sep
Oct
Nov
Dec
Jan
Feb
Week of
Cumulative Out-Of-Sample Error
(Post Intercept Shift)
Apr
May
Jun
Jul
Med Abs Err
Med Err
1.5
1.0
0.5
0.0
1
2
3
4
5
6
Weeks Forecasting
All Model Versions
Delphi
Los Alamos Nat Lab
Imperial
SIKJalpha
IHME - MS SEIR
May
Jul
Sep
Nov
Jan
All Cumulative Errors
0.15
0.10
0.05
0.00
-0.05
Mar

### Namibia

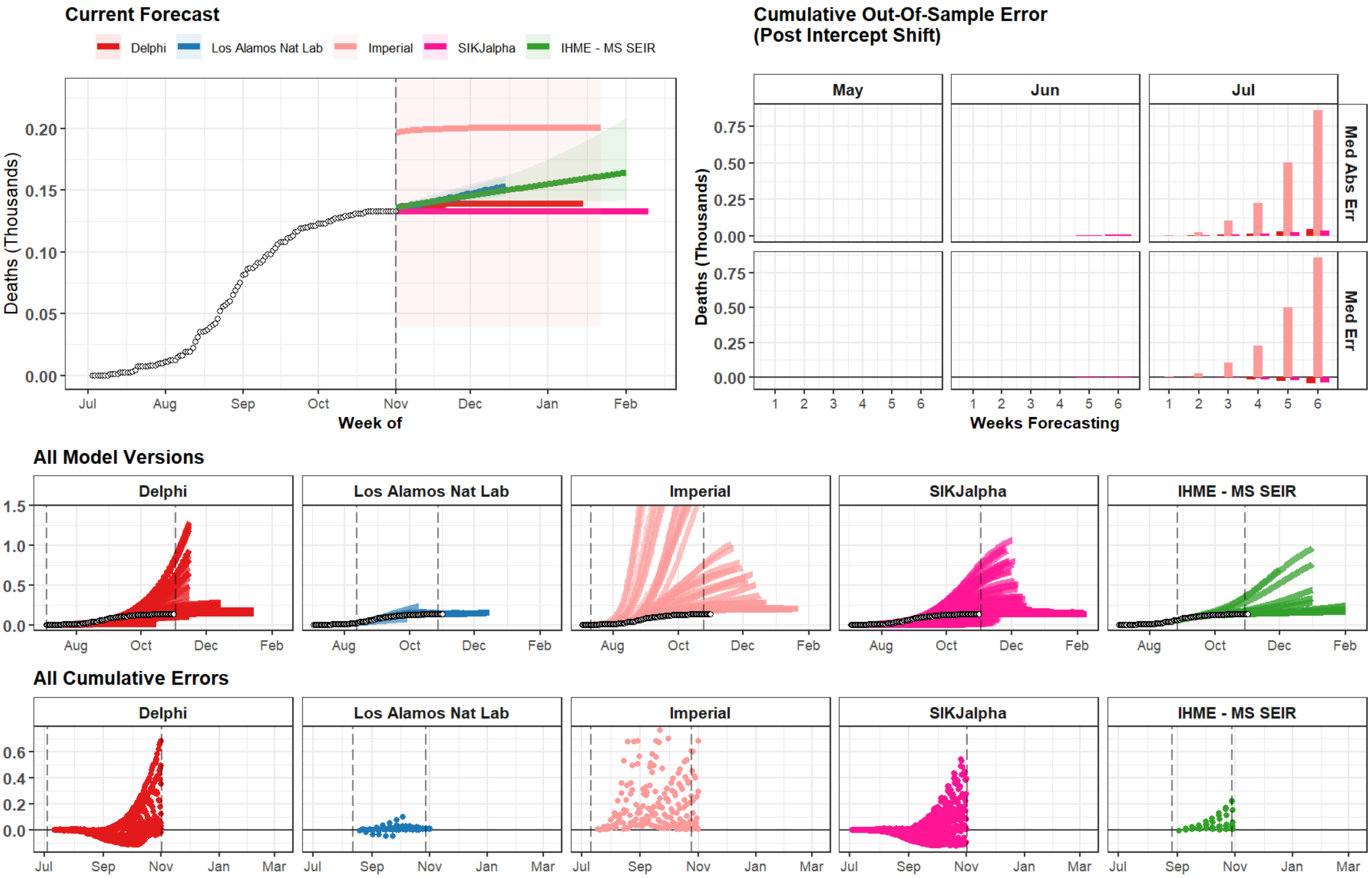

Current Forecast
Delphi
Los Alamos Nat Lab
Imperial
SIKJalpha
IHME - MS SEIR
Deaths (Thousands)
0.00
0.05
0.10
0.15
0.20
Jul
Aug
Sep
Oct
Nov
Dec
Jan
Feb
Week of
Cumulative Out-Of-Sample Error
(Post Intercept Shift)
May
Jun
Jul
Med Abs Err
Med Err
0.00
0.25
0.50
0.75
1
2
3
4
5
6
Weeks Forecasting
All Model Versions
Delphi
Los Alamos Nat Lab
Imperial
SIKJalpha
IHME - MS SEIR
0.0
0.5
1.0
1.5
Aug
Oct
Dec
Feb
All Cumulative Errors
Delphi
Los Alamos Nat Lab
Imperial
SIKJalpha
IHME - MS SEIR
0.0
0.2
0.4
0.6
Jul
Sep
Nov
Jan
Mar

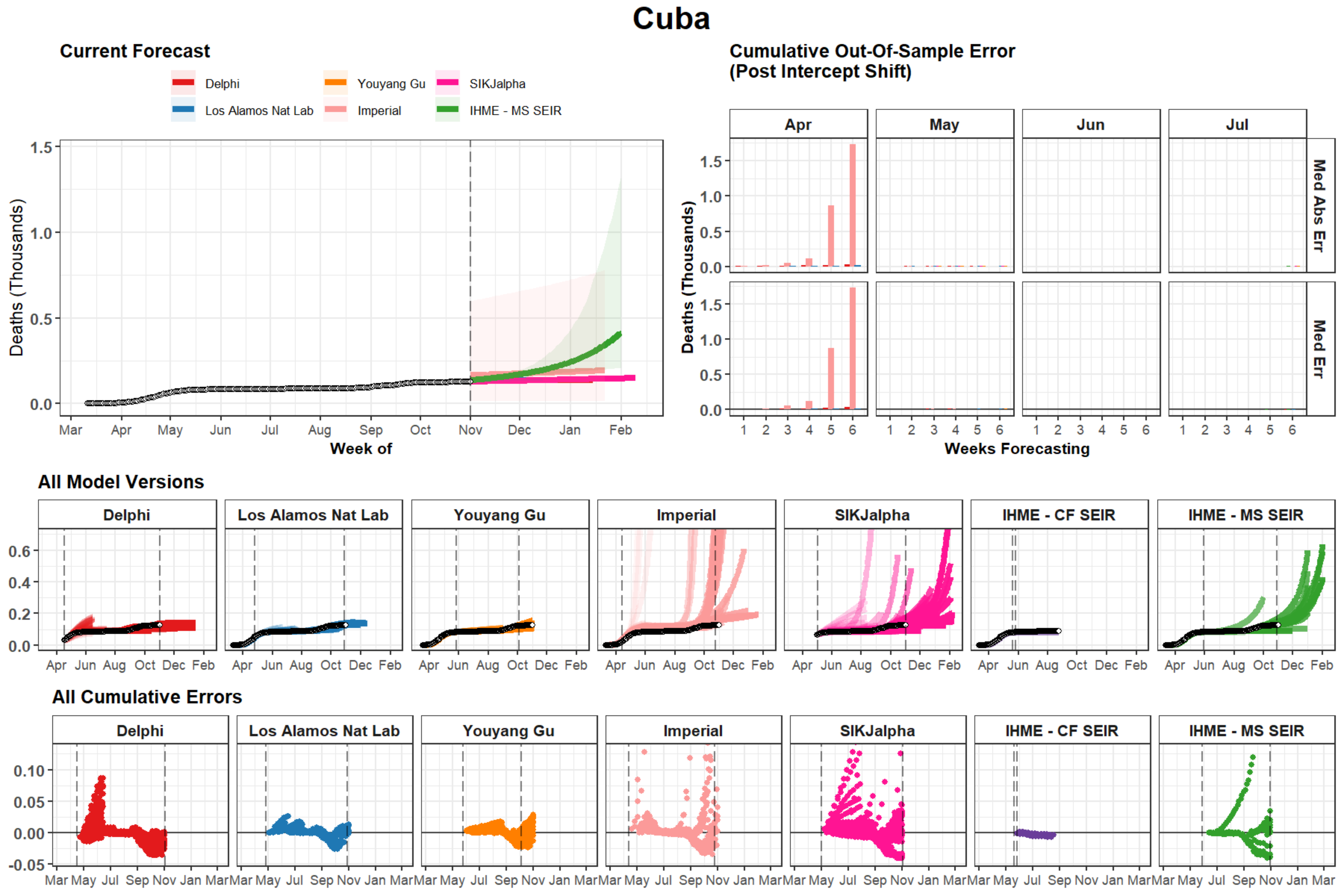

Cuba
Current Forecast
Delphi
Los Alamos Nat Lab
Youyang Gu
Imperial
SIKJalpha
IHME - MS SEIR
Deaths (Thousands)
Week of
Cumulative Out-Of-Sample Error
(Post Intercept Shift)
Apr
May
Jun
Jul
Med Abs Err
Med Err
Weeks Forecasting
All Model Versions
IHME - CF SEIR
All Cumulative Errors

### Cote d'Ivoire

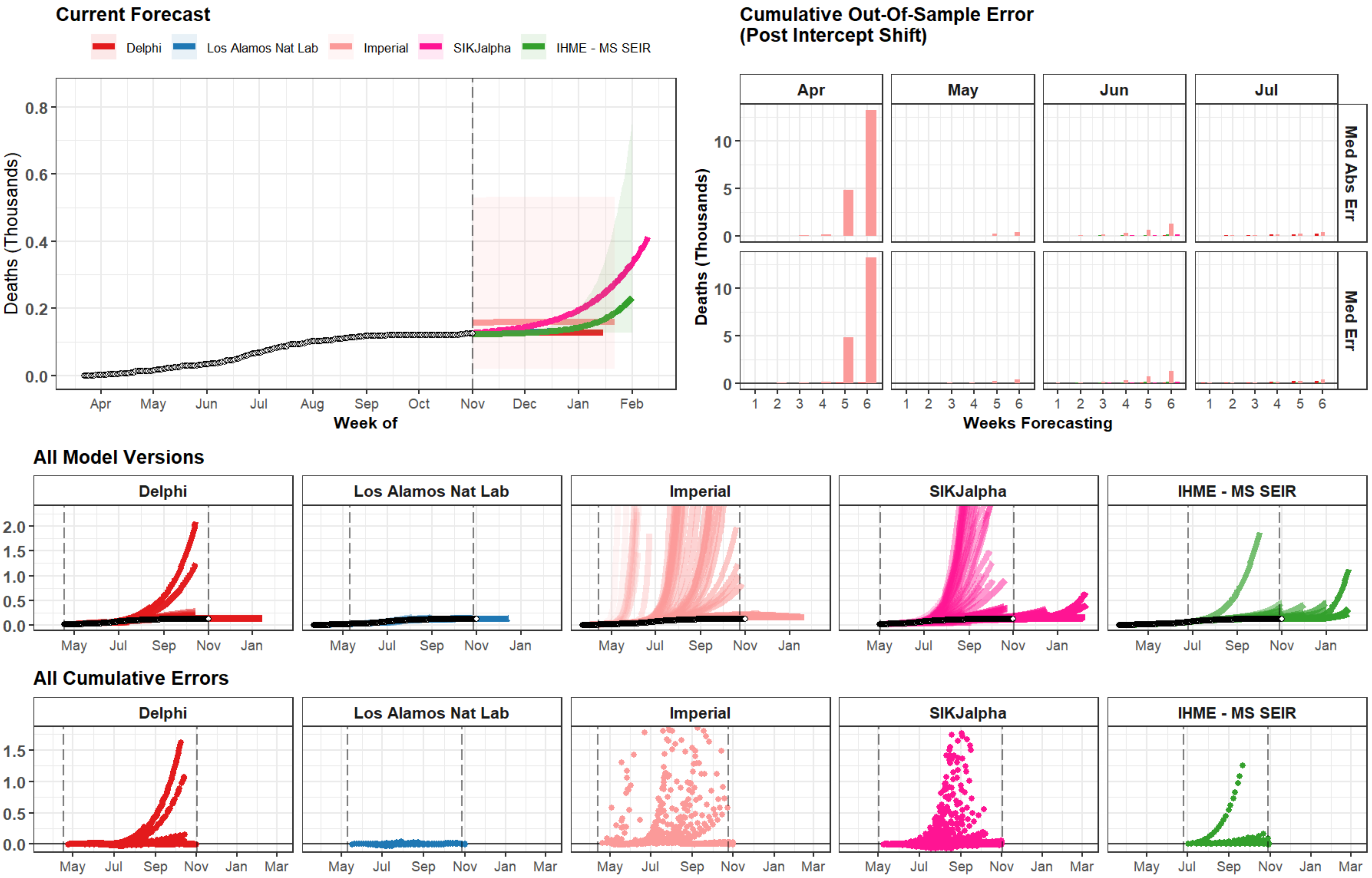

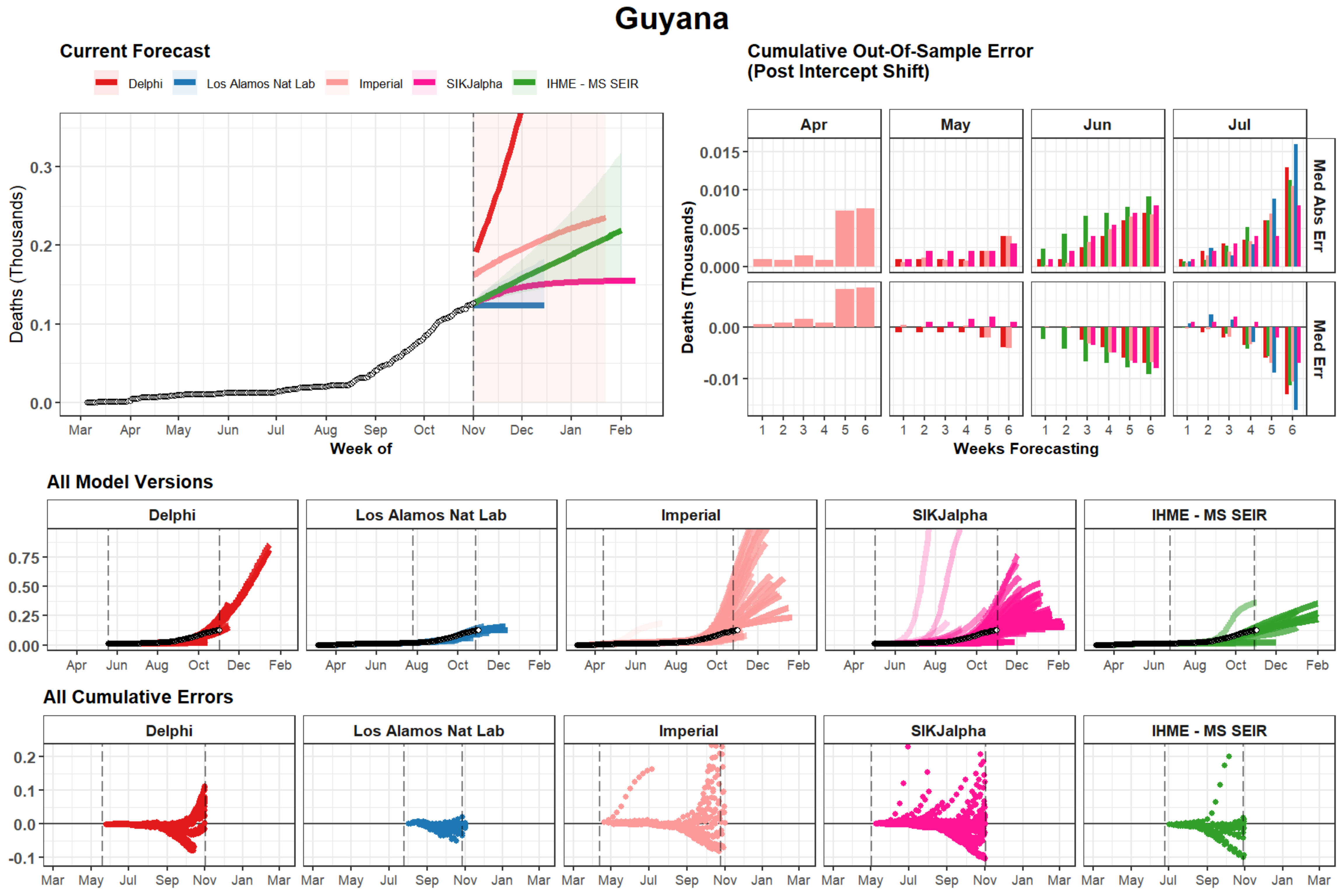

Guyana
Current Forecast
Delphi
Los Alamos Nat Lab
Imperial
SIKJalpha
IHME - MS SEIR
Deaths (Thousands)
Week of
Cumulative Out-Of-Sample Error
(Post Intercept Shift)
Apr
May
Jun
Jul
Med Abs Err
Med Err
Weeks Forecasting
All Model Versions
All Cumulative Errors

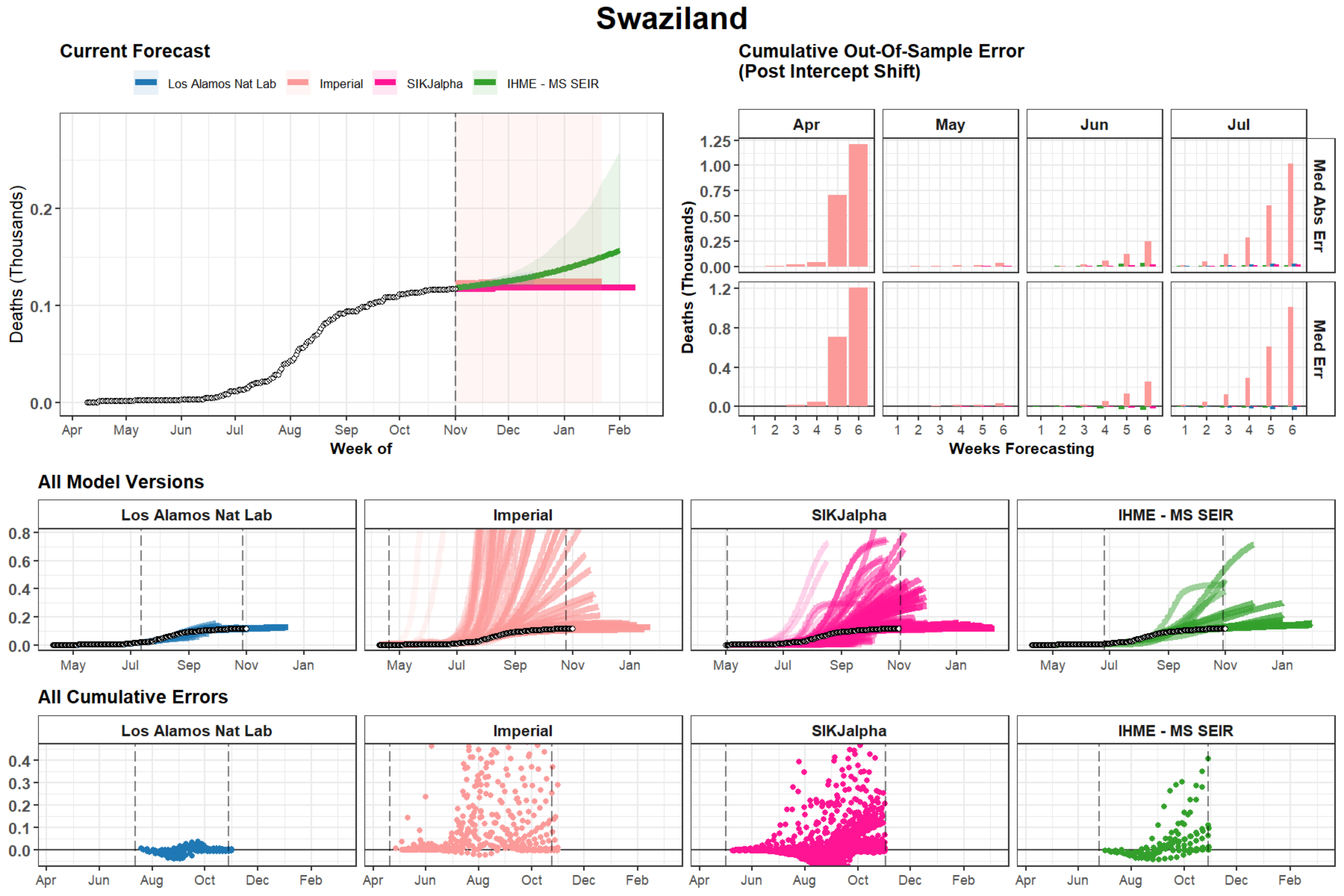

Swaziland
Current Forecast
Los Alamos Nat Lab
Imperial
SIKJalpha
IHME - MS SEIR
Deaths (Thousands)
0.0
0.1
0.2
Apr
May
Jun
Jul
Aug
Sep
Oct
Nov
Dec
Jan
Feb
Week of
Cumulative Out-Of-Sample Error
(Post Intercept Shift)
Apr
May
Jun
Jul
Med Abs Err
Med Err
0.00
0.25
0.50
0.75
1.00
1.25
0.0
0.4
0.8
1.2
1
2
3
4
5
6
Weeks Forecasting
All Model Versions
Los Alamos Nat Lab
Imperial
SIKJalpha
IHME - MS SEIR
0.0
0.2
0.4
0.6
0.8
May
Jul
Sep
Nov
Jan
All Cumulative Errors
Los Alamos Nat Lab
Imperial
SIKJalpha
IHME - MS SEIR
0.0
0.1
0.2
0.3
0.4
Apr
Jun
Aug
Oct
Dec
Feb

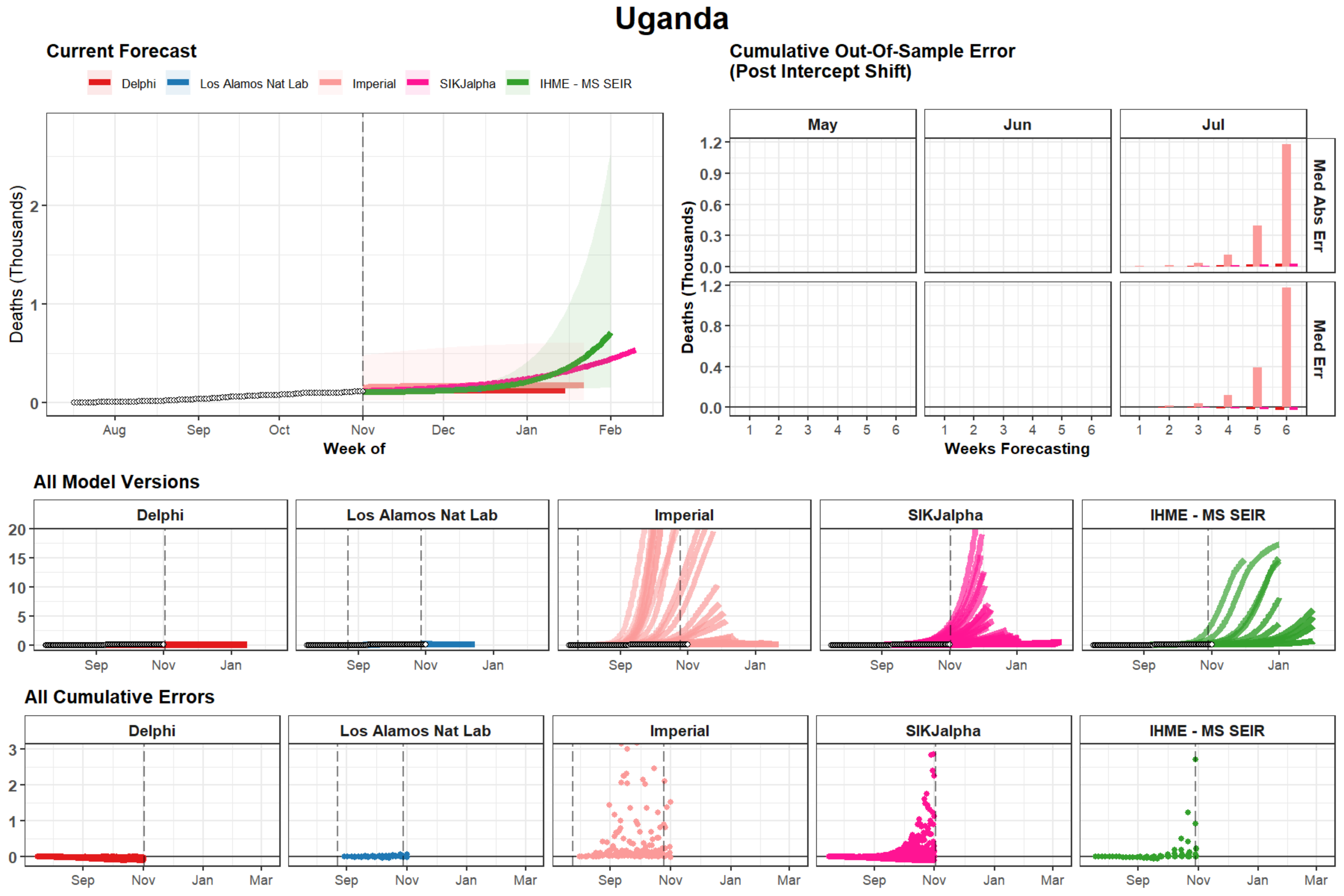

Uganda
Current Forecast
Delphi
Los Alamos Nat Lab
Imperial
SIKJalpha
IHME - MS SEIR
Deaths (Thousands)
Week of
Aug
Sep
Oct
Nov
Dec
Jan
Feb
Cumulative Out-Of-Sample Error
(Post Intercept Shift)
May
Jun
Jul
Med Abs Err
Med Err
Weeks Forecasting
All Model Versions
All Cumulative Errors
Mar

### Suriname

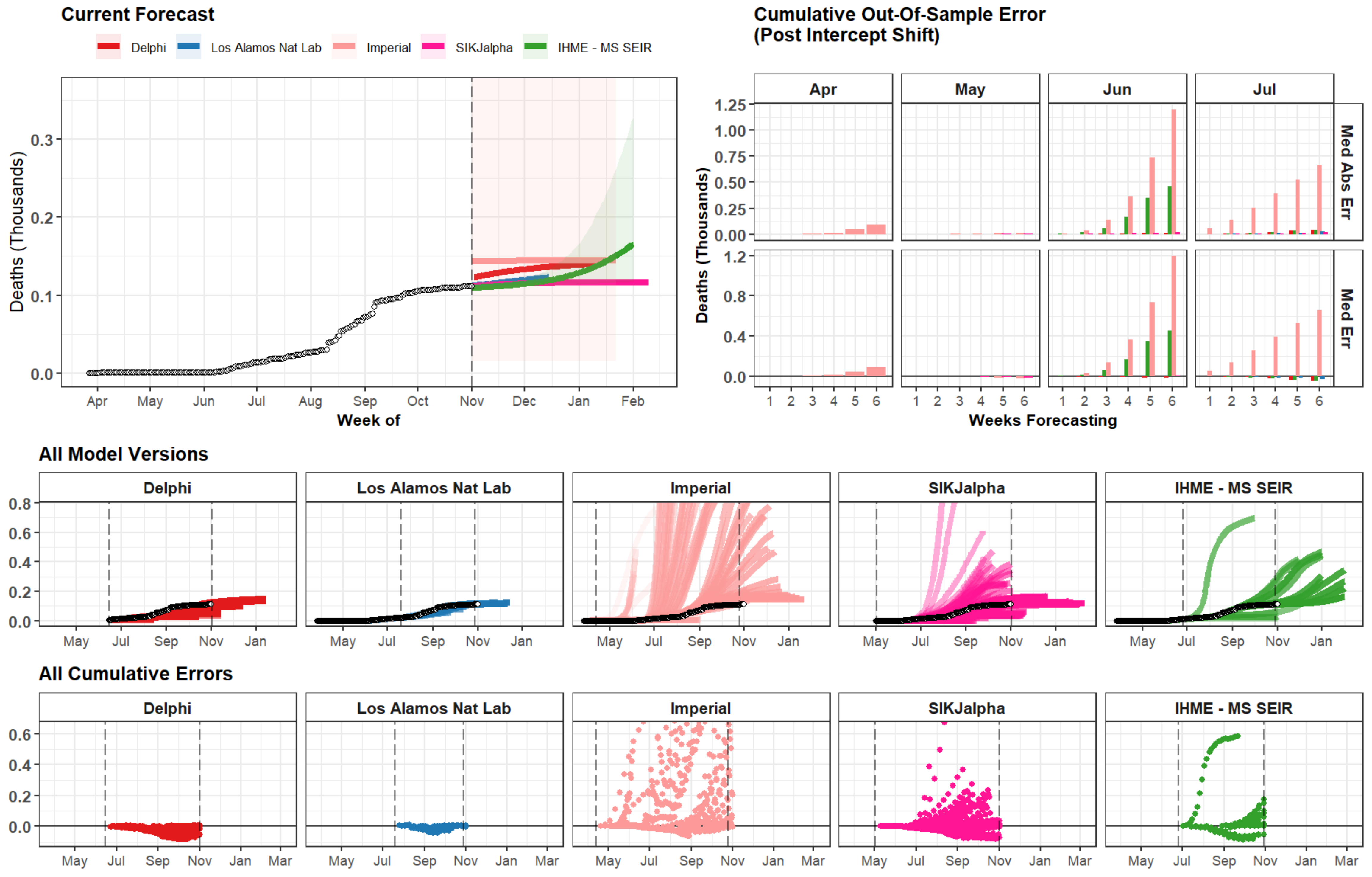

Current Forecast
Delphi
Los Alamos Nat Lab
Imperial
SIKJalpha
IHME - MS SEIR
Deaths (Thousands)
Week of
Apr
May
Jun
Jul
Aug
Sep
Oct
Nov
Dec
Jan
Feb
Cumulative Out-Of-Sample Error
(Post Intercept Shift)
Med Abs Err
Med Err
Weeks Forecasting
All Model Versions
All Cumulative Errors
Mar

### Trinidad and Tobago

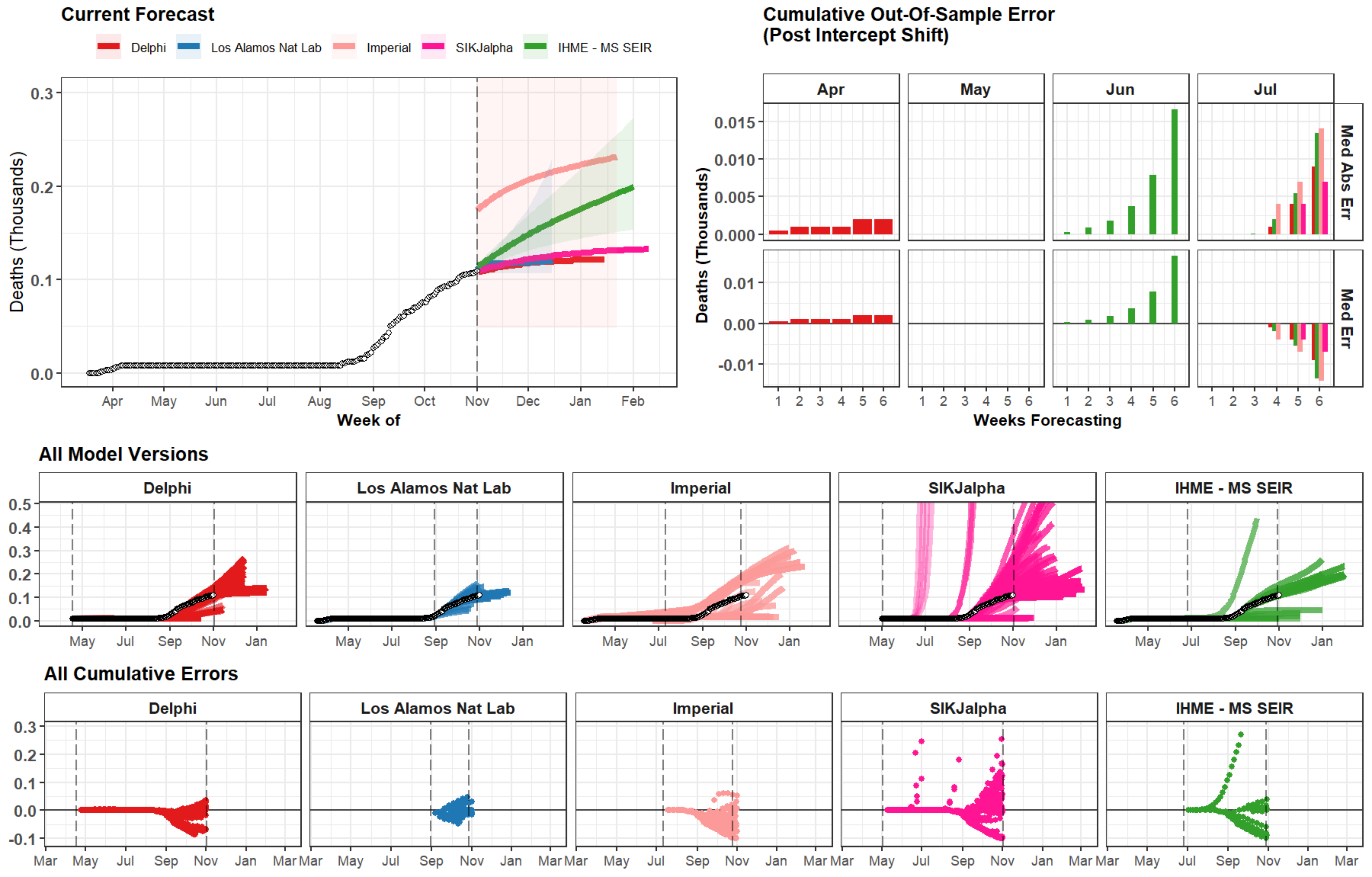

### Somalia

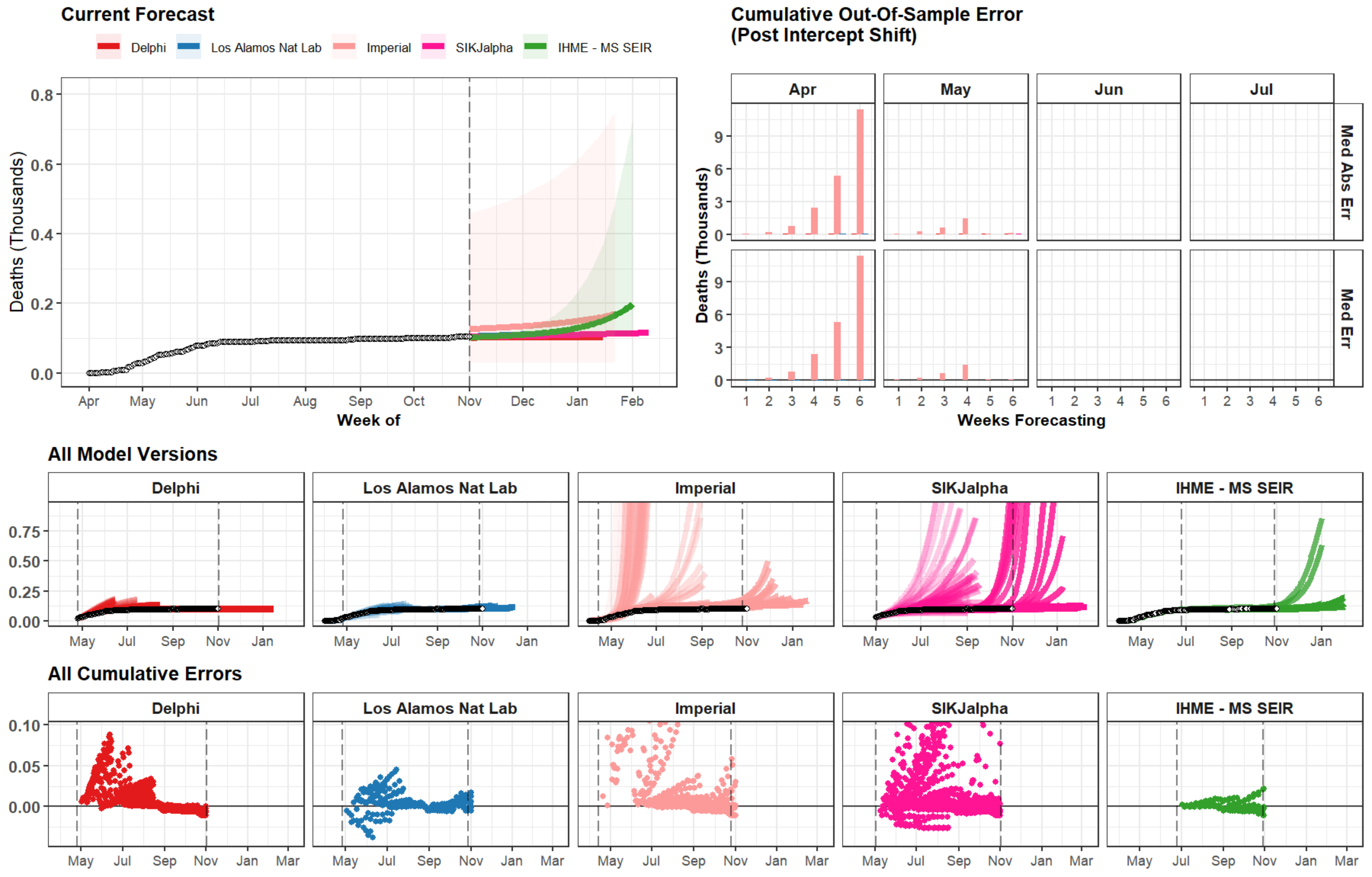
Current Forecast
Delphi
Los Alamos Nat Lab
Imperial
SIKJalpha
IHME - MS SEIR
Deaths (Thousands)
0.0
0.2
0.4
0.6
0.8
Apr
May
Jun
Jul
Aug
Sep
Oct
Nov
Dec
Jan
Feb
Week of
Cumulative Out-Of-Sample Error
(Post Intercept Shift)
Apr
May
Jun
Jul
Med Abs Err
Med Err
0
3
6
9
1 2 3 4 5 6
Weeks Forecasting
All Model Versions
Delphi
Los Alamos Nat Lab
Imperial
SIKJalpha
IHME - MS SEIR
0.00
0.25
0.50
0.75
May
Jul
Sep
Nov
Jan
All Cumulative Errors
0.00
0.05
0.10
Mar

### Chad

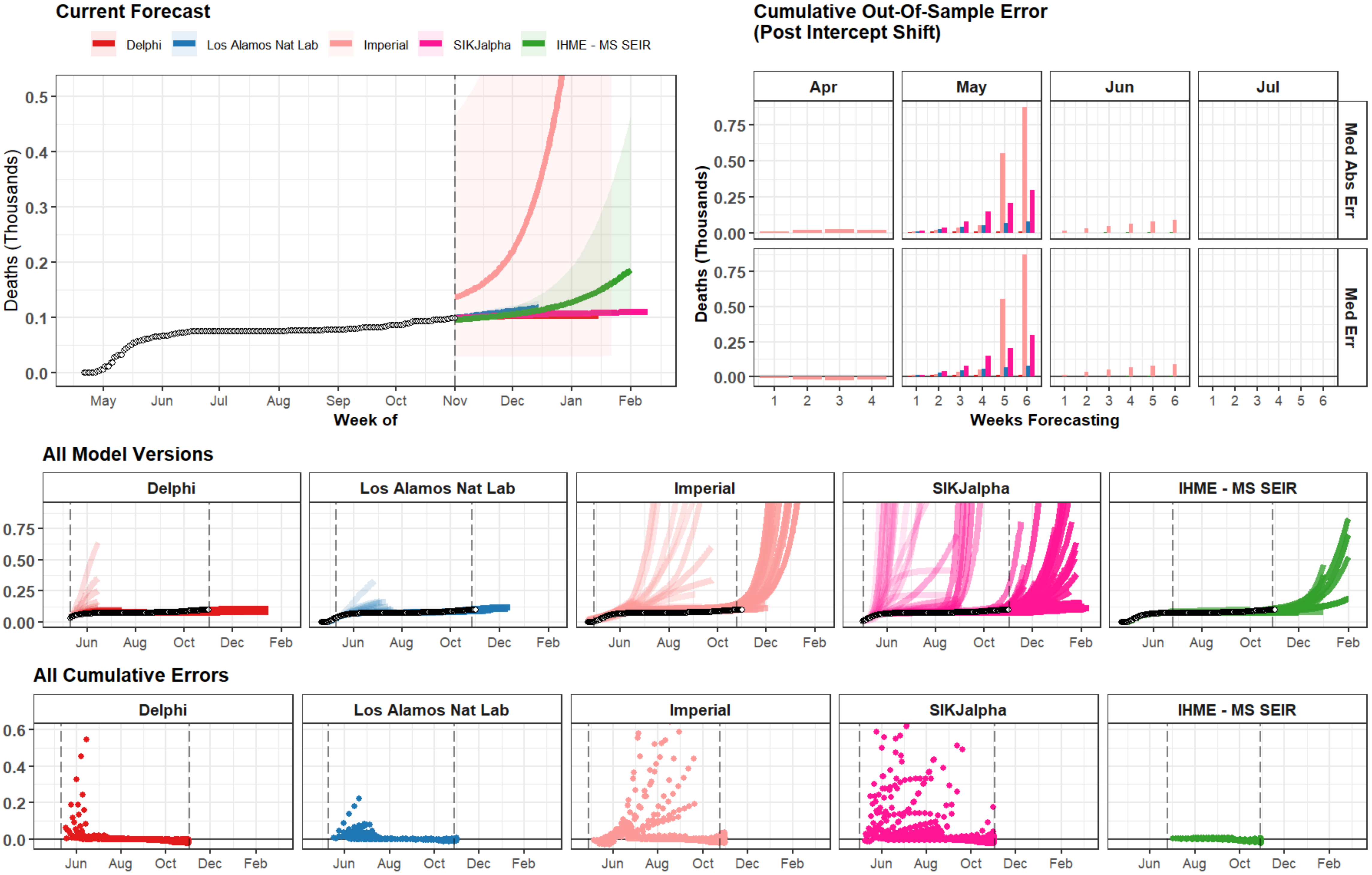
Current Forecast
Delphi
Los Alamos Nat Lab
Imperial
SIKJalpha
IHME - MS SEIR
Deaths (Thousands)
Week of
Cumulative Out-Of-Sample Error
(Post Intercept Shift)
Apr
May
Jun
Jul
Med Abs Err
Med Err
Weeks Forecasting
All Model Versions
All Cumulative Errors

### Cape Verde

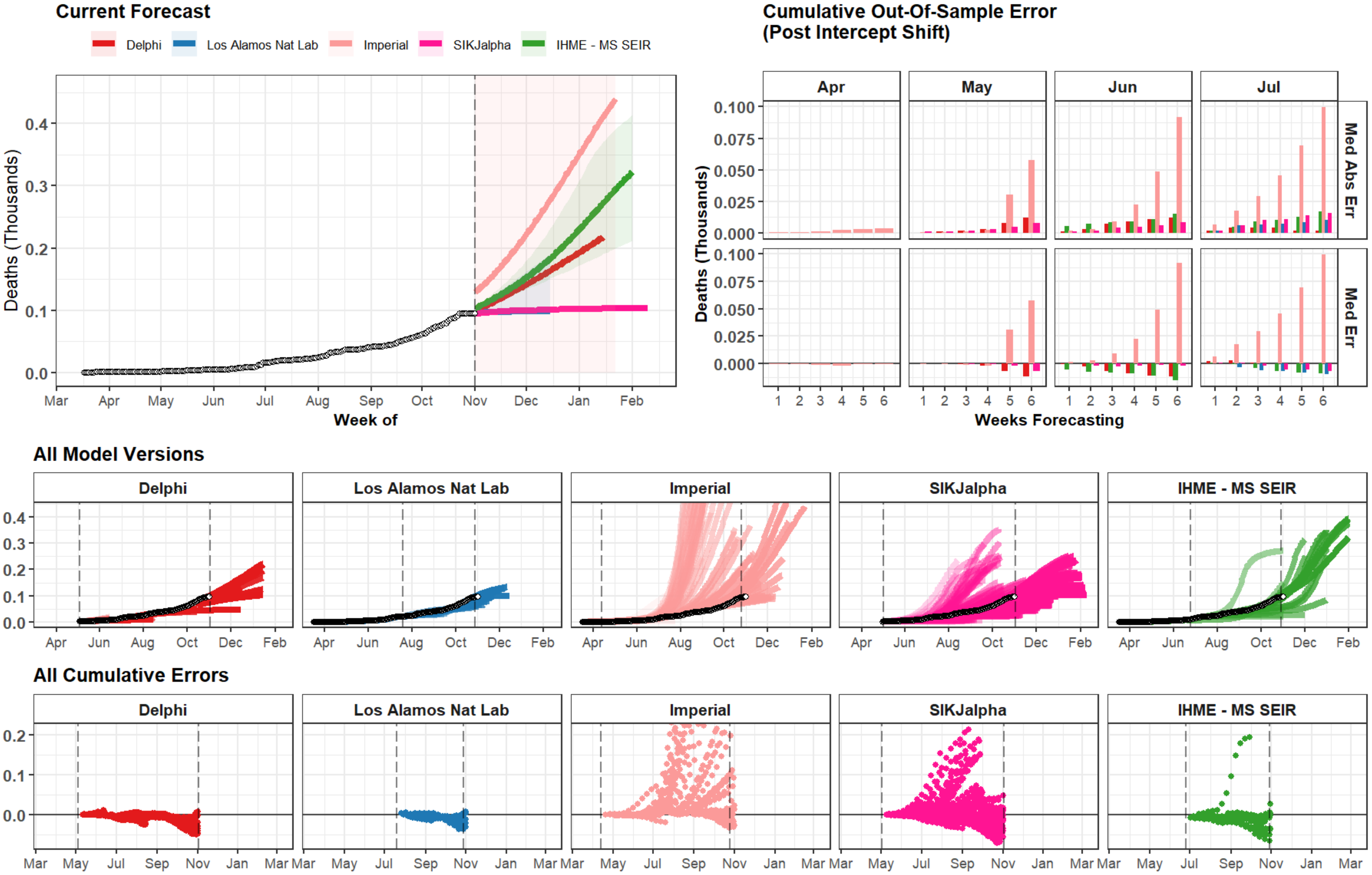
Current Forecast
Delphi
Los Alamos Nat Lab
Imperial
SIKJalpha
IHME - MS SEIR
Deaths (Thousands)
Week of
Cumulative Out-Of-Sample Error
(Post Intercept Shift)
Apr
May
Jun
Jul
Med Abs Err
Med Err
Weeks Forecasting
All Model Versions
All Cumulative Errors

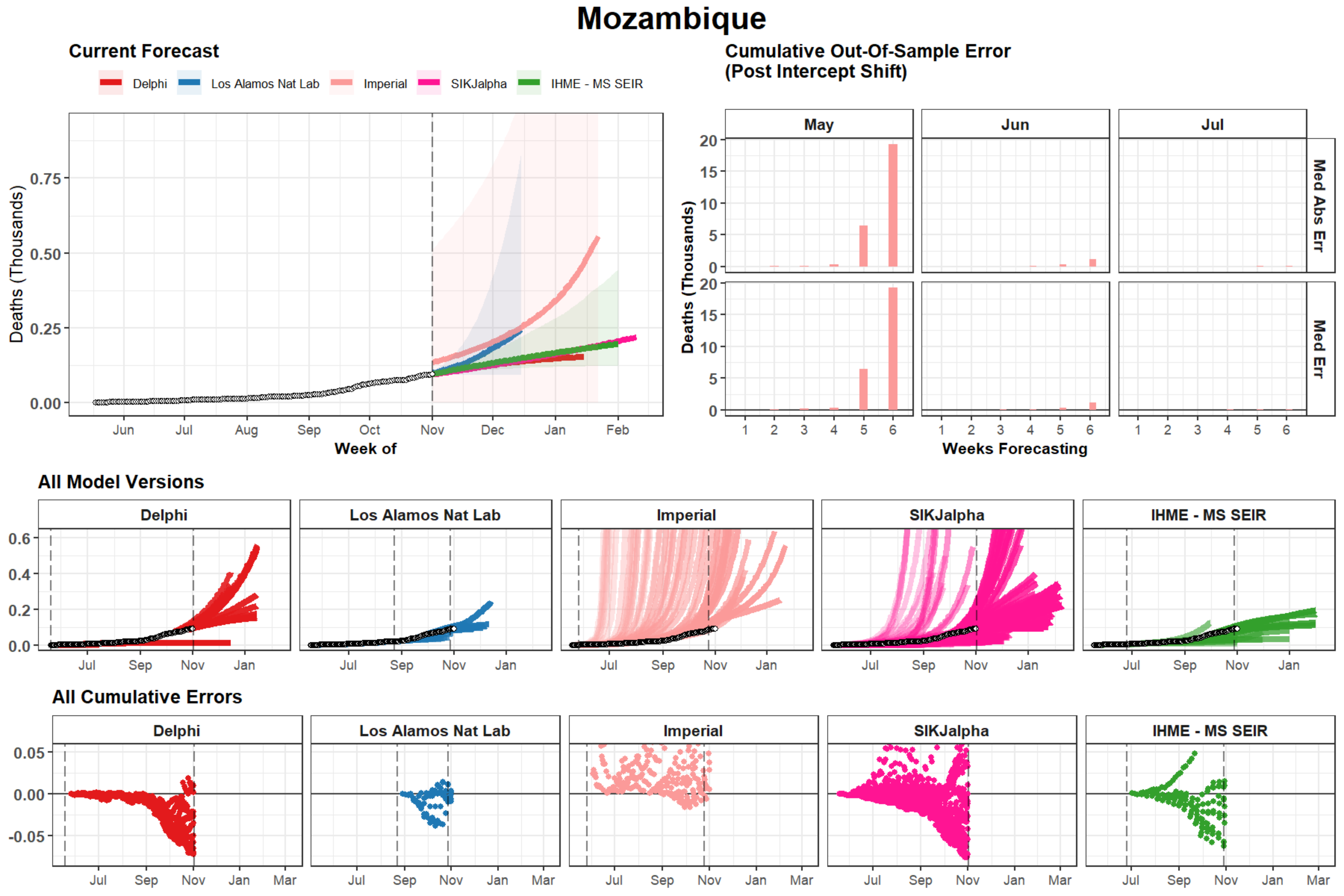
Mozambique
Current Forecast
Delphi
Los Alamos Nat Lab
Imperial
SIKJalpha
IHME - MS SEIR
Deaths (Thousands)
0.00
0.25
0.50
0.75
Jun
Jul
Aug
Sep
Oct
Nov
Dec
Jan
Feb
Week of
Cumulative Out-Of-Sample Error
(Post Intercept Shift)
May
Jun
Jul
Med Abs Err
Med Err
Deaths (Thousands)
0
5
10
15
20
1
2
3
4
5
6
Weeks Forecasting
All Model Versions
Delphi
Los Alamos Nat Lab
Imperial
SIKJalpha
IHME - MS SEIR
0.0
0.2
0.4
0.6
Jul
Sep
Nov
Jan
All Cumulative Errors
Delphi
Los Alamos Nat Lab
Imperial
SIKJalpha
IHME - MS SEIR
-0.05
0.00
0.05
Jul
Sep
Nov
Jan
Mar

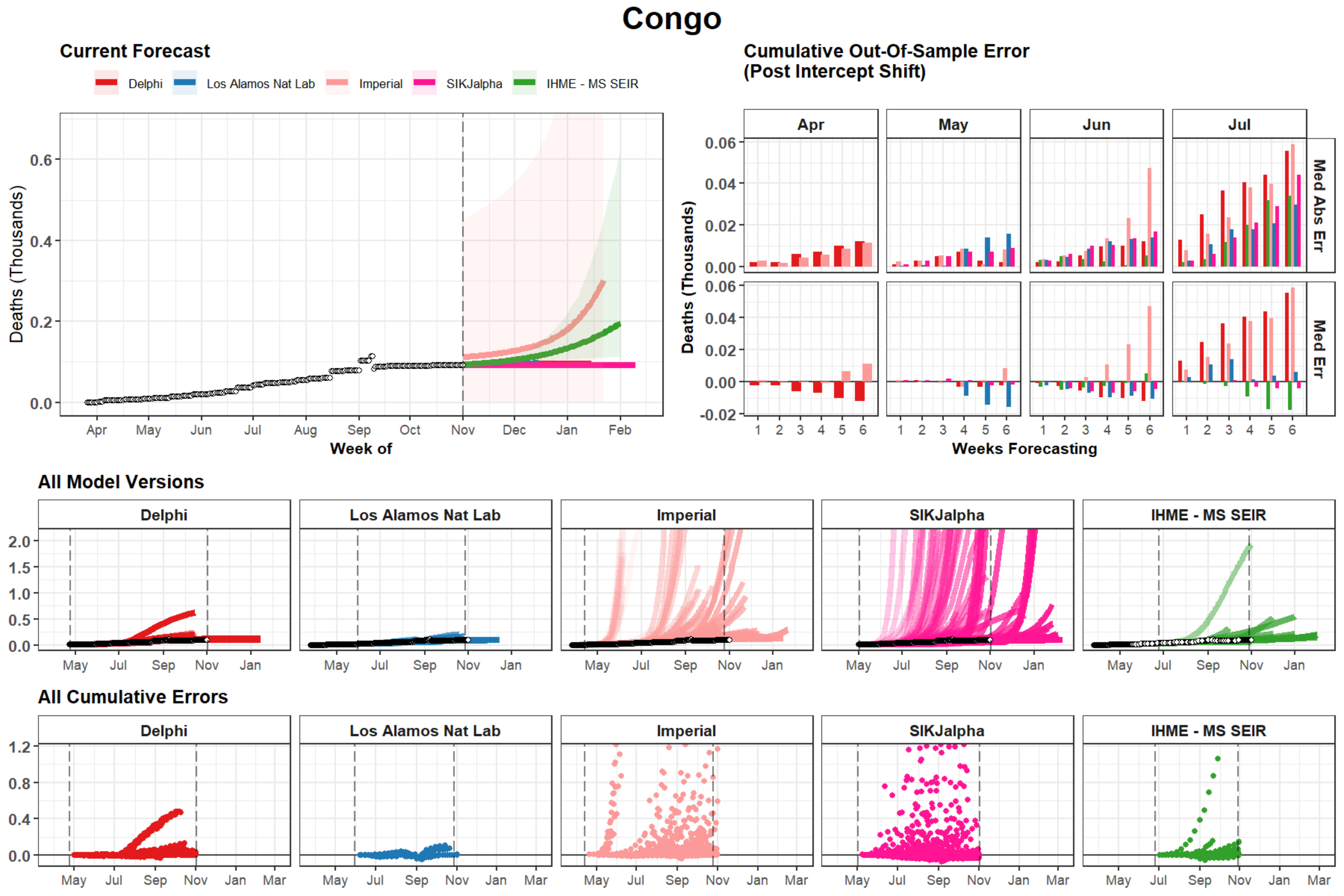

Congo
Current Forecast
Delphi
Los Alamos Nat Lab
Imperial
SIKJalpha
IHME - MS SEIR
Deaths (Thousands)
Week of
Cumulative Out-Of-Sample Error
(Post Intercept Shift)
Apr
May
Jun
Jul
Med Abs Err
Med Err
Weeks Forecasting
All Model Versions
All Cumulative Errors

### Wyoming

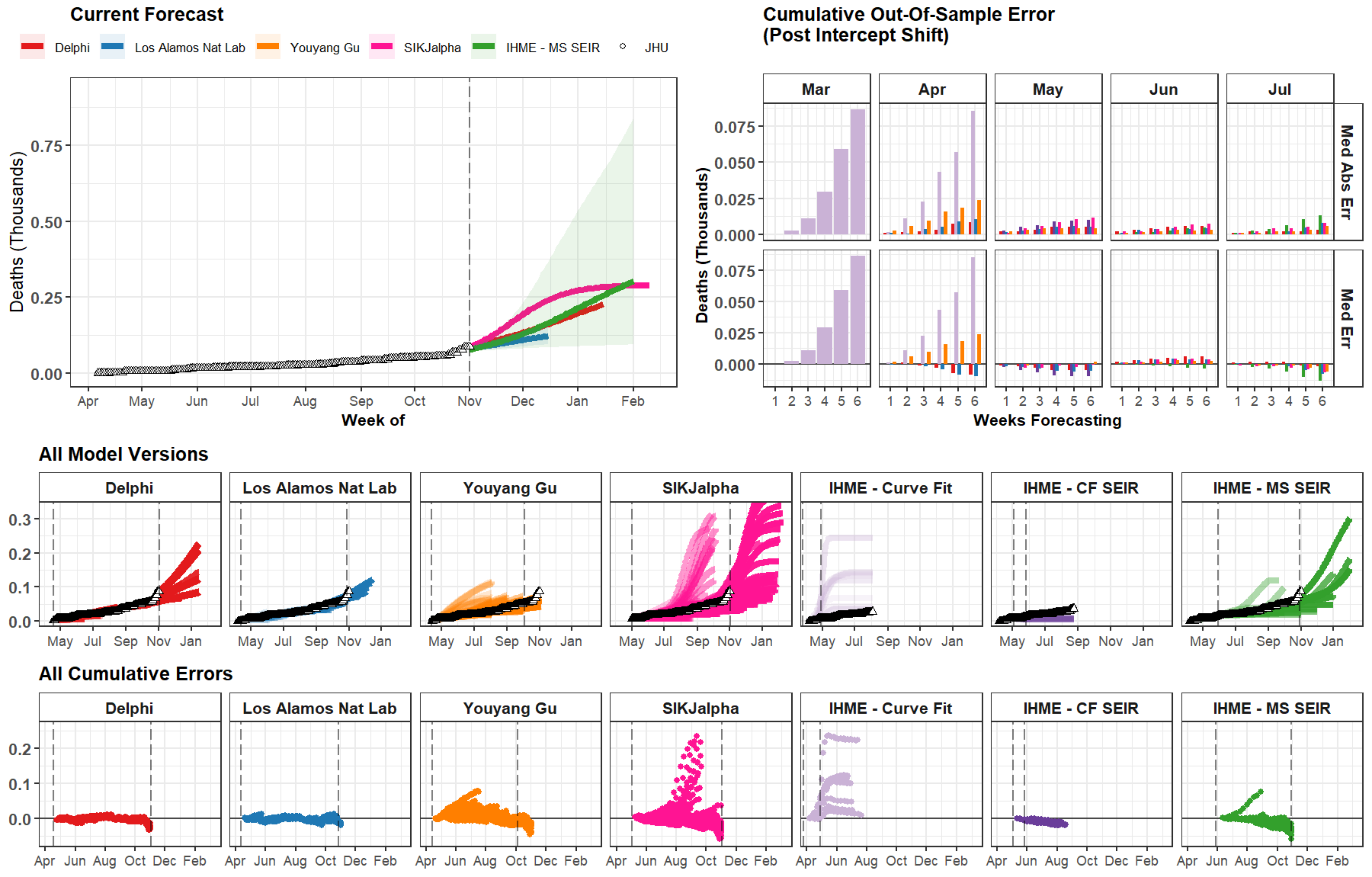
Current Forecast
Delphi
Los Alamos Nat Lab
Youyang Gu
SIKJalpha
IHME - MS SEIR
JHU
Deaths (Thousands)
Week of
Apr May Jun Jul Aug Sep Oct Nov Dec Jan Feb
Cumulative Out-Of-Sample Error
(Post Intercept Shift)
Mar Apr May Jun Jul
Med Abs Err
Med Err
Weeks Forecasting
All Model Versions
Delphi
Los Alamos Nat Lab
Youyang Gu
SIKJalpha
IHME - Curve Fit
IHME - CF SEIR
IHME - MS SEIR
All Cumulative Errors

### Equatorial Guinea

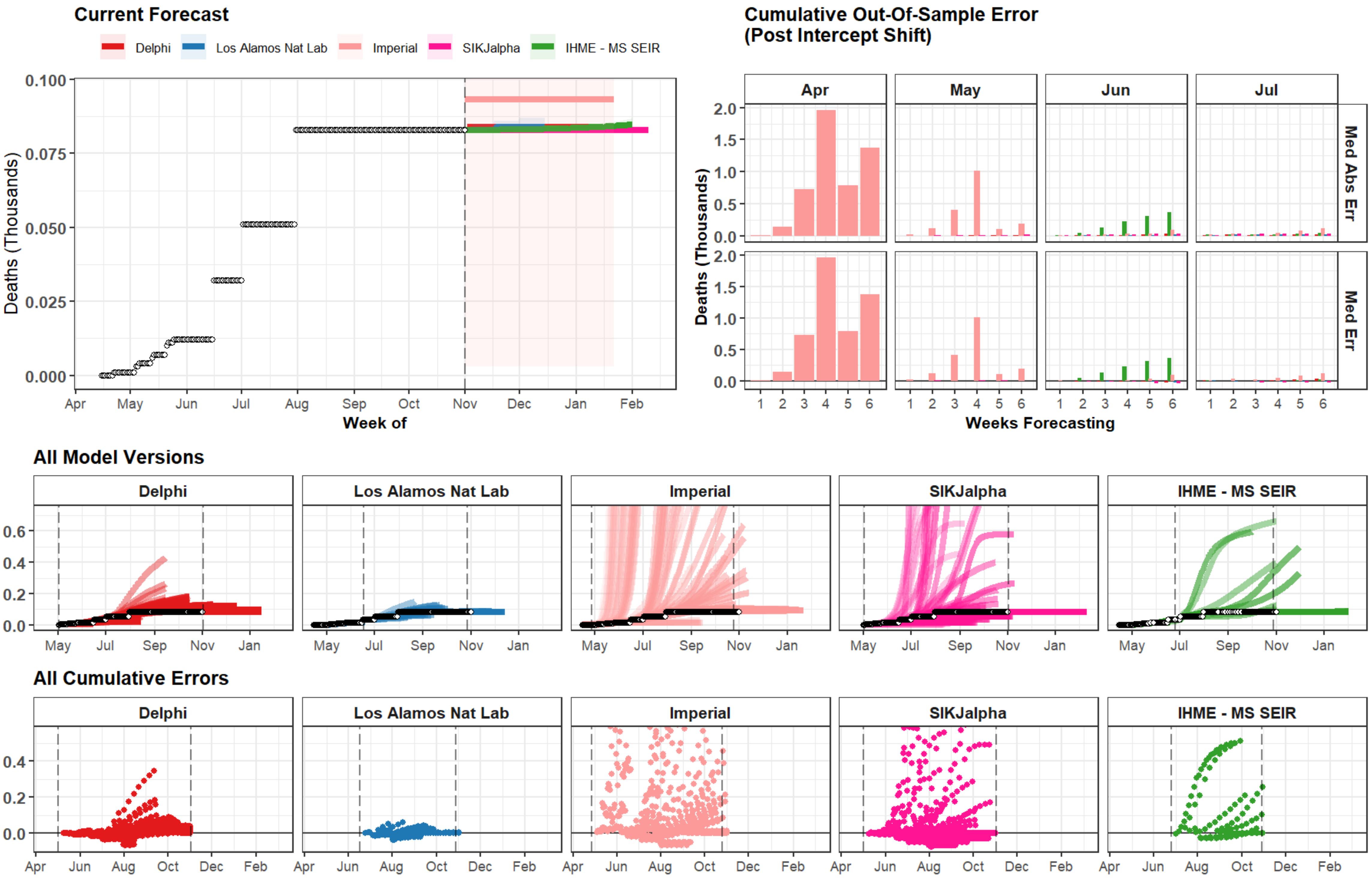

### Liberia

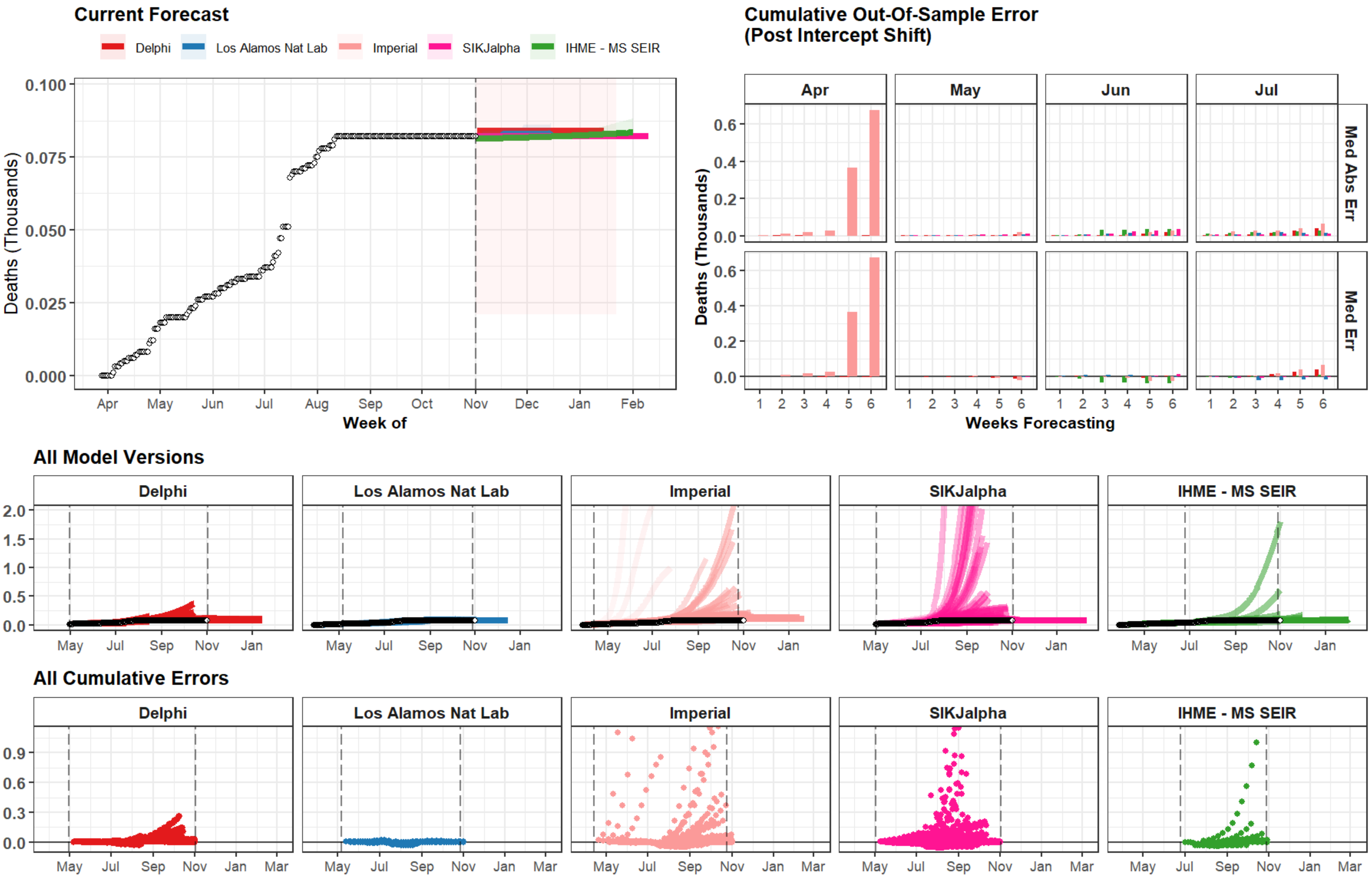

Current Forecast
Delphi
Los Alamos Nat Lab
Imperial
SIKJalpha
IHME - MS SEIR
Deaths (Thousands)
0.100
0.075
0.050
0.025
0.000
Apr
May
Jun
Jul
Aug
Sep
Oct
Nov
Dec
Jan
Feb
Week of
Cumulative Out-Of-Sample Error
(Post Intercept Shift)
Apr
May
Jun
Jul
Med Abs Err
Med Err
Deaths (Thousands)
0.6
0.4
0.2
0.0
1
2
3
4
5
6
Weeks Forecasting
All Model Versions
Delphi
Los Alamos Nat Lab
Imperial
SIKJalpha
IHME - MS SEIR
2.0
1.5
1.0
0.5
0.0
May
Jul
Sep
Nov
Jan
All Cumulative Errors
Delphi
Los Alamos Nat Lab
Imperial
SIKJalpha
IHME - MS SEIR
0.9
0.6
0.3
0.0
May
Jul
Sep
Nov
Jan
Mar

### Tajikistan

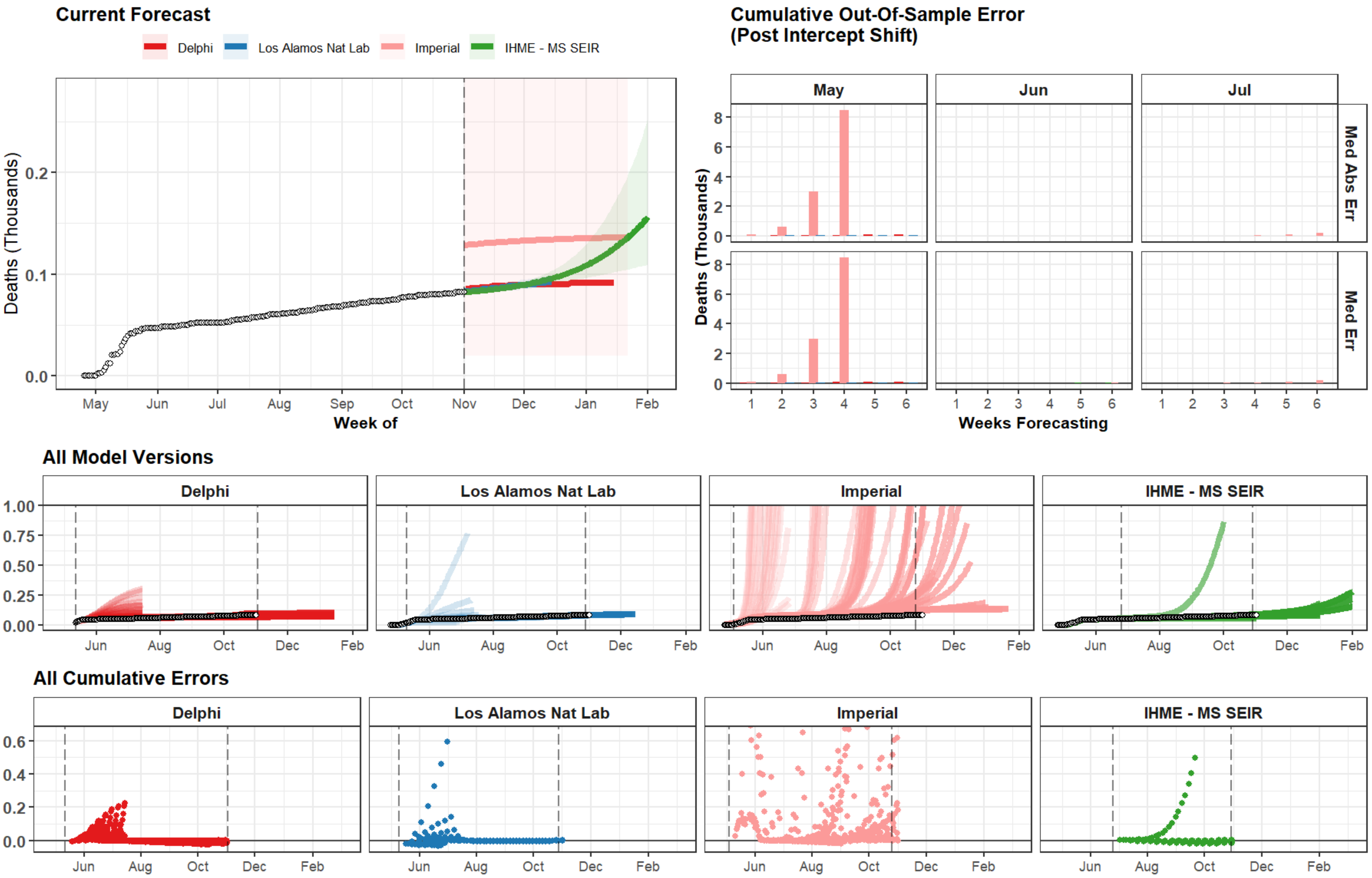

Current Forecast
Delphi
Los Alamos Nat Lab
Imperial
IHME - MS SEIR
Deaths (Thousands)
0.0
0.1
0.2
May
Jun
Jul
Aug
Sep
Oct
Nov
Dec
Jan
Feb
Week of
Cumulative Out-Of-Sample Error
(Post Intercept Shift)
May
Jun
Jul
Med Abs Err
Med Err
Deaths (Thousands)
0
2
4
6
8
1
2
3
4
5
6
Weeks Forecasting
All Model Versions
Delphi
Los Alamos Nat Lab
Imperial
IHME - MS SEIR
0.00
0.25
0.50
0.75
1.00
Jun
Aug
Oct
Dec
Feb
All Cumulative Errors
Delphi
Los Alamos Nat Lab
Imperial
IHME - MS SEIR
0.0
0.2
0.4
0.6
Jun
Aug
Oct
Dec
Feb

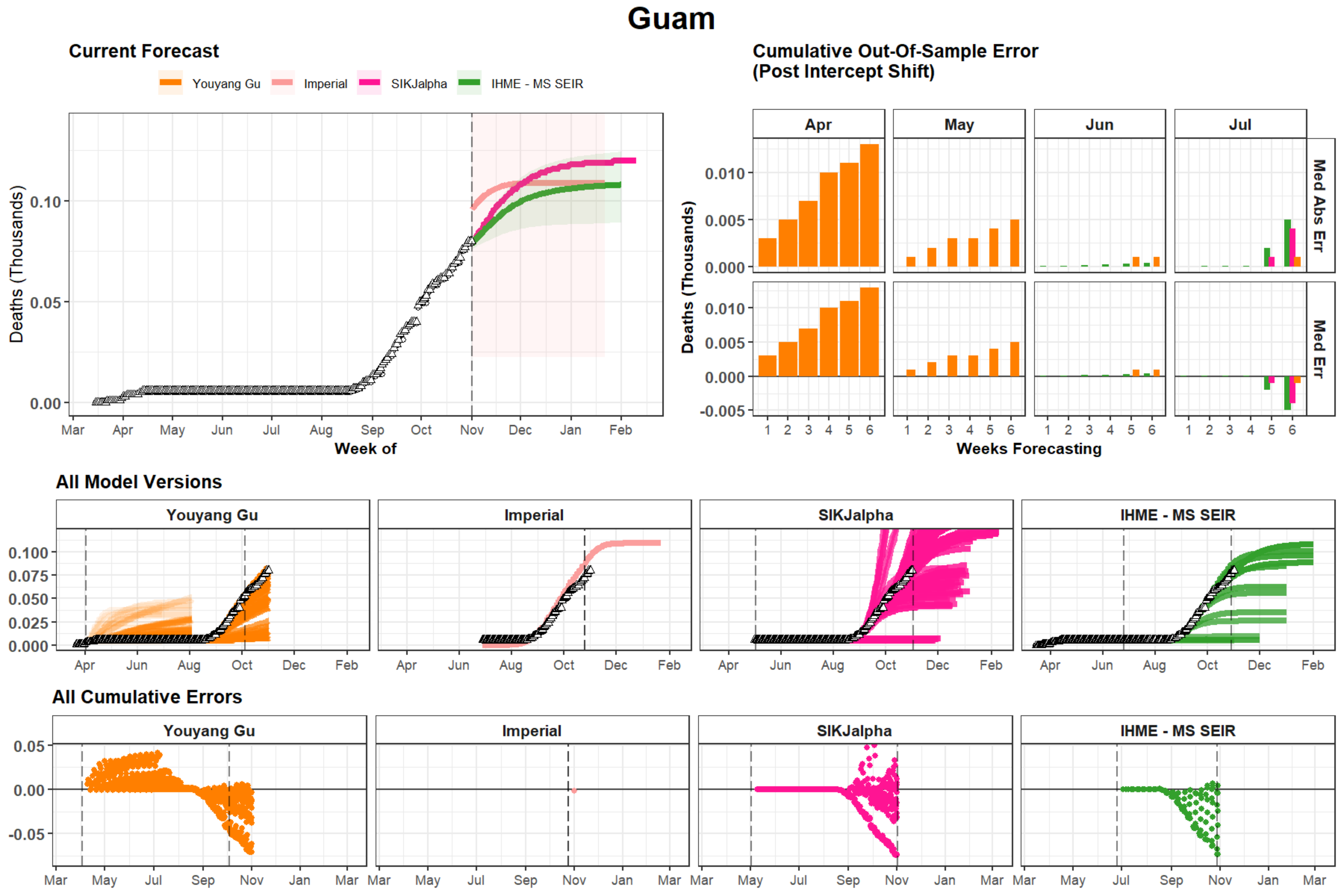

Guam
Current Forecast
Youyang Gu
Imperial
SIKJalpha
IHME - MS SEIR
Deaths (Thousands)
0.10
0.05
0.00
Mar
Apr
May
Jun
Jul
Aug
Sep
Oct
Nov
Dec
Jan
Feb
Week of
Cumulative Out-Of-Sample Error
(Post Intercept Shift)
Apr
May
Jun
Jul
Med Abs Err
Med Err
0.010
0.005
0.000
-0.005
1
2
3
4
5
6
Weeks Forecasting
All Model Versions
0.100
0.075
0.050
0.025
0.000
All Cumulative Errors
0.05
0.00
-0.05

### Alaska

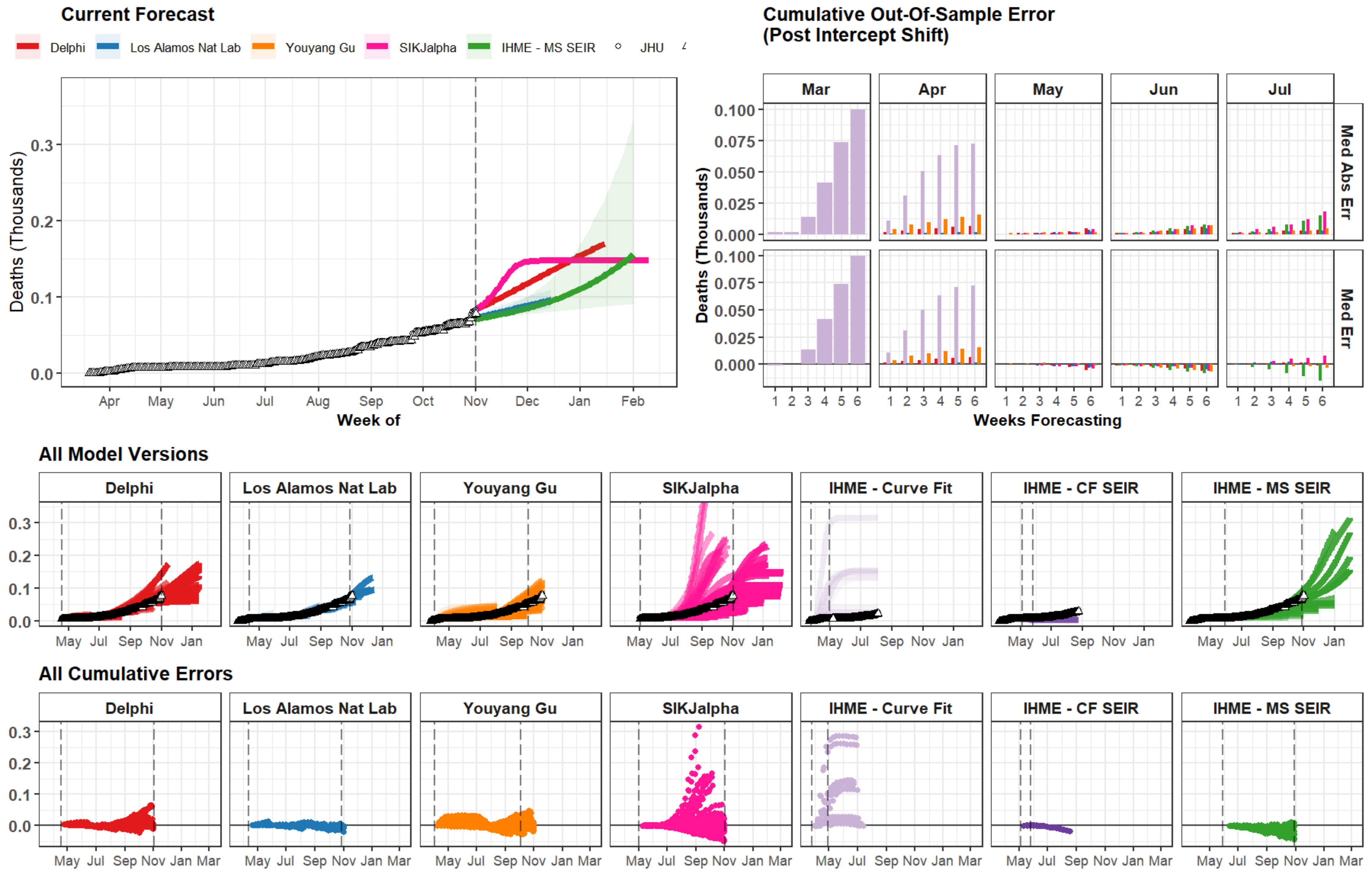
Current Forecast
Delphi
Los Alamos Nat Lab
Youyang Gu
SIKJalpha
IHME - MS SEIR
JHU
Deaths (Thousands)
Week of
Apr May Jun Jul Aug Sep Oct Nov Dec Jan Feb
Cumulative Out-Of-Sample Error
(Post Intercept Shift)
Mar Apr May Jun Jul
Med Abs Err
Med Err
Weeks Forecasting
All Model Versions
Delphi
Los Alamos Nat Lab
Youyang Gu
SIKJalpha
IHME - Curve Fit
IHME - CF SEIR
IHME - MS SEIR
All Cumulative Errors

### Andorra

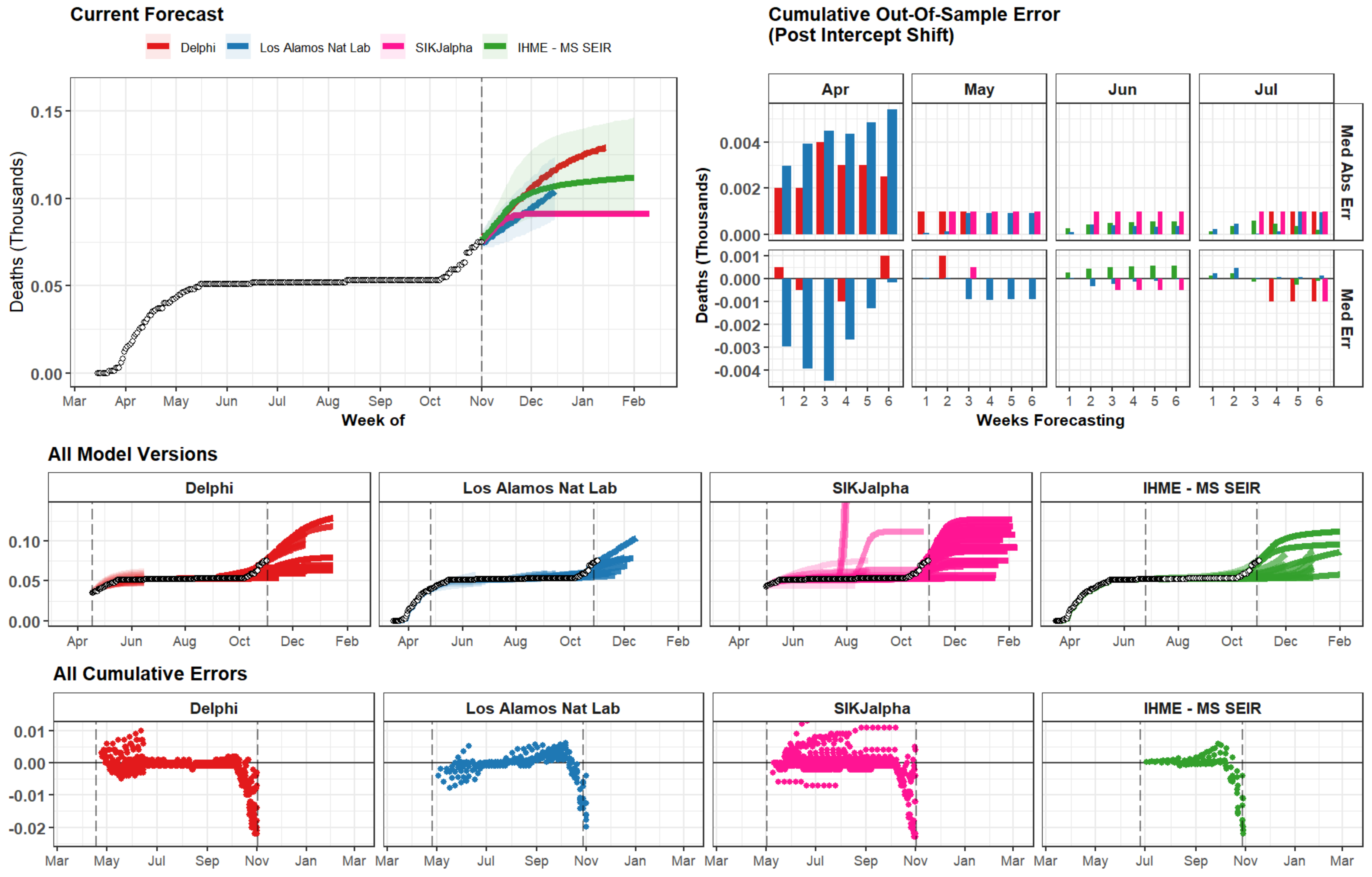
Current Forecast
Delphi
Los Alamos Nat Lab
SIKJalpha
IHME - MS SEIR
Deaths (Thousands)
0.15
0.10
0.05
0.00
Mar
Apr
May
Jun
Jul
Aug
Sep
Oct
Nov
Dec
Jan
Feb
Week of
Cumulative Out-Of-Sample Error
(Post Intercept Shift)
Apr
May
Jun
Jul
Med Abs Err
Med Err
0.004
0.002
0.000
0.001
-0.001
-0.002
-0.003
-0.004
1
2
3
4
5
6
Weeks Forecasting
All Model Versions
Delphi
Los Alamos Nat Lab
SIKJalpha
IHME - MS SEIR
All Cumulative Errors
0.01
0.00
-0.01
-0.02

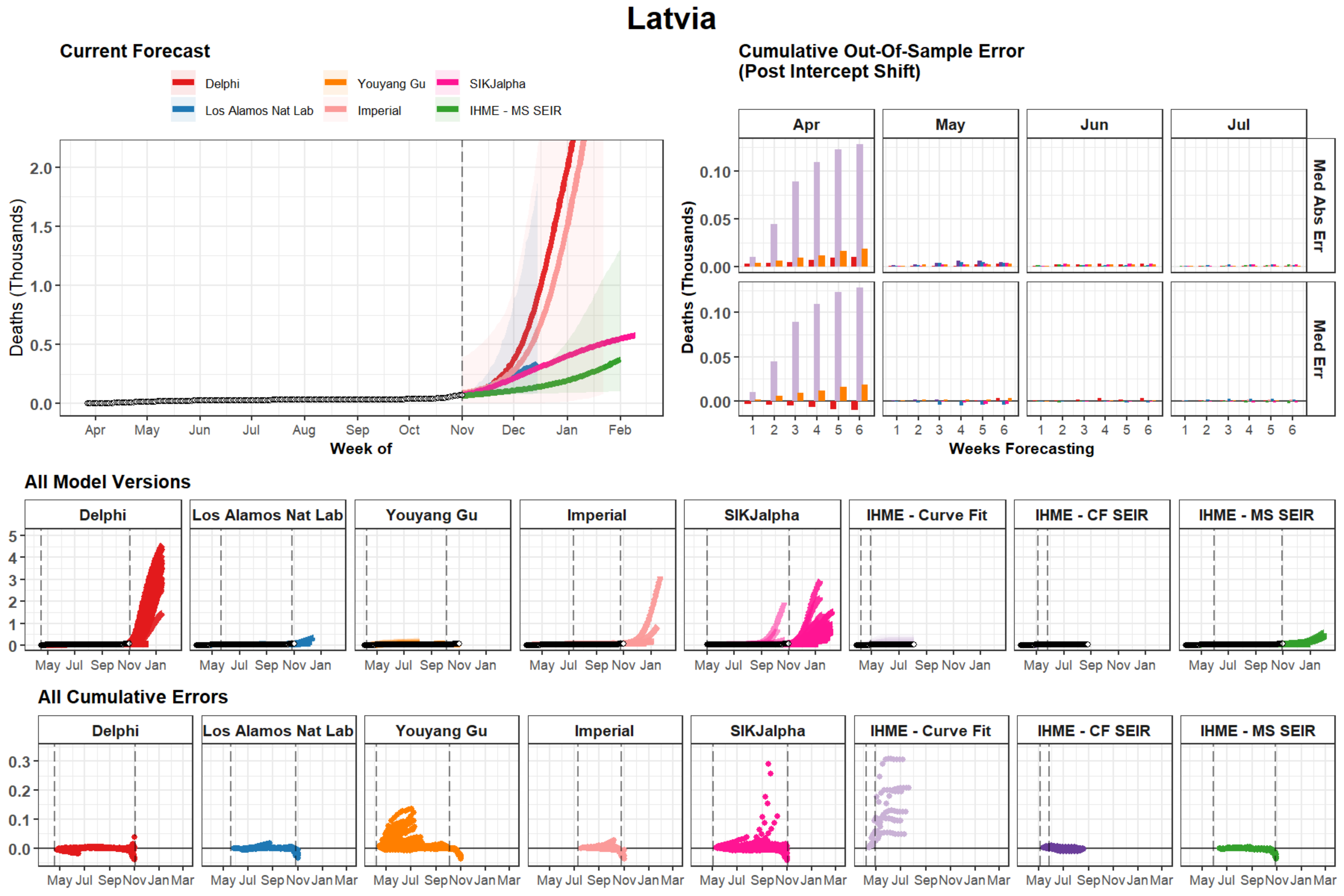

Latvia
Current Forecast
Delphi
Los Alamos Nat Lab
Youyang Gu
Imperial
SIKJalpha
IHME - MS SEIR
Deaths (Thousands)
Week of
Apr
May
Jun
Jul
Aug
Sep
Oct
Nov
Dec
Jan
Feb
Cumulative Out-Of-Sample Error
(Post Intercept Shift)
Med Abs Err
Med Err
Weeks Forecasting
All Model Versions
IHME - Curve Fit
IHME - CF SEIR
All Cumulative Errors

### Sierra Leone

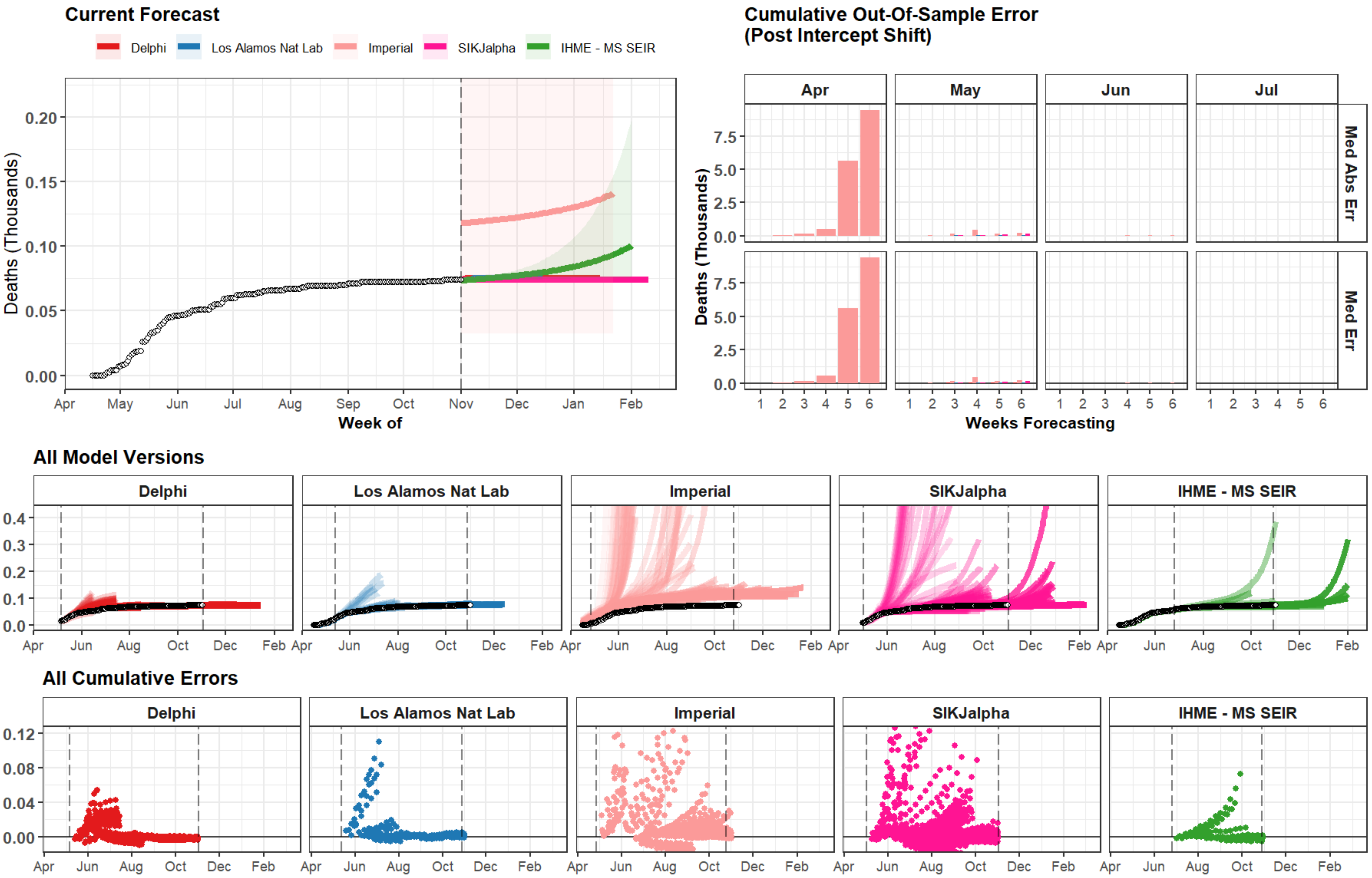

### Estonia

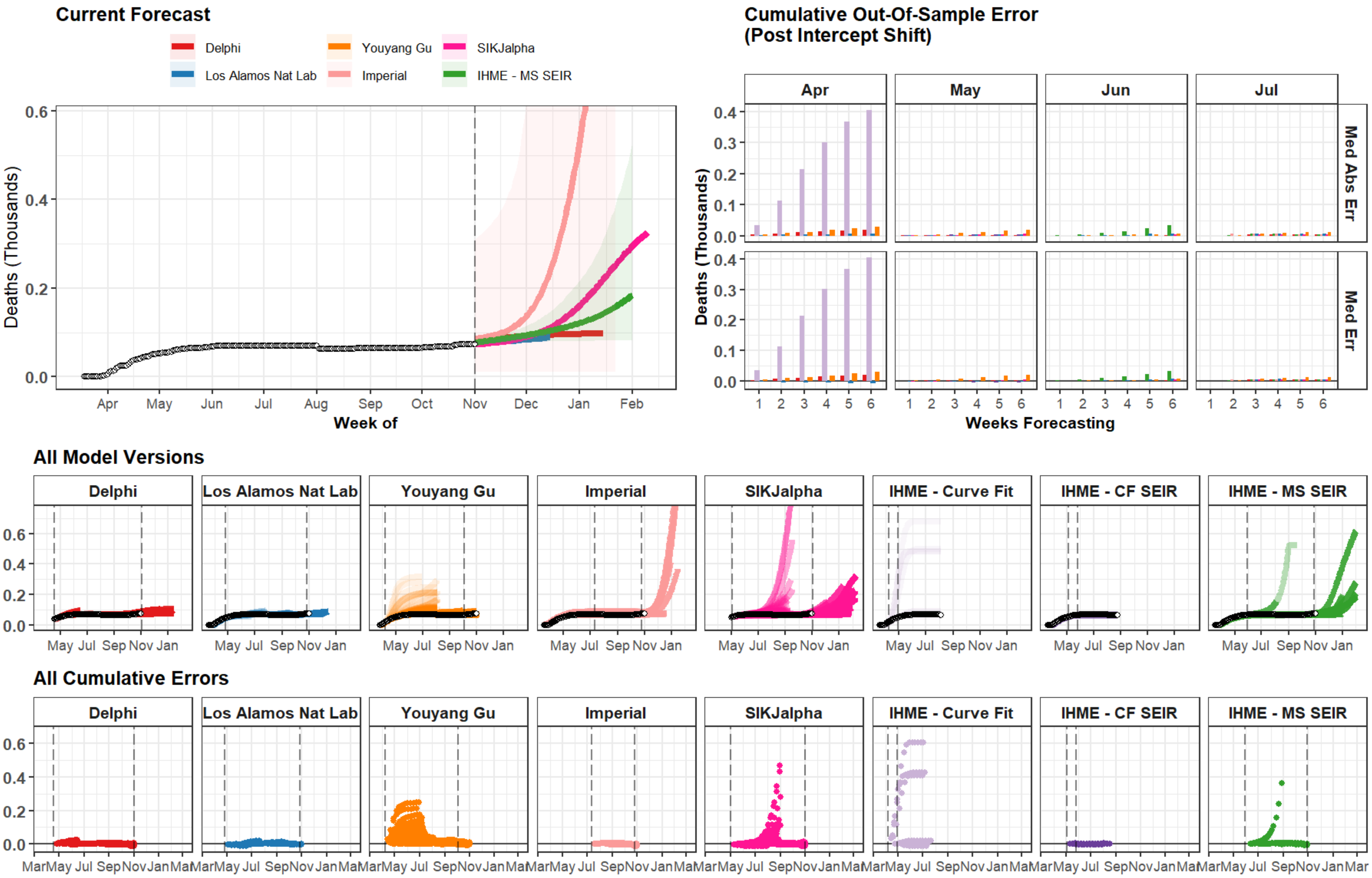
Current Forecast
Delphi
Youyang Gu
SIKJalpha
Los Alamos Nat Lab
Imperial
IHME - MS SEIR
Deaths (Thousands)
Week of
Apr May Jun Jul Aug Sep Oct Nov Dec Jan Feb
Cumulative Out-Of-Sample Error
(Post Intercept Shift)
Apr
May
Jun
Jul
Med Abs Err
Med Err
Weeks Forecasting
All Model Versions
Delphi
Los Alamos Nat Lab
Youyang Gu
Imperial
SIKJalpha
IHME - Curve Fit
IHME - CF SEIR
IHME - MS SEIR
All Cumulative Errors

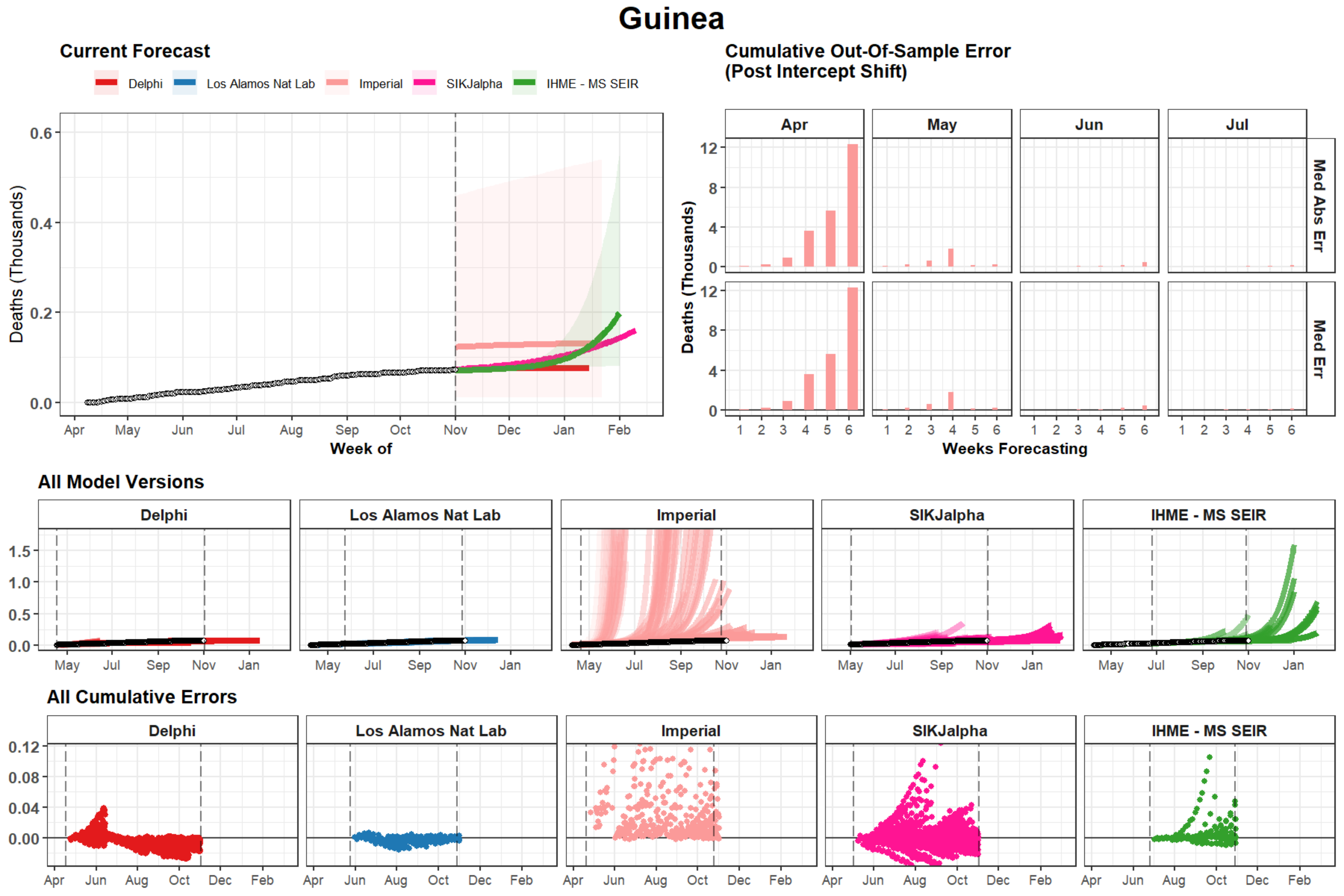

Guinea
Current Forecast
Delphi
Los Alamos Nat Lab
Imperial
SIKJalpha
IHME - MS SEIR
Deaths (Thousands)
Week of
Cumulative Out-Of-Sample Error
(Post Intercept Shift)
Apr
May
Jun
Jul
Med Abs Err
Med Err
Weeks Forecasting
All Model Versions
All Cumulative Errors

### Niger

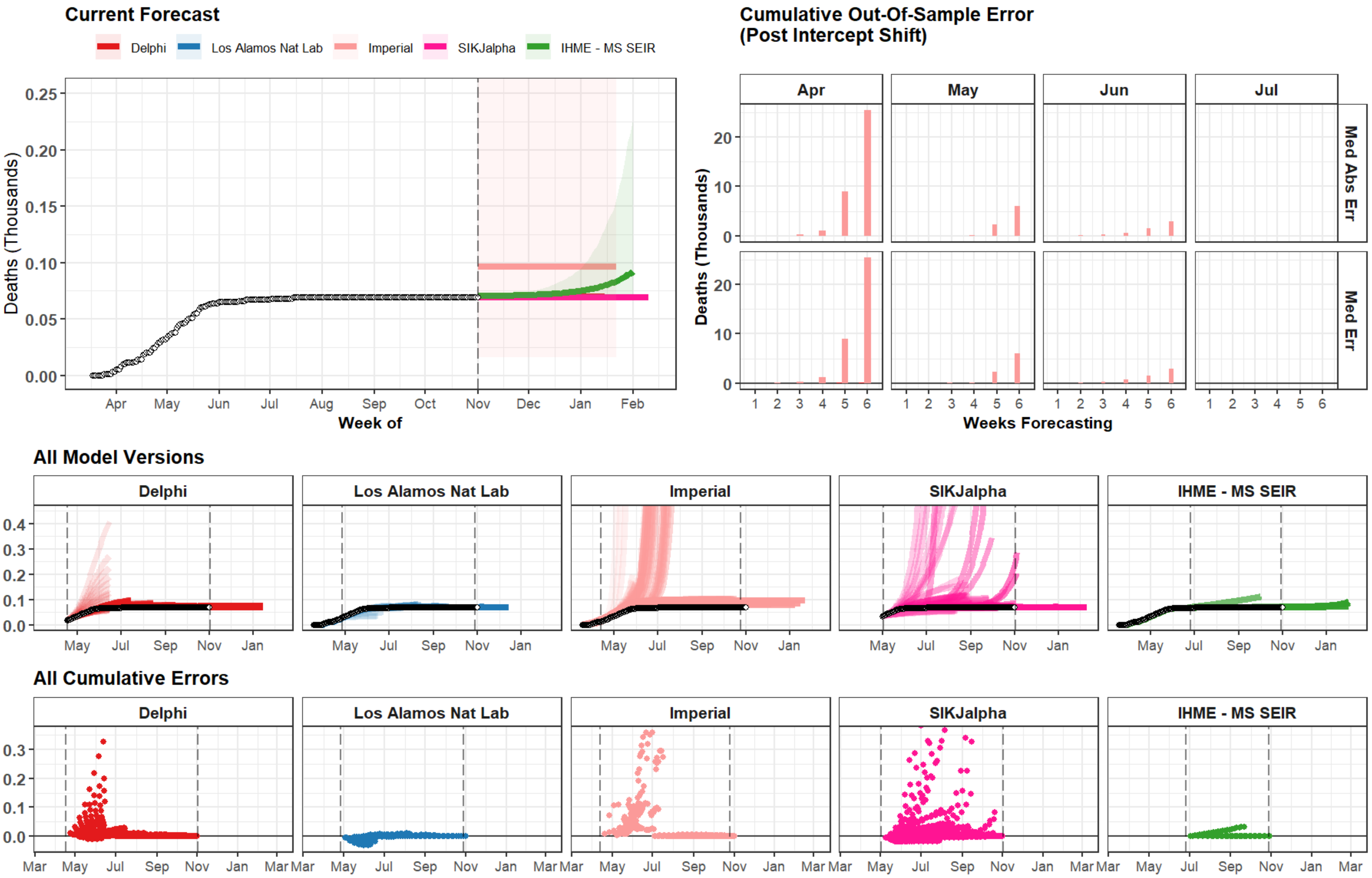
Current Forecast
Delphi
Los Alamos Nat Lab
Imperial
SIKJalpha
IHME - MS SEIR
Deaths (Thousands)
Week of
Cumulative Out-Of-Sample Error
(Post Intercept Shift)
Apr
May
Jun
Jul
Med Abs Err
Med Err
Weeks Forecasting
All Model Versions
All Cumulative Errors

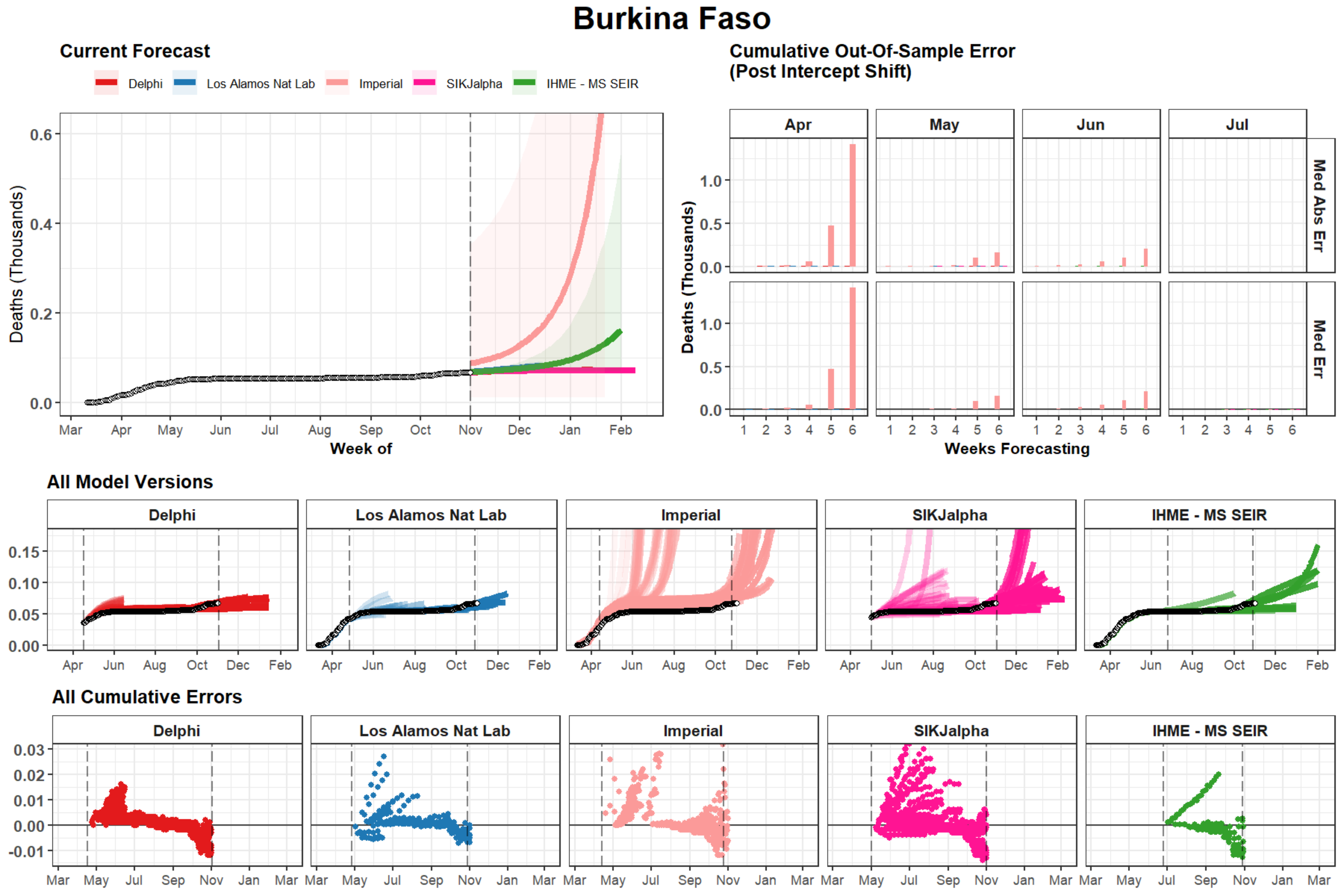
Burkina Faso
Current Forecast
Delphi
Los Alamos Nat Lab
Imperial
SIKJalpha
IHME - MS SEIR
Deaths (Thousands)
Week of
Cumulative Out-Of-Sample Error
(Post Intercept Shift)
Apr
May
Jun
Jul
Med Abs Err
Med Err
Weeks Forecasting
All Model Versions
All Cumulative Errors

### Malta

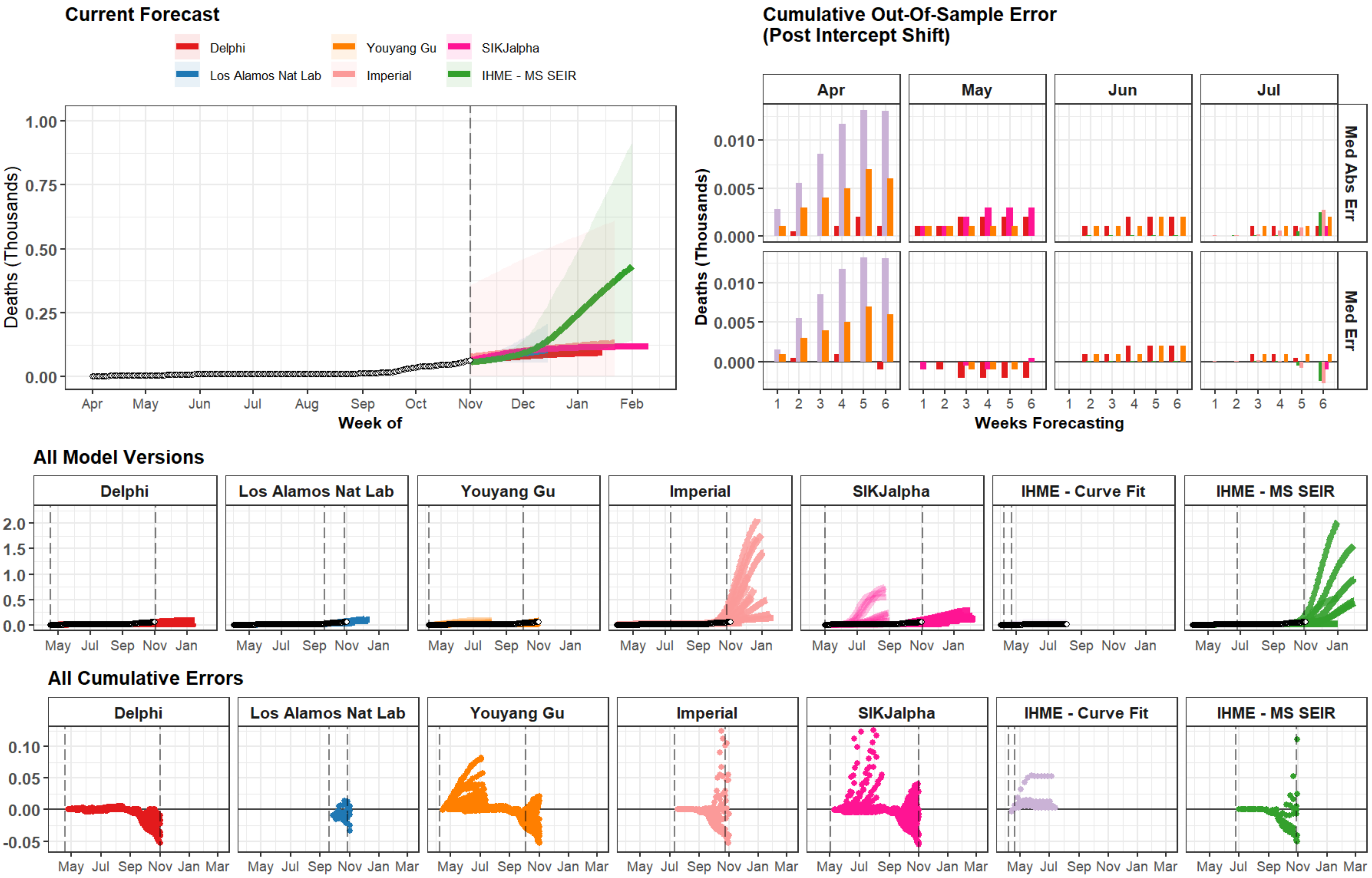

Current Forecast
Delphi
Youyang Gu
SIKJalpha
Los Alamos Nat Lab
Imperial
IHME - MS SEIR
Deaths (Thousands)
1.00
0.75
0.50
0.25
0.00
Apr
May
Jun
Jul
Aug
Sep
Oct
Nov
Dec
Jan
Feb
Week of
Cumulative Out-Of-Sample Error
(Post Intercept Shift)
Apr
May
Jun
Jul
Med Abs Err
Med Err
0.010
0.005
0.000
Weeks Forecasting
All Model Versions
Delphi
Los Alamos Nat Lab
Youyang Gu
Imperial
SIKJalpha
IHME - Curve Fit
IHME - MS SEIR
2.0
1.5
1.0
0.5
0.0
All Cumulative Errors
0.10
0.05
0.00
-0.05
May
Jul
Sep
Nov
Jan
Mar

### Central African Republic

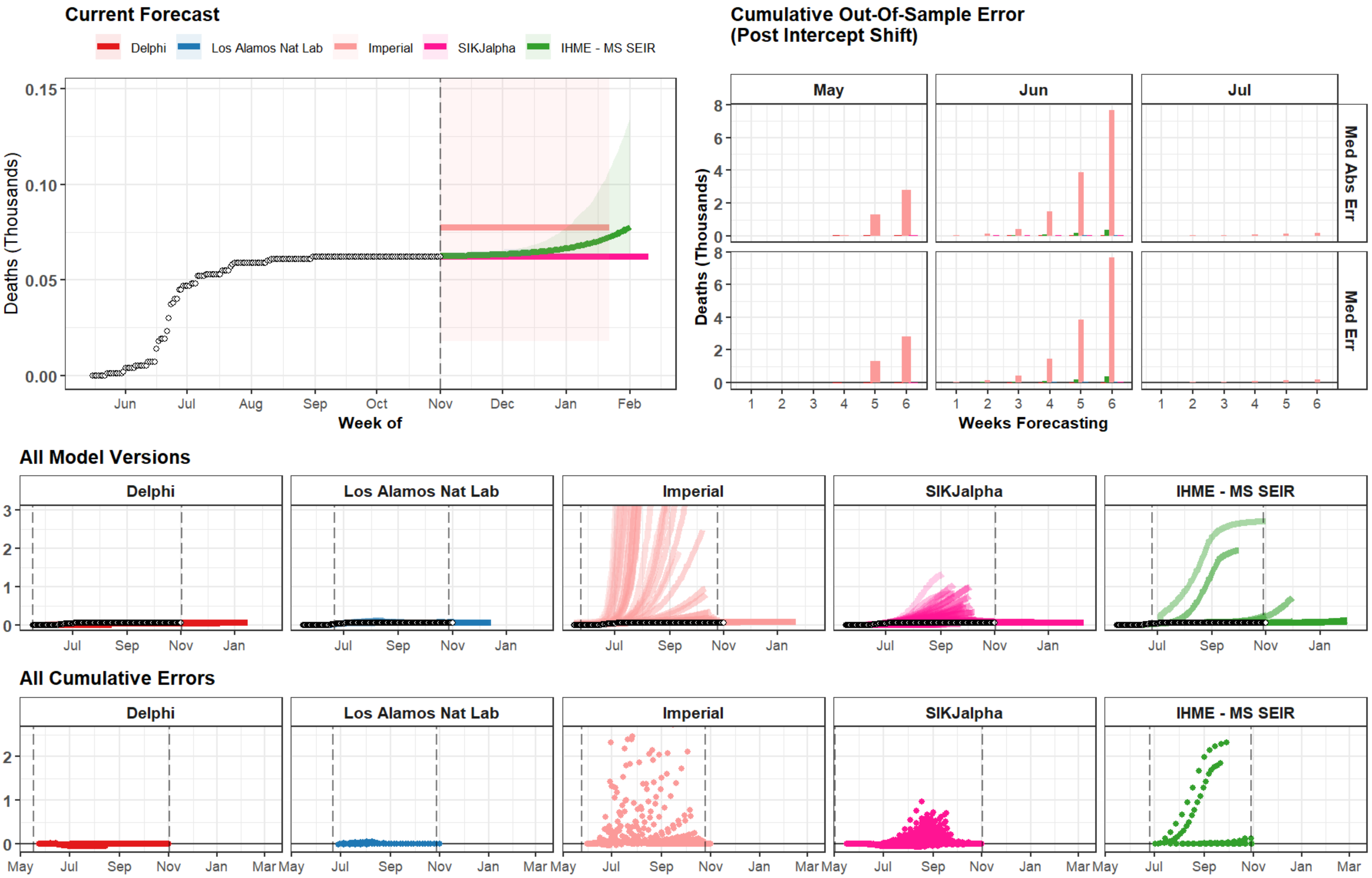

### Djibouti

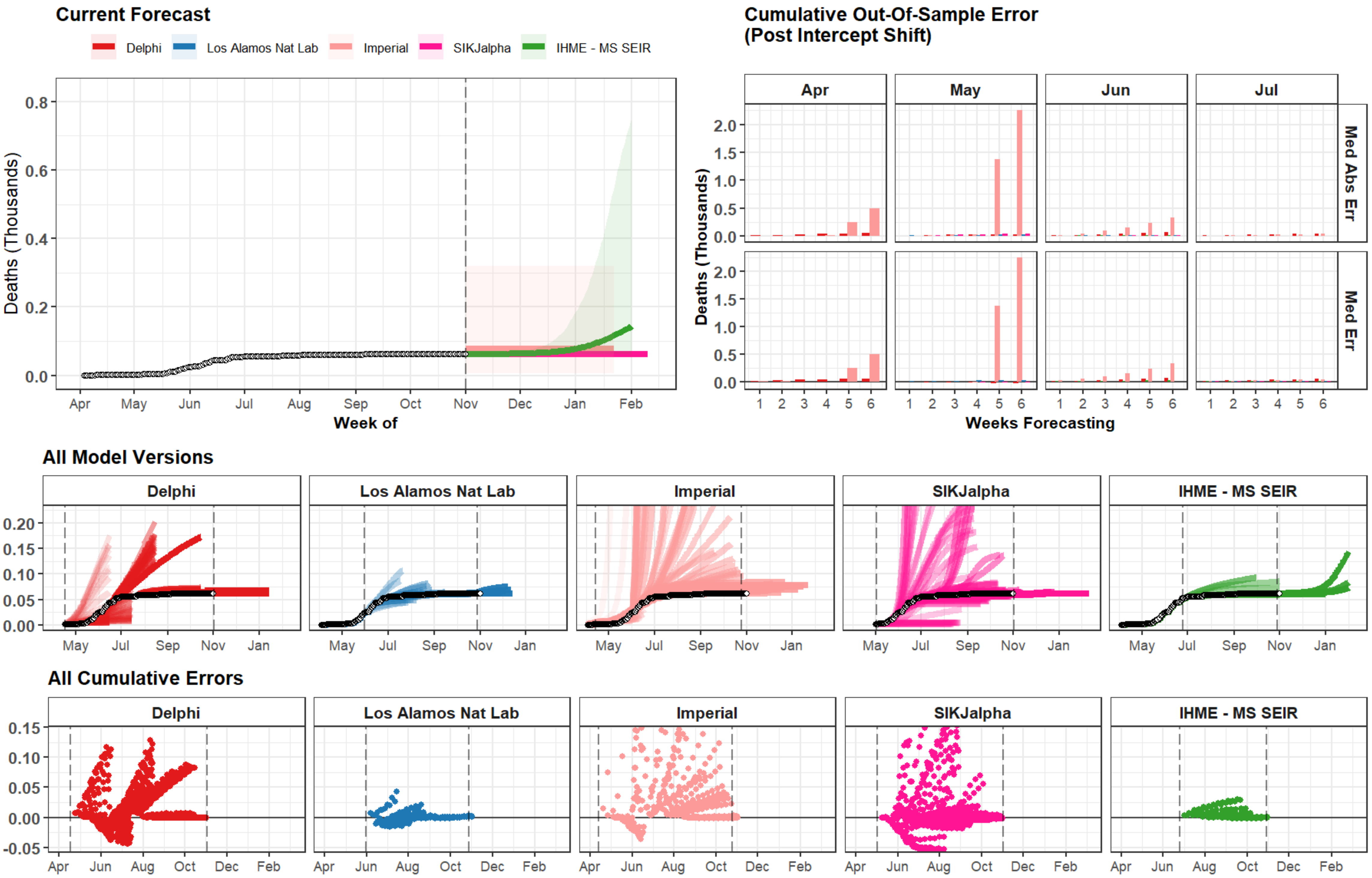

Current Forecast
Delphi
Los Alamos Nat Lab
Imperial
SIKJalpha
IHME - MS SEIR
Deaths (Thousands)
0.0
0.2
0.4
0.6
0.8
Apr
May
Jun
Jul
Aug
Sep
Oct
Nov
Dec
Jan
Feb
Week of
Cumulative Out-Of-Sample Error
(Post Intercept Shift)
Apr
May
Jun
Jul
Med Abs Err
Med Err
0.0
0.5
1.0
1.5
2.0
1
2
3
4
5
6
Weeks Forecasting
All Model Versions
Delphi
Los Alamos Nat Lab
Imperial
SIKJalpha
IHME - MS SEIR
0.00
0.05
0.10
0.15
0.20
May
Jul
Sep
Nov
Jan
All Cumulative Errors
Delphi
Los Alamos Nat Lab
Imperial
SIKJalpha
IHME - MS SEIR
-0.05
0.00
0.05
0.10
0.15
Apr
Jun
Aug
Oct
Dec
Feb

### Belize

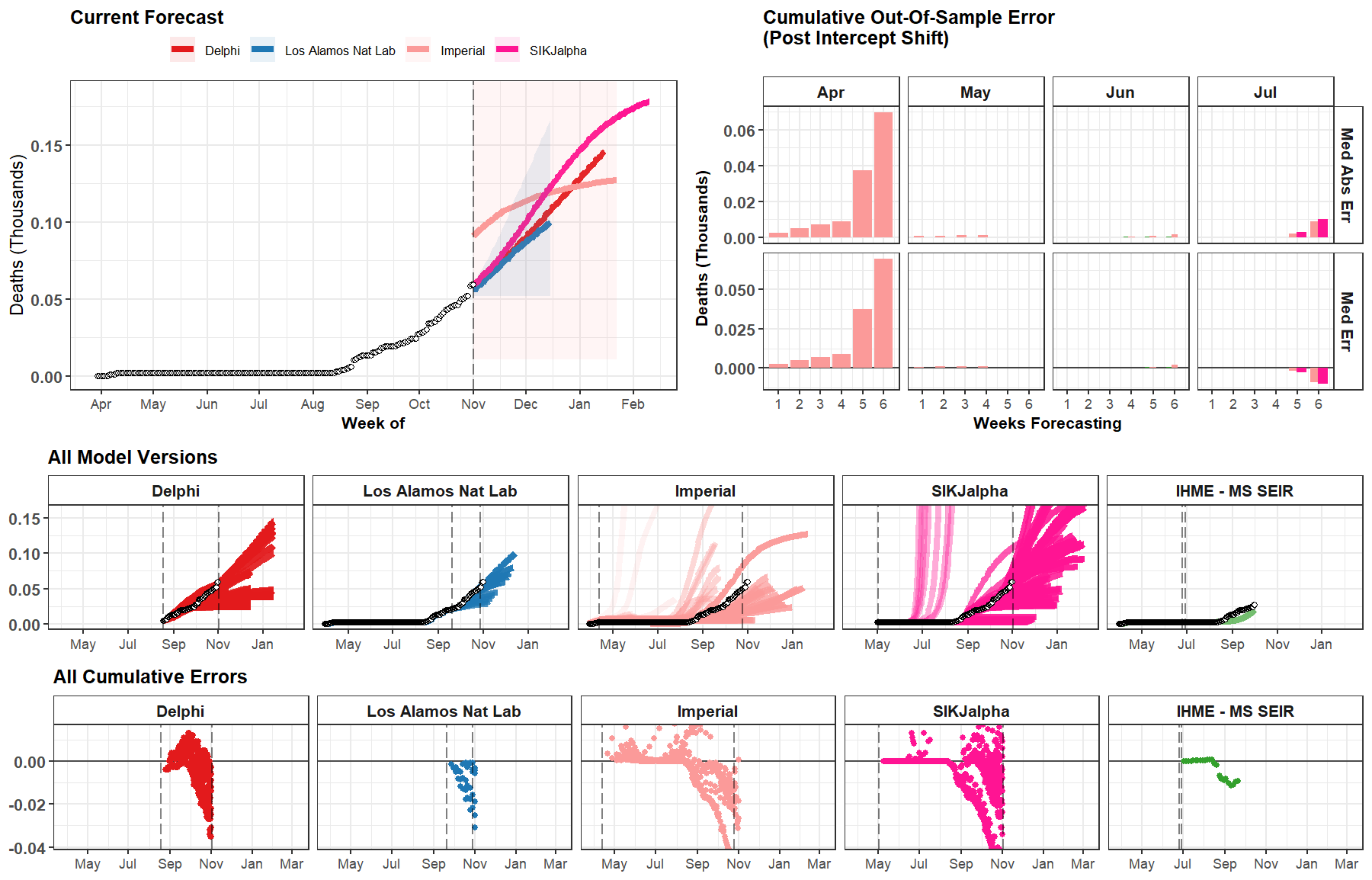
Current Forecast
Delphi
Los Alamos Nat Lab
Imperial
SIKJalpha
Deaths (Thousands)
Week of
Cumulative Out-Of-Sample Error
(Post Intercept Shift)
Apr
May
Jun
Jul
Med Abs Err
Med Err
Weeks Forecasting
All Model Versions
Delphi
Los Alamos Nat Lab
Imperial
SIKJalpha
IHME - MS SEIR
All Cumulative Errors
Delphi
Los Alamos Nat Lab
Imperial
SIKJalpha
IHME - MS SEIR

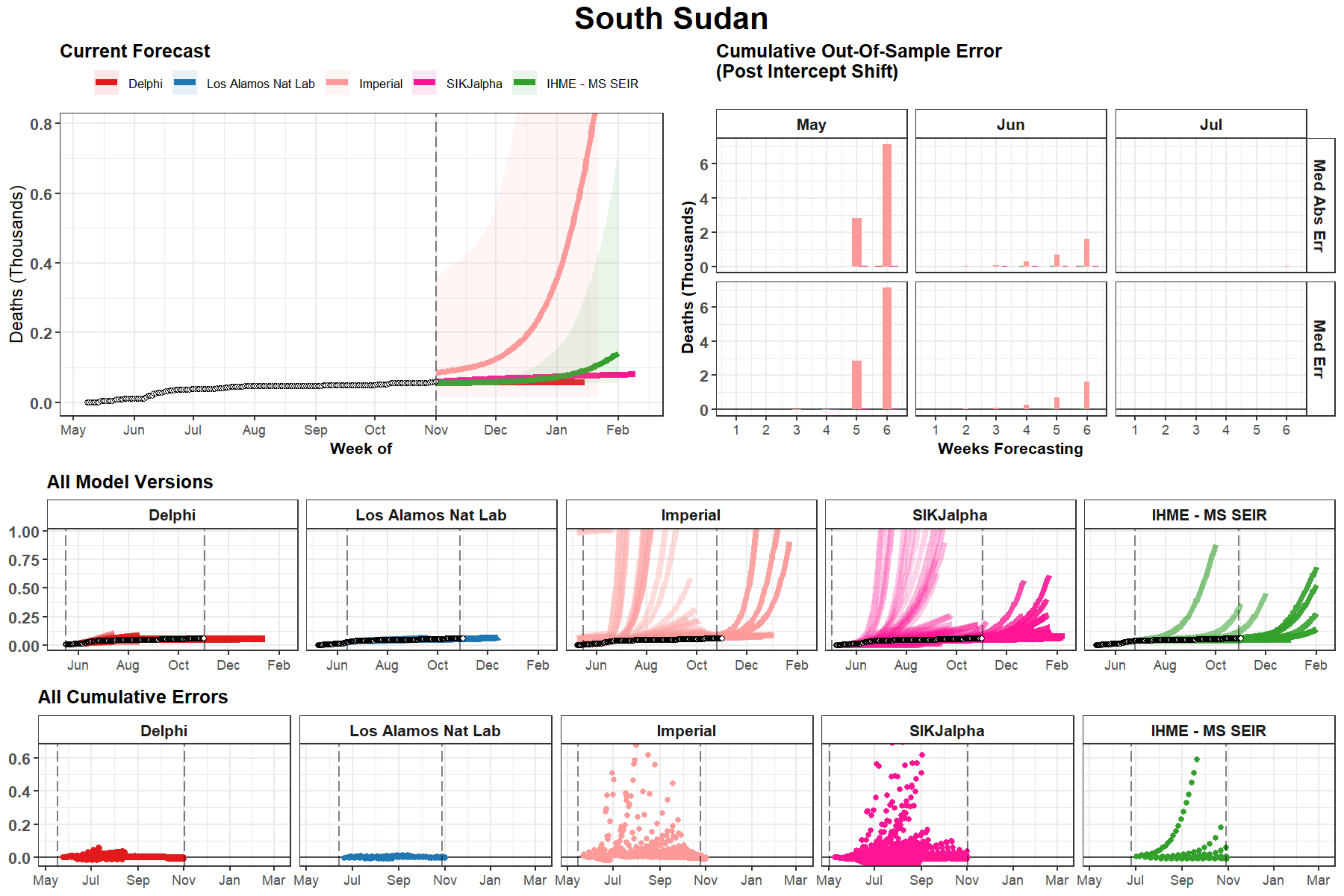
South Sudan
Current Forecast
Delphi
Los Alamos Nat Lab
Imperial
SIKJalpha
IHME - MS SEIR
Deaths (Thousands)
Week of
Cumulative Out-Of-Sample Error
(Post Intercept Shift)
May
Jun
Jul
Med Abs Err
Med Err
Weeks Forecasting
All Model Versions
All Cumulative Errors

### Thailand

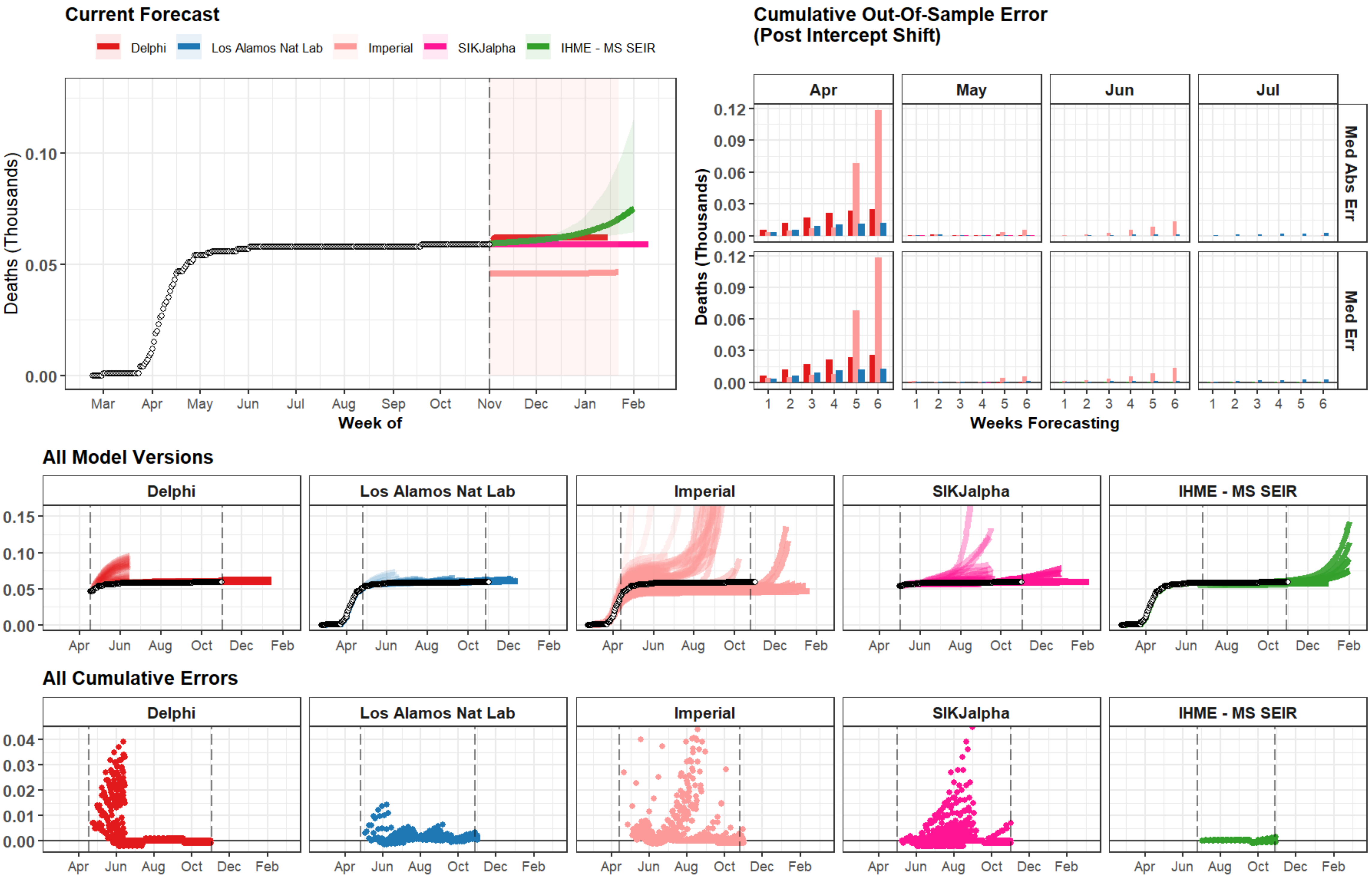

Current Forecast
Delphi
Los Alamos Nat Lab
Imperial
SIKJalpha
IHME - MS SEIR
Deaths (Thousands)
0.10
0.05
0.00
Mar
Apr
May
Jun
Jul
Aug
Sep
Oct
Nov
Dec
Jan
Feb
Week of
Cumulative Out-Of-Sample Error
(Post Intercept Shift)
Apr
May
Jun
Jul
0.12
0.09
0.06
0.03
0.00
Med Abs Err
Med Err
Deaths (Thousands)
1
2
3
4
5
6
Weeks Forecasting
All Model Versions
Delphi
Los Alamos Nat Lab
Imperial
SIKJalpha
IHME - MS SEIR
0.15
0.10
0.05
0.00
Apr
Jun
Aug
Oct
Dec
Feb
All Cumulative Errors
Delphi
Los Alamos Nat Lab
Imperial
SIKJalpha
IHME - MS SEIR
0.04
0.03
0.02
0.01
0.00
Apr
Jun
Aug
Oct
Dec
Feb

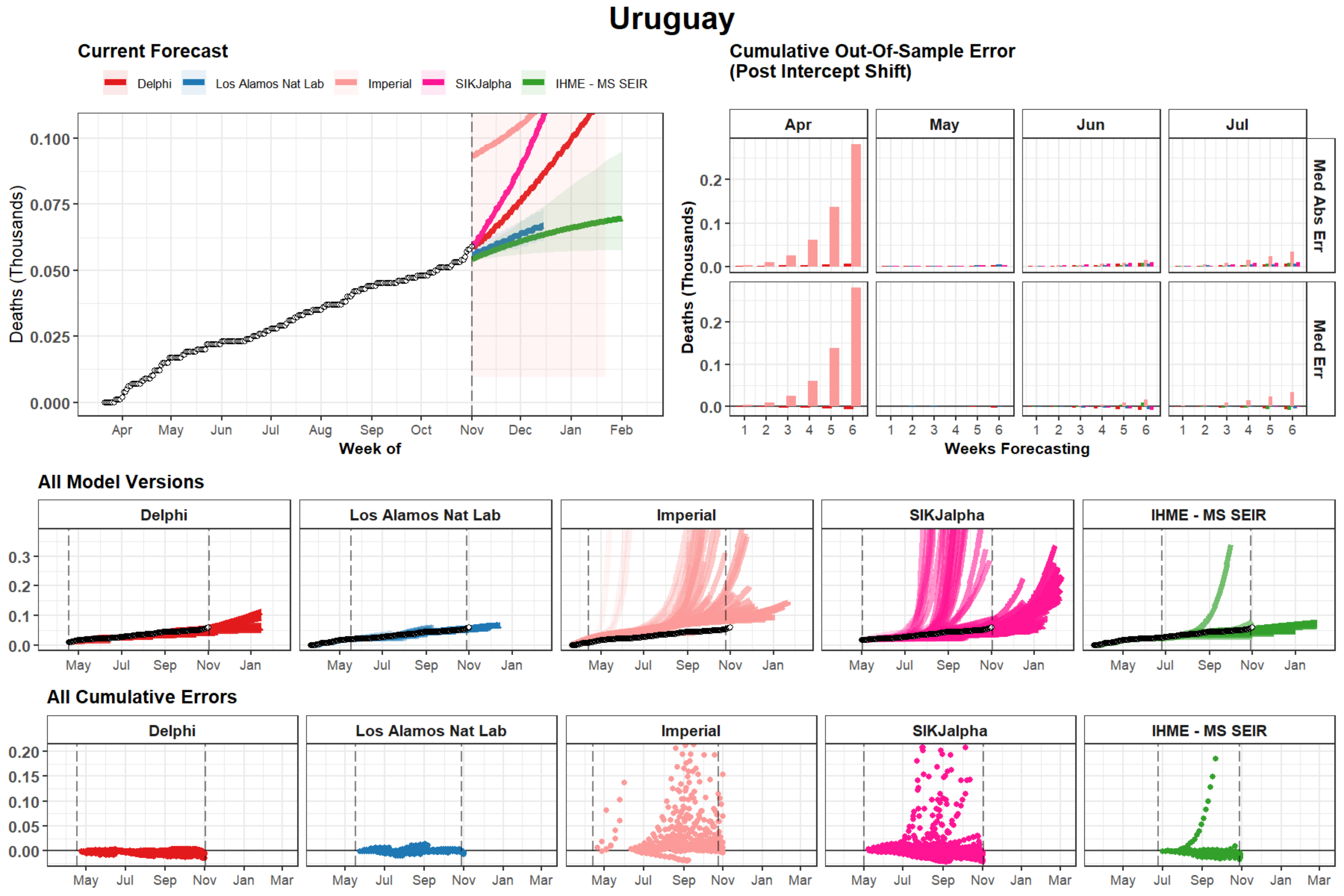

Uruguay
Current Forecast
Delphi
Los Alamos Nat Lab
Imperial
SIKJalpha
IHME - MS SEIR
Deaths (Thousands)
Week of
Apr
May
Jun
Jul
Aug
Sep
Oct
Nov
Dec
Jan
Feb
Cumulative Out-Of-Sample Error
(Post Intercept Shift)
Med Abs Err
Med Err
Weeks Forecasting
All Model Versions
All Cumulative Errors

### Vermont

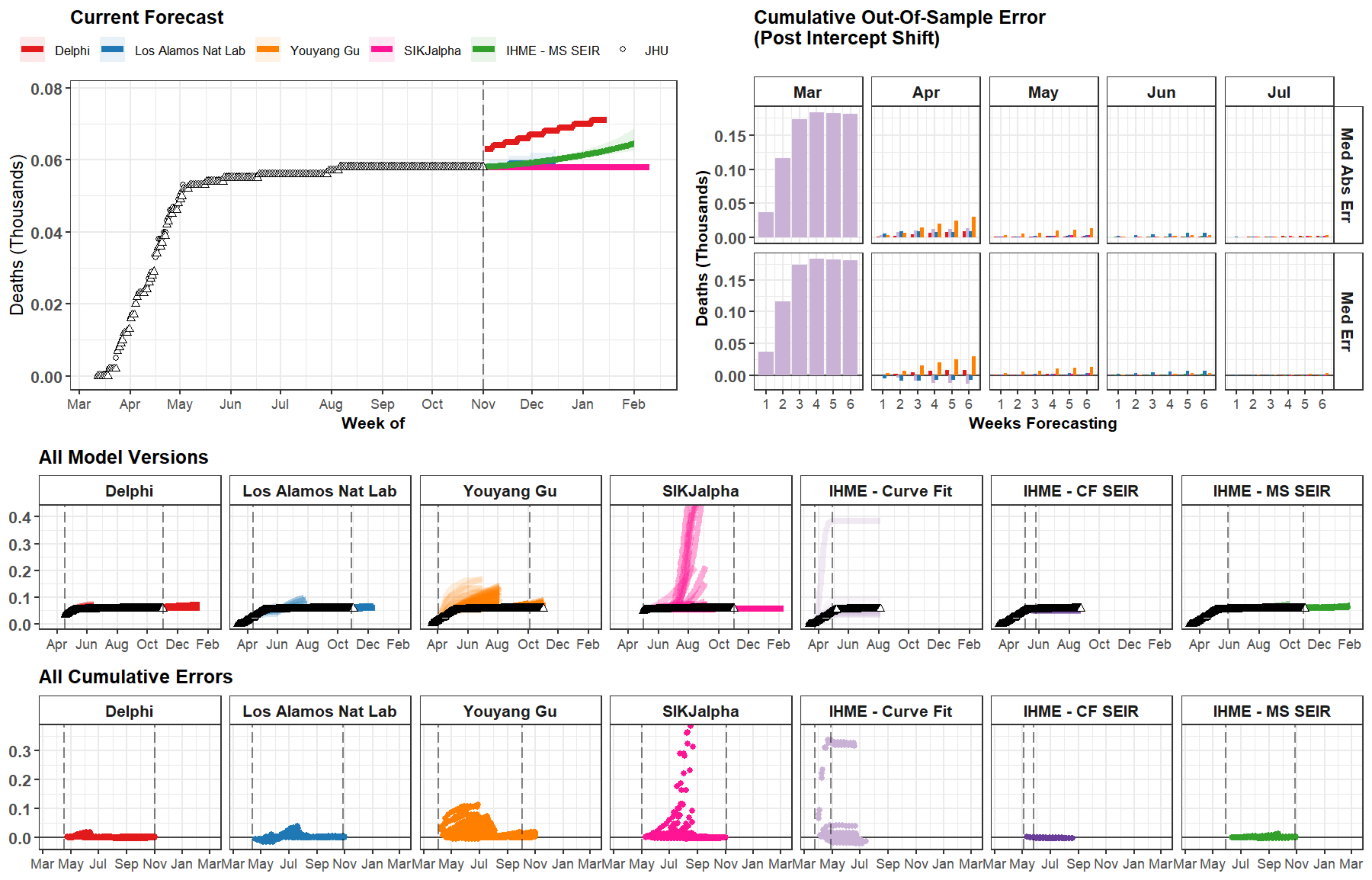

Current Forecast
Delphi
Los Alamos Nat Lab
Youyang Gu
SIKJalpha
IHME - MS SEIR
JHU
Deaths (Thousands)
0.08
0.06
0.04
0.02
0.00
Mar
Apr
May
Jun
Jul
Aug
Sep
Oct
Nov
Dec
Jan
Feb
Week of
Cumulative Out-Of-Sample Error
(Post Intercept Shift)
Mar
Apr
May
Jun
Jul
Med Abs Err
Med Err
0.15
0.10
0.05
0.00
1 2 3 4 5 6
Weeks Forecasting
All Model Versions
Delphi
Los Alamos Nat Lab
Youyang Gu
SIKJalpha
IHME - Curve Fit
IHME - CF SEIR
IHME - MS SEIR
0.4
0.3
0.2
0.1
0.0
Apr Jun Aug Oct Dec Feb
All Cumulative Errors
0.3
0.2
0.1
0.0
Mar May Jul Sep Nov Jan Mar

### Togo

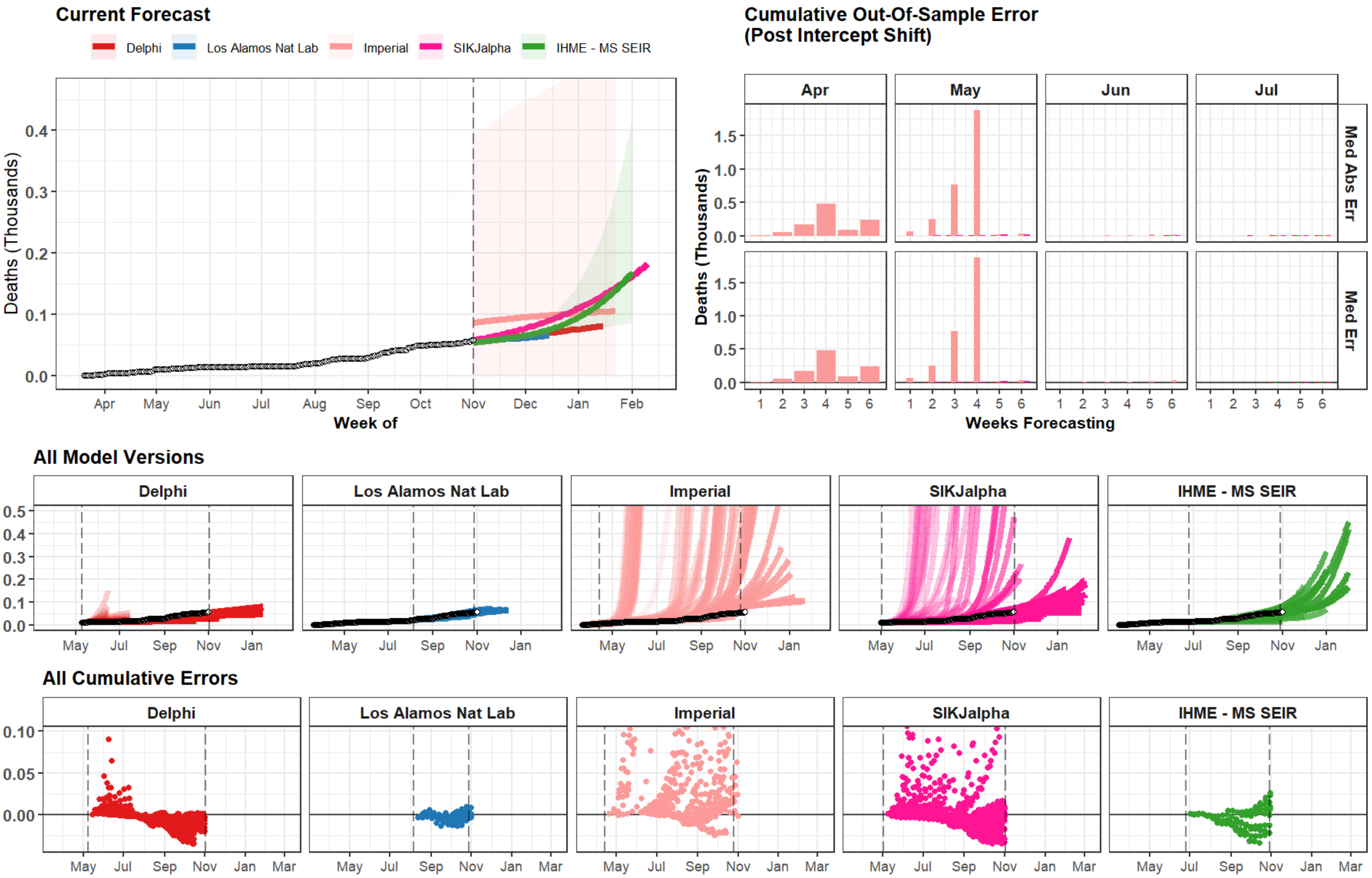
Current Forecast
Delphi
Los Alamos Nat Lab
Imperial
SIKJalpha
IHME - MS SEIR
Deaths (Thousands)
Week of
Apr
May
Jun
Jul
Aug
Sep
Oct
Nov
Dec
Jan
Feb
Cumulative Out-Of-Sample Error
(Post Intercept Shift)
Med Abs Err
Med Err
Weeks Forecasting
All Model Versions
All Cumulative Errors
Mar

### Gabon

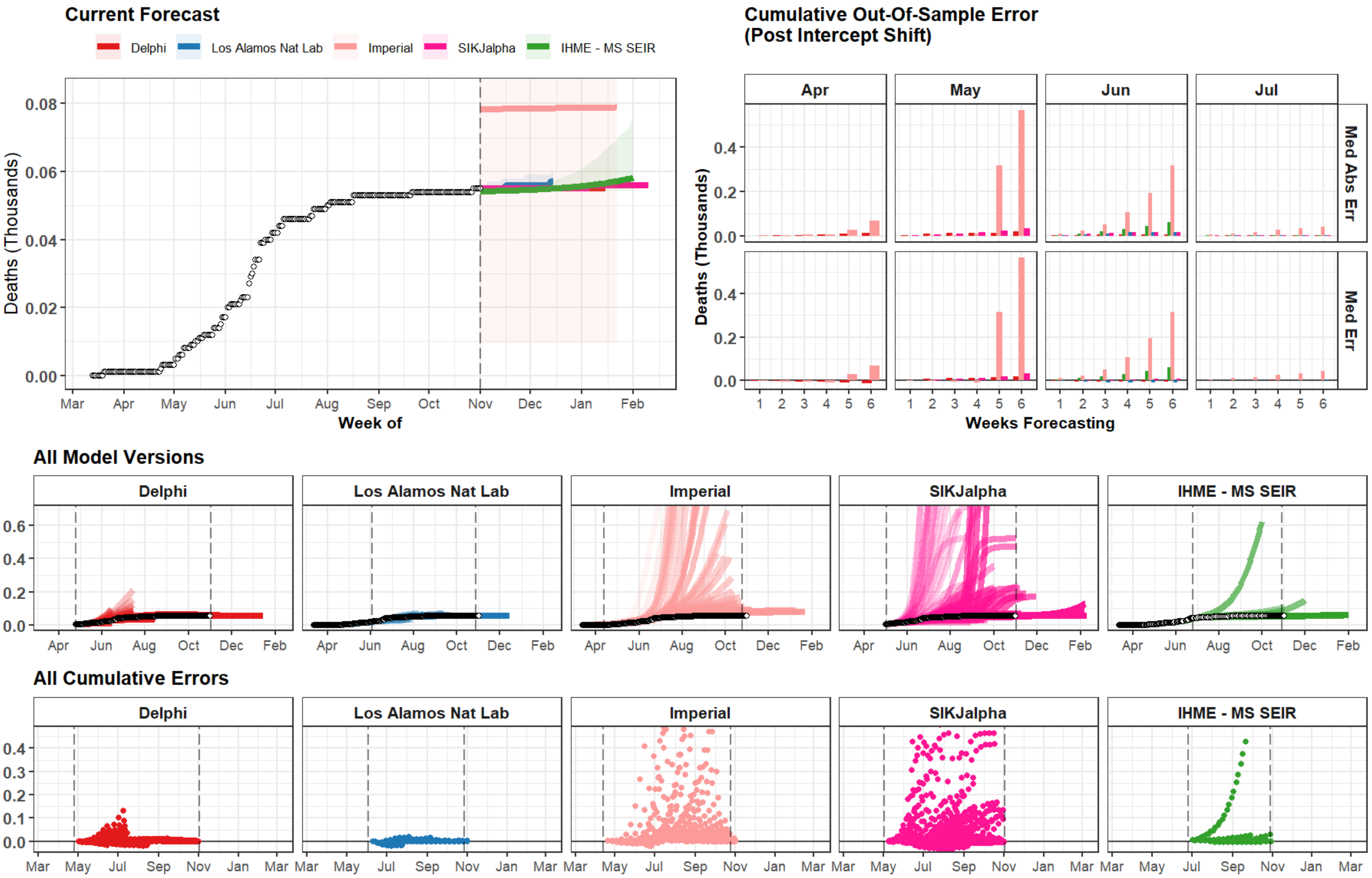

Current Forecast
Delphi
Los Alamos Nat Lab
Imperial
SIKJalpha
IHME - MS SEIR
Deaths (Thousands)
Week of
Cumulative Out-Of-Sample Error
(Post Intercept Shift)
Apr
May
Jun
Jul
Med Abs Err
Med Err
Weeks Forecasting
All Model Versions
All Cumulative Errors

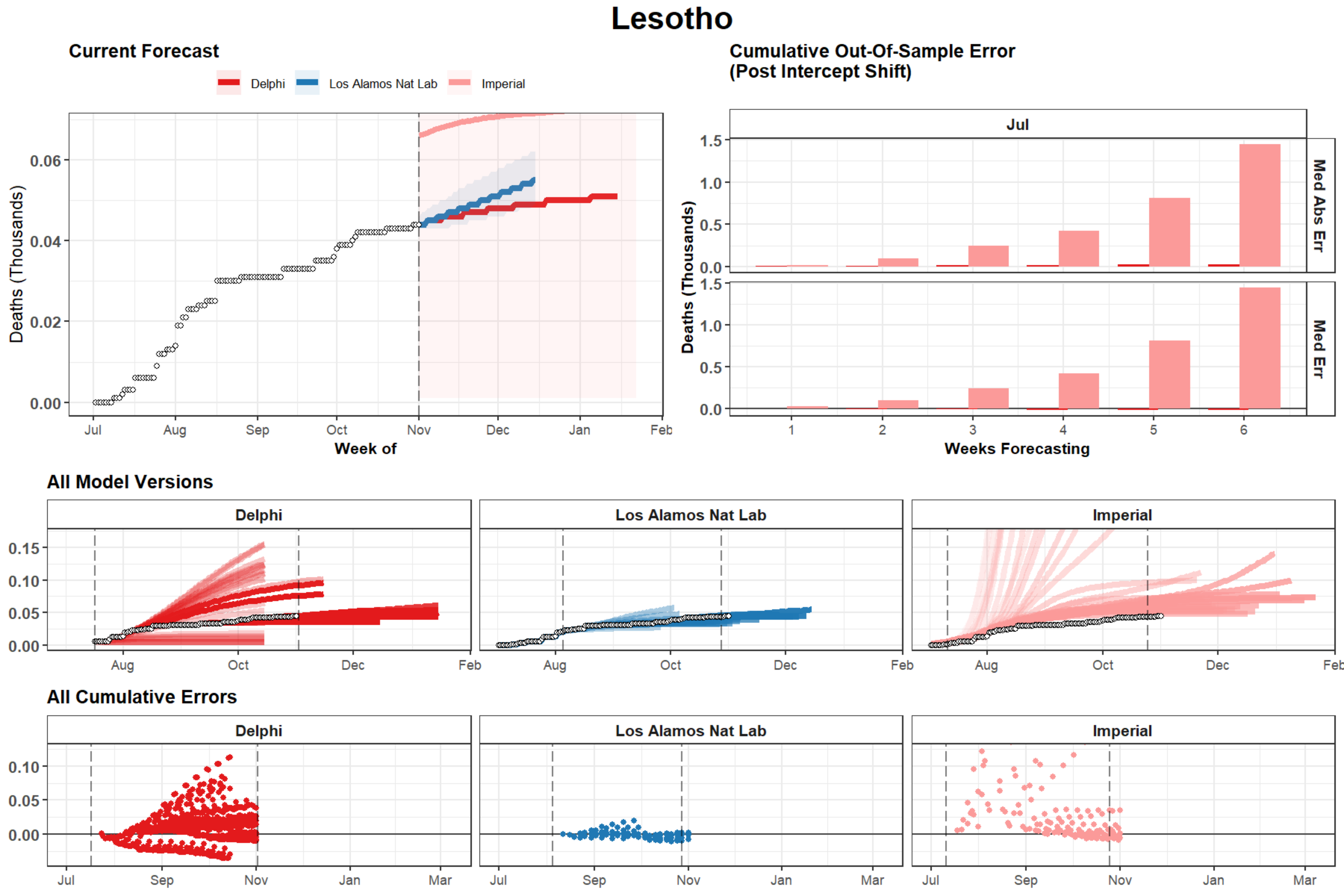
Lesotho
Current Forecast
Delphi
Los Alamos Nat Lab
Imperial
Deaths (Thousands)
0.06
0.04
0.02
0.00
Jul
Aug
Sep
Oct
Nov
Dec
Jan
Feb
Week of
Cumulative Out-Of-Sample Error
(Post Intercept Shift)
Jul
Med Abs Err
Med Err
Deaths (Thousands)
1.5
1.0
0.5
0.0
1
2
3
4
5
6
Weeks Forecasting
All Model Versions
Delphi
Los Alamos Nat Lab
Imperial
0.15
0.10
0.05
0.00
Aug
Oct
Dec
Feb
All Cumulative Errors
Delphi
Los Alamos Nat Lab
Imperial
0.10
0.05
0.00
Jul
Sep
Nov
Jan
Mar

### Benin

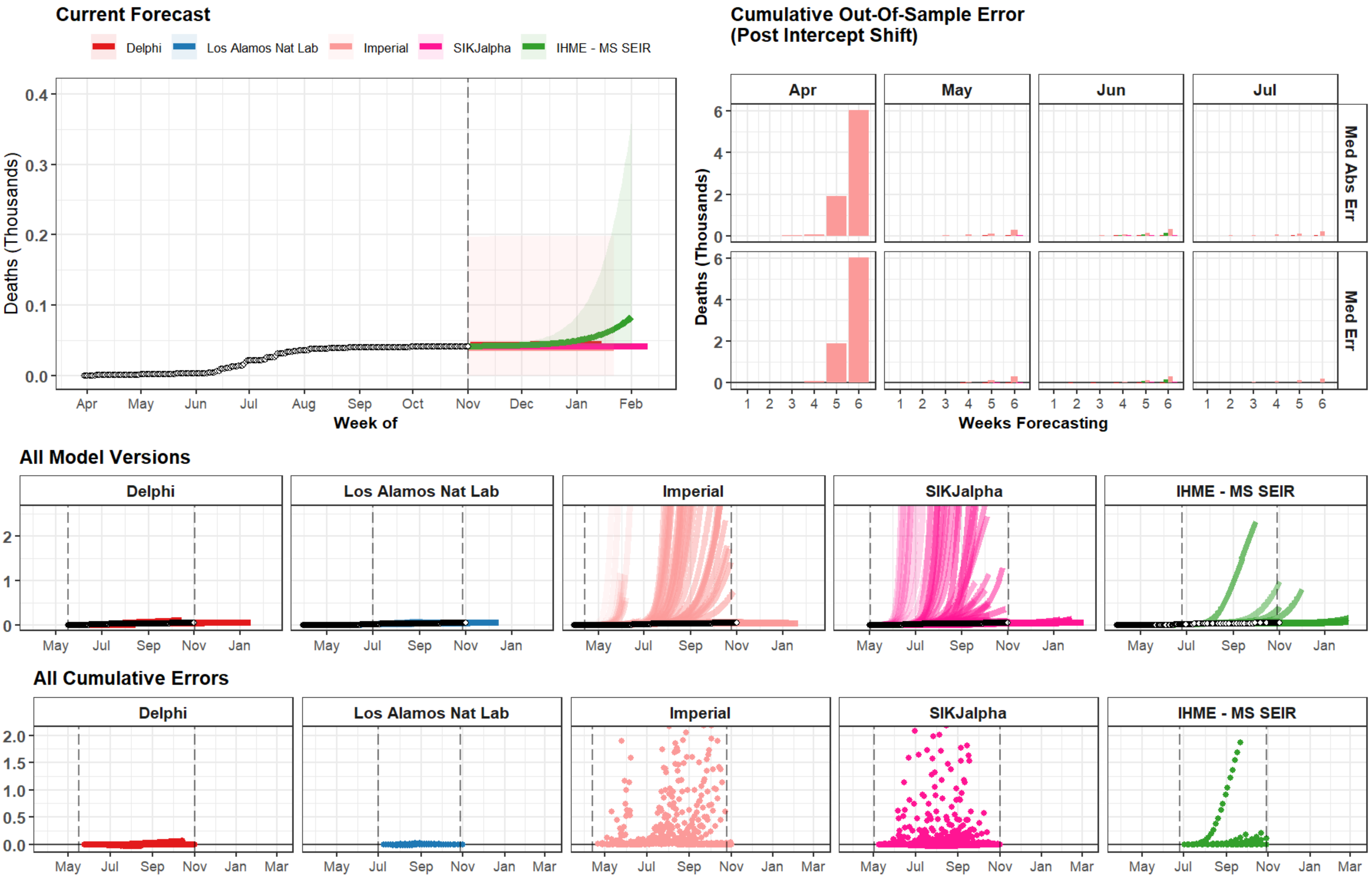
Current Forecast
Delphi
Los Alamos Nat Lab
Imperial
SIKJalpha
IHME - MS SEIR
Deaths (Thousands)
Week of
Cumulative Out-Of-Sample Error
(Post Intercept Shift)
Apr
May
Jun
Jul
Med Abs Err
Med Err
Weeks Forecasting
All Model Versions
All Cumulative Errors

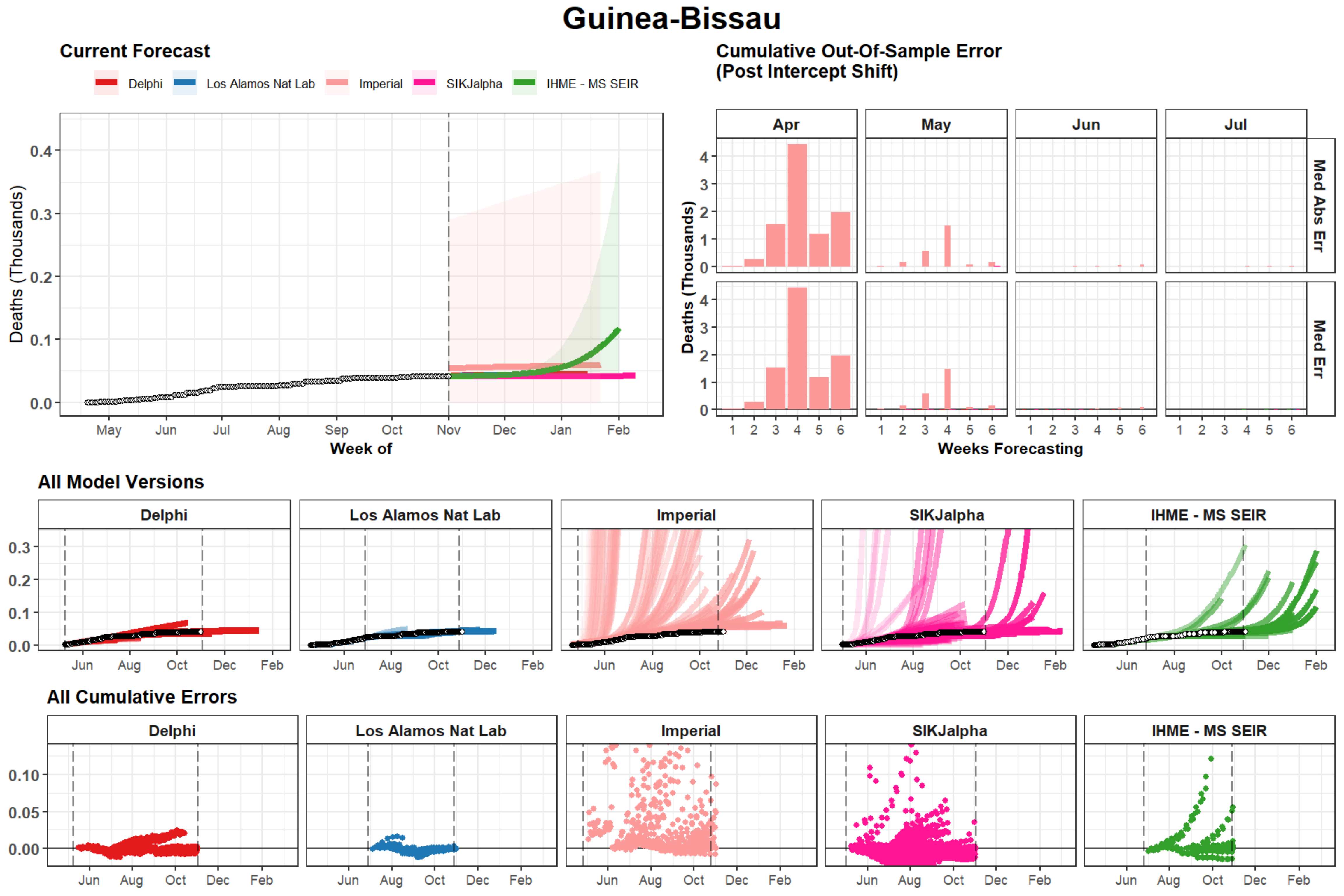
Guinea-Bissau
Current Forecast
Delphi
Los Alamos Nat Lab
Imperial
SIKJalpha
IHME - MS SEIR
Deaths (Thousands)
Week of
Cumulative Out-Of-Sample Error
(Post Intercept Shift)
Apr
May
Jun
Jul
Med Abs Err
Med Err
Weeks Forecasting
All Model Versions
All Cumulative Errors

### Maldives

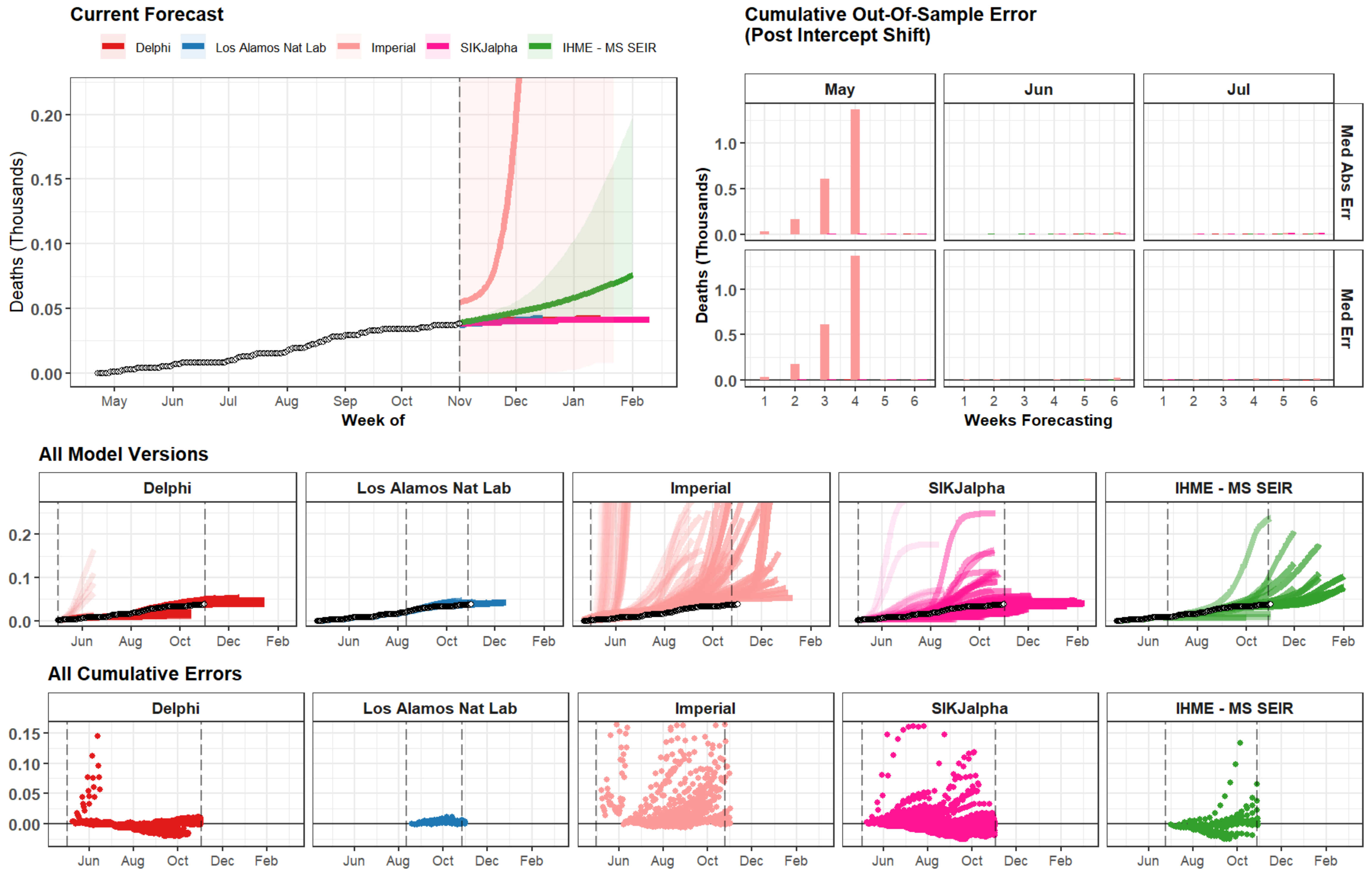
Current Forecast
Delphi
Los Alamos Nat Lab
Imperial
SIKJalpha
IHME - MS SEIR
Deaths (Thousands)
0.00
0.05
0.10
0.15
0.20
May
Jun
Jul
Aug
Sep
Oct
Nov
Dec
Jan
Feb
Week of
Cumulative Out-Of-Sample Error
(Post Intercept Shift)
May
Jun
Jul
Med Abs Err
Med Err
Deaths (Thousands)
0.0
0.5
1.0
1
2
3
4
5
6
Weeks Forecasting
All Model Versions
Delphi
Los Alamos Nat Lab
Imperial
SIKJalpha
IHME - MS SEIR
0.0
0.1
0.2
Jun
Aug
Oct
Dec
Feb
All Cumulative Errors
Delphi
Los Alamos Nat Lab
Imperial
SIKJalpha
IHME - MS SEIR
0.00
0.05
0.10
0.15
Jun
Aug
Oct
Dec
Feb

### Rwanda

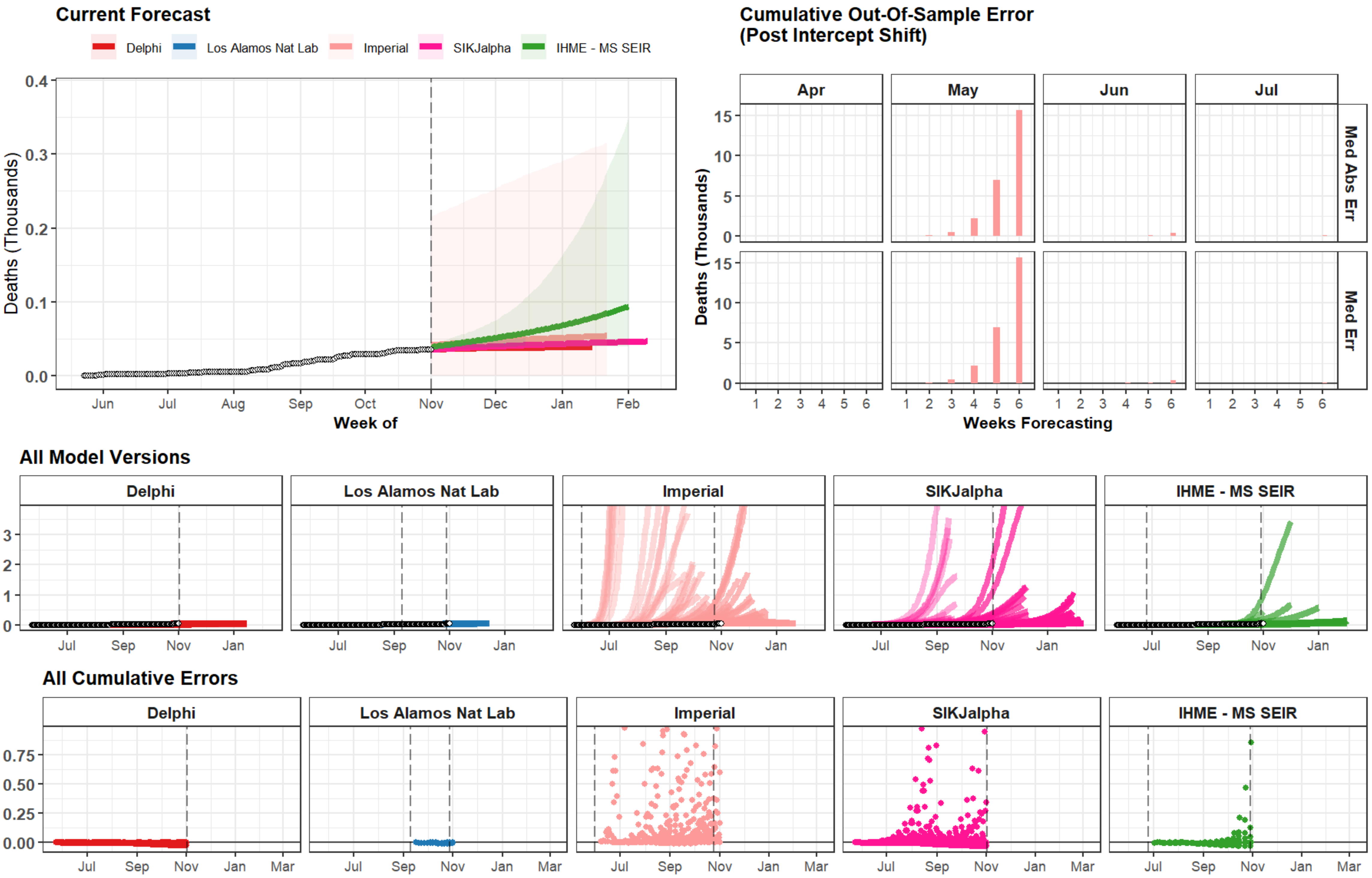
Current Forecast
Delphi
Los Alamos Nat Lab
Imperial
SIKJalpha
IHME - MS SEIR
Deaths (Thousands)
0.4
0.3
0.2
0.1
0.0
Jun
Jul
Aug
Sep
Oct
Nov
Dec
Jan
Feb
Week of
Cumulative Out-Of-Sample Error
(Post Intercept Shift)
Apr
May
Jun
Jul
Med Abs Err
Med Err
Deaths (Thousands)
15
10
5
0
1 2 3 4 5 6
Weeks Forecasting
All Model Versions
Delphi
Los Alamos Nat Lab
Imperial
SIKJalpha
IHME - MS SEIR
3
2
1
0
Jul
Sep
Nov
Jan
All Cumulative Errors
Delphi
Los Alamos Nat Lab
Imperial
SIKJalpha
IHME - MS SEIR
0.75
0.50
0.25
0.00
Jul
Sep
Nov
Jan
Mar

### Vietnam

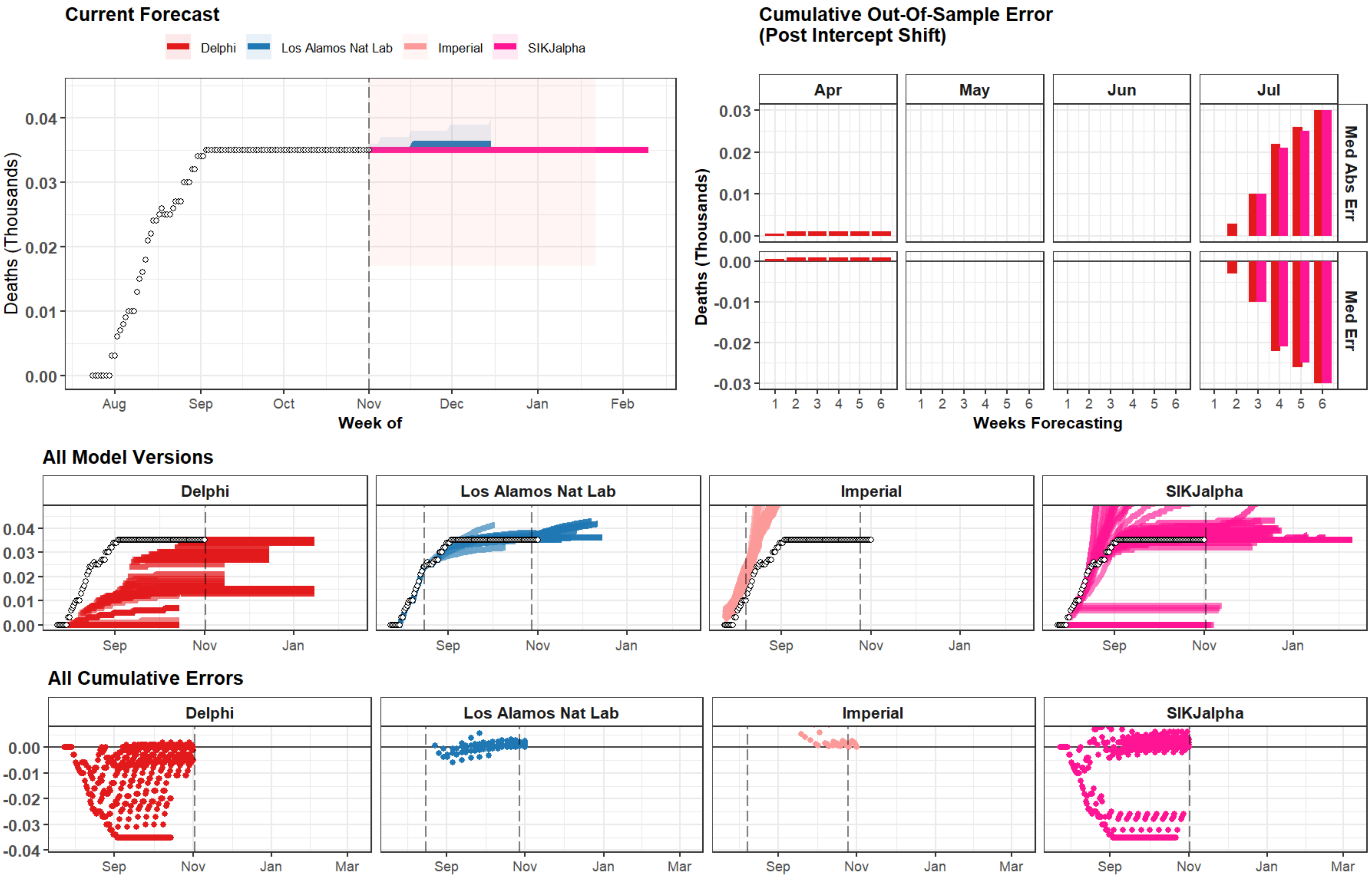
Current Forecast
Delphi
Los Alamos Nat Lab
Imperial
SIKJalpha
Deaths (Thousands)
Week of
Cumulative Out-Of-Sample Error
(Post Intercept Shift)
Apr
May
Jun
Jul
Med Abs Err
Med Err
Weeks Forecasting
All Model Versions
All Cumulative Errors

### Singapore

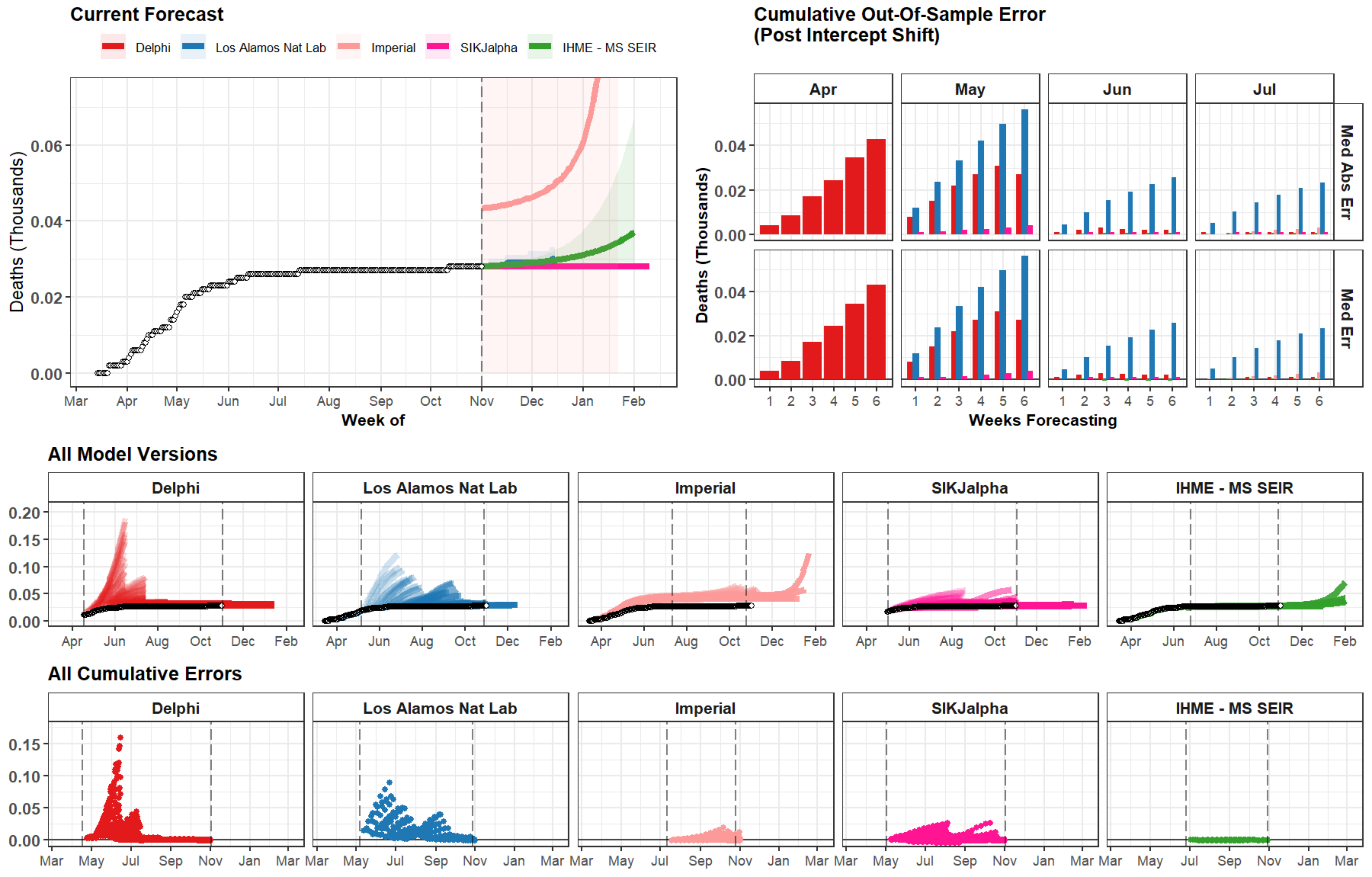
Current Forecast
Delphi
Los Alamos Nat Lab
Imperial
SIKJalpha
IHME - MS SEIR
Deaths (Thousands)
0.00
0.02
0.04
0.06
Mar
Apr
May
Jun
Jul
Aug
Sep
Oct
Nov
Dec
Jan
Feb
Week of
Cumulative Out-Of-Sample Error
(Post Intercept Shift)
Apr
May
Jun
Jul
Med Abs Err
Med Err
Weeks Forecasting
All Model Versions
0.20
0.15
0.10
0.05
All Cumulative Errors

### Cyprus

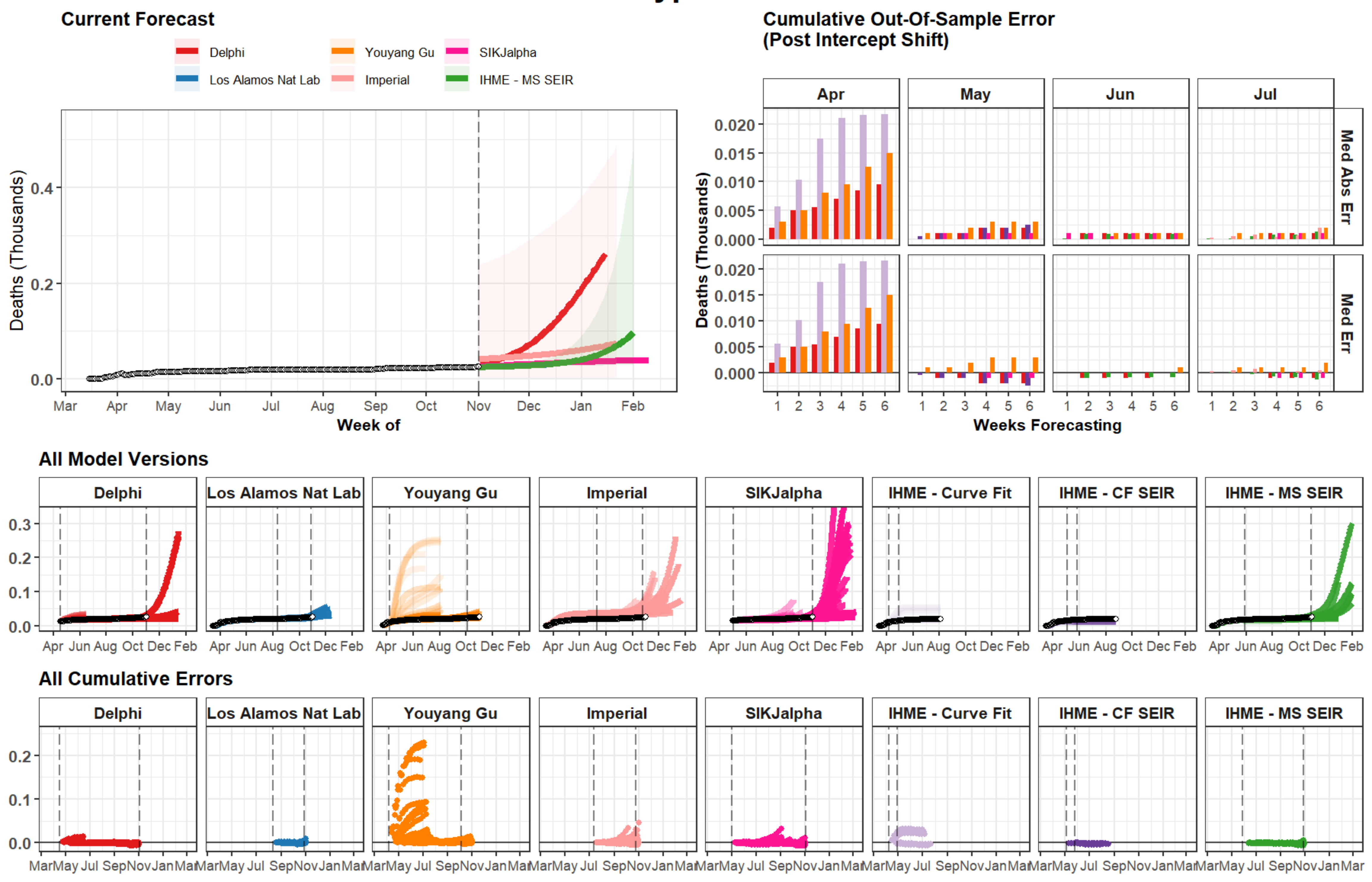

Current Forecast
Delphi
Youyang Gu
SIKJalpha
Los Alamos Nat Lab
Imperial
IHME - MS SEIR
Deaths (Thousands)
Week of
Mar Apr May Jun Jul Aug Sep Oct Nov Dec Jan Feb
Cumulative Out-Of-Sample Error
(Post Intercept Shift)
Apr
May
Jun
Jul
Med Abs Err
Med Err
Weeks Forecasting
All Model Versions
Delphi
Los Alamos Nat Lab
Youyang Gu
Imperial
SIKJalpha
IHME - Curve Fit
IHME - CF SEIR
IHME - MS SEIR
All Cumulative Errors

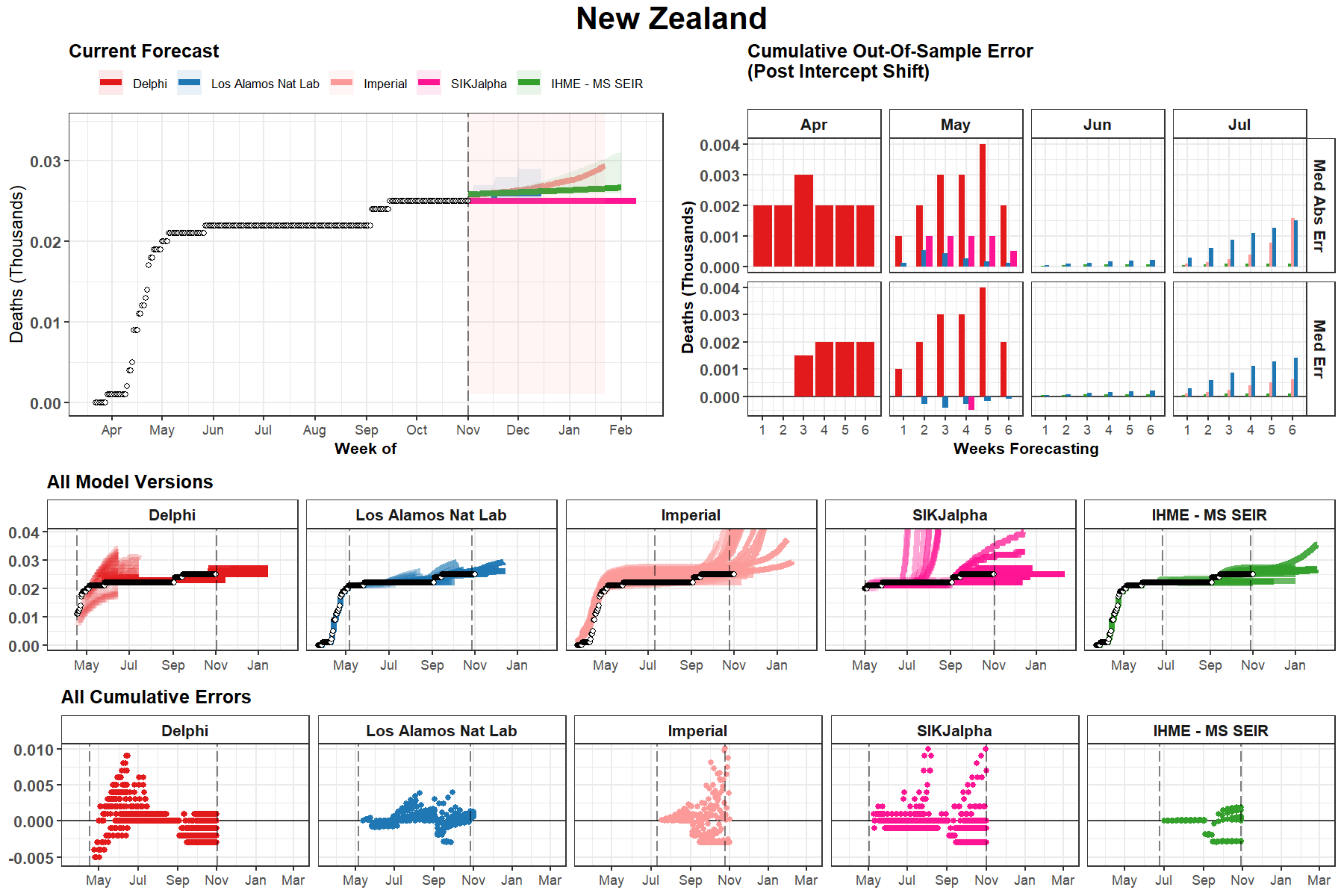
New Zealand
Current Forecast
Delphi
Los Alamos Nat Lab
Imperial
SIKJalpha
IHME - MS SEIR
Deaths (Thousands)
0.03
0.02
0.01
0.00
Apr
May
Jun
Jul
Aug
Sep
Oct
Nov
Dec
Jan
Feb
Week of
Cumulative Out-Of-Sample Error
(Post Intercept Shift)
Apr
May
Jun
Jul
Med Abs Err
Med Err
0.004
0.003
0.002
0.001
0.000
Weeks Forecasting
All Model Versions
All Cumulative Errors
0.010
0.005
0.000
-0.005
Mar

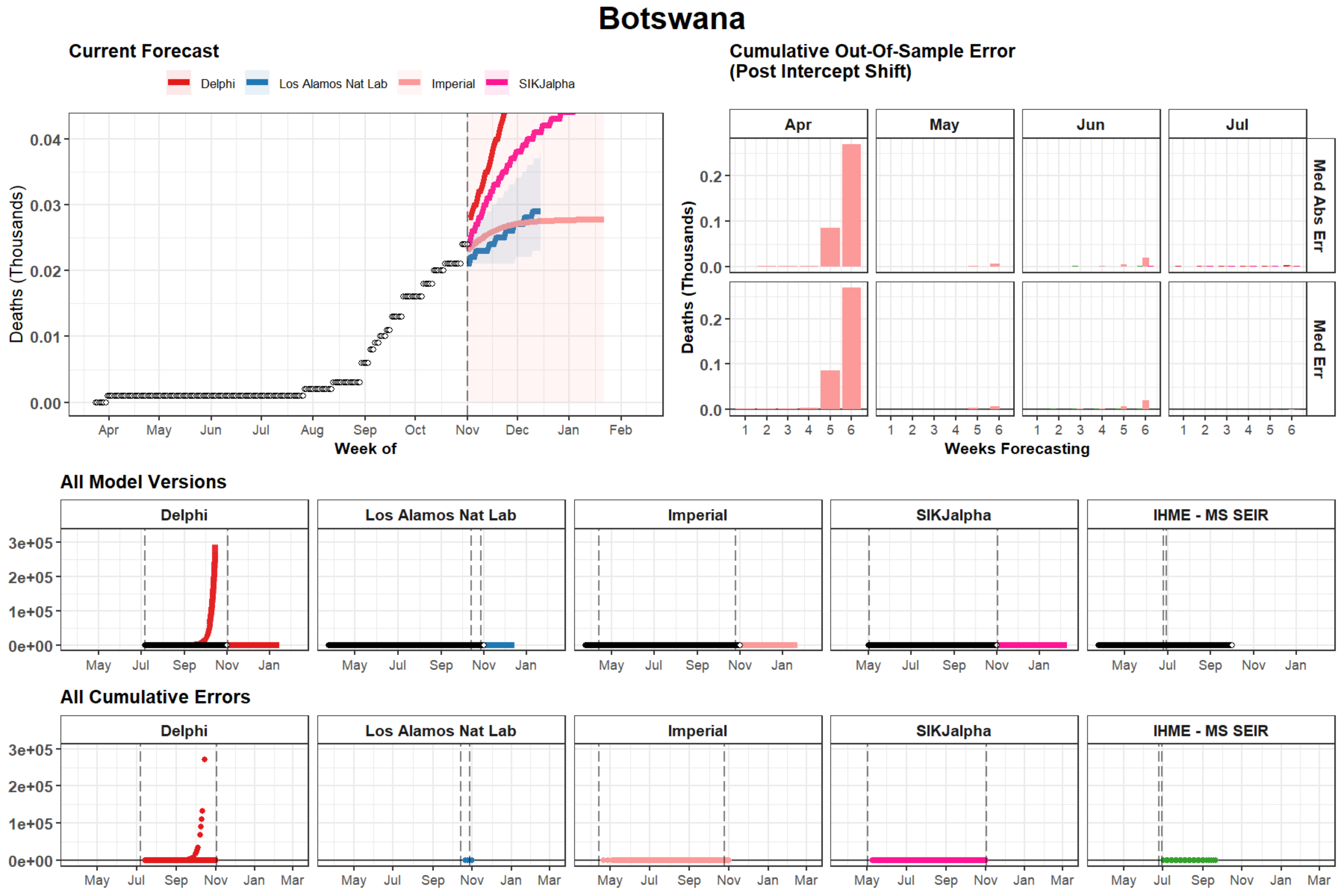

Botswana
Current Forecast
Delphi
Los Alamos Nat Lab
Imperial
SIKJalpha
Deaths (Thousands)
0.04
0.03
0.02
0.01
0.00
Apr May Jun Jul Aug Sep Oct Nov Dec Jan Feb
Week of
Cumulative Out-Of-Sample Error (Post Intercept Shift)
Apr May Jun Jul
Med Abs Err
Med Err
Deaths (Thousands)
0.2
0.1
0.0
1 2 3 4 5 6
Weeks Forecasting
All Model Versions
Delphi
Los Alamos Nat Lab
Imperial
SIKJalpha
IHME - MS SEIR
3e+05
2e+05
1e+05
0e+00
May Jul Sep Nov Jan
All Cumulative Errors
Delphi
Los Alamos Nat Lab
Imperial
SIKJalpha
IHME - MS SEIR
3e+05
2e+05
1e+05
0e+00
May Jul Sep Nov Jan Mar

### Sri Lanka

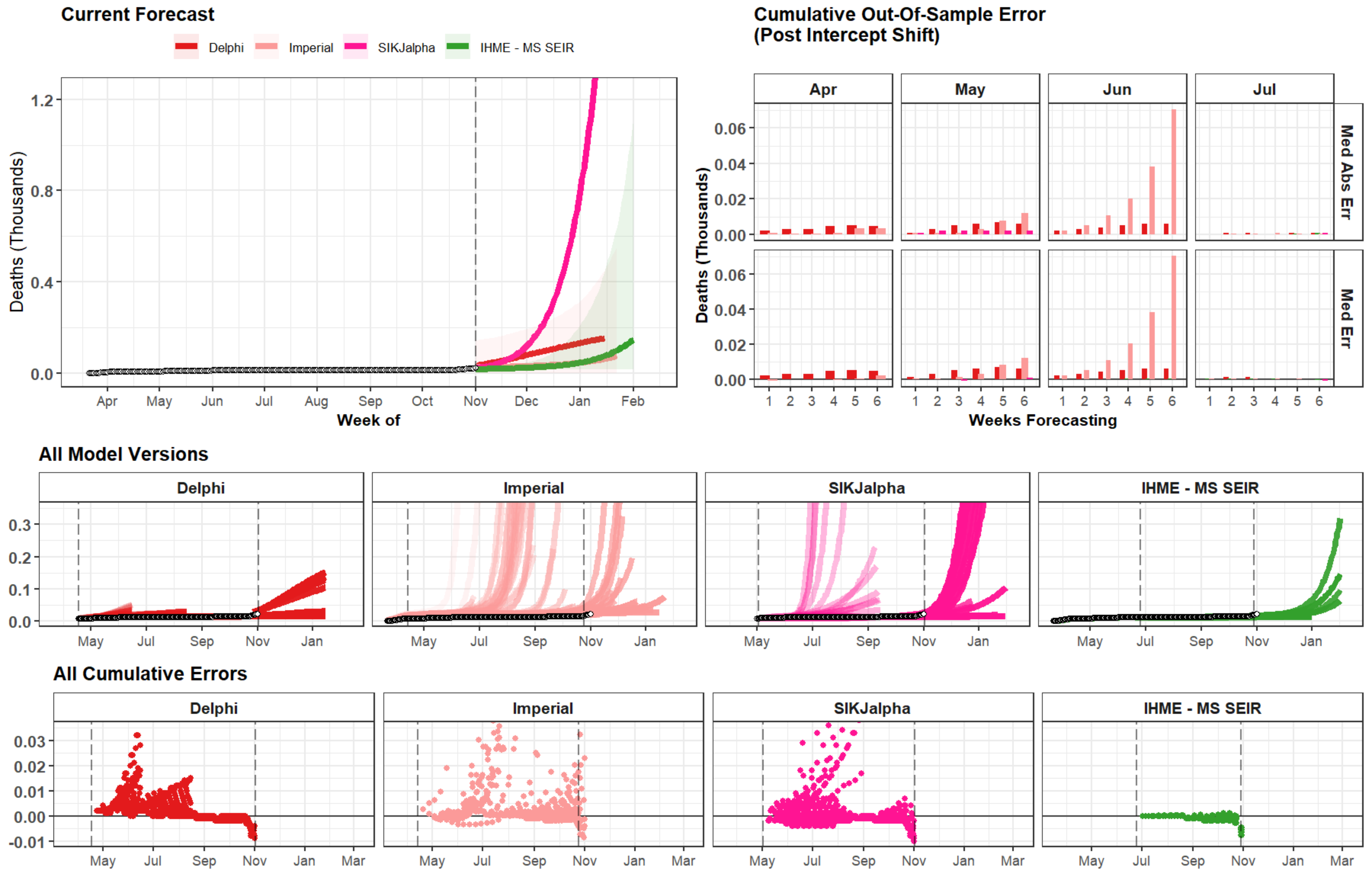

Current Forecast
Delphi
Imperial
SIKJalpha
IHME - MS SEIR
Deaths (Thousands)
1.2
0.8
0.4
0.0
Apr
May
Jun
Jul
Aug
Sep
Oct
Nov
Dec
Jan
Feb
Week of
Cumulative Out-Of-Sample Error
(Post Intercept Shift)
Apr
May
Jun
Jul
Med Abs Err
Med Err
0.06
0.04
0.02
0.00
1
2
3
4
5
6
Weeks Forecasting
All Model Versions
Delphi
Imperial
SIKJalpha
IHME - MS SEIR
0.3
0.2
0.1
0.0
May
Jul
Sep
Nov
Jan
All Cumulative Errors
Delphi
Imperial
SIKJalpha
IHME - MS SEIR
0.03
0.02
0.01
0.00
-0.01
May
Jul
Sep
Nov
Jan
Mar

### Tanzania

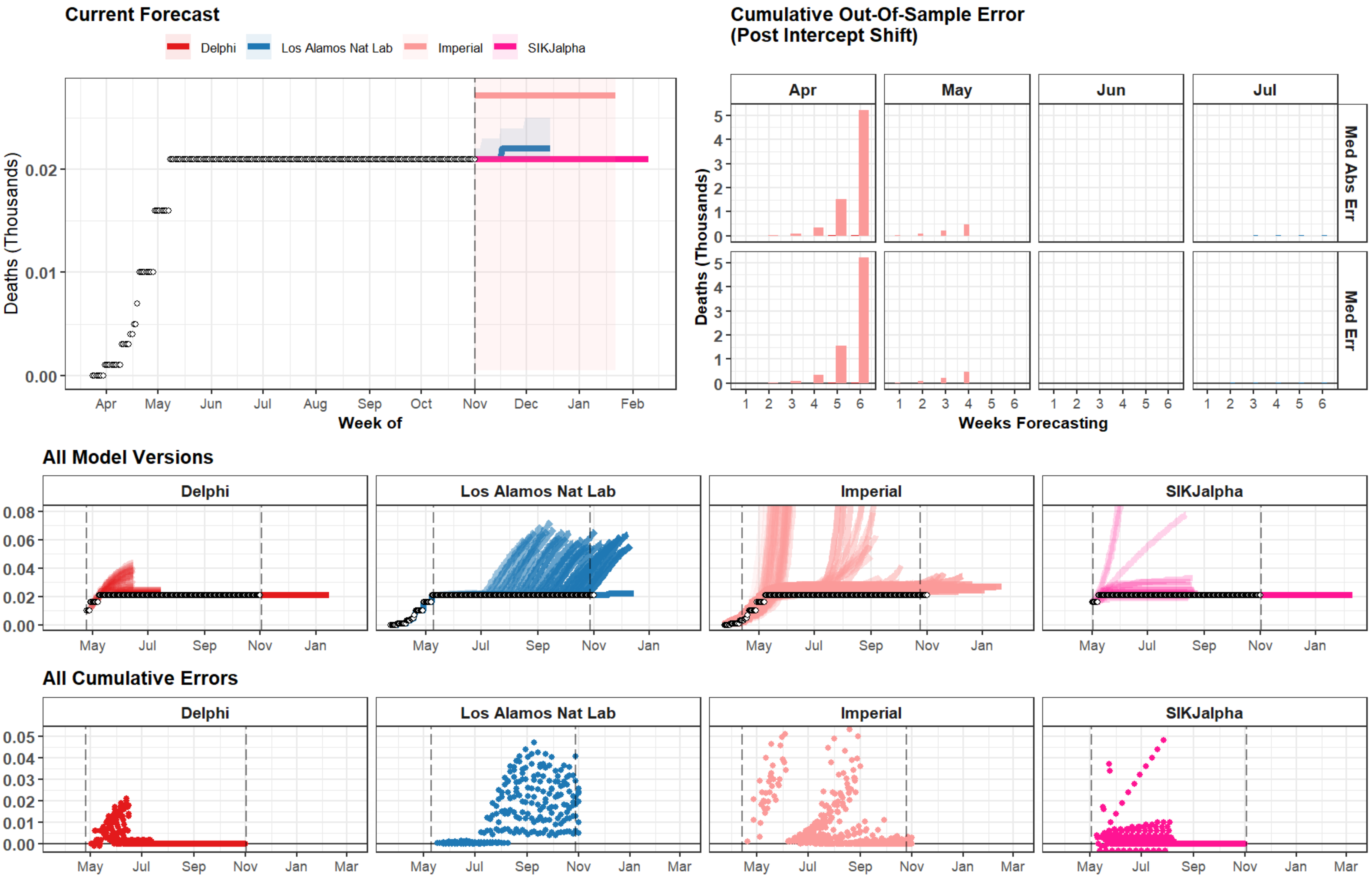

Current Forecast
Delphi
Los Alamos Nat Lab
Imperial
SIKJalpha
Deaths (Thousands)
0.00
0.01
0.02
Apr
May
Jun
Jul
Aug
Sep
Oct
Nov
Dec
Jan
Feb
Week of
Cumulative Out-Of-Sample Error
(Post Intercept Shift)
Apr
May
Jun
Jul
Med Abs Err
Med Err
Deaths (Thousands)
0
1
2
3
4
5
1 2 3 4 5 6
Weeks Forecasting
All Model Versions
Delphi
Los Alamos Nat Lab
Imperial
SIKJalpha
0.00
0.02
0.04
0.06
0.08
May
Jul
Sep
Nov
Jan
All Cumulative Errors
Delphi
Los Alamos Nat Lab
Imperial
SIKJalpha
0.00
0.01
0.02
0.03
0.04
0.05
May
Jul
Sep
Nov
Jan
Mar

### Virgin Islands, U.S.

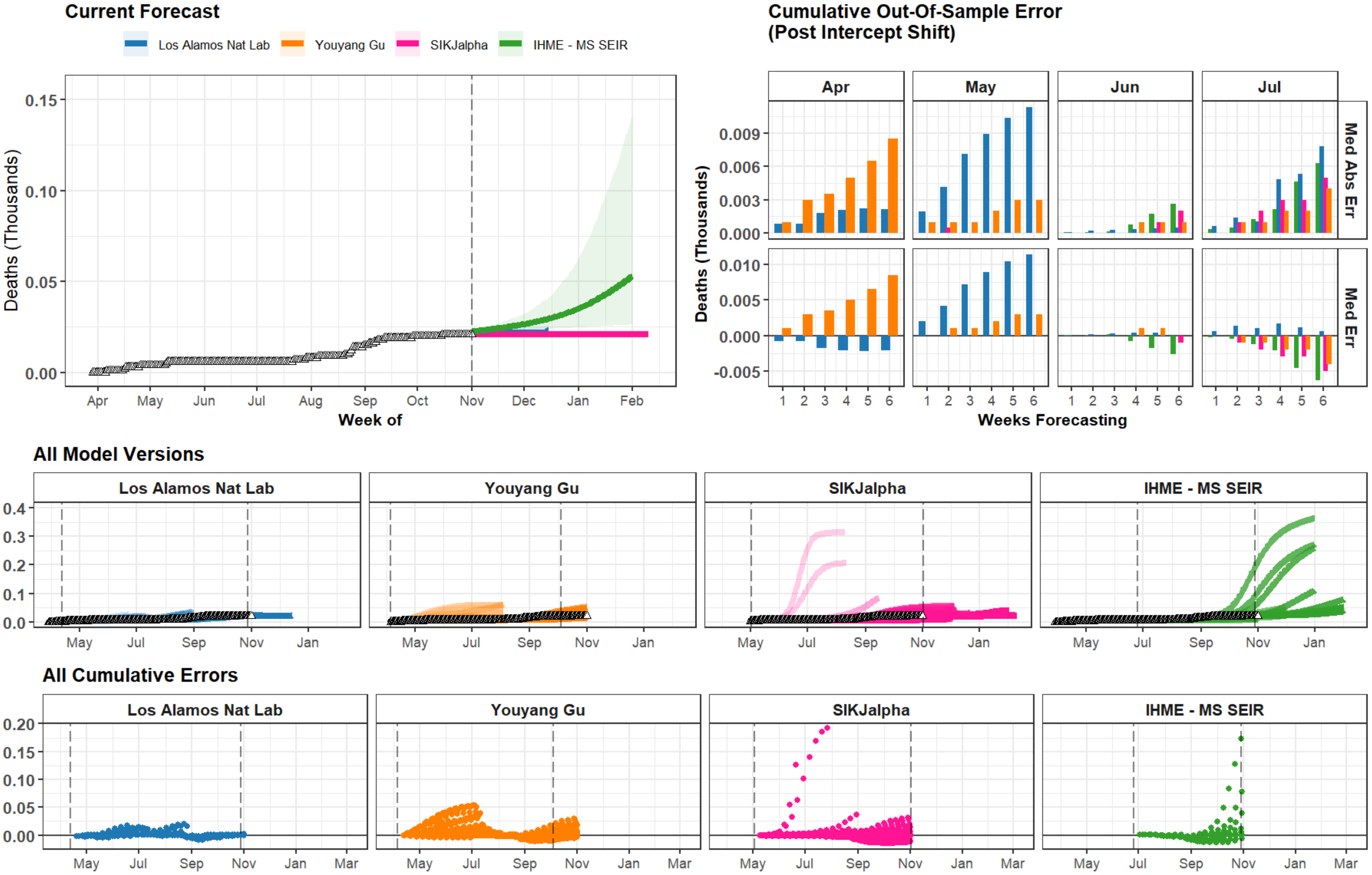

### Sao Tome and Principe

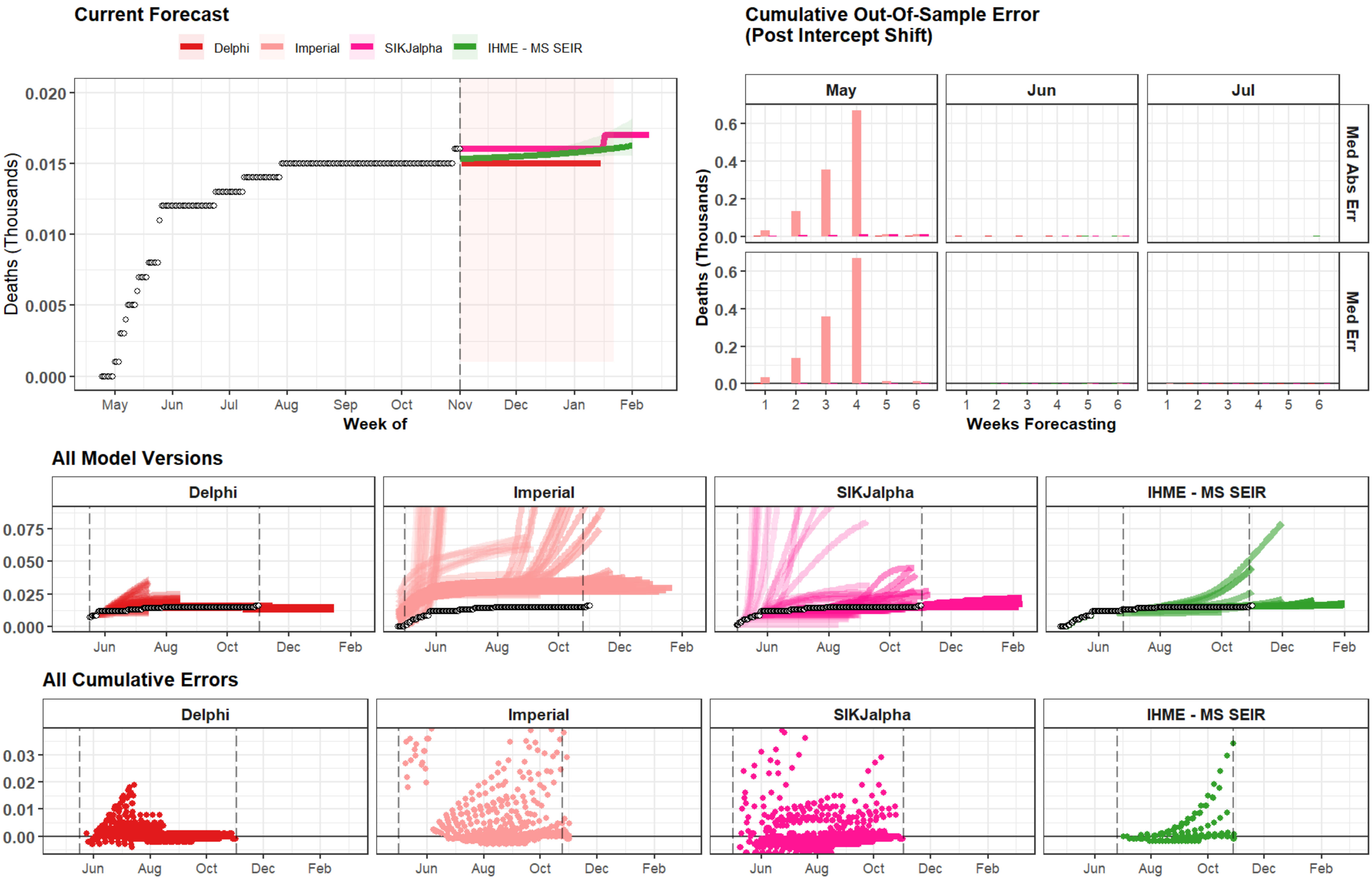

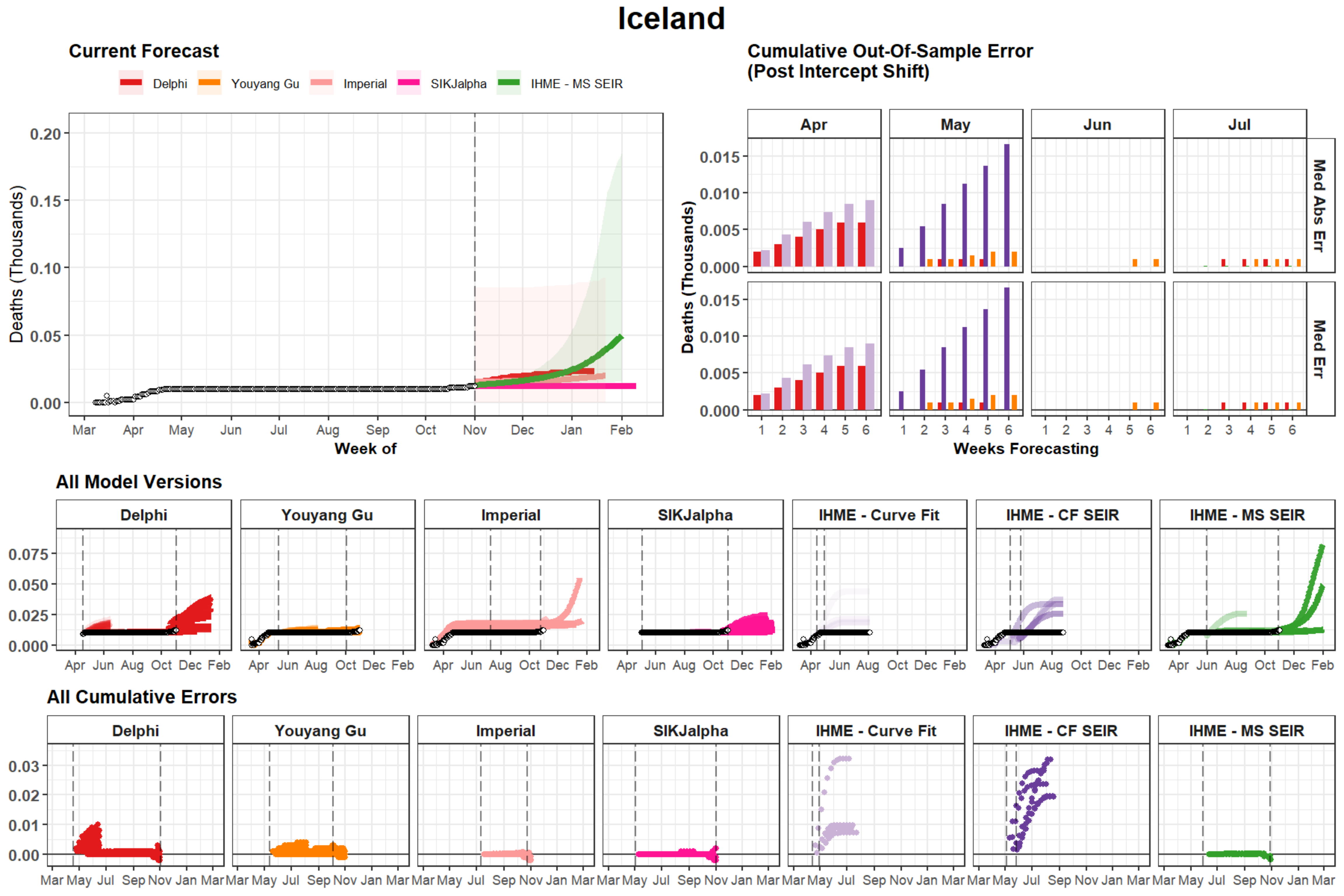

Iceland
Current Forecast
Delphi
Youyang Gu
Imperial
SIKJalpha
IHME - MS SEIR
Deaths (Thousands)
0.20
0.15
0.10
0.05
0.00
Mar
Apr
May
Jun
Jul
Aug
Sep
Oct
Nov
Dec
Jan
Feb
Week of
Cumulative Out-Of-Sample Error
(Post Intercept Shift)
Apr
May
Jun
Jul
Med Abs Err
Med Err
0.015
0.010
0.005
0.000
Weeks Forecasting
All Model Versions
Delphi
Youyang Gu
Imperial
SIKJalpha
IHME - Curve Fit
IHME - CF SEIR
IHME - MS SEIR
0.075
0.050
0.025
0.000
All Cumulative Errors
0.03
0.02
0.01
0.00
