## Supplemental Figures - Peak Timing for "Predictive performance of international COVID-19 mortality forecasting models"

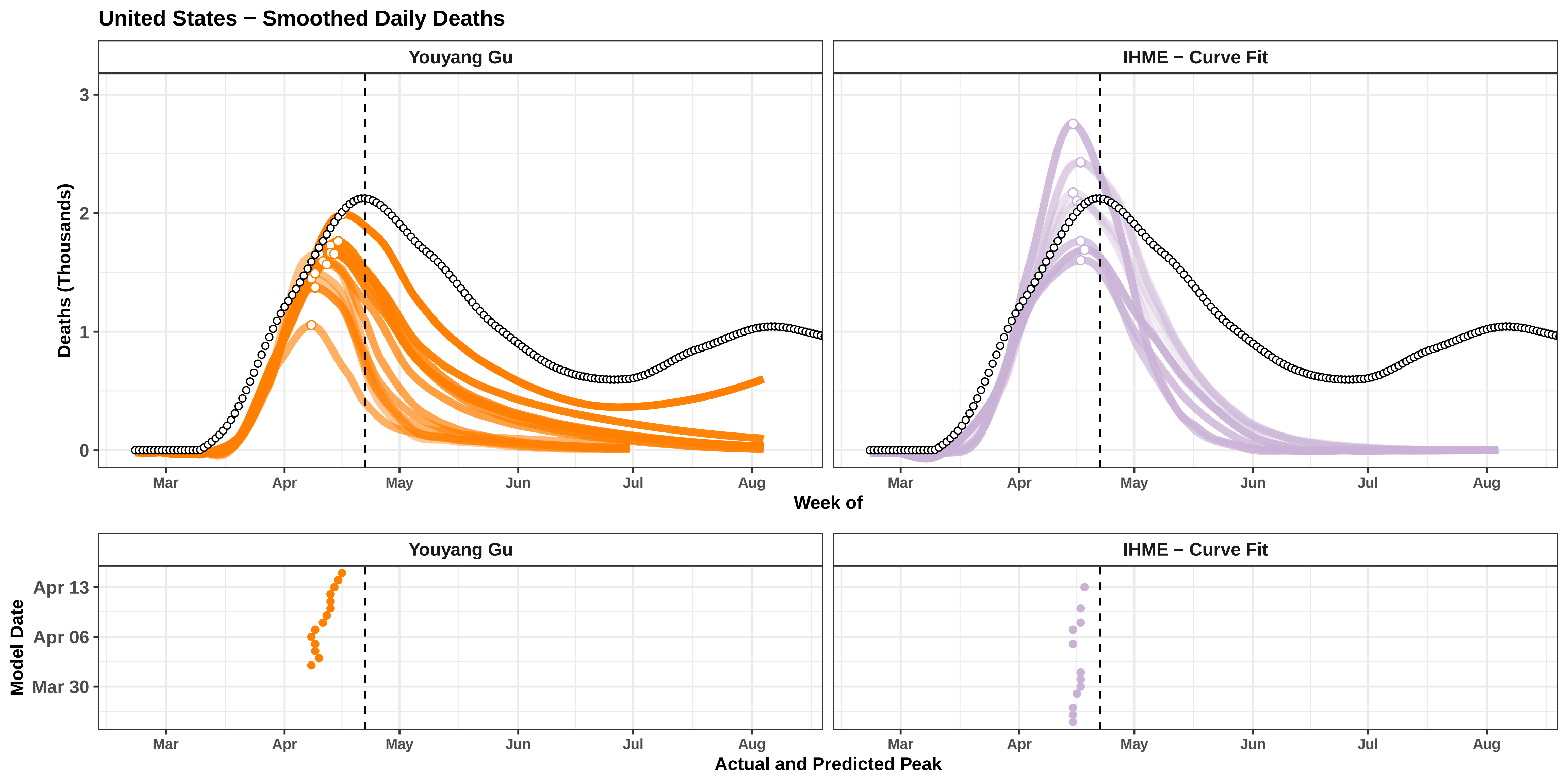
United States − Smoothed Daily Deaths
Youyang Gu
IHME − Curve Fit
Deaths (Thousands)
0
1
2
3
Mar
Apr
May
Jun
Jul
Aug
Week of
Model Date
Apr 13
Apr 06
Mar 30
Actual and Predicted Peak

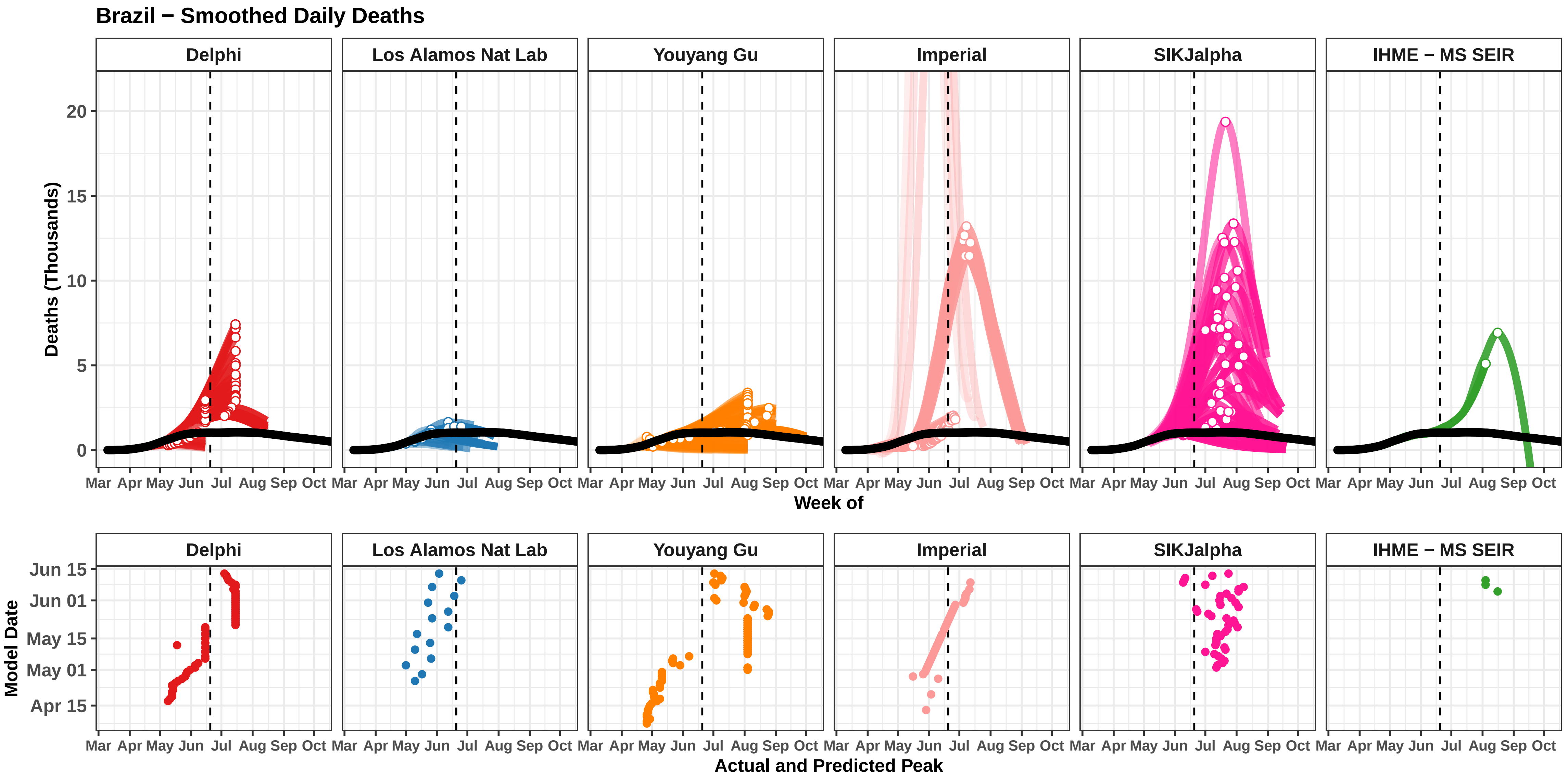
Brazil − Smoothed Daily Deaths
Delphi
Los Alamos Nat Lab
Youyang Gu
Imperial
SIKJalpha
IHME − MS SEIR
Deaths (Thousands)
0
5
10
15
20
Mar Apr May Jun Jul Aug Sep Oct
Week of
Model Date
Jun 15
Jun 01
May 15
May 01
Apr 15
Actual and Predicted Peak

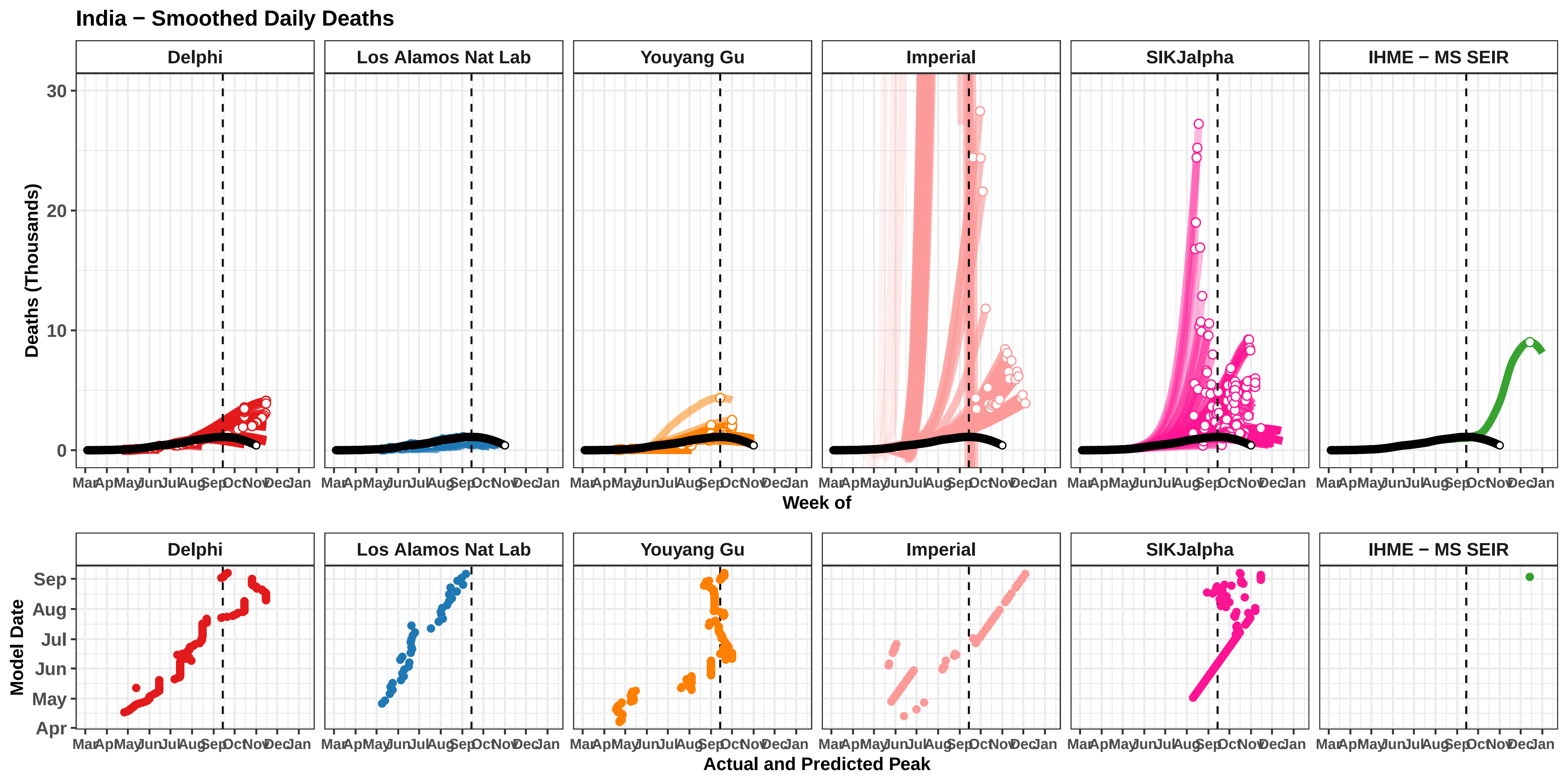

India − Smoothed Daily Deaths
Delphi
Los Alamos Nat Lab
Youyang Gu
Imperial
SIKJalpha
IHME − MS SEIR
Deaths (Thousands)
30
20
10
0
Mar Apr May Jun Jul Aug Sep Oct Nov Dec Jan
Week of
Model Date
Sep
Aug
Jul
Jun
May
Apr
Actual and Predicted Peak

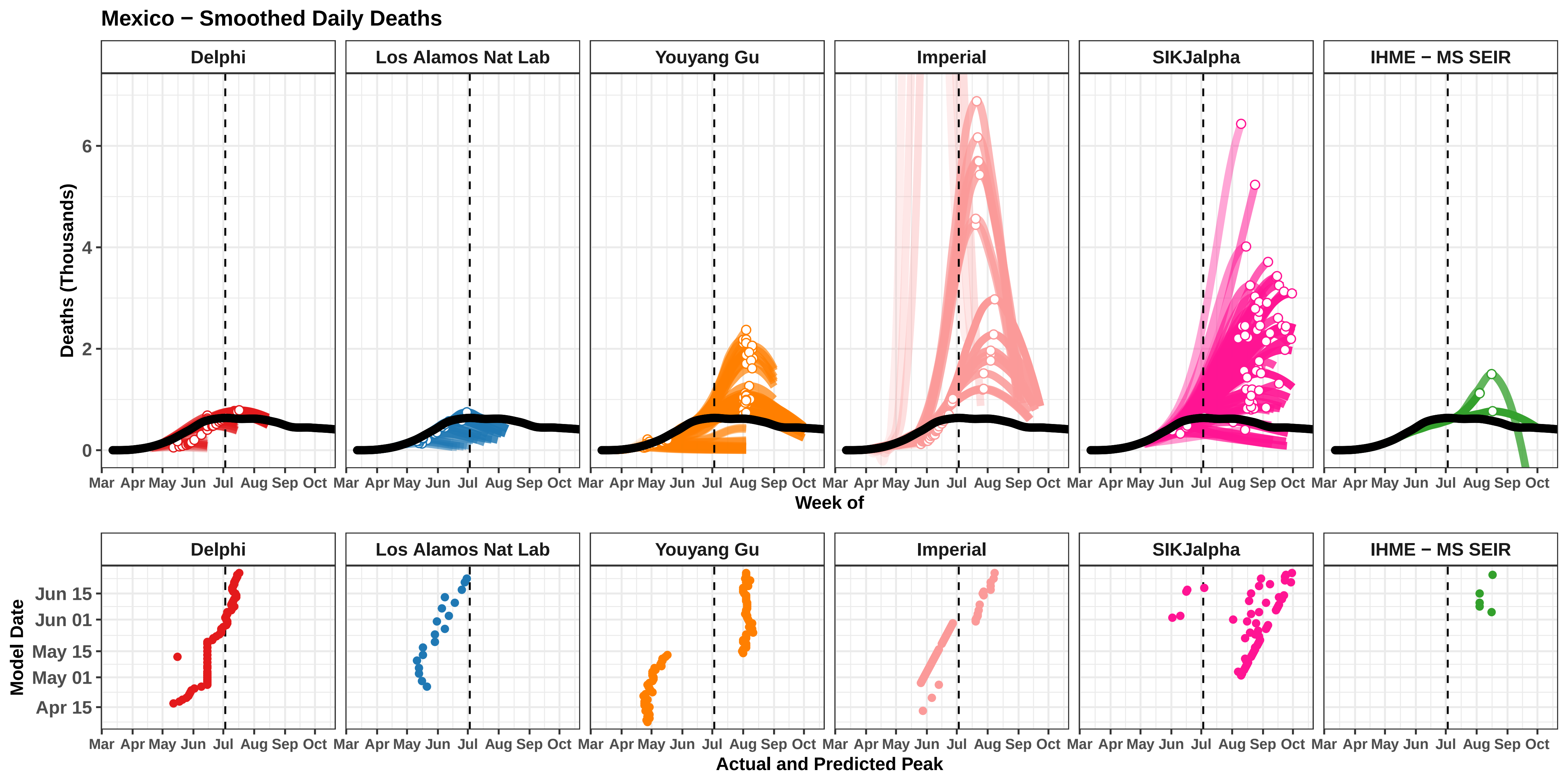

Mexico − Smoothed Daily Deaths
Delphi
Los Alamos Nat Lab
Youyang Gu
Imperial
SIKJalpha
IHME − MS SEIR
Deaths (Thousands)
0
2
4
6
Mar Apr May Jun Jul Aug Sep Oct
Week of
Model Date
Jun 15
Jun 01
May 15
May 01
Apr 15
Actual and Predicted Peak

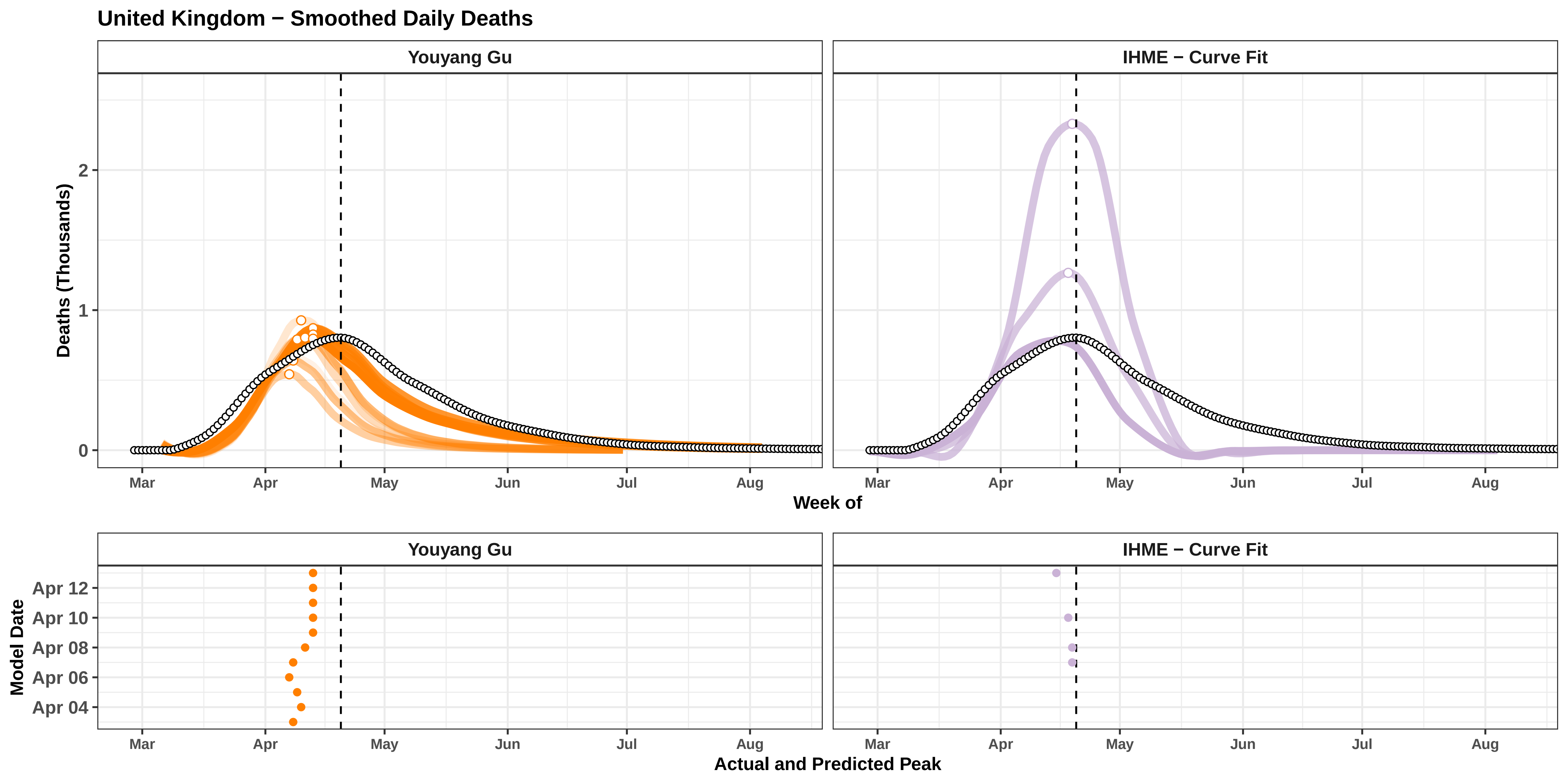

United Kingdom − Smoothed Daily Deaths
Youyang Gu
IHME − Curve Fit
Deaths (Thousands)
0
1
2
Mar
Apr
May
Jun
Jul
Aug
Week of
Model Date
Apr 04
Apr 06
Apr 08
Apr 10
Apr 12
Actual and Predicted Peak

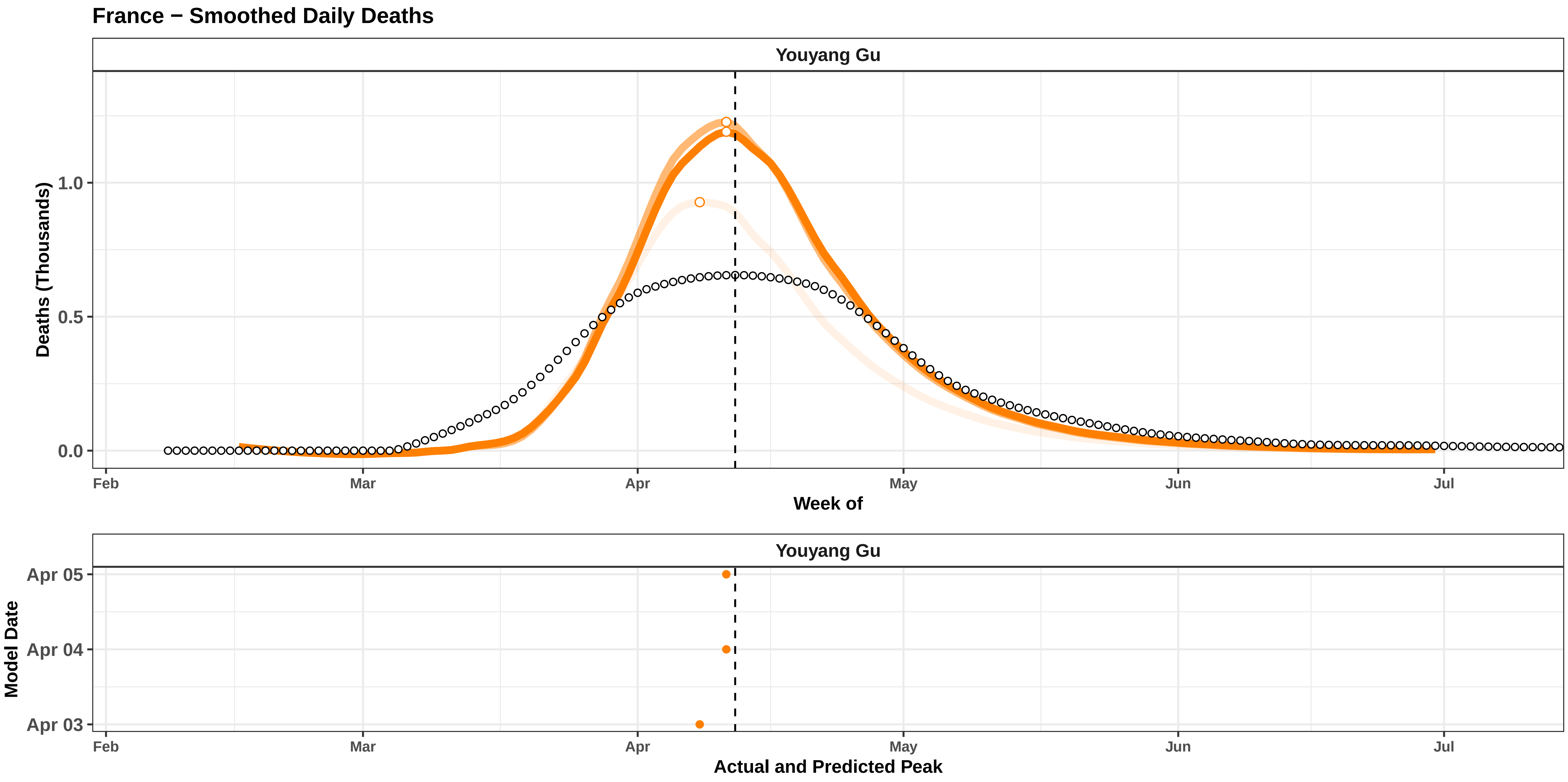
France − Smoothed Daily Deaths
Youyang Gu
Deaths (Thousands)
1.0
0.5
0.0
Feb
Mar
Apr
May
Jun
Jul
Week of
Youyang Gu
Model Date
Apr 05
Apr 04
Apr 03
Feb
Mar
Apr
May
Jun
Jul
Actual and Predicted Peak

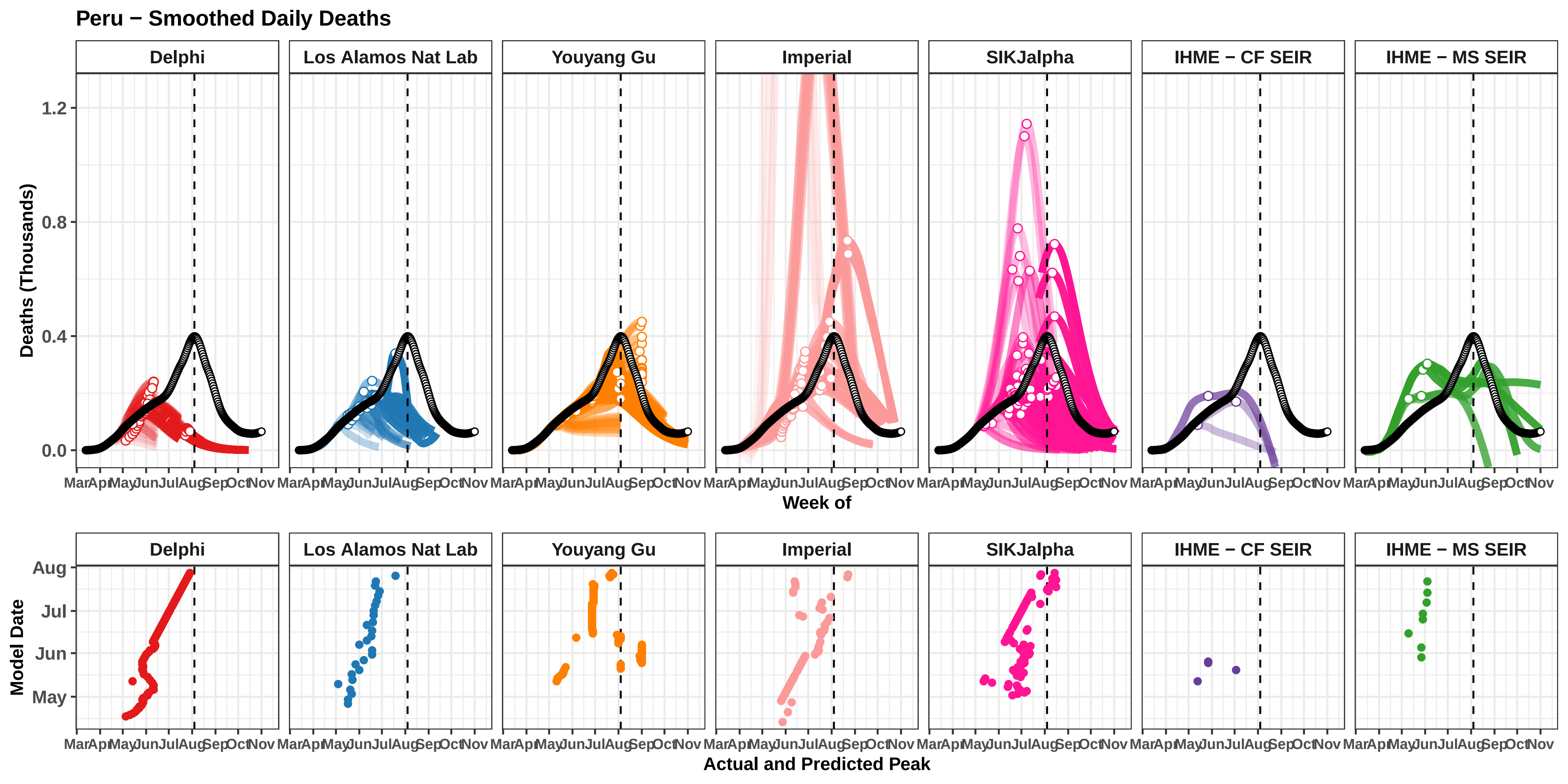
Peru − Smoothed Daily Deaths
Delphi
Los Alamos Nat Lab
Youyang Gu
Imperial
SIKJalpha
IHME − CF SEIR
IHME − MS SEIR
Deaths (Thousands)
0.0
0.4
0.8
1.2
Mar Apr May Jun Jul Aug Sep Oct Nov
Week of
Model Date
May
Jun
Jul
Aug
Actual and Predicted Peak

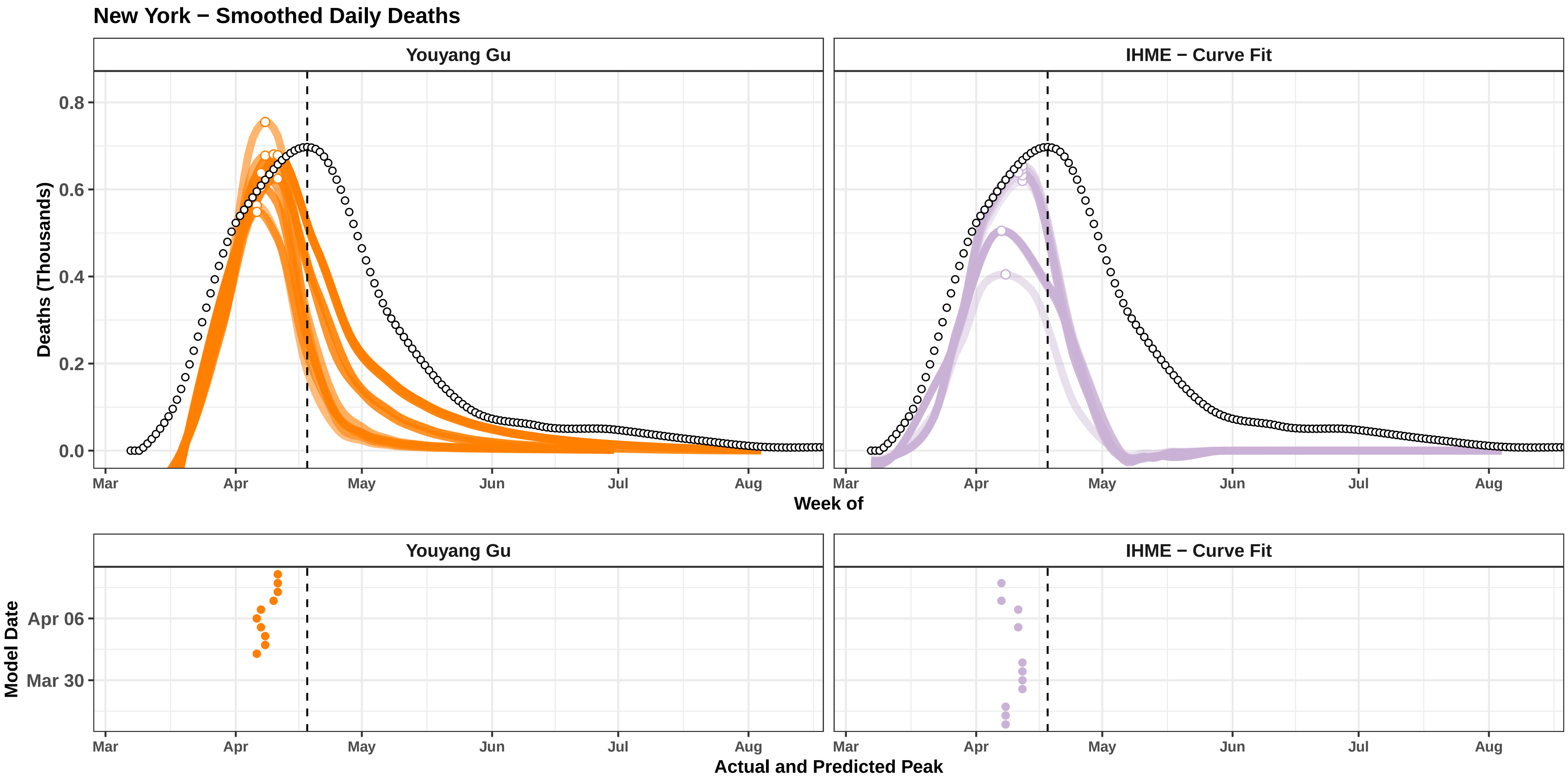

New York − Smoothed Daily Deaths
Youyang Gu
IHME − Curve Fit
Deaths (Thousands)
0.8
0.6
0.4
0.2
0.0
Mar
Apr
May
Jun
Jul
Aug
Week of
Model Date
Apr 06
Mar 30
Actual and Predicted Peak

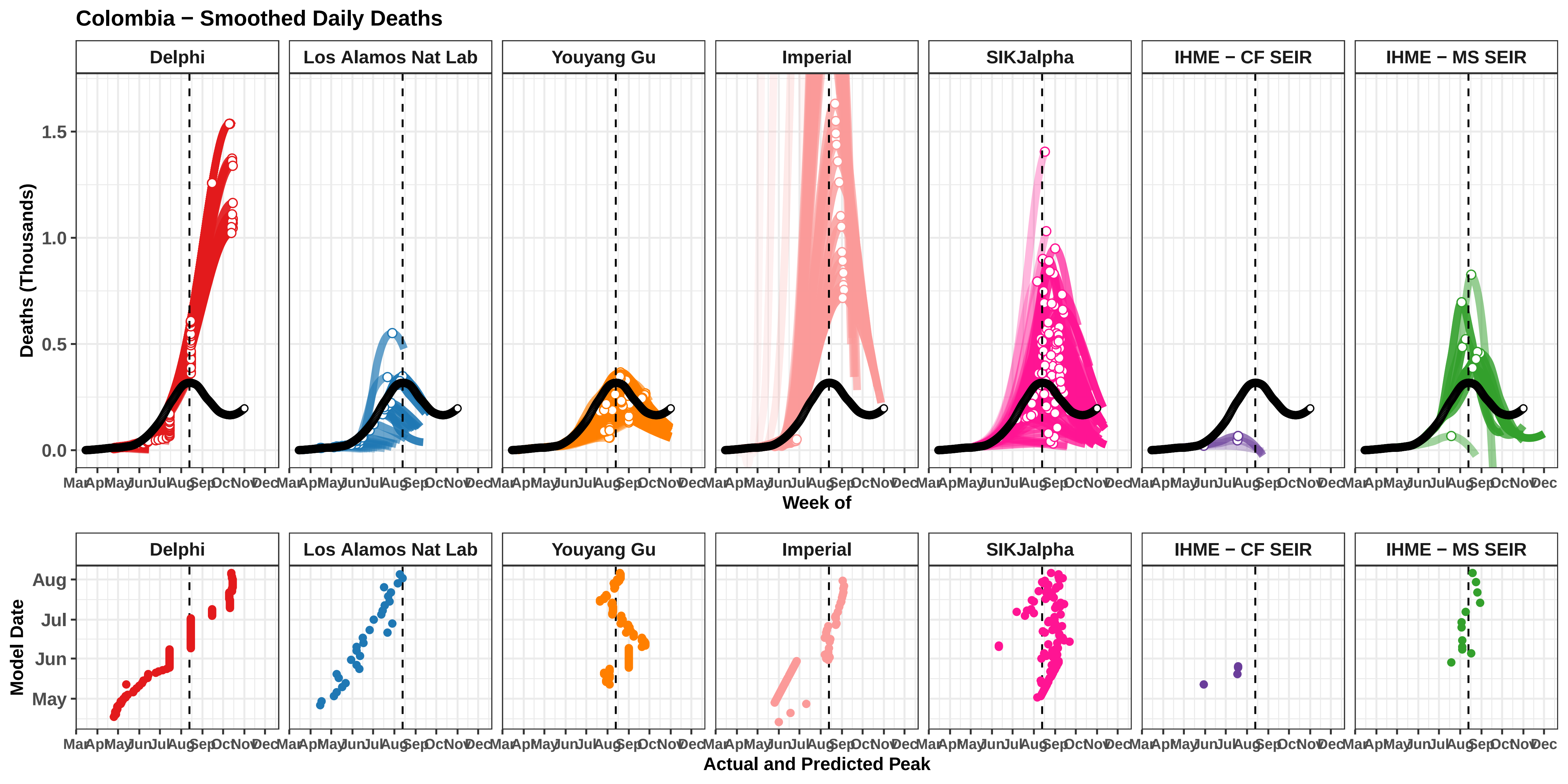

Colombia − Smoothed Daily Deaths
Delphi
Los Alamos Nat Lab
Youyang Gu
Imperial
SIKJalpha
IHME − CF SEIR
IHME − MS SEIR
Deaths (Thousands)
0.0
0.5
1.0
1.5
Mar Apr May Jun Jul Aug Sep Oct Nov Dec
Week of
Model Date
May
Jun
Jul
Aug
Actual and Predicted Peak

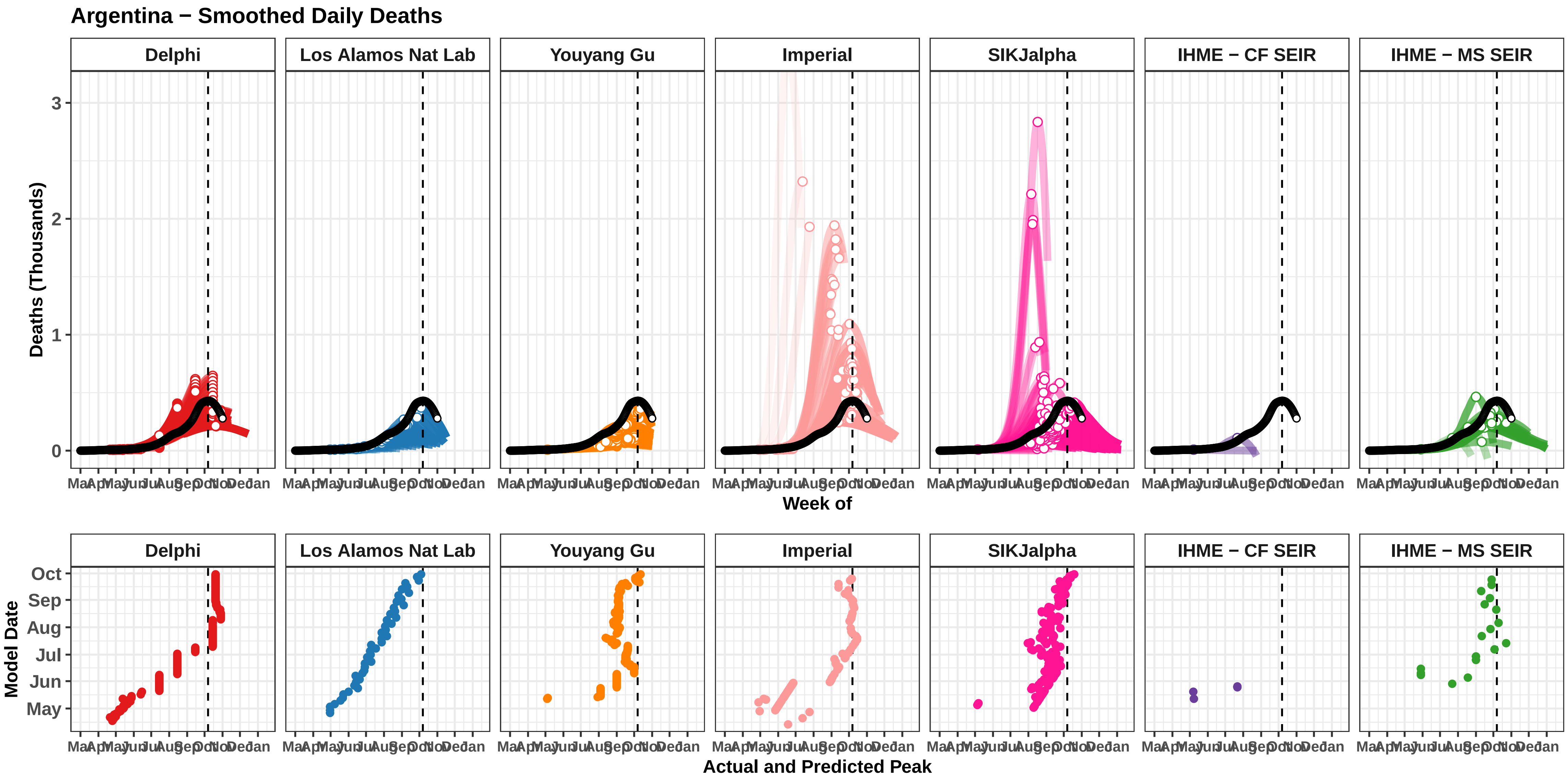

Argentina − Smoothed Daily Deaths
Delphi
Los Alamos Nat Lab
Youyang Gu
Imperial
SIKJalpha
IHME − CF SEIR
IHME − MS SEIR
Deaths (Thousands)
0
1
2
3
Week of
Model Date
Oct
Sep
Aug
Jul
Jun
May
Actual and Predicted Peak

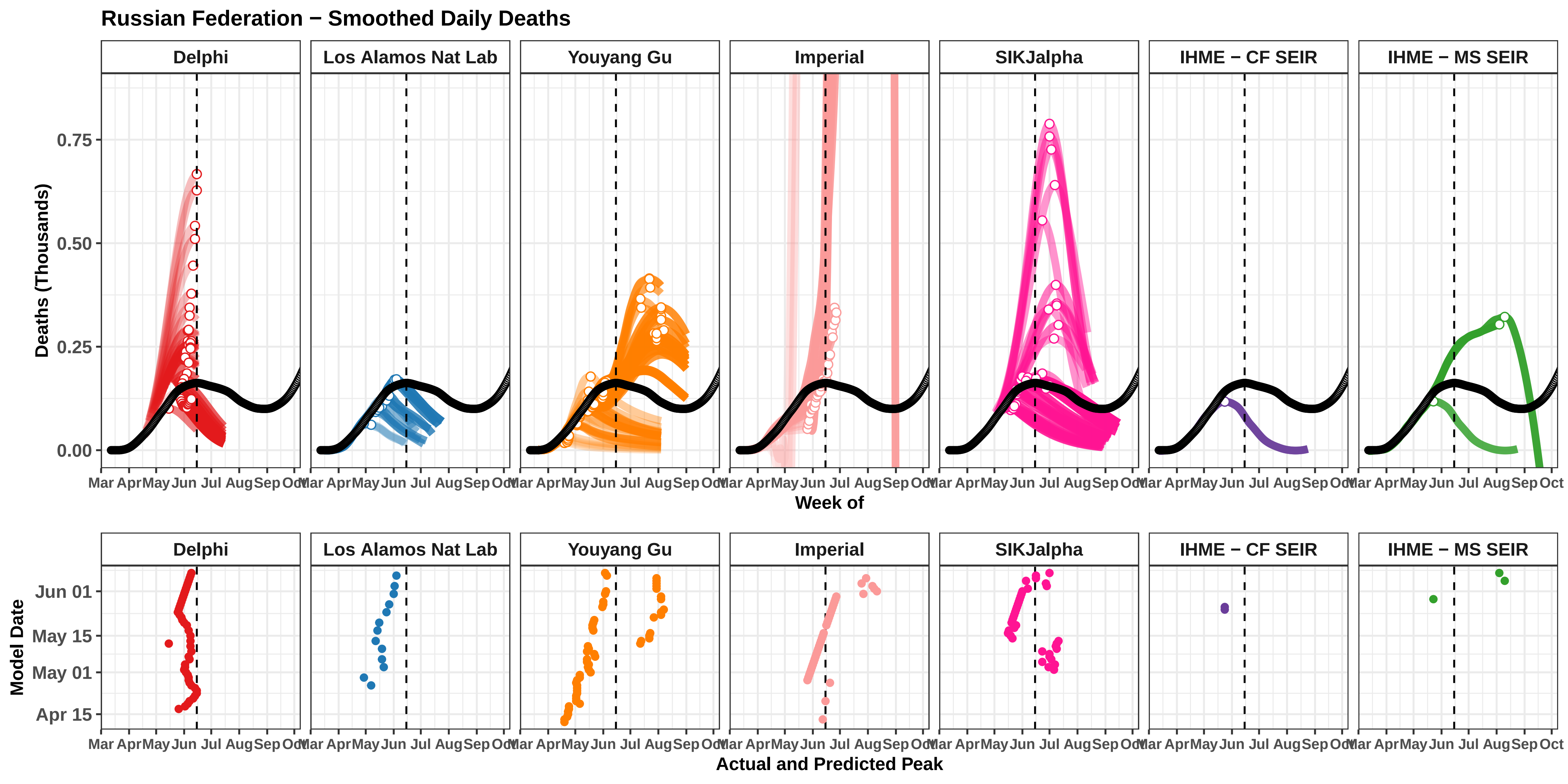

Russian Federation − Smoothed Daily Deaths
Delphi
Los Alamos Nat Lab
Youyang Gu
Imperial
SIKJalpha
IHME − CF SEIR
IHME − MS SEIR
Deaths (Thousands)
0.75
0.50
0.25
0.00
Mar Apr May Jun Jul Aug Sep Oct
Week of
Model Date
Jun 01
May 15
May 01
Apr 15
Actual and Predicted Peak

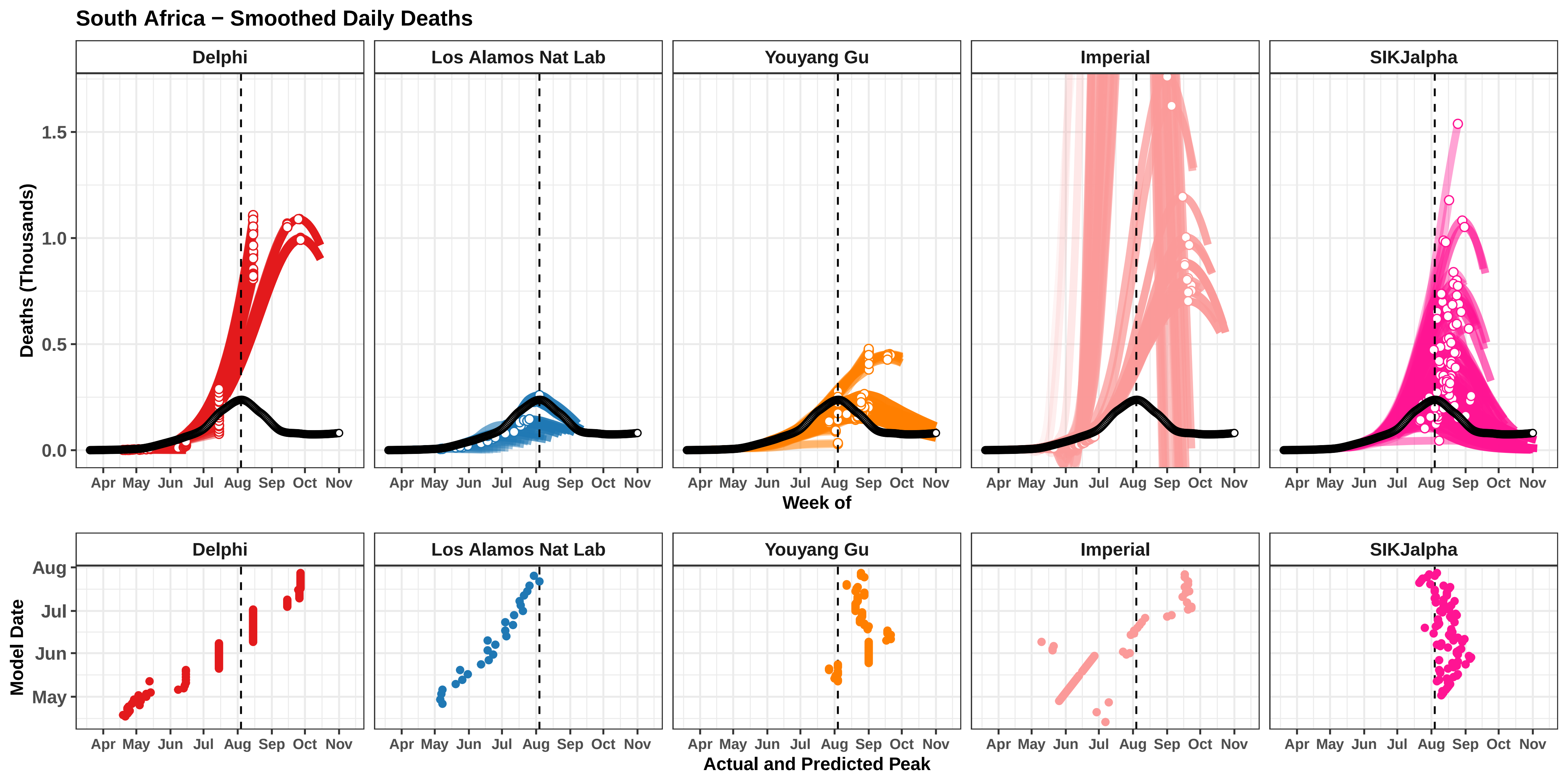

South Africa − Smoothed Daily Deaths
Delphi
Los Alamos Nat Lab
Youyang Gu
Imperial
SIKJalpha
Deaths (Thousands)
0.0
0.5
1.0
1.5
Apr May Jun Jul Aug Sep Oct Nov
Week of
Model Date
May
Jun
Jul
Aug
Actual and Predicted Peak

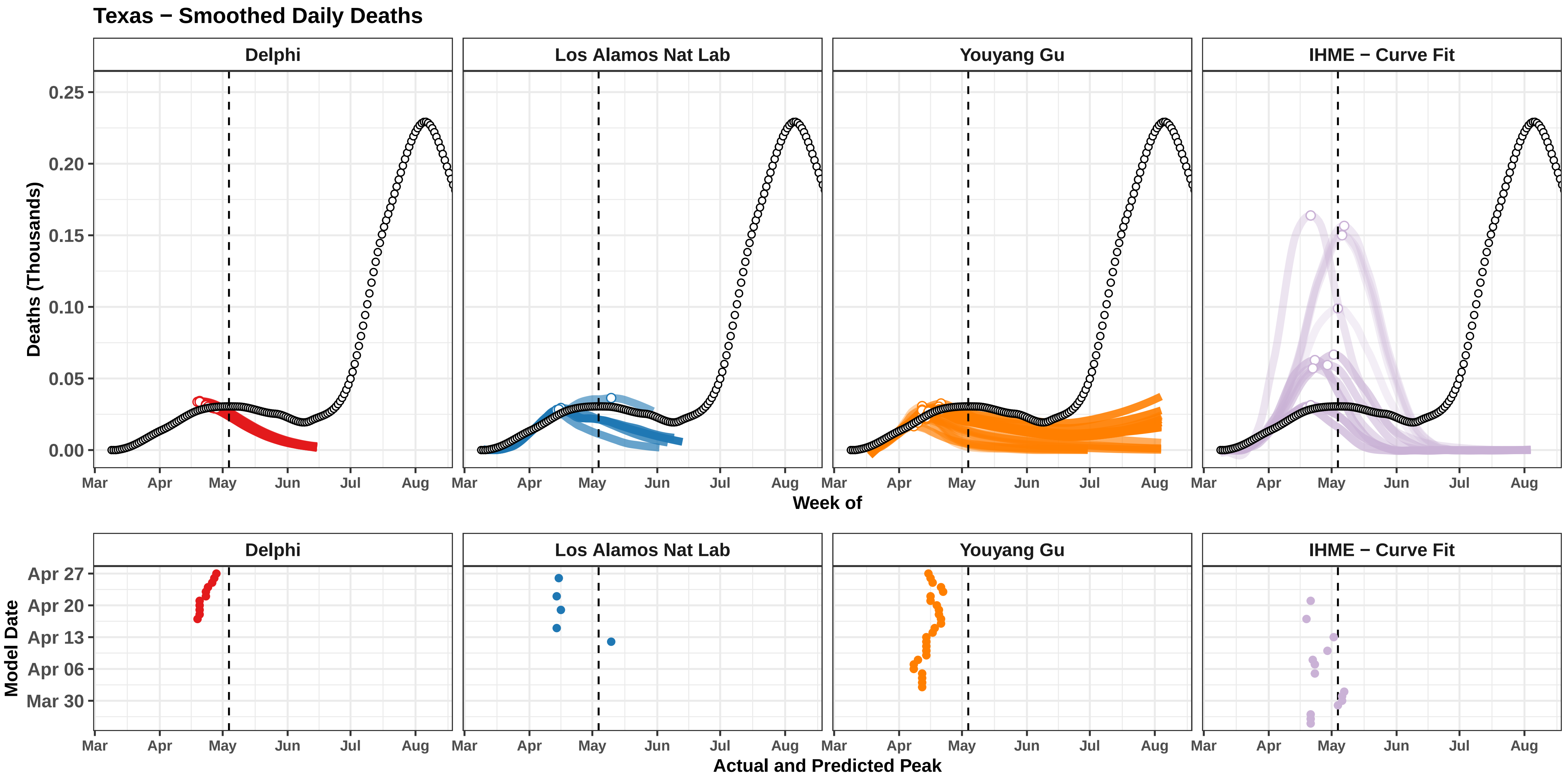

Texas − Smoothed Daily Deaths
Delphi
Los Alamos Nat Lab
Youyang Gu
IHME − Curve Fit
Deaths (Thousands)
0.25
0.20
0.15
0.10
0.05
0.00
Mar
Apr
May
Jun
Jul
Aug
Week of
Model Date
Apr 27
Apr 20
Apr 13
Apr 06
Mar 30
Actual and Predicted Peak

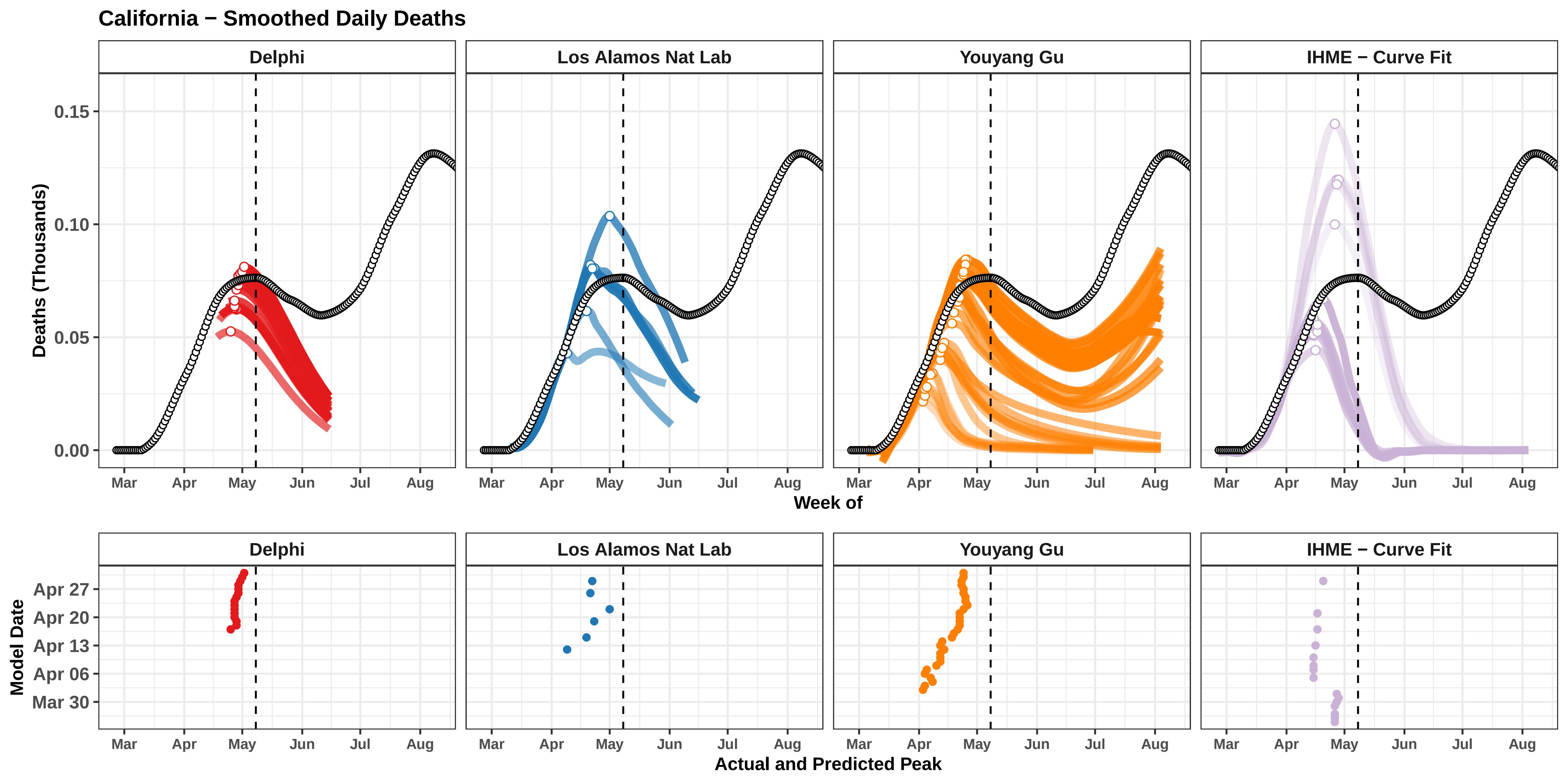
California − Smoothed Daily Deaths
Delphi
Los Alamos Nat Lab
Youyang Gu
IHME − Curve Fit
Deaths (Thousands)
0.15
0.10
0.05
0.00
Mar
Apr
May
Jun
Jul
Aug
Week of
Model Date
Apr 27
Apr 20
Apr 13
Apr 06
Mar 30
Actual and Predicted Peak

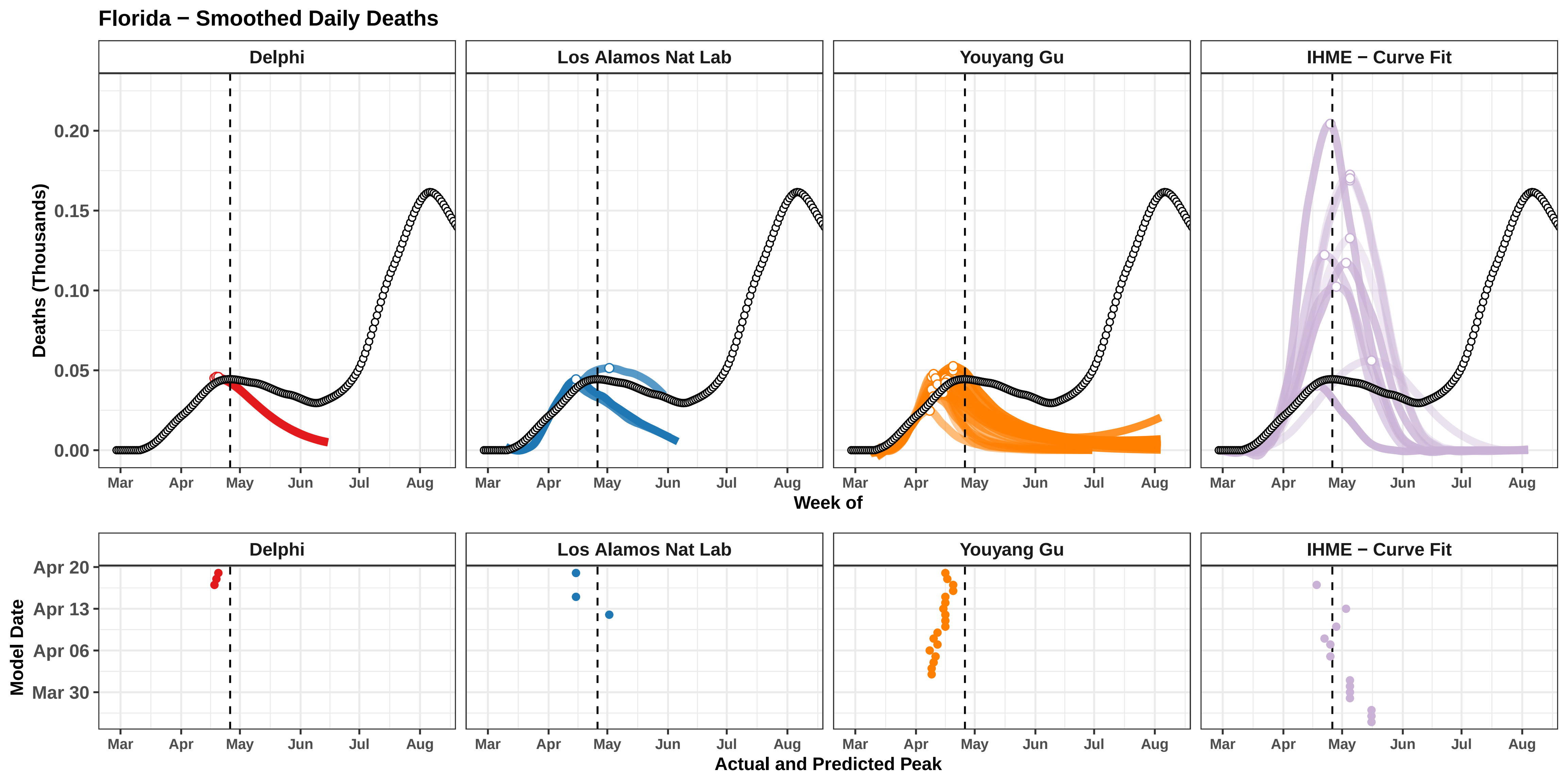

Florida − Smoothed Daily Deaths
Delphi
Los Alamos Nat Lab
Youyang Gu
IHME − Curve Fit
Deaths (Thousands)
0.20
0.15
0.10
0.05
0.00
Mar
Apr
May
Jun
Jul
Aug
Week of
Model Date
Apr 20
Apr 13
Apr 06
Mar 30
Actual and Predicted Peak

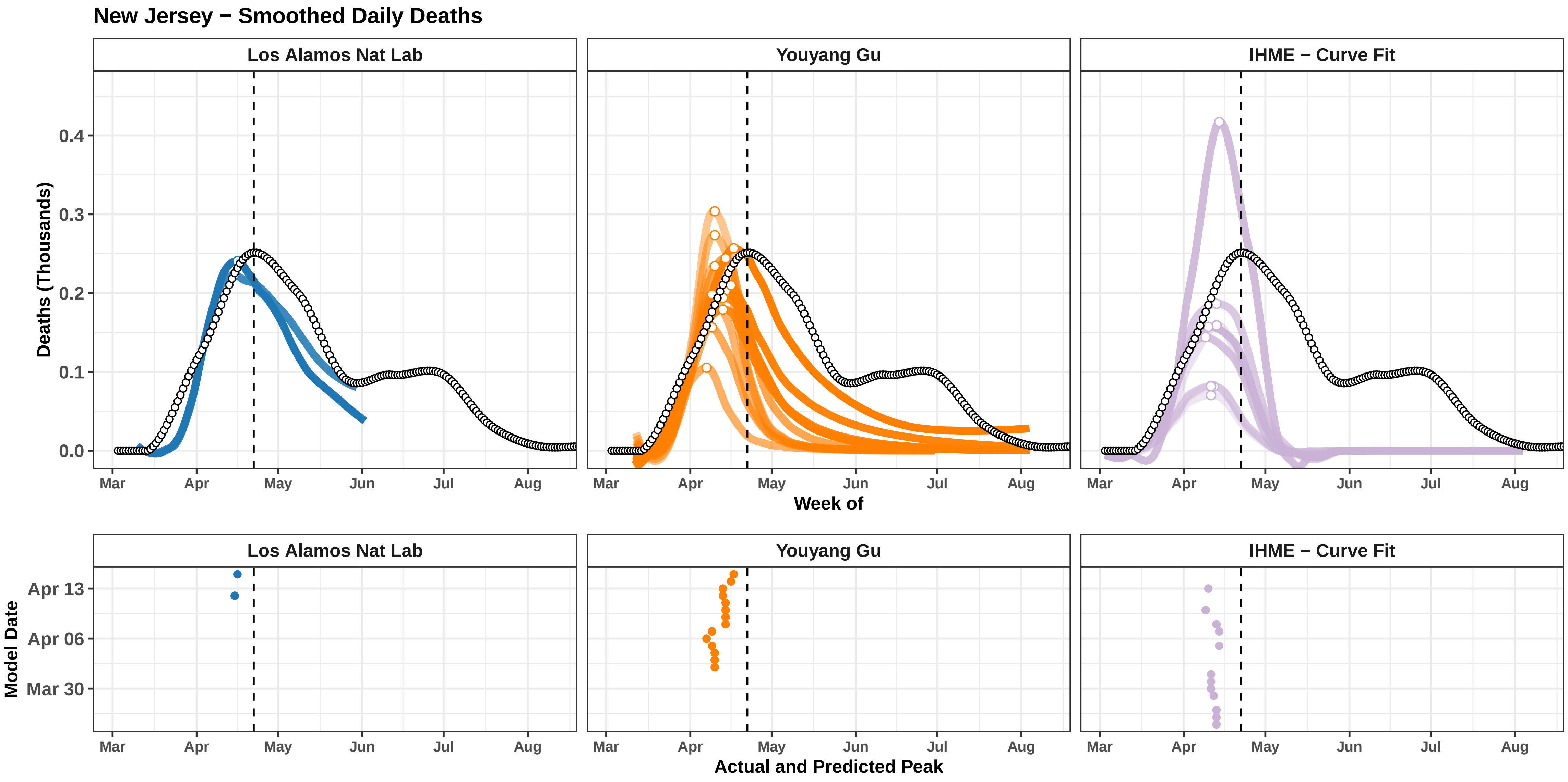
New Jersey − Smoothed Daily Deaths
Los Alamos Nat Lab
Youyang Gu
IHME − Curve Fit
Deaths (Thousands)
0.0
0.1
0.2
0.3
0.4
Mar
Apr
May
Jun
Jul
Aug
Week of
Model Date
Apr 13
Apr 06
Mar 30
Actual and Predicted Peak

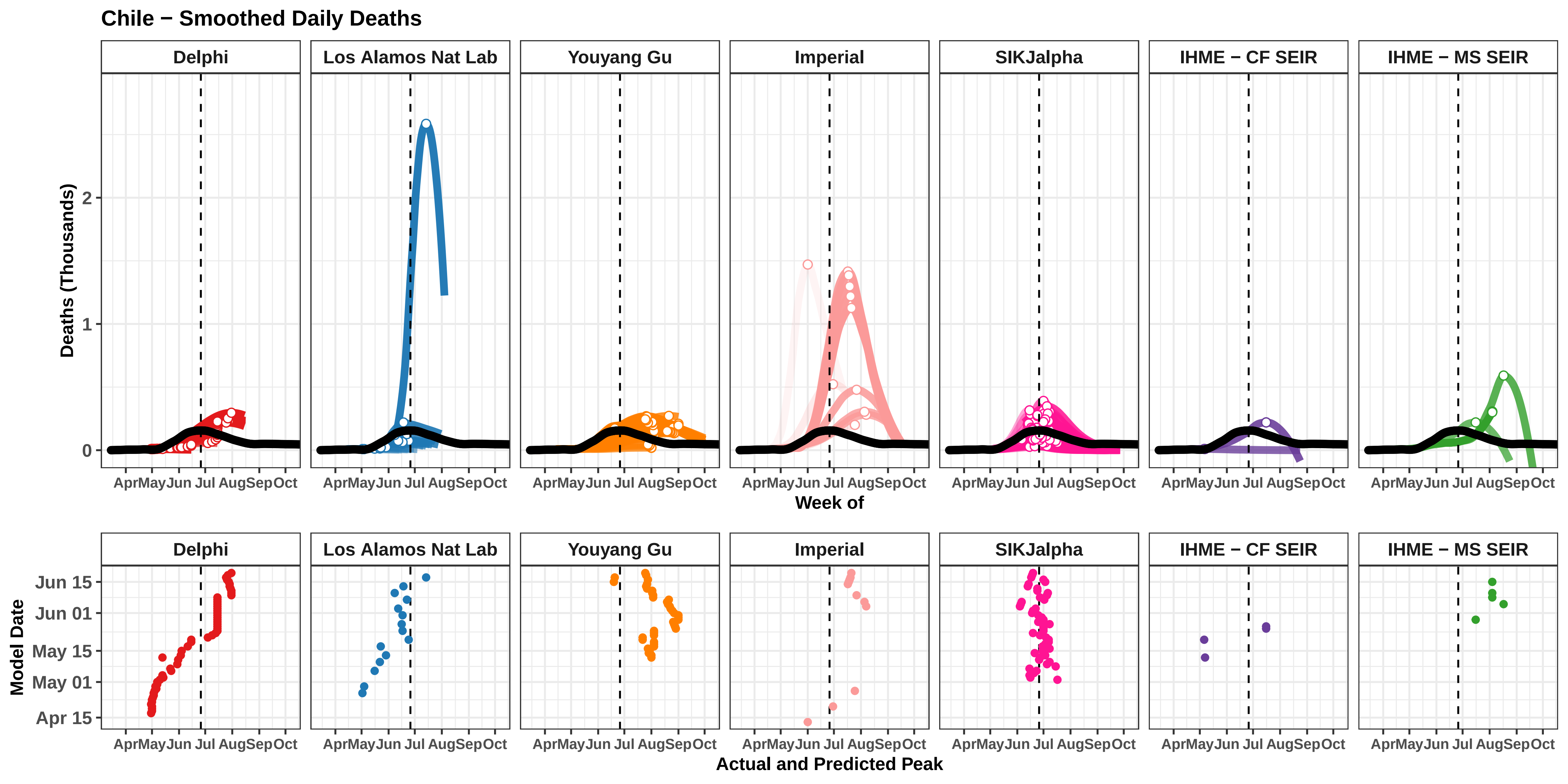
Chile − Smoothed Daily Deaths
Delphi
Los Alamos Nat Lab
Youyang Gu
Imperial
SIKJalpha
IHME − CF SEIR
IHME − MS SEIR
Deaths (Thousands)
0
1
2
Apr May Jun Jul Aug Sep Oct
Week of
Model Date
Jun 15
Jun 01
May 15
May 01
Apr 15
Actual and Predicted Peak

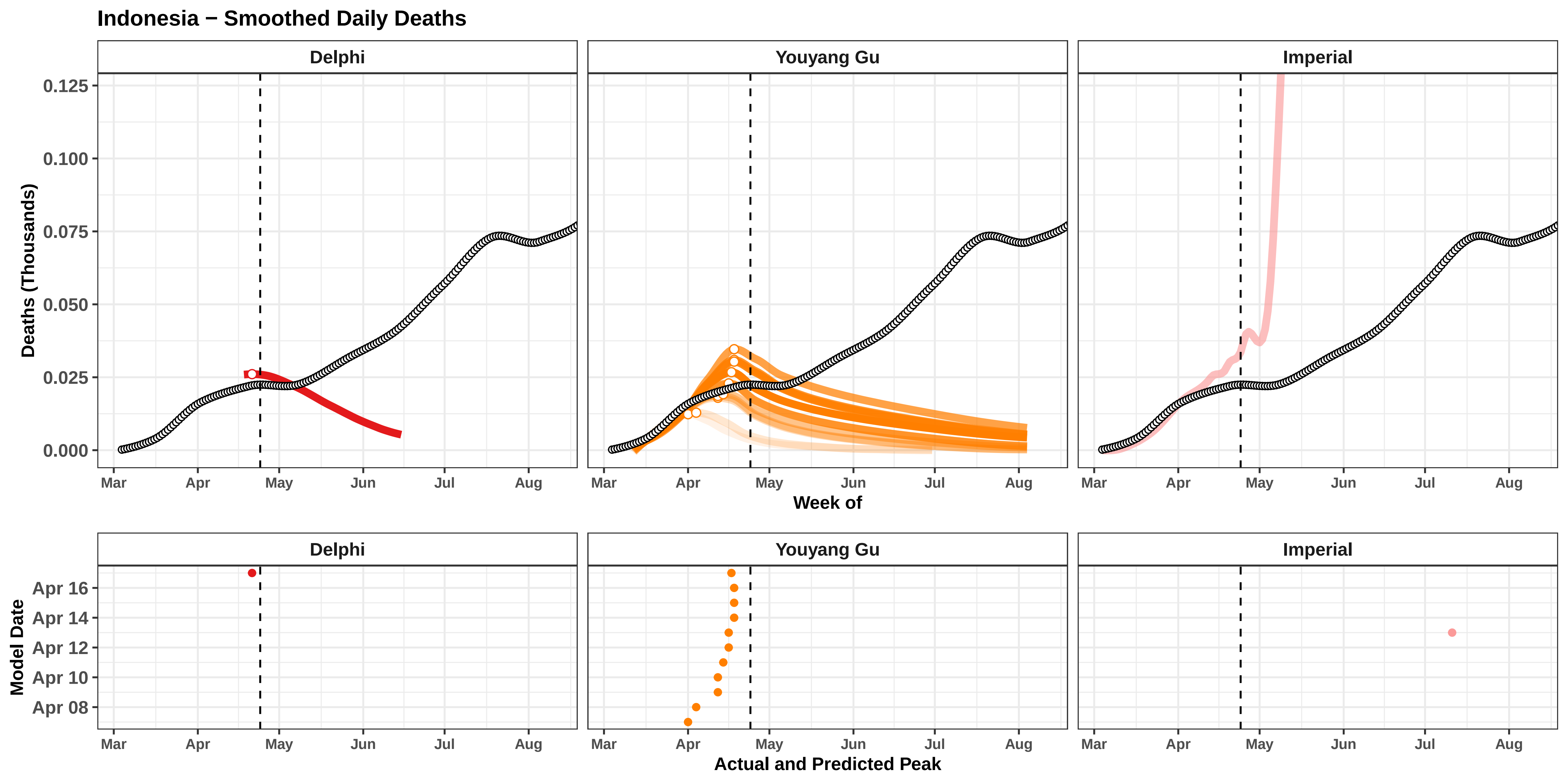

Indonesia − Smoothed Daily Deaths
Delphi
Youyang Gu
Imperial
Deaths (Thousands)
0.125
0.100
0.075
0.050
0.025
0.000
Mar
Apr
May
Jun
Jul
Aug
Week of
Model Date
Apr 16
Apr 14
Apr 12
Apr 10
Apr 08
Actual and Predicted Peak

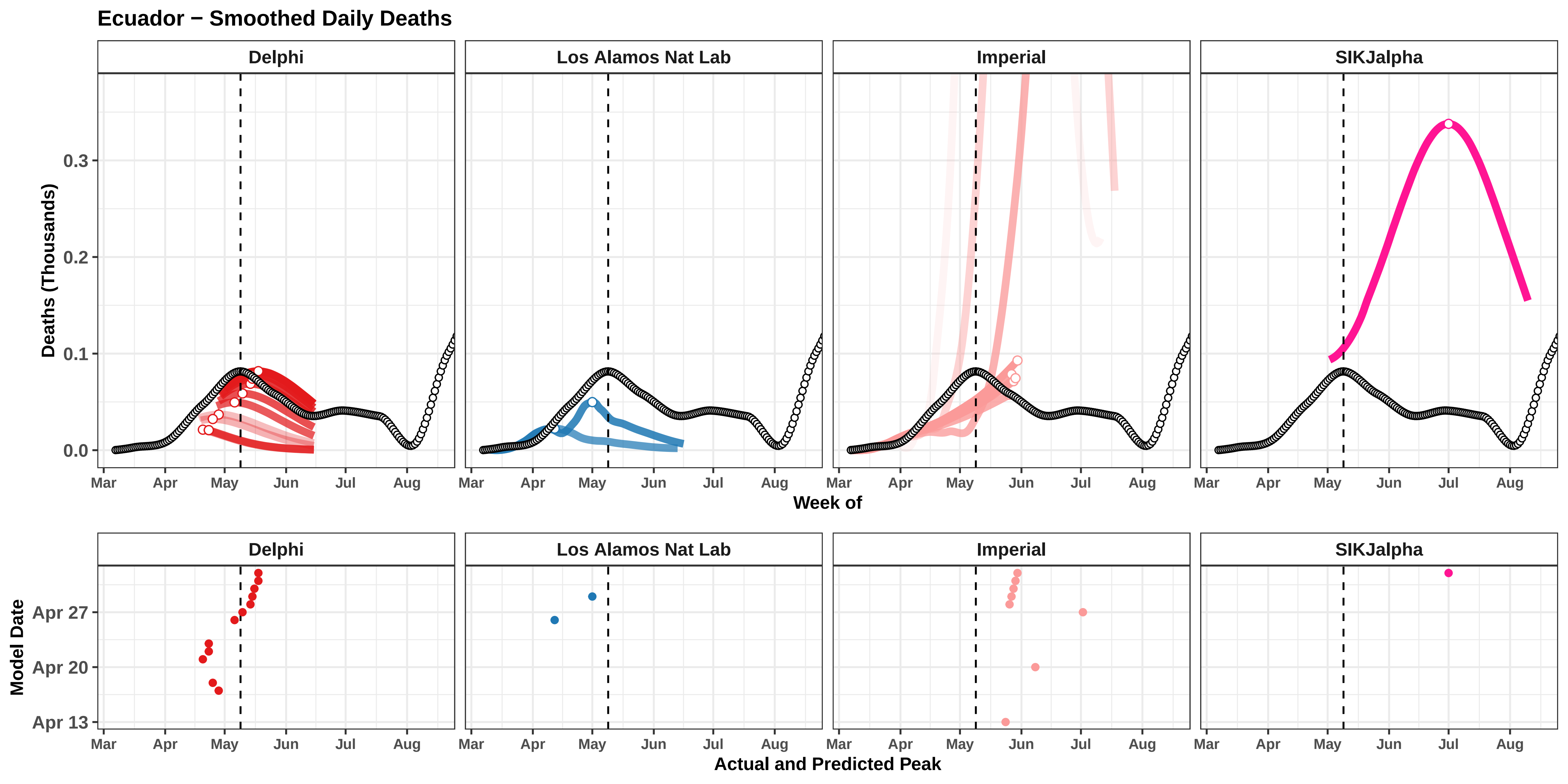
Ecuador − Smoothed Daily Deaths
Delphi
Los Alamos Nat Lab
Imperial
SIKJalpha
Deaths (Thousands)
0.0
0.1
0.2
0.3
Mar
Apr
May
Jun
Jul
Aug
Week of
Model Date
Apr 13
Apr 20
Apr 27
Actual and Predicted Peak

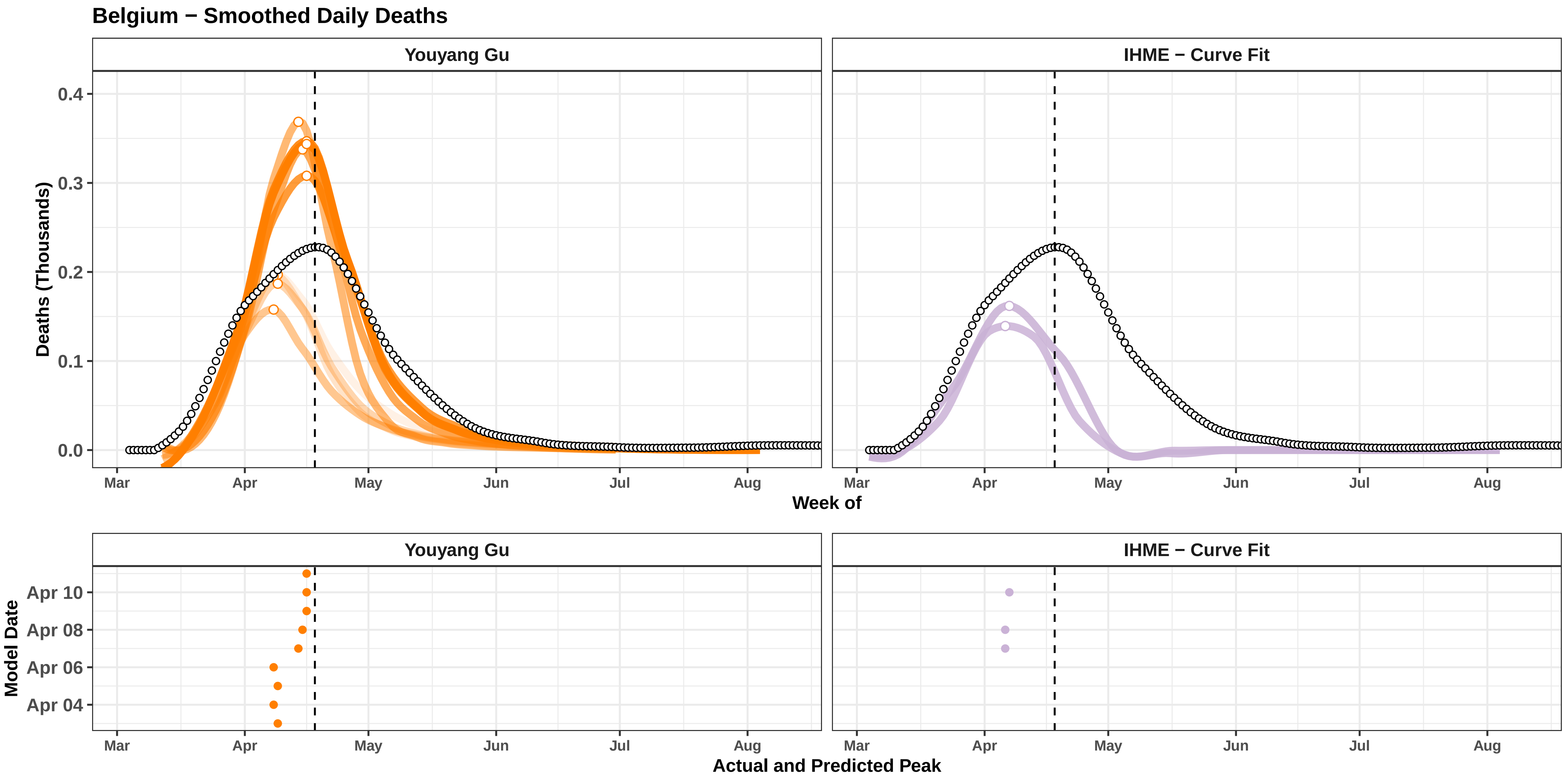

Belgium − Smoothed Daily Deaths
Youyang Gu
IHME − Curve Fit
Deaths (Thousands)
0.0
0.1
0.2
0.3
0.4
Mar
Apr
May
Jun
Jul
Aug
Week of
Model Date
Apr 04
Apr 06
Apr 08
Apr 10
Actual and Predicted Peak

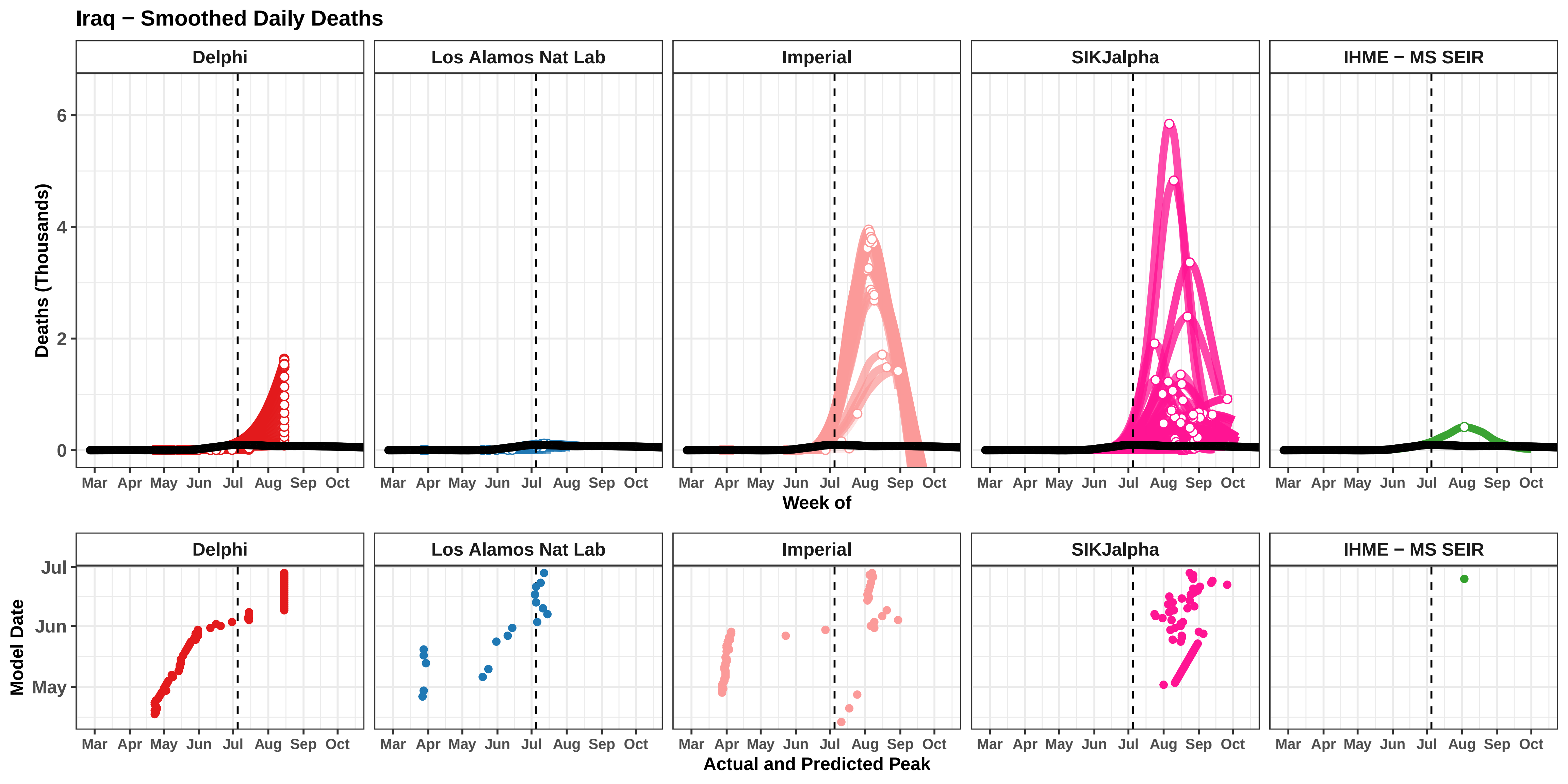
Iraq − Smoothed Daily Deaths
Delphi
Los Alamos Nat Lab
Imperial
SIKJalpha
IHME − MS SEIR
Deaths (Thousands)
0
2
4
6
Mar Apr May Jun Jul Aug Sep Oct
Week of
Model Date
Jul
Jun
May
Actual and Predicted Peak

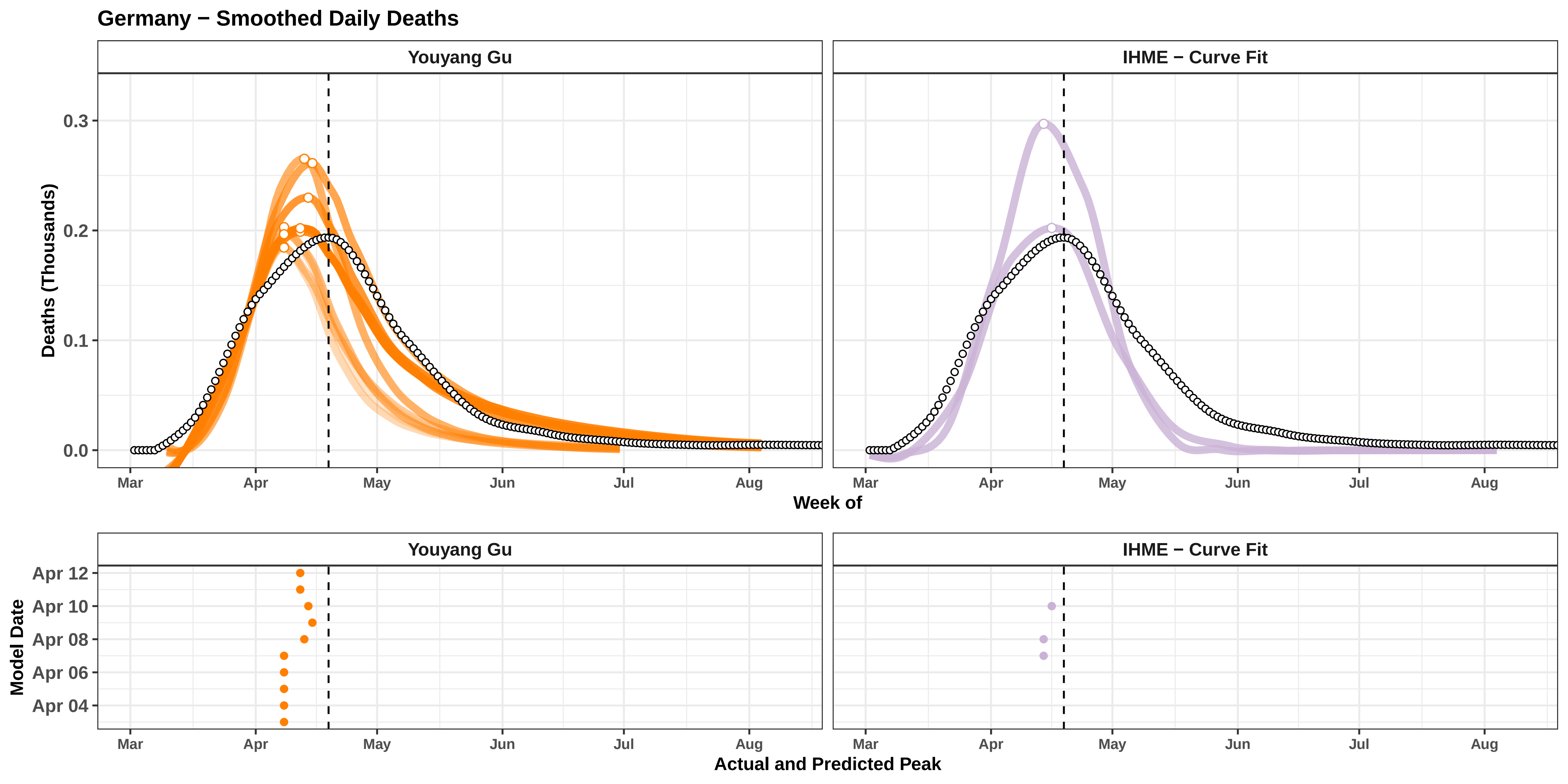

Germany − Smoothed Daily Deaths
Youyang Gu
IHME − Curve Fit
Deaths (Thousands)
0.3
0.2
0.1
0.0
Mar
Apr
May
Jun
Jul
Aug
Week of
Model Date
Apr 12
Apr 10
Apr 08
Apr 06
Apr 04
Actual and Predicted Peak

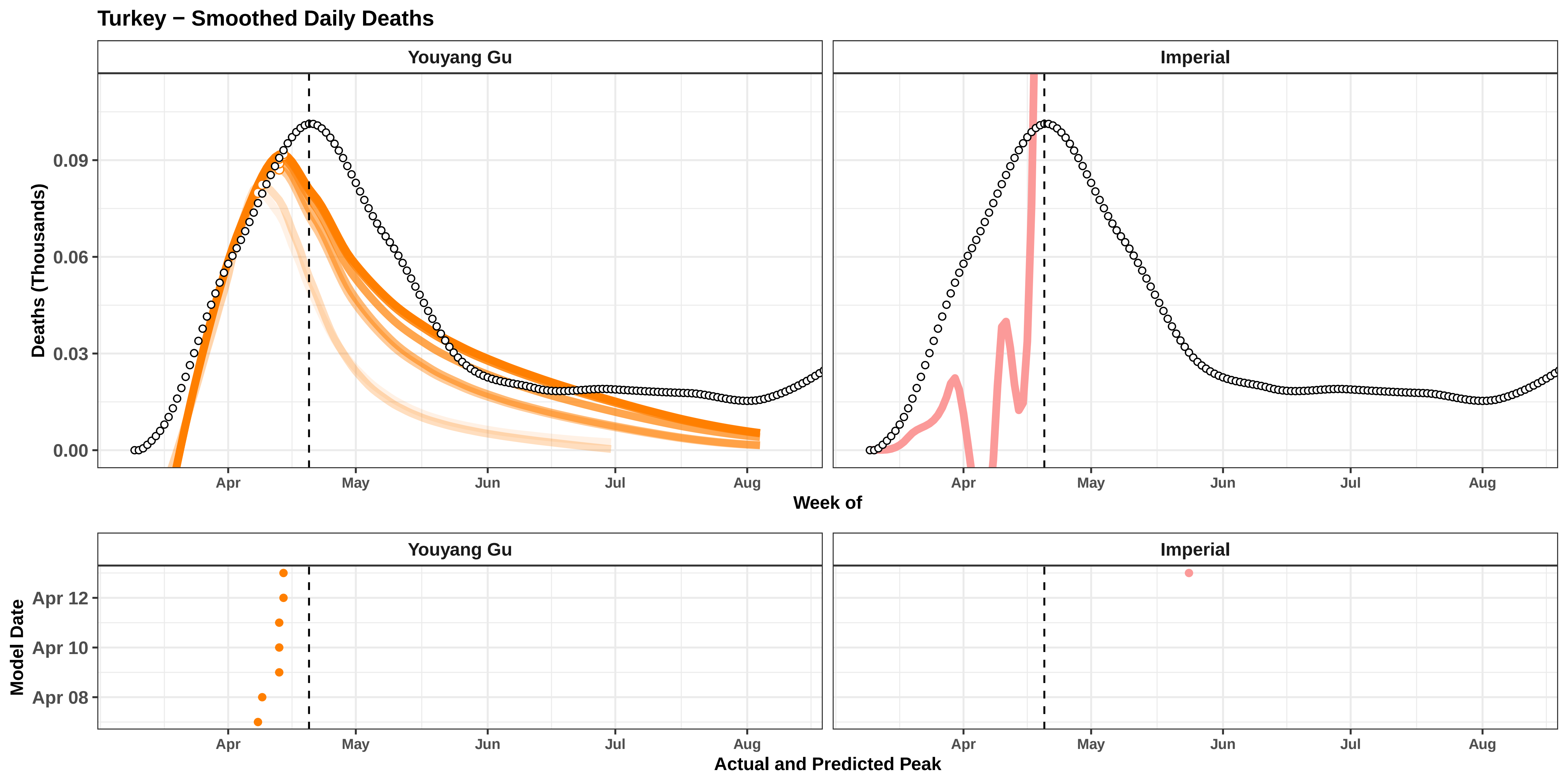

Turkey − Smoothed Daily Deaths
Youyang Gu
Imperial
Deaths (Thousands)
0.09
0.06
0.03
0.00
Apr
May
Jun
Jul
Aug
Week of
Youyang Gu
Imperial
Model Date
Apr 12
Apr 10
Apr 08
Actual and Predicted Peak

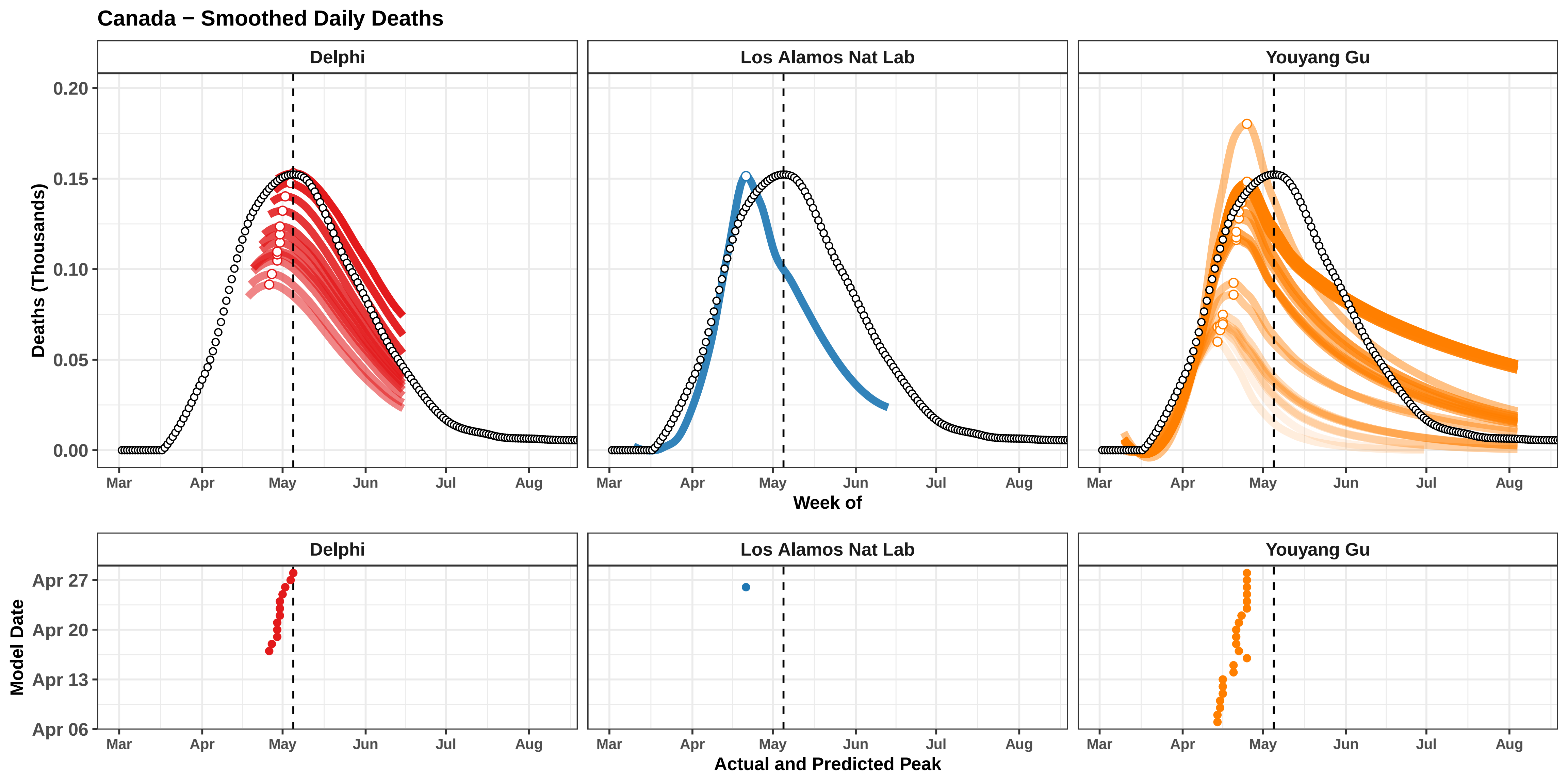

Canada − Smoothed Daily Deaths
Delphi
Los Alamos Nat Lab
Youyang Gu
Deaths (Thousands)
0.20
0.15
0.10
0.05
0.00
Mar
Apr
May
Jun
Jul
Aug
Week of
Model Date
Apr 27
Apr 20
Apr 13
Apr 06
Actual and Predicted Peak

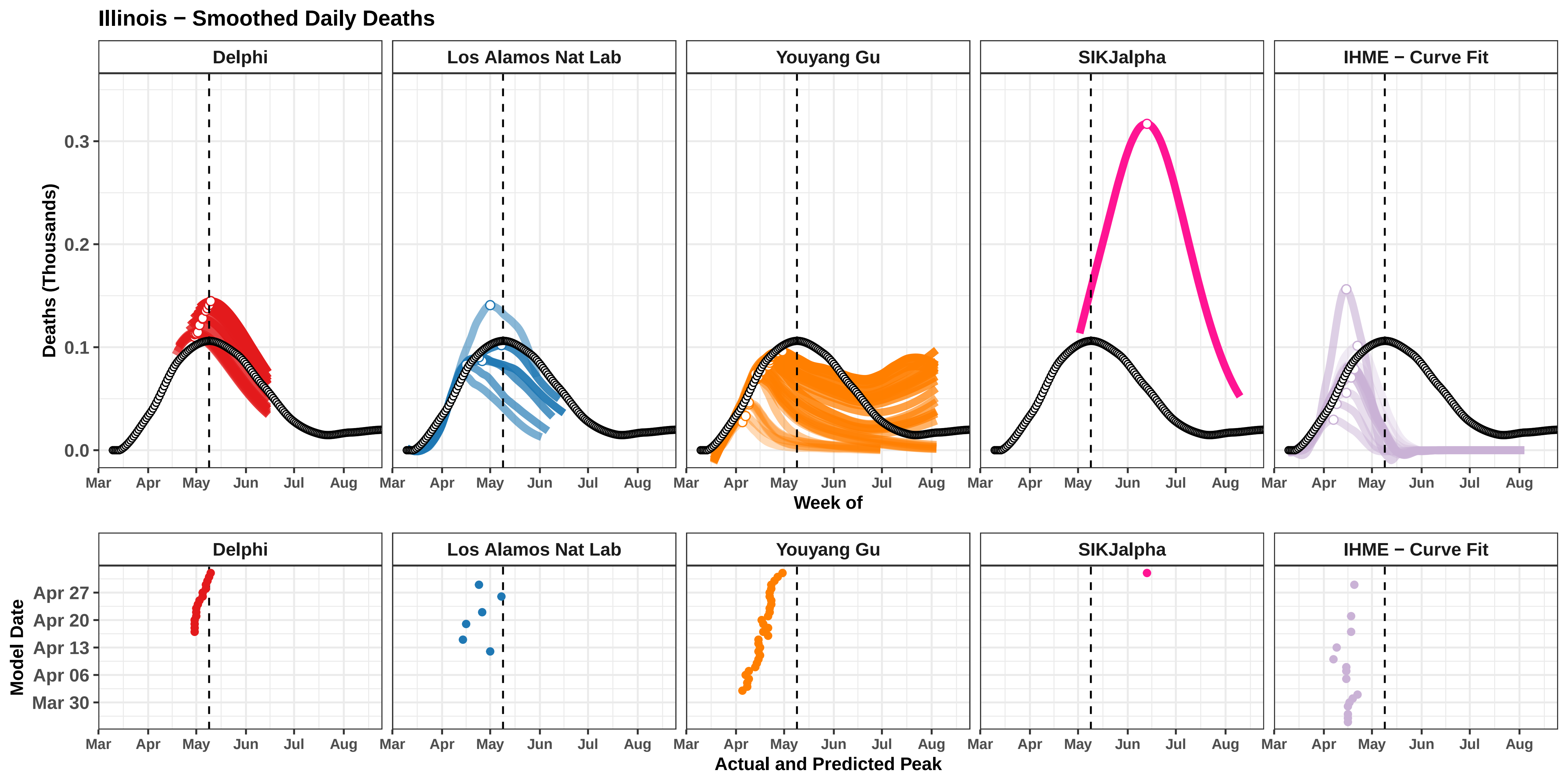

Illinois − Smoothed Daily Deaths
Delphi
Los Alamos Nat Lab
Youyang Gu
SIKJalpha
IHME − Curve Fit
Deaths (Thousands)
0.0
0.1
0.2
0.3
Mar
Apr
May
Jun
Jul
Aug
Week of
Model Date
Apr 27
Apr 20
Apr 13
Apr 06
Mar 30
Actual and Predicted Peak

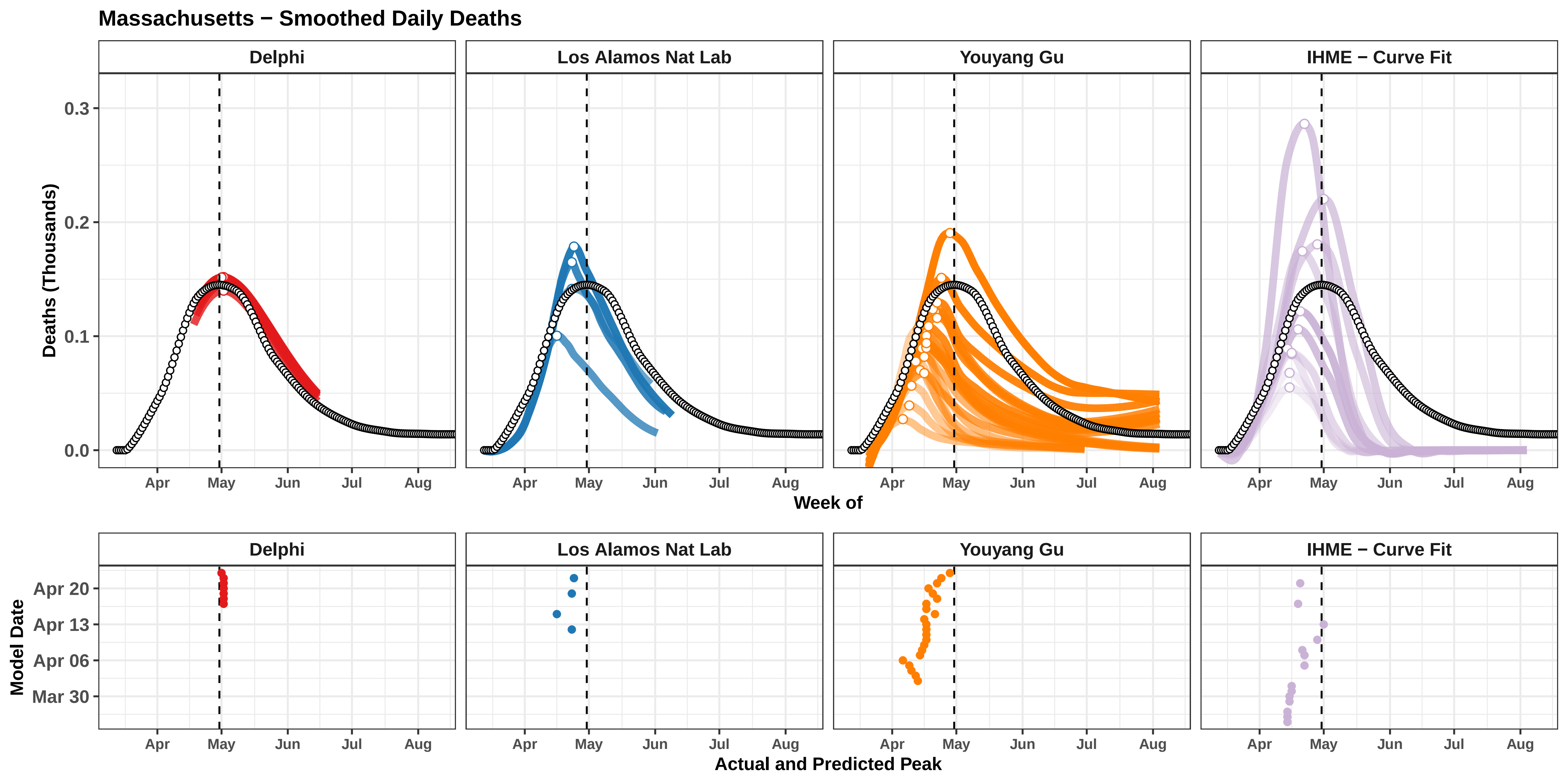
Massachusetts − Smoothed Daily Deaths
Delphi
Los Alamos Nat Lab
Youyang Gu
IHME − Curve Fit
Deaths (Thousands)
0.3
0.2
0.1
0.0
Apr
May
Jun
Jul
Aug
Week of
Model Date
Apr 20
Apr 13
Apr 06
Mar 30
Actual and Predicted Peak

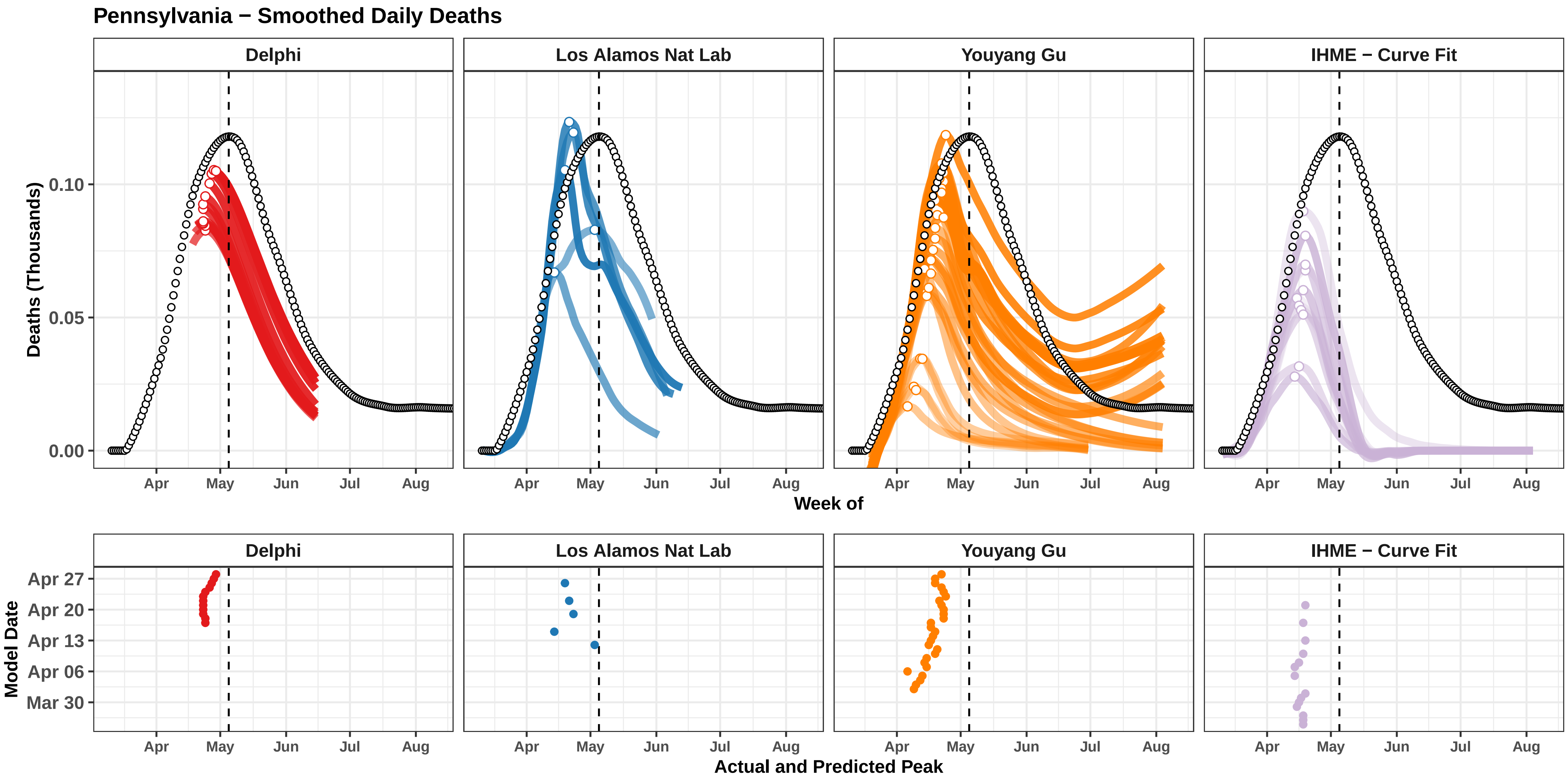

Pennsylvania − Smoothed Daily Deaths
Delphi
Los Alamos Nat Lab
Youyang Gu
IHME − Curve Fit
Deaths (Thousands)
0.10
0.05
0.00
Apr
May
Jun
Jul
Aug
Week of
Model Date
Apr 27
Apr 20
Apr 13
Apr 06
Mar 30
Actual and Predicted Peak

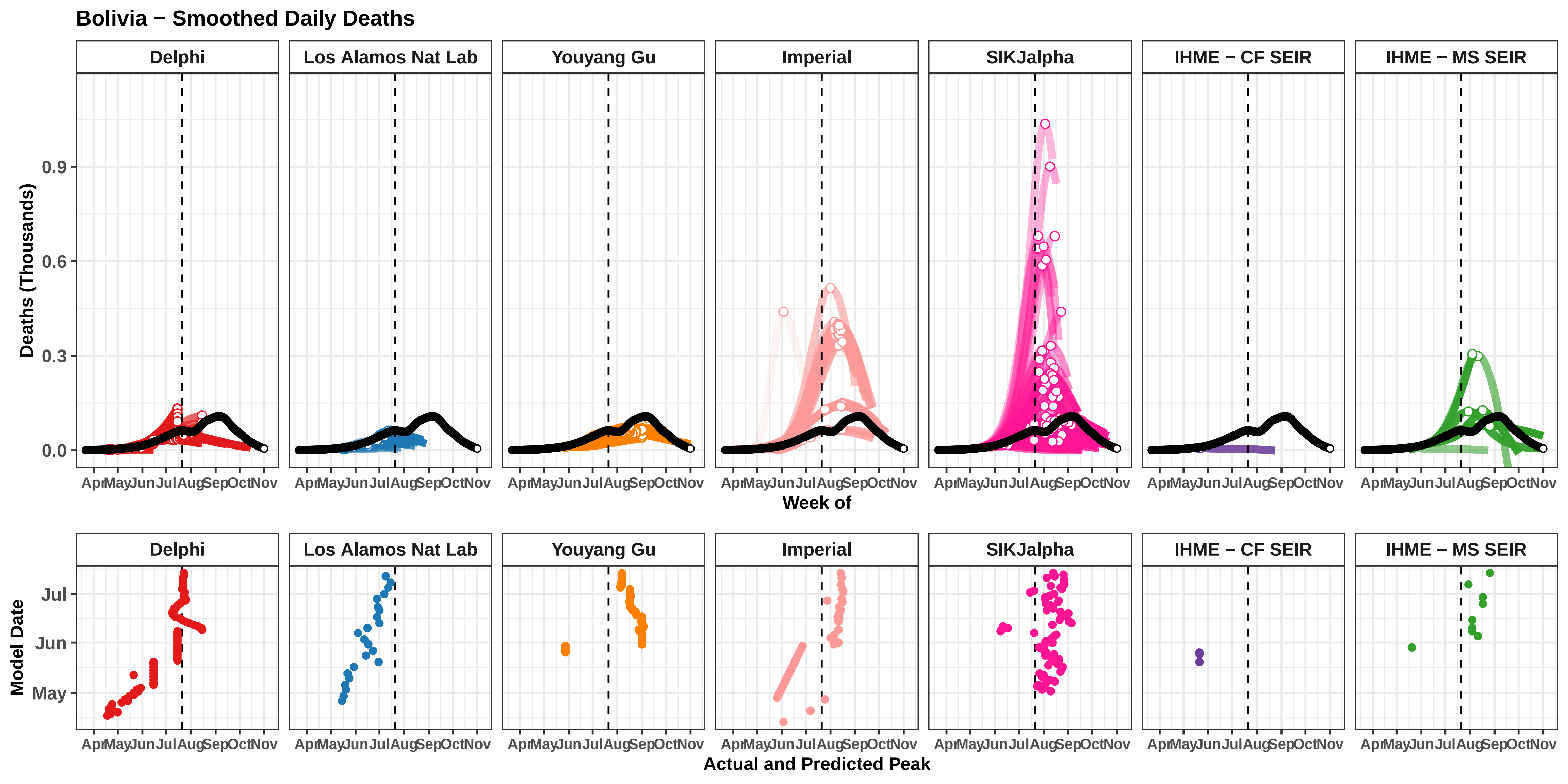
Bolivia − Smoothed Daily Deaths
Delphi
Los Alamos Nat Lab
Youyang Gu
Imperial
SIKJalpha
IHME − CF SEIR
IHME − MS SEIR
Deaths (Thousands)
0.0
0.3
0.6
0.9
Apr May Jun Jul Aug Sep Oct Nov
Week of
Model Date
May
Jun
Jul
Actual and Predicted Peak

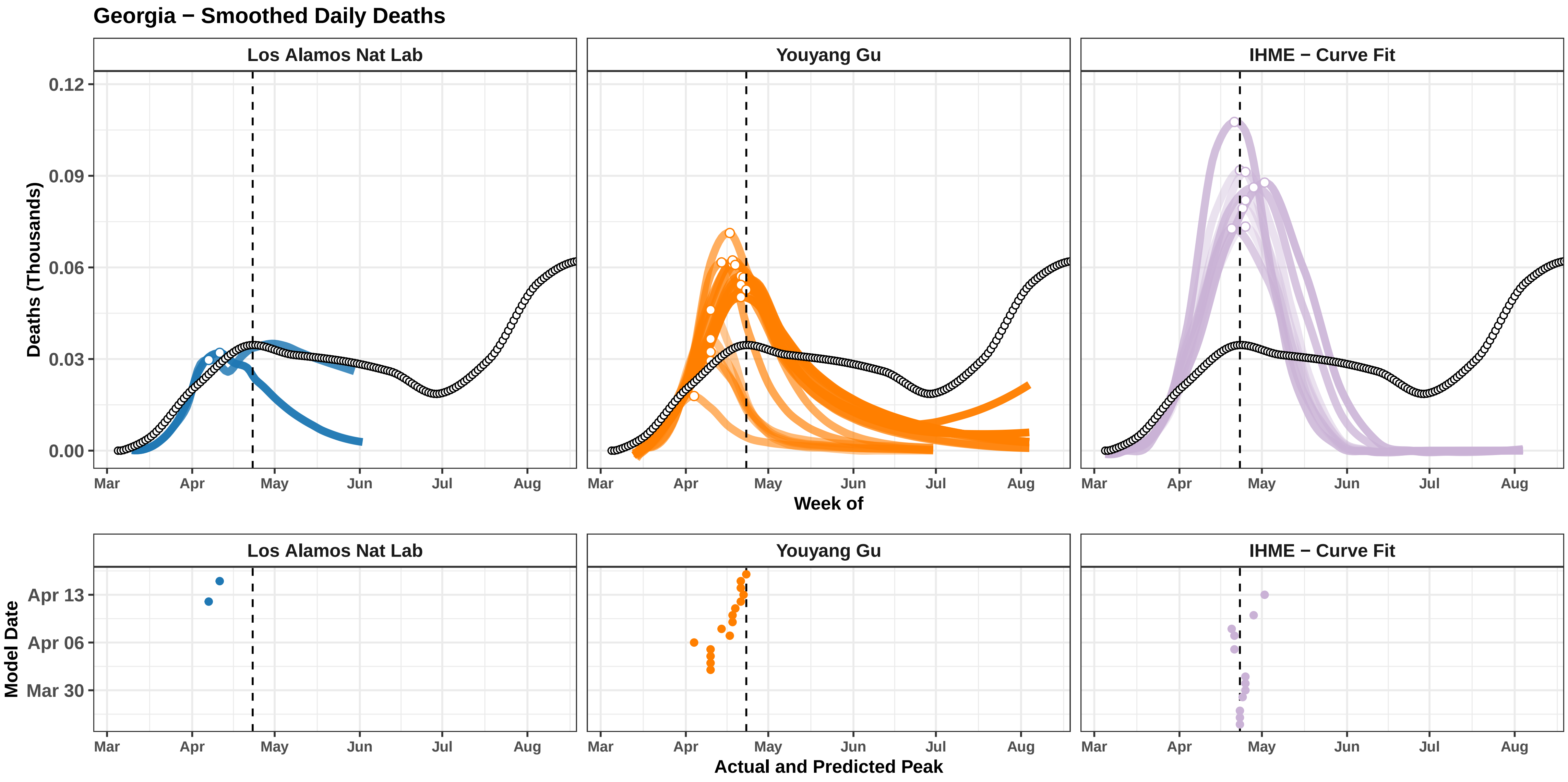

Georgia − Smoothed Daily Deaths
Los Alamos Nat Lab
Youyang Gu
IHME − Curve Fit
Deaths (Thousands)
0.12
0.09
0.06
0.03
0.00
Mar
Apr
May
Jun
Jul
Aug
Week of
Model Date
Apr 13
Apr 06
Mar 30
Actual and Predicted Peak

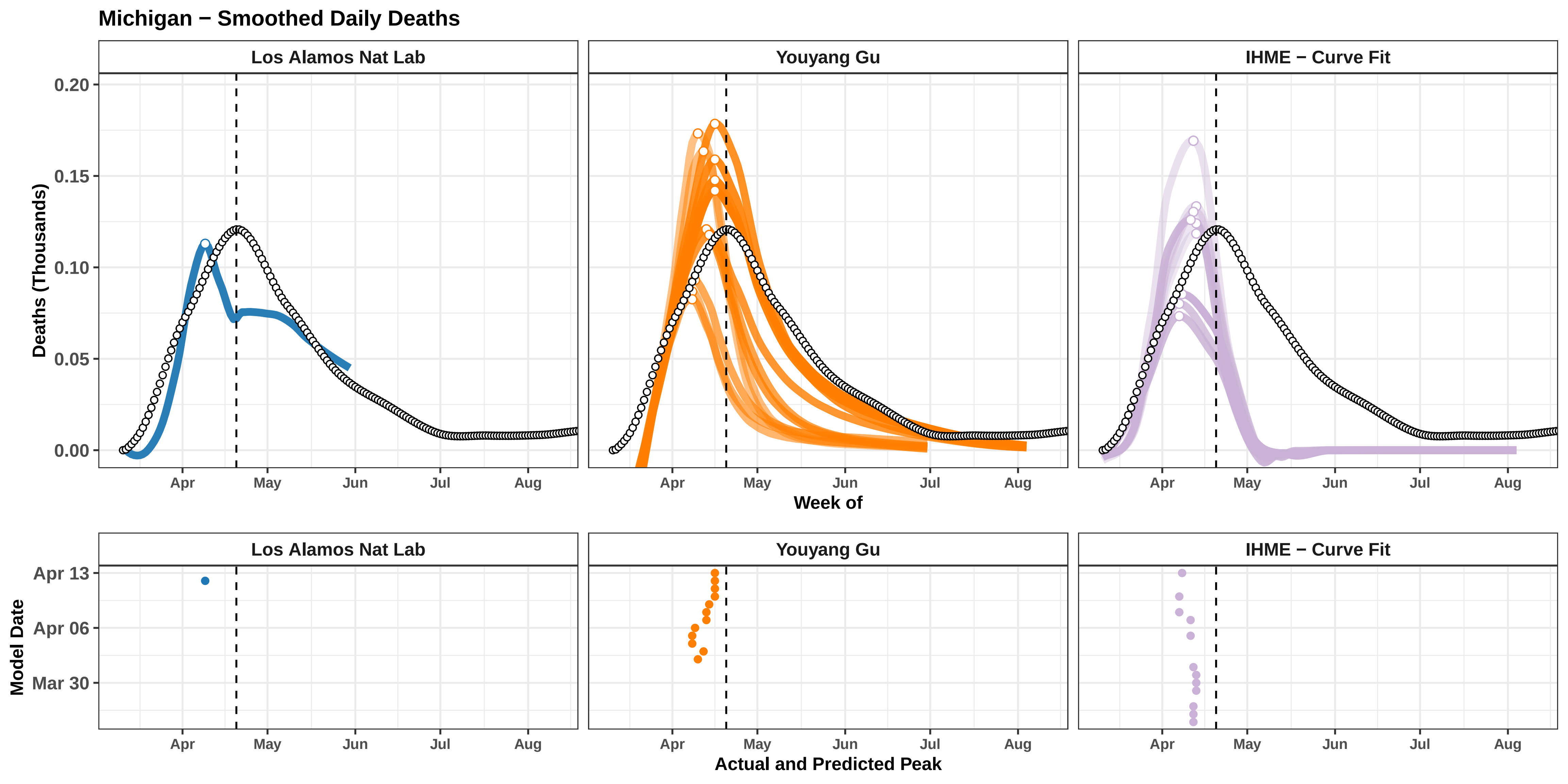

Michigan − Smoothed Daily Deaths
Los Alamos Nat Lab
Youyang Gu
IHME − Curve Fit
Deaths (Thousands)
0.20
0.15
0.10
0.05
0.00
Apr
May
Jun
Jul
Aug
Week of
Model Date
Apr 13
Apr 06
Mar 30
Actual and Predicted Peak

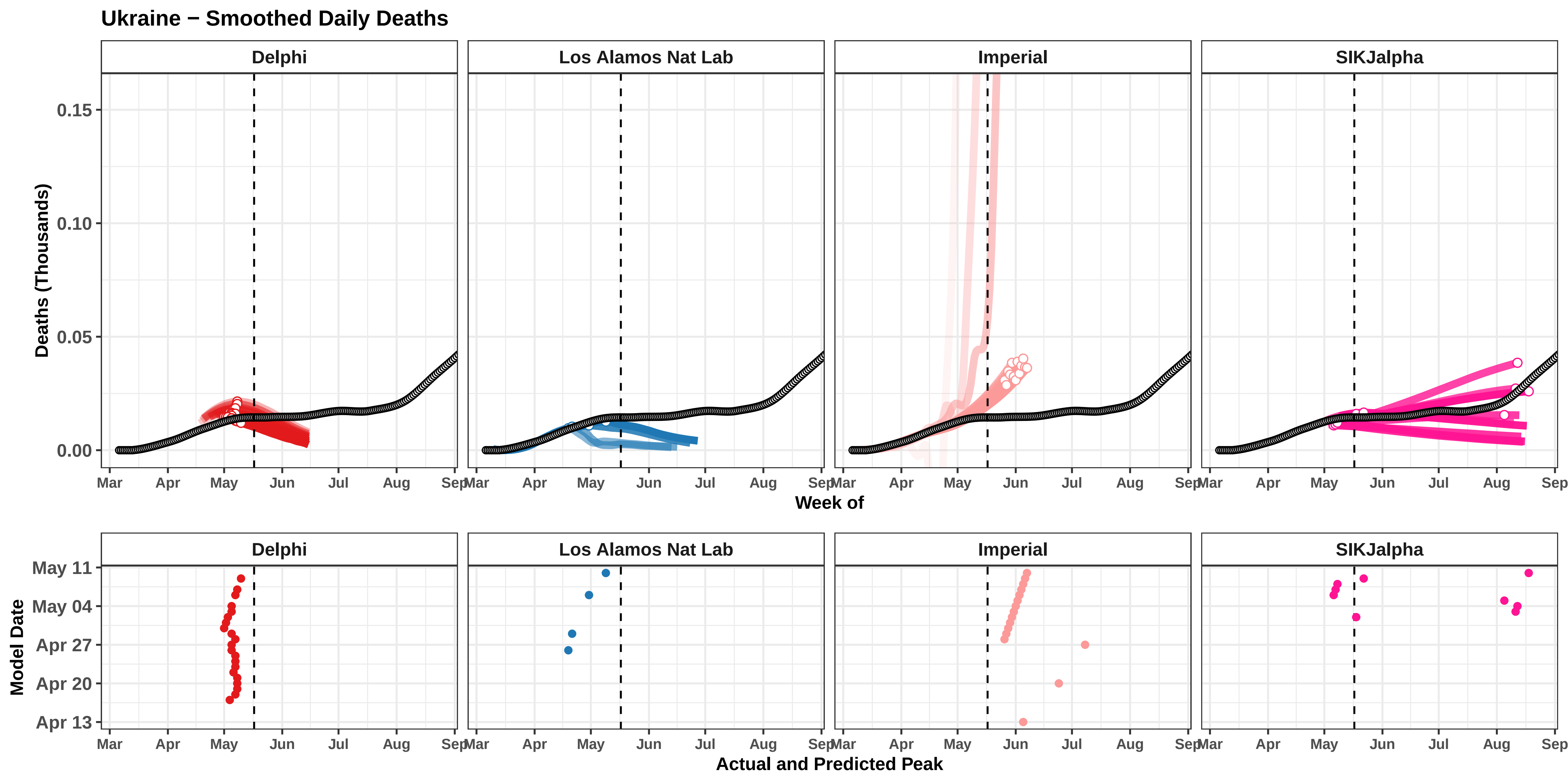

Ukraine − Smoothed Daily Deaths
Delphi
Los Alamos Nat Lab
Imperial
SIKJalpha
Deaths (Thousands)
0.15
0.10
0.05
0.00
Mar
Apr
May
Jun
Jul
Aug
Sep
Week of
Model Date
May 11
May 04
Apr 27
Apr 20
Apr 13
Actual and Predicted Peak

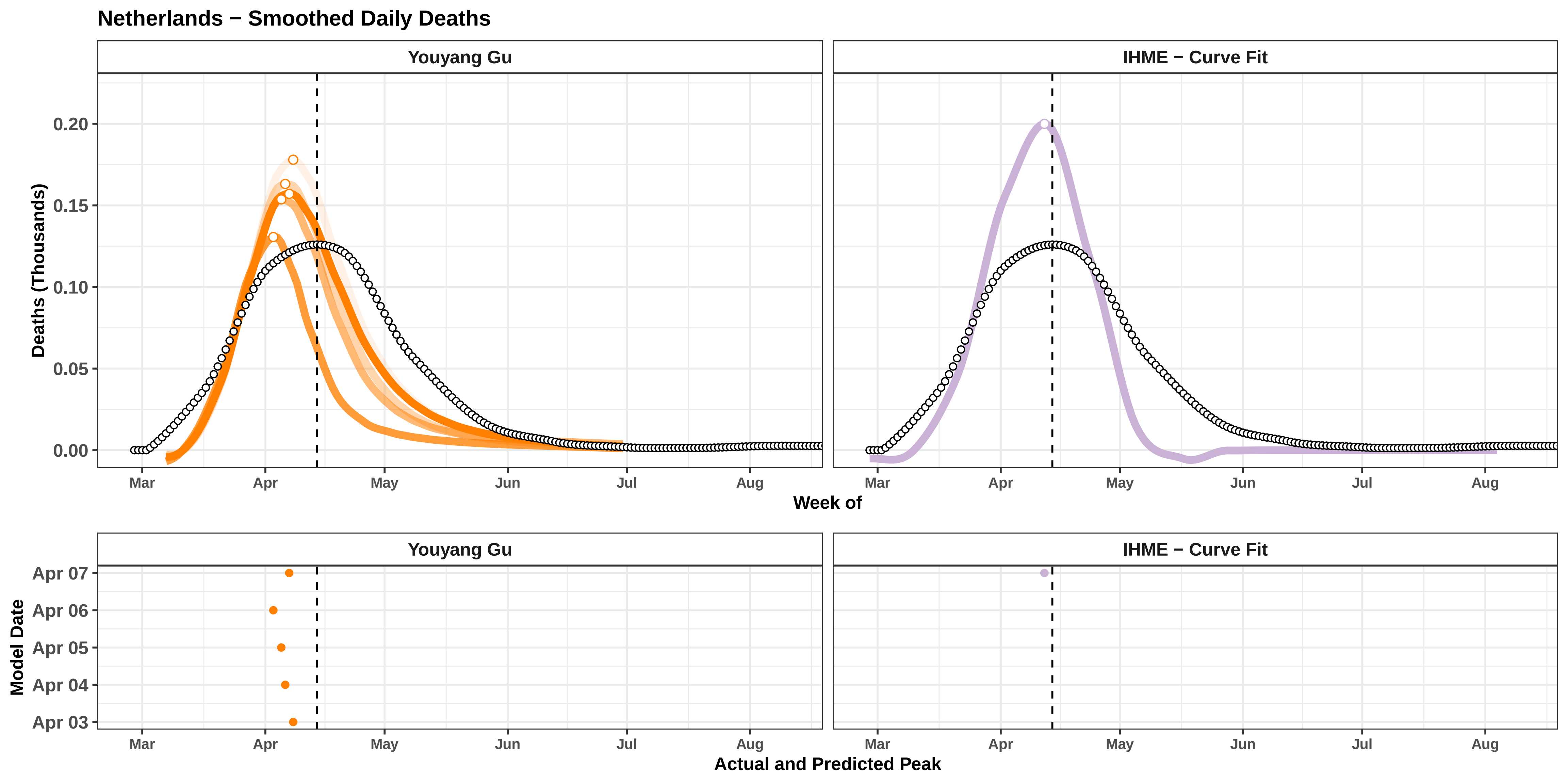

Netherlands − Smoothed Daily Deaths
Youyang Gu
IHME − Curve Fit
Deaths (Thousands)
0.20
0.15
0.10
0.05
0.00
Mar
Apr
May
Jun
Jul
Aug
Week of
Model Date
Apr 07
Apr 06
Apr 05
Apr 04
Apr 03
Actual and Predicted Peak

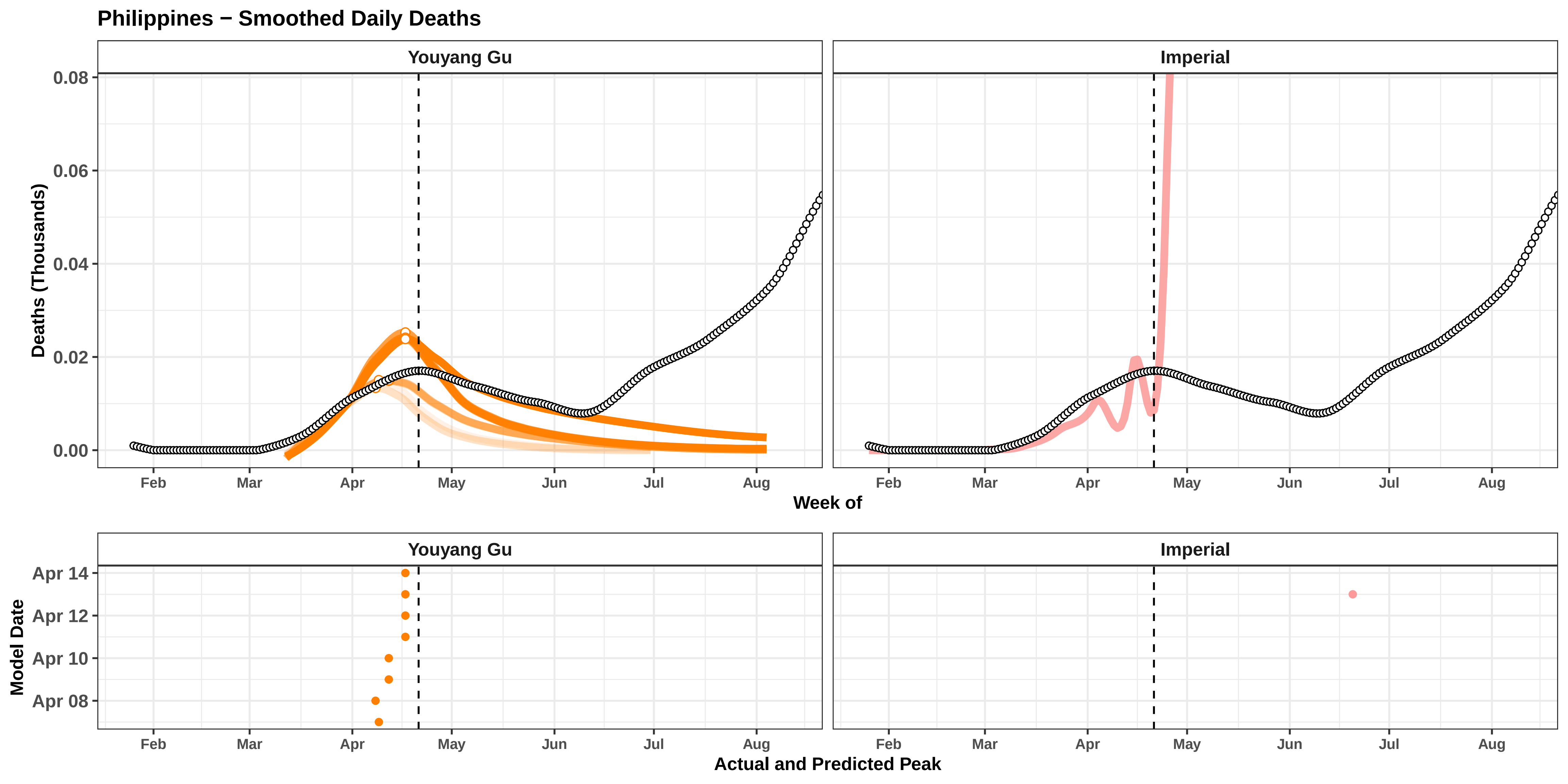

Philippines − Smoothed Daily Deaths
Youyang Gu
Imperial
Deaths (Thousands)
0.08
0.06
0.04
0.02
0.00
Feb
Mar
Apr
May
Jun
Jul
Aug
Week of
Model Date
Apr 14
Apr 12
Apr 10
Apr 08
Actual and Predicted Peak

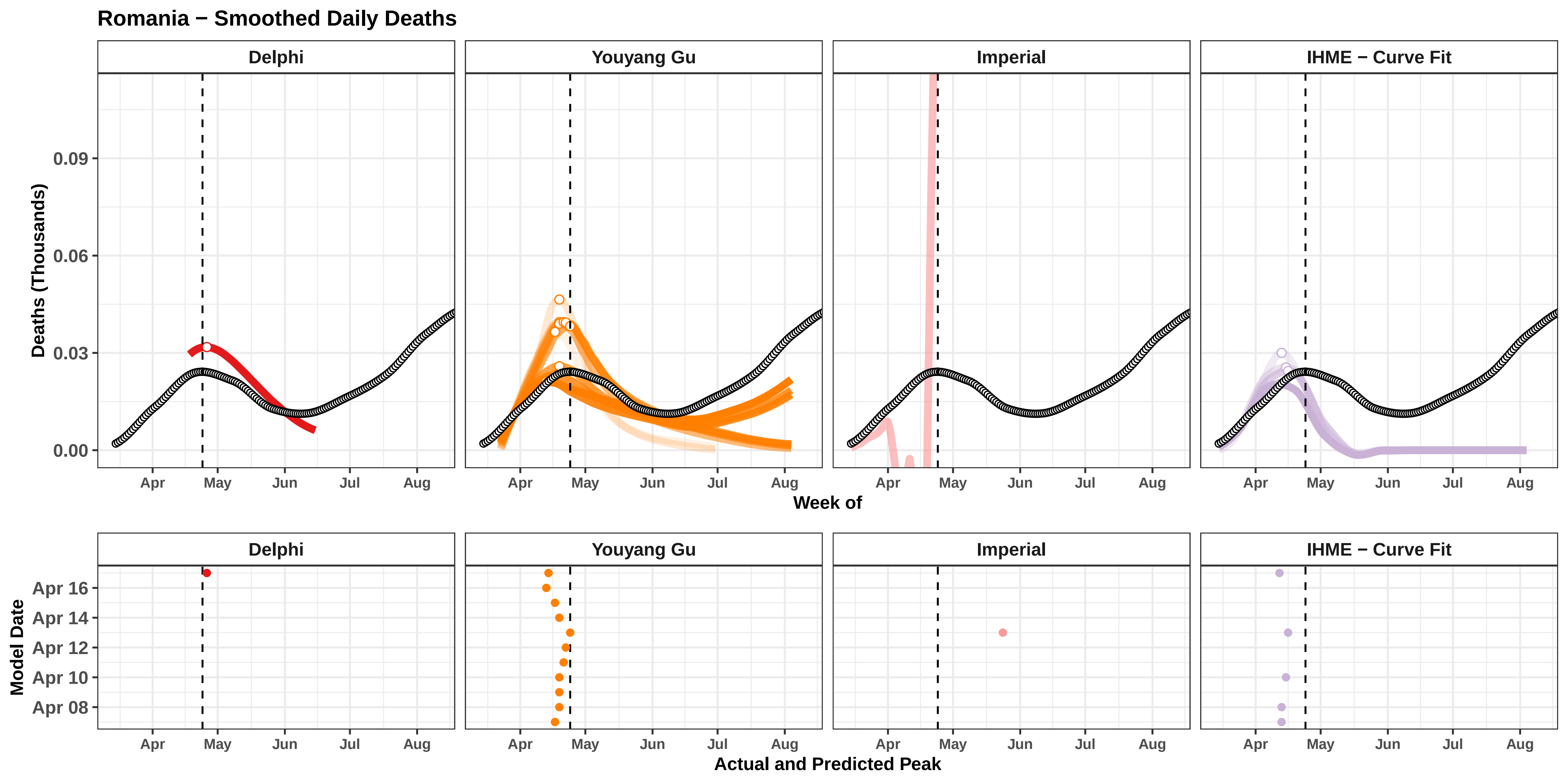

Romania − Smoothed Daily Deaths
Delphi
Youyang Gu
Imperial
IHME − Curve Fit
Deaths (Thousands)
0.09
0.06
0.03
0.00
Apr
May
Jun
Jul
Aug
Week of
Model Date
Apr 16
Apr 14
Apr 12
Apr 10
Apr 08
Actual and Predicted Peak

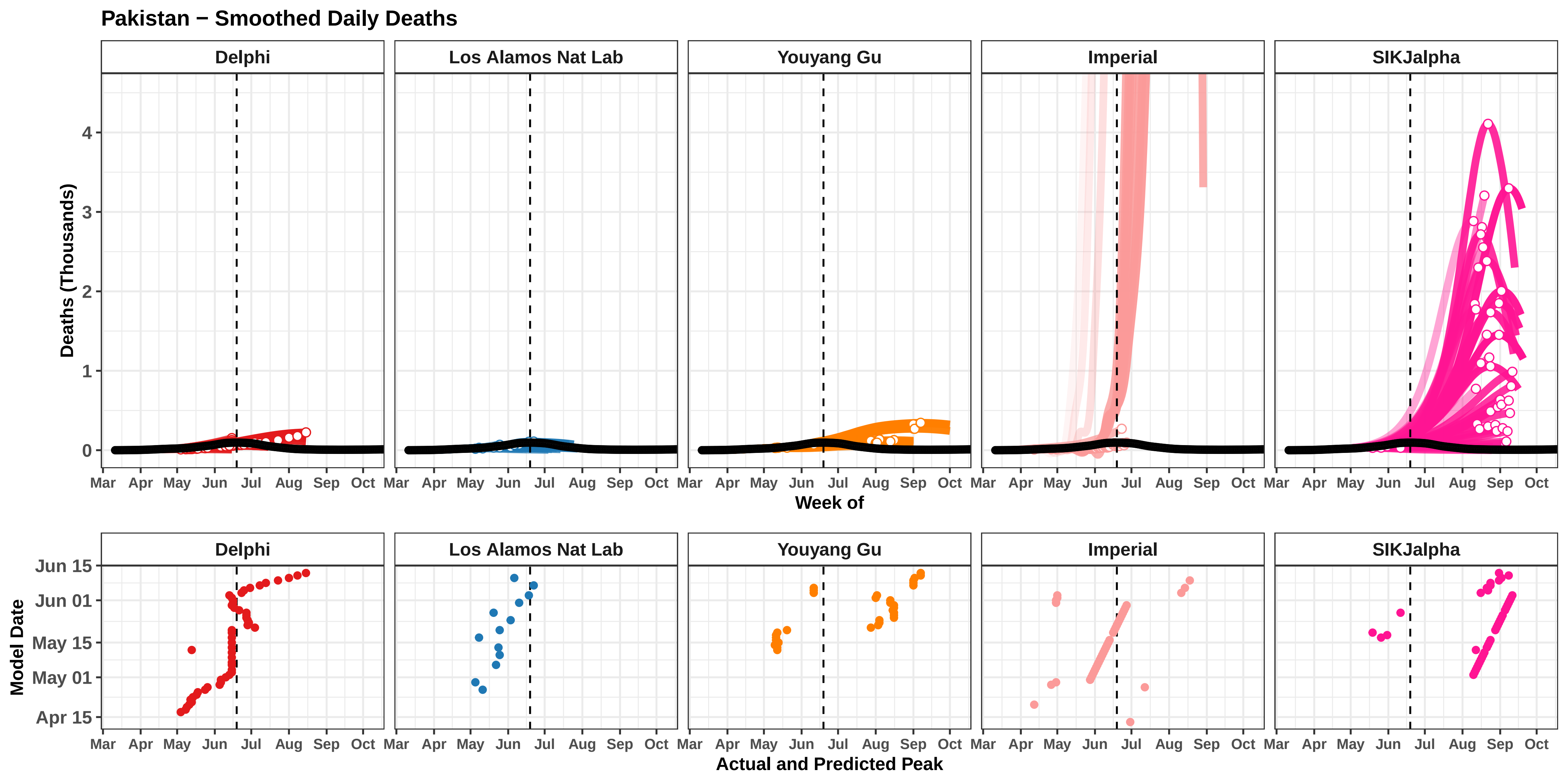

Pakistan − Smoothed Daily Deaths
Delphi
Los Alamos Nat Lab
Youyang Gu
Imperial
SIKJalpha
Deaths (Thousands)
0
1
2
3
4
Mar
Apr
May
Jun
Jul
Aug
Sep
Oct
Week of
Model Date
Jun 15
Jun 01
May 15
May 01
Apr 15
Actual and Predicted Peak

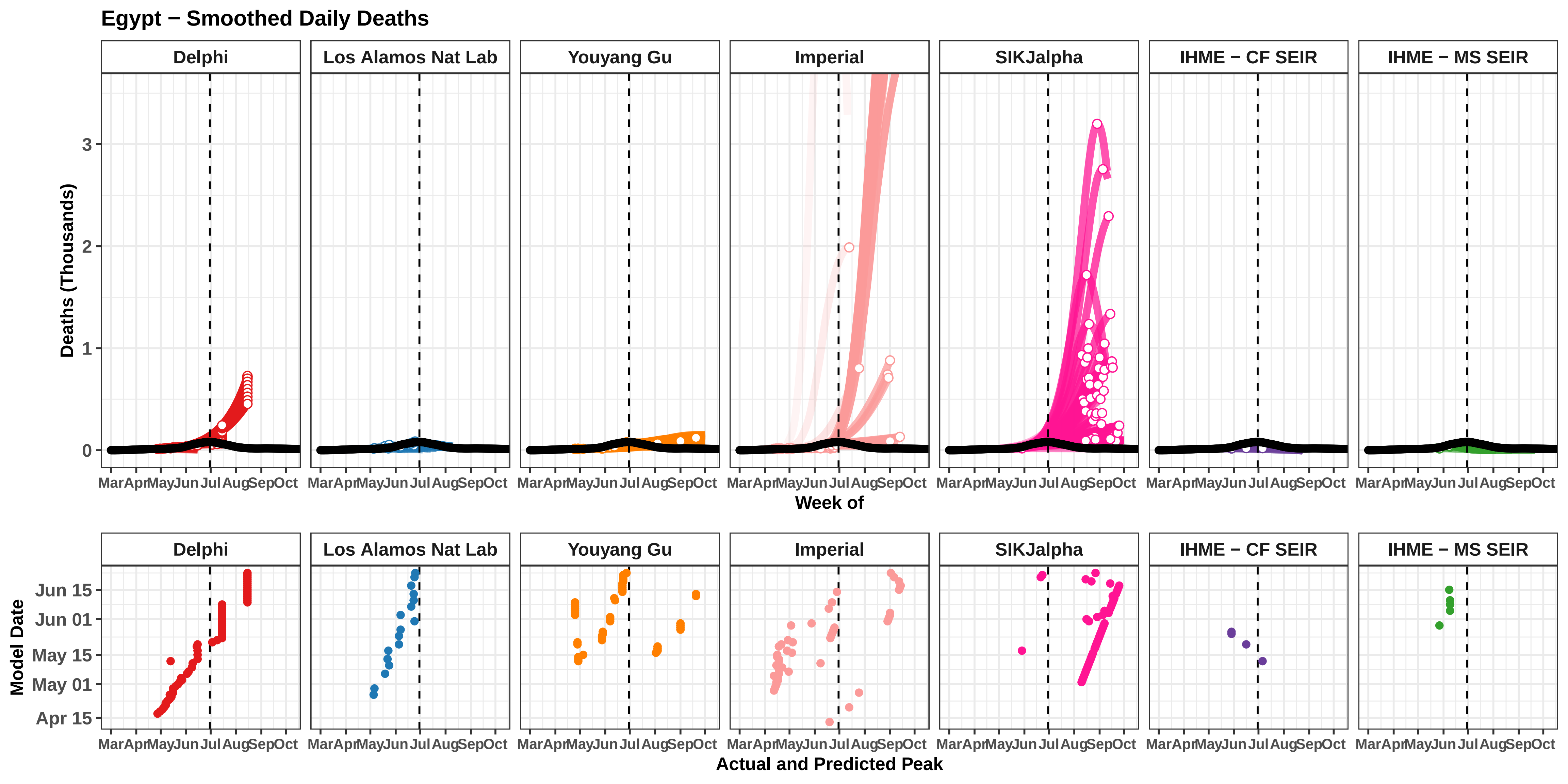

Egypt − Smoothed Daily Deaths
Delphi
Los Alamos Nat Lab
Youyang Gu
Imperial
SIKJalpha
IHME − CF SEIR
IHME − MS SEIR
Deaths (Thousands)
0
1
2
3
Mar Apr May Jun Jul Aug Sep Oct
Week of
Model Date
Jun 15
Jun 01
May 15
May 01
Apr 15
Actual and Predicted Peak

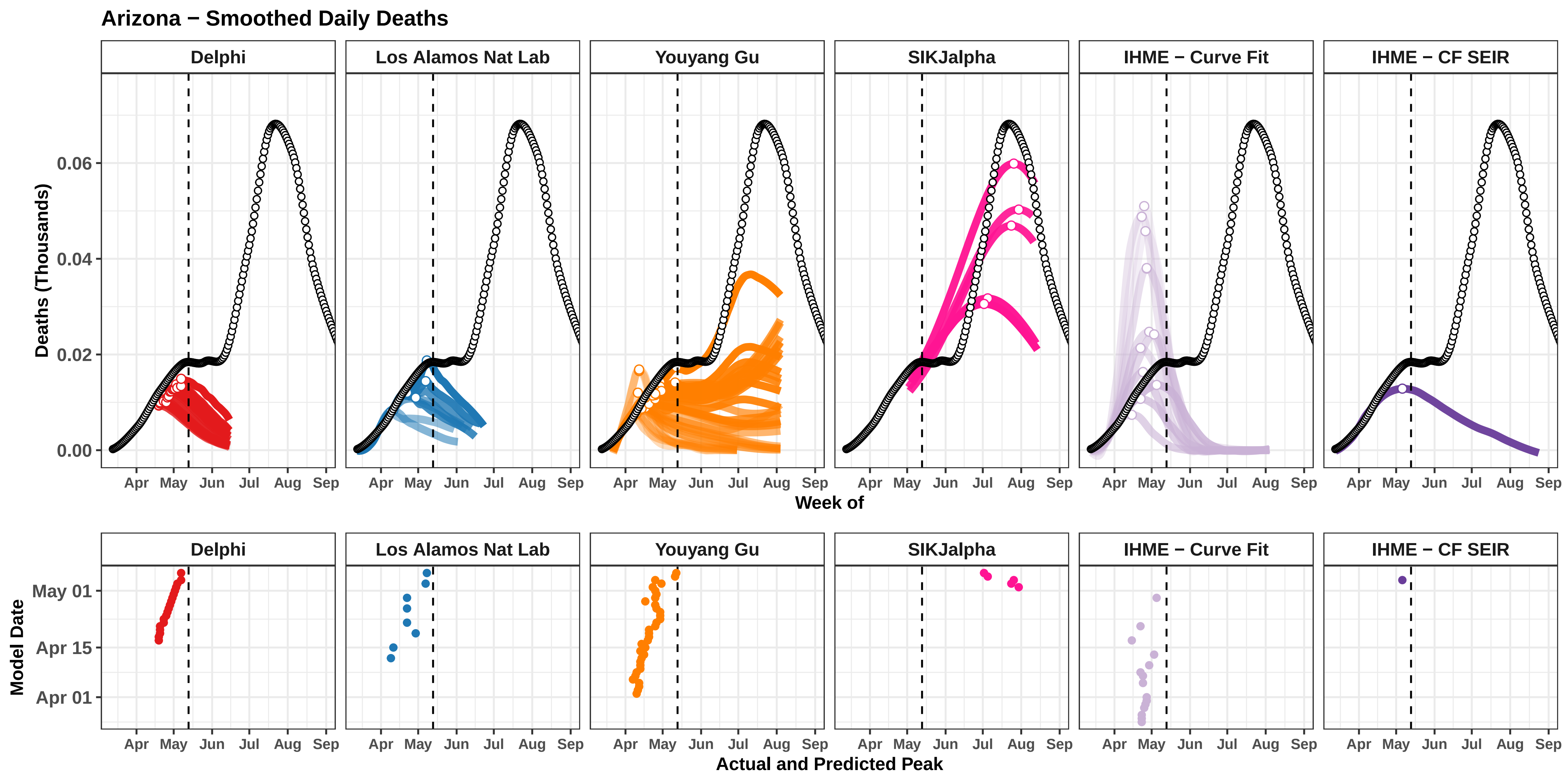

Arizona − Smoothed Daily Deaths
Delphi
Los Alamos Nat Lab
Youyang Gu
SIKJalpha
IHME − Curve Fit
IHME − CF SEIR
Deaths (Thousands)
0.00
0.02
0.04
0.06
Apr
May
Jun
Jul
Aug
Sep
Week of
Model Date
May 01
Apr 15
Apr 01
Actual and Predicted Peak

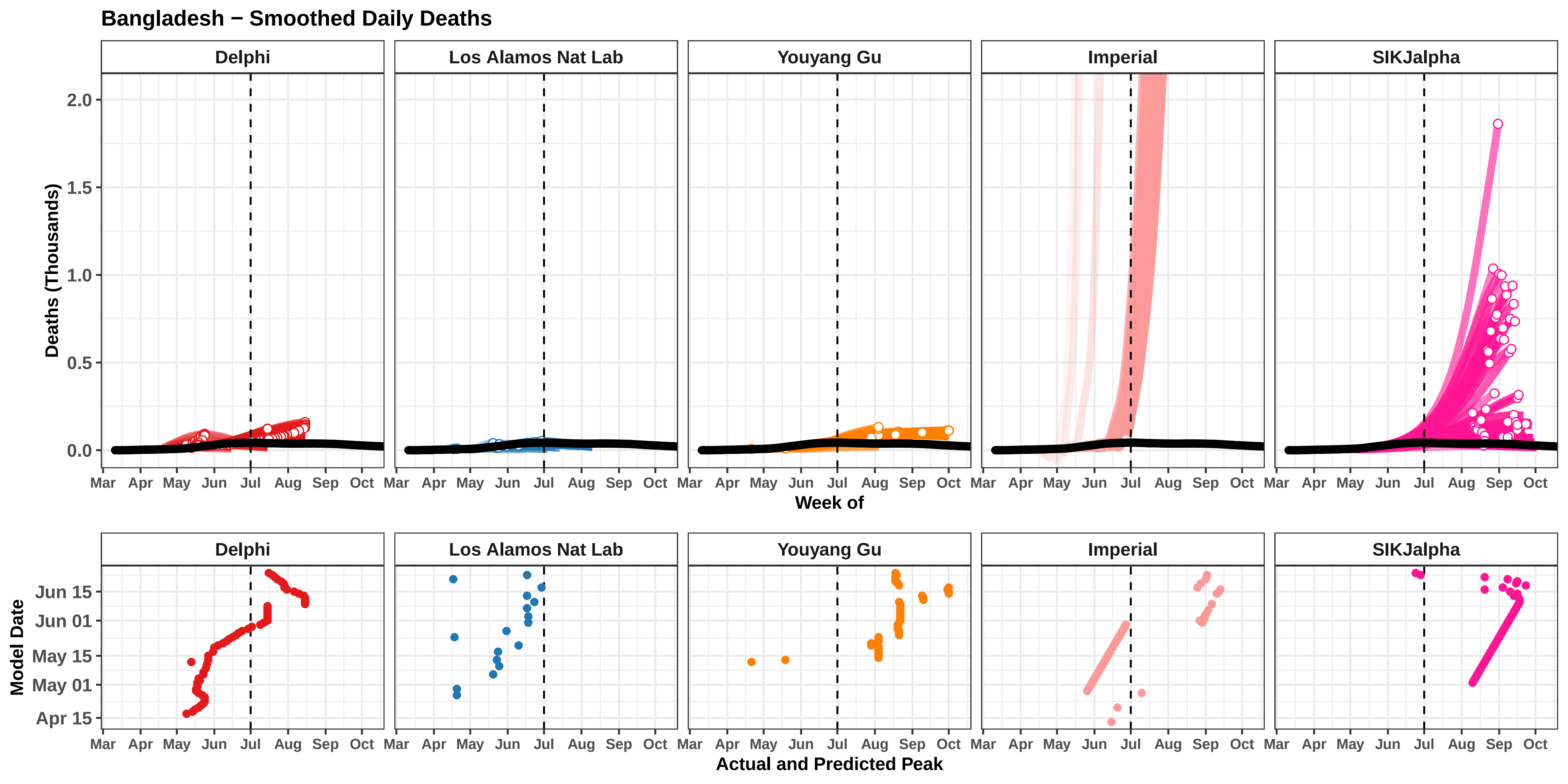

Bangladesh − Smoothed Daily Deaths
Delphi
Los Alamos Nat Lab
Youyang Gu
Imperial
SIKJalpha
Deaths (Thousands)
0.0
0.5
1.0
1.5
2.0
Mar
Apr
May
Jun
Jul
Aug
Sep
Oct
Week of
Model Date
Apr 15
May 01
May 15
Jun 01
Jun 15
Actual and Predicted Peak

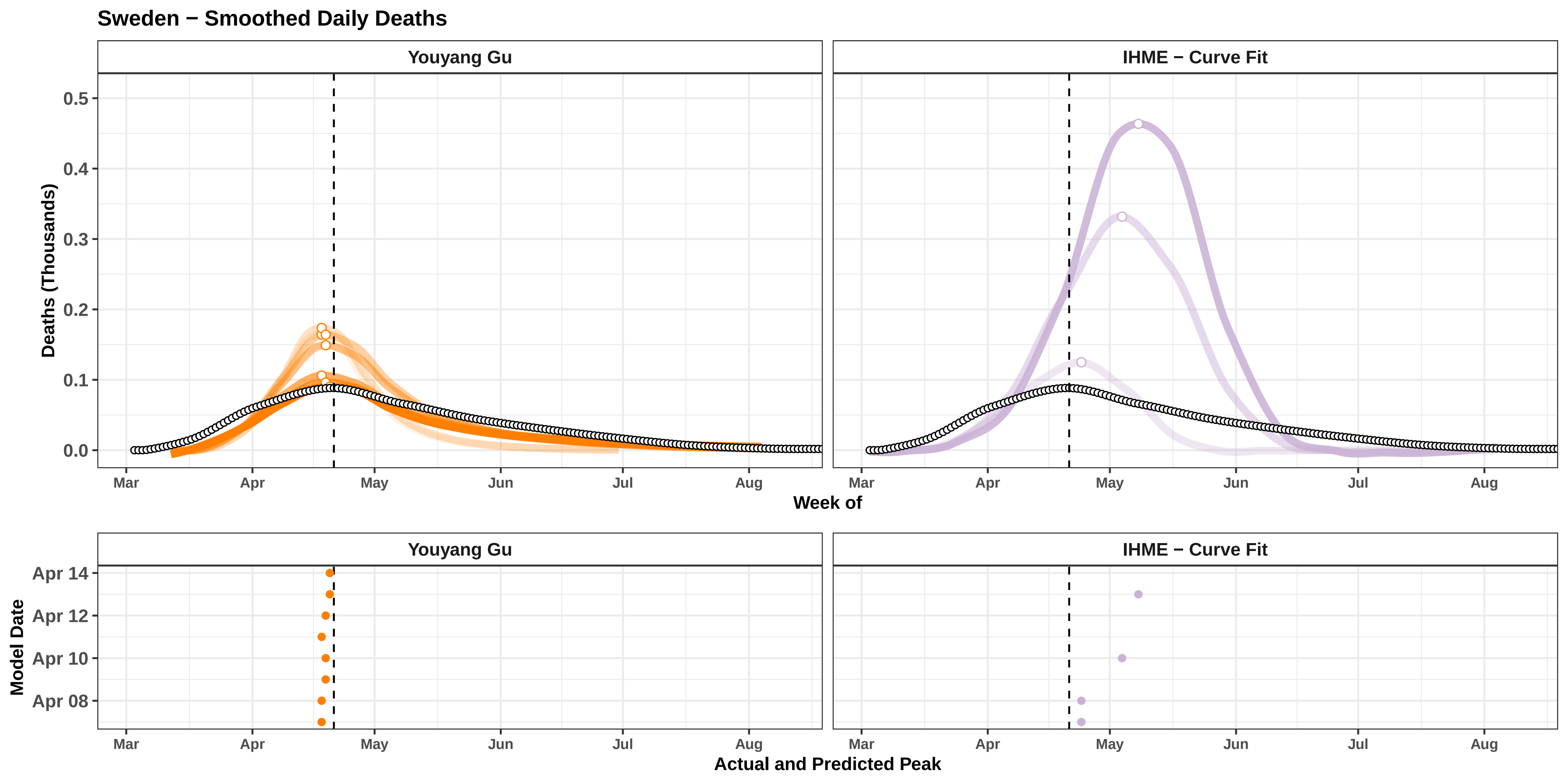

Sweden − Smoothed Daily Deaths
Youyang Gu
IHME − Curve Fit
Deaths (Thousands)
0.0
0.1
0.2
0.3
0.4
0.5
Mar
Apr
May
Jun
Jul
Aug
Week of
Model Date
Apr 08
Apr 10
Apr 12
Apr 14
Actual and Predicted Peak

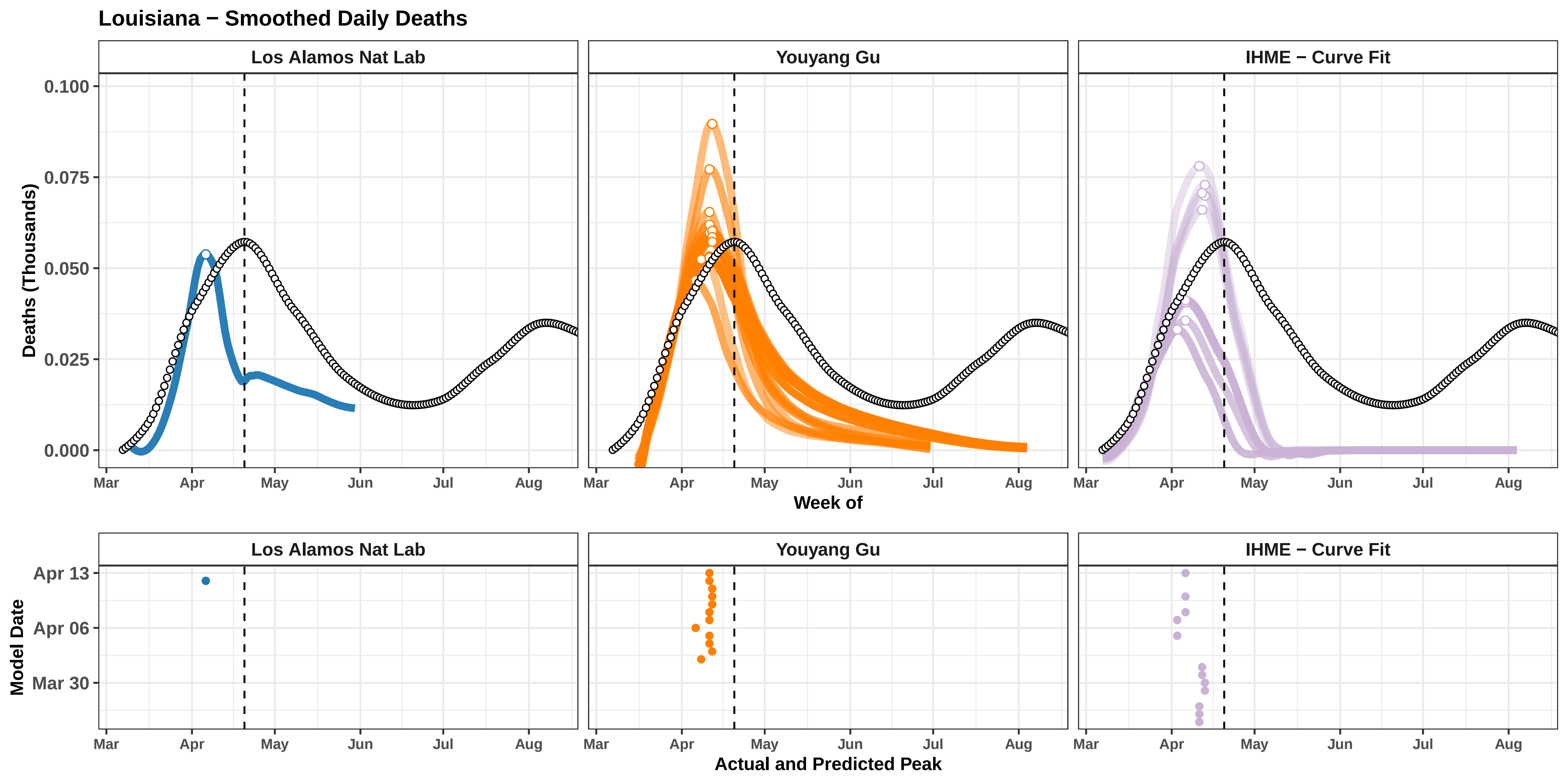

Louisiana − Smoothed Daily Deaths
Los Alamos Nat Lab
Youyang Gu
IHME − Curve Fit
Deaths (Thousands)
0.100
0.075
0.050
0.025
0.000
Mar
Apr
May
Jun
Jul
Aug
Week of
Model Date
Apr 13
Apr 06
Mar 30
Actual and Predicted Peak

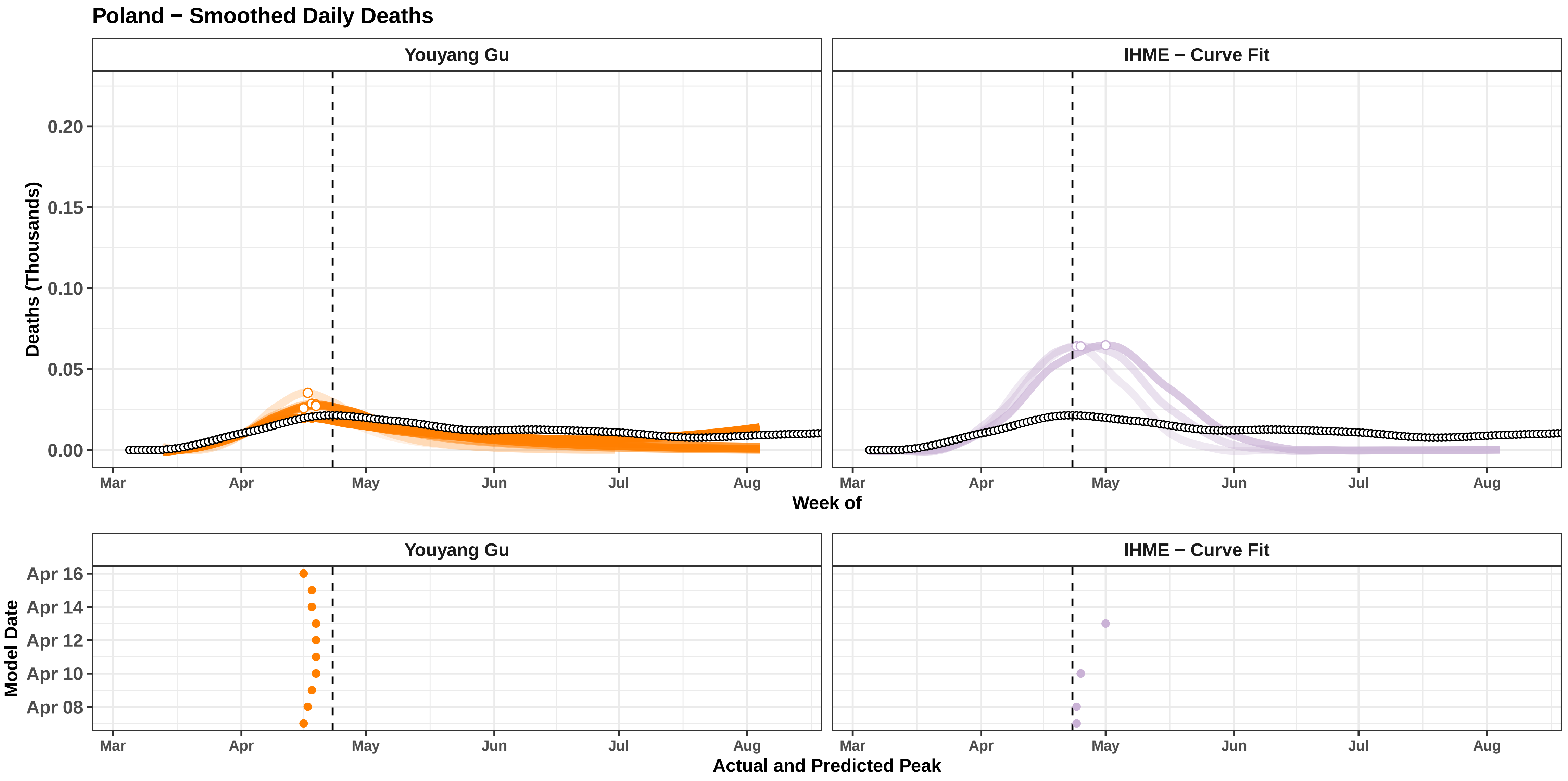

Poland − Smoothed Daily Deaths
Youyang Gu
IHME − Curve Fit
Deaths (Thousands)
0.20
0.15
0.10
0.05
0.00
Mar
Apr
May
Jun
Jul
Aug
Week of
Model Date
Apr 16
Apr 14
Apr 12
Apr 10
Apr 08
Actual and Predicted Peak

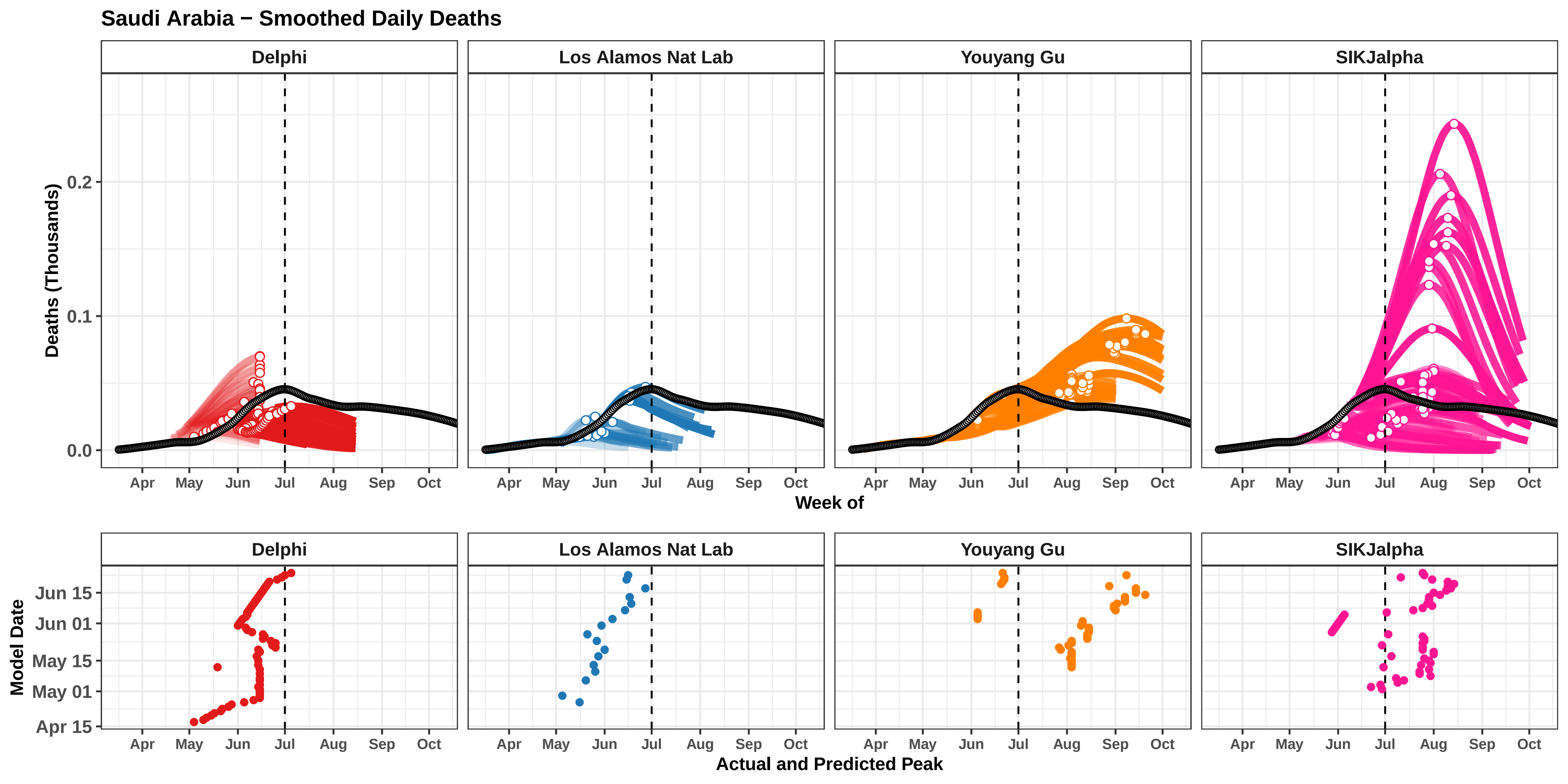
Saudi Arabia − Smoothed Daily Deaths
Delphi
Los Alamos Nat Lab
Youyang Gu
SIKJalpha
Deaths (Thousands)
0.0
0.1
0.2
Apr
May
Jun
Jul
Aug
Sep
Oct
Week of
Model Date
Apr 15
May 01
May 15
Jun 01
Jun 15
Actual and Predicted Peak

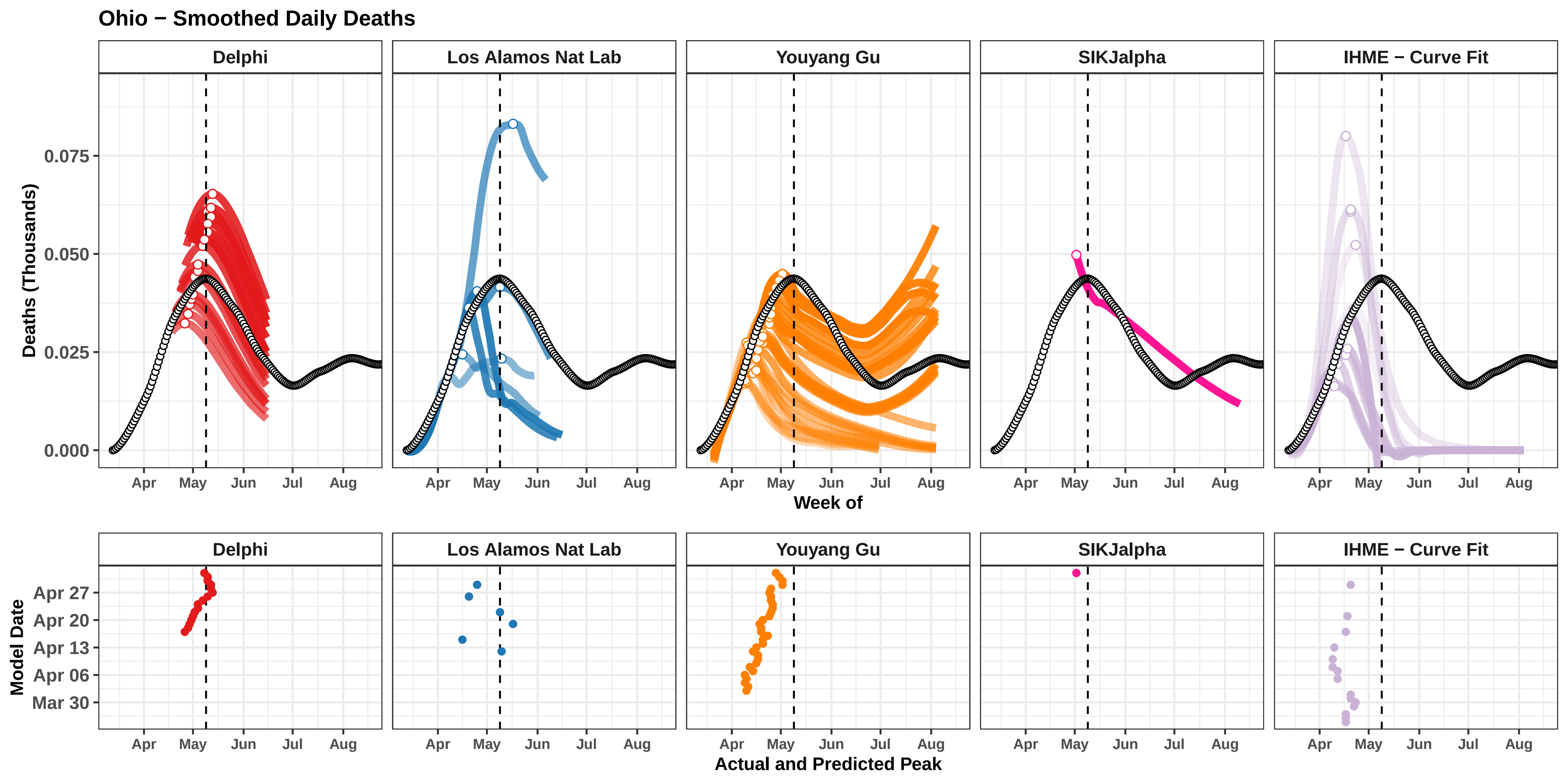

Ohio − Smoothed Daily Deaths
Delphi
Los Alamos Nat Lab
Youyang Gu
SIKJalpha
IHME − Curve Fit
Deaths (Thousands)
0.075
0.050
0.025
0.000
Apr
May
Jun
Jul
Aug
Week of
Model Date
Apr 27
Apr 20
Apr 13
Apr 06
Mar 30
Actual and Predicted Peak

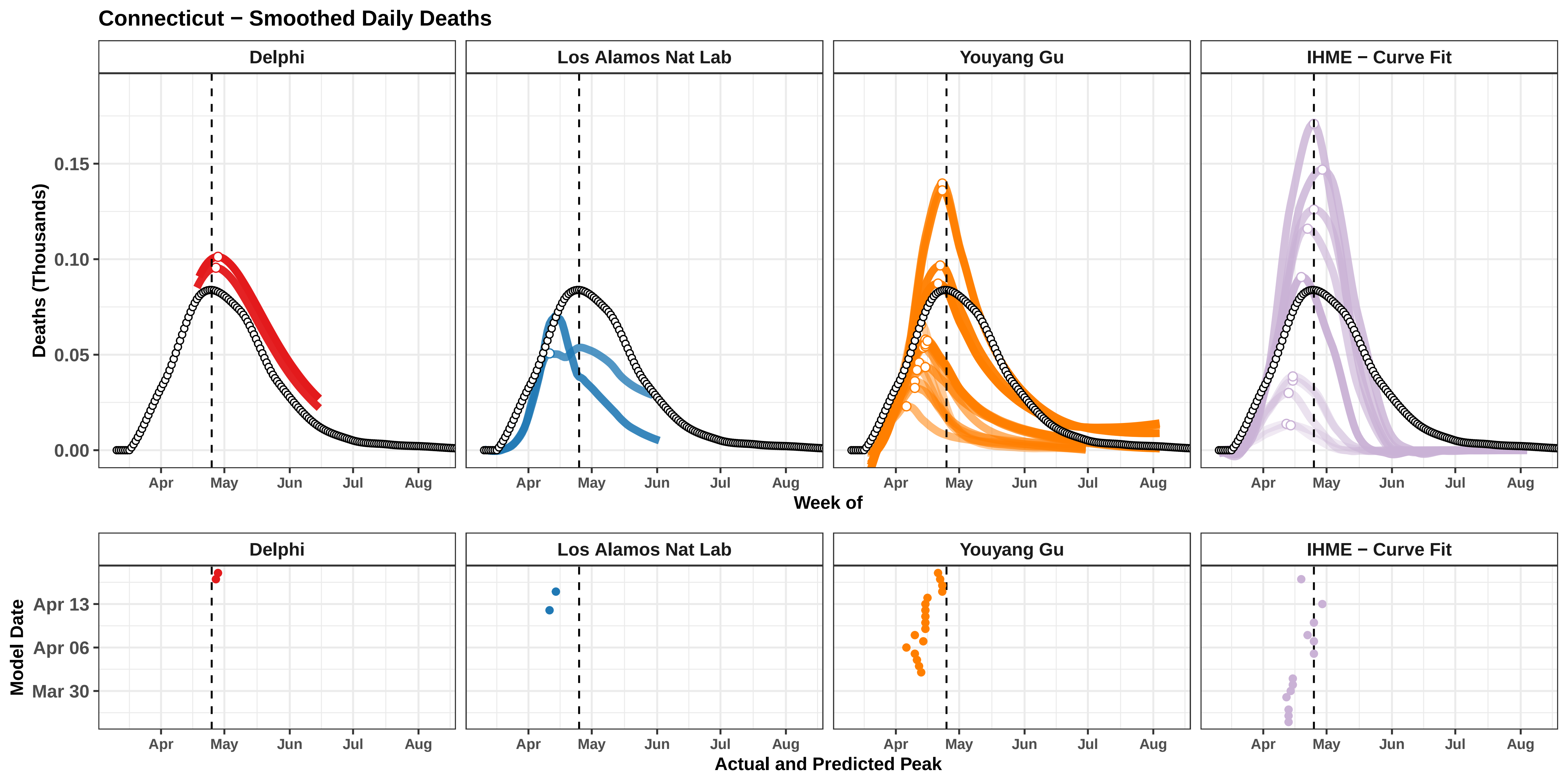

Connecticut − Smoothed Daily Deaths
Delphi
Los Alamos Nat Lab
Youyang Gu
IHME − Curve Fit
Deaths (Thousands)
0.15
0.10
0.05
0.00
Apr
May
Jun
Jul
Aug
Week of
Model Date
Apr 13
Apr 06
Mar 30
Actual and Predicted Peak

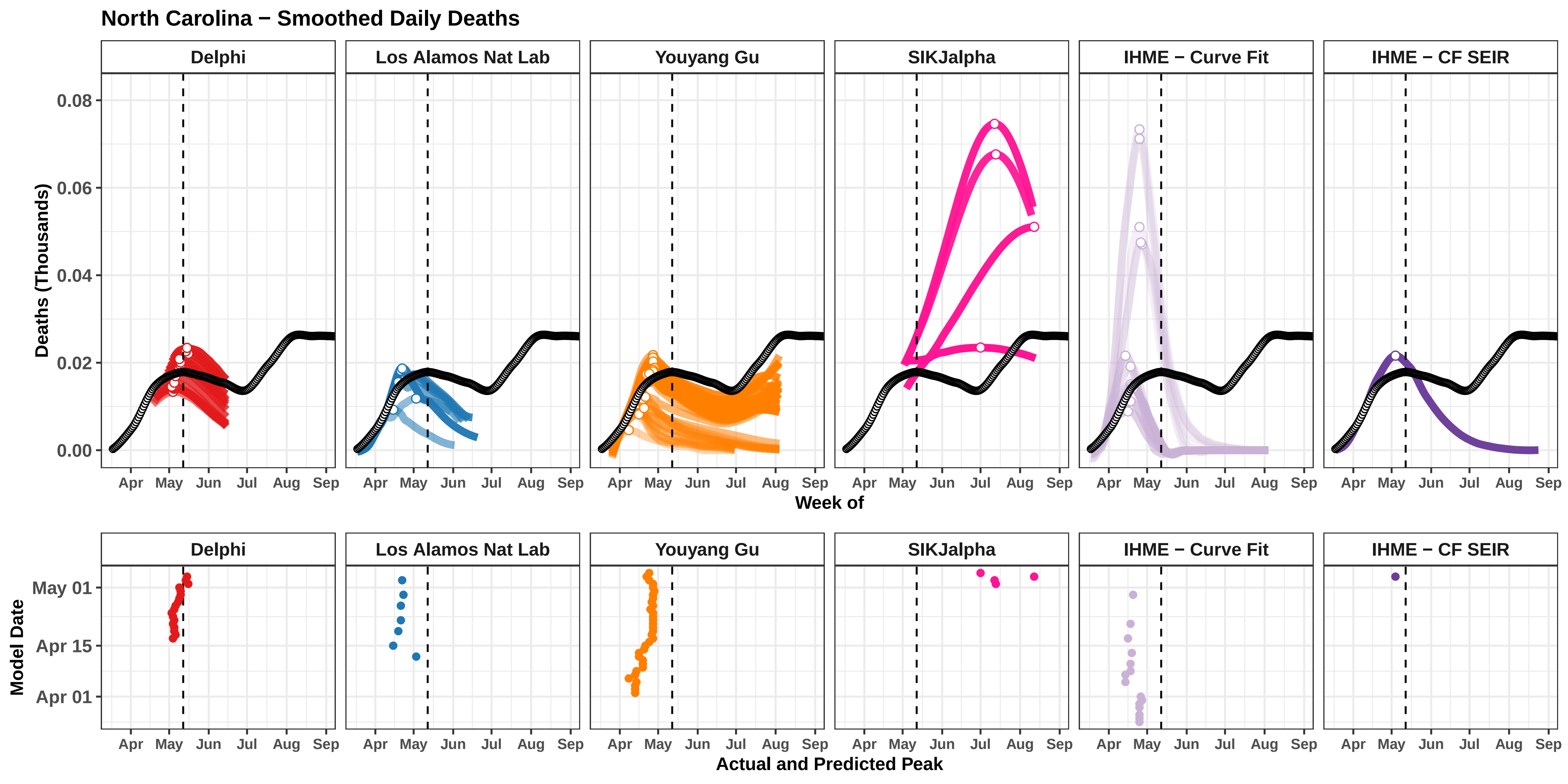

North Carolina − Smoothed Daily Deaths
Delphi
Los Alamos Nat Lab
Youyang Gu
SIKJalpha
IHME − Curve Fit
IHME − CF SEIR
Deaths (Thousands)
0.08
0.06
0.04
0.02
0.00
Apr May Jun Jul Aug Sep
Week of
Model Date
May 01
Apr 15
Apr 01
Actual and Predicted Peak

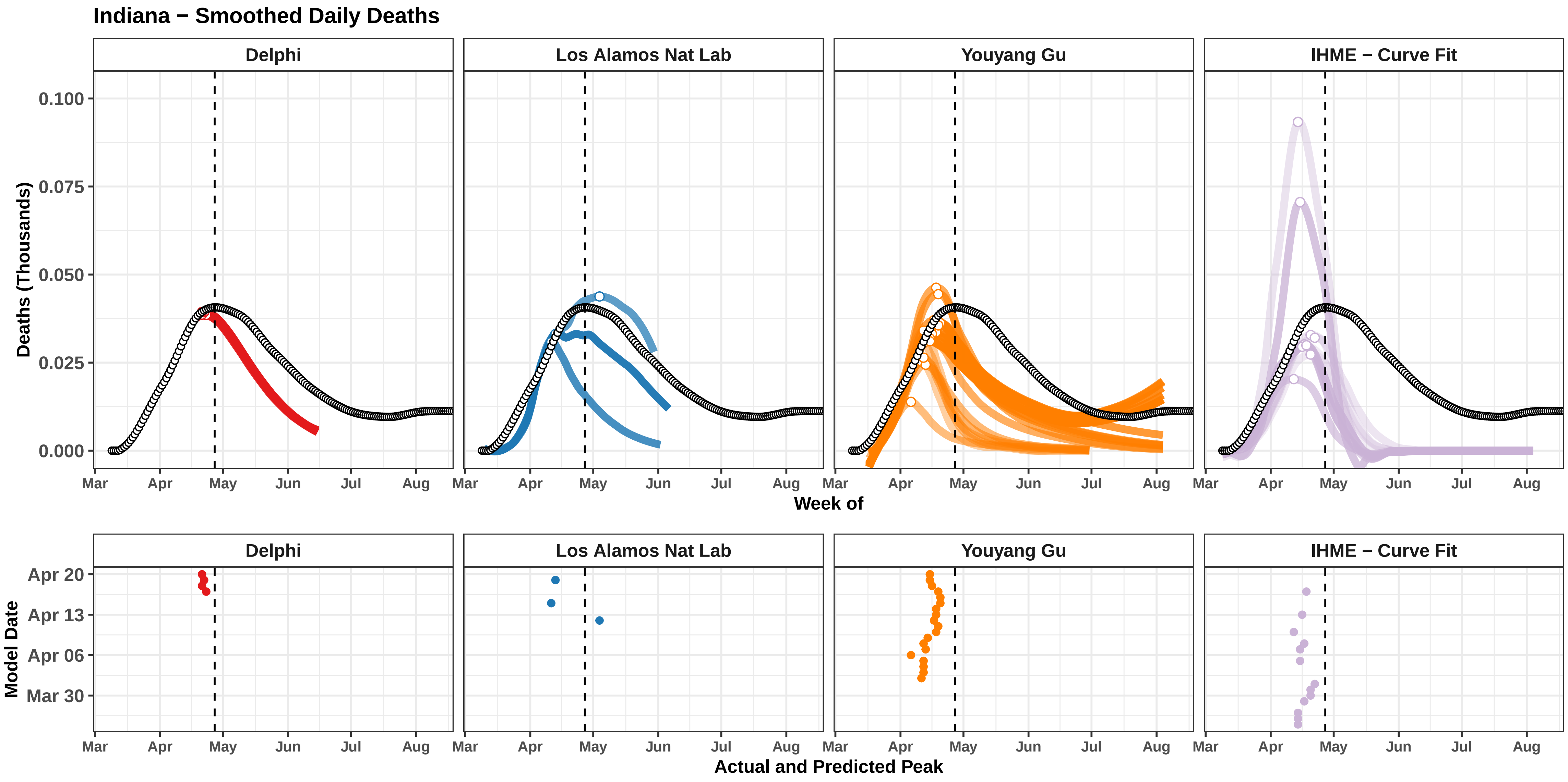

Indiana − Smoothed Daily Deaths
Delphi
Los Alamos Nat Lab
Youyang Gu
IHME − Curve Fit
Deaths (Thousands)
0.100
0.075
0.050
0.025
0.000
Mar
Apr
May
Jun
Jul
Aug
Week of
Model Date
Apr 20
Apr 13
Apr 06
Mar 30
Actual and Predicted Peak

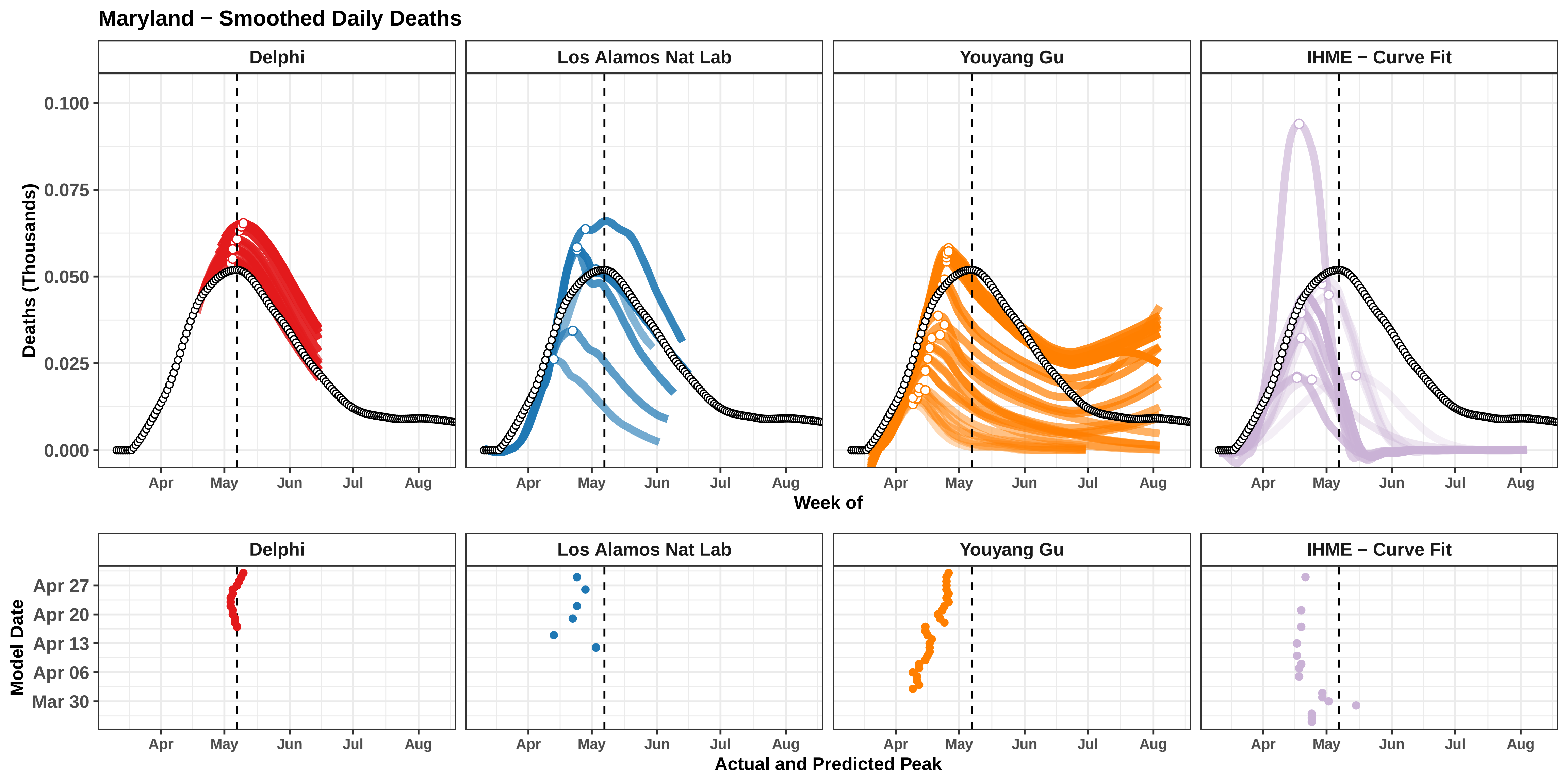

Maryland − Smoothed Daily Deaths
Delphi
Los Alamos Nat Lab
Youyang Gu
IHME − Curve Fit
Deaths (Thousands)
0.100
0.075
0.050
0.025
0.000
Apr
May
Jun
Jul
Aug
Week of
Model Date
Apr 27
Apr 20
Apr 13
Apr 06
Mar 30
Actual and Predicted Peak

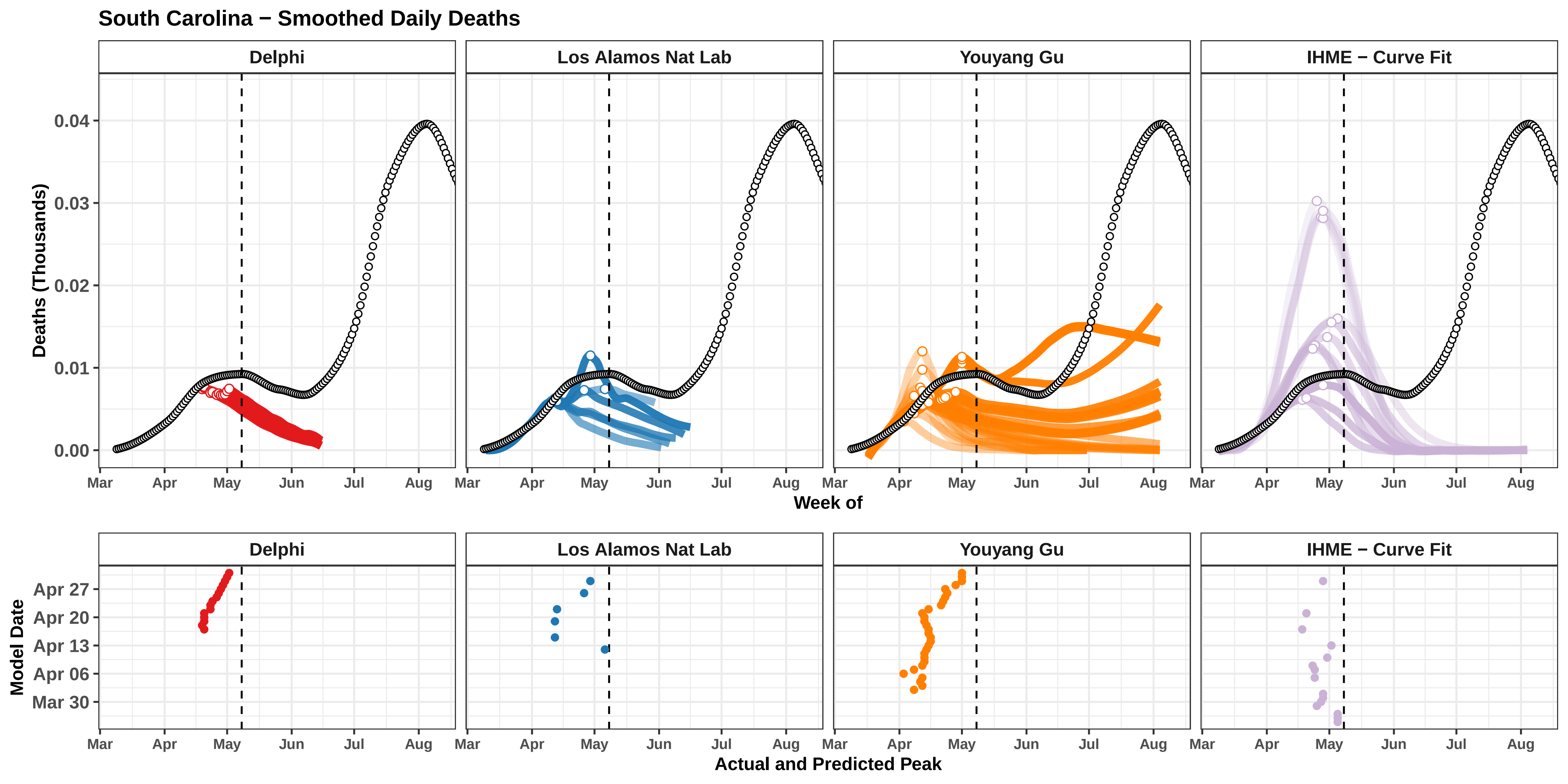

South Carolina − Smoothed Daily Deaths
Delphi
Los Alamos Nat Lab
Youyang Gu
IHME − Curve Fit
Deaths (Thousands)
0.00
0.01
0.02
0.03
0.04
Mar
Apr
May
Jun
Jul
Aug
Week of
Model Date
Apr 27
Apr 20
Apr 13
Apr 06
Mar 30
Actual and Predicted Peak

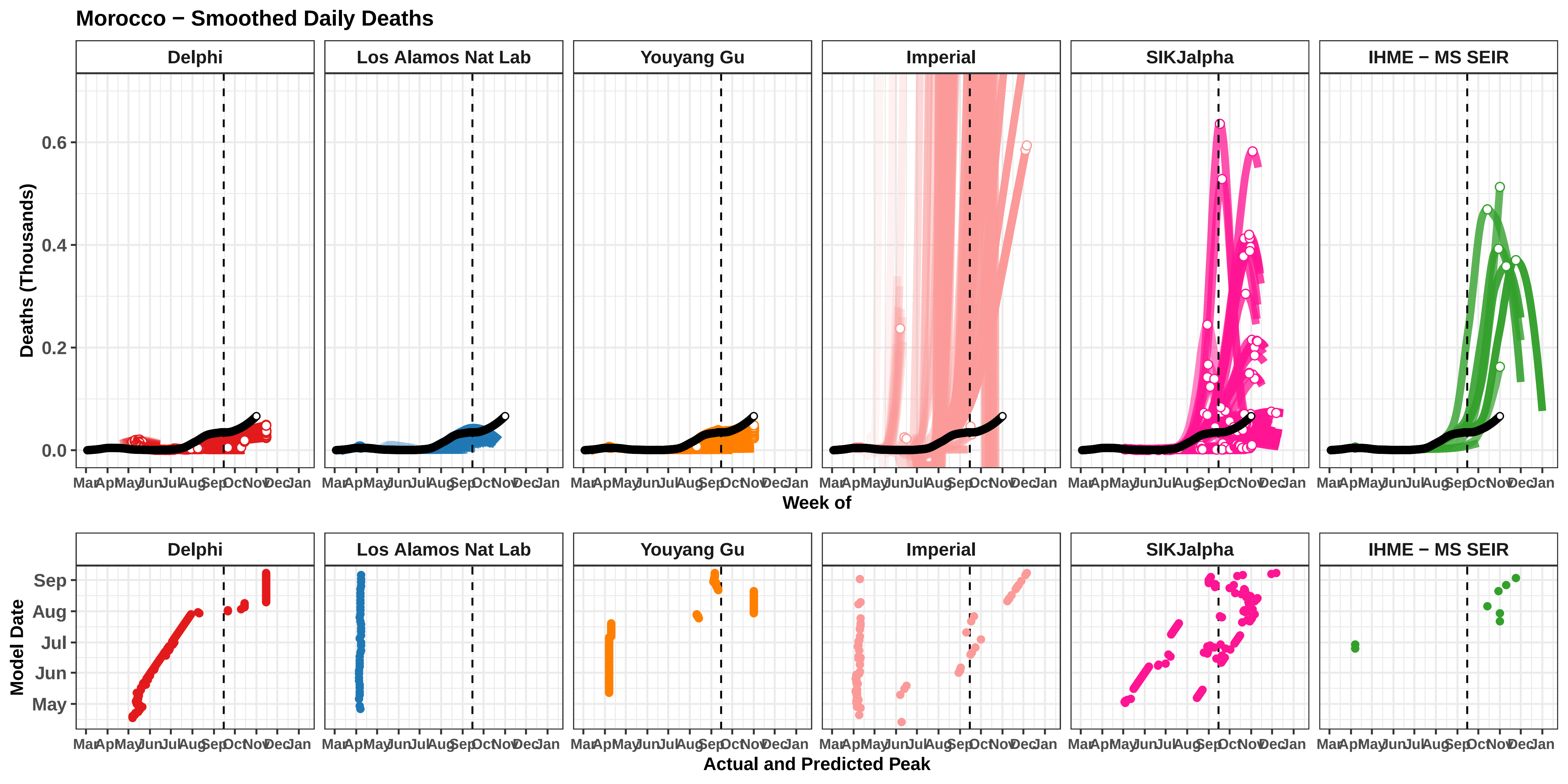

Morocco − Smoothed Daily Deaths
Delphi
Los Alamos Nat Lab
Youyang Gu
Imperial
SIKJalpha
IHME − MS SEIR
Deaths (Thousands)
0.0
0.2
0.4
0.6
Mar
Apr
May
Jun
Jul
Aug
Sep
Oct
Nov
Dec
Jan
Week of
Model Date
May
Jun
Jul
Aug
Sep
Actual and Predicted Peak

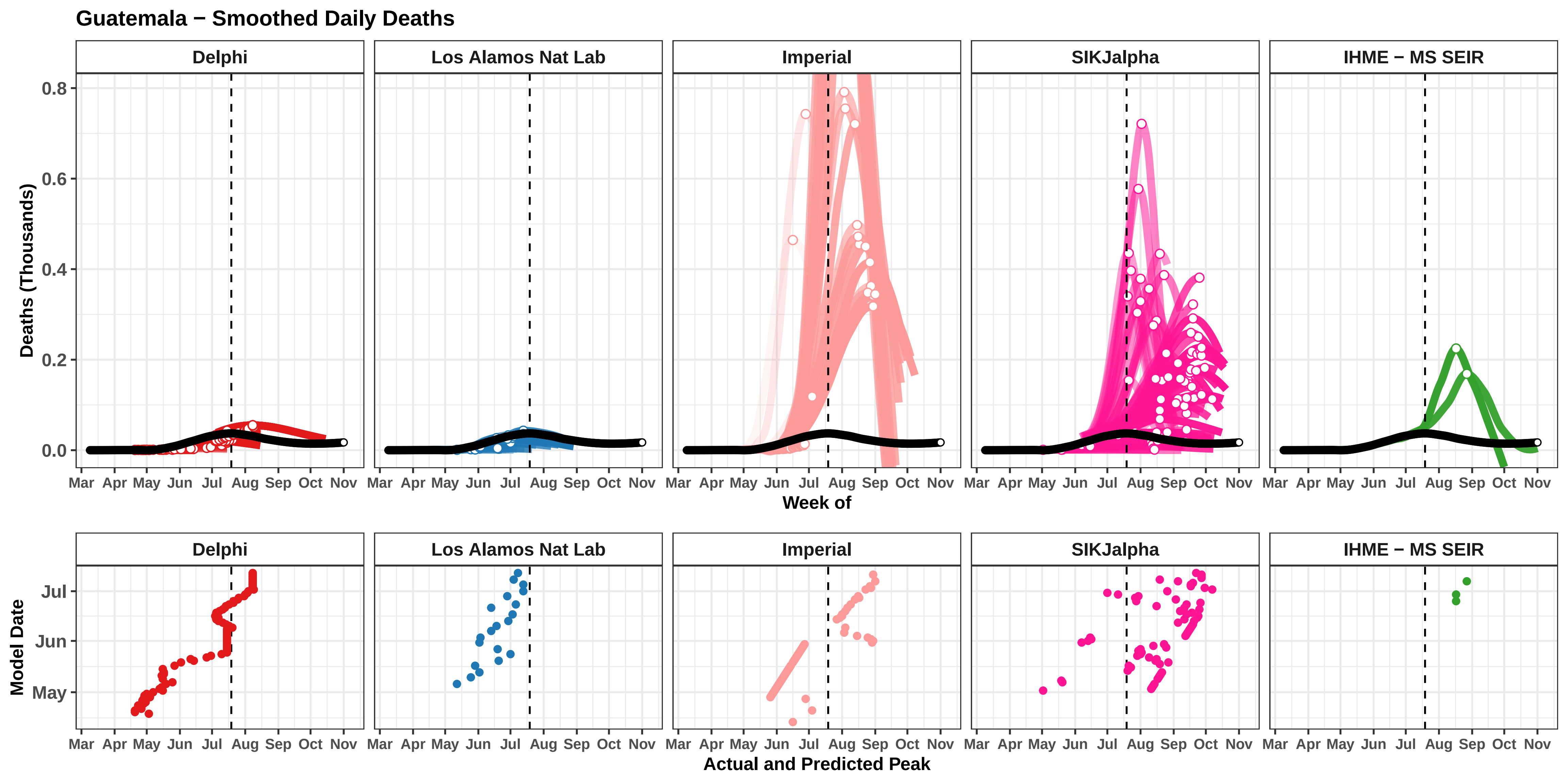
Guatemala − Smoothed Daily Deaths
Delphi
Los Alamos Nat Lab
Imperial
SIKJalpha
IHME − MS SEIR
Deaths (Thousands)
0.0
0.2
0.4
0.6
0.8
Mar
Apr
May
Jun
Jul
Aug
Sep
Oct
Nov
Week of
Model Date
May
Jun
Jul
Actual and Predicted Peak

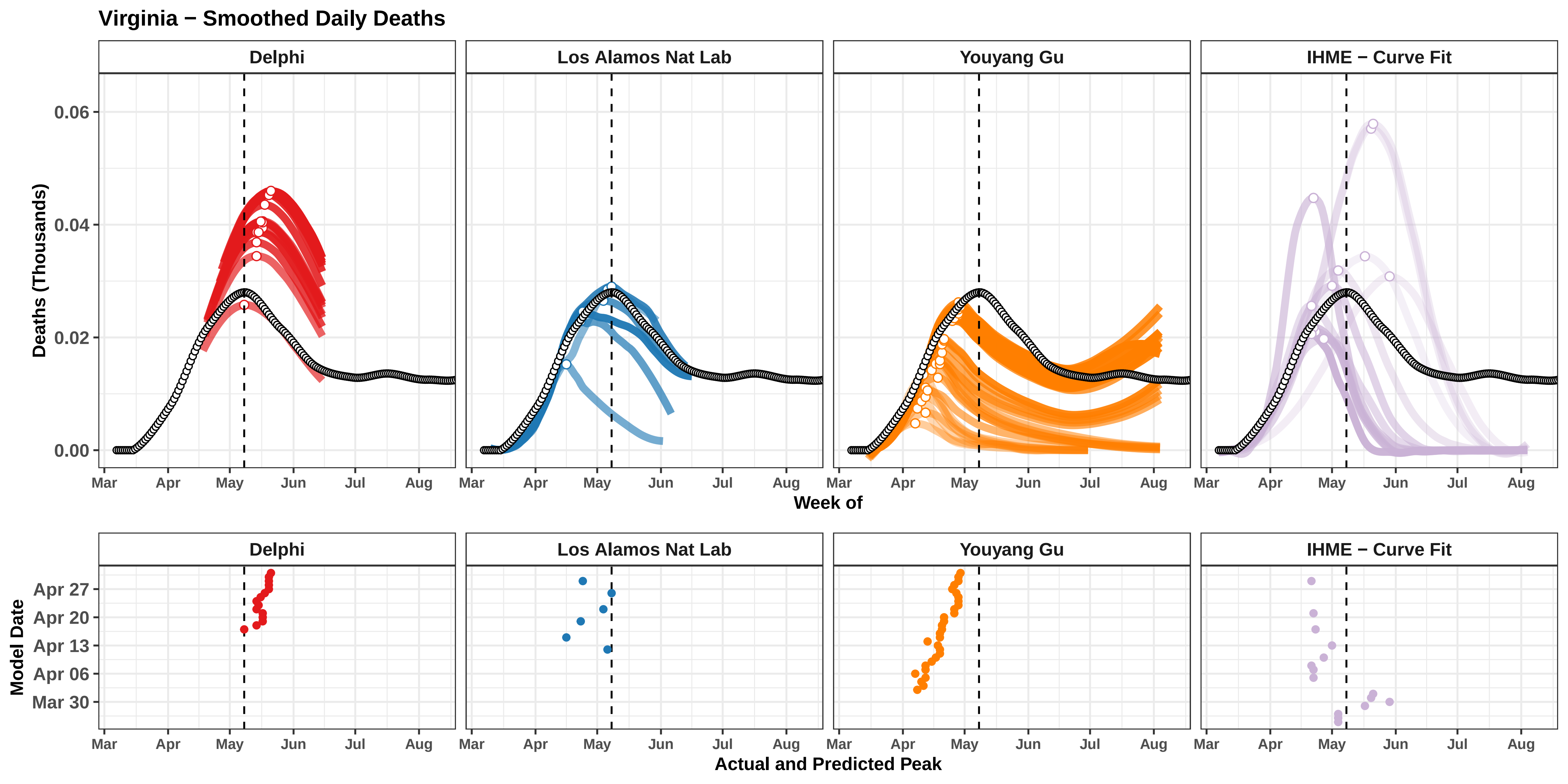

Virginia − Smoothed Daily Deaths
Delphi
Los Alamos Nat Lab
Youyang Gu
IHME − Curve Fit
Deaths (Thousands)
0.06
0.04
0.02
0.00
Mar
Apr
May
Jun
Jul
Aug
Week of
Model Date
Apr 27
Apr 20
Apr 13
Apr 06
Mar 30
Actual and Predicted Peak

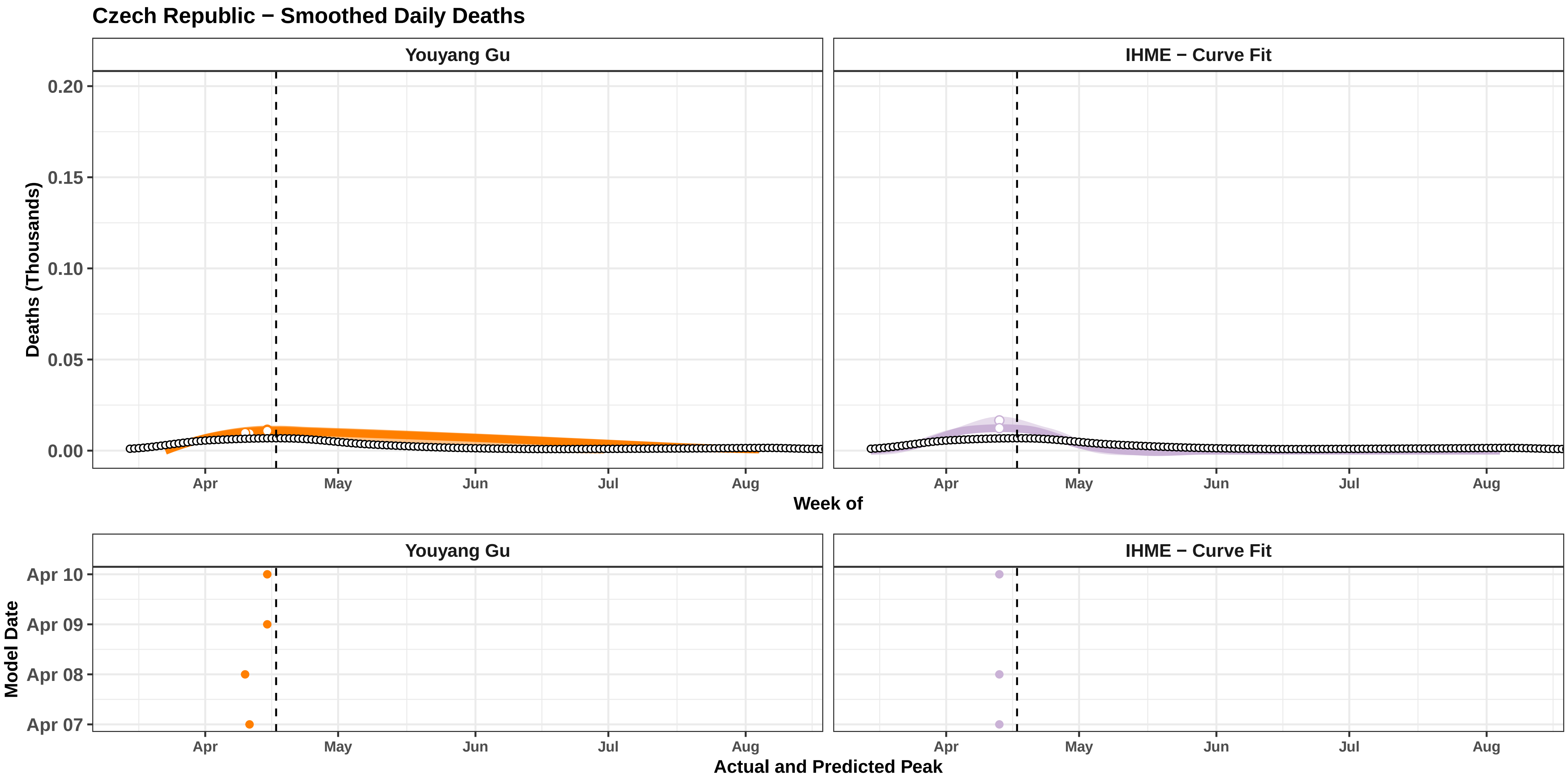

Czech Republic − Smoothed Daily Deaths
Youyang Gu
IHME − Curve Fit
Deaths (Thousands)
0.20
0.15
0.10
0.05
0.00
Apr
May
Jun
Jul
Aug
Week of
Model Date
Apr 10
Apr 09
Apr 08
Apr 07
Actual and Predicted Peak

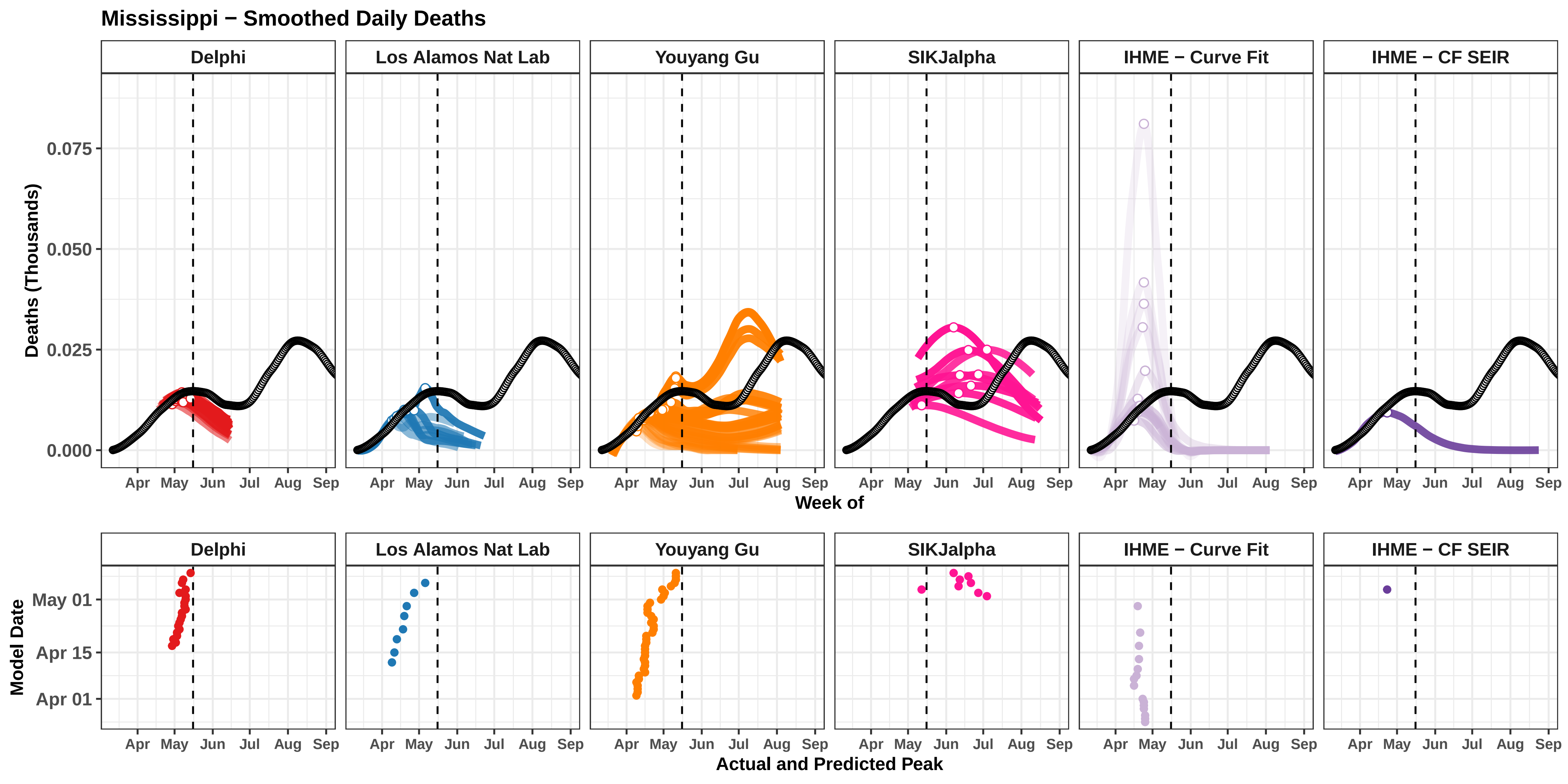
Mississippi − Smoothed Daily Deaths
Delphi
Los Alamos Nat Lab
Youyang Gu
SIKJalpha
IHME − Curve Fit
IHME − CF SEIR
Deaths (Thousands)
0.075
0.050
0.025
0.000
Apr
May
Jun
Jul
Aug
Sep
Week of
Model Date
May 01
Apr 15
Apr 01
Actual and Predicted Peak

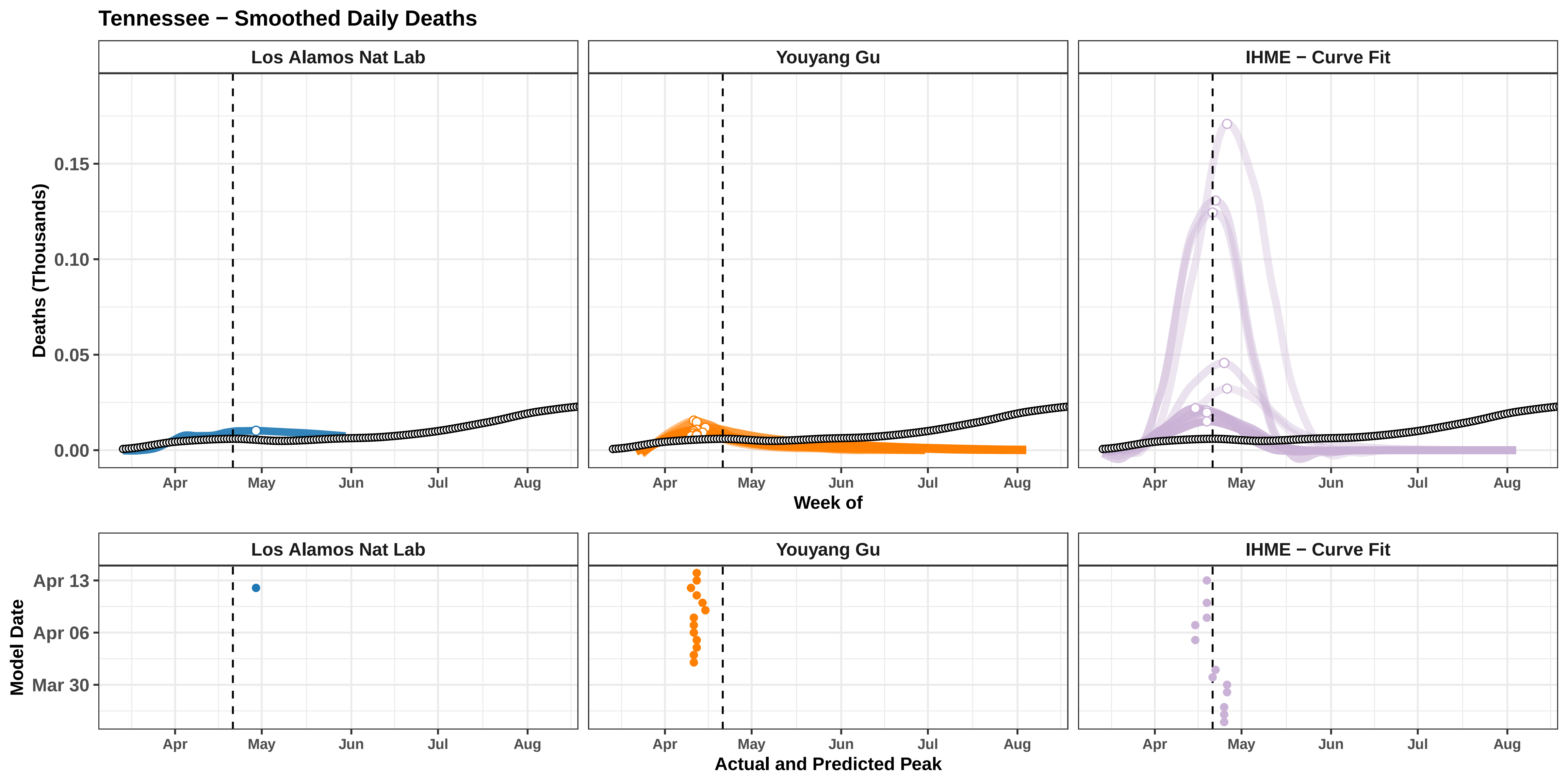

Tennessee − Smoothed Daily Deaths
Los Alamos Nat Lab
Youyang Gu
IHME − Curve Fit
Deaths (Thousands)
0.15
0.10
0.05
0.00
Apr
May
Jun
Jul
Aug
Week of
Model Date
Apr 13
Apr 06
Mar 30
Actual and Predicted Peak

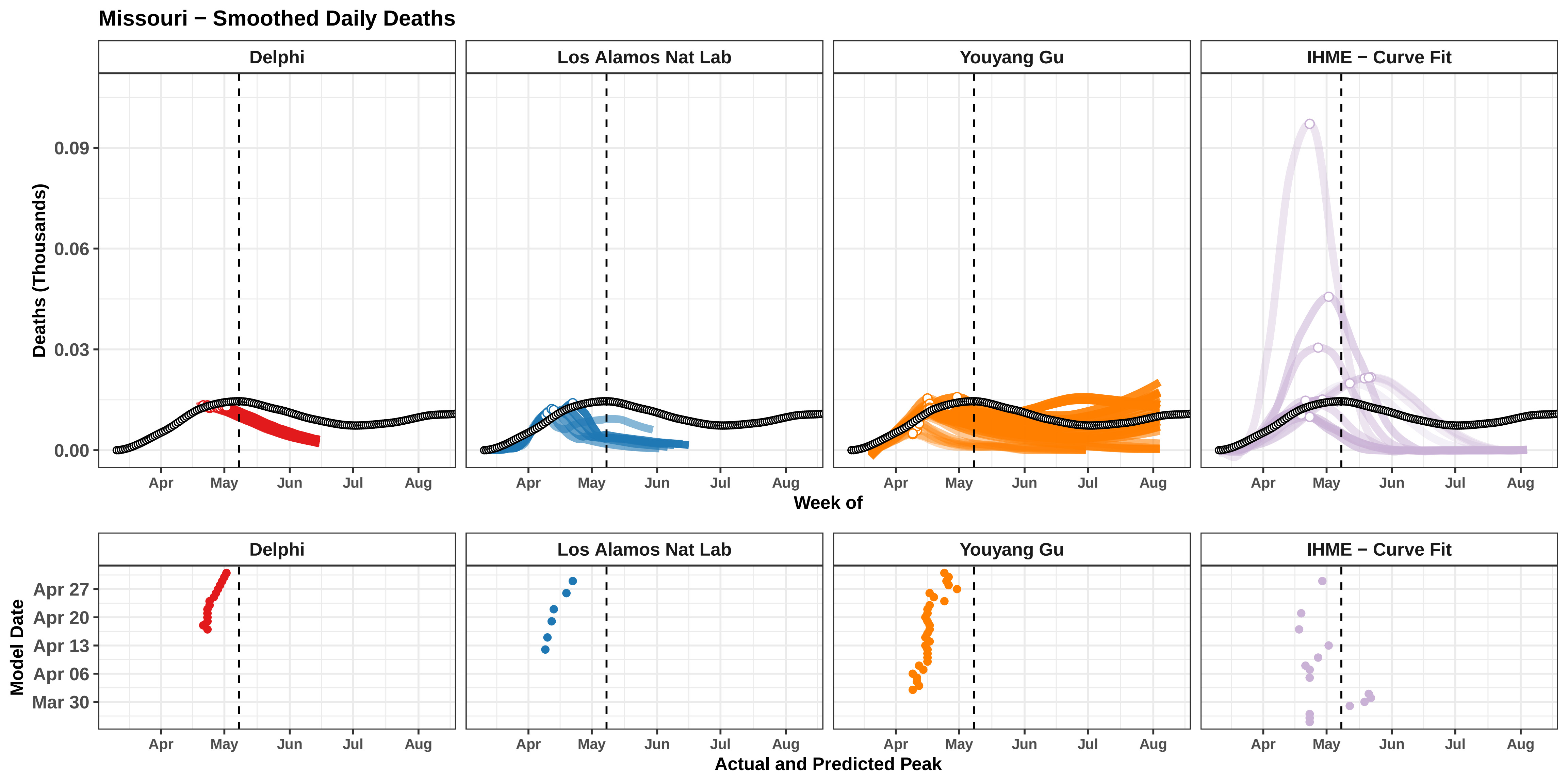

Missouri − Smoothed Daily Deaths
Delphi
Los Alamos Nat Lab
Youyang Gu
IHME − Curve Fit
Deaths (Thousands)
0.09
0.06
0.03
0.00
Apr
May
Jun
Jul
Aug
Week of
Model Date
Apr 27
Apr 20
Apr 13
Apr 06
Mar 30
Actual and Predicted Peak

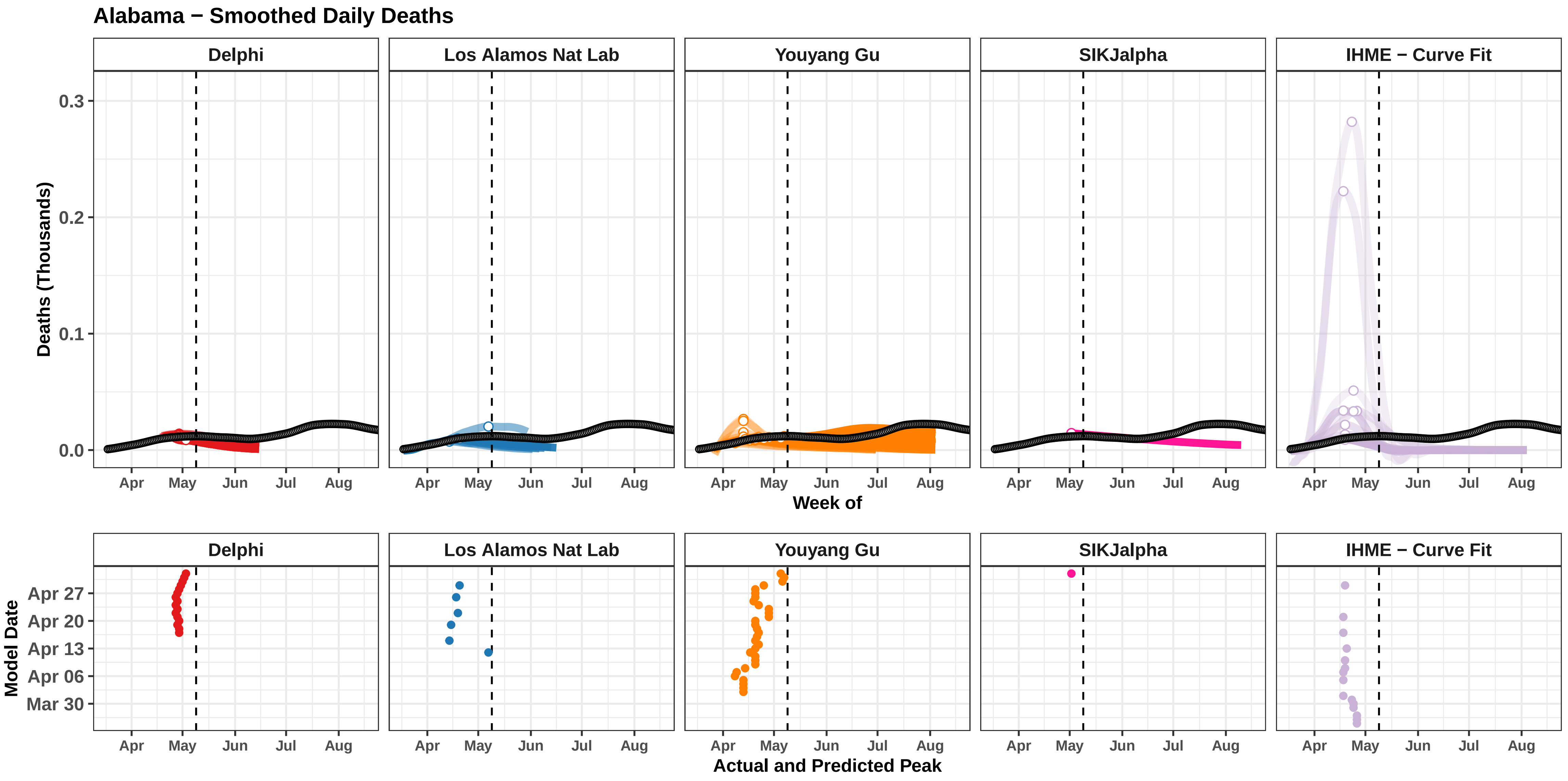

Alabama − Smoothed Daily Deaths
Delphi
Los Alamos Nat Lab
Youyang Gu
SIKJalpha
IHME − Curve Fit
Deaths (Thousands)
0.3
0.2
0.1
0.0
Apr
May
Jun
Jul
Aug
Week of
Model Date
Apr 27
Apr 20
Apr 13
Apr 06
Mar 30
Actual and Predicted Peak

### Panama − Smoothed Daily Deaths

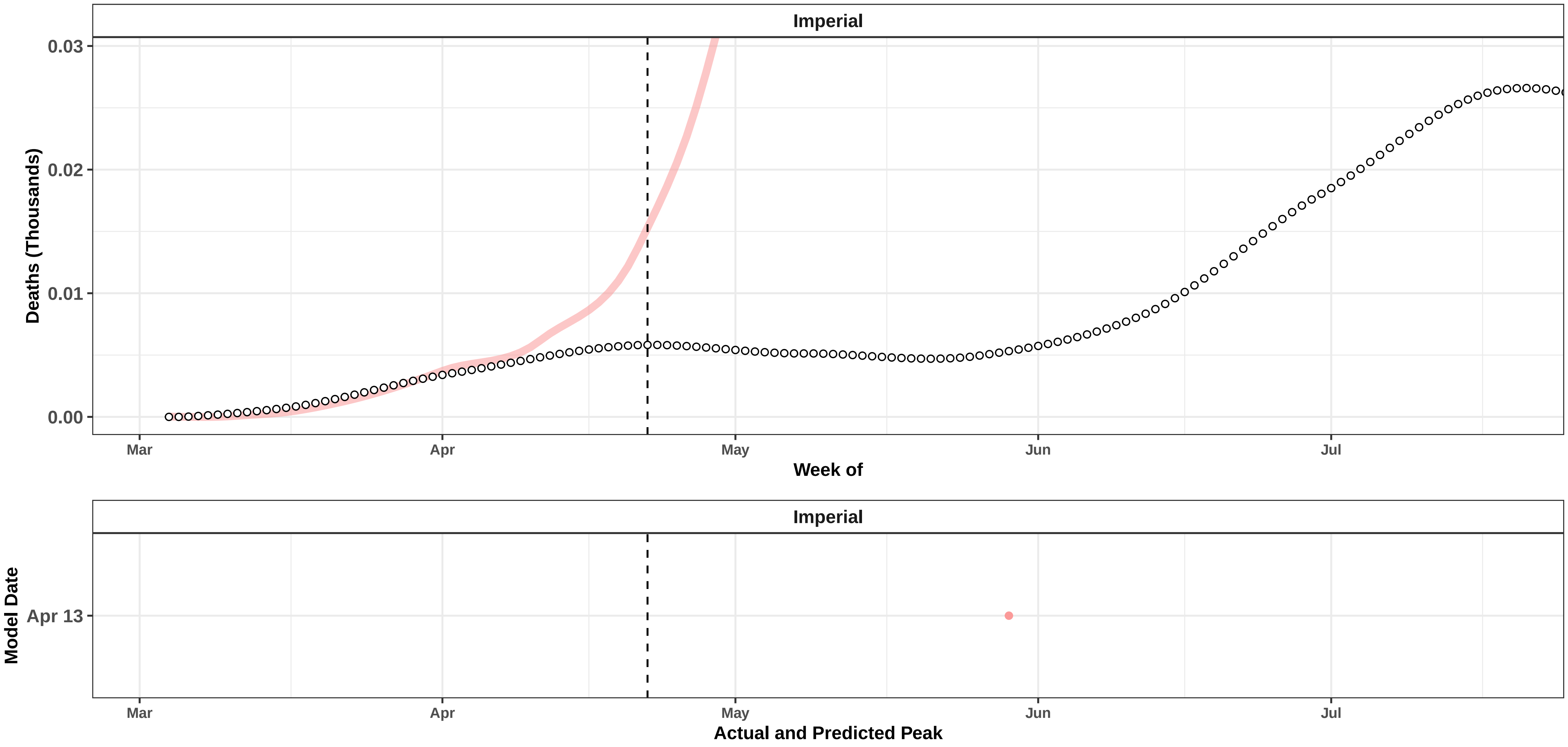

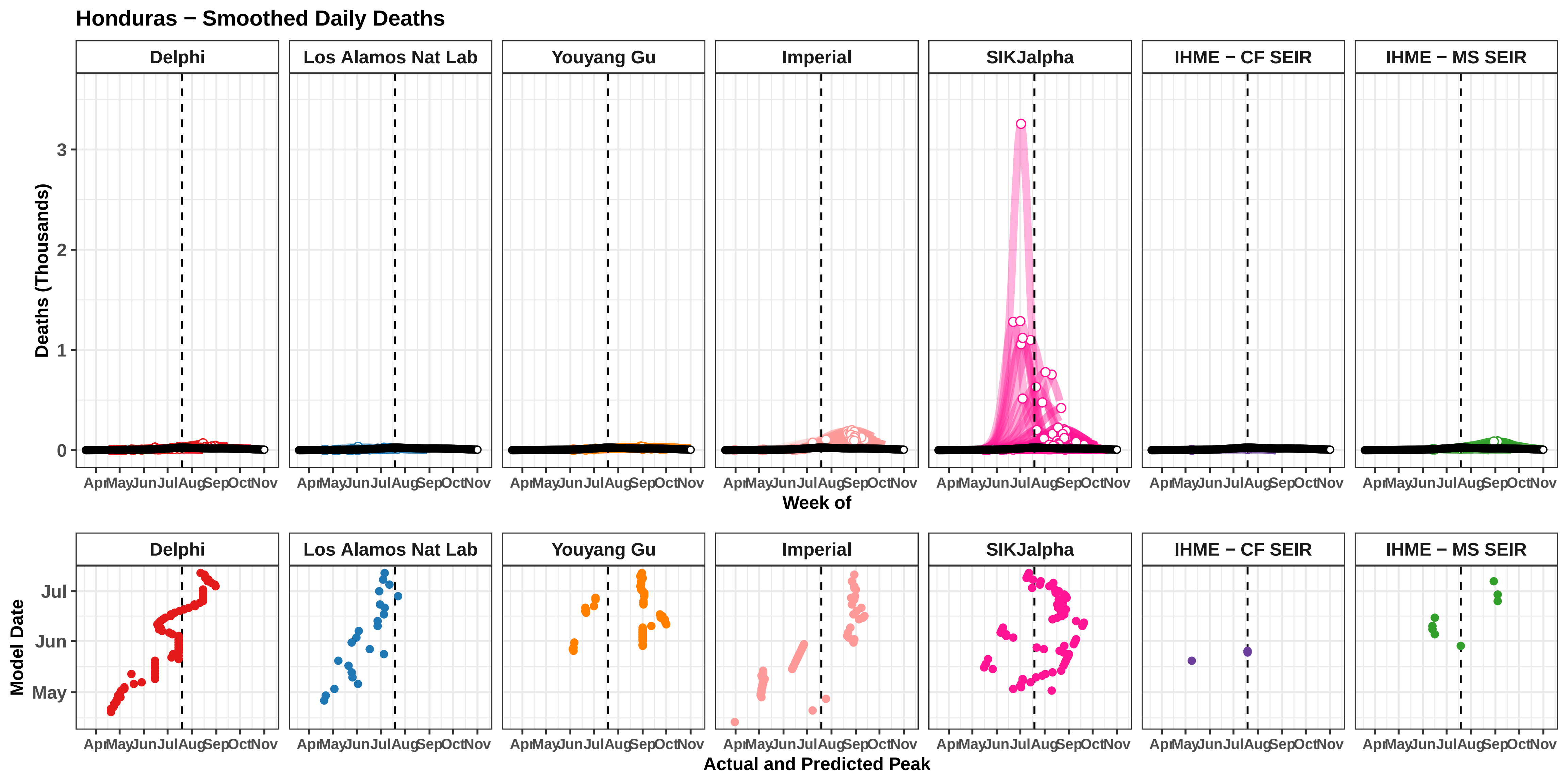
Honduras − Smoothed Daily Deaths
Delphi
Los Alamos Nat Lab
Youyang Gu
Imperial
SIKJalpha
IHME − CF SEIR
IHME − MS SEIR
Deaths (Thousands)
0
1
2
3
Apr May Jun Jul Aug Sep Oct Nov
Week of
Model Date
May
Jun
Jul
Actual and Predicted Peak

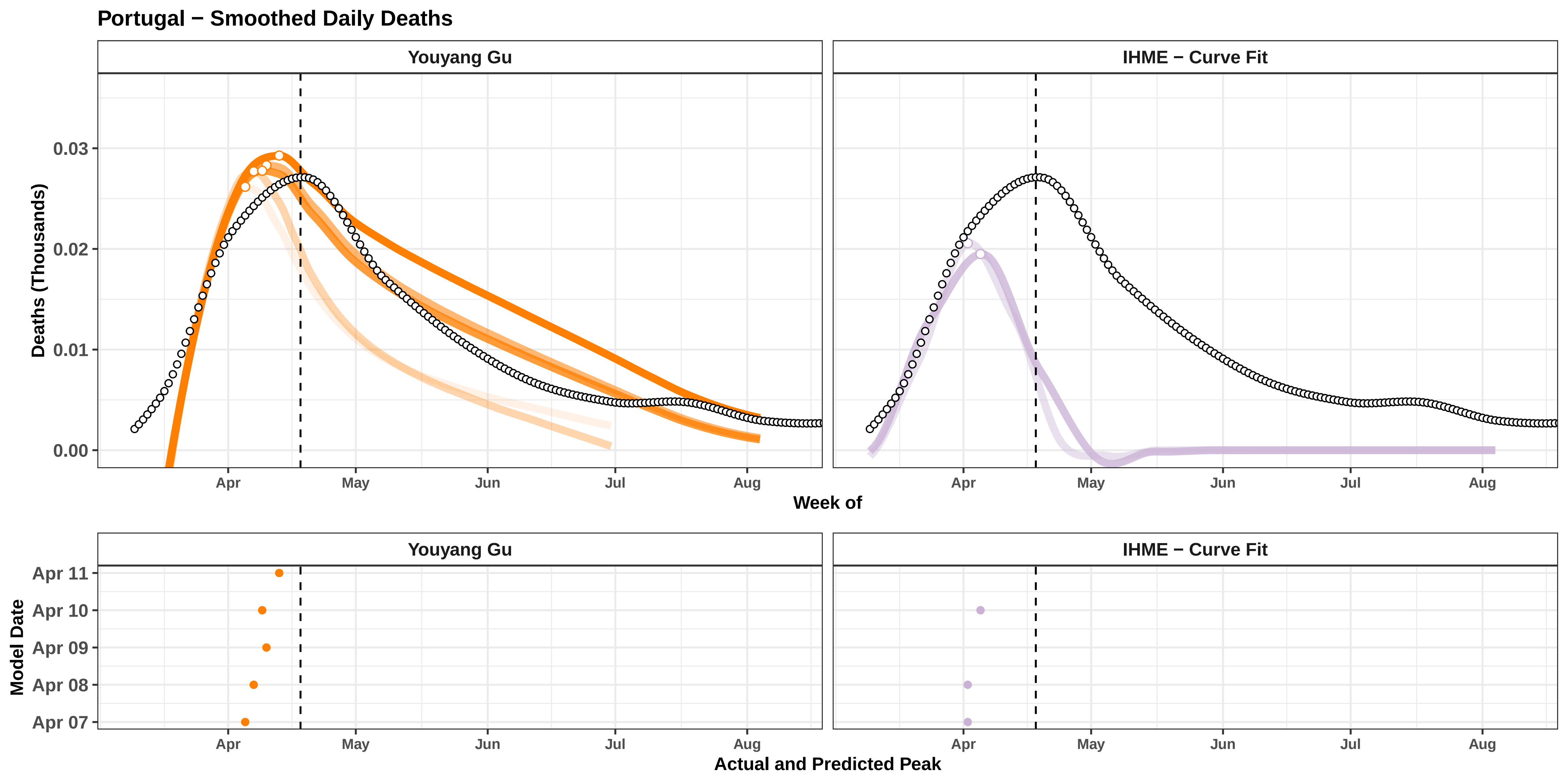

Portugal − Smoothed Daily Deaths
Youyang Gu
IHME − Curve Fit
Deaths (Thousands)
0.03
0.02
0.01
0.00
Apr
May
Jun
Jul
Aug
Week of
Youyang Gu
IHME − Curve Fit
Model Date
Apr 11
Apr 10
Apr 09
Apr 08
Apr 07
Apr
May
Jun
Jul
Aug
Actual and Predicted Peak

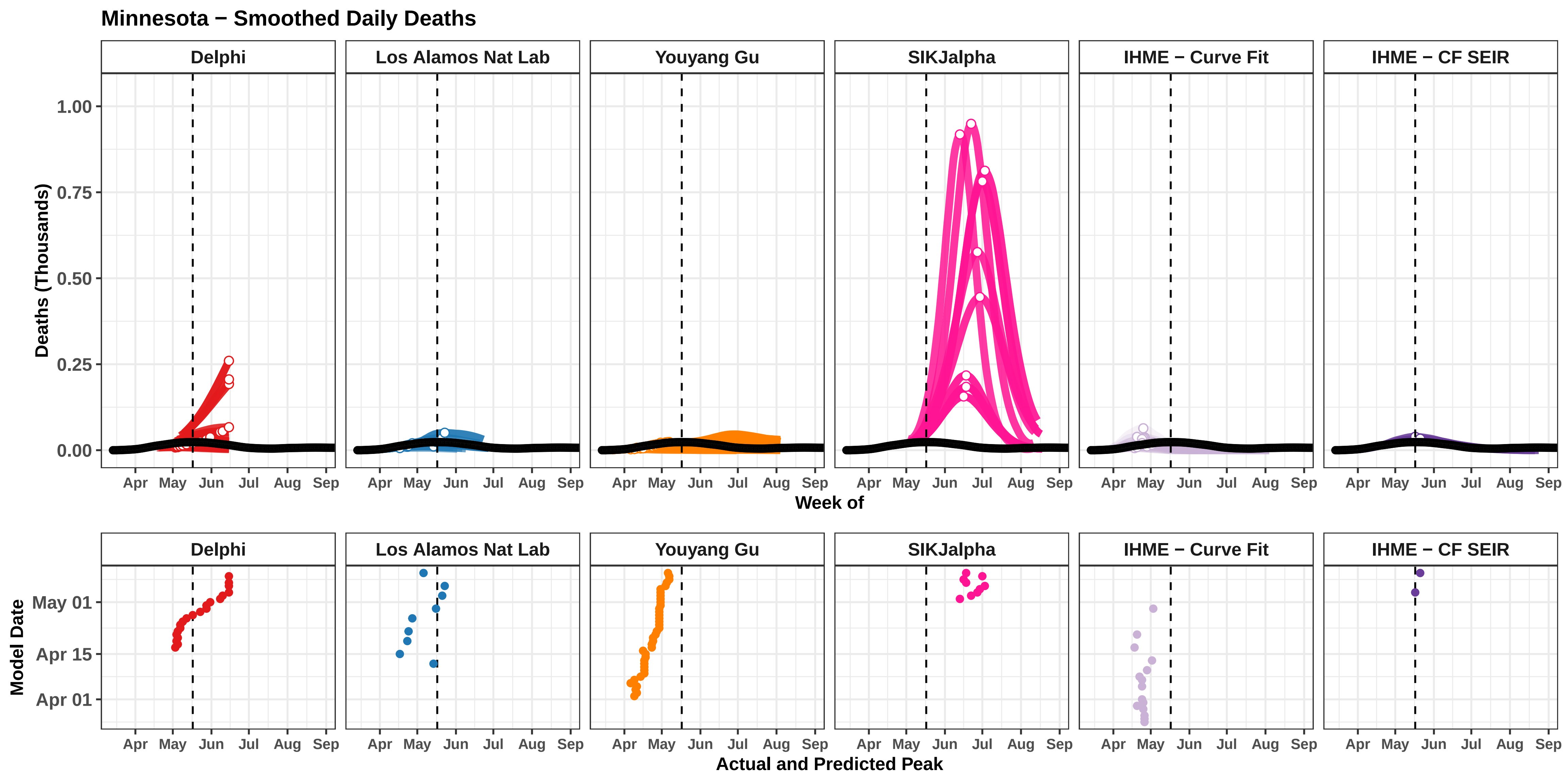

Minnesota − Smoothed Daily Deaths
Delphi
Los Alamos Nat Lab
Youyang Gu
SIKJalpha
IHME − Curve Fit
IHME − CF SEIR
Deaths (Thousands)
1.00
0.75
0.50
0.25
0.00
Apr
May
Jun
Jul
Aug
Sep
Week of
Model Date
May 01
Apr 15
Apr 01
Actual and Predicted Peak

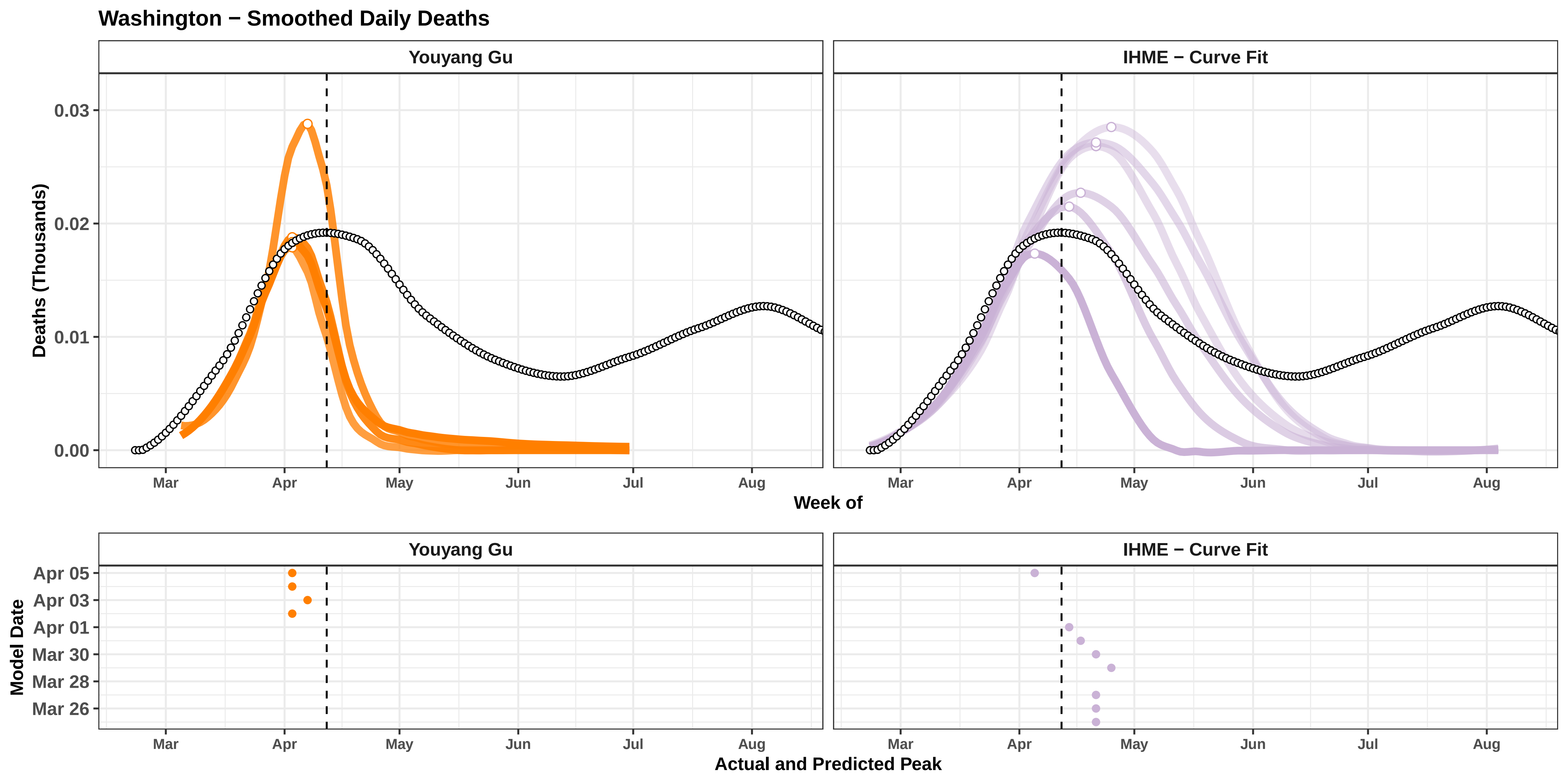
Washington − Smoothed Daily Deaths
Youyang Gu
IHME − Curve Fit
Deaths (Thousands)
0.03
0.02
0.01
0.00
Mar
Apr
May
Jun
Jul
Aug
Week of
Model Date
Apr 05
Apr 03
Apr 01
Mar 30
Mar 28
Mar 26
Actual and Predicted Peak

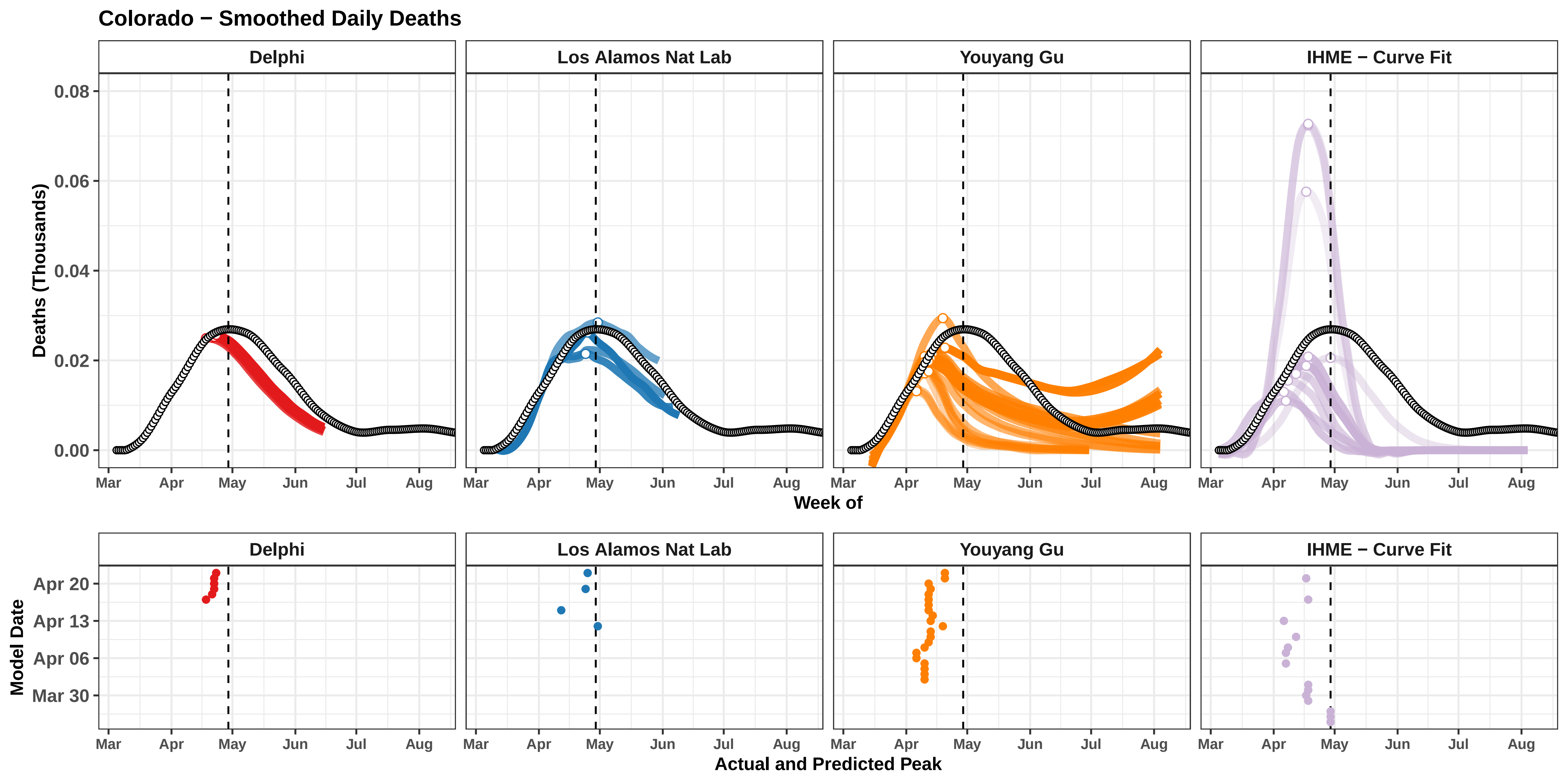
Colorado − Smoothed Daily Deaths
Delphi
Los Alamos Nat Lab
Youyang Gu
IHME − Curve Fit
Deaths (Thousands)
0.08
0.06
0.04
0.02
0.00
Mar
Apr
May
Jun
Jul
Aug
Week of
Model Date
Apr 20
Apr 13
Apr 06
Mar 30
Actual and Predicted Peak

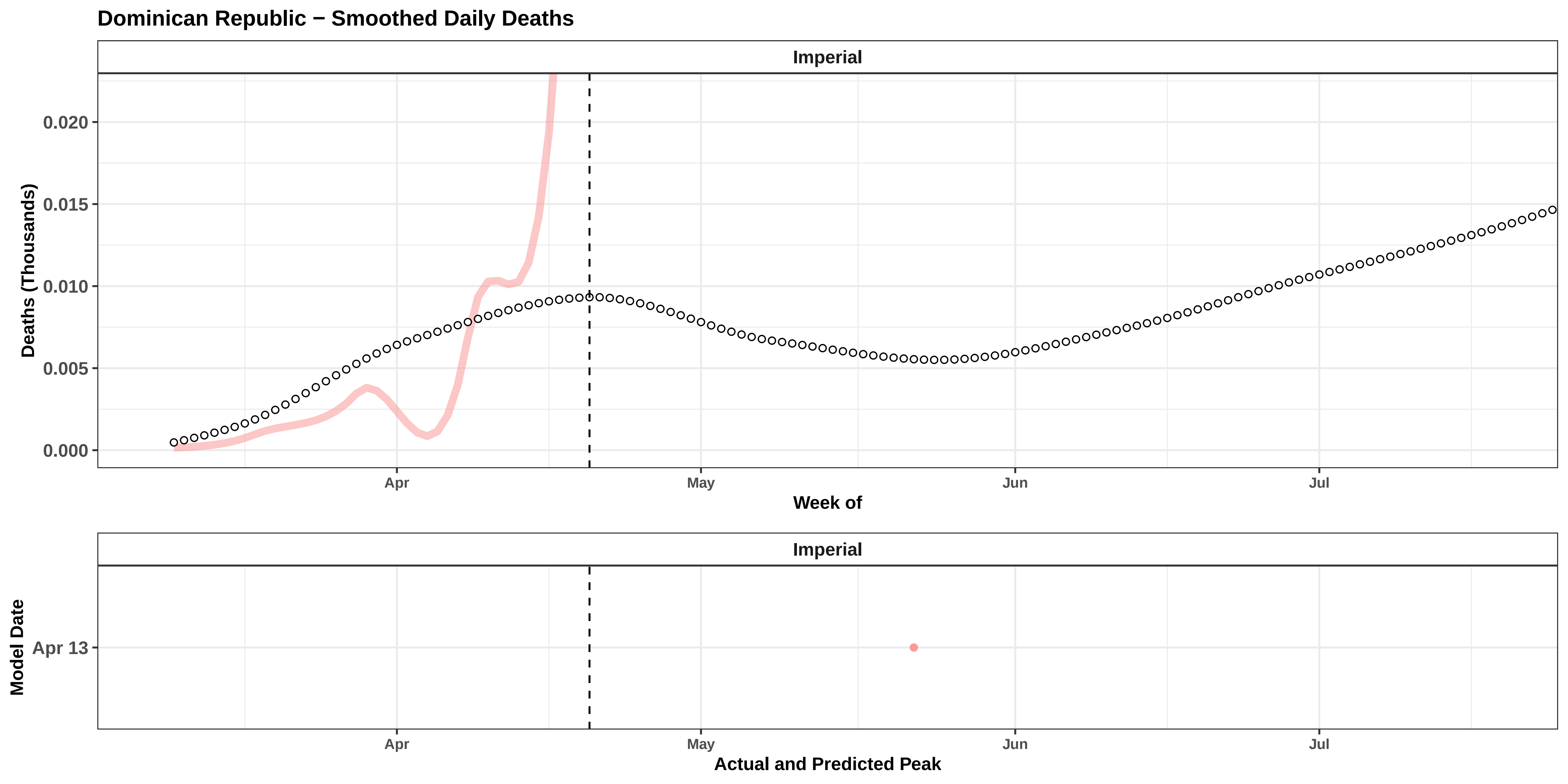

Dominican Republic − Smoothed Daily Deaths
Imperial
0.020
0.015
0.010
0.005
0.000
Deaths (Thousands)
Apr
May
Jun
Jul
Week of
Imperial
Model Date
Apr 13
Apr
May
Jun
Jul
Actual and Predicted Peak

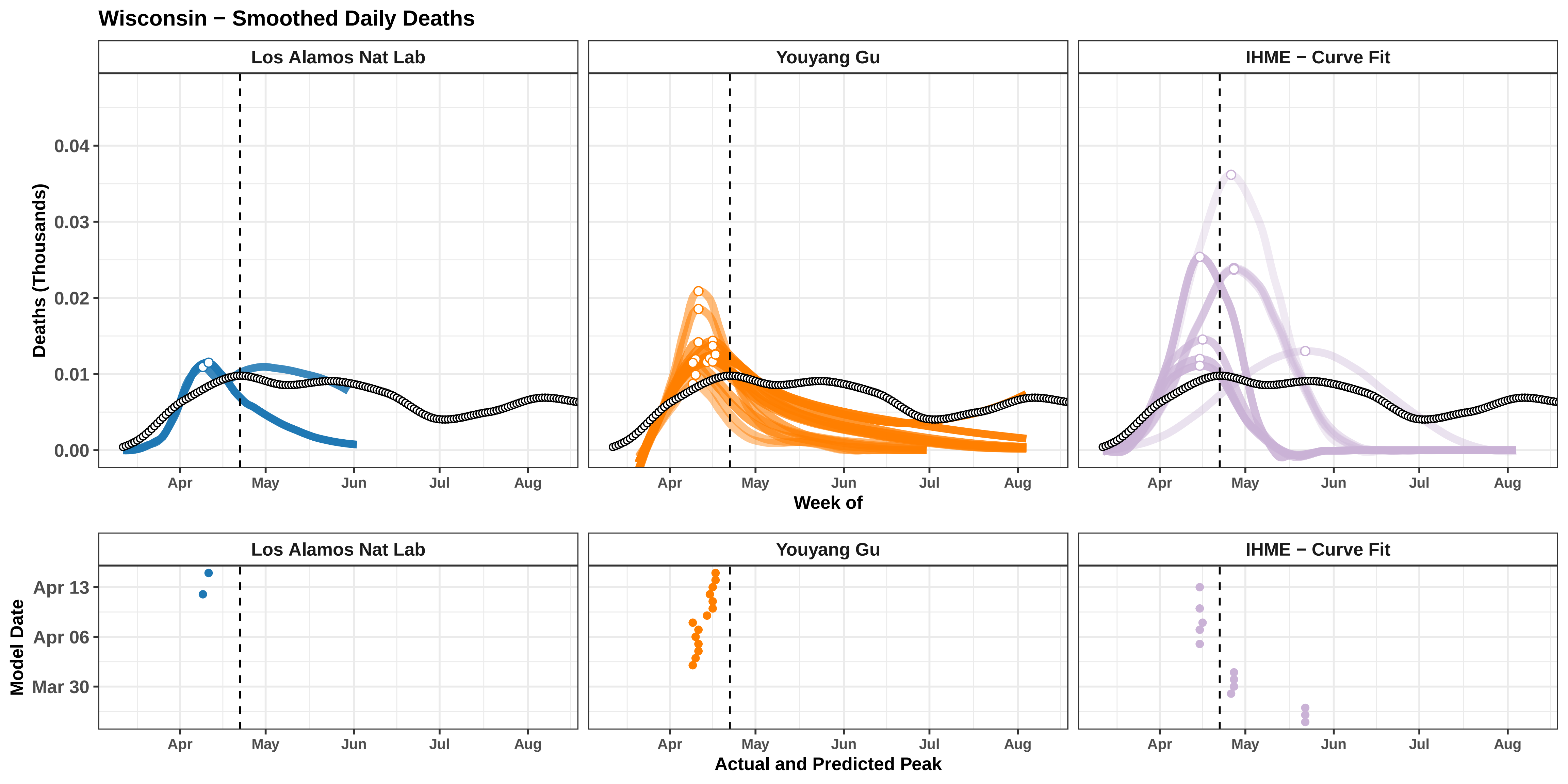

Wisconsin − Smoothed Daily Deaths
Los Alamos Nat Lab
Youyang Gu
IHME − Curve Fit
Deaths (Thousands)
0.04
0.03
0.02
0.01
0.00
Apr
May
Jun
Jul
Aug
Week of
Model Date
Apr 13
Apr 06
Mar 30
Actual and Predicted Peak

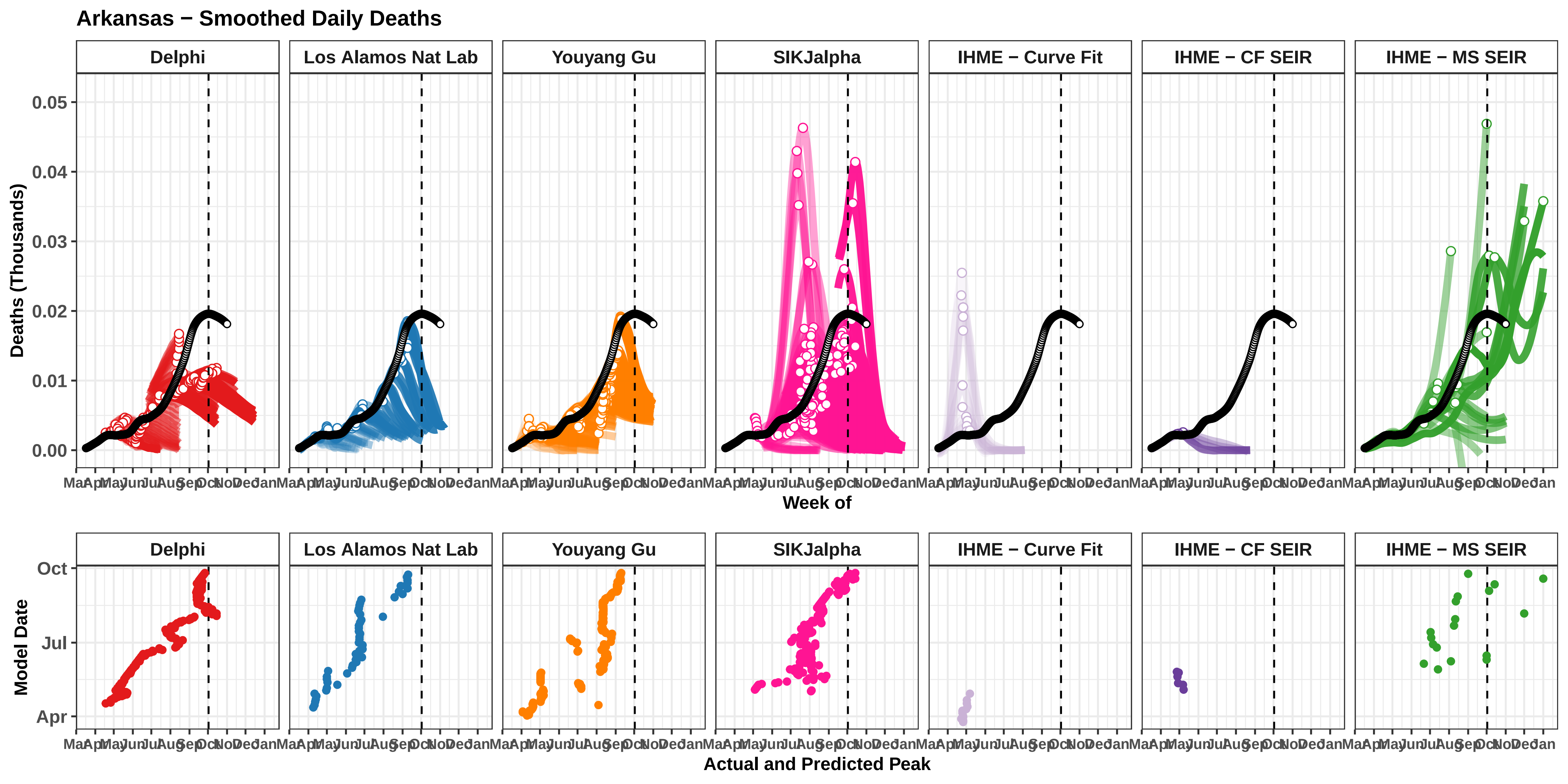

Arkansas − Smoothed Daily Deaths
Delphi
Los Alamos Nat Lab
Youyang Gu
SIKJalpha
IHME − Curve Fit
IHME − CF SEIR
IHME − MS SEIR
Deaths (Thousands)
0.05
0.04
0.03
0.02
0.01
0.00
Mar Apr May Jun Jul Aug Sep Oct Nov Dec Jan
Week of
Model Date
Oct
Jul
Apr
Actual and Predicted Peak

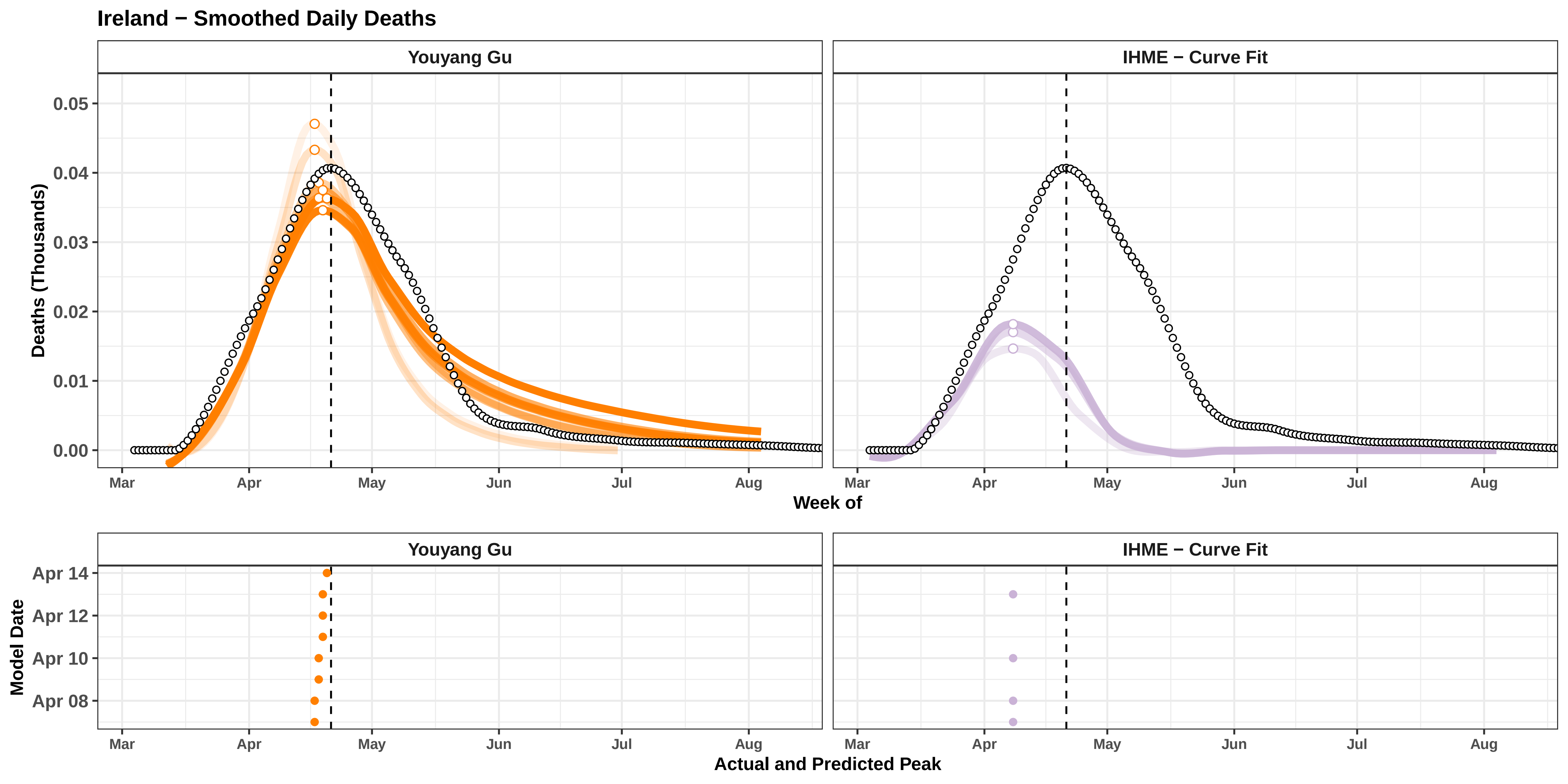
Ireland − Smoothed Daily Deaths
Youyang Gu
IHME − Curve Fit
Deaths (Thousands)
0.05
0.04
0.03
0.02
0.01
0.00
Mar
Apr
May
Jun
Jul
Aug
Week of
Model Date
Apr 14
Apr 12
Apr 10
Apr 08
Actual and Predicted Peak

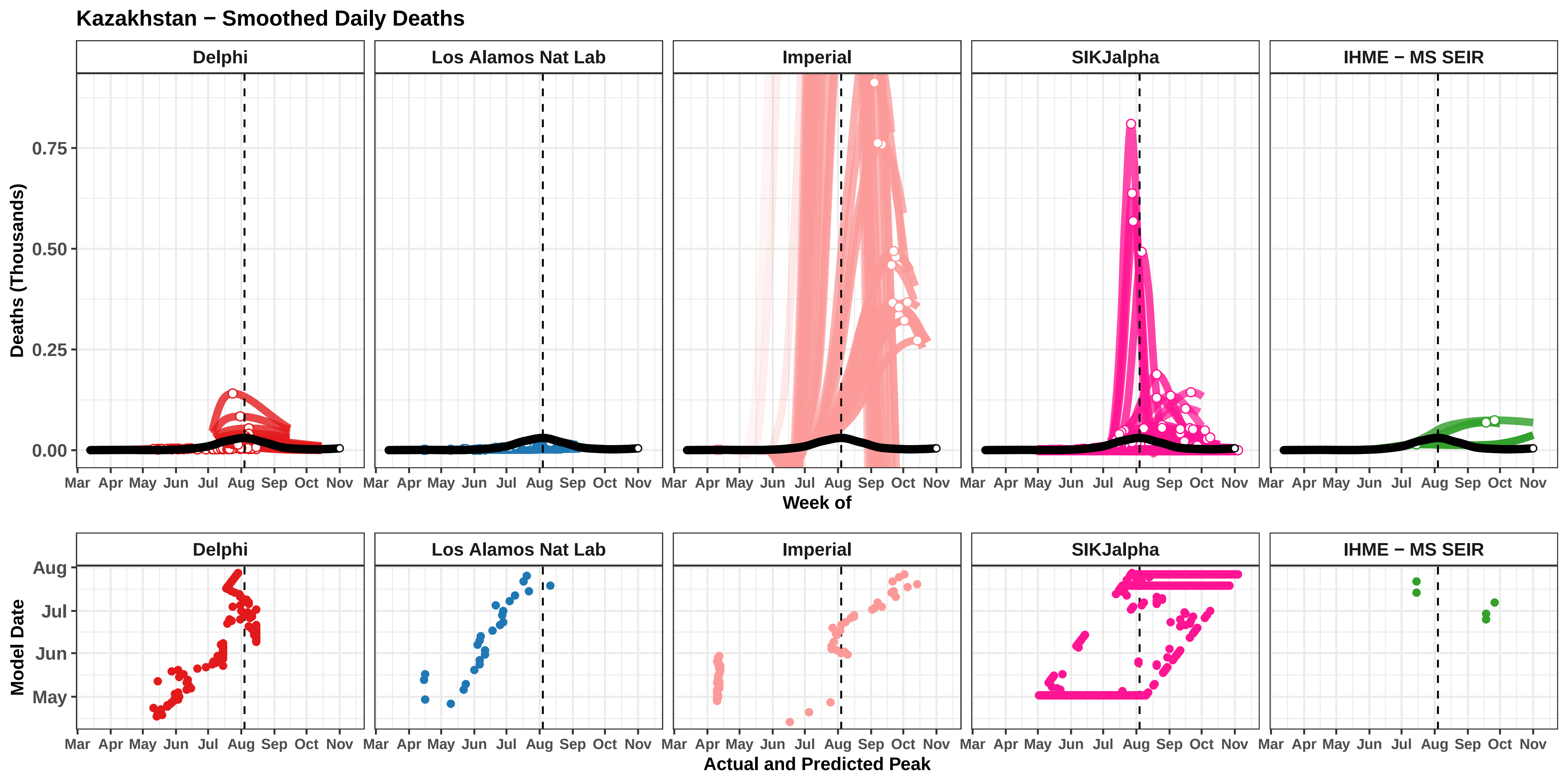

Kazakhstan − Smoothed Daily Deaths
Delphi
Los Alamos Nat Lab
Imperial
SIKJalpha
IHME − MS SEIR
Deaths (Thousands)
0.75
0.50
0.25
0.00
Mar Apr May Jun Jul Aug Sep Oct Nov
Week of
Model Date
Aug
Jul
Jun
May
Actual and Predicted Peak

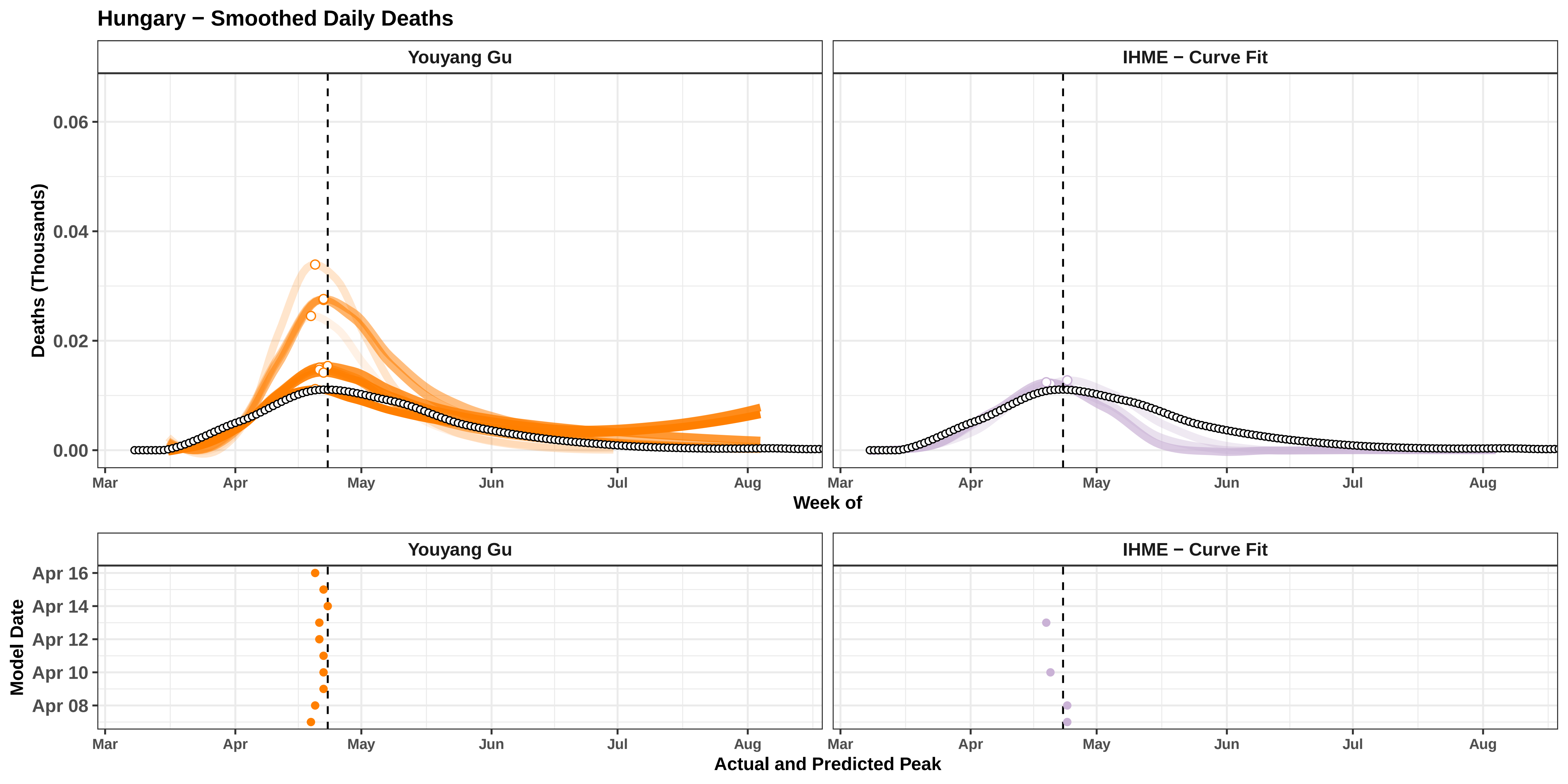

Hungary − Smoothed Daily Deaths
Youyang Gu
IHME − Curve Fit
Deaths (Thousands)
0.06
0.04
0.02
0.00
Mar
Apr
May
Jun
Jul
Aug
Week of
Model Date
Apr 16
Apr 14
Apr 12
Apr 10
Apr 08
Actual and Predicted Peak

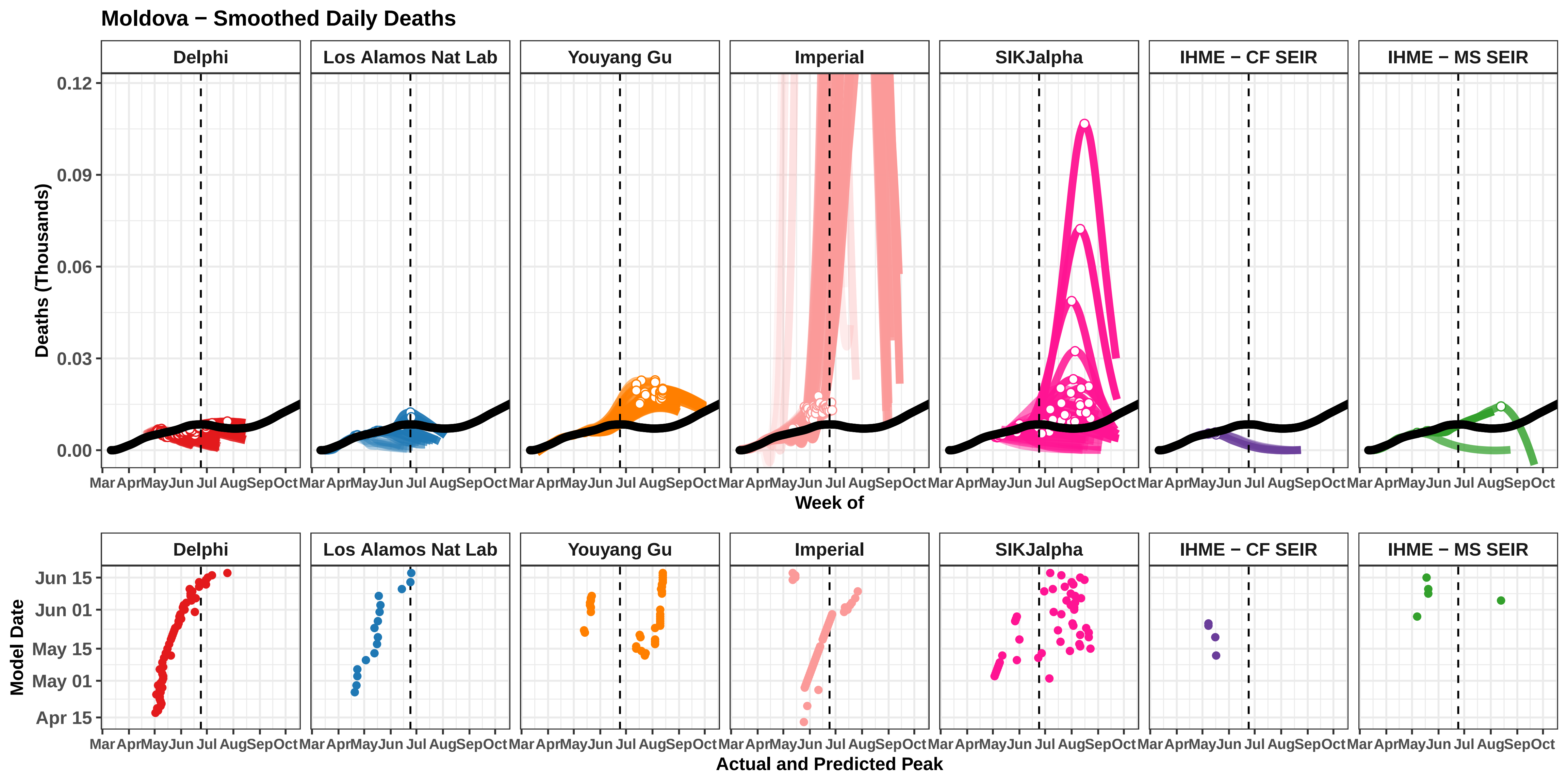
Moldova − Smoothed Daily Deaths
Delphi
Los Alamos Nat Lab
Youyang Gu
Imperial
SIKJalpha
IHME − CF SEIR
IHME − MS SEIR
Deaths (Thousands)
0.12
0.09
0.06
0.03
0.00
Mar Apr May Jun Jul Aug Sep Oct
Week of
Model Date
Jun 15
Jun 01
May 15
May 01
Apr 15
Actual and Predicted Peak

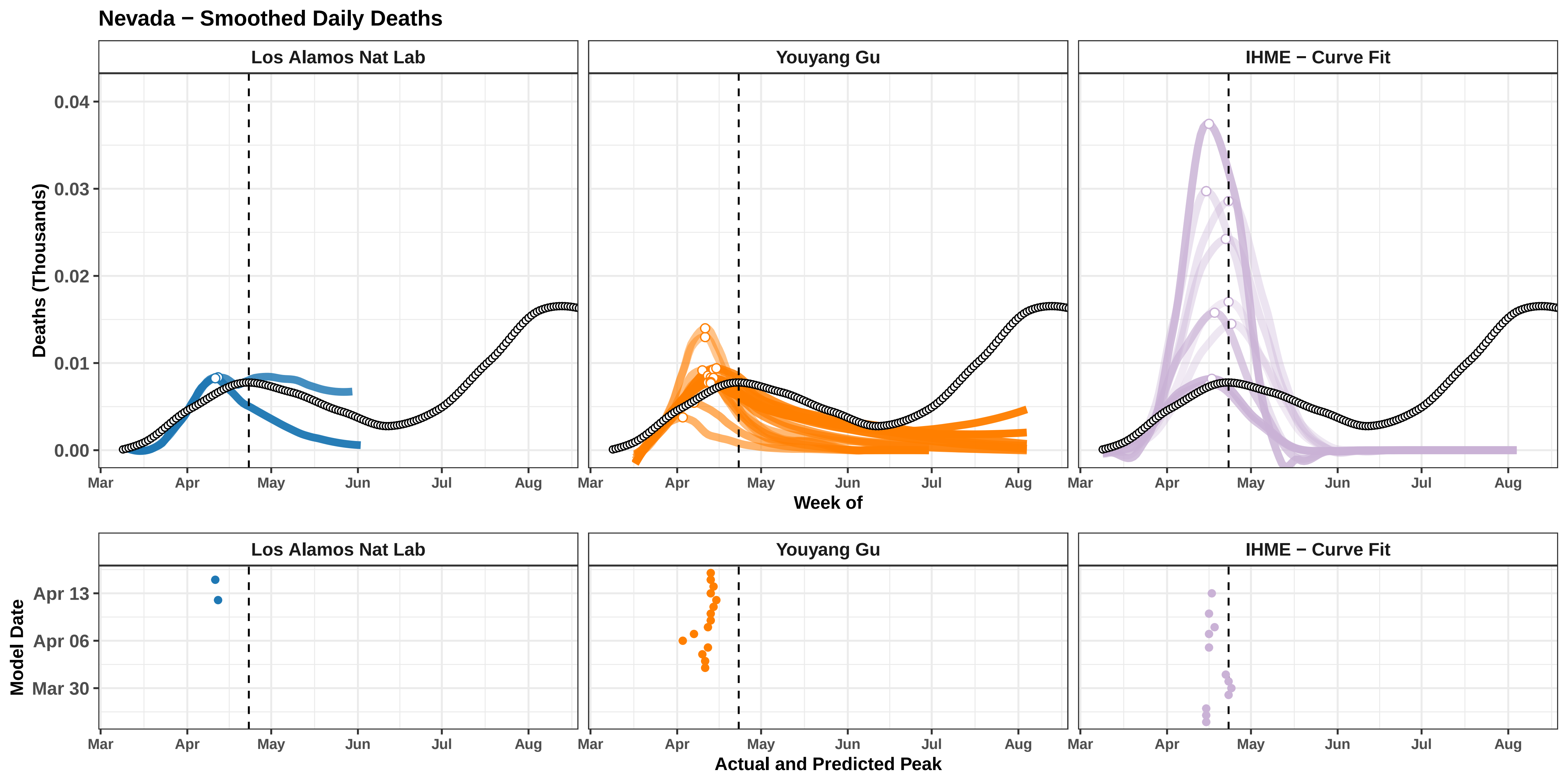

Nevada − Smoothed Daily Deaths
Los Alamos Nat Lab
Youyang Gu
IHME − Curve Fit
Deaths (Thousands)
0.04
0.03
0.02
0.01
0.00
Mar
Apr
May
Jun
Jul
Aug
Week of
Model Date
Apr 13
Apr 06
Mar 30
Actual and Predicted Peak

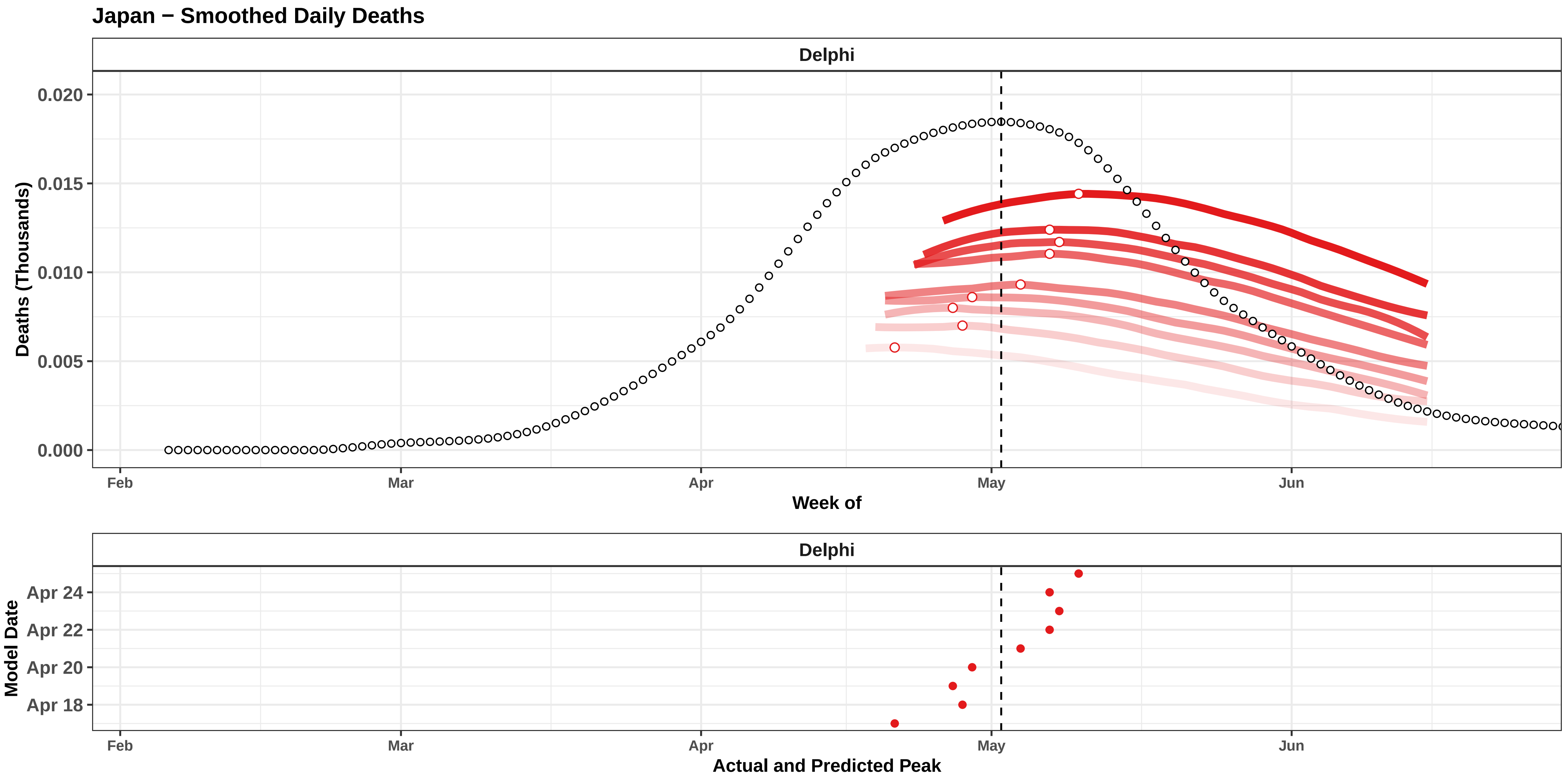
Japan − Smoothed Daily Deaths
Delphi
0.020
0.015
0.010
0.005
0.000
Deaths (Thousands)
Feb
Mar
Apr
May
Jun
Week of
Delphi
Apr 24
Apr 22
Apr 20
Apr 18
Model Date
Feb
Mar
Apr
May
Jun
Actual and Predicted Peak

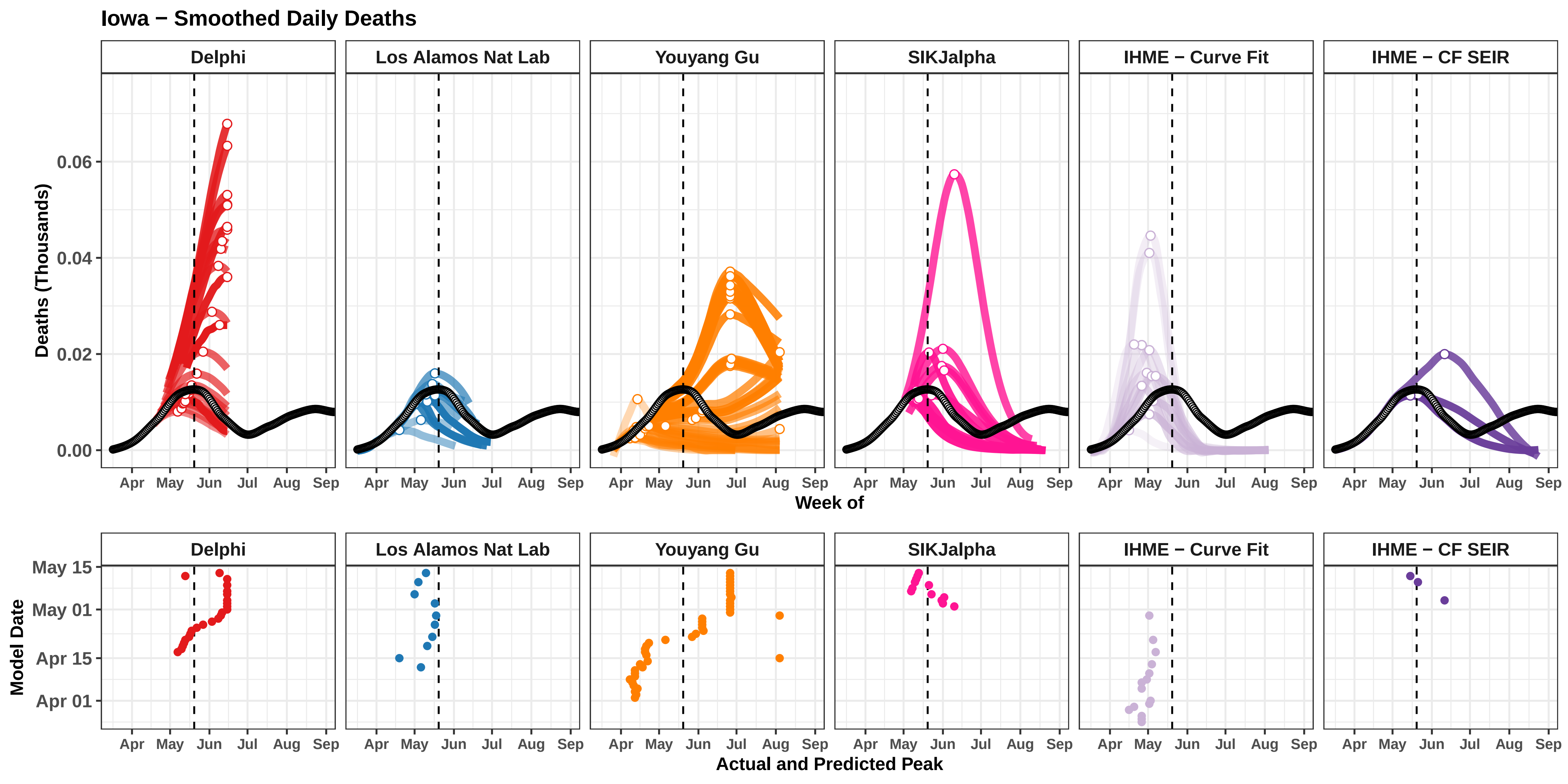

Iowa − Smoothed Daily Deaths
Delphi
Los Alamos Nat Lab
Youyang Gu
SIKJalpha
IHME − Curve Fit
IHME − CF SEIR
Deaths (Thousands)
0.00
0.02
0.04
0.06
Apr
May
Jun
Jul
Aug
Sep
Week of
Model Date
May 15
May 01
Apr 15
Apr 01
Actual and Predicted Peak

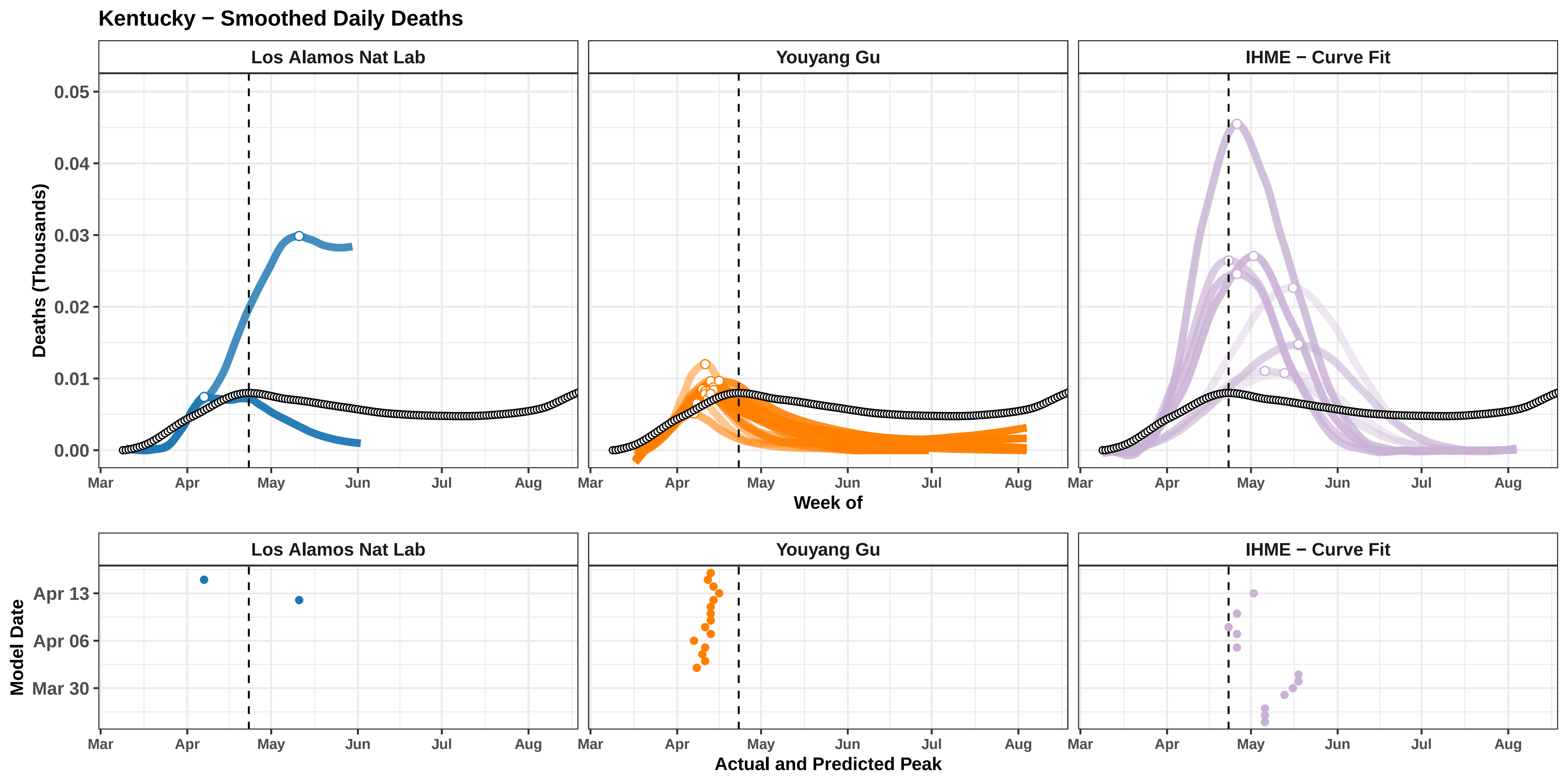

Kentucky − Smoothed Daily Deaths
Los Alamos Nat Lab
Youyang Gu
IHME − Curve Fit
Deaths (Thousands)
0.00
0.01
0.02
0.03
0.04
0.05
Mar
Apr
May
Jun
Jul
Aug
Week of
Model Date
Mar 30
Apr 06
Apr 13
Actual and Predicted Peak

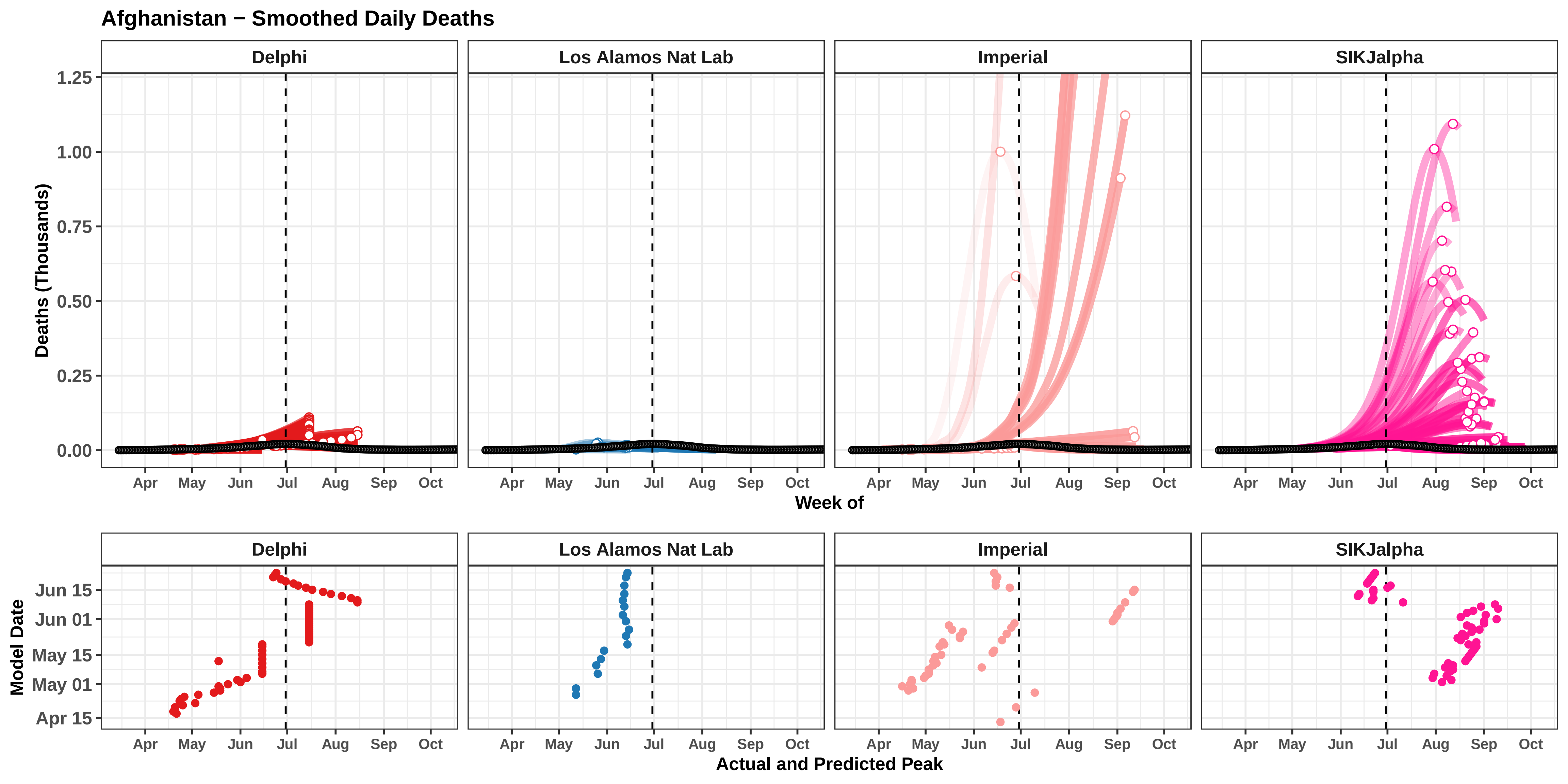

Afghanistan − Smoothed Daily Deaths
Delphi
Los Alamos Nat Lab
Imperial
SIKJalpha
Deaths (Thousands)
1.25
1.00
0.75
0.50
0.25
0.00
Apr
May
Jun
Jul
Aug
Sep
Oct
Week of
Model Date
Jun 15
Jun 01
May 15
May 01
Apr 15
Actual and Predicted Peak

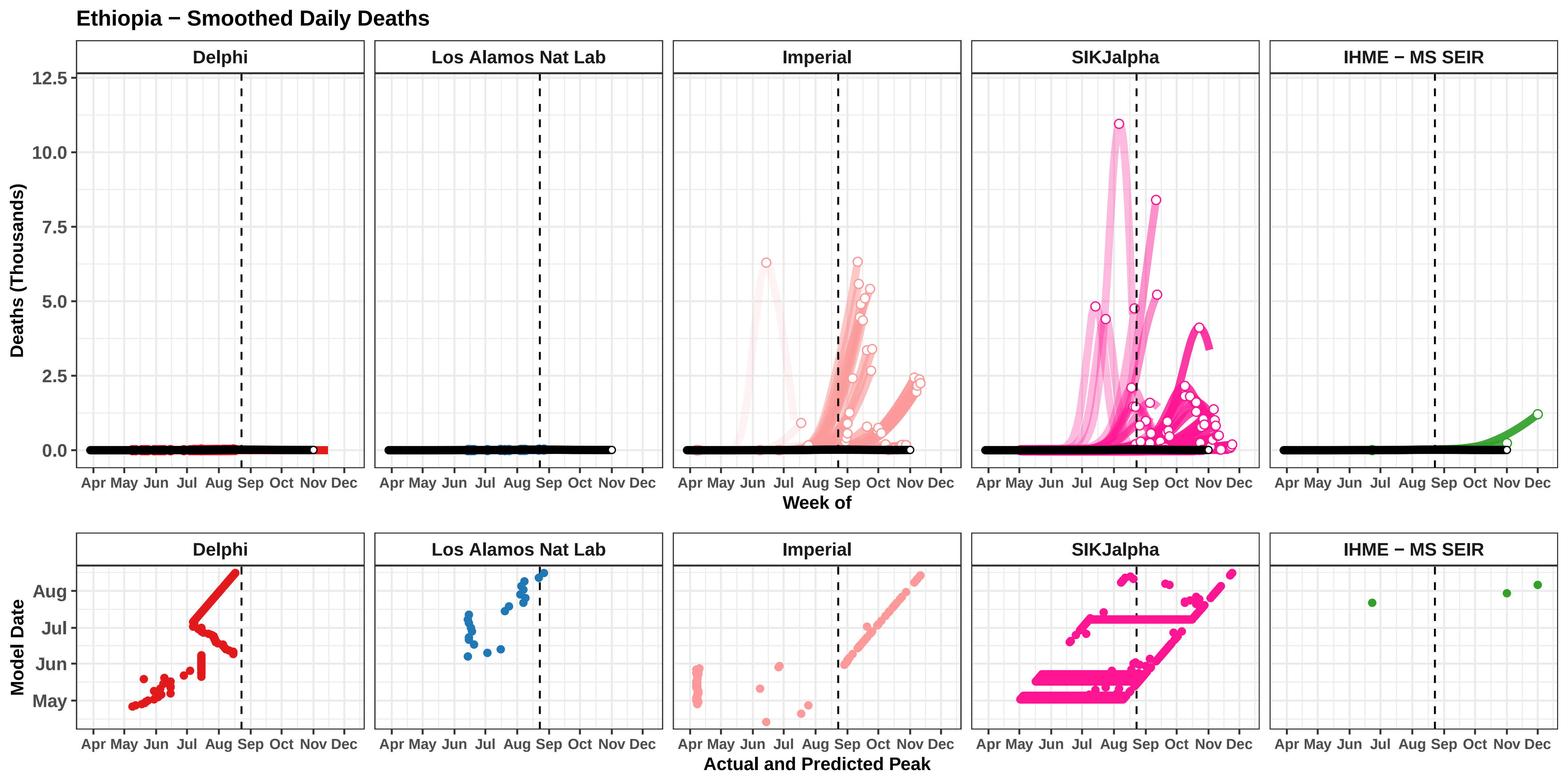

Ethiopia − Smoothed Daily Deaths
Delphi
Los Alamos Nat Lab
Imperial
SIKJalpha
IHME − MS SEIR
Deaths (Thousands)
12.5
10.0
7.5
5.0
2.5
0.0
Apr May Jun Jul Aug Sep Oct Nov Dec
Week of
Model Date
Aug
Jul
Jun
May
Actual and Predicted Peak

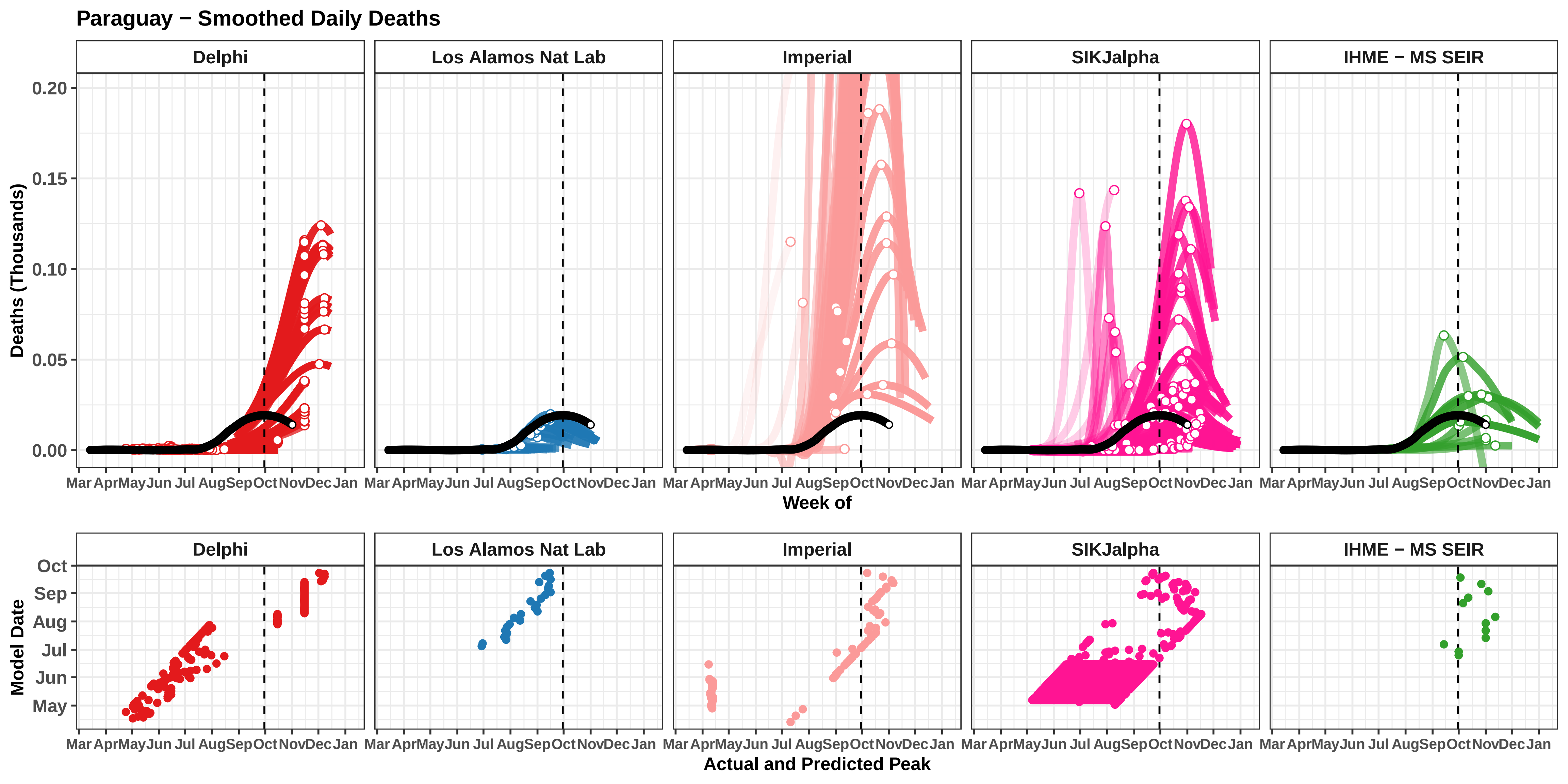
Paraguay − Smoothed Daily Deaths
Delphi
Los Alamos Nat Lab
Imperial
SIKJalpha
IHME − MS SEIR
Deaths (Thousands)
0.00
0.05
0.10
0.15
0.20
Mar Apr May Jun Jul Aug Sep Oct Nov Dec Jan
Week of
Model Date
May
Jun
Jul
Aug
Sep
Oct
Actual and Predicted Peak

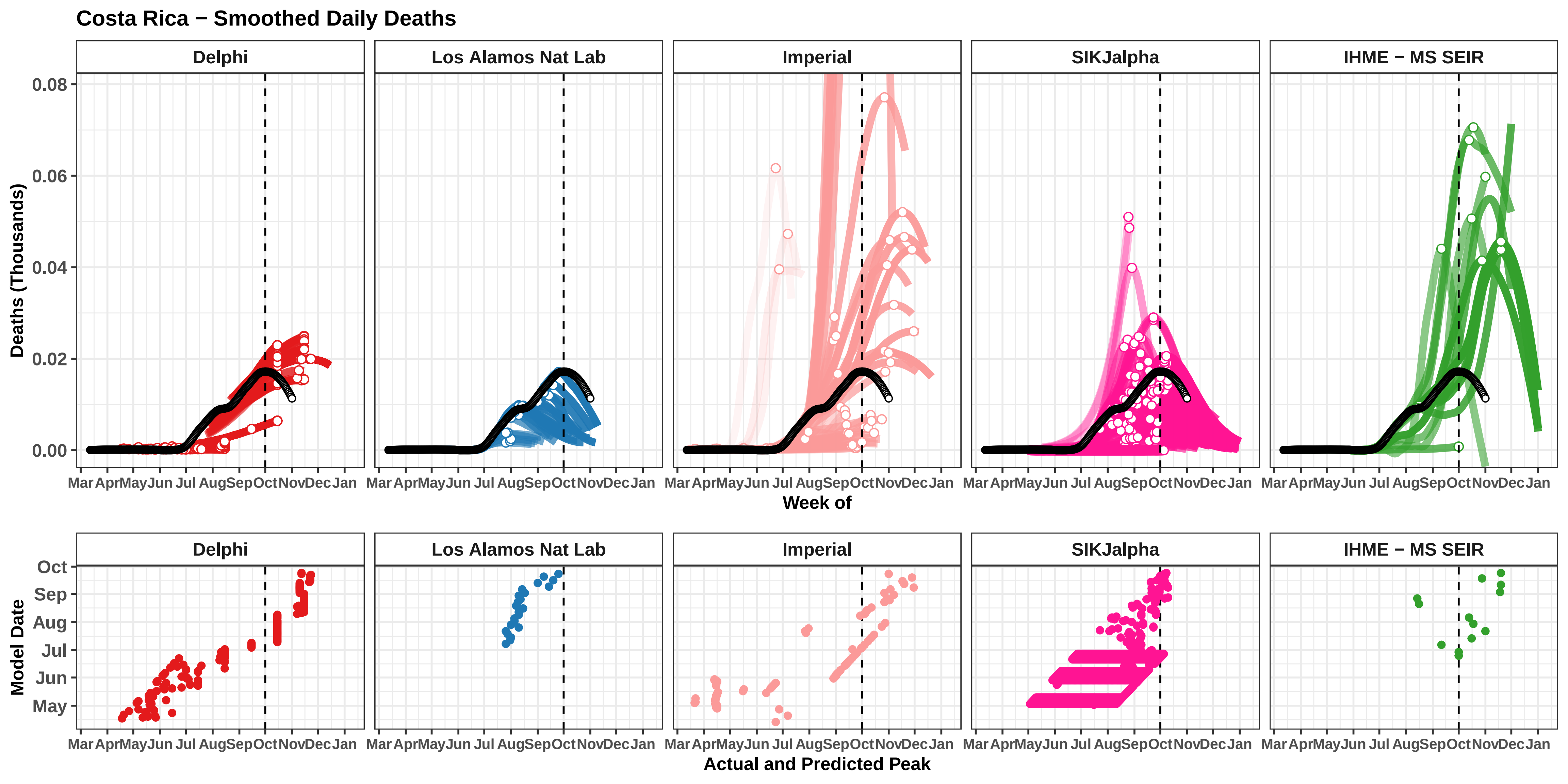

Costa Rica − Smoothed Daily Deaths
Delphi
Los Alamos Nat Lab
Imperial
SIKJalpha
IHME − MS SEIR
Deaths (Thousands)
0.08
0.06
0.04
0.02
0.00
Mar Apr May Jun Jul Aug Sep Oct Nov Dec Jan
Week of
Model Date
Oct
Sep
Aug
Jul
Jun
May
Actual and Predicted Peak

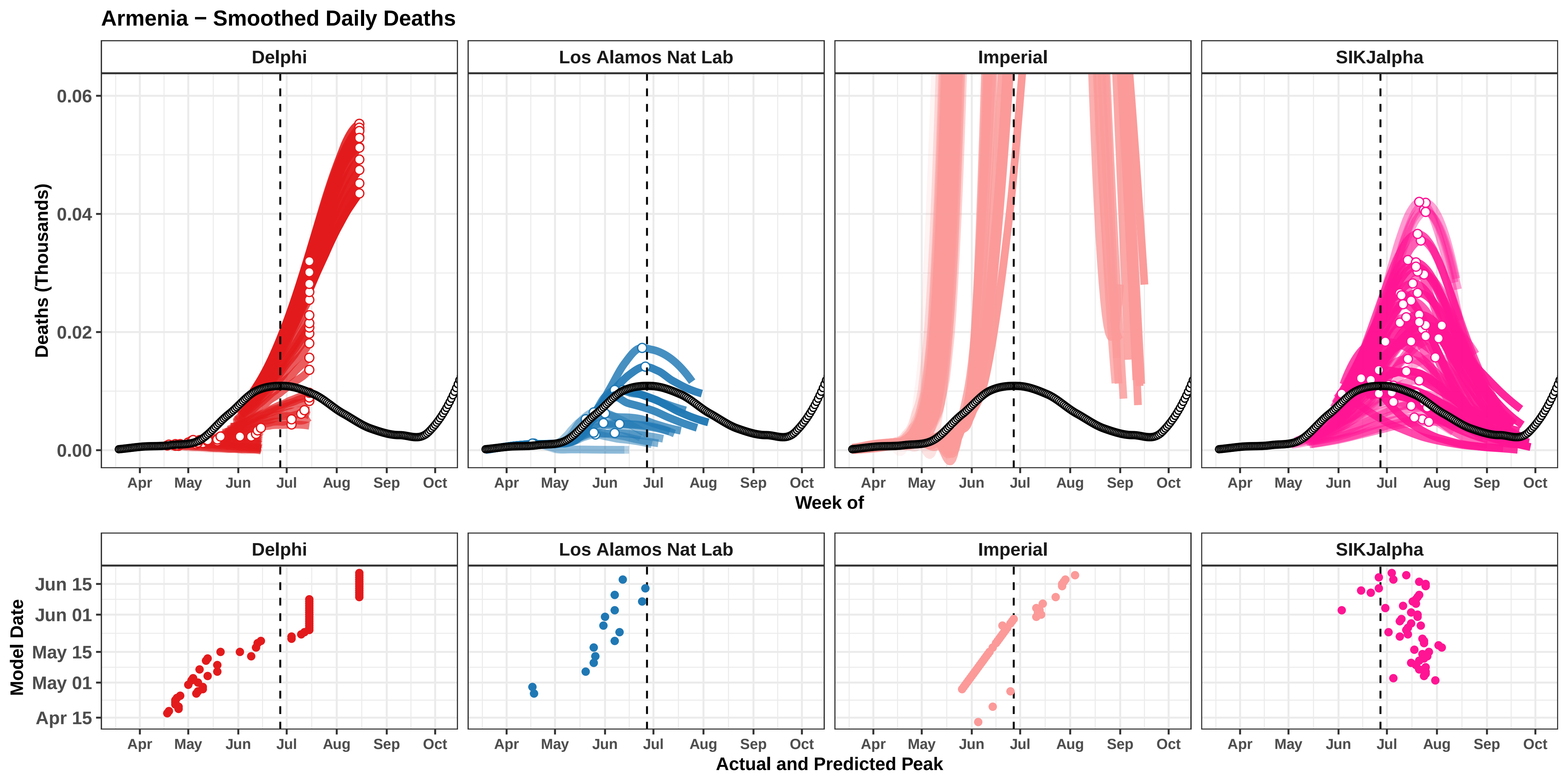

Armenia − Smoothed Daily Deaths
Delphi
Los Alamos Nat Lab
Imperial
SIKJalpha
Deaths (Thousands)
0.06
0.04
0.02
0.00
Apr
May
Jun
Jul
Aug
Sep
Oct
Week of
Model Date
Jun 15
Jun 01
May 15
May 01
Apr 15
Actual and Predicted Peak

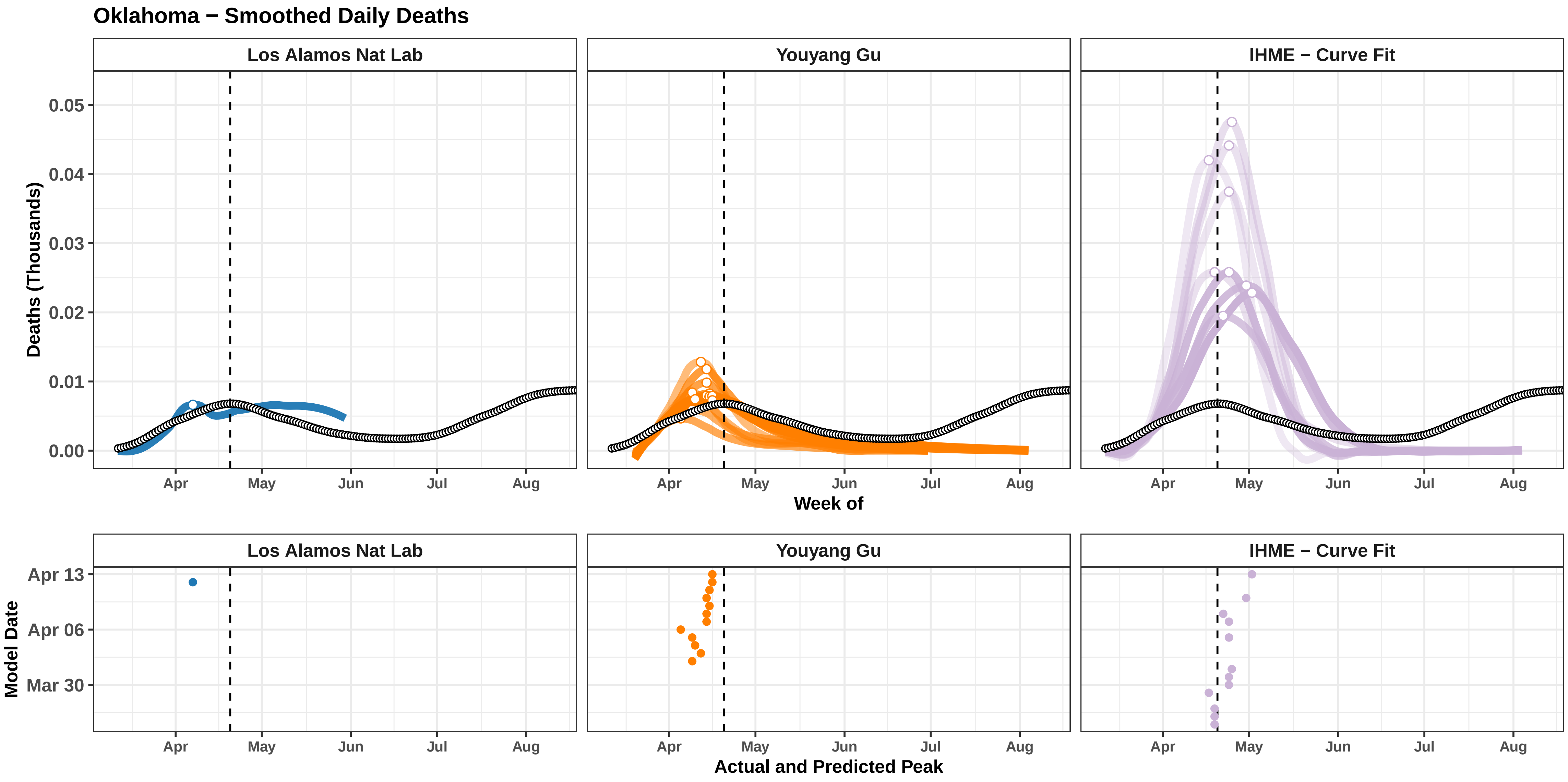

Oklahoma − Smoothed Daily Deaths
Los Alamos Nat Lab
Youyang Gu
IHME − Curve Fit
Deaths (Thousands)
0.05
0.04
0.03
0.02
0.01
0.00
Apr
May
Jun
Jul
Aug
Week of
Model Date
Apr 13
Apr 06
Mar 30
Actual and Predicted Peak

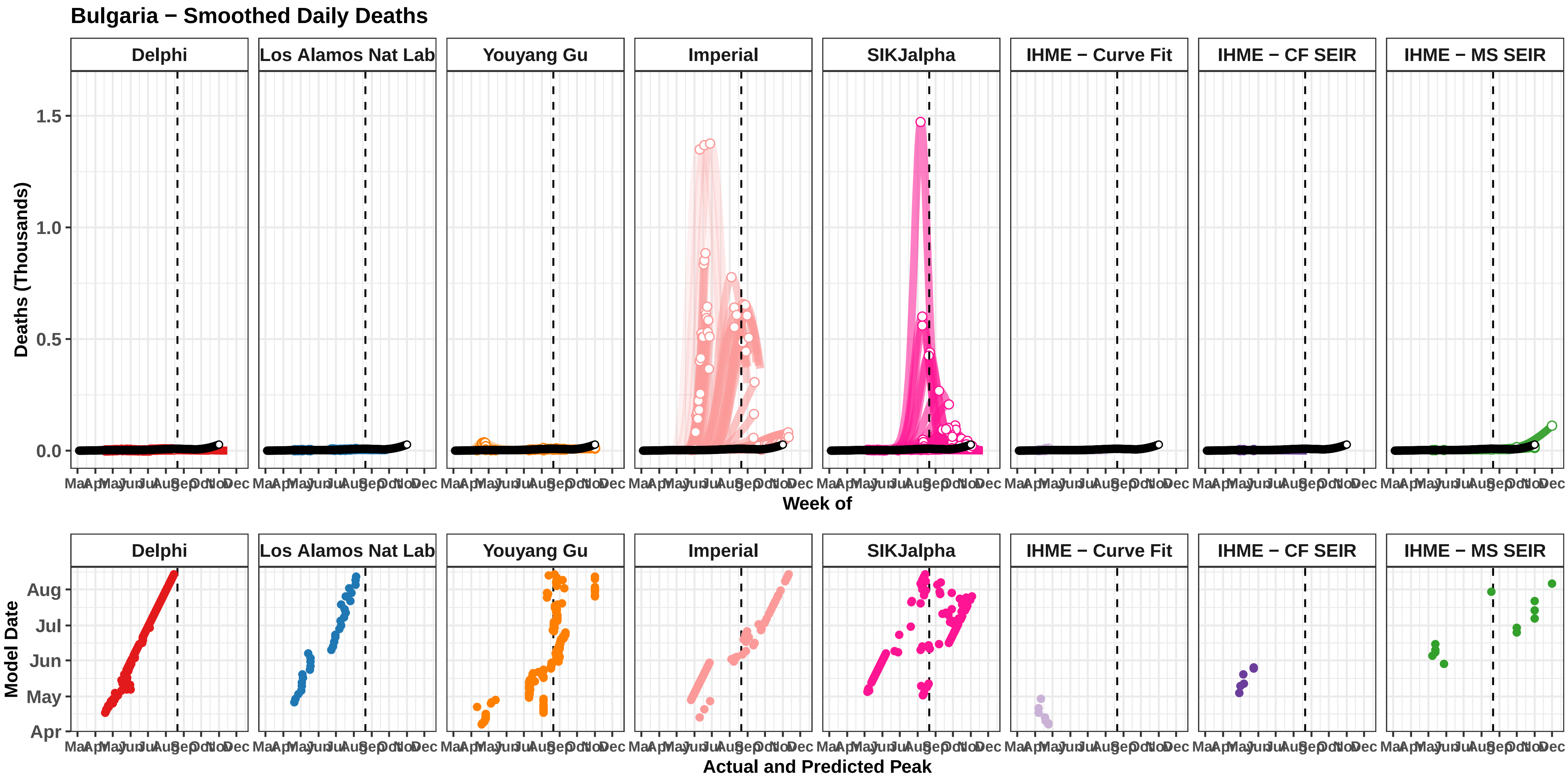

Bulgaria − Smoothed Daily Deaths
Delphi
Los Alamos Nat Lab
Youyang Gu
Imperial
SIKJalpha
IHME − Curve Fit
IHME − CF SEIR
IHME − MS SEIR
Deaths (Thousands)
0.0
0.5
1.0
1.5
Mar Apr May Jun Jul Aug Sep Oct Nov Dec
Week of
Model Date
Apr
May
Jun
Jul
Aug
Actual and Predicted Peak

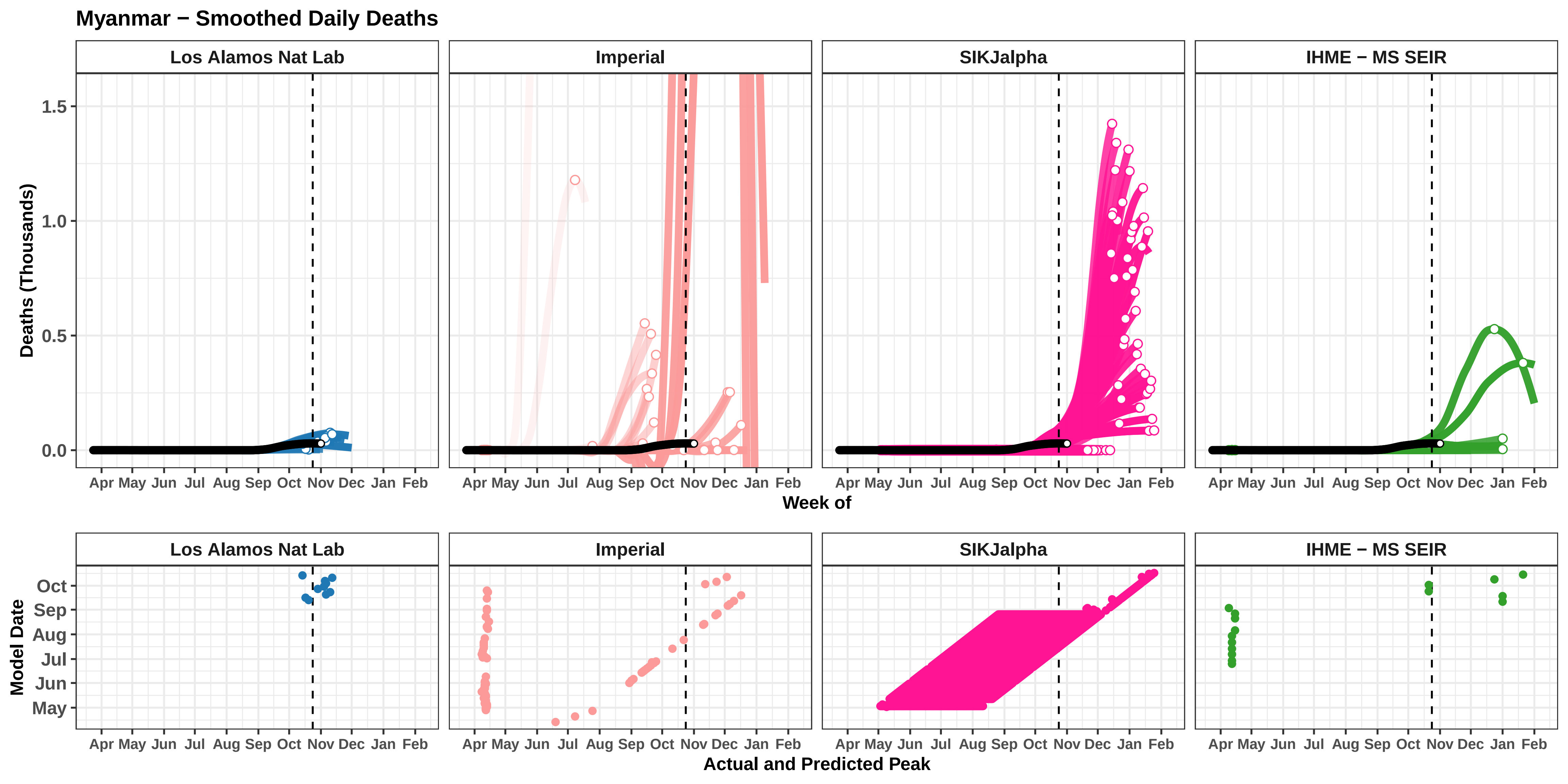

Myanmar − Smoothed Daily Deaths
Los Alamos Nat Lab
Imperial
SIKJalpha
IHME − MS SEIR
Deaths (Thousands)
0.0
0.5
1.0
1.5
Apr May Jun Jul Aug Sep Oct Nov Dec Jan Feb
Week of
Model Date
May
Jun
Jul
Aug
Sep
Oct
Actual and Predicted Peak

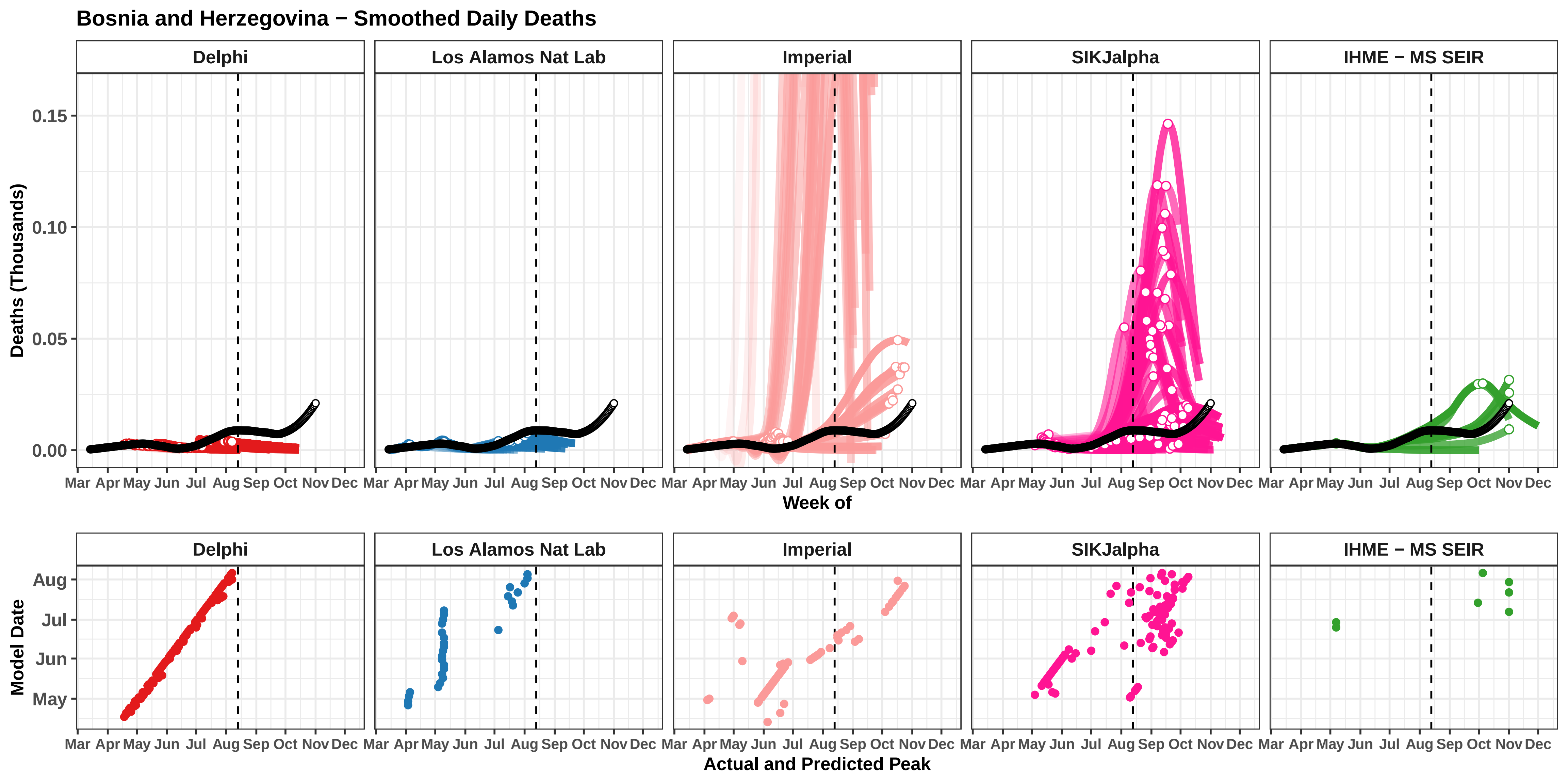

Bosnia and Herzegovina − Smoothed Daily Deaths
Delphi
Los Alamos Nat Lab
Imperial
SIKJalpha
IHME − MS SEIR
Deaths (Thousands)
0.15
0.10
0.05
0.00
Mar Apr May Jun Jul Aug Sep Oct Nov Dec
Week of
Model Date
Aug
Jul
Jun
May
Actual and Predicted Peak

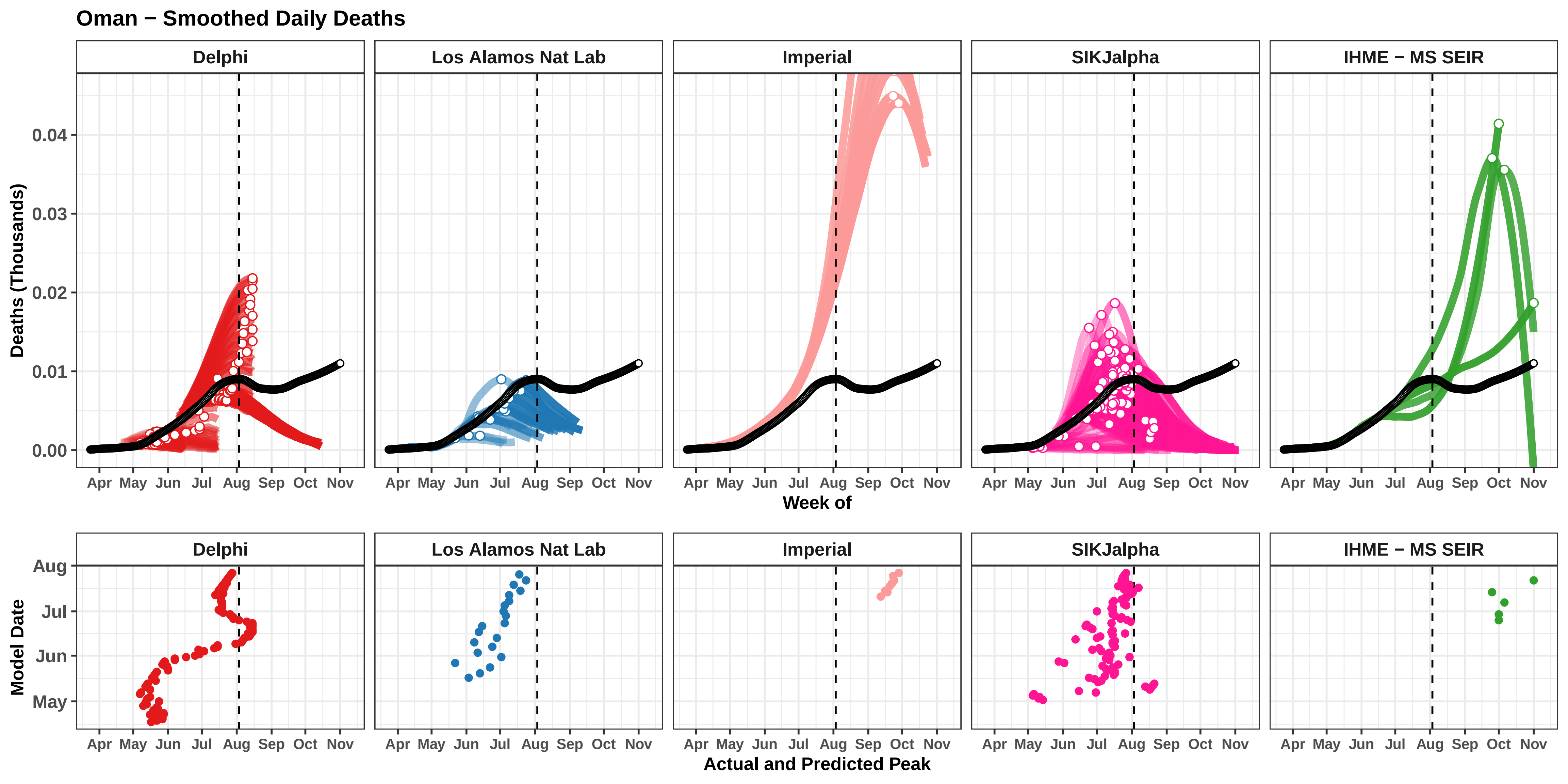

Oman − Smoothed Daily Deaths
Delphi
Los Alamos Nat Lab
Imperial
SIKJalpha
IHME − MS SEIR
Deaths (Thousands)
0.00
0.01
0.02
0.03
0.04
Apr May Jun Jul Aug Sep Oct Nov
Week of
Model Date
Aug
Jul
Jun
May
Actual and Predicted Peak

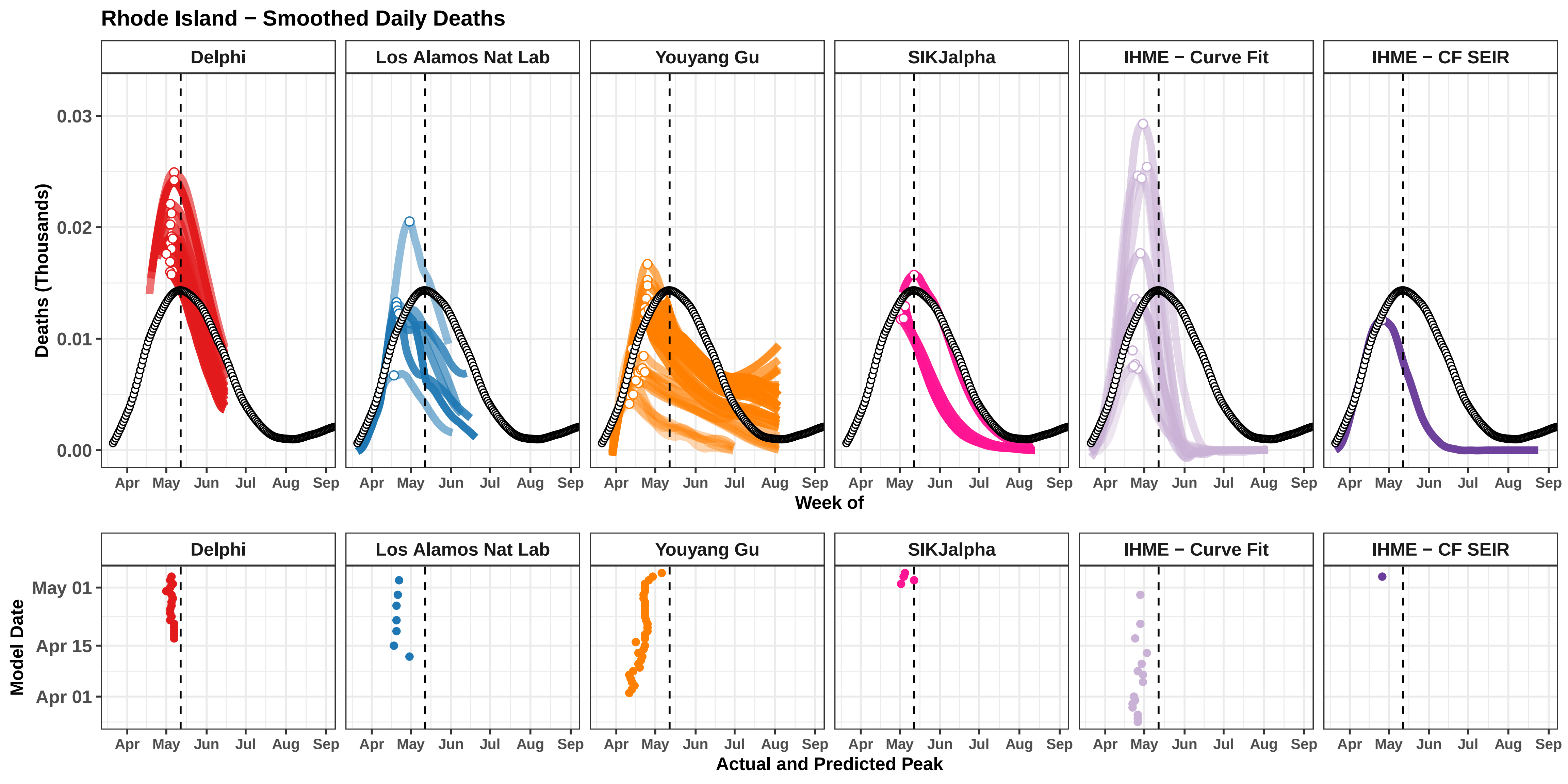

Rhode Island − Smoothed Daily Deaths
Delphi
Los Alamos Nat Lab
Youyang Gu
SIKJalpha
IHME − Curve Fit
IHME − CF SEIR
Deaths (Thousands)
0.03
0.02
0.01
0.00
Apr May Jun Jul Aug Sep
Week of
Model Date
May 01
Apr 15
Apr 01
Actual and Predicted Peak

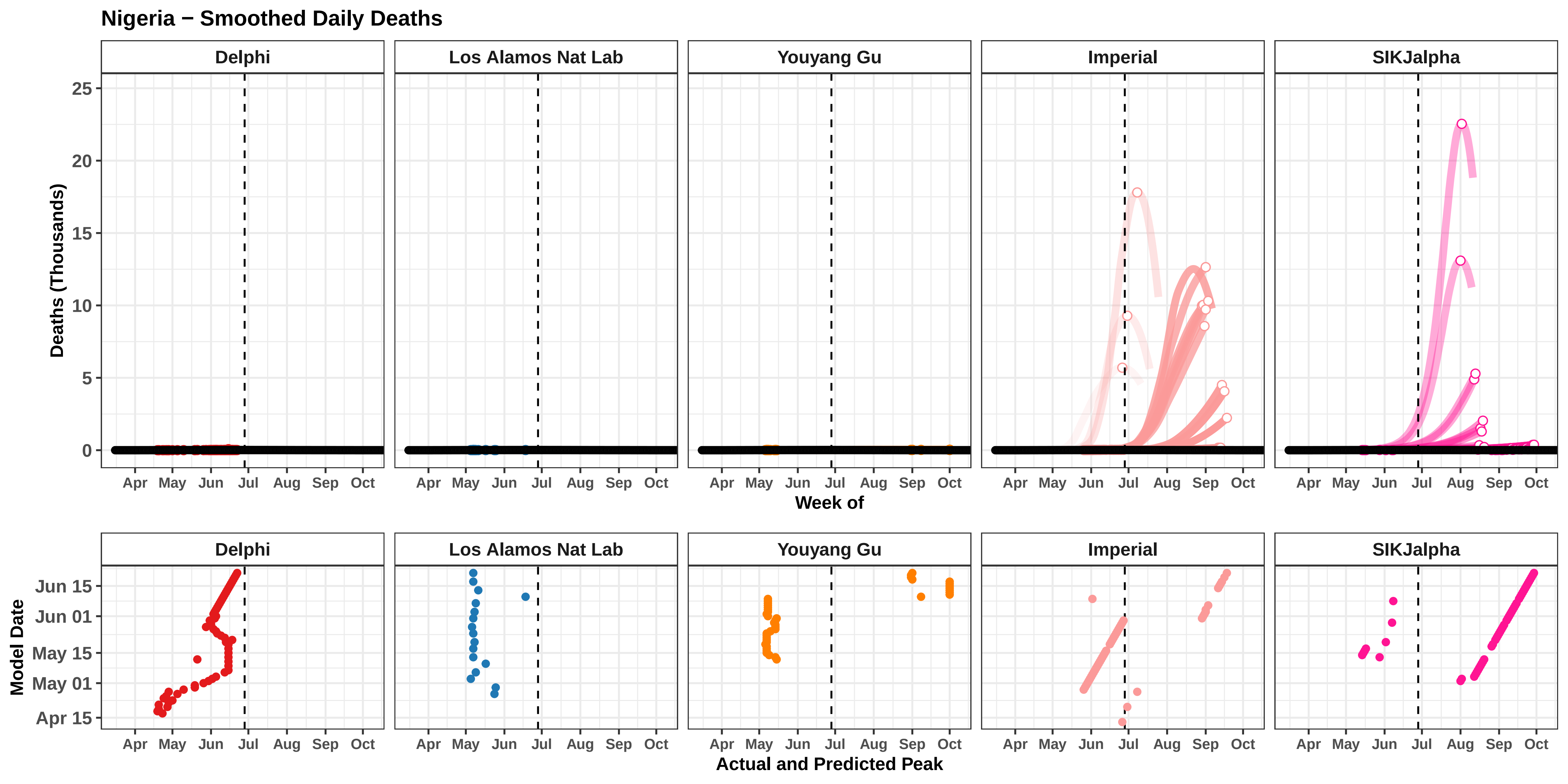

Nigeria − Smoothed Daily Deaths
Delphi
Los Alamos Nat Lab
Youyang Gu
Imperial
SIKJalpha
Deaths (Thousands)
25
20
15
10
5
0
Apr
May
Jun
Jul
Aug
Sep
Oct
Week of
Model Date
Jun 15
Jun 01
May 15
May 01
Apr 15
Actual and Predicted Peak

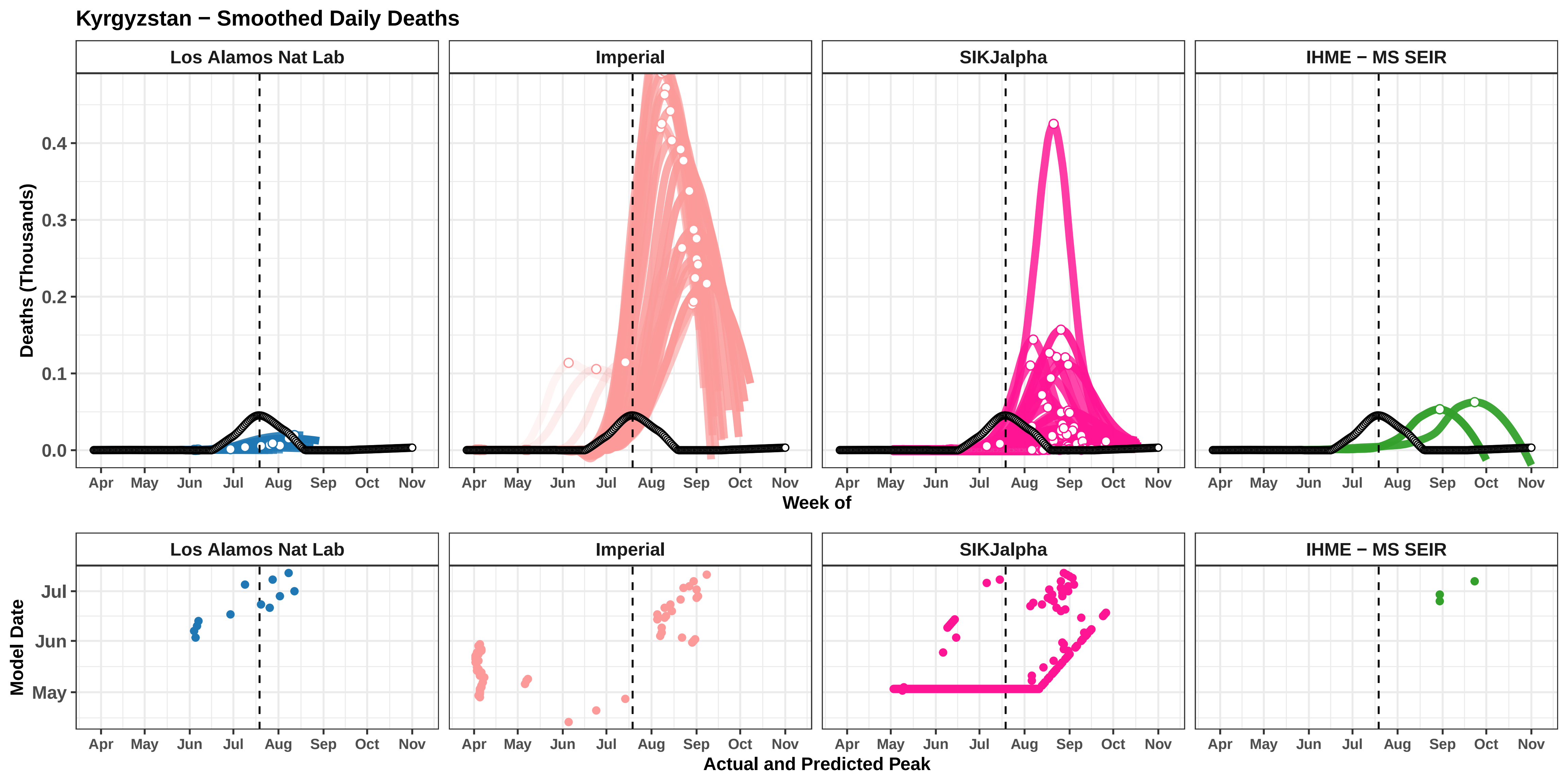

Kyrgyzstan − Smoothed Daily Deaths
Los Alamos Nat Lab
Imperial
SIKJalpha
IHME − MS SEIR
Deaths (Thousands)
0.4
0.3
0.2
0.1
0.0
Apr
May
Jun
Jul
Aug
Sep
Oct
Nov
Week of
Model Date
Jul
Jun
May
Actual and Predicted Peak

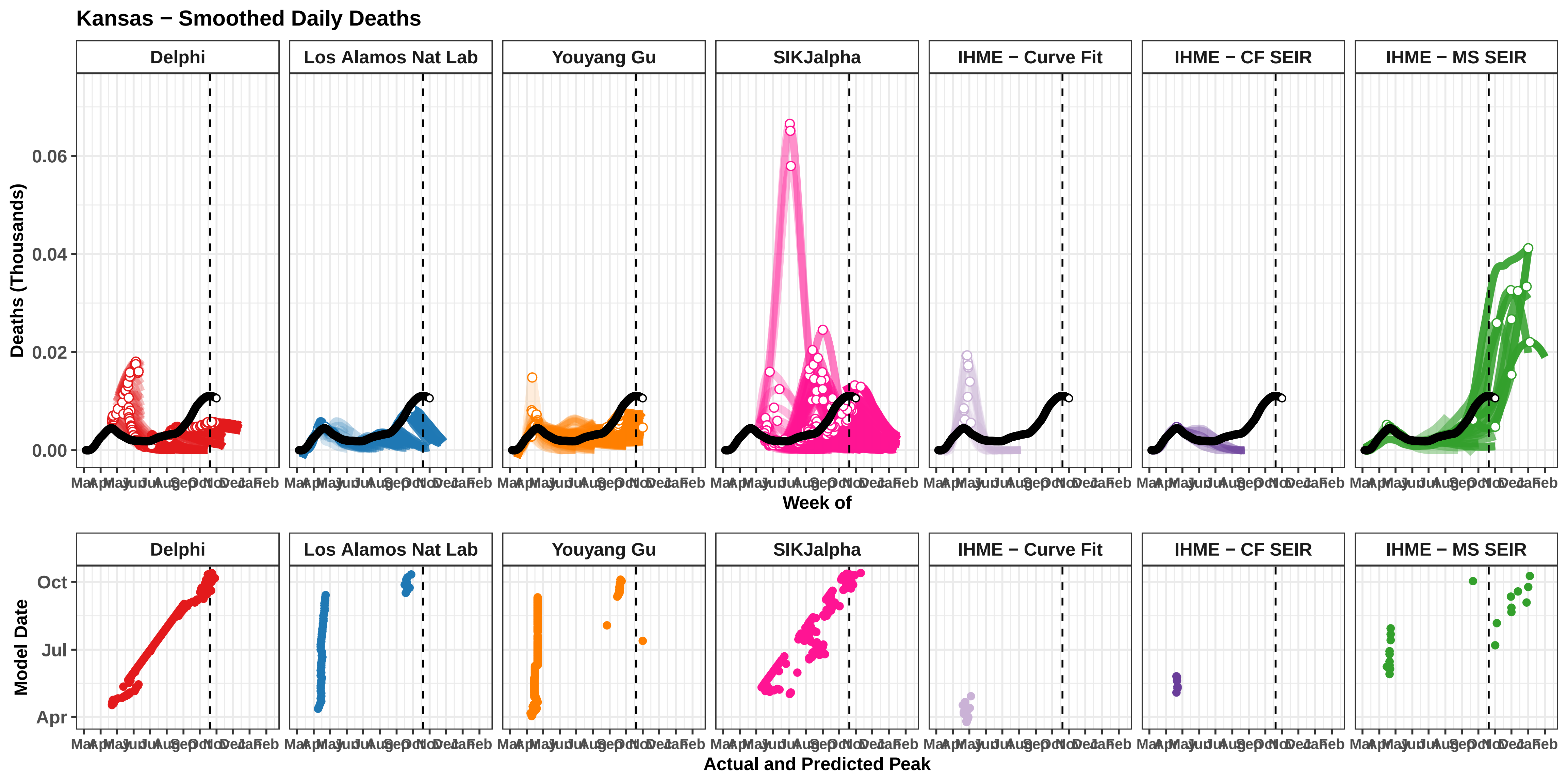

Kansas − Smoothed Daily Deaths
Delphi
Los Alamos Nat Lab
Youyang Gu
SIKJalpha
IHME − Curve Fit
IHME − CF SEIR
IHME − MS SEIR
Deaths (Thousands)
0.06
0.04
0.02
0.00
Mar Apr May Jun Jul Aug Sep Oct Nov Dec Jan Feb
Week of
Model Date
Oct
Jul
Apr
Actual and Predicted Peak

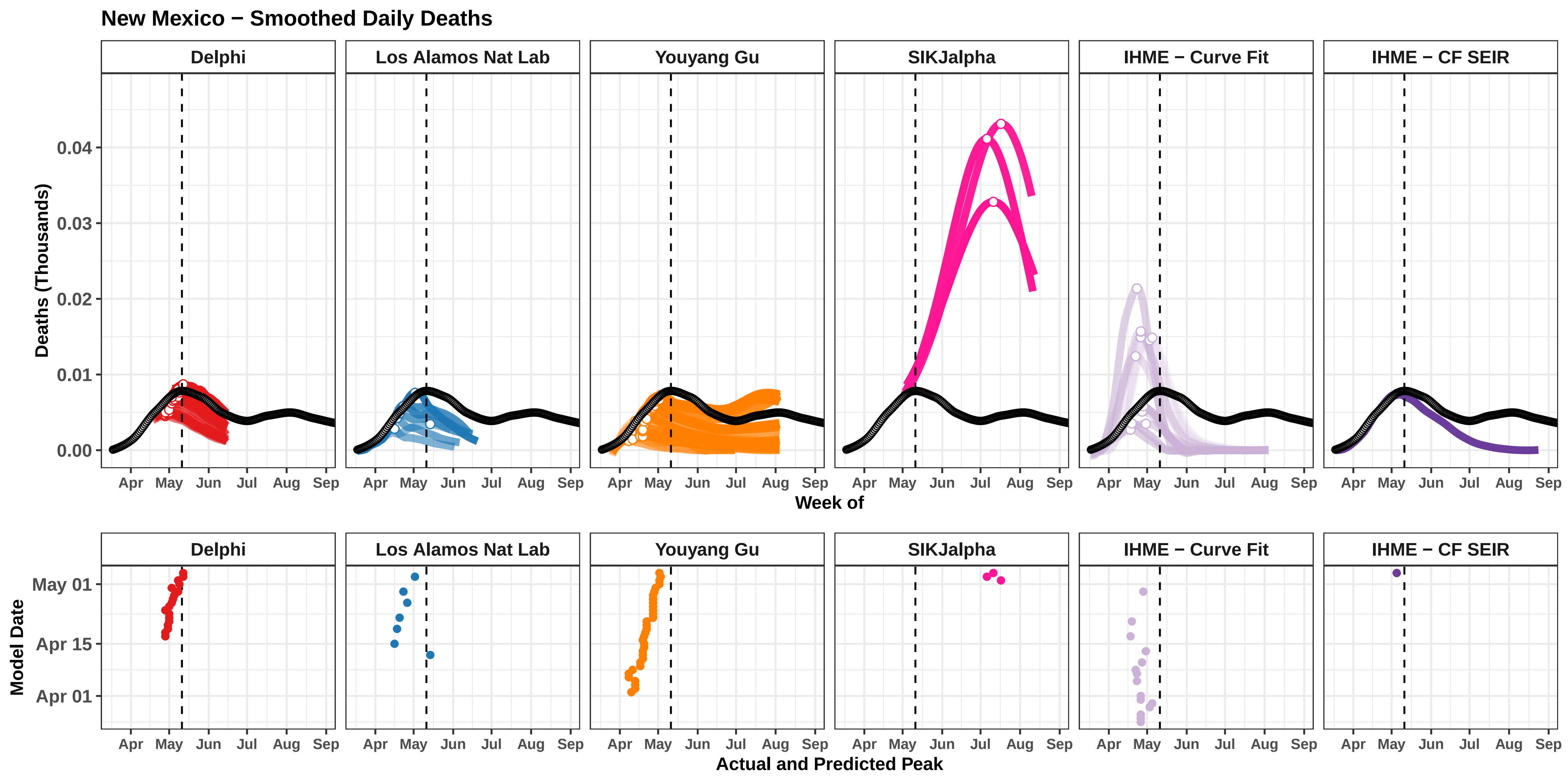

New Mexico − Smoothed Daily Deaths
Delphi
Los Alamos Nat Lab
Youyang Gu
SIKJalpha
IHME − Curve Fit
IHME − CF SEIR
Deaths (Thousands)
0.04
0.03
0.02
0.01
0.00
Apr
May
Jun
Jul
Aug
Sep
Week of
Model Date
May 01
Apr 15
Apr 01
Actual and Predicted Peak

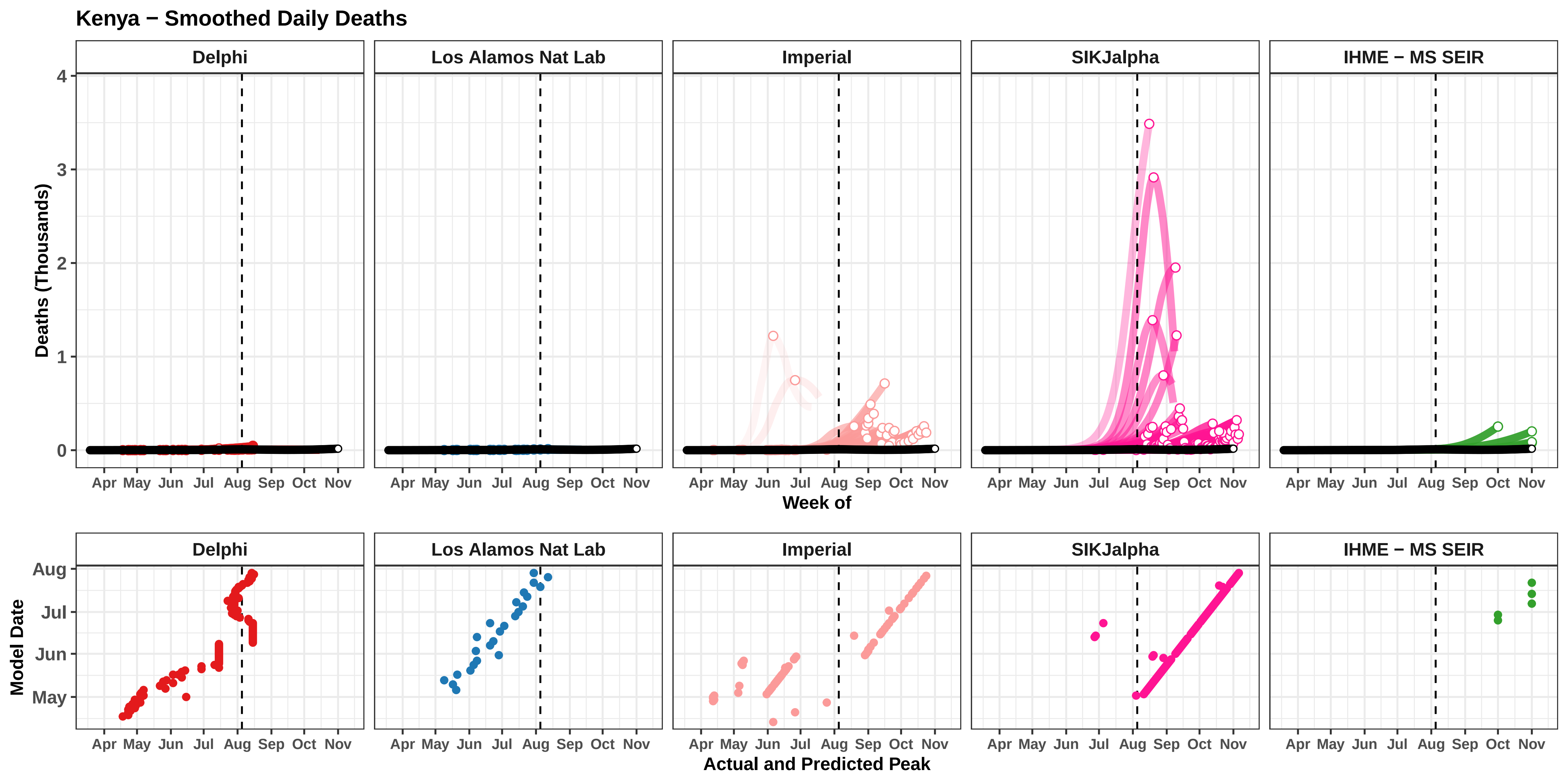

Kenya − Smoothed Daily Deaths
Delphi
Los Alamos Nat Lab
Imperial
SIKJalpha
IHME − MS SEIR
Deaths (Thousands)
0
1
2
3
4
Apr May Jun Jul Aug Sep Oct Nov
Week of
Model Date
May
Jun
Jul
Aug
Actual and Predicted Peak

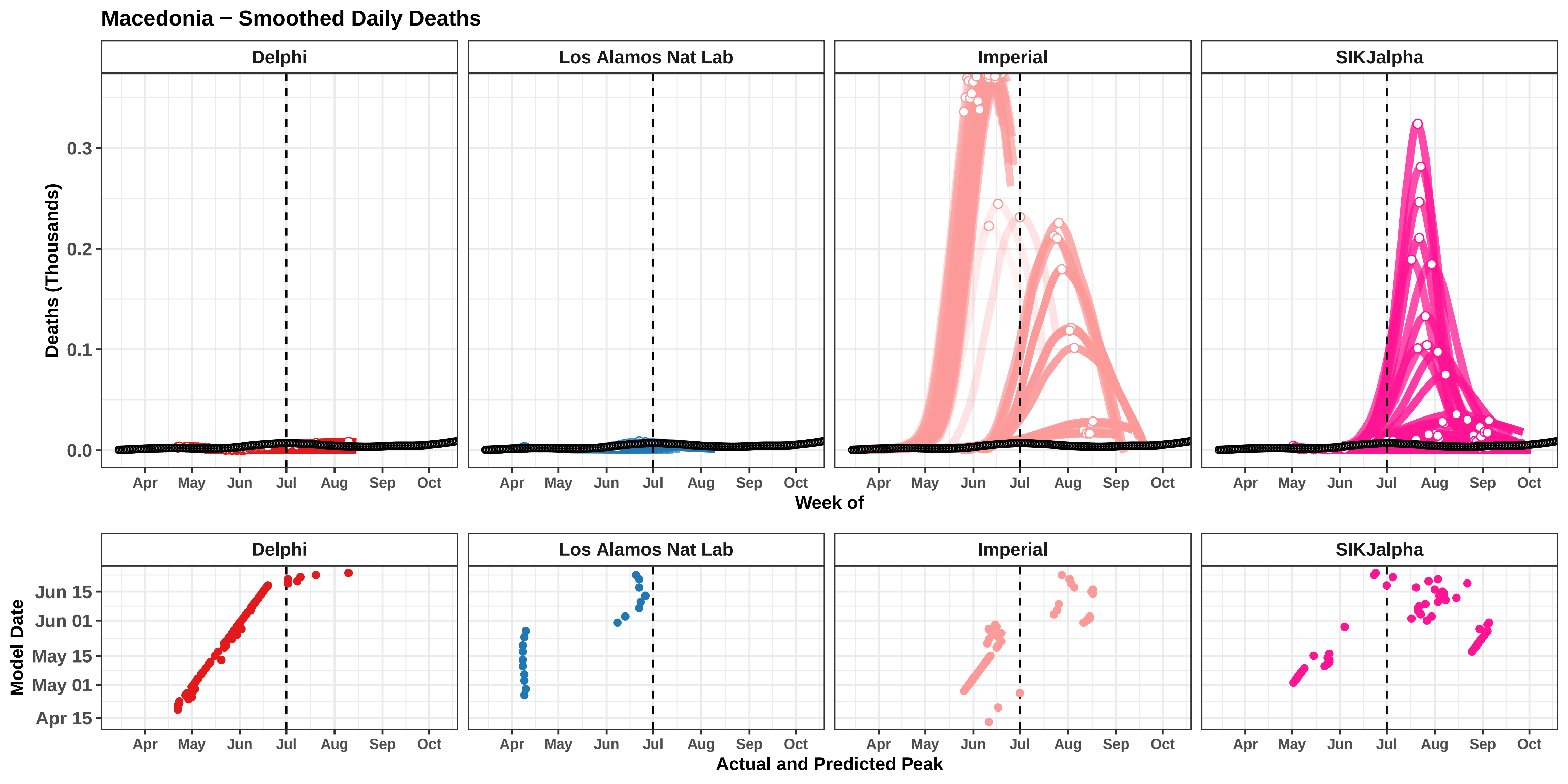

Macedonia − Smoothed Daily Deaths
Delphi
Los Alamos Nat Lab
Imperial
SIKJalpha
Deaths (Thousands)
0.0
0.1
0.2
0.3
Apr
May
Jun
Jul
Aug
Sep
Oct
Week of
Model Date
Jun 15
Jun 01
May 15
May 01
Apr 15
Actual and Predicted Peak

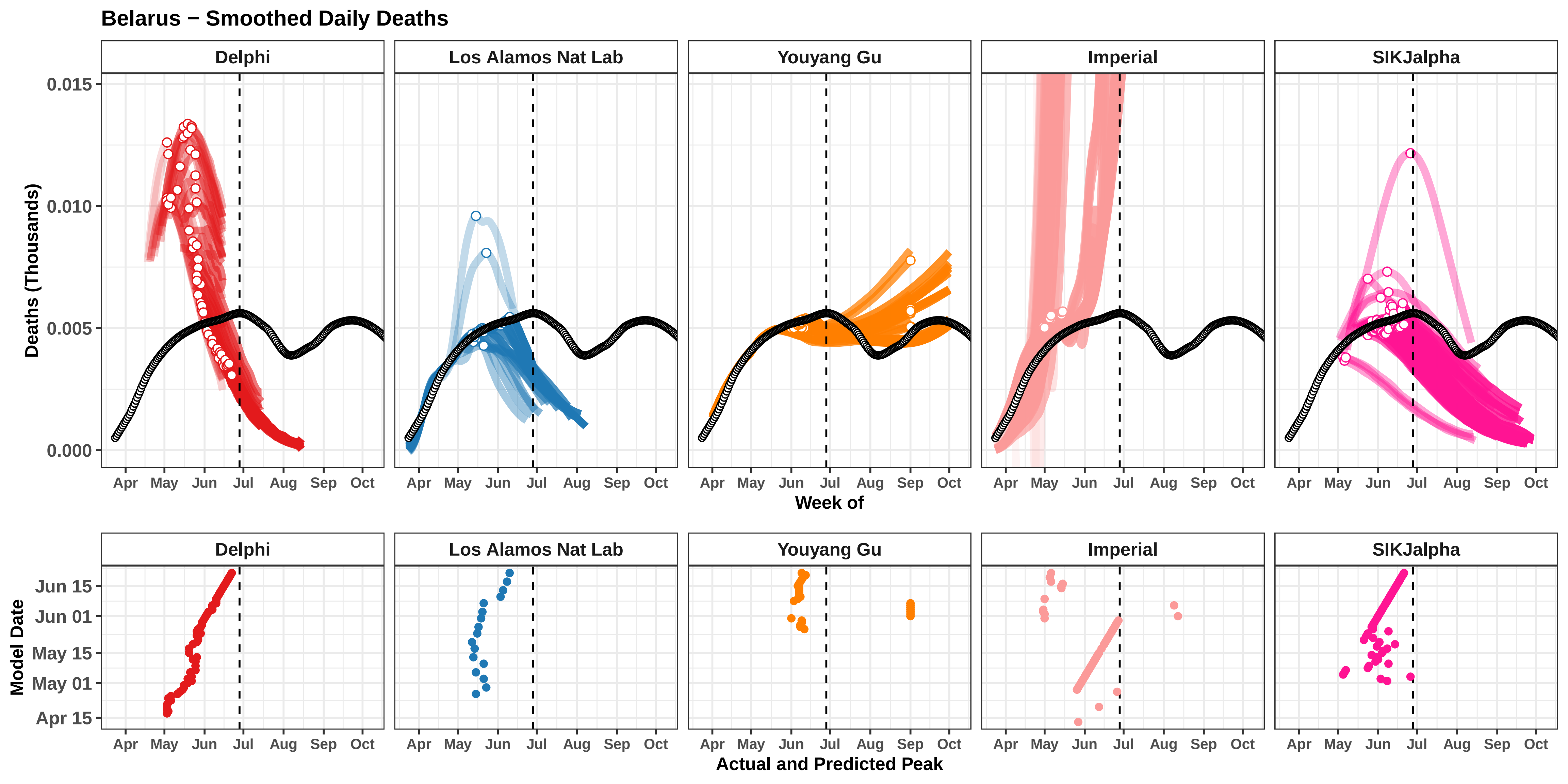

Belarus − Smoothed Daily Deaths
Delphi
Los Alamos Nat Lab
Youyang Gu
Imperial
SIKJalpha
Deaths (Thousands)
0.015
0.010
0.005
0.000
Apr
May
Jun
Jul
Aug
Sep
Oct
Week of
Model Date
Jun 15
Jun 01
May 15
May 01
Apr 15
Actual and Predicted Peak

### El Salvador − Smoothed Daily Deaths

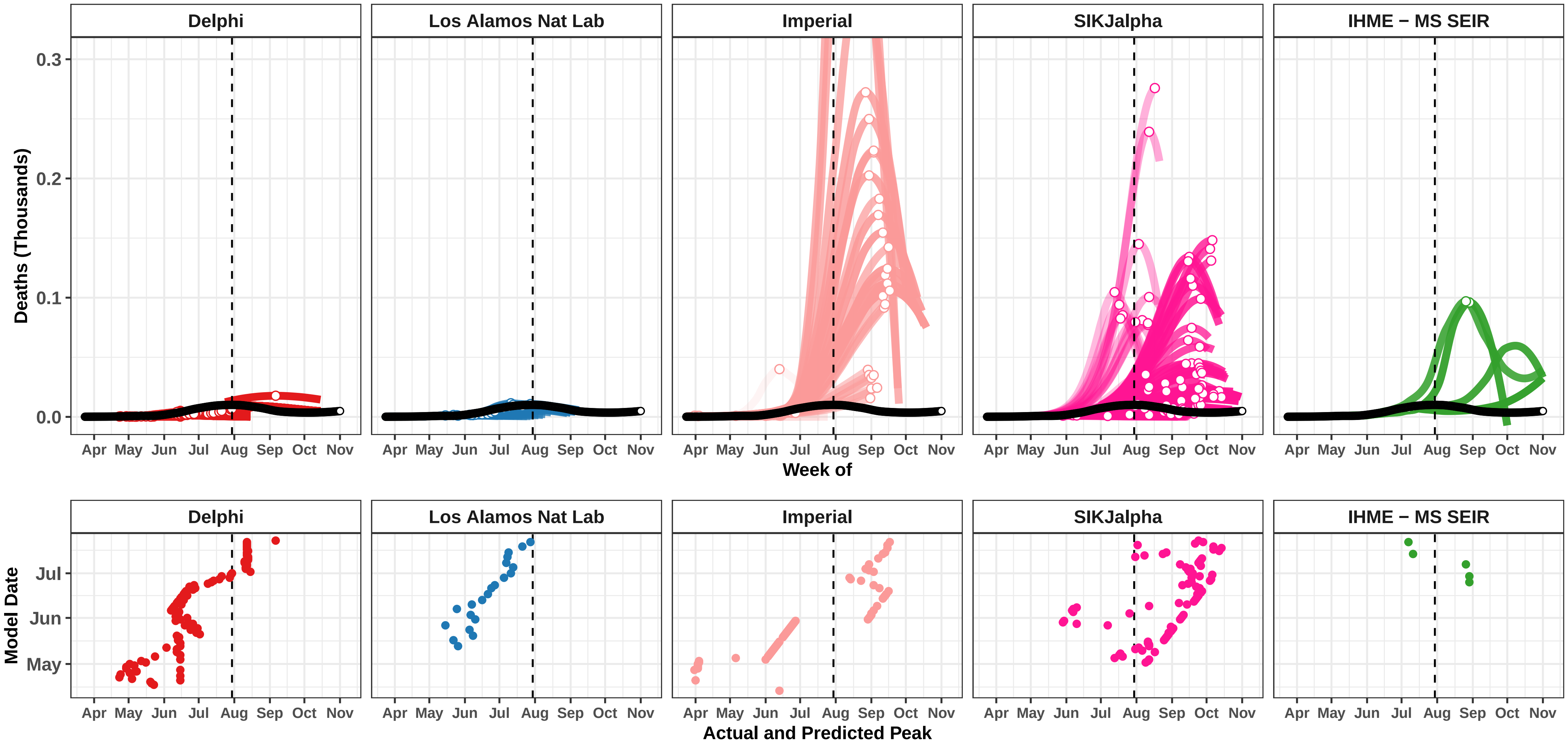

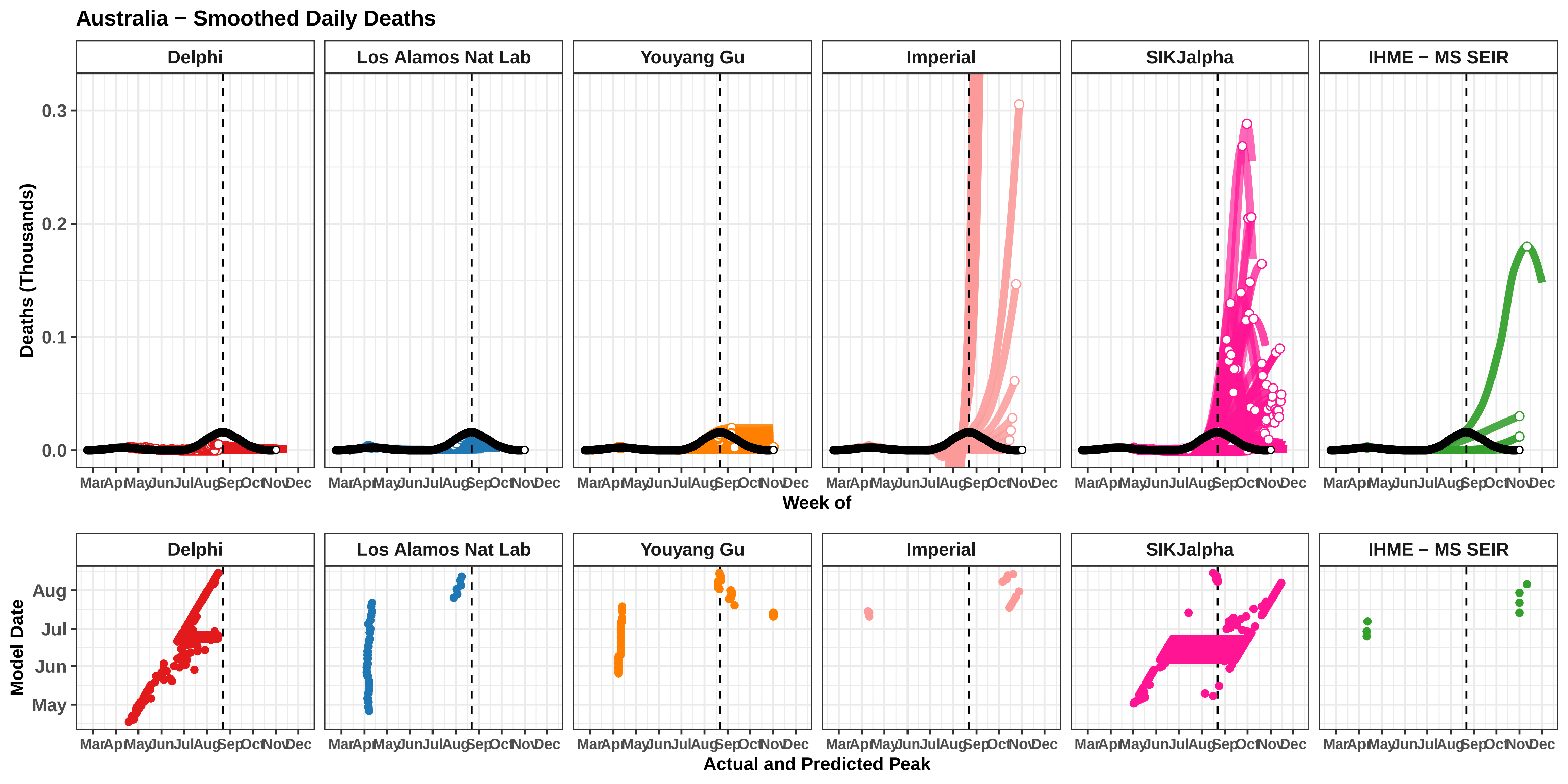

Australia − Smoothed Daily Deaths
Delphi
Los Alamos Nat Lab
Youyang Gu
Imperial
SIKJalpha
IHME − MS SEIR
Deaths (Thousands)
0.3
0.2
0.1
0.0
Mar Apr May Jun Jul Aug Sep Oct Nov Dec
Week of
Model Date
Aug
Jul
Jun
May
Actual and Predicted Peak

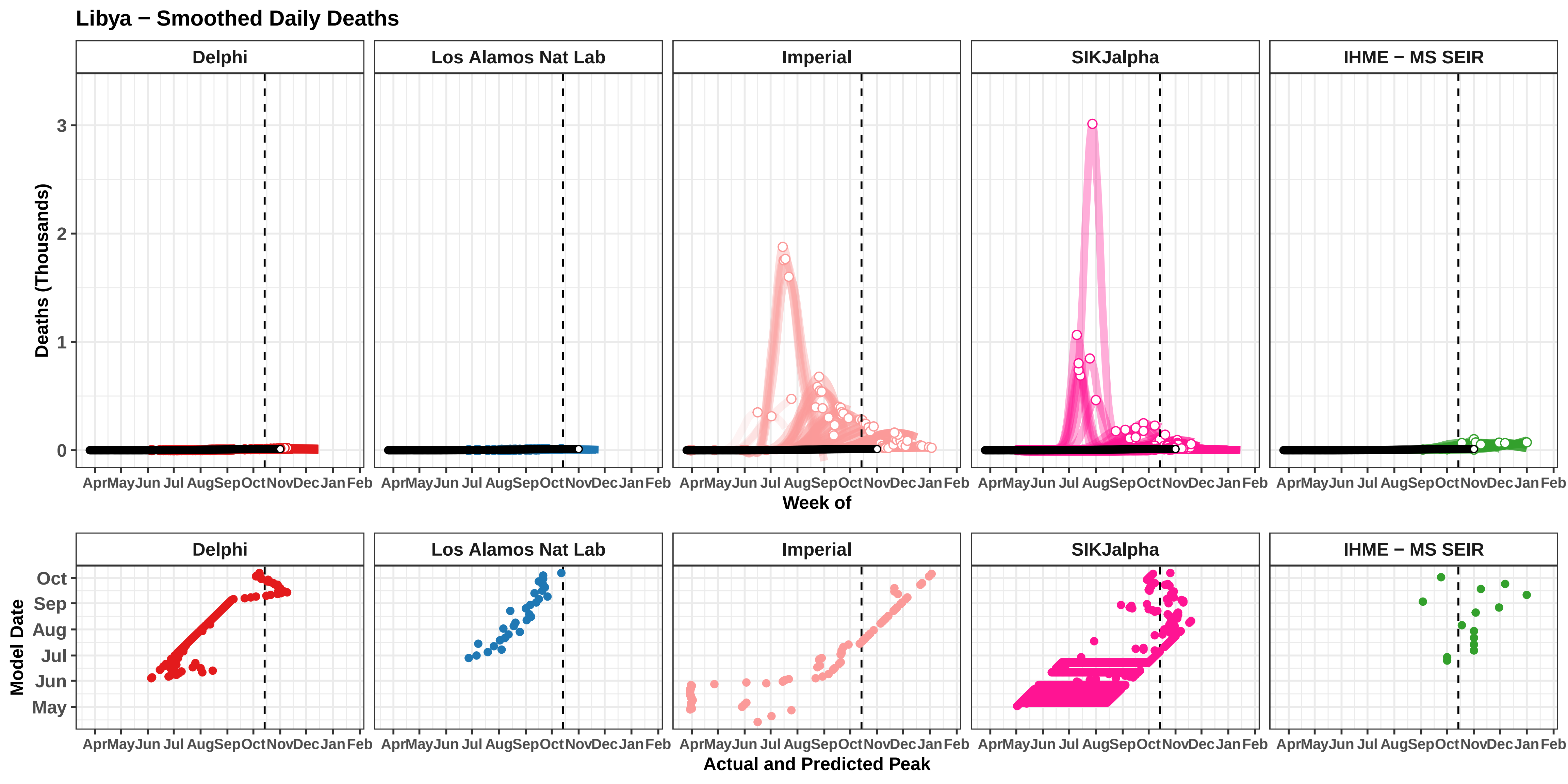

Libya − Smoothed Daily Deaths
Delphi
Los Alamos Nat Lab
Imperial
SIKJalpha
IHME − MS SEIR
Deaths (Thousands)
0
1
2
3
Apr May Jun Jul Aug Sep Oct Nov Dec Jan Feb
Week of
Model Date
May
Jun
Jul
Aug
Sep
Oct
Actual and Predicted Peak

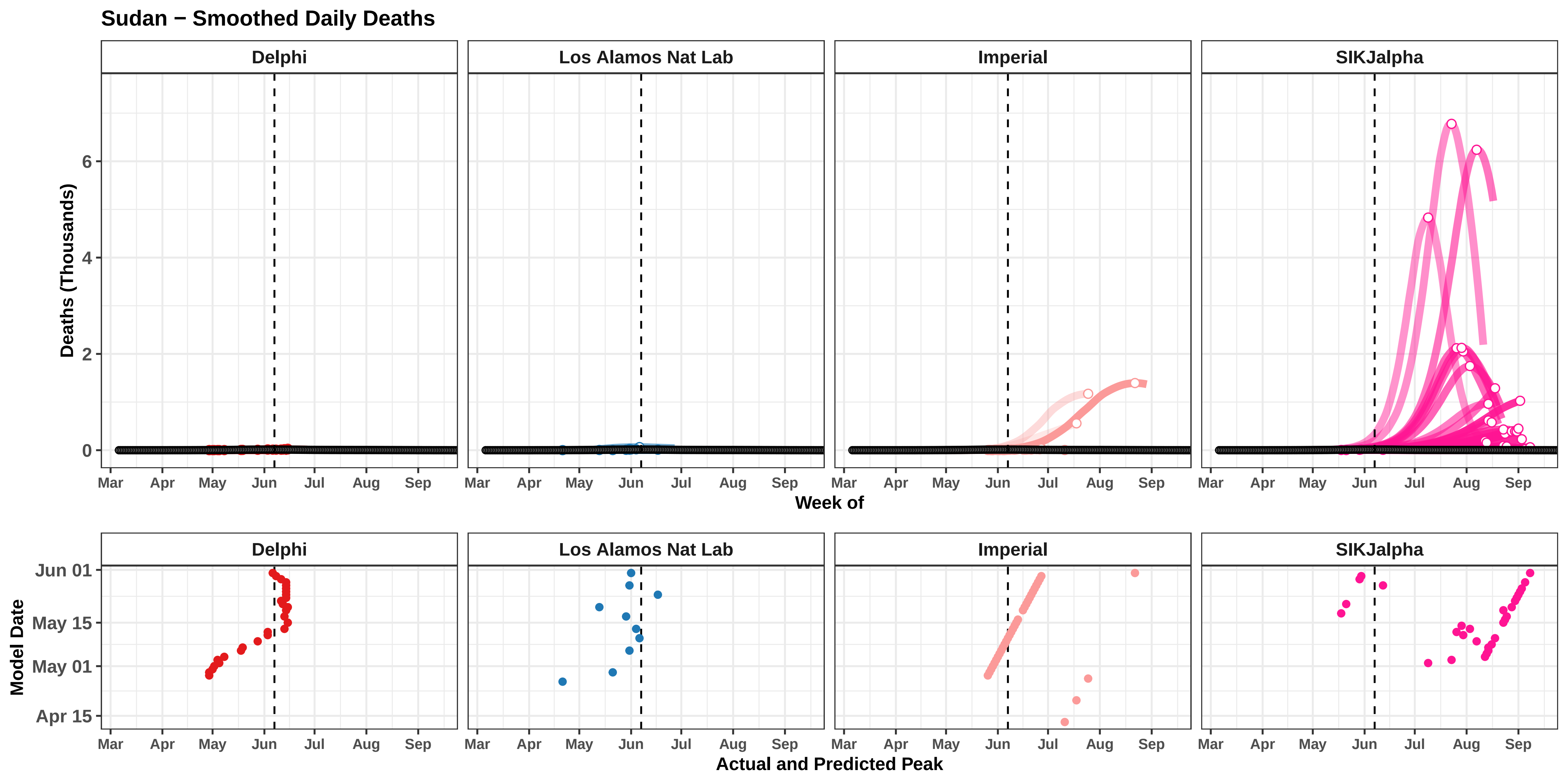

Sudan − Smoothed Daily Deaths
Delphi
Los Alamos Nat Lab
Imperial
SIKJalpha
Deaths (Thousands)
0
2
4
6
Mar
Apr
May
Jun
Jul
Aug
Sep
Week of
Model Date
Jun 01
May 15
May 01
Apr 15
Actual and Predicted Peak

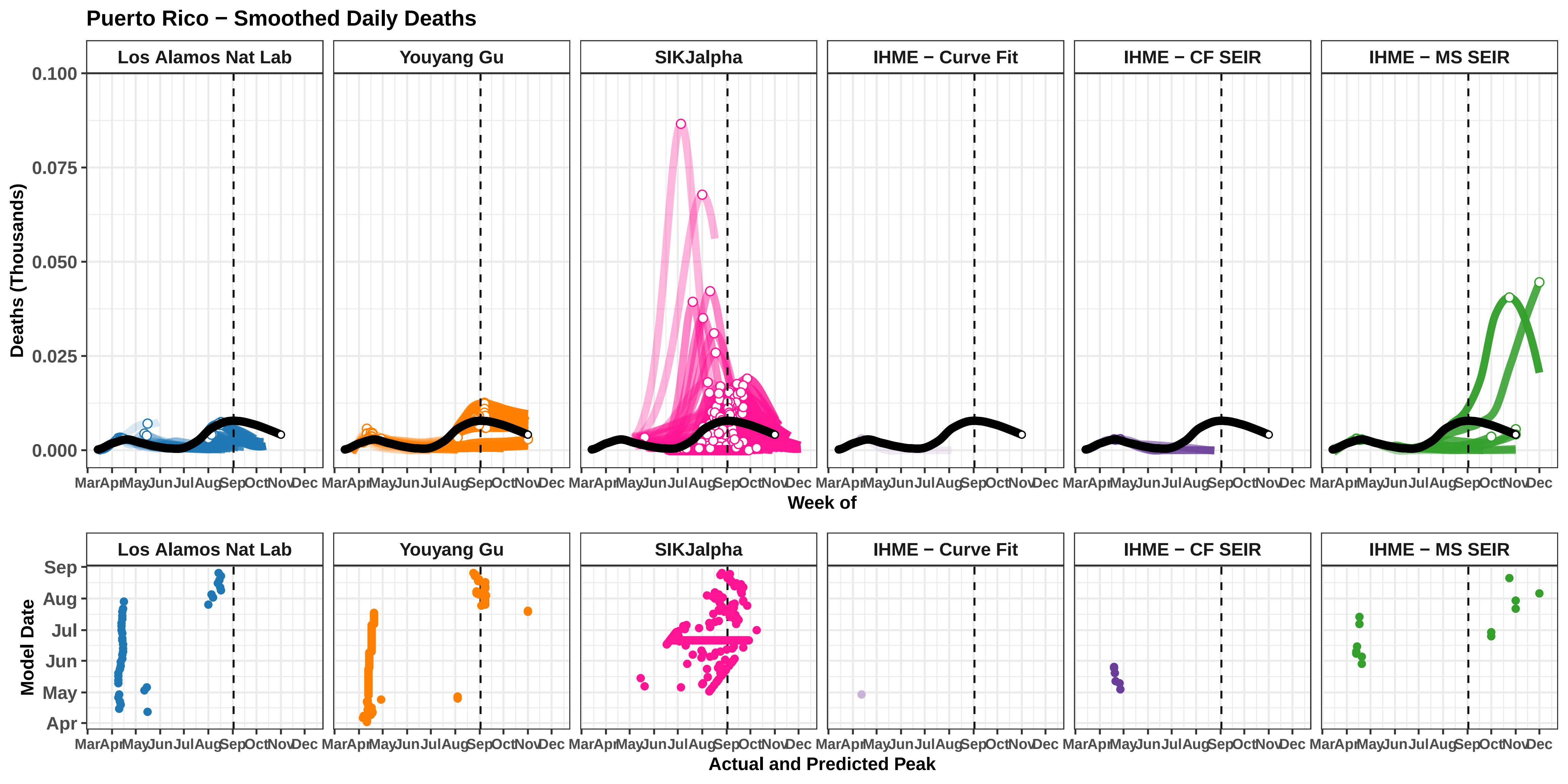

Puerto Rico − Smoothed Daily Deaths
Los Alamos Nat Lab
Youyang Gu
SIKJalpha
IHME − Curve Fit
IHME − CF SEIR
IHME − MS SEIR
Deaths (Thousands)
0.100
0.075
0.050
0.025
0.000
Mar Apr May Jun Jul Aug Sep Oct Nov Dec
Week of
Model Date
Sep
Aug
Jul
Jun
May
Apr
Actual and Predicted Peak

### Serbia − Smoothed Daily Deaths

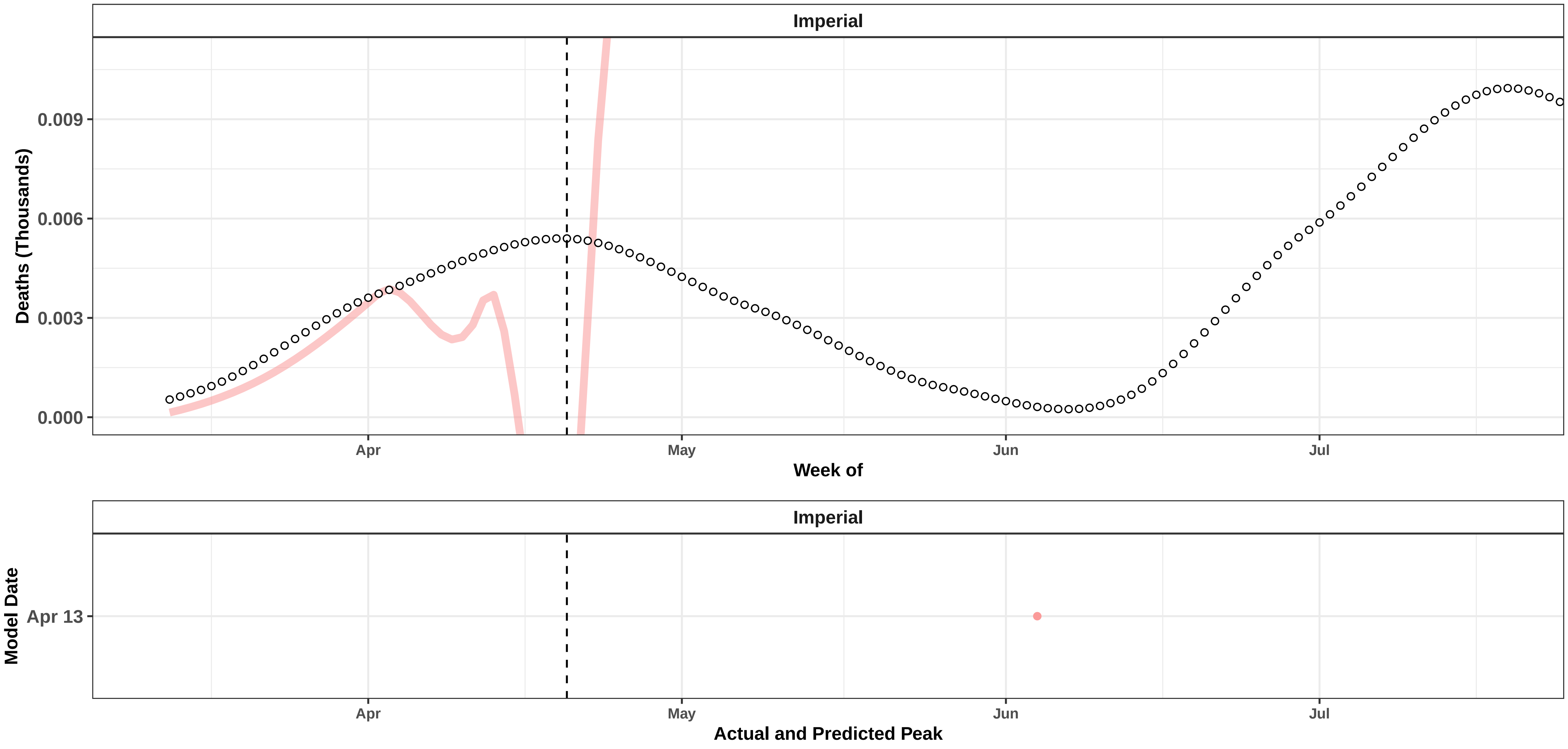

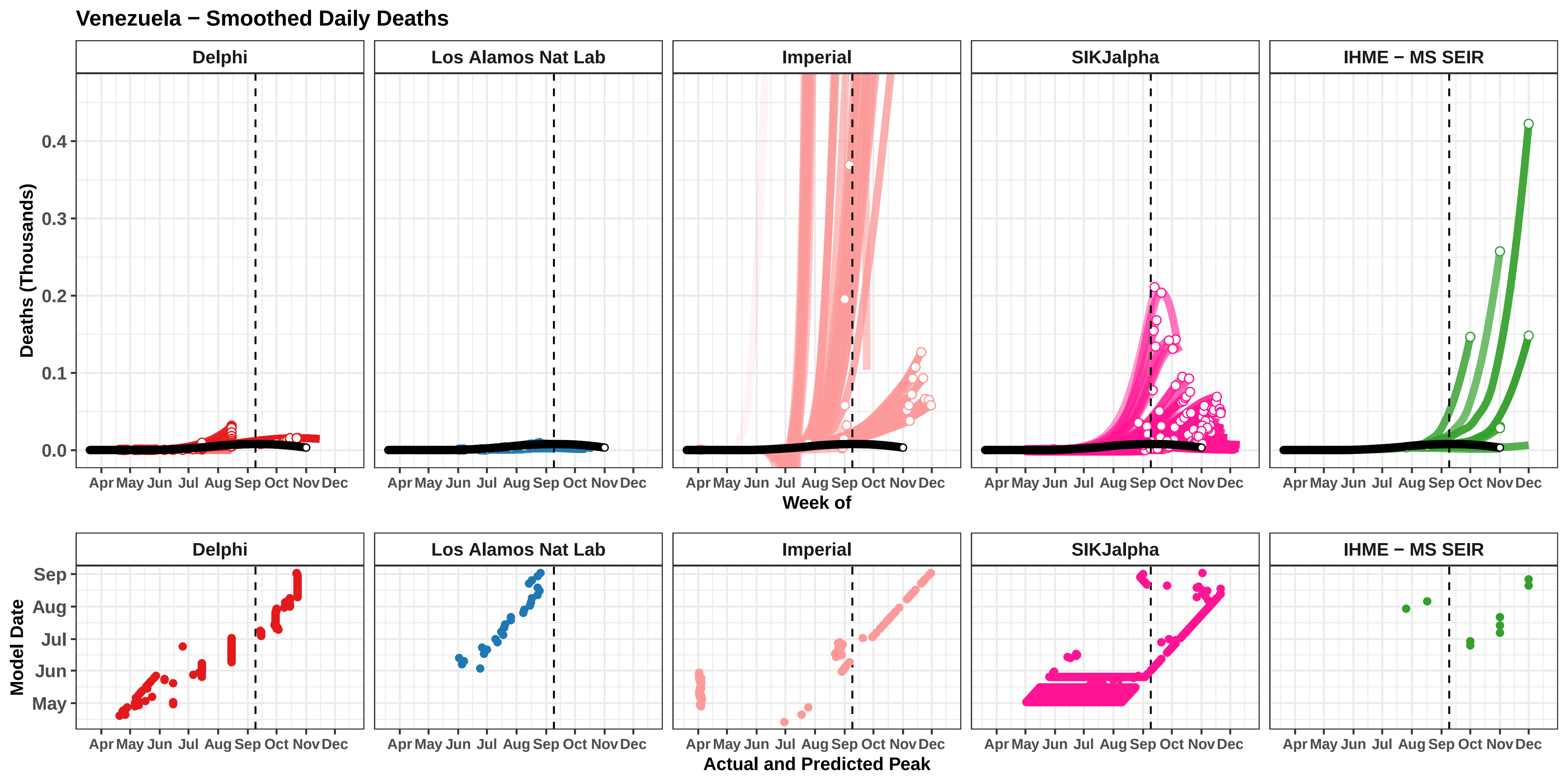

Venezuela − Smoothed Daily Deaths
Delphi
Los Alamos Nat Lab
Imperial
SIKJalpha
IHME − MS SEIR
Deaths (Thousands)
0.0
0.1
0.2
0.3
0.4
Apr May Jun Jul Aug Sep Oct Nov Dec
Week of
Model Date
May
Jun
Jul
Aug
Sep
Actual and Predicted Peak

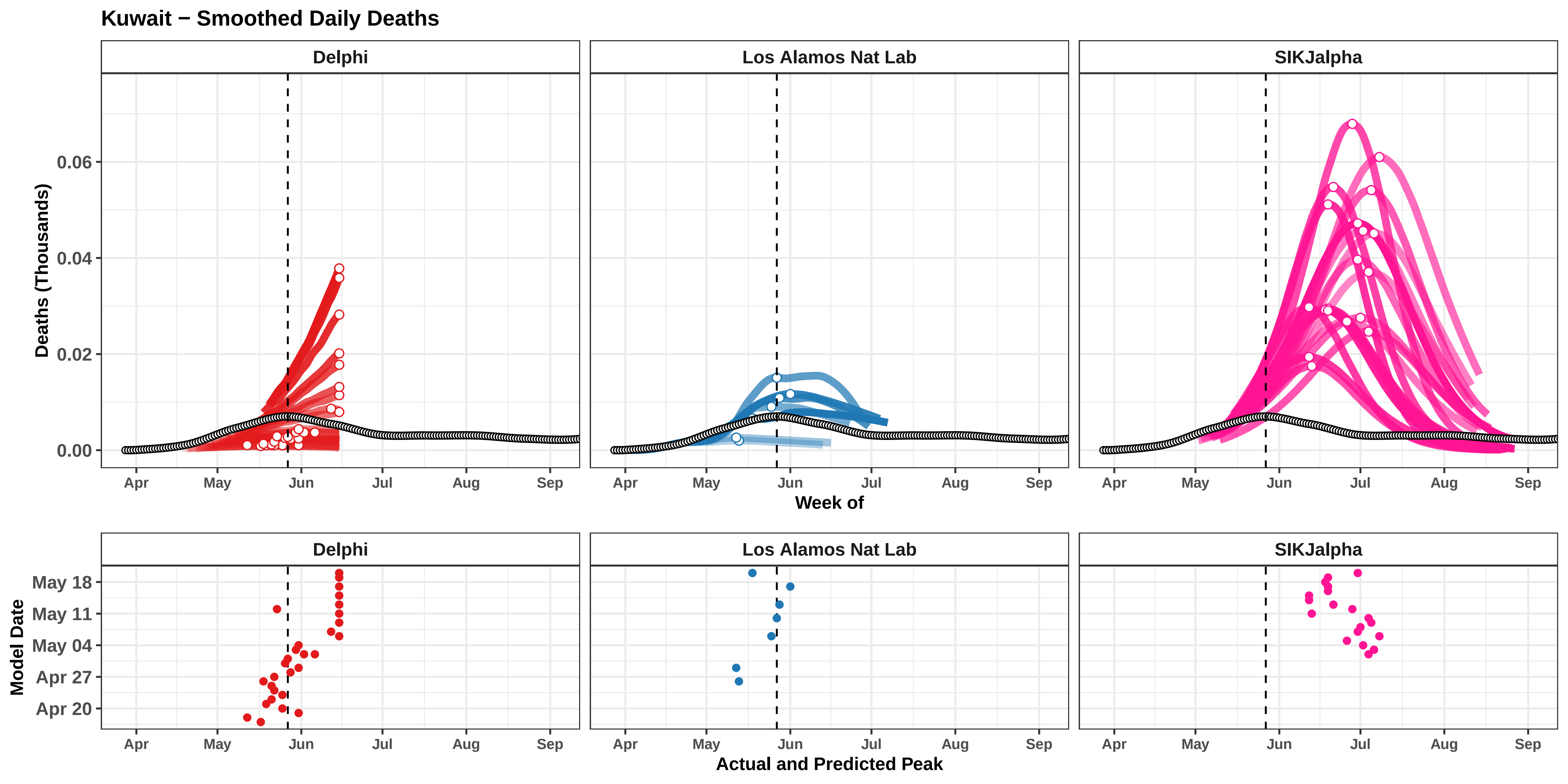

Kuwait − Smoothed Daily Deaths
Delphi
Los Alamos Nat Lab
SIKJalpha
Deaths (Thousands)
0.00
0.02
0.04
0.06
Apr
May
Jun
Jul
Aug
Sep
Week of
Model Date
May 18
May 11
May 04
Apr 27
Apr 20
Actual and Predicted Peak

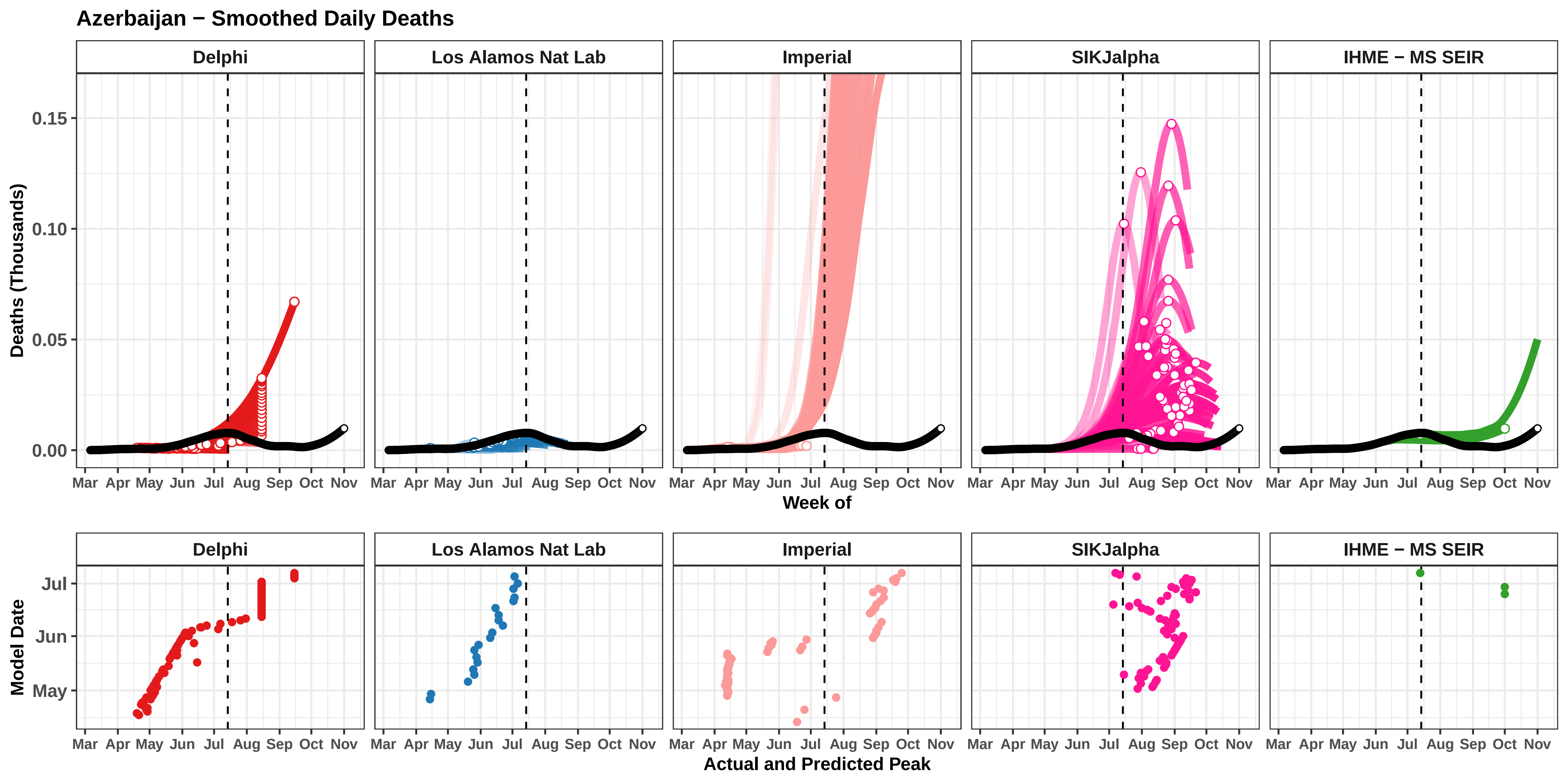

Azerbaijan − Smoothed Daily Deaths
Delphi
Los Alamos Nat Lab
Imperial
SIKJalpha
IHME − MS SEIR
Deaths (Thousands)
0.15
0.10
0.05
0.00
Mar Apr May Jun Jul Aug Sep Oct Nov
Week of
Model Date
Jul
Jun
May
Actual and Predicted Peak

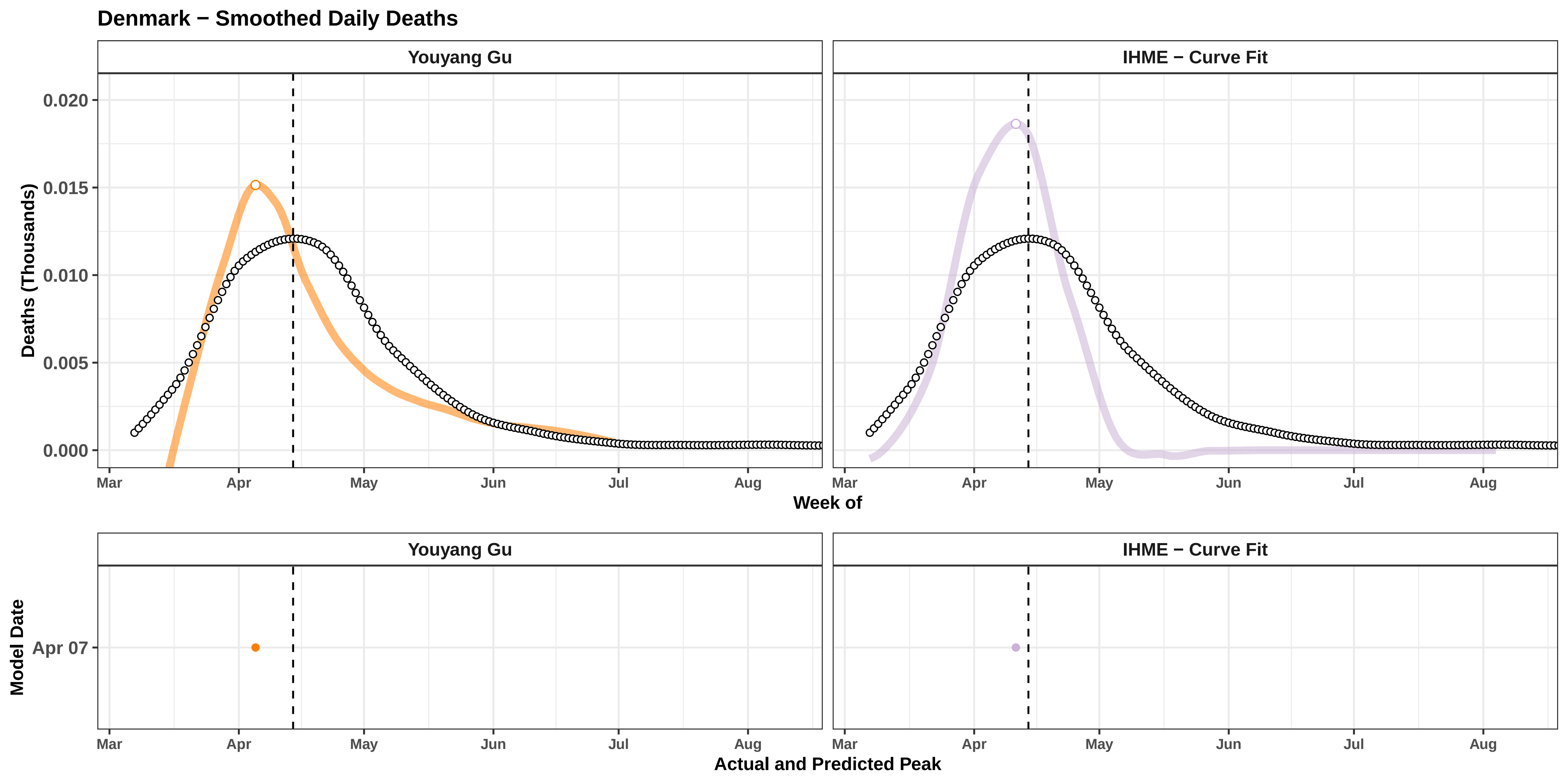

Denmark − Smoothed Daily Deaths
Youyang Gu
IHME − Curve Fit
Deaths (Thousands)
0.020
0.015
0.010
0.005
0.000
Mar
Apr
May
Jun
Jul
Aug
Week of
Model Date
Apr 07
Actual and Predicted Peak

**Delaware − Smoothed Daily Deaths**

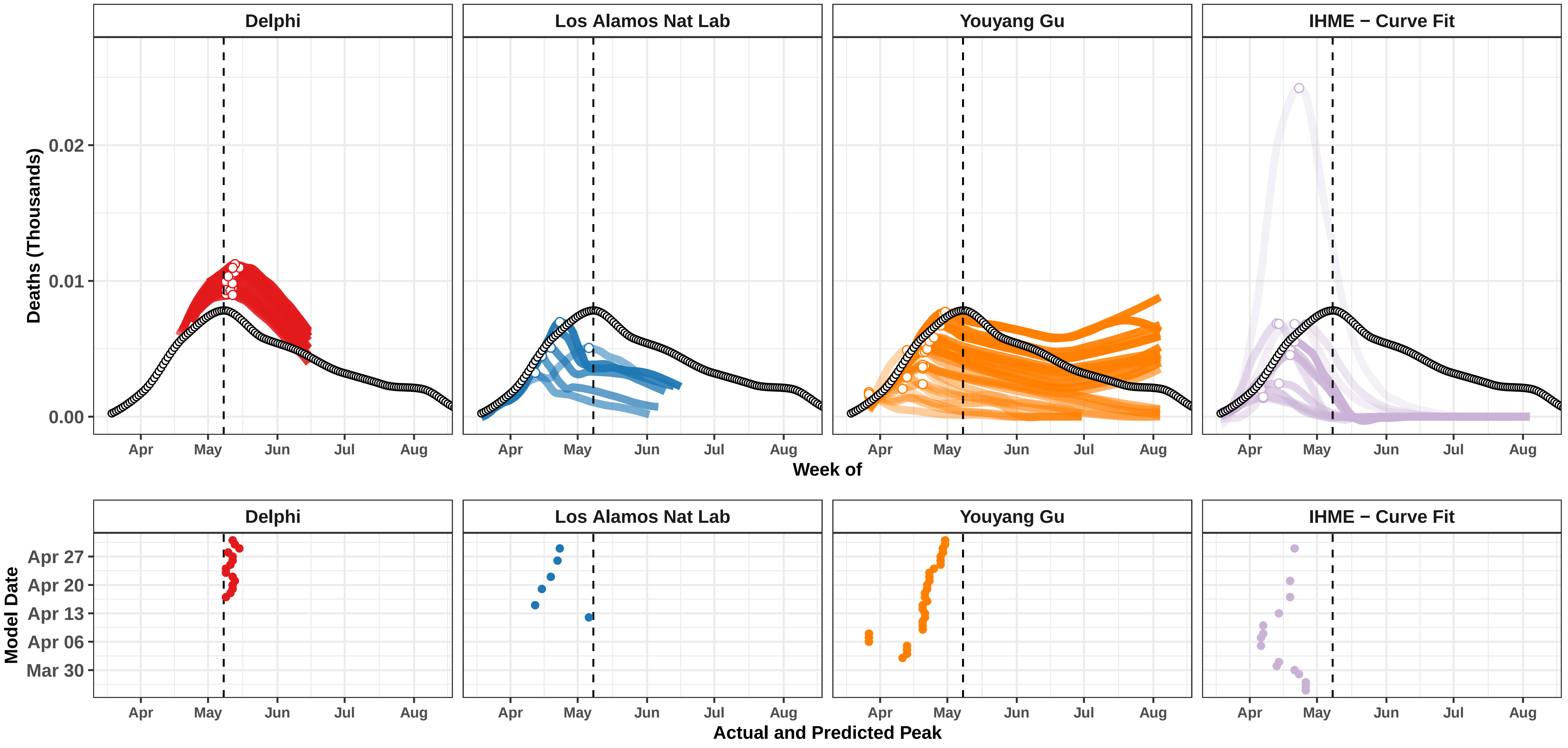

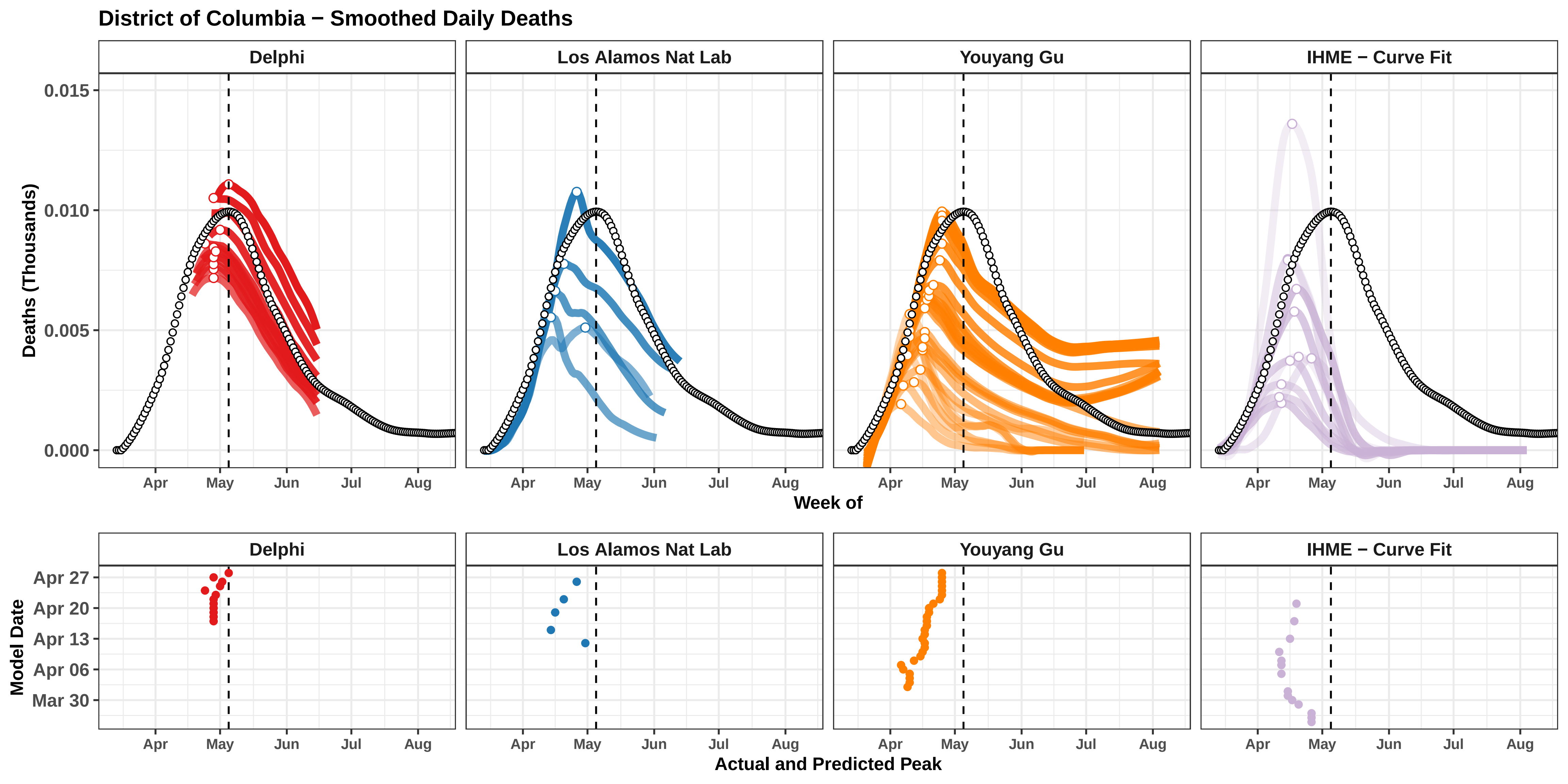

District of Columbia − Smoothed Daily Deaths
Delphi
Los Alamos Nat Lab
Youyang Gu
IHME − Curve Fit
Deaths (Thousands)
0.015
0.010
0.005
0.000
Apr
May
Jun
Jul
Aug
Week of
Model Date
Apr 27
Apr 20
Apr 13
Apr 06
Mar 30
Actual and Predicted Peak

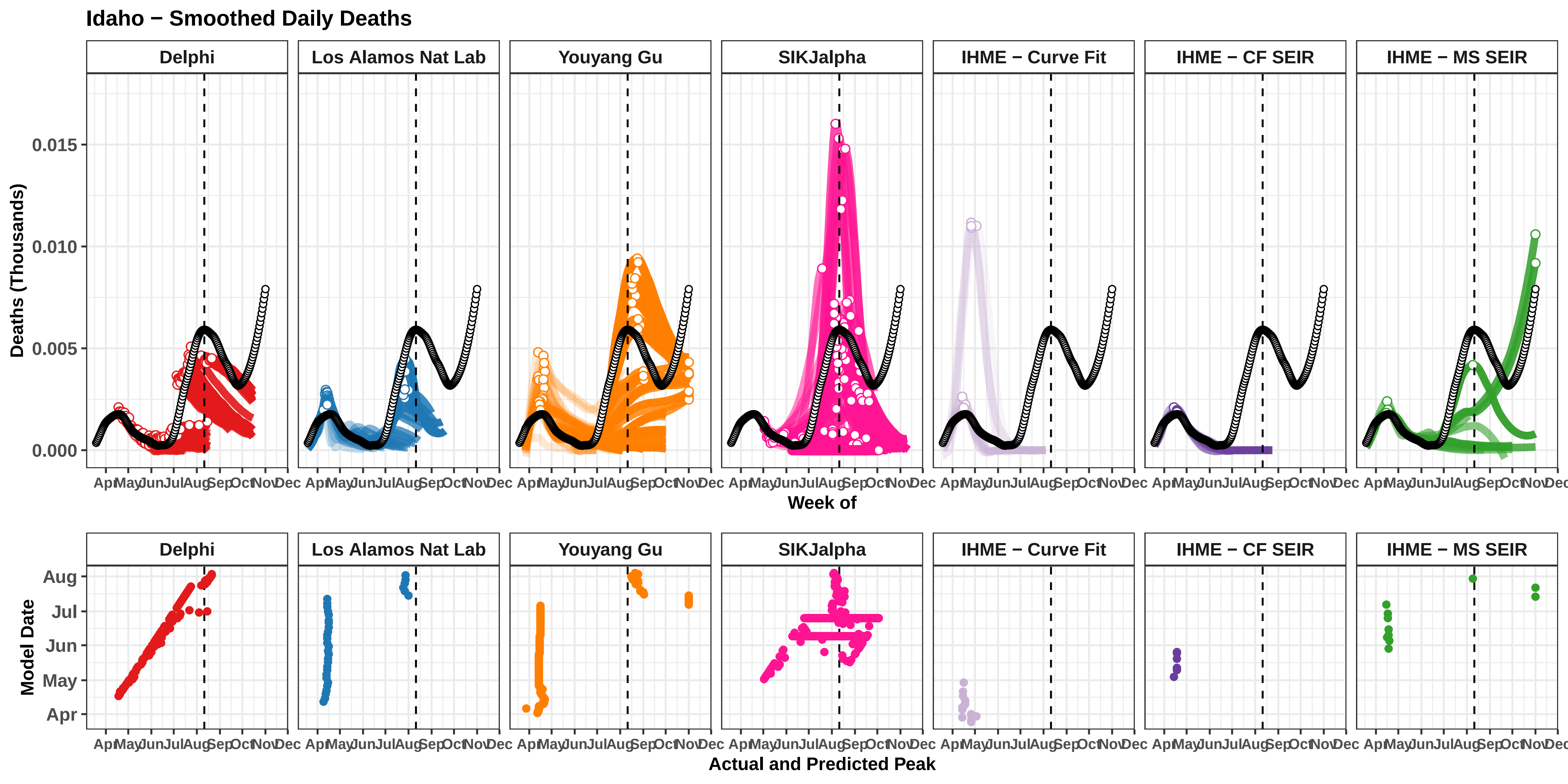

Idaho − Smoothed Daily Deaths
Delphi
Los Alamos Nat Lab
Youyang Gu
SIKJalpha
IHME − Curve Fit
IHME − CF SEIR
IHME − MS SEIR
Deaths (Thousands)
0.015
0.010
0.005
0.000
Apr May Jun Jul Aug Sep Oct Nov Dec
Week of
Model Date
Aug
Jul
Jun
May
Apr
Actual and Predicted Peak

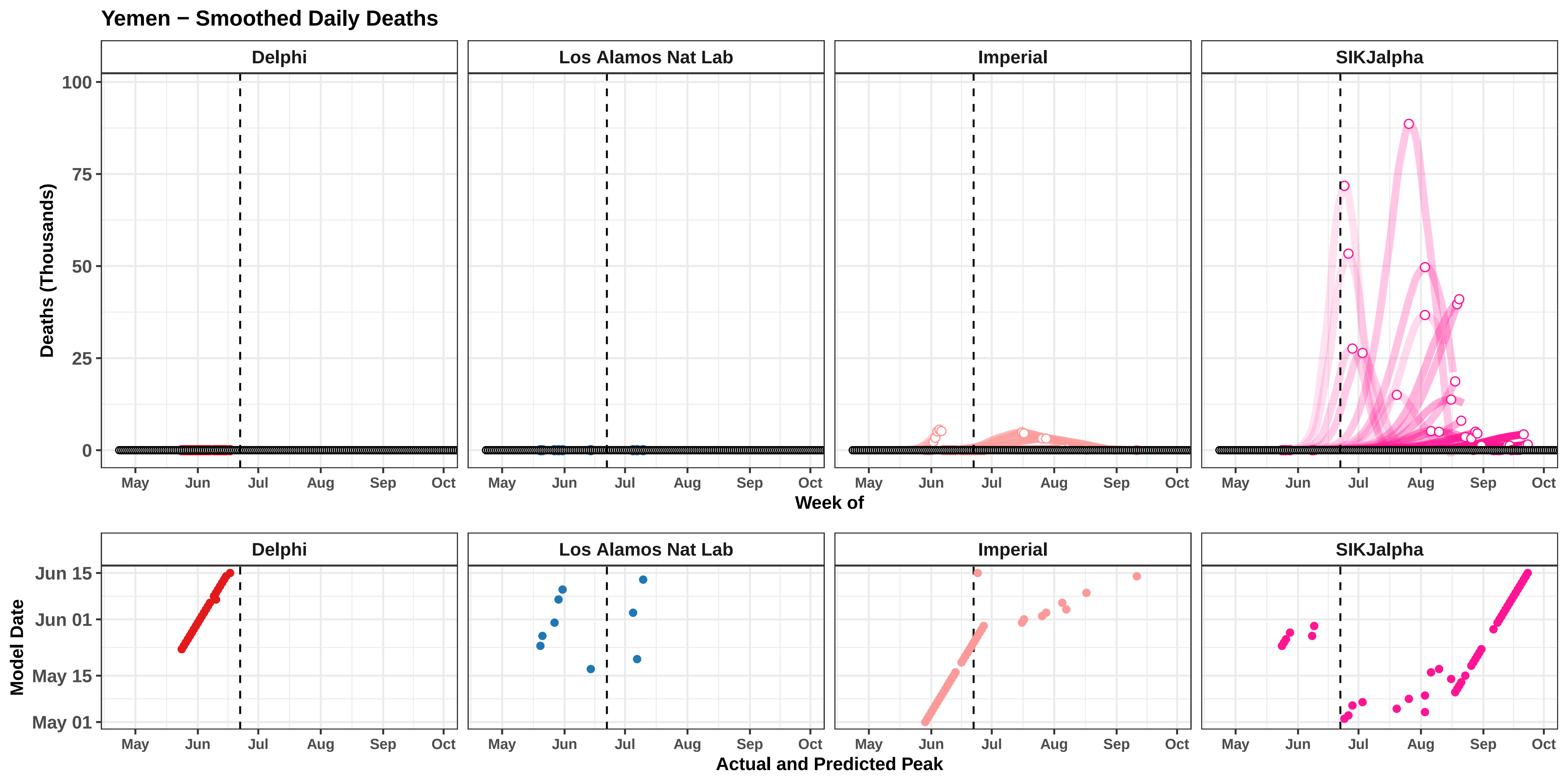
Yemen − Smoothed Daily Deaths
Delphi
Los Alamos Nat Lab
Imperial
SIKJalpha
Deaths (Thousands)
100
75
50
25
0
May
Jun
Jul
Aug
Sep
Oct
Week of
Model Date
Jun 15
Jun 01
May 15
May 01
Actual and Predicted Peak

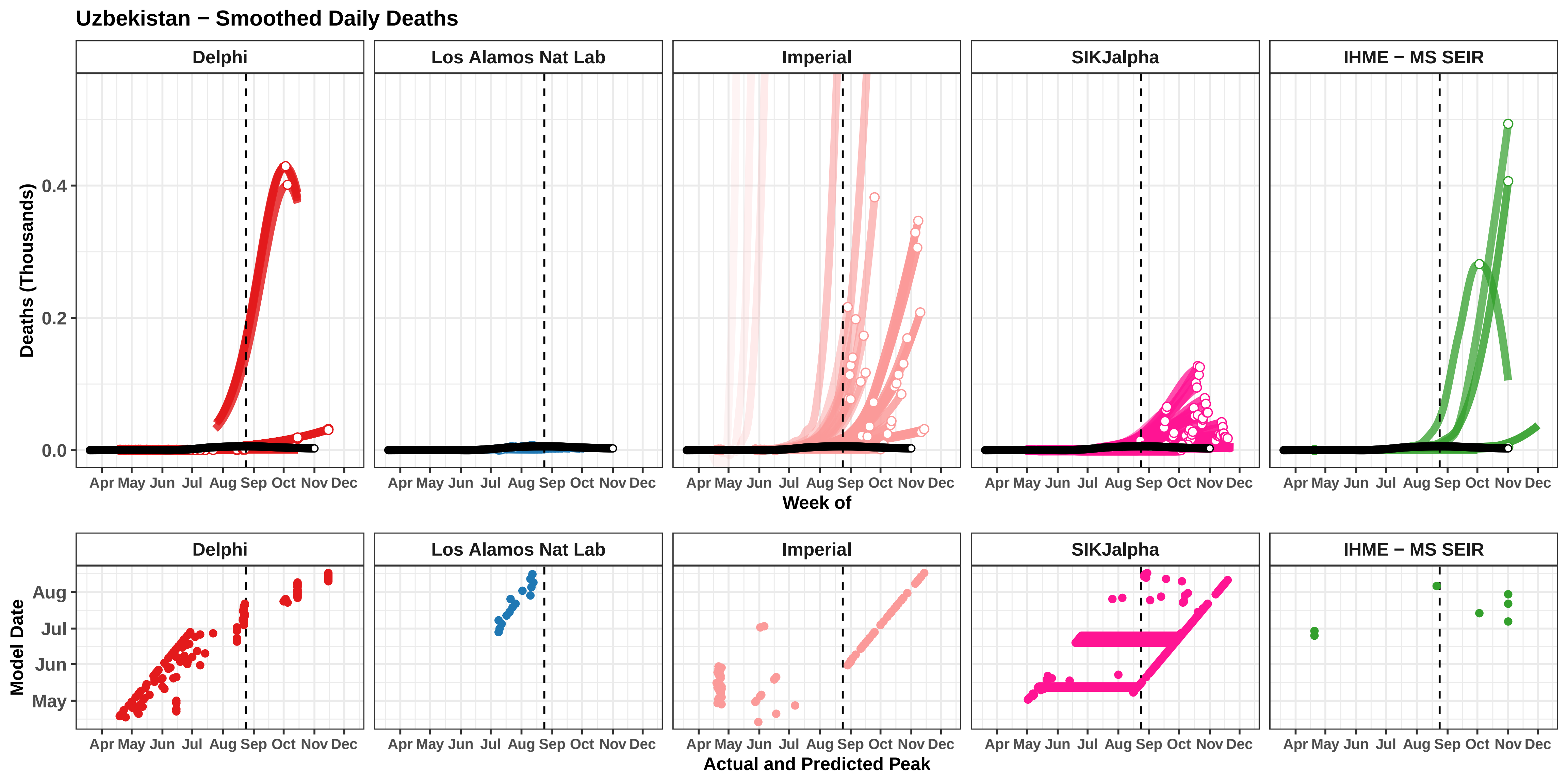

Uzbekistan − Smoothed Daily Deaths
Delphi
Los Alamos Nat Lab
Imperial
SIKJalpha
IHME − MS SEIR
Deaths (Thousands)
0.0
0.2
0.4
Apr May Jun Jul Aug Sep Oct Nov Dec
Week of
Model Date
May
Jun
Jul
Aug
Actual and Predicted Peak

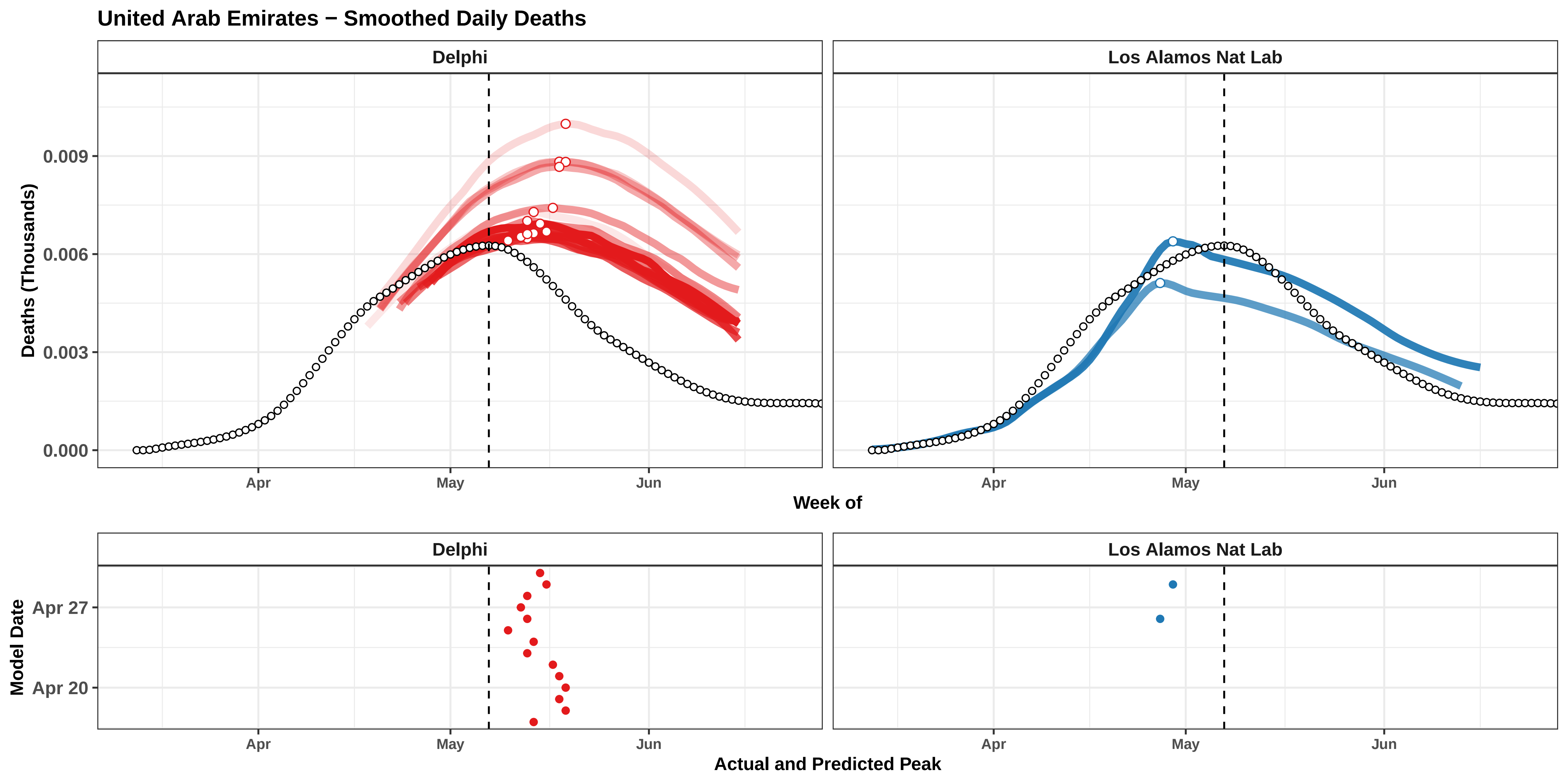

United Arab Emirates − Smoothed Daily Deaths
Delphi
Los Alamos Nat Lab
Deaths (Thousands)
0.009
0.006
0.003
0.000
Apr
May
Jun
Week of
Delphi
Los Alamos Nat Lab
Model Date
Apr 27
Apr 20
Apr
May
Jun
Actual and Predicted Peak

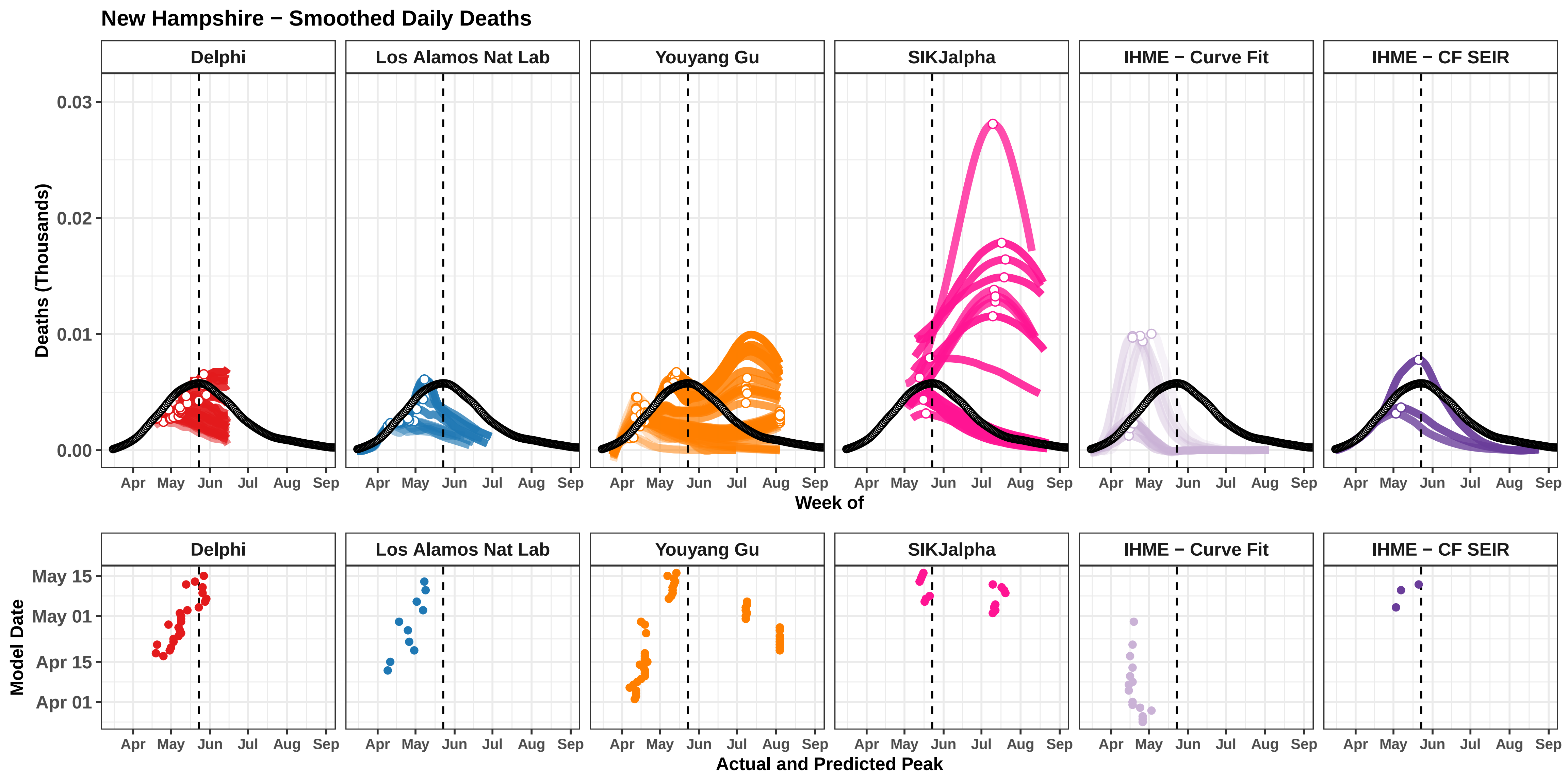

New Hampshire − Smoothed Daily Deaths
Delphi
Los Alamos Nat Lab
Youyang Gu
SIKJalpha
IHME − Curve Fit
IHME − CF SEIR
Deaths (Thousands)
0.03
0.02
0.01
0.00
Apr
May
Jun
Jul
Aug
Sep
Week of
Model Date
May 15
May 01
Apr 15
Apr 01
Actual and Predicted Peak

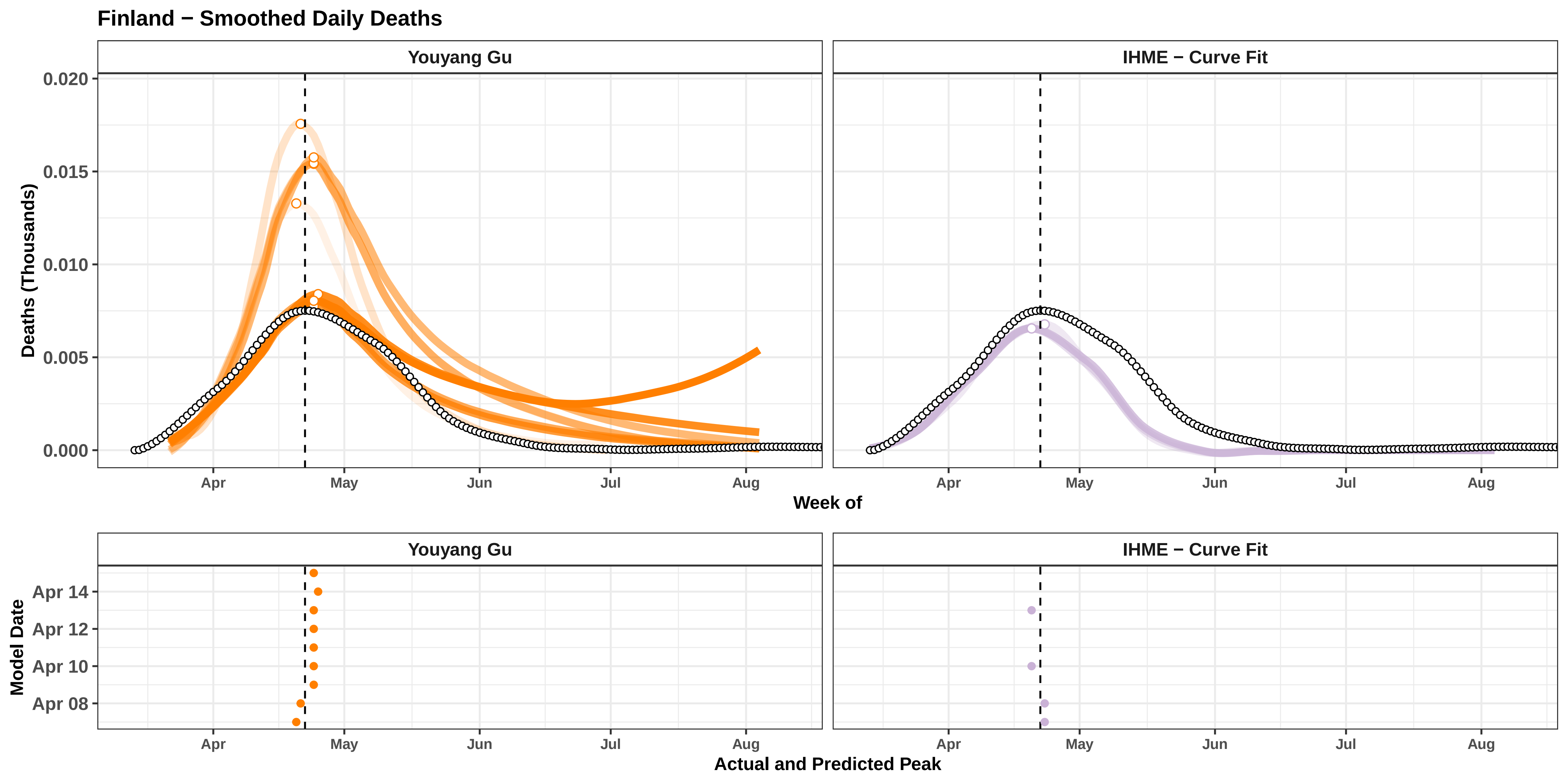

Finland − Smoothed Daily Deaths
Youyang Gu
IHME − Curve Fit
Deaths (Thousands)
0.020
0.015
0.010
0.005
0.000
Apr
May
Jun
Jul
Aug
Week of
Youyang Gu
IHME − Curve Fit
Model Date
Apr 14
Apr 12
Apr 10
Apr 08
Apr
May
Jun
Jul
Aug
Actual and Predicted Peak

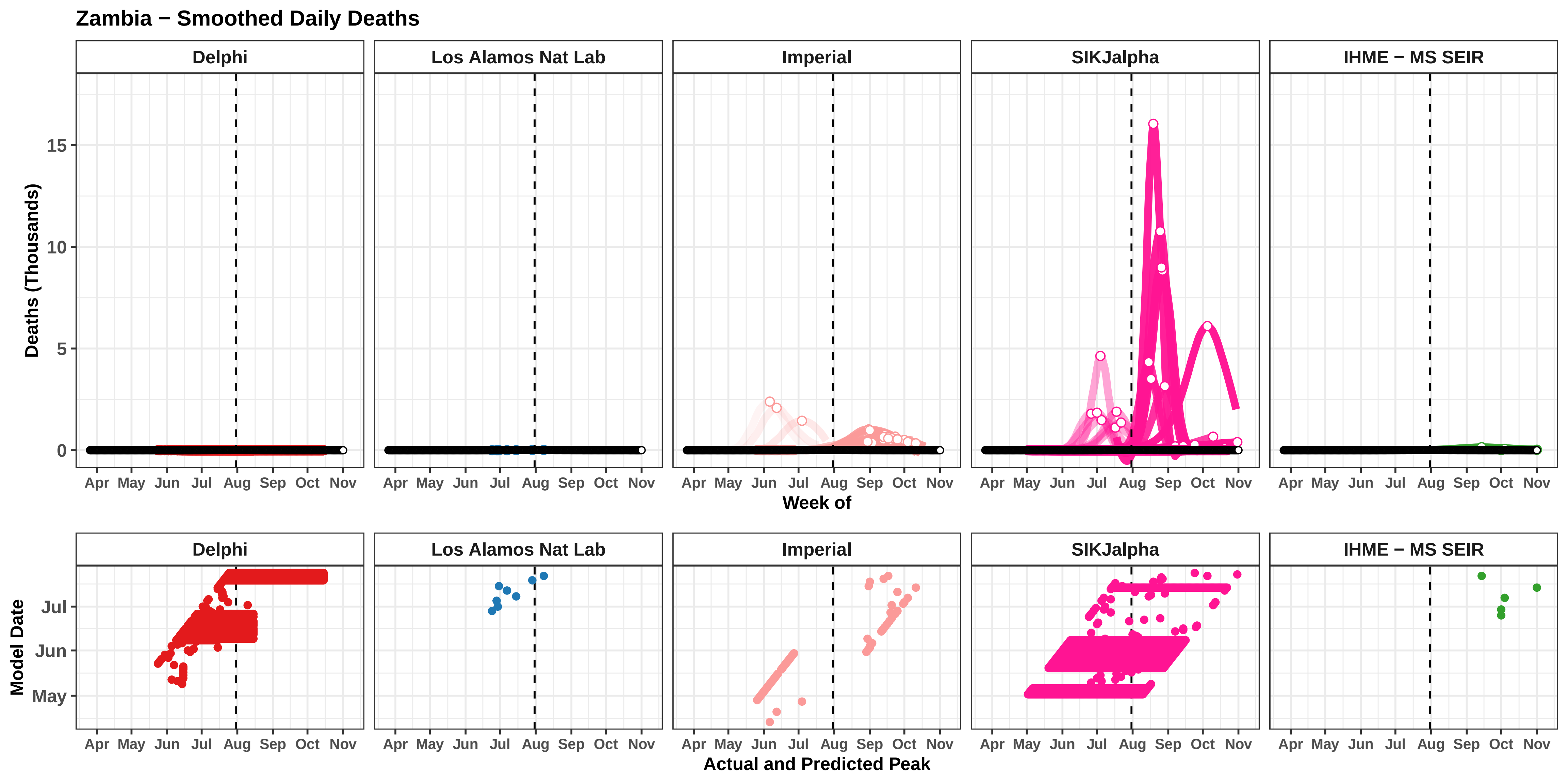

Zambia − Smoothed Daily Deaths
Delphi
Los Alamos Nat Lab
Imperial
SIKJalpha
IHME − MS SEIR
Deaths (Thousands)
0
5
10
15
Apr May Jun Jul Aug Sep Oct Nov
Week of
Model Date
May
Jun
Jul
Actual and Predicted Peak
