## Supplemental Figures - Smoothed Daily Deaths for "Predictive performance of international COVID-19 mortality forecasting models"

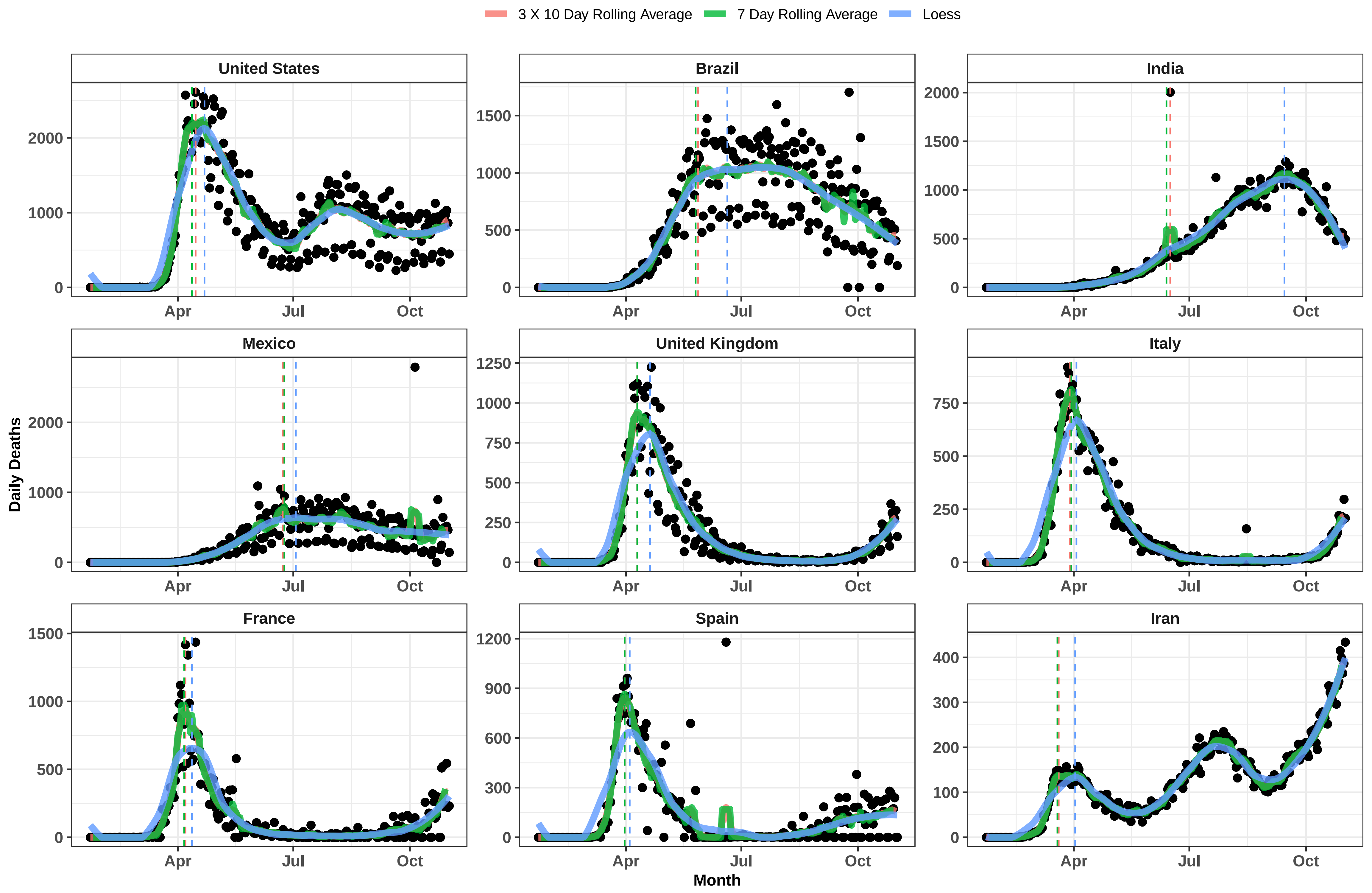

3 X 10 Day Rolling Average
7 Day Rolling Average
Loess
United States
Brazil
India
Mexico
United Kingdom
Italy
France
Spain
Iran
Daily Deaths
Month
Apr
Jul
Oct

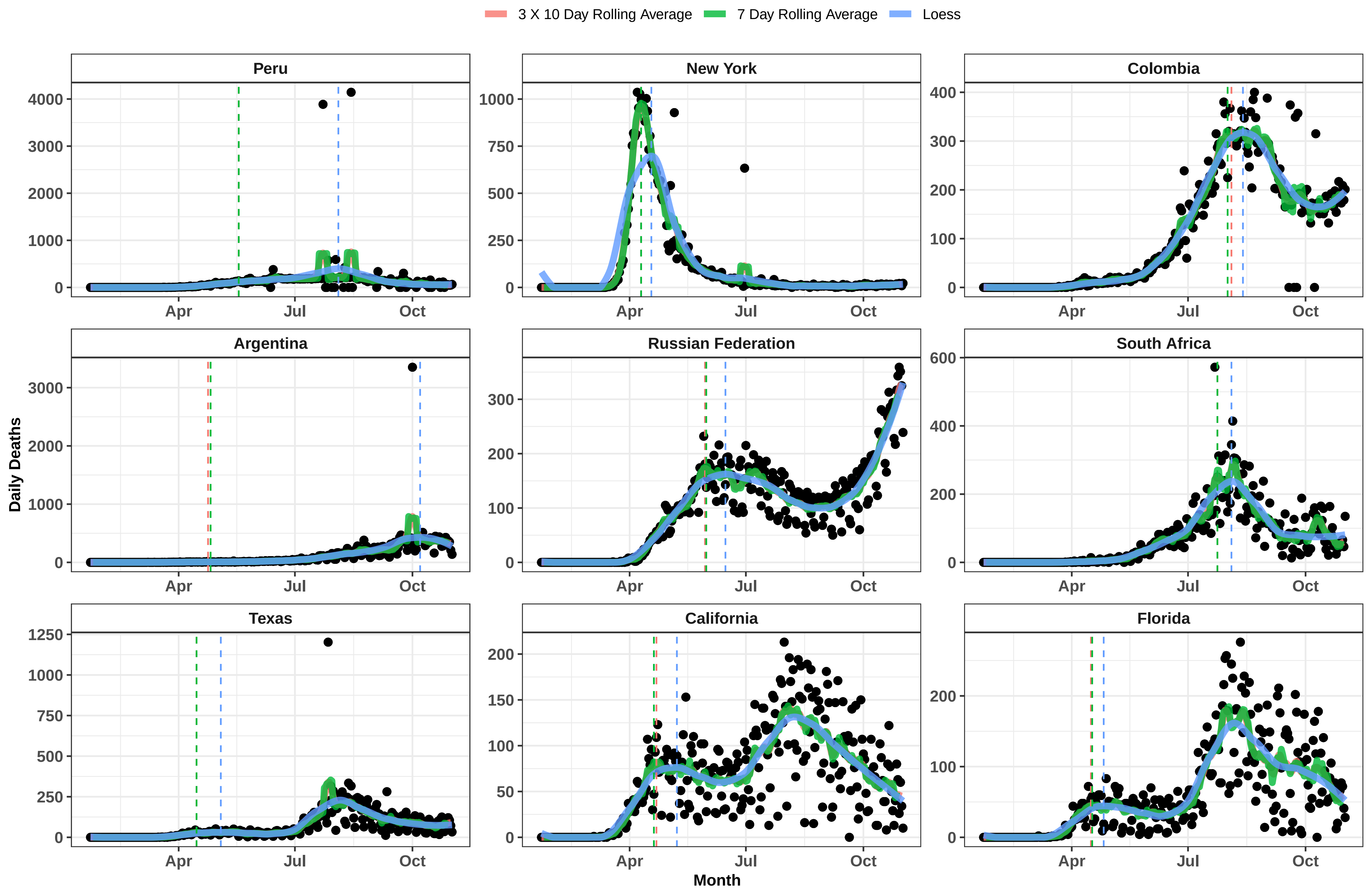

3 X 10 Day Rolling Average
7 Day Rolling Average
Loess
Peru
New York
Colombia
Argentina
Russian Federation
South Africa
Texas
California
Florida
Daily Deaths
Month
Apr
Jul
Oct

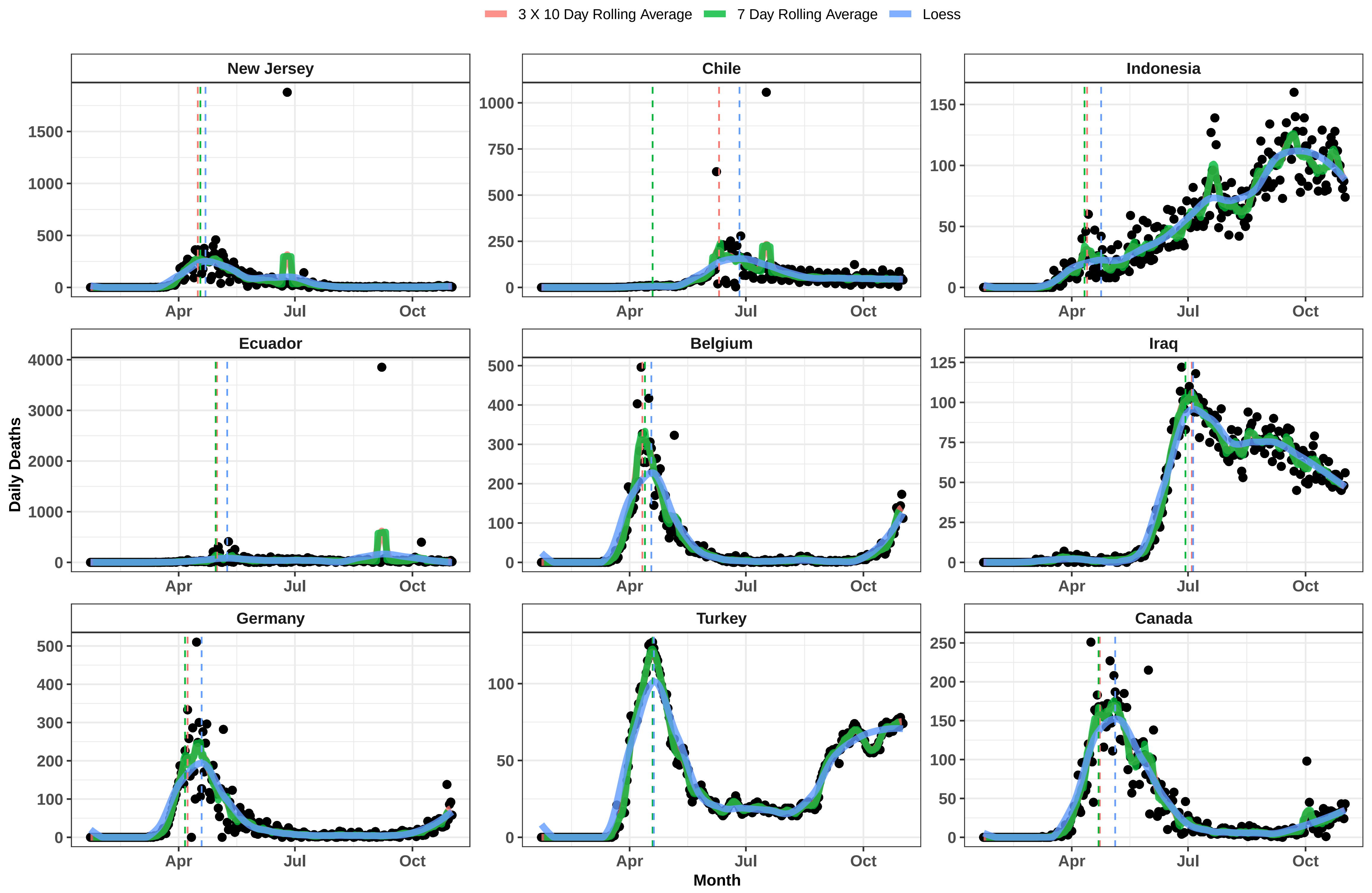

3 X 10 Day Rolling Average
7 Day Rolling Average
Loess
New Jersey
Chile
Indonesia
Ecuador
Belgium
Iraq
Germany
Turkey
Canada
Daily Deaths
Month
Apr
Jul
Oct

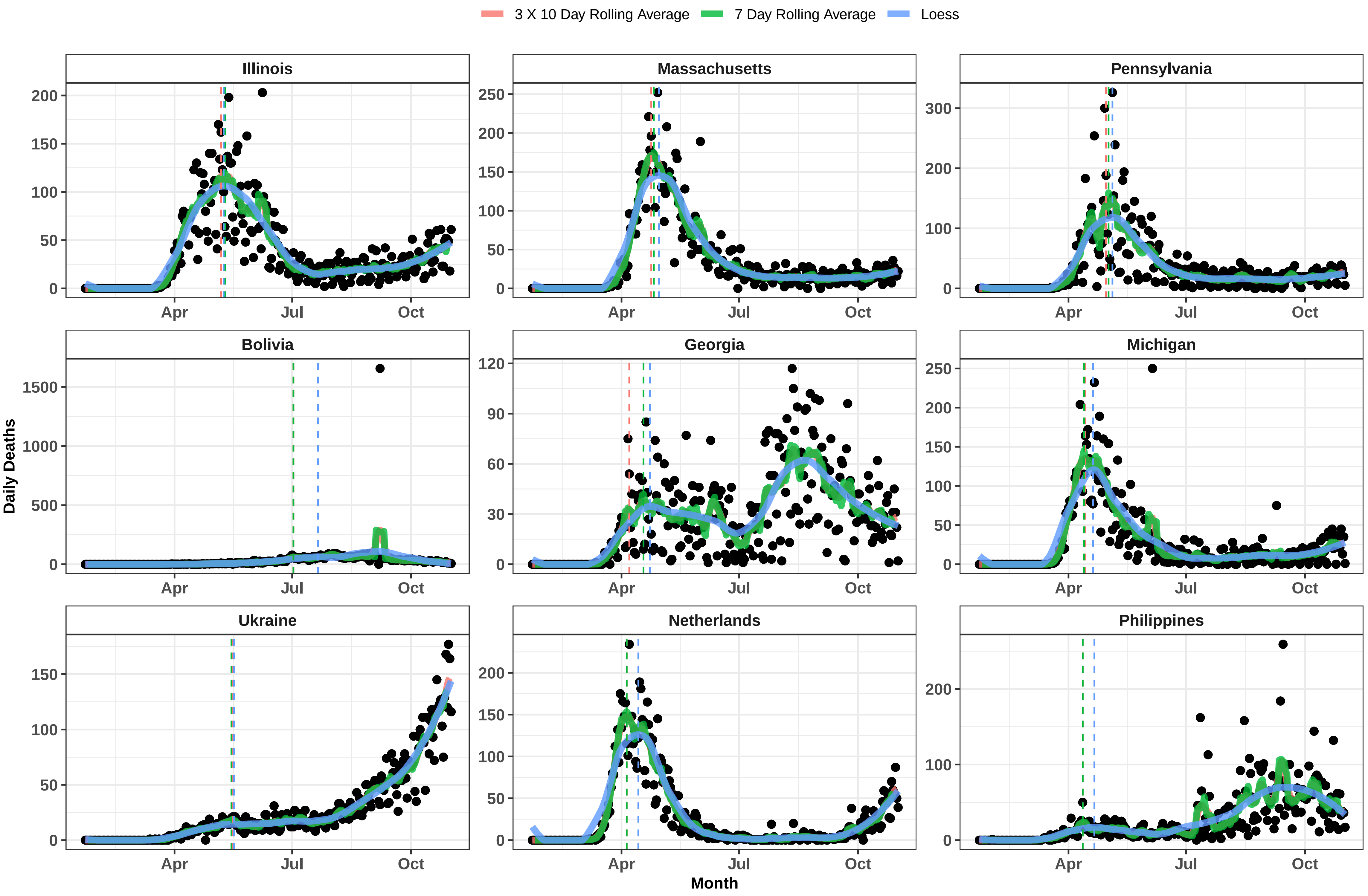

3 X 10 Day Rolling Average
7 Day Rolling Average
Loess
Illinois
Massachusetts
Pennsylvania
Bolivia
Georgia
Michigan
Ukraine
Netherlands
Philippines
Daily Deaths
Month
Apr
Jul
Oct

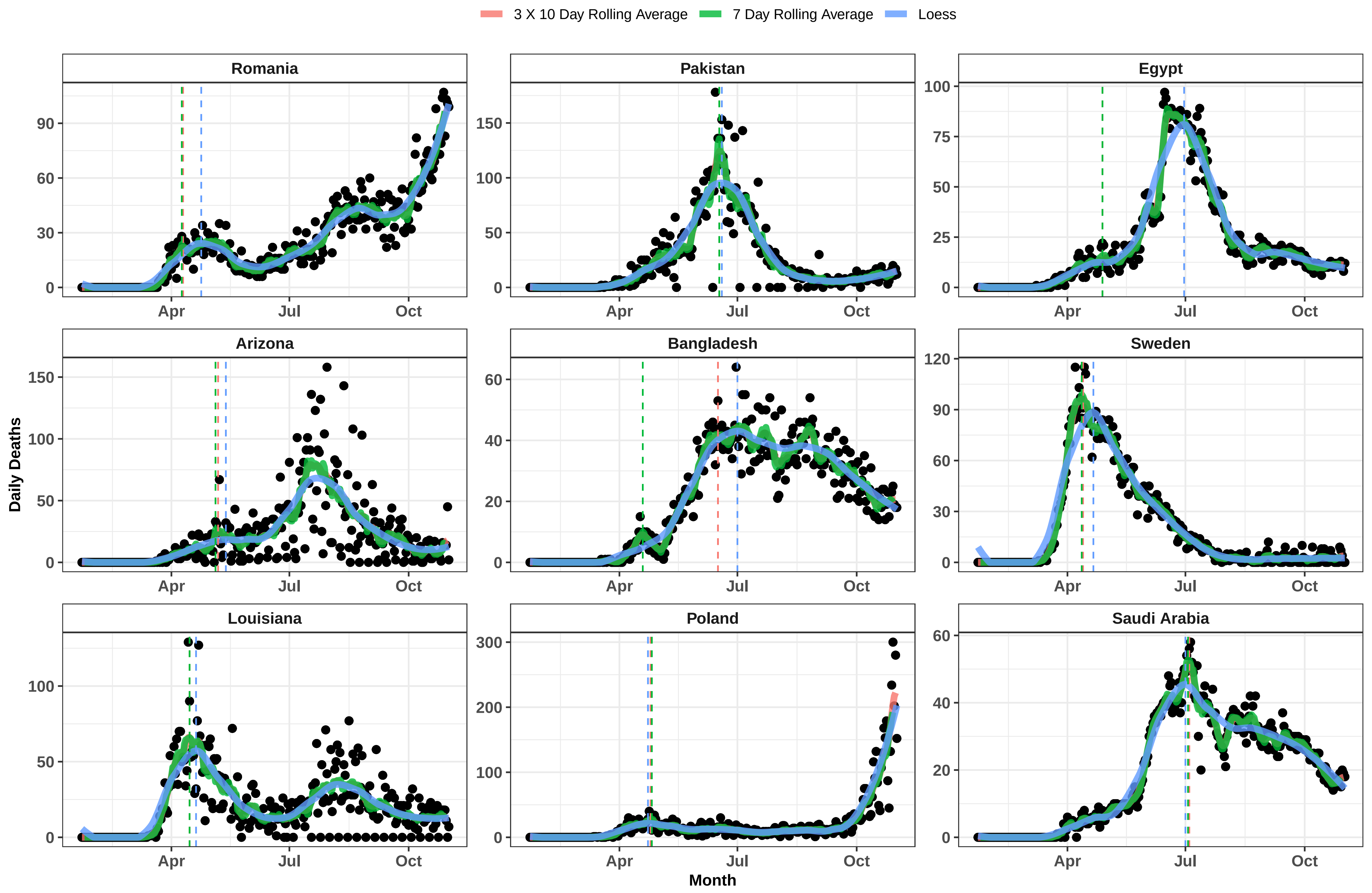

3 X 10 Day Rolling Average
7 Day Rolling Average
Loess
Romania
Pakistan
Egypt
Arizona
Bangladesh
Sweden
Louisiana
Poland
Saudi Arabia
Daily Deaths
Month
Apr
Jul
Oct

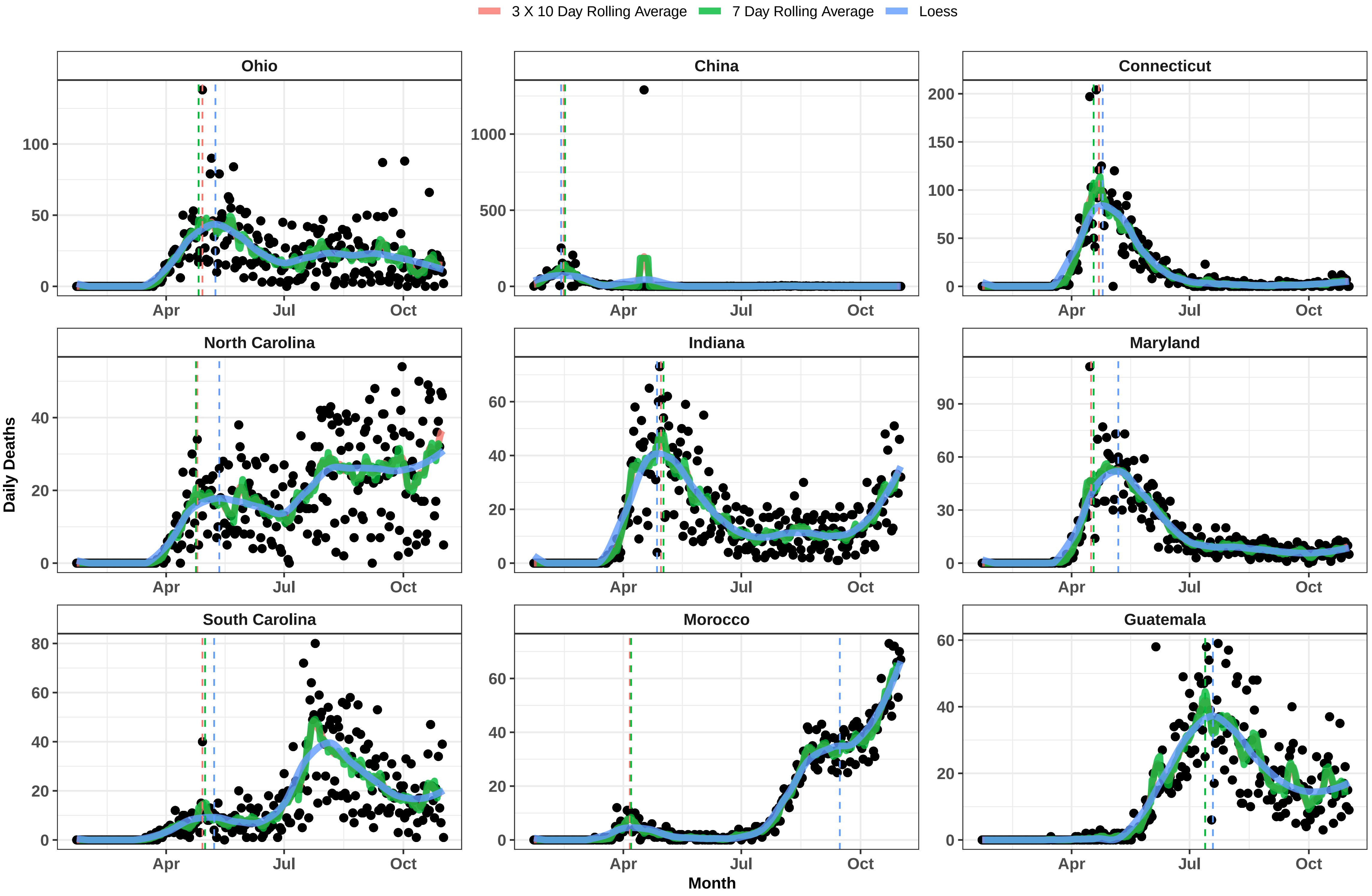

3 X 10 Day Rolling Average
7 Day Rolling Average
Loess
Ohio
China
Connecticut
North Carolina
Indiana
Maryland
South Carolina
Morocco
Guatemala
Daily Deaths
Month
Apr
Jul
Oct

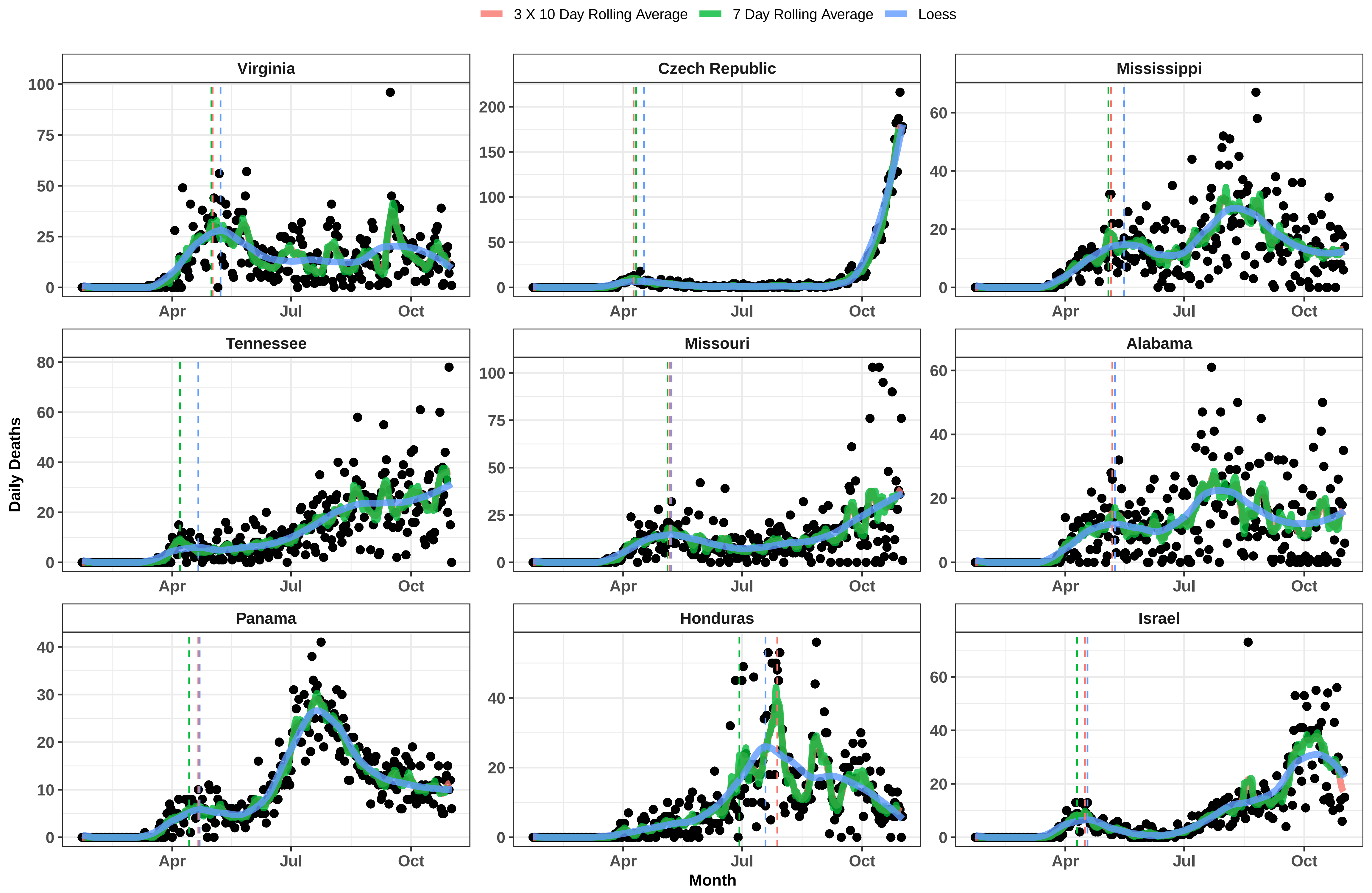

3 X 10 Day Rolling Average
7 Day Rolling Average
Loess
Virginia
Czech Republic
Mississippi
Tennessee
Missouri
Alabama
Panama
Honduras
Israel
Daily Deaths
Month
Apr
Jul
Oct

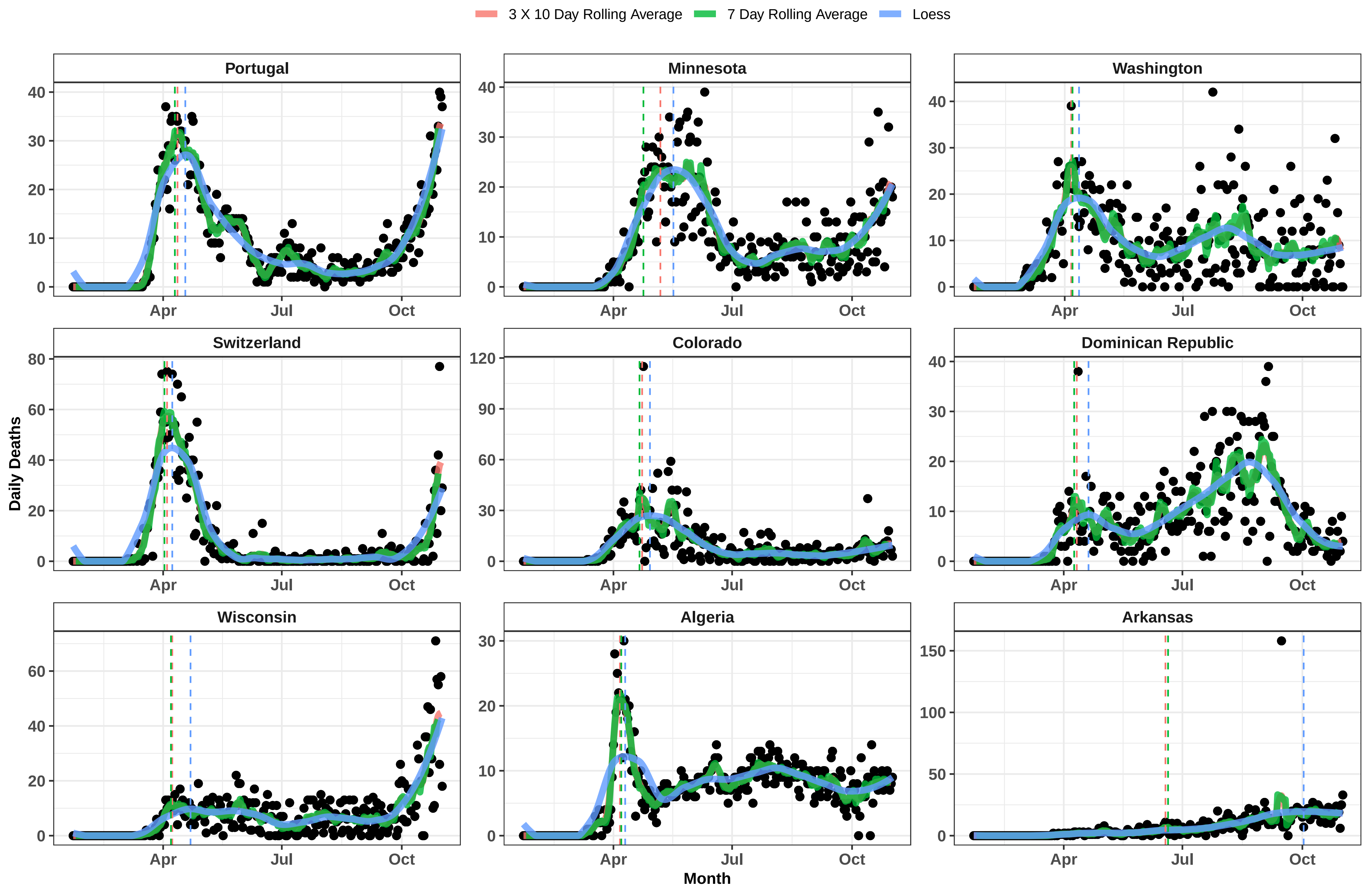

3 X 10 Day Rolling Average
7 Day Rolling Average
Loess
Portugal
Minnesota
Washington
Switzerland
Colorado
Dominican Republic
Wisconsin
Algeria
Arkansas
Daily Deaths
Month
Apr
Jul
Oct

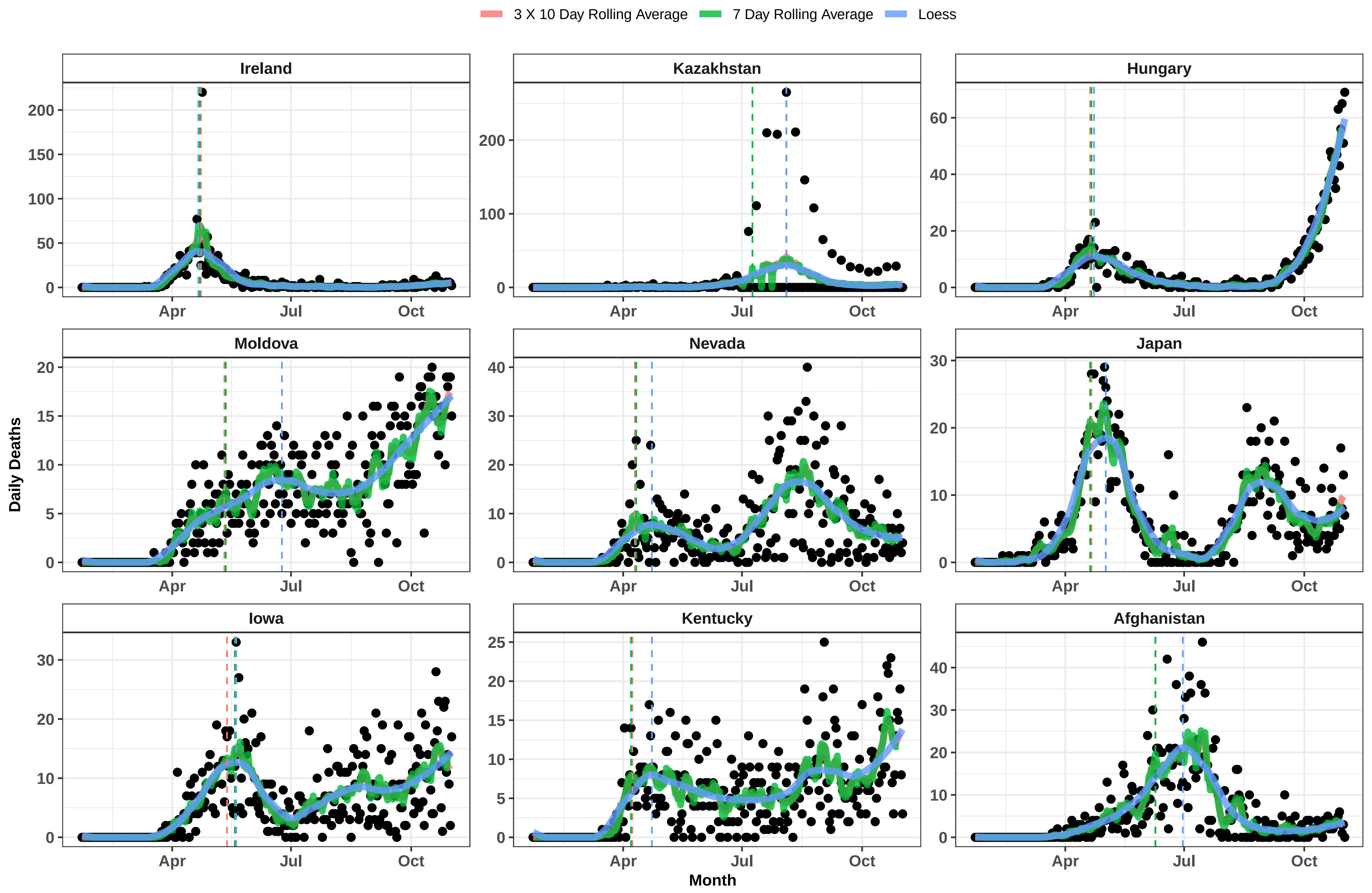
3 X 10 Day Rolling Average
7 Day Rolling Average
Loess
Ireland
Kazakhstan
Hungary
Moldova
Nevada
Japan
Iowa
Kentucky
Afghanistan
Daily Deaths
Month
Apr
Jul
Oct

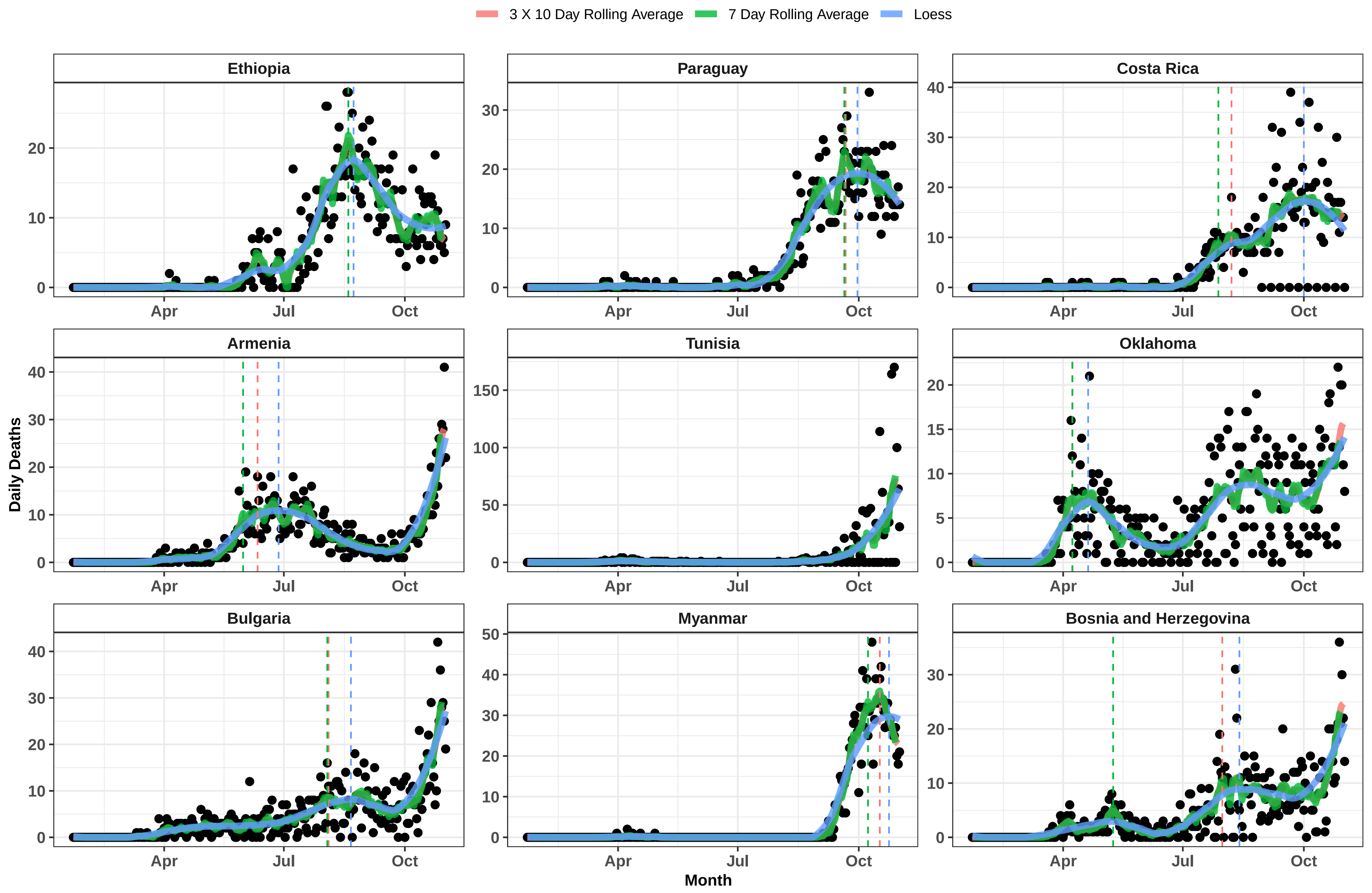
3 X 10 Day Rolling Average
7 Day Rolling Average
Loess
Ethiopia
Paraguay
Costa Rica
Armenia
Tunisia
Oklahoma
Bulgaria
Myanmar
Bosnia and Herzegovina
Daily Deaths
Month
Apr
Jul
Oct

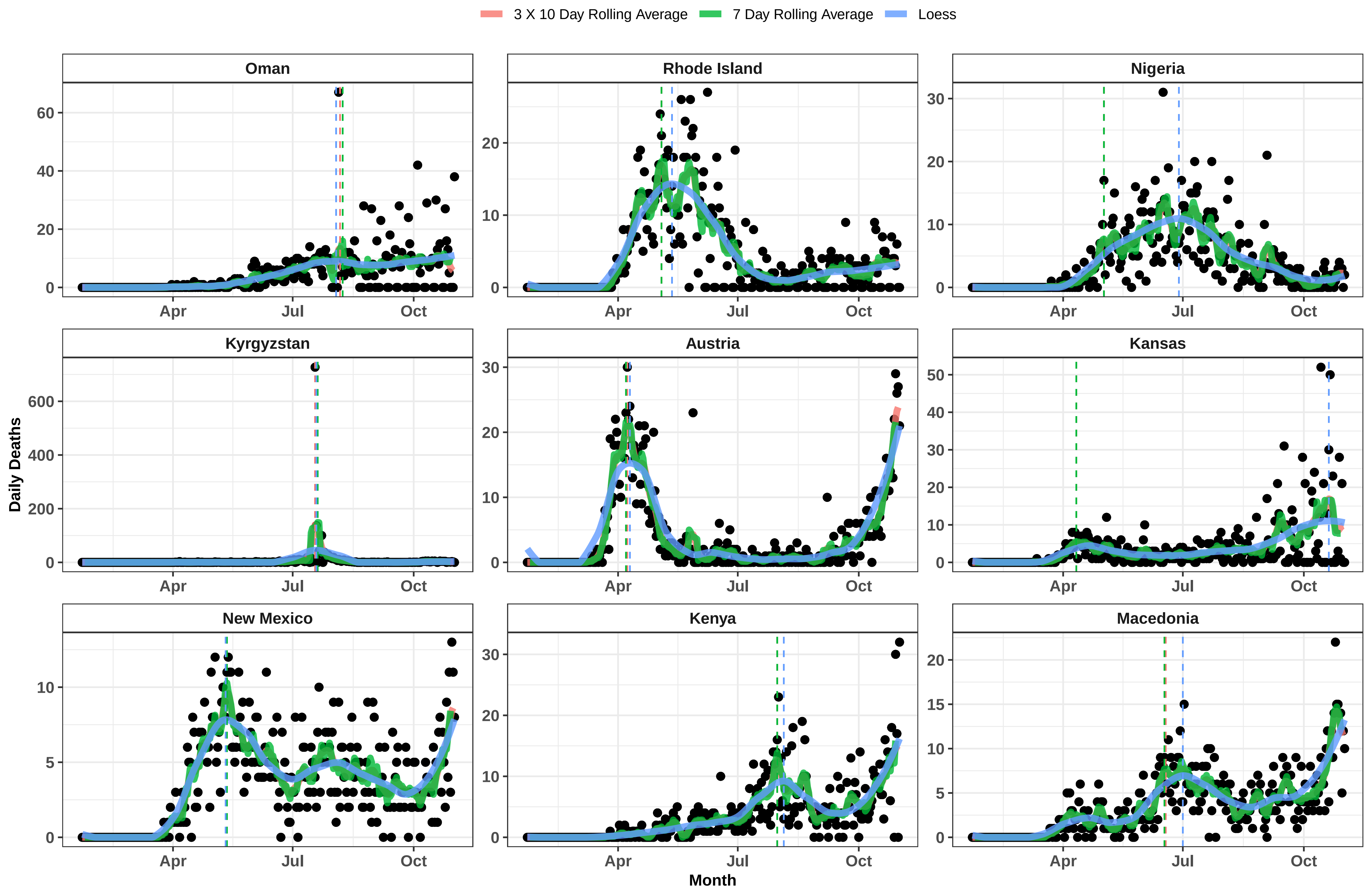

3 X 10 Day Rolling Average
7 Day Rolling Average
Loess
Oman
Rhode Island
Nigeria
Kyrgyzstan
Austria
Kansas
New Mexico
Kenya
Macedonia
Daily Deaths
Month
Apr
Jul
Oct

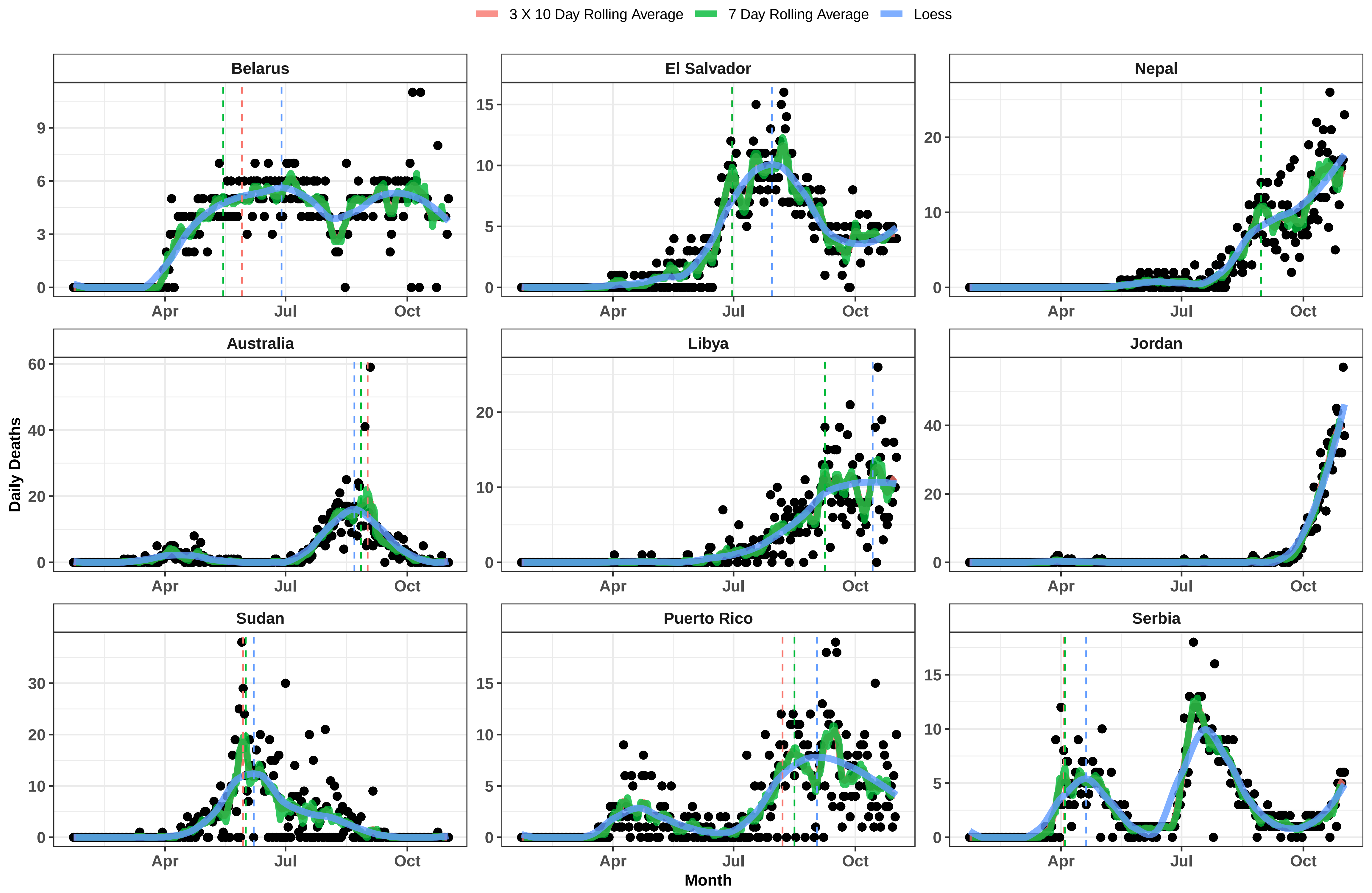

3 X 10 Day Rolling Average
7 Day Rolling Average
Loess
Belarus
El Salvador
Nepal
Australia
Libya
Jordan
Sudan
Puerto Rico
Serbia
Daily Deaths
Month
Apr
Jul
Oct

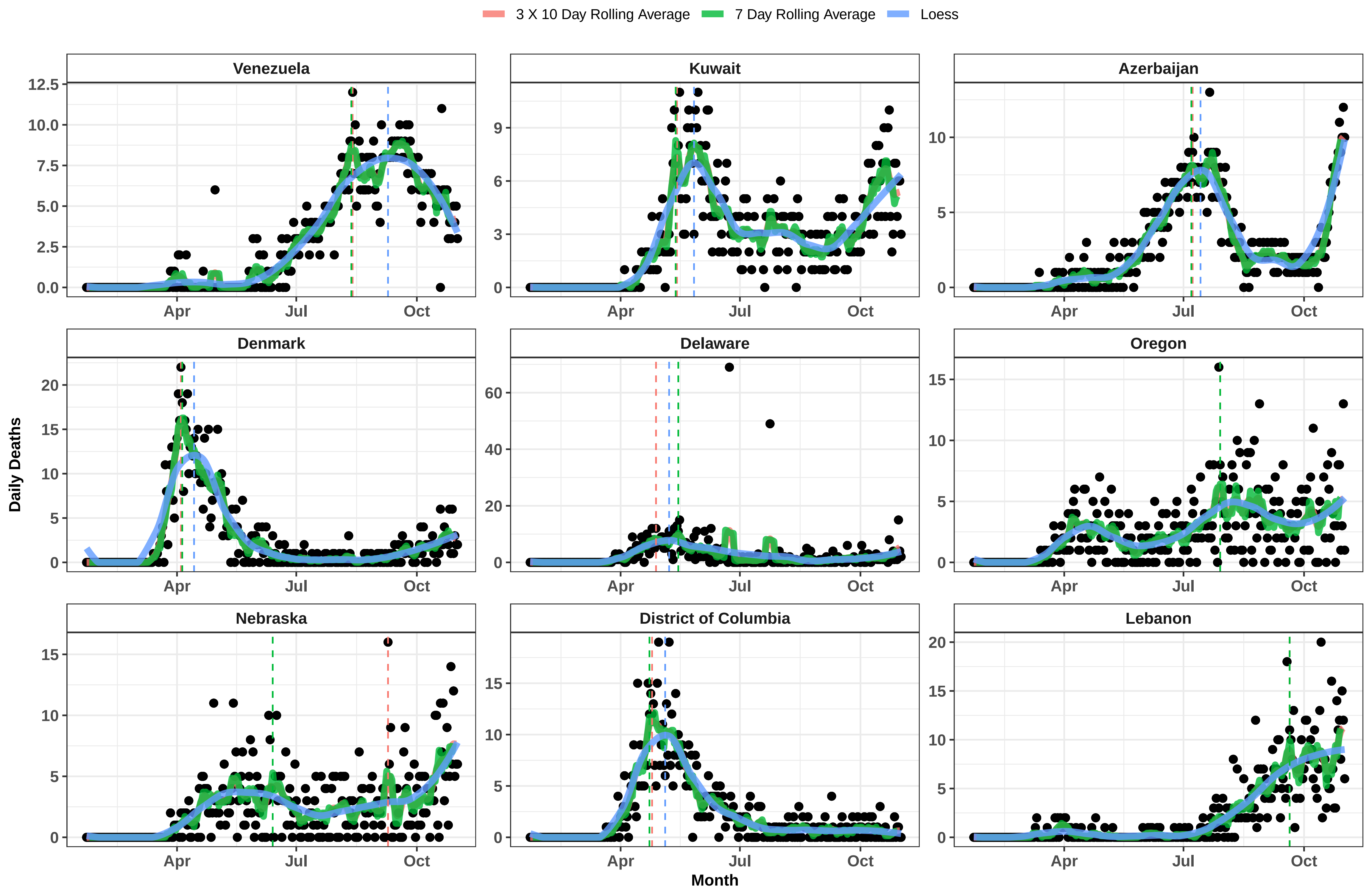

3 X 10 Day Rolling Average
7 Day Rolling Average
Loess
Venezuela
Kuwait
Azerbaijan
Denmark
Delaware
Oregon
Nebraska
District of Columbia
Lebanon
Daily Deaths
Month
Apr
Jul
Oct

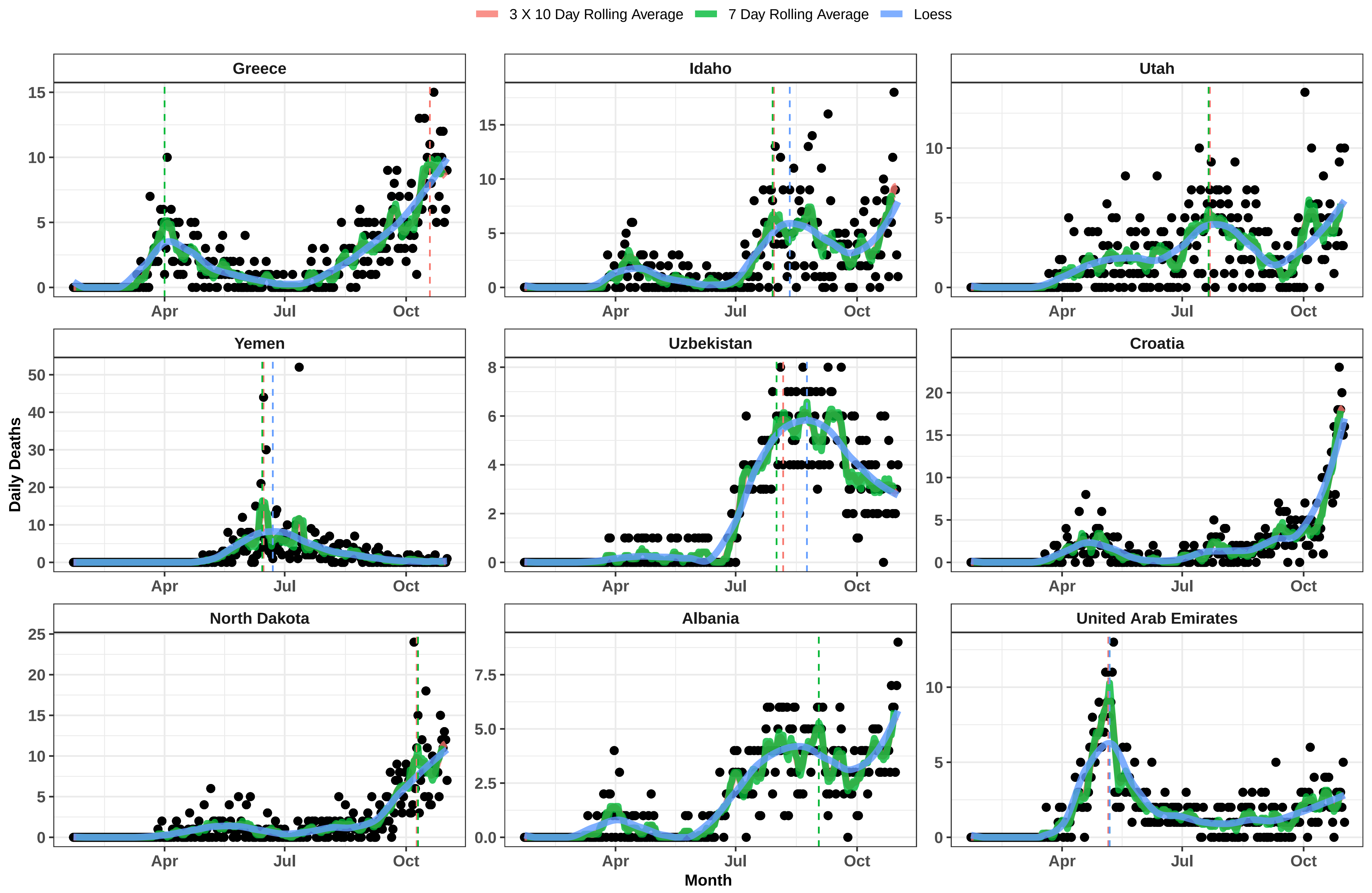

3 X 10 Day Rolling Average
7 Day Rolling Average
Loess
Greece
Idaho
Utah
Yemen
Uzbekistan
Croatia
North Dakota
Albania
United Arab Emirates
Daily Deaths
Month
Apr
Jul
Oct

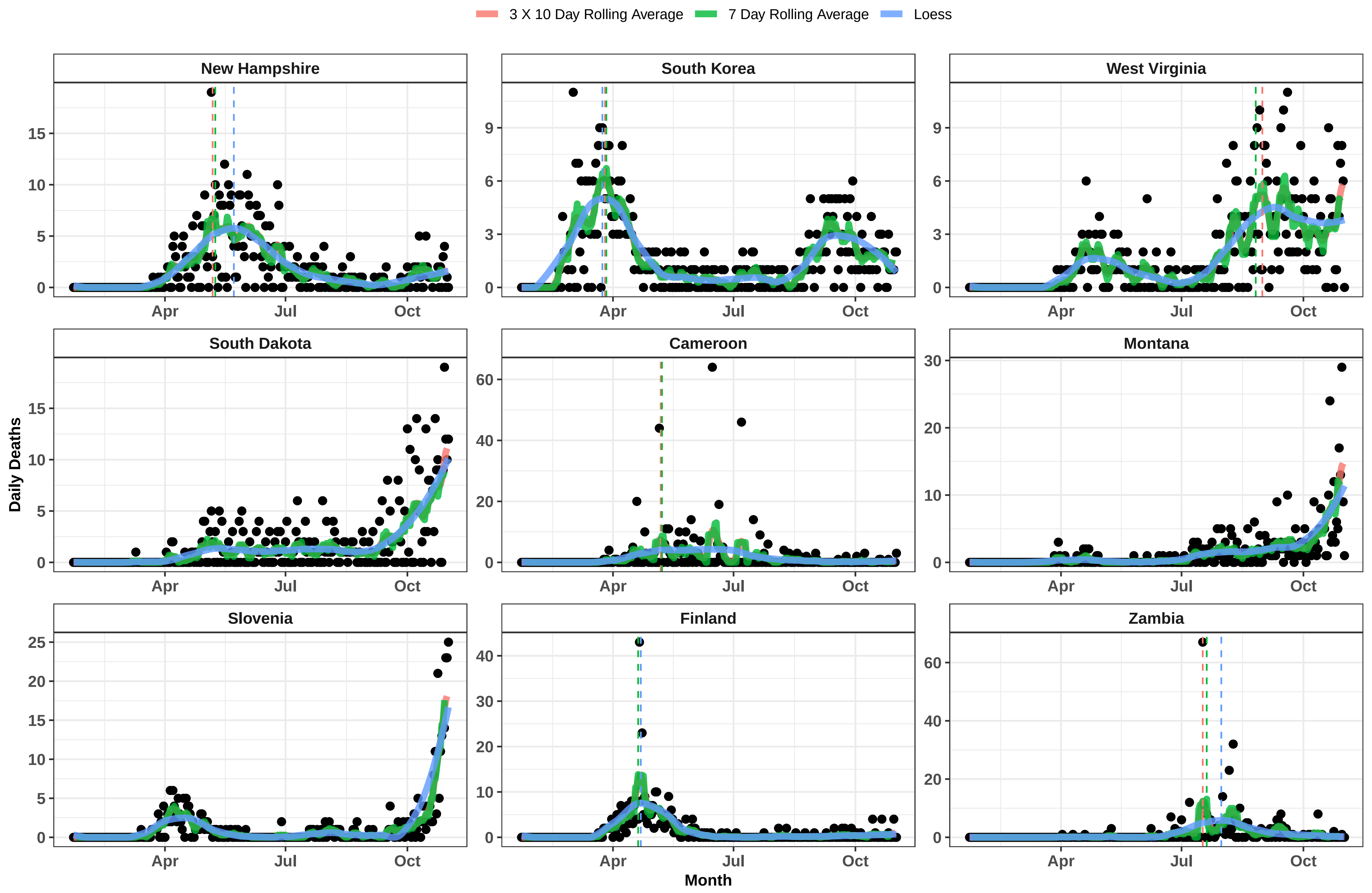

3 X 10 Day Rolling Average
7 Day Rolling Average
Loess
New Hampshire
South Korea
West Virginia
South Dakota
Cameroon
Montana
Slovenia
Finland
Zambia
Daily Deaths
Month
Apr
Jul
Oct

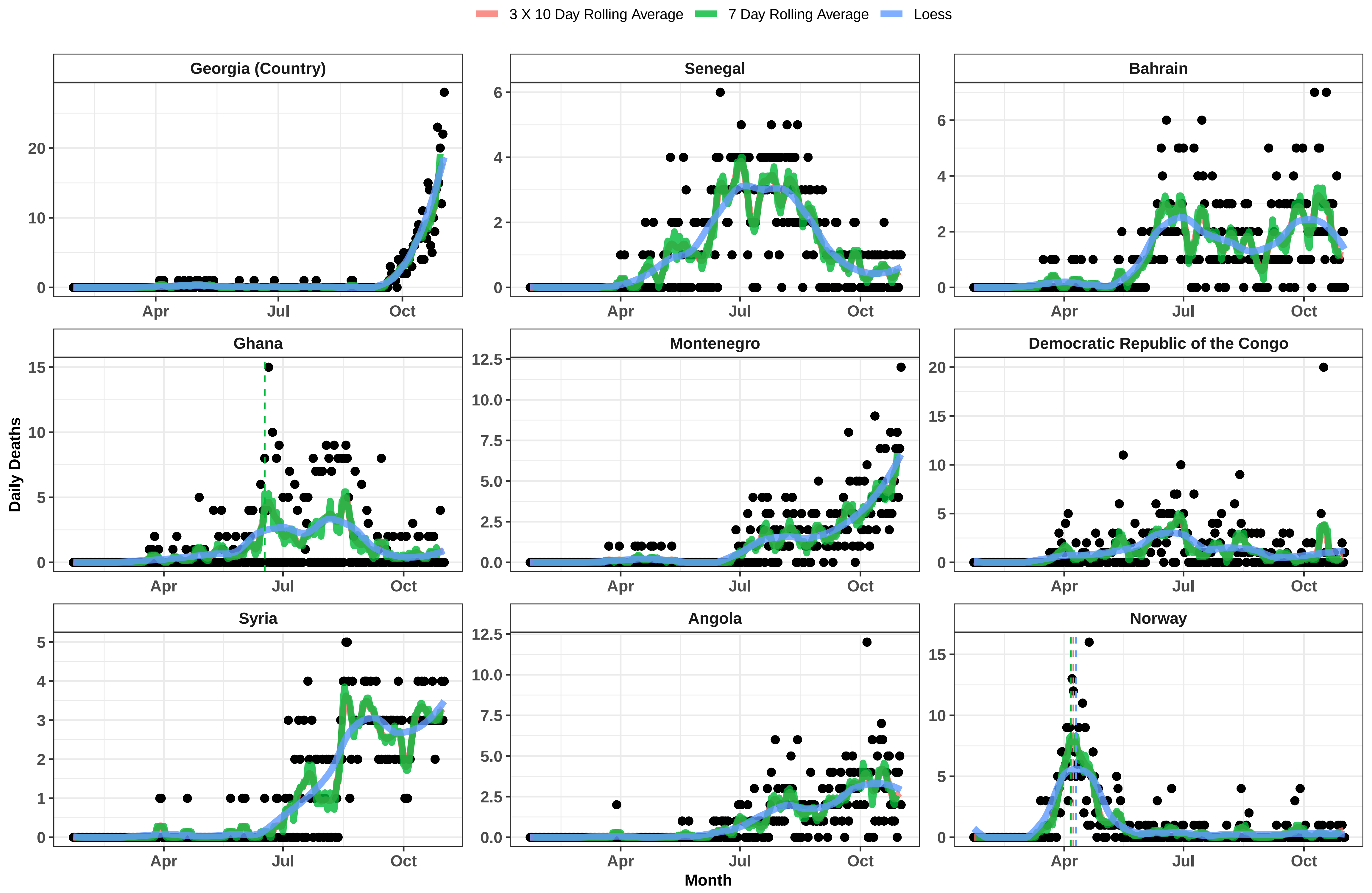

3 X 10 Day Rolling Average
7 Day Rolling Average
Loess
Georgia (Country)
Senegal
Bahrain
Ghana
Montenegro
Democratic Republic of the Congo
Syria
Angola
Norway
Daily Deaths
Month
Apr
Jul
Oct

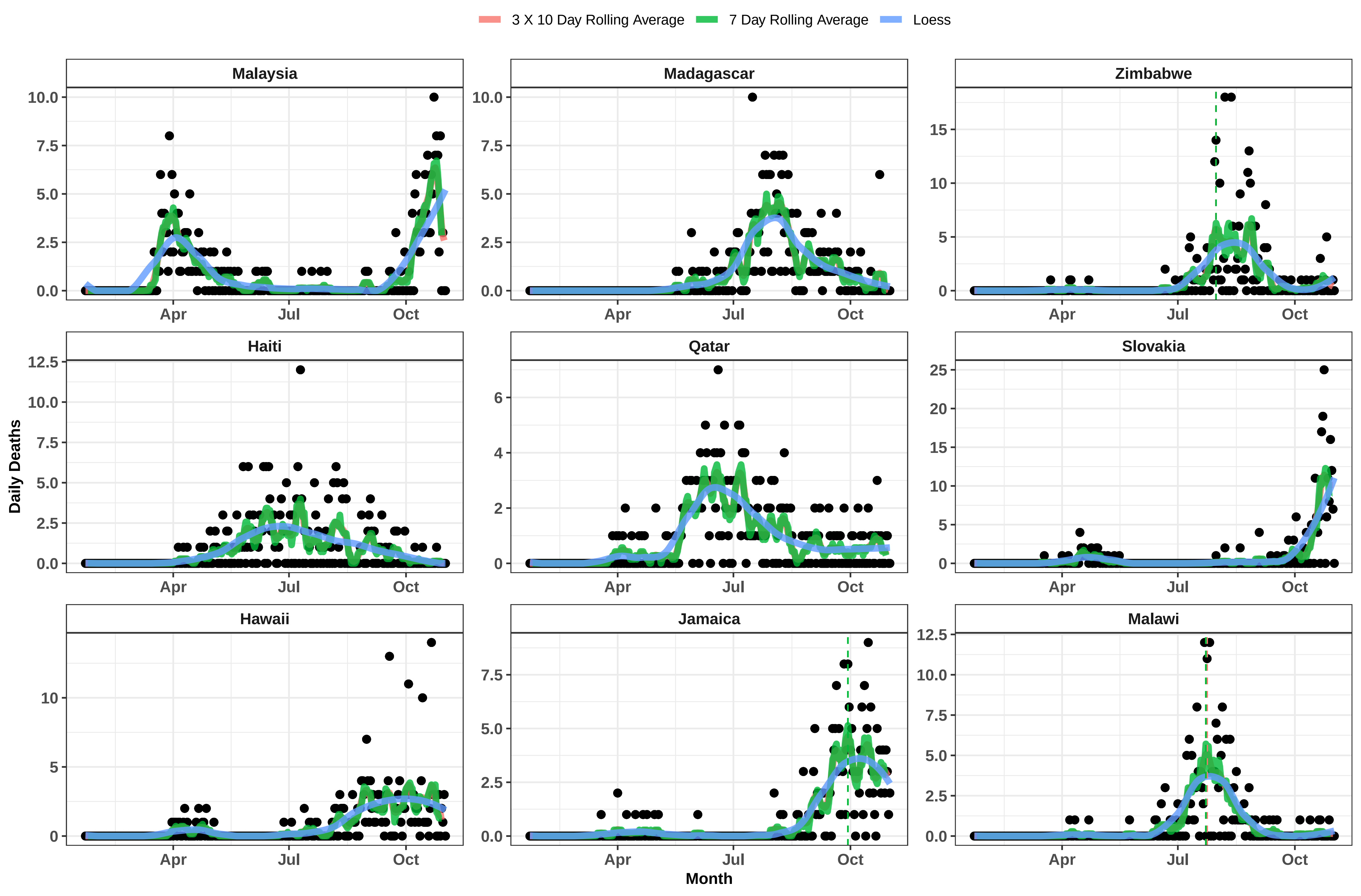

3 X 10 Day Rolling Average
7 Day Rolling Average
Loess
Malaysia
Madagascar
Zimbabwe
Haiti
Qatar
Slovakia
Hawaii
Jamaica
Malawi
Daily Deaths
Month
Apr
Jul
Oct

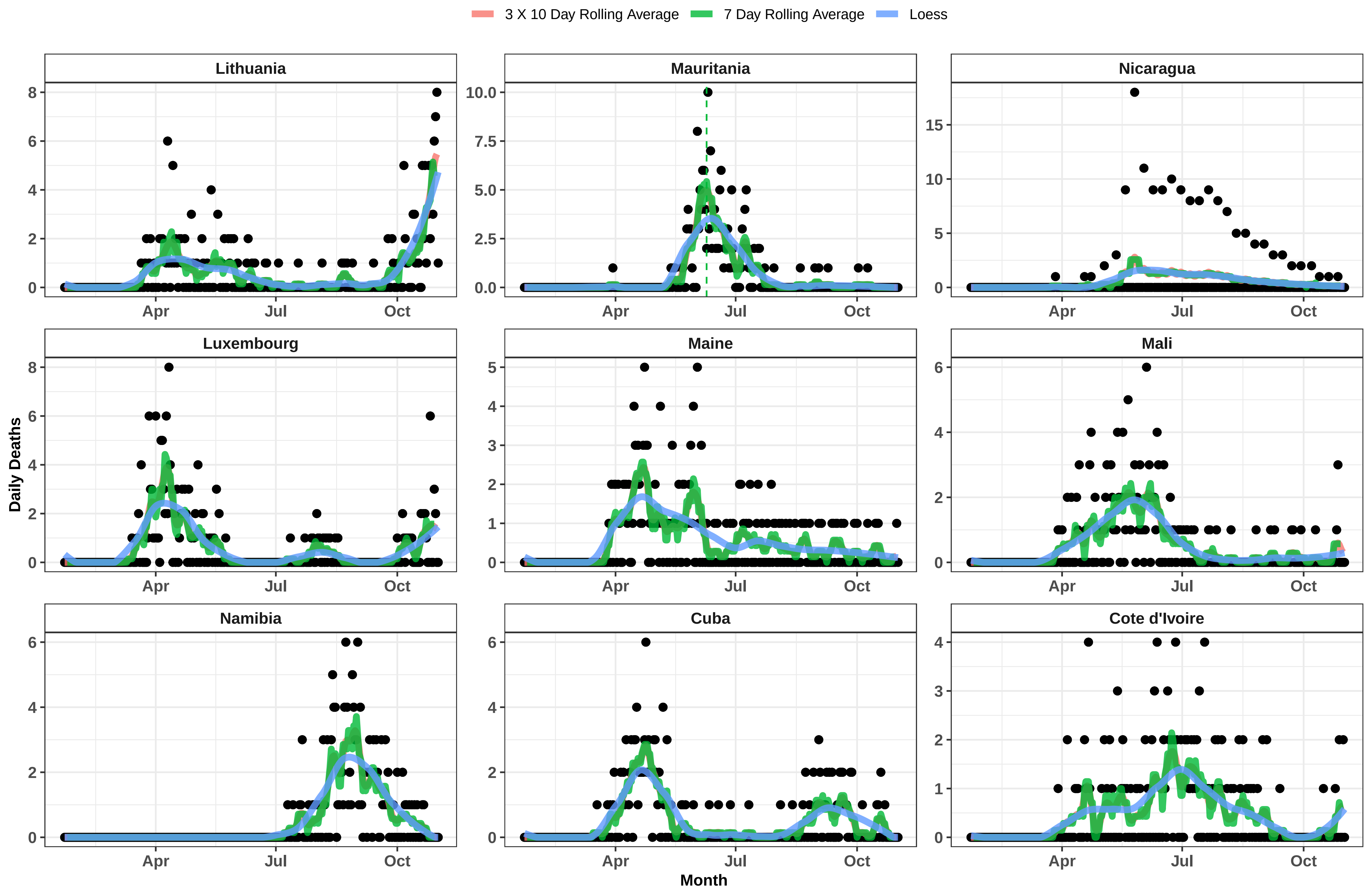

3 X 10 Day Rolling Average
7 Day Rolling Average
Loess
Lithuania
Mauritania
Nicaragua
Luxembourg
Maine
Mali
Namibia
Cuba
Cote d'Ivoire
Daily Deaths
Month
Apr
Jul
Oct

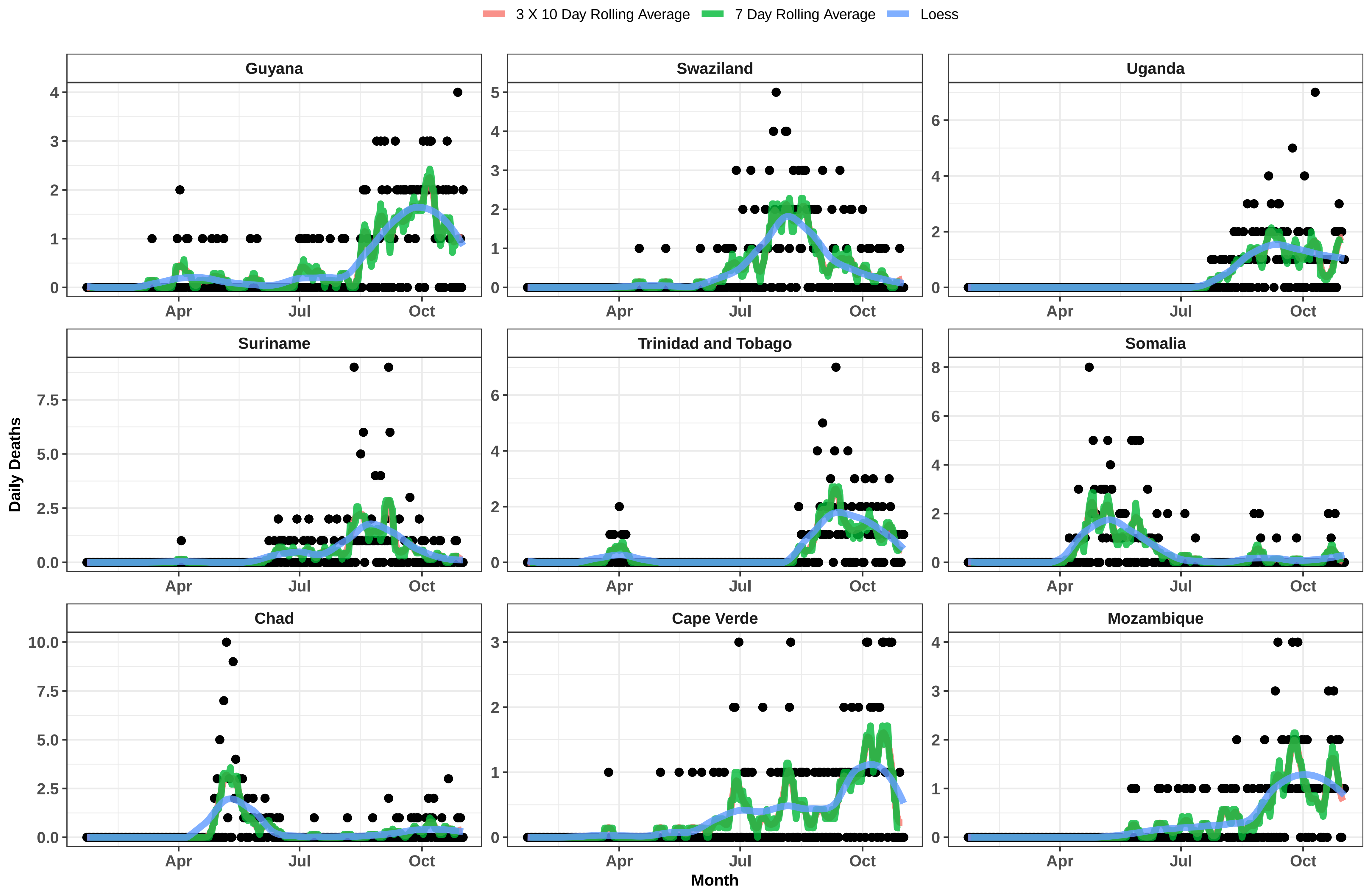

3 X 10 Day Rolling Average
7 Day Rolling Average
Loess
Guyana
Swaziland
Uganda
Suriname
Trinidad and Tobago
Somalia
Chad
Cape Verde
Mozambique
Daily Deaths
Month
Apr
Jul
Oct

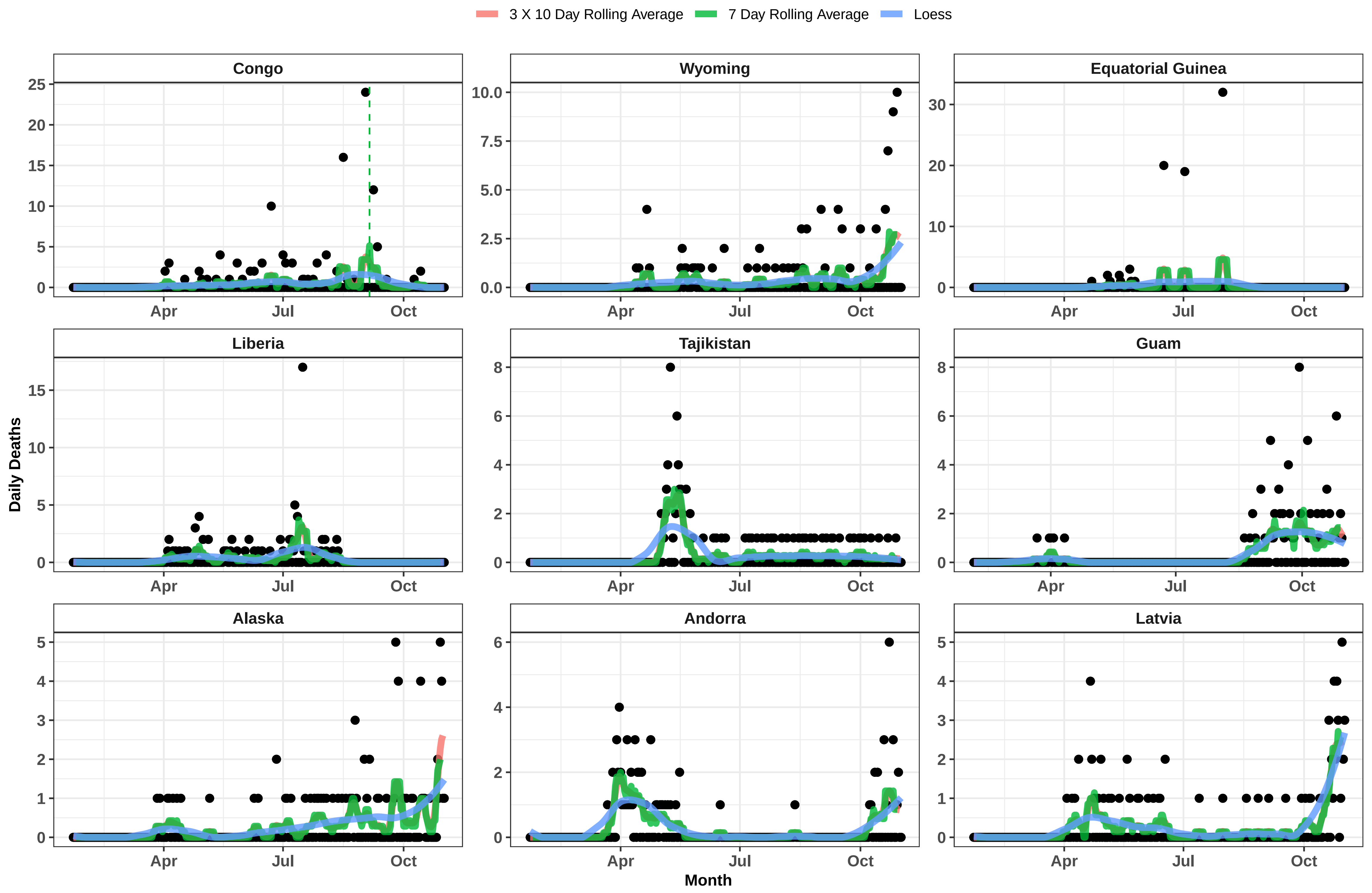

3 X 10 Day Rolling Average
7 Day Rolling Average
Loess
Congo
Wyoming
Equatorial Guinea
Liberia
Tajikistan
Guam
Alaska
Andorra
Latvia
Daily Deaths
Month
Apr
Jul
Oct

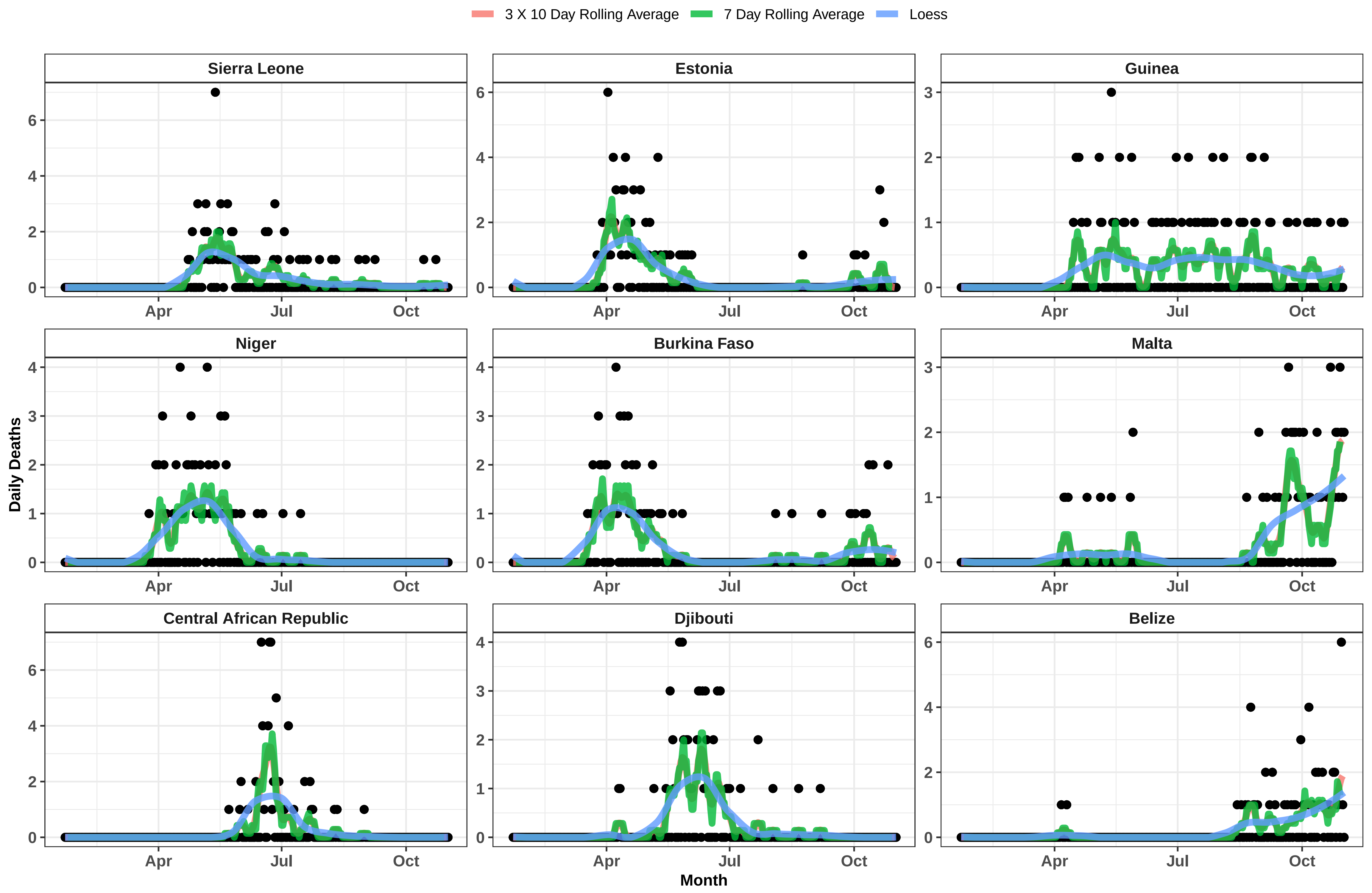
3 X 10 Day Rolling Average
7 Day Rolling Average
Loess
Sierra Leone
Estonia
Guinea
Niger
Burkina Faso
Malta
Central African Republic
Djibouti
Belize
Daily Deaths
Month
Apr
Jul
Oct

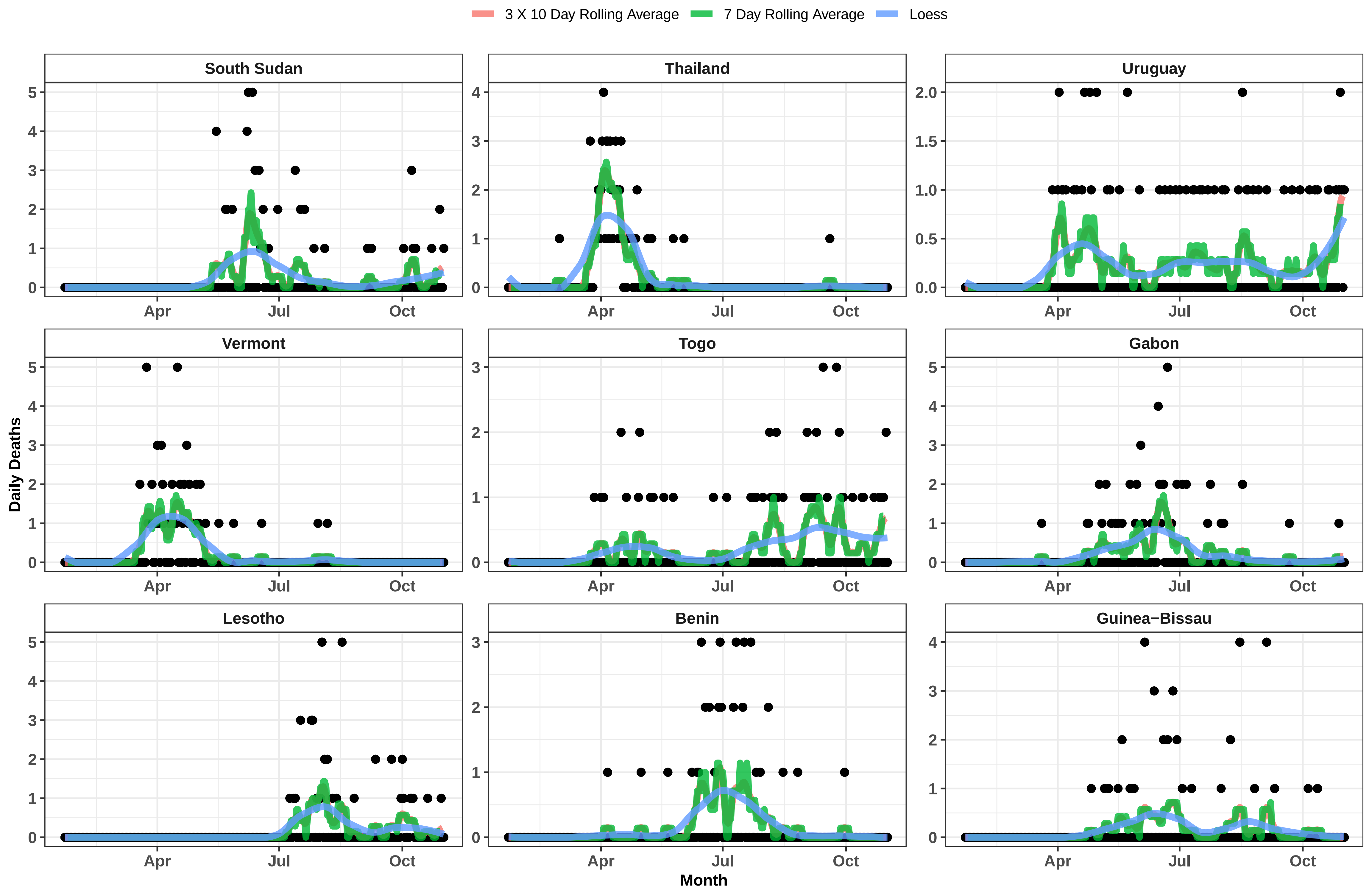

3 X 10 Day Rolling Average
7 Day Rolling Average
Loess
South Sudan
Thailand
Uruguay
Vermont
Togo
Gabon
Lesotho
Benin
Guinea−Bissau
Daily Deaths
Month
Apr
Jul
Oct

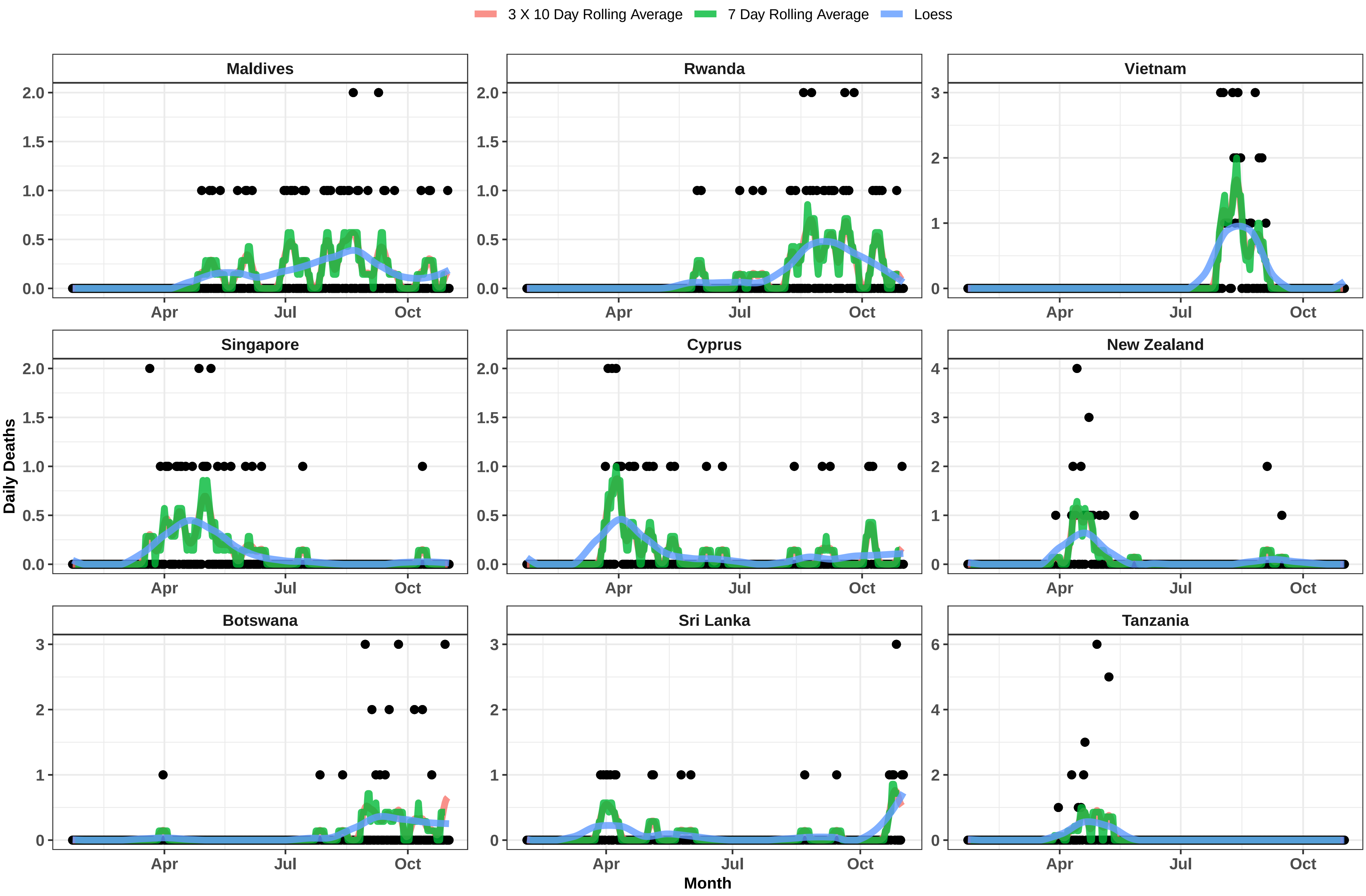

3 X 10 Day Rolling Average
7 Day Rolling Average
Loess
Maldives
Rwanda
Vietnam
Singapore
Cyprus
New Zealand
Botswana
Sri Lanka
Tanzania
Daily Deaths
Month
Apr
Jul
Oct

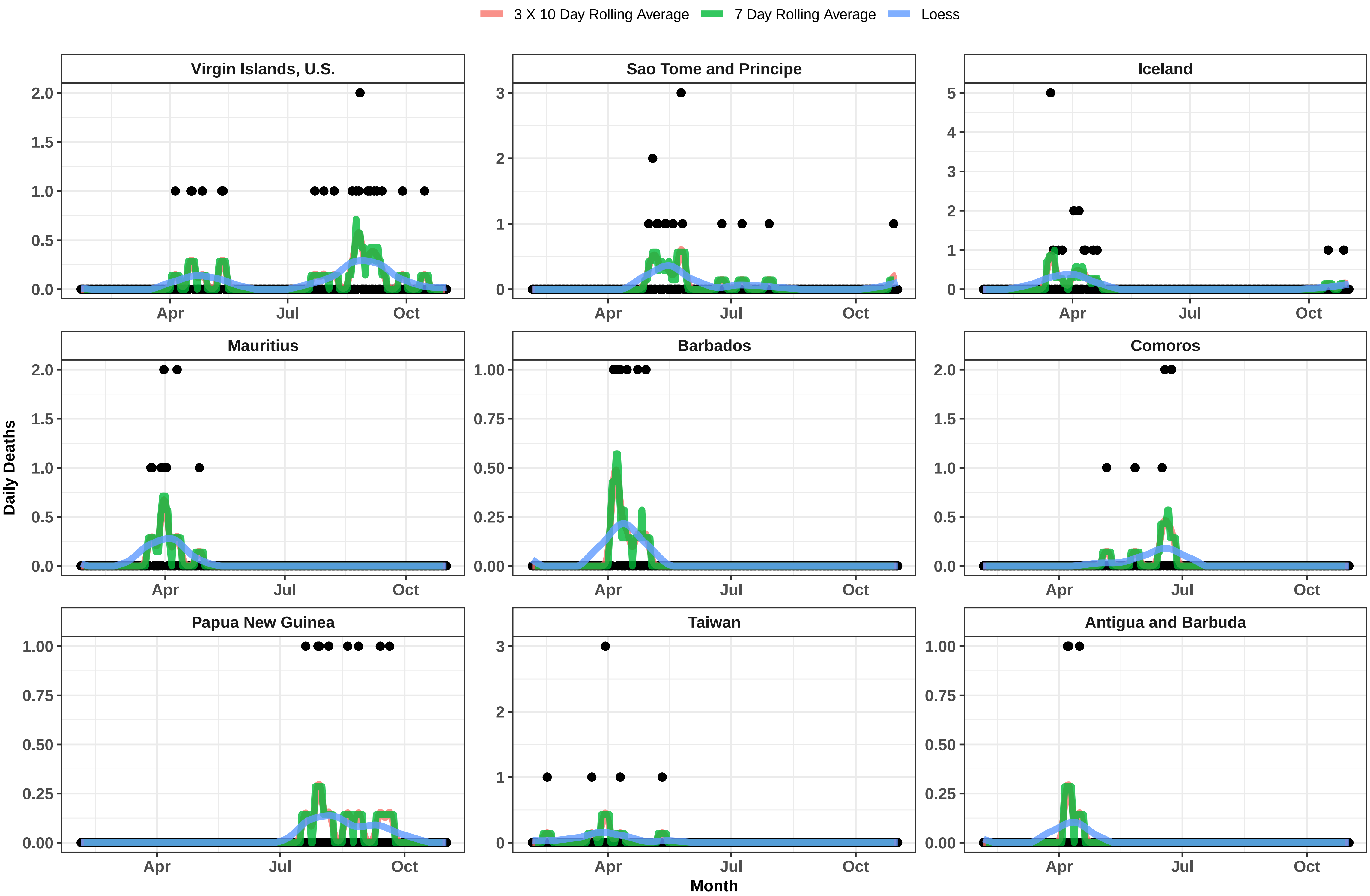

3 X 10 Day Rolling Average
7 Day Rolling Average
Loess
Virgin Islands, U.S.
Sao Tome and Principe
Iceland
Mauritius
Barbados
Comoros
Papua New Guinea
Taiwan
Antigua and Barbuda
Daily Deaths
Month
Apr
Jul
Oct

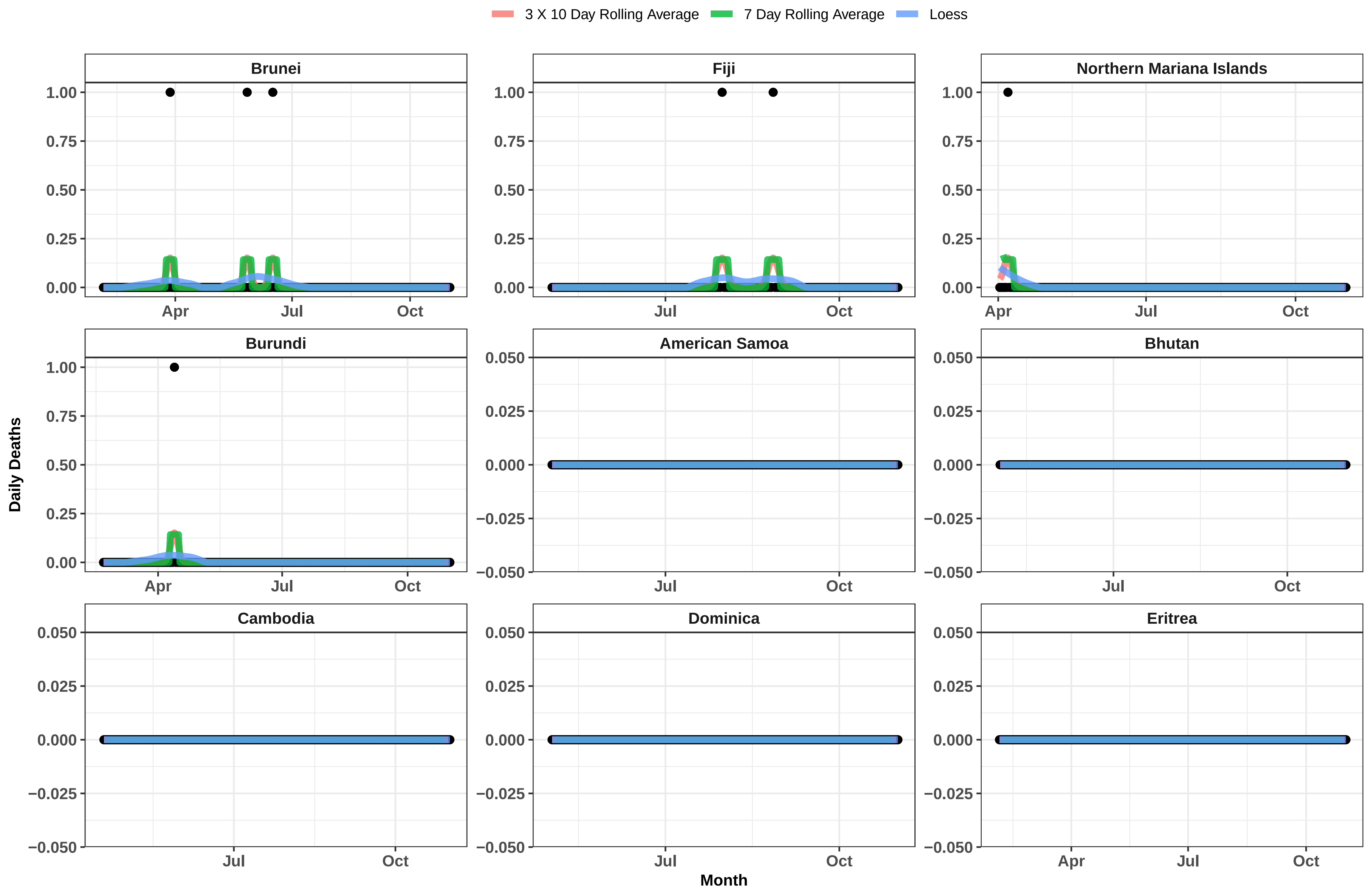

3 X 10 Day Rolling Average
7 Day Rolling Average
Loess
Brunei
Fiji
Northern Mariana Islands
Burundi
American Samoa
Bhutan
Cambodia
Dominica
Eritrea
1.00
0.75
0.50
0.25
0.00
0.050
0.025
0.000
−0.025
−0.050
Apr
Jul
Oct
Daily Deaths
Month

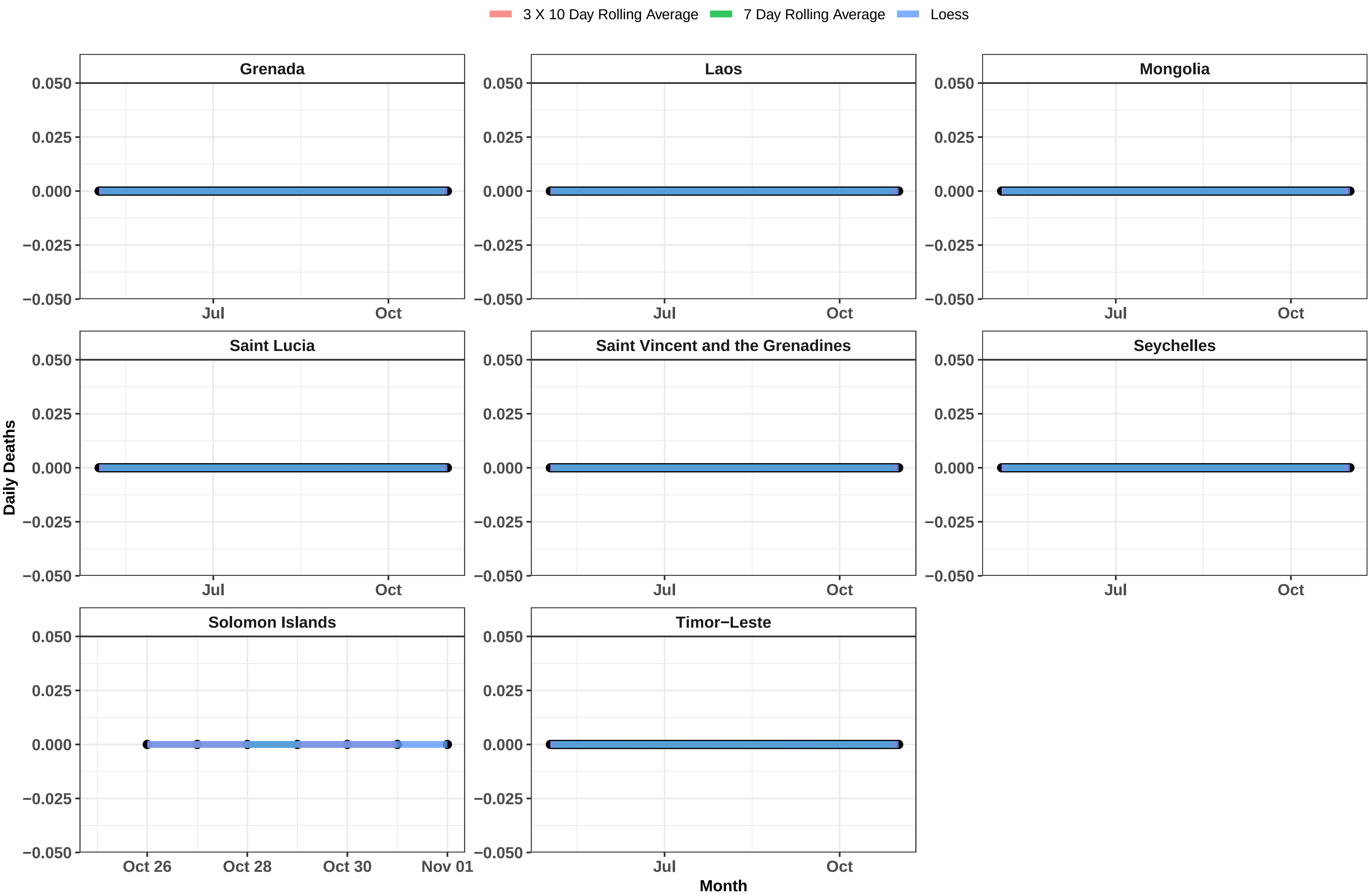
3 X 10 Day Rolling Average
7 Day Rolling Average
Loess
Grenada
Laos
Mongolia
Saint Lucia
Saint Vincent and the Grenadines
Seychelles
Solomon Islands
Timor−Leste
0.050
0.025
0.000
−0.025
−0.050
Jul
Oct
Oct 26
Oct 28
Oct 30
Nov 01
Daily Deaths
Month
